# Scoring respondents on a semantic frame of reference identifies a group that is burdened across many questionnaires and reaches care less easily

**DOI:** 10.64898/2026.09.03.26362158

**Authors:** Manu Aggarwal, Vipul Periwal

## Abstract

Cohorts and health systems give people batteries of questionnaires on symptoms and life circumstances, and a person’s difficulties can show up in several at once. Finding who is faring badly means scoring the whole battery. Methods that span a battery estimate its structure from responses rather than from what the questions mean, and work on question wording does not score the people who answered. Here we build a semantic frame of reference from published question text alone, and score respondents on it. Language models group 577 questions from 50 questionnaires into components fixed before any cohort answers, each interpretable because it rests on a few questions. We place any battery on these components from its question text and score each respondent by how unusual their position is. On an All of Us battery answered by 22,332 participants, we find a group burdened across many questionnaires. In health records and access surveys that never entered the scoring, this group reaches care less easily than the rest of the cohort, and being more distressed does not explain the difference. Built once, the frame scores any battery, and each score shows which questions produced it.

## 1. Introduction

Research cohorts and health systems ask people about symptoms and about the circumstances of their lives—food insecurity, discrimination, neighborhood safety, interpersonal violence—and record the answers in a *battery* [1]. A battery is a set of published *instruments*, and an instrument is a set of questions, or *items*. Finding who is faring badly means asking how a person stands across everything they were asked, the symptoms and the circumstances together.

The established tool for flagging a person from their answers is a validated threshold, a fixed rule applied to one instrument. That leaves a battery short in two ways. Many of its instruments have no threshold at all. Those that record social exposures usually have none, so they describe a person’s circumstances but cannot flag anyone. Perceived discrimination, for example, is associated with worse health across many outcomes [2], yet no instrument that records it has a validated threshold. And the thresholds that do exist are not combined, so a person who is moderately burdened on several instruments meets none, even though staying below one threshold does not by itself mean faring well [3]. No threshold, then, says how a person stands across everything they were asked.

Two approaches do span a whole battery, and both estimate its structure—how its questions relate to one another—from the responses. First, a factor model estimates how strongly the responses to each item track a few underlying dimensions [4, 5]. Second, a network estimates how strongly the responses to two items are associated once the rest are accounted for [6]. But that structure depends on the sample and needs enough respondents to be stable [7, 8], so it does not exist before a cohort has answered. It does not say whether two questions go together because they ask about the same thing or because their answers happen to covary in that sample. A screen for the whole battery needs a structure that spans instruments, is fixed before any cohort answers, and can be interpreted by what the questions ask.

Such a structure can come from what the questions mean rather than from how the answers covary. A language model places a question by what it asks about, so questions asking about the same thing sit near one another [9]. Any set of items is therefore grouped by semantic content without curation and without responses. The closest prior work embeds items, instruments and construct labels in one vector space across thousands of published items, and the embeddings detect where a label and the content under it come apart [10]. The two do come apart in practice. Across seven common depression instruments, 40% of the symptoms they cover appear in only one of the seven [11]. Two items answered under one label can therefore ask about different things, and two items from unrelated instruments can ask about the same thing. Content also predicts how an item’s answers behave [12, 13], though not what any one person reports. None of that prior work scores the people who answered.

Here we build a semantic frame of reference from published question text alone. We assemble a pool of 577 published questions spanning 50 instruments and 54 documented constructs. We compress that pool into a small number of directions of content, which we call *components*. Each rests on a handful of questions rather than on all of them, its *core items* (Fig. 1a). For example, one groups questions about food insecurity and another questions about psychosis. That set of components is the *reference basis*. It exists before any cohort answers and belongs to no battery in particular. We then place any battery on it without manual curation, and neither the battery nor its instruments need to have been in the pool. Batteries given to different cohorts at different times are described on the same components, a first step toward comparing them with no shared respondents. Each participant’s answers become a position on those components, and a single number says how unusual that position is within the cohort (Fig. 1b). That number splits into one contribution per component, each interpretable through the few questions it rests on. The resulting screen reports the content that accounts for each statistical outlier.

**Figure 1:**
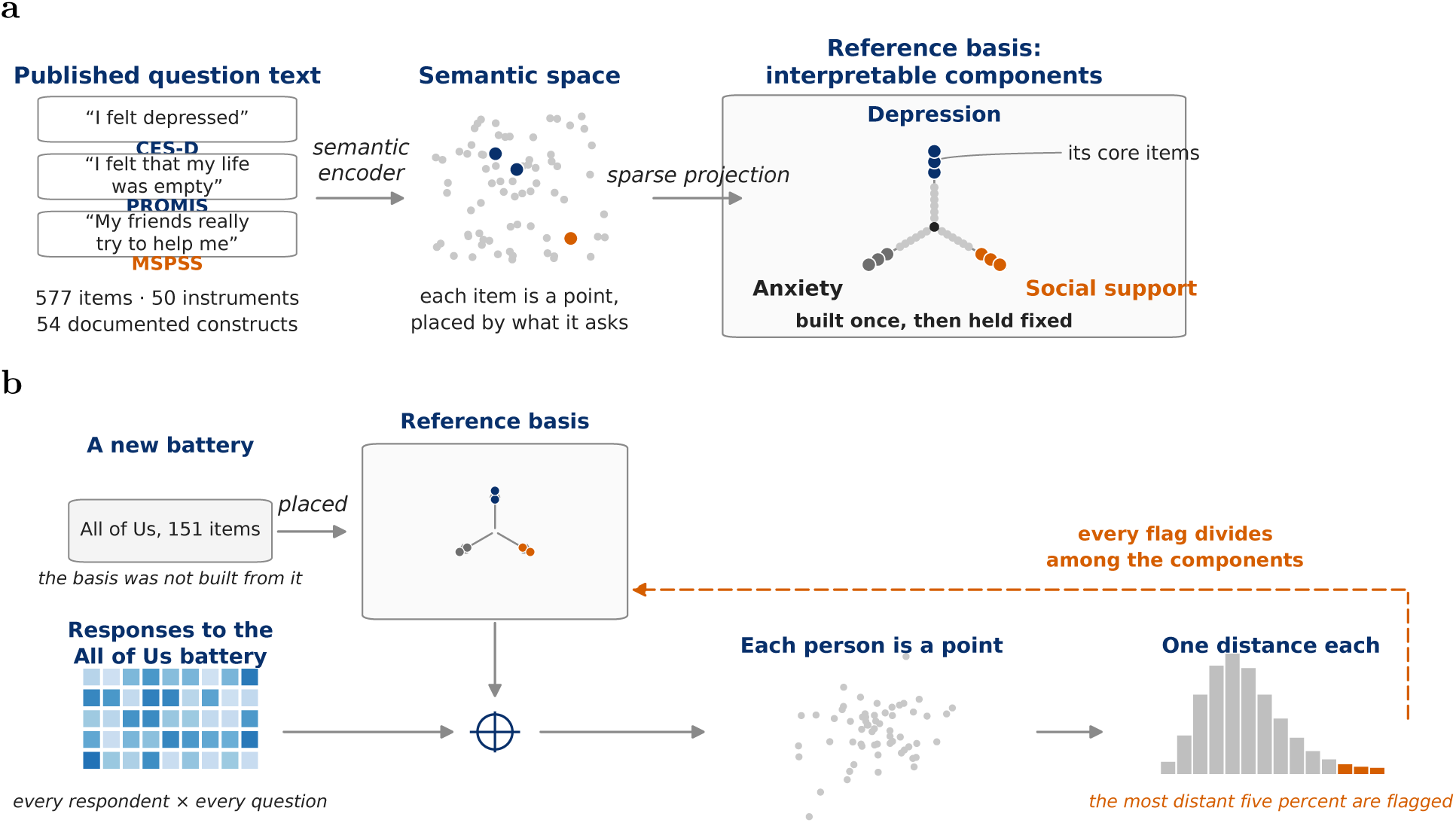
A reference basis built from published question text, and what it is for. **(a)** How the basis is built. The published text of the pool’s 577 questions is embedded by a sentence encoder, so each question is a point placed by what it asks, and a sparse projection turns those points into a small set of components, each dominated by a few items, its core items. Three components are drawn as directions and named by the documented constructs of their core items. The basis is then held fixed. **(b)** What it is for. The All of Us battery, which the basis was not built from, is placed on those same components. The basis and a respondent’s answers together give that respondent one position and one distance, each distance measured against that respondent’s own cohort. The most distant five percent are flagged, and each distance splits into one contribution per component. The three questions in (a) are pool items shown verbatim. The point clouds, the response grid and the distribution are schematic.

We demonstrate this on two batteries that include instruments and questions outside the pool. The first is 151 questions we selected from 17 instruments and 13 standalone questions across six surveys, answered by 22,332 All of Us participants [14]. The second is a much smaller battery from the Health and Retirement Study (HRS), which the basis places where its documentation says it belongs. Scoring the All of Us participants on the basis returns a group of statistical outliers whose burden is spread across many instruments rather than concentrated in one. Nearly all of them also meet at least one of the battery’s eight validated clinical criteria, and on average they meet 5.0 of the eight against 1.4 across the cohort. Five participants meet none, and the screen traces all five to discrimination, which the battery asks about and no instrument in it scores. The group as a whole reports more barriers to reaching care and concentrates more of its care in the emergency department, and being more distressed does not explain either.

## 2. Results

### 2.1. A reference basis computed from item text alone groups items by content

We computed the *reference basis* on a pool of 577 validated items from 50 published instruments covering 54 documented constructs (Methods §4.2, Supplementary Note S17 and Data 4), under six sentence encoders (Supplementary Note S3 and Table S5). Single-encoder results below are given for gte-large-en-v1.5, taken as the exemplar. That encoder returns a reference basis of 55 components. Each component is named by the documented constructs of the items that *core* it, in the instruments’ vocabulary rather than one of our own (Methods §4.2, §4.9). Thirteen components rest on a single construct and seven on two (Fig. 2a gives those resting on at most three constructs, Supplementary Fig. S5 all 55, and Supplementary Data 4 every encoder).

**Figure 2:**
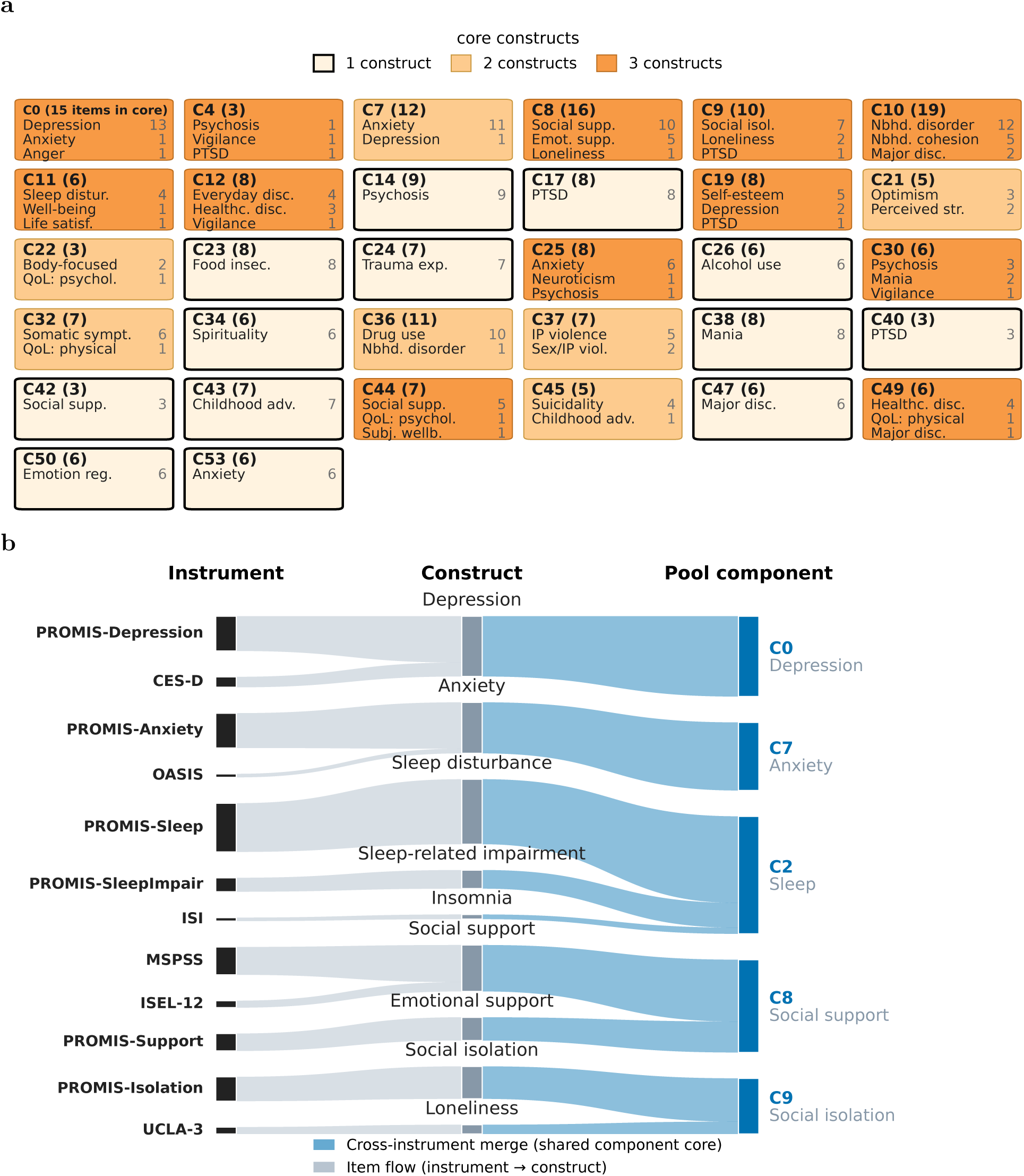
What the components rest on. **(a)** The 32 components whose cores rest on at most three documented constructs. Each card gives the component, its core size in parentheses, and the constructs of its core items with the number of items assigned to each, colored by the number of distinct constructs in the core (lightest = a single construct). All 55 are in Supplementary Fig. S5 and, with every core item’s question text, in Supplementary Data 4. **(b)** For five constructs, item flow from source instrument (left) through construct (middle) to the component its items land on (right), gte-large exemplar.

Computing the basis used no documented construct labels, yet items that share a construct land in the same component’s core about 18 times more often than items whose constructs differ (18% against 1%) (Methods §4.8, Supplementary Note S16). Where the grouping departs from those constructs, it departs by content. Some constructs split across components, depression and anxiety most of all, and items from different instruments meet on one component. The depression items of the Center for Epidemiologic Studies Depression Scale and of the PROMIS depression bank land on a single component. The social-support items of the Interpersonal Support Evaluation List and of the Multidimensional Scale of Perceived Social Support land on another (Fig. 2b).

From the item side, each item is dominated by its loadings on a few components, its *core components* (Methods §4.2). The De Jong Gierveld loneliness item *I often feel rejected* is a core item of no component, and its four core components still fit what it asks, led by social isolation, life satisfaction, depression and optimism (Supplementary Fig. S4a). The constructs of an item’s core components name it, and we compare that name with the item’s own documented construct. The documented construct is among those constructs 87% of the time against 40% expected by chance (Methods §4.9, Supplementary Note S11 and Fig. S4). Where label and placement disagree, the placement often follows the question rather than the label, as when the PROMIS Sleep Disturbance item *I felt worried at bedtime* lands among the anxiety components rather than the sleep ones. The remaining disagreements are items whose construct the pool covers with a single instrument or with none at all (Supplementary Note S11, Supplementary Data 6). A dense basis of the same size on the same items shows that sparsity, not the reduction in dimension, makes a component interpretable (Methods §4.10, Supplementary Note S19 and Table S16).

The results so far are for one encoder. Components are matched between every pair of the six by the overlap of their core items alone, with no construct labels entering. Of the 55, 39 reproduce in at least four encoders and 27 in all six (Fig. 3a, Methods §4.11, Supplementary Note S12 and Fig. S6). The content that recurs is socially and clinically central, among it depression, psychosis, food insecurity, everyday discrimination and adverse childhood experiences, while the 16 that reproduce less widely are mostly components whose cores spread over many constructs at once (Supplementary Table S8, Fig. S5, Supplementary Data 5). The construct profiles of an item’s core components agree across encoders as well, overlapping at 0.38 against 0.11 expected by chance (where 1 means full overlap and 0 means none), and most closely for items with a single core component (Fig. 3b, Supplementary Note S12 and Table S9).

**Figure 3:**
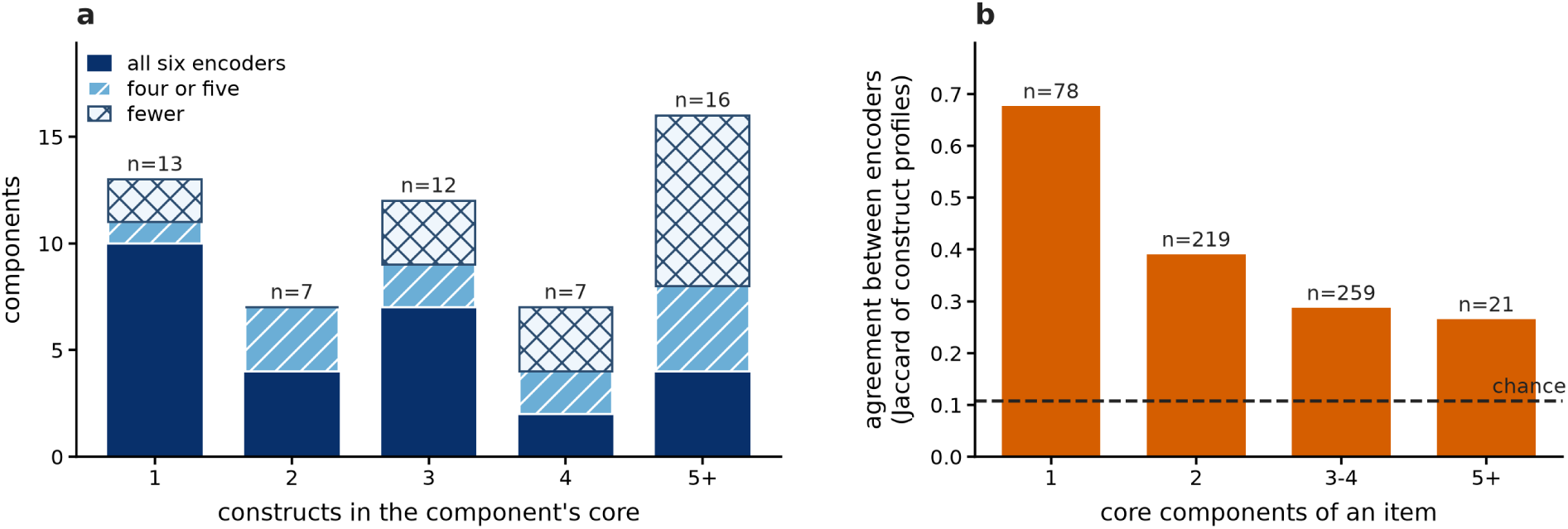
The components reproduce under different encoders. **(a)** The 55 gte-large components, binned by the number of distinct documented constructs in the core, with each bar split by the number of the six encoders in which it reproduces. A component counts as reproduced in an encoder when its matched core there overlaps it beyond chance (Supplementary Note S12). Components with up to two-construct cores almost always recur, and reproducibility falls as a core spans more constructs. **(b)** From the item side, how well the construct profiles of an item’s core components agree between encoders, by the number of core components the item has, against the permutation chance level (dashed). Agreement is the overlap of two profiles, 1 for identical profiles and 0 for profiles sharing no construct.

No responses enter the basis, so it does not depend on who answered. In contrast, a structure estimated from responses is a property of the sample that produced it. Fitting a factor model to the 22,332 All of Us responses gives item loadings that agree between two disjoint halves of the cohort at 0.85, and a principal component analysis gives the same. With one hundred respondents in each half, the agreement falls to 0.29. The text-derived basis is the same at every sample size (Methods §4.12, Supplementary Note S13 and Fig. S7).

### 2.2. The basis places instruments outside the pool where their documentation says they belong

Because the basis is fixed, we place a new battery on it without refitting. We embed its items with the same encoder, project them onto the same components, keep the components its own items reach, and name each kept component by the documented constructs of its core items in the battery (Methods §4.3).

We start with All of Us’s own battery, the 151 items we selected from 17 validated instruments and 13 standalone questions across six surveys, spanning 30 documented constructs (Methods §4.4, Supplementary Data 1). It keeps 43 of the 55 components under the exemplar encoder and 41 to 49 across the six encoders. Three of its instruments are external to the reference pool, which holds none of their item text, and they are the test of transfer.

All three land on components that different instruments core within the pool, documented for the same content (Fig. 4, drawn in red on the right axis, with the items that form every component of this battery under each encoder in Supplementary Data 3). The Big Five Inventory-2 personality items, documented in All of Us as open-mindedness, land on a component the International Personality Item Pool markers core, whose construct profile is led by openness. The childhood adversity items land on a component cored entirely by the Adverse Childhood Experiences Questionnaire, whose items are documented as adverse childhood experiences. The perceived social support items land on a component cored by the Interpersonal Support Evaluation List, documented as social support. Each pair is two instruments written by different authors, with no item text in common and a different documented name for the content, so the agreement rests on the semantic content alone.

**Figure 4:**
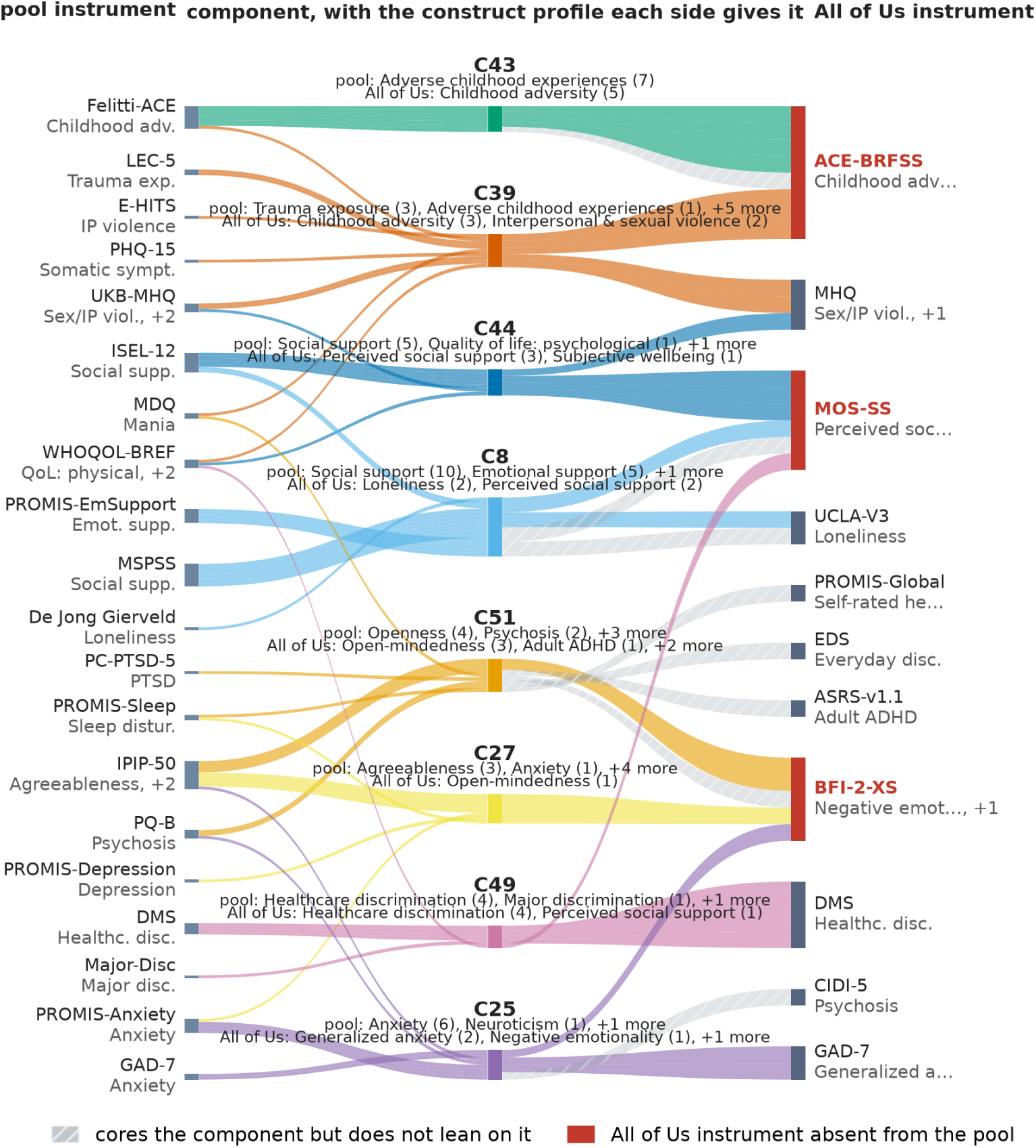
Independently written instruments core the same components. Eight components of the basis, each shown from both sides (gte-large exemplar). The component is in the center, with two profiles printed one above the other: the *pool-side* profile, from the constructs of its core items within the pool, above the *battery-side* profile, from the All of Us items that core it within that battery. On the left are the pool instruments whose items form its core, on the right the All of Us instruments whose items form it, each labeled with the documented construct of the items it contributes. Each side is named in its own documentation, so the two names differ where the content does not. The three All of Us instruments the pool never contains are drawn in red on the right axis. Where the same instrument appears on both sides it is a shared instrument rather than a transfer. Hatching marks an item that joins a component’s core within the battery without itself being one of the items that keep that component (Methods §4.3).

The HRS 2022 questionnaire [15] is a much smaller battery, 35 social items across five instruments spanning six documented constructs, and the reference pool contains none of the text of its eight neighborhood items. Six of those eight land on a single component whose pool-side construct profile is led by neighborhood disorder and neighborhood cohesion. The battery keeps 21 of the 55 components under the exemplar encoder and 18 to 21 across the six encoders. The components it drops cover content HRS does not ask about, and under every encoder each of its six constructs is in the construct profile of at least one component it keeps (Supplementary Note S14, Tables S10 to S12, Figs. S8 to S14). Supplementary Note S21, Fig. S15 and Tables S18 to S21 place every question of both batteries item by item.

Placement could still follow the wording rather than the content. The pool asks 92 of the 151 All of Us items and 18 of the 35 HRS items itself, some in the same words and some in its own, so the same content can be compared in two wordings. Of the 92, the 35 whose wording differs from the pool’s all share at least one core component with their pool version, at a mean overlap of 0.73 under the exemplar encoder against 0.03 for a random pool item, where 1 means full overlap and 0 means none. Their differences are most often a recall window such as *In the past 7 days*, which the pool strips before embedding. The 14 HRS items that differ are bare fragments against the pool’s interrogative frame, and 13 of the 14 still share a core component, at a mean overlap of 0.49. Both overlaps hold across the six encoders (Supplementary Note S18, Table S15). The grouping therefore follows the semantic content much more than the syntax.

For a stronger test of whether the grouping is driven by semantic content, we independently vary content and syntax rather than rely on the different syntax the instruments happen to use. We rewrote each of the 151 All of Us items twice, once carrying its own content in another item’s syntax and once carrying another item’s content in its own syntax, and placed the resulting variants, about 3,000 in all, on the same basis (Methods §4.14, Supplementary Note S5 and Fig. S1). The variants land closer to the item whose semantic content they carry than to the item whose syntax they took, under every one of the six encoders. The exception is syntax that itself names a construct, as in *… in my neighborhood*, where the semantic content of *neighborhood* cannot be separated from the syntax by design and the content item and the syntax item come closer together without changing places.

### 2.3. The basis scores respondents, and the most distant are burdened across many instruments at once

We now score the 22,332 All of Us participants who answered all 151 items on the components the battery reached in §2.2. This turns each person’s whole pattern of answers into a single position in the basis, and we give each participant one *distance*, how far that position sits from the cohort’s mean (Methods §4.15). We flag the most distant five percent under each of the six encoders, about 1,100 participants each. Those six sets overlap at a mean of 0.55 over the fifteen encoder pairs, where 1 means the same set of people and 0 means no one in common, against 0.026 expected if the six screens were unrelated. That margin holds at every threshold from the 90th to the 99th percentile (Supplementary Note S6). The six independently trained models therefore overlap far more than chance in whom they flag, and that overlap is not an artifact of where the cut is made. The 510 **participants** flagged under all six, 2.3% of the cohort, are the *consensus outliers*, called simply the outliers from here on.

Since neither the basis nor the screen uses any clinical label, score or cutoff, we check the screen against the criteria clinicians use. Eight of the instruments in the battery have a validated criterion (Supplementary Note S20 and Table S17). 505 of the 510 outliers meet at least one of the eight criteria, 99% of the group against 58% of the cohort as a whole. Counting how many of the eight a participant meets at once, rather than whether they meet any, turns the same criteria into a measure of how many kinds of burden they report. An outlier meets a mean of 5.0 **of the eight**, against 1.4 across the cohort, with five or more met by 62% of the group and all eight by 11% (Fig. 5). That breadth is not a matter of how the outliers use the response scale or of severity alone (Supplementary Note S7).

**Figure 5:**
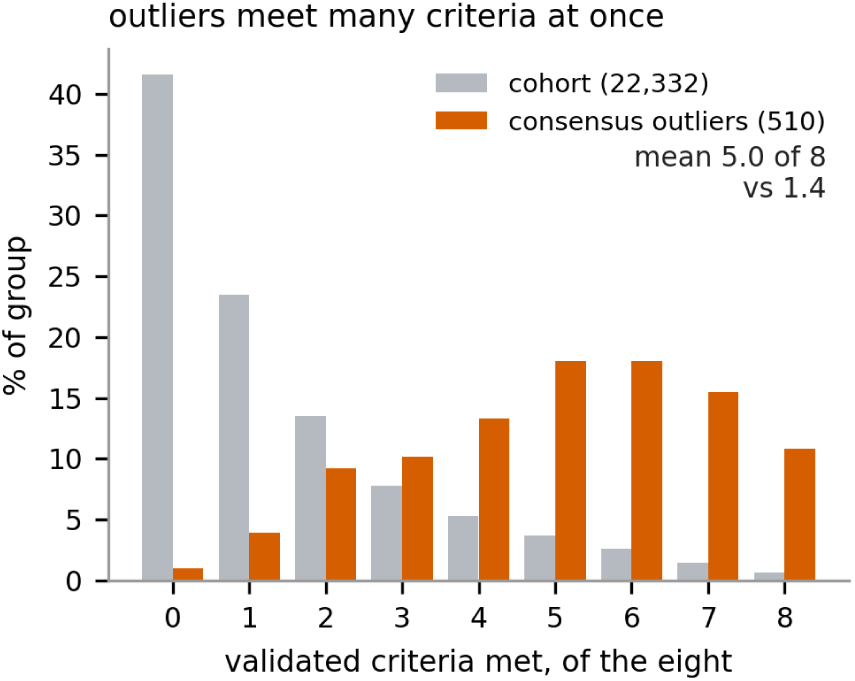
The outliers meet more validated criteria at once than the cohort. For each participant, how many of the eight validated criteria they meet, as a percentage of the group, for the cohort (*N* = 22,332) and for the consensus outliers (*N* = 510). The cohort concentrates at zero or one, the outliers at five, and 11% of them meet all eight. The agreement of the six encoders across outlier thresholds is shown in Supplementary Fig. S2.

### 2.4. Five outliers meet no validated criterion, and discrimination carries all five

Five of the 510 outliers meet none of the eight validated criteria, so the content that puts them in the group lies outside those criteria. We call these five the *clinically invisible* outliers. The distance itself locates that content, because it splits exactly into one contribution per component (Methods §4.7). A component’s *share* is how much of the participant’s distance from the cohort mean it carries. We say a component *carries* a participant when it is among those that reach half of that distance. Each of the five is carried by a discrimination component under the exemplar encoder, and under each of the other five encoders a discrimination construct carries all five as well (Supplementary Note S22, Table S22). None of the eight validated criteria covers discrimination.

The five differ in how that component makes them an outlier. A component can be far from the cohort in its own right, or unremarkable on its own and far from what the participant’s other components predict. A participant’s *integration* is the share of their distance carried by components of the second kind. That part exists only because the components are taken together. For participant 1, the component with the largest share is far from the cohort in its own right, and components of the second kind carry only 12% of their distance. For participant 2 the largest component is unremarkable on its own, and components of the second kind carry 98%. **Participant 2 is therefore unusual in how their components combine, not in any one component.** Table 1 interprets the five. Still, none of the five is beyond the reach of a single instrument. Each is no longer flagged once we set the items on which they are individually extreme to the cohort mean, a median of one to four items per participant (Supplementary Note S9).

**Table 1:** The five clinically invisible outliers, who meet none of the eight validated criteria. For each, the component carrying the largest share of their squared distance and the next largest, each named by the documented constructs of the All of Us items that core it within the battery, under the exemplar encoder. *|z|* alone is how far the participant sits from the cohort on that component with the others ignored, and *|z|* given the rest is how far they sit from what their other components predict. Integration is the share of the distance carried by components that are extreme given the rest (Methods §4.7). Participants are ordered by their largest share and numbered as in Supplementary Table S22. The same five under all six encoders are Supplementary Note S22.

|  | Largest share |  |  |  |  | Next largest |  |  |
| --- | --- | --- | --- | --- | --- | --- | --- | --- |
| | Constructs of its core items | Share | $\vert z\vert $ alone | $\vert z\vert $ given rest | | Constructs | Share | Integration |
| 1 | everyday discrimination, healthcare discrimination | 44.9% | 6.5 | 3.3 |  | perceived social support, subjective wellbeing | 13.9% | 12% |
| 2 | open-mindedness, self-rated health, everyday discrimination, adult ADHD | 28.5% | 3.2 | 6.0 |  | everyday discrimination, healthcare discrimination | 20.5% | 98% |
| 3 | everyday discrimination, healthcare discrimination | 26.0% | 4.4 | 2.9 |  | perceived social support, loneliness | 12.5% | 66% |
| 4 | healthcare discrimination, perceived social support | 21.5% | 3.0 | 3.1 |  | interpersonal & sexual violence | 19.0% | 44% |
| 5 | substance use, neighborhood disorder & safety, childhood adversity | 19.1% | 2.3 | 4.0 |  | adult ADHD, healthcare discrimination, religious participation, everyday discrimination and others | 18.4% | 78% |

### 2.5. The group divides by content, much of it specific to a handful of participants

The same decomposition applied to all 510 shows that the content carrying a participant varies from one to the next (Fig. 6). For 268 the component with the largest share is one of only six, each of them largest for at least twenty participants. For another 143 it is one of 29 that are largest for fewer than ten each (Supplementary Note S26), and the remaining 99 sit on eight components of intermediate frequency. This tail of rare content is specific rather than residual. Two of the 29 are cored on interpersonal and sexual violence alone, and one on suicidality and self-harm. Across all 29, that content carries the few it reaches nearly as far as common content carries anyone, a median 20.3% of the distance against 25.5% for the six. Per-instrument scoring returns one number per instrument against a threshold set for that instrument alone, and those numbers are not commensurable, so it cannot report which content carries a participant.

**Figure 6:**
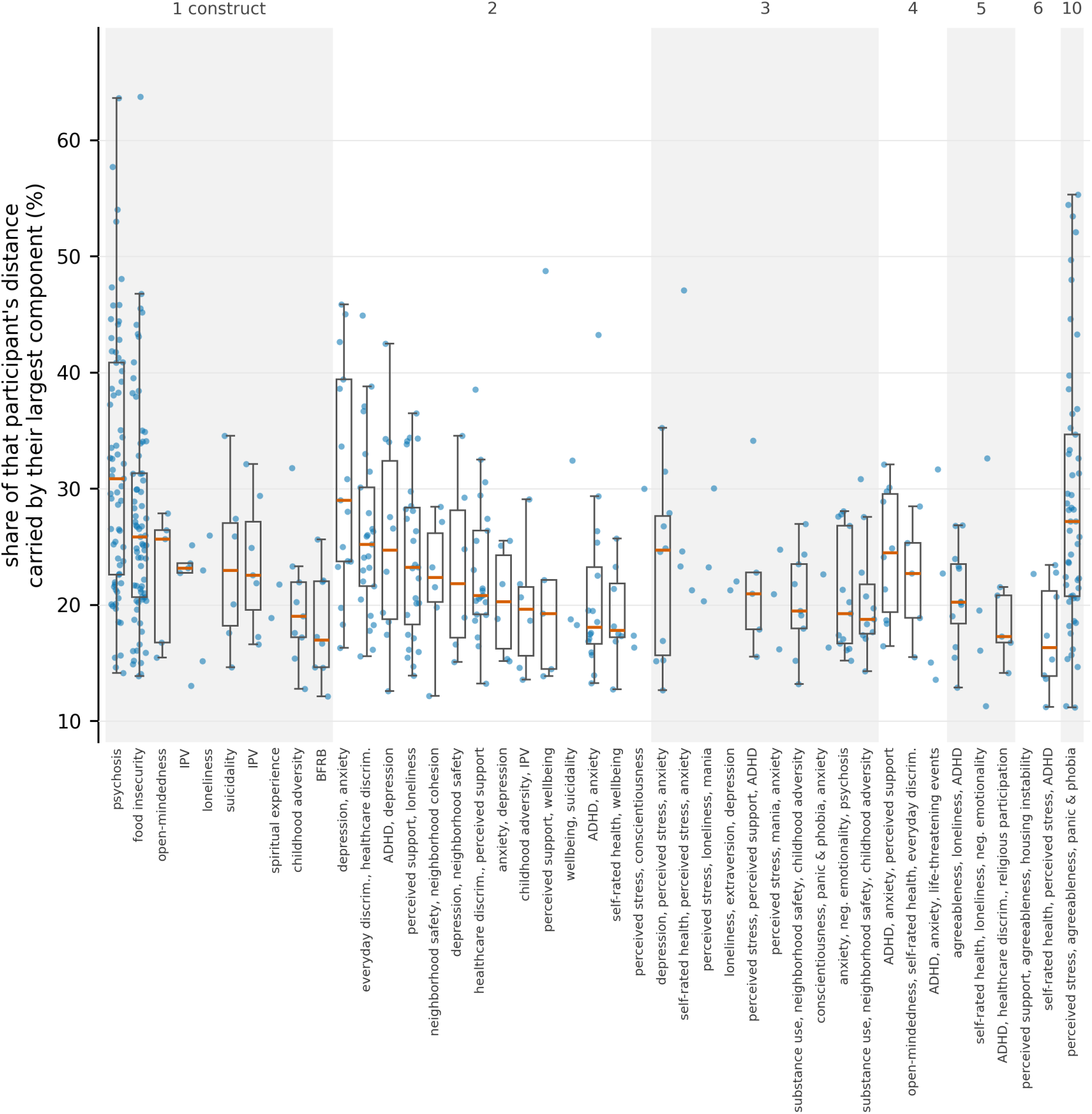
A few components are largest for most outliers, and many more for only a few each. Each of the 510 appears once, at the component carrying the largest share of their squared distance, on the exemplar encoder. Components are grouped by how many documented constructs core them within the battery, numbered above the panel and ordered within a group by decreasing median. Each is named on the axis by up to three of those constructs. Boxes are drawn only for components that are largest for at least five participants, and give the median and quartiles.

Naming that content does not depend on the encoder. Component indices are not comparable across encoders, since each fits its own basis, but the documented constructs are the same objects under all six. Summing a participant’s item contributions within constructs therefore partitions their distance with no weighting choice (Methods §4.7). The construct carrying the most of a participant’s distance agrees across at least four of the six encoders for 82% of the group. At the component layer, where indices are local to an encoder and only the construct profiles can be compared, that agreement instead tracks how concentrated a participant’s distance is (Supplementary Note S24 and Fig. S16). The construct names are the documented constructs of the battery’s own items, taken from the instruments’ documentation rather than from the responses. We therefore name what carries a participant by construct rather than by component index, and divide the group by those names.

### 2.6. Reaching care is harder for this group, and not because they are more distressed

Neither the access survey nor the health records entered the screen. In the access survey, the outliers are about twice as likely as the rest of the cohort to report a barrier to care (82% against 44%) or a cost barrier (67% against 28%). They are more than three times as likely to have delayed or gone without care because of a provider’s race or religion (32% against 9%). In their electronic health records, their care is about twice as concentrated in the emergency department (2.4% against 1.2% of visits). The gap remains when we hold the total amount of care a person receives fixed (Methods §4.17). Neither pattern is confined to the flagged group. Across the whole linked cohort both rise steadily with the score rather than appearing at a threshold (Fig. 7, with the barrier-by-barrier breakdown and the visit-count caveat in Supplementary Note S8, Table S7 and Fig. S3).

**Figure 7:**
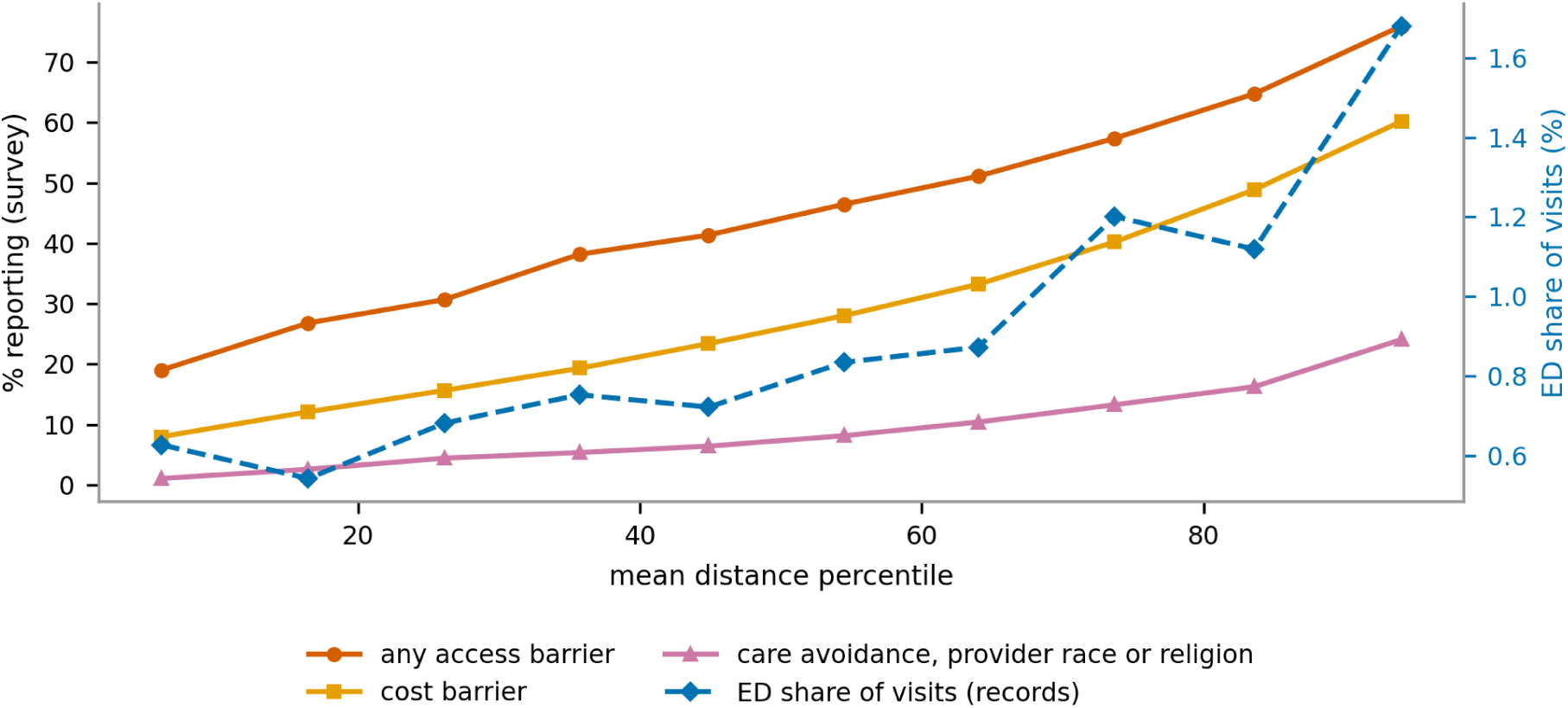
The further a participant sits across instruments, the more barriers to care and the more care in the emergency department. Participants grouped into ten equal-size bins by their cross-instrument score, each marker at its bin’s mean percentile (electronic-health-srecord cohort, *N* = 20,972). The share reporting access, cost, and race/religion care-avoidance barriers rises (left axis, survey self-report), and the emergency-department share of a participant’s visits rises (right axis, records). Two different data streams, the same under-served group.

Being more distressed in general does not account for this. Two simpler measures built from the same 151 answers make the comparison. The first is how far a participant’s answers depart from the cohort’s on average, with no cross-instrument structure in it at all. The second is the same cross-instrument score with the correlations between components discarded, which keeps the reference structure and drops what taking the instruments together adds. Each is associated with the same outcomes on its own. When we compare the three measures together, only the full cross-instrument score remains associated with emergency-department use, and neither simpler measure is distinguishable from no association. For the self-reported barriers the full score accounts for most of the association and the average departure for a smaller part (Supplementary Note S25, Table S23). The three measures identify overlapping but different people, agreeing on only 618 of their respective most extreme five percent. They differ in which participants they pick out, not in how strongly they rank the same ones. The effects are small and the comparison is cross-sectional. Within those limits, a participant’s position across instruments accounts for the association with reaching care, and their average departure for less of it.

## 3. Discussion

We built one frame of reference from the published text of 577 questions spanning 50 instruments, held it fixed, and placed two batteries on it from their item text alone. Scoring the All of Us participants on that frame returns a group whose difficulties are spread across many instruments rather than concentrated in one. The validated criteria identify most of them and miss a few entirely, and the group reaches care less easily than the rest of the cohort.

No label defines that group. Its members meet several validated criteria at once rather than one (§2.3), and the content that carries them varies from person to person, much of it specific to a handful of participants (§2.5). Five meet no criterion at all, and all five are carried by discrimination, which the battery asks about and no instrument in it scores (§2.4).

Two simpler measures built from the same answers test whether the group’s poorer access to care is severity in another guise. One has no cross-instrument structure, the other discards the relations between components, and each accounts for less of the association than the full score does (§2.6). The effects are small. Within that limit, breadth across instruments, not distress in general, tracks reaching care.

That breadth is measurable because one structure spans every instrument in the battery, and we recovered that structure from the semantic content of the items rather than from the responses. A structure recovered that way is the same for any cohort and exists before anyone answers. A factor model estimates the structure from the responses instead, so the structure it finds belongs to the sample that produced it, and shifts when it is re-estimated for the next cohort (§2.1). The routes that carry structure between instruments have the same dependence, and each is rebuilt for the pair in front of it. Integrative data analysis and item-response-theory linking align instruments through shared response data or co-calibration samples [16, 17], and the calibrated items go into banks a study can draw from [18]. Experts align instruments instead by comparing what the items ask, which is how retrospective harmonization across cohorts works [19]. That route needs no responses either, but each alignment is a judgment about the studies at hand and produces nothing a further battery could use.

The same problem recurs within one cohort measured over decades. Batteries change, their items overlap only in part, and a change in a person’s score means a change in the person only when the two measurements are comparable. In long-running cohorts they rarely are. The National Health Interview Survey redesigned its questionnaire in 2019, estimates from that year on are not comparable with earlier ones [20], and comparisons across the change rely on variables harmonized one at a time [21]. A structure built from item text needs neither shared respondents nor shared response data, and it is the same for every pair. It places every version of a changing battery from its item text, with no year serving as the reference, so repeated measurements of one person sit on the same components.

Scoring people in that structure turns it into a screen, but detection is not the contribution. A factor model scored with the same distance also uses the whole battery, so cutoff-free screening is not unique to a structure built from item text. The components add something else. Each rests on a few readable questions, so a score divides among the questions that produced it. The screen reports not only that someone is unusual but what makes them so. An outlier attributed to discrimination or food insecurity is visible in a way the same outlier, dispersed across every item, is not.

Fixing the frame sets two limits on the screen. The first is that the frame transfers the structure between batteries, not yet a score. We place two batteries on the same components, but we score each cohort against its own mean and covariance. A distance therefore says where someone sits within their cohort, and nothing about how they compare with anyone outside it. A score two cohorts share would need a reference distribution belonging to neither. The components make that possible and this work does not attempt it.

The second is coverage. The frame holds only what the pool covered. A battery can ask about content that none of the 577 questions raised, and the placement retains little of such an item, so its distinctive content is not represented. We report that retained share for every question of both batteries (Supplementary Note S21 and Fig. S15). Two batteries placed on the same components can be compared only on content that both retain, a check this work reports item by item but does not make. Widening the pool needs more published item text and no new responses, but a wider pool gives a different frame, so a frame’s coverage is settled when the frame is fixed.

Beyond the frame, two parts of the pipeline are taken as given rather than fitted. The first is how responses enter. We take each item on its own instrument’s response scale and rescale it to [*−*1, 1], which treats its ordinal categories as equally spaced and gives the origin a different meaning across items. Treating ordinal responses as continuous is defensible above about five categories and less so below [22], and metric models fitted to ordinal data can distort what they estimate [23]. A screen keyed on distance is less exposed than a regression coefficient, since any order-preserving recoding leaves the same people far from the cohort. But the distance still depends on where each category is placed. Estimating where each category sits rather than assuming it [24], as item-response theory does, is the clearest improvement available here. The second is the encoders. We use them off the shelf, and models fine-tuned on psychometric items [10] could sharpen both the structure and the screen.

The cohort sets a further limit on what the associations can mean. All of Us is a volunteer sample that is not statistically representative of the US population, and the response-level analyses are cross-sectional, so they describe associations rather than causal pathways. They are also restricted to participants who completed all 151 items and, for the health records, to the linked subset, so completers may differ from non-completers. These analyses are exploratory and were not preregistered, and we guard against overfitting by requiring consensus across six encoders. Validating the screen out of sample, on new item sets and longitudinally against clinical outcomes, is the natural next step. With repeated measurements placed on the same components, the question after that is whether a person’s recent movement predicts where they go next.

The text of a battery’s questions is enough to build a compact representation of that battery and to score respondents on it, with every outlier traceable to the questions behind them. The approach asks for no new data collection, only the batteries that cohorts and health systems already administer. Applied to those batteries, the approach returns a group that no single instrument defines, together with the content that puts each person there, and that group reaches care less easily.

## 4. Methods

### 4.1. Overview

We compute the reference basis once from the published text of the pool items and then hold it fixed (§4.2). We place a battery on it by projecting its items, refit nothing, and keep the components its items reach (§4.3). We score respondents on those kept components and flag the unusual ones (§4.6), and each outlier’s distance partitions exactly over the items, and so over the constructs, that produced it (§4.7). The subsections below give that pipeline, then the All of Us battery, then the analyses in the order the Results report them.

### 4.2. The reference pool and basis

We assembled a reference pool of 577 validated psychosocial items from 50 published instruments spanning 54 documented constructs, taken from published item text (Supplementary Note S17, Table S14, Supplementary Data 4). All of Us also administered fourteen of these instruments, kept as shared instruments, and the other 36 are independent instruments of the same constructs, so most constructs are measured by more than one unrelated instrument. Each item’s construct label is the one documented in its instrument’s own codebook or manual, never inferred from the item text. We stripped only the leading recall-window phrase from each item.

Embedding the published text of the pool’s *p* = 577 items with a sentence encoder gives an embedding matrix *M ∈* ℝ*^p^*^×^*^D^*, whose rows are the items and whose *D* columns are the encoder’s embedding dimensions, and we mean-center its columns to *M̃*. We use six encoders—bge-m3, bge-large-en-v1.5, all-mpnet-base-v2, e5-large-v2, gte-large-en-v1.5 and qwen3-embedding-0.6b—each with its own embedding dimension *D* (768 for all-mpnet-base-v2, 1024 for the rest), each yielding its own basis, and we process all six identically (Supplementary Note S3, Table S5). gte-large-en-v1.5 serves as the exemplar for single-encoder figures. The stage returns *k* orthonormal directions *W ∈* ℝ*^D^*^×^*^k^* and the item projection

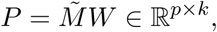

whose entry *P_ic_*is the weight of item *i* on component *c*. Each column of *P* is a *component*, a weighting over all *p* items. *P* is built from item text alone, with no responses and no manual cross-walk between instruments.

We select the number of components *k* per encoder by parallel analysis on permuted data [25, 26]. Parallel analysis retains a component only when its eigenvalue exceeds the eigenvalue from column-permuted embeddings, which keep each dimension’s marginal distribution but destroy the cross-dimension covariance. This calibrates *k* against chance, necessary because *p* items in hundreds of embedding dimensions inflate sample eigenvalues, and it replaces an arbitrary variance or scree cutoff with a data-determined threshold.

We concentrate the projection with ROSS-PCA’s fully differentiable (log-sum, reweighted-*ℓ*_1_) sparsity penalty [27, 28], applied to the projection *P* rather than to the directions *W*, with an orthonormality term holding the columns of *W* orthogonal. We penalize *P* because *P* is the object we interpret, whereas *W* lies in the embedding space and is not human-readable. The penalty therefore acts on the items’ coordinates along the directions, not on the directions themselves, so the fit searches over *W* for a set of orthonormal directions whose induced item projections are sparse. Finding the directions and concentrating the projection are one optimization rather than a variance-maximizing fit followed by pruning. The penalty shrinks small coordinates without zeroing them.

Each component’s *core* is the fixed point of a pruning iteration on *P* : keep the top-round(PR) items by weight—where the participation ratio 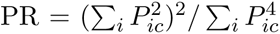 is the effective number of items that make up component *c*—then recompute PR on the survivors and re-prune, iterating to convergence. This yields the self-consistent set whose size equals its own participation ratio, and the iteration converges because any set *S* satisfies PR(*S*) *≤ |S|*, so the size never increases. A component’s *core energy* is the fraction of its squared weight that its core holds, 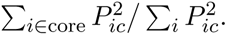 Supplementary Table S14. gives the number of components and the size of their cores under each encoder.

The objective is non-convex, so its solution depends on the initialization, and every fit reported here starts from the principal components of the same embeddings, which is the solution the penalty weight *λ*_score_ recovers as it goes to zero. We set that weight by a sweep over {0.001, 0.005, 0.01, 0.05, 0.1, 0.5, 1, 5} for each encoder, refitting from the principal components at each value with *λ*_ortho_ = 0.1. We take the value that maximizes median core energy while *W* stays orthonormal and the variance retained relative to principal component analysis holds. That value is *λ*_score_ = 1 for every encoder (Supplementary Note S15 and Table S13). The construct labels take no part in this choice.

The same pruning, applied across an item’s row rather than down a component’s column, defines the item’s *core components*. Because the columns of *M̃* are centered, each component of *P* has zero mean over the items, so we scale each to unit variance, *P̂_ic_* = *P_ic_*/*s_c_* with 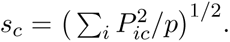 The core components of item *i* are the fixed point of the same top-round(PR) iteration run on the standardized row *P̂_i_*_·_, the small set of components on which the item departs most from the rest of the pool.

We scale because the components have unequal spread, about a factor of three in *s_c_*. On the raw row the fixed point selects the components an item is *reconstructed* from, and the widest component dominates that choice whatever the item asks. On the standardized row it selects the components that *distinguish* the item from the rest of the pool (Supplementary Note S10). The scaling is not a whitening of *P*, and the divisors *s_c_* are fixed on the pool and stored with *W* and the centering, so an item projected onto the basis later is standardized against the same reference.

### 4.3. Placing a battery on the basis

The portable object is the triple (*µ*_pool_*, W*_pool_*, σ*_pool_), the pool’s mean, its component loadings, and its per-column standard deviations, computed once on the pool and stored with it. A new battery contributes to none of the three, and nothing is refit.

Placing a battery *S* takes three steps. We embed its items with the same encoder, without the pool’s stem removal, so a battery is placed on the basis exactly as its items are published (Supplementary Note S17). We project them onto the stored components, 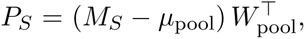, and divide column-wise by the pool’s standard deviations, *Z_S_* = *P_S_/σ*_pool_, so the battery is expressed in the pool’s units and not in its own. Each item’s *core components* are then the fixed point of the participation-ratio rule applied across its row of *Z_S_*, and its *modal component* is the single one it stands out on most. A component’s *core-item band* on the pool is the smallest standardized loading among the pool items that form its core there. A component is *kept* for *S* when at least one item of *S* clears that band, or when it is the modal component of at least one item of *S*. Components meeting neither condition are dropped when the basis is used for *S*. Both conditions are evaluated one item at a time against divisors and bands fixed on the pool, and the modal condition contributes at most one component per item, so the kept set stays below the battery’s own item count. The screen needs that bound. If the number of kept components reached the number of items, the scored subspace would span the responses themselves whichever components defined it, and the distance would be the one a response-based screen computes. Finally we interpret each kept component by its core items within *S*, the fixed point of the same rule applied down that component’s column of *Z_S_*, and label it by the documented constructs of those items.

Two properties follow. An item’s core components are computed from its own row against fixed divisors, so they do not depend on what else is in *S*, and adding items to a battery can keep further components but cannot change the ones already kept. The interpretation step is a selection down a single component’s column, where the constant divisor cancels exactly, so it does not depend on the calibration but does depend on the composition of *S*. We interpret a component with the constructs documented for the items of *S*, so this analysis never assumes or uses a correspondence between the vocabulary of *S* and the pool’s own labels.

One caveat follows from keeping the pool’s origin. On the pool the components are zero-mean, so being extreme on one and being far from the battery’s own center coincide. On a new battery they need not, and a battery displaced along a component can have its core items there picked out by the displacement. No All of Us component sits more than half a pool standard deviation off center, while seven HRS components exceed that and one exceeds a full standard deviation, so the caveat is immaterial for All of Us and a stated limitation for the smaller battery. Assembly, provenance, per-encoder tables, and the reproducibility map are provided in Supplementary Data 4.

### 4.4. The All of Us battery

The battery placed on the basis is 151 items selected from six All of Us survey modules. Its items are embedded by their *prompt text* (not responses) with the same six encoders and placed on the basis without refitting (§4.3). For grouping we use the authoritative item-to-instrument map of Supplementary Note S1 and Table S1 (17 multi-item instruments [29–45] plus 13 single items, with the three suicide items treated as one unit).

### 4.5. Construct assignment

To organize the items by what they measure rather than by their source instrument, we assigned each of the 151 items to a single *construct*, the specific attribute its prompt asks about (Supplementary Table S2, full assignment in Supplementary Data 1, items_full.csv, instrument-by-instrument grounding in Supplementary Note S2 and Tables S3 and S4). Where the source instrument documents its own subscales, those decide the assignment. Unidimensional instruments map all of their items to one construct, and we split multidimensional instruments by their validated subscales, as with the BFI-2-XS and its five trait factors. Two composite screeners, the UK Biobank MHQ and the WMH-CIDI, document no subscales, so we grouped their 21 items by topic, as we did the 13 standalone items that belong to no multi-item instrument. Those 34 of 151 items are the only ones whose construct we assign ourselves, item by item in Supplementary Data 1. This yields 30 constructs.

These labels name what carries a participant’s distance (§4.7). They take no part in computing the basis, in placing a battery on it, or in checking placement against documentation, all of which use the pool’s labels and never the battery’s (§4.2).

### 4.6. Scoring and flagging respondents

We apply the concentrated projection to respondent data. We take each item on its source instrument’s response scale and rescale it to [*−*1, 1]. The origin is therefore the middle category on an odd-length scale and no observable response on an even-length one. We reverse-code an item only when the published scoring criteria of a validated instrument mark it as reverse-keyed, so its direction is the one its own instrument defines rather than one we assign. We leave items from sources with no published scoring key in their original direction. This gives the response matrix *X ∈* ℝ*^N^*^×^*^p^* over *N* respondents and the component scores *Z* = *XP ∈* ℝ*^N^*^×^*^k^*. We residualize *Z* on age and sex at birth, age as a single linear term and sex as a binary indicator, by survey-weighted least squares, which reduces to ordinary least squares under the unit weights of this cohort. We then score each respondent by the survey-weighted Mahalanobis distance [46] of the residualized scores, with the score covariance regularized by Ledoit-Wolf shrinkage [47]. Because the responses are bounded and discrete, we define outliers by the empirical upper tail of the cohort distance distribution rather than a nominal *χ*^2^ cutoff. We run the screen separately for each encoder’s projection and take as *consensus outliers* the respondents flagged under every encoder. For score-gradient and rank analyses we also retain each respondent’s continuous *outlier score*, the mean of their empirical Mahalanobis percentile across the encoders.

### 4.7. Attributing an outlier

An outlier’s score is a single Mahalanobis distance, and we decompose it into the components that produce it. Writing a respondent’s residualized component scores as *y* and the Ledoit-Wolf precision of the cohort scores as Θ (§4.6), the squared distance splits exactly into one contribution per component,

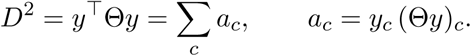

Each contribution *a_c_* is covariance-aware. It measures how far a component departs from the value its correlated neighbors predict rather than the raw size of the score. A component with a moderate score can therefore contribute more than a larger but expected one, and a contribution can even be negative (Supplementary Note S23). Ranking components by *a_c_*is therefore not the same as ranking them by score magnitude.

A component can also be extreme in either of two ways. Writing Σ for the score covariance, 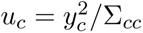 measures how extreme the component is on its own and 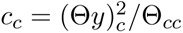 how extreme it is given the others. The component is *joint* when *c_c_ > u_c_*. Both reduce to *a_c_* when the components are uncorrelated, so the attribution adapts to the data rather than assuming an answer, and we report *integration*, the share of *D*^2^ carried by joint components.

Because the component scores are linear in the responses, the same distance also partitions exactly over the items. With *P*_kept_ the kept columns of the projection and *x* the respondent’s covariate-residualized responses,

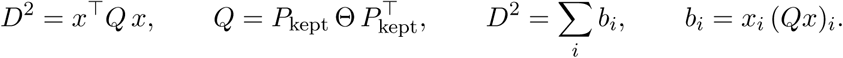

This partition is well posed, *p* items in *p* dimensions with no rank deficit, and it is the layer the item-level test of Supplementary Note S9 operates on. We report the component attribution, so an outlier comes back in the same objects the basis is built from and §2.2 interprets.

A component *carries* a participant when it belongs to the smallest set of components whose shares cumulate to half of their distance. We also report the component with the largest share, so the two counts can be compared across the group. Because every encoder fits its own basis, a component index under one is not the same component under another, so we compare components across encoders by the documented constructs of the battery items that core them. Agreement is the mean overlap of those construct sets over the fifteen encoder pairs, 1 for identical sets and 0 for sets sharing nothing, and we compare it against a floor computed the same way between different participants.

Two limits apply. We do not report the pairwise terms Θ*_cd_y_c_y_d_*, because on a battery whose kept components approach its item count they measure redundancy among components the battery cannot separate rather than anything about the respondent. And per-participant attribution is reproducible in proportion to how concentrated a participant’s distance is rather than uniformly across a group (Supplementary Note S24), a relation that holds on the item side as well (Supplementary Note S17).

### 4.8. Whether the basis separates unlike content

We measure separation pool-wide rather than component by component. Over every unordered pair of pool items we ask how often two items that share at least one component’s core also share a documented construct, and compare it with the rate for two items drawn at random, which is fixed by the sizes of the constructs alone. We report the reverse conditional as well, how often a pair shares a core given that the two items do or do not share a documented construct, which says whether unlike content is placed apart. Alongside it we report a permutation null that shuffles the construct labels over items while holding the cores fixed. It recovers the random rate, so no part of the baseline comes from the core structure. The statistic is a single number for the whole pool under each encoder rather than a per-component measure.

### 4.9. Checking the naming against documentation

Each component is named by the documented constructs of its core items, its *construct profile*, and each item has its own core components (§4.2). We call an item *faithful* when its own documented construct appears among the constructs of the core items of its core components, counting any core component rather than only the largest, because a diffuse item spreads its characteristic weight over several related components. An item never counts its own membership of a core, which would otherwise make every core item faithful by construction, so the measure is leave-one-out throughout. We report the rate against a null that permutes the item-to-construct labels and recomputes it, over 200 permutations. We report it over all items, and separately in Supplementary Note S11 for the items that core some component and those that core none, whose null rates differ because the two groups reach profiles of different size. We compare against each item’s own documented construct, with no coarser grouping.

### 4.10. Dense-basis control

To test whether sparsity rather than the reduction in dimension makes a component interpretable, we fit a dense basis on the same pool items by principal component analysis with no sparsity, with the same encoder and the same number of components. We scored it for faithfulness by the same rule (§4.9). Two things differ. A dense component has no core, so we give each the same item budget the sparse basis uses, its top *m* items by absolute standardized loading with *m* the mean sparse core size. Applying the sparse core-item band to dense loadings instead would select most of the pool. We compare the two bases on the margin of the faithfulness rate over each basis’s own null, never the raw rate, because a basis whose components span more constructs attains a higher raw rate by construction.

### 4.11. Cross-encoder reproducibility of the pool components

We assessed how far the pool components depend on the encoder by matching them across all six. For each pair of encoders we scored every pair of components by the Jaccard overlap of their fixed-point cores (the item sets) and found the optimal one-to-one matching with the Hungarian algorithm on 1 *−* Jaccard. Because the number of components differs across encoders (49 to 55), the assignment is rectangular—the smaller set is fully matched and the surplus components in the larger set are left unmatched. A null distribution comes from permuting item identities before matching, and a match counts as reproduced only when its Jaccard exceeds the 95th percentile of that null (*τ* = 0.15). A component reproduces across a set of encoders when it has a *τ* -passing match in each of them. The matching uses the item cores alone and no construct labels. We take the number of core constructs used to stratify reproducibility afterward from the construct assignment (Supplementary Data 4), and it plays no part in the matching.

We assessed the item side of the projection the same way. For each item we took its core components under every encoder and collected the constructs of their core items, which gives the item a set of constructs under each encoder. We measured the agreement of these sets between encoders by their mean pairwise Jaccard against a null that permutes item identities. A component contributes the whole construct profile of its core rather than a single label for it, so the agreement is a set comparison throughout. We report this agreement overall, stratified by the number of core components an item has, and under a share-weighted form of the same comparison that uses the item counts within each profile (Supplementary Note S12).

### 4.12. Response-based baseline and portability

To situate the text-derived structure against the standard alternative, we compare it with the two response-based structures a psychometrician would estimate from the same data: principal component analysis and common-factor analysis of the 22,332 *×* 151 response matrix. Both return the same pair of objects as the text-derived projection, a loading per item and a score per respondent, so we compare the three on those shared outputs rather than on their internal vectors. We measure the stability of the response-based structures under resampling. For each subsample size *n ∈* {100, 250, 500, 1000, 2000, 5000, 11,000} we draw two disjoint random subsamples of *n* respondents, fit the structure independently on each, match its components one-to-one across the two fits to maximize total absolute loading correlation, and record the mean matched absolute loading correlation, averaged over three repeats (Supplementary Note S13). The text-derived structure is the fixed item embeddings and does not depend on the responses, so it is identical for every subsample and defined at *n* = 0. We fit principal component analysis and factor analysis (with a varimax rotation) with scikit-learn on the same rescaled responses the screen uses (§4.6).

### 4.13. Transfer to the Health and Retirement Study

As a further test of portability we placed a second, independently administered battery on the fixed reference basis. The 2022 HRS self-administered (Leave-Behind) questionnaire [15, 48] comes from a nationally representative panel of older adults in the United States. It administers five social instruments written and administered independently of All of Us and of the pool: everyday and healthcare discrimination [36], loneliness (the eleven-item revised UCLA Loneliness Scale), perceived stress (PSS-10), and neighborhood disorder and social cohesion. We took the 35 item texts as administered—the row of each grid question, without the shared header the codebook repeats onto every item—and used the text alone, with no responses. We embedded the 35 items with the same six encoders and, holding each encoder’s pool basis fixed, projected the HRS embeddings onto it, 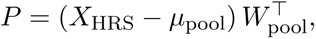 exactly as for the All of Us items (§4.3).

We interpret each kept component twice, once by the pool items that form its core within the pool and once by the HRS items that form its core within HRS. Supplementary Fig. S9 draws the components whose pool-side core includes a construct HRS also measures, the subset on which the two sides can be compared instrument by instrument. We measured cross-encoder agreement of the placements from each HRS item’s core components, exactly as for the pool (§4.11, Supplementary Note S14), and record two labeling decisions there. The placement uses item text only, so screening the HRS cohort forms no part of this test.

### 4.14. Semantic content versus syntax

An item’s *semantic content* is what it asks about, and its *syntax* is the form it is asked in, its stem and grammatical frame. Two tests separate them, one on the wordings the instruments happen to use and one on wordings we generate.

Both batteries ask questions the pool also asks, so the effect of a difference in syntax can be measured rather than assumed. We match an item of *S* to a pool item when their texts agree after case, punctuation and spacing are removed and one is a suffix of the other. That agreement survives when a battery keeps a stem the pool strips or administers a question as a bare fragment. We split matched pairs by whether the two texts are byte-identical, and for each pair compare the set of core components the two versions reach on the same basis by the Jaccard index, against a baseline that pairs each battery item with a random pool item. Byte-identical text must place identically, so that group is a check on the procedure rather than a result. The matching uses text alone, with no construct labels, no instrument identity and no responses.

For the stronger test we generated counterfactual variants that recombine the items’ own semantic content and syntax. We assigned each of the 151 items to one of 13 syntax families, the reusable scaffold with a content slot, labeling each family *neutral* or *content-bearing* by whether the template itself names a construct (Supplementary Note S4, Table S6), and to constructs as in §4.5. Every item served as a seed. For each seed we drew ten donors per variant type, each differing from the seed in both syntax family and construct so that no swap is a no-op, with the donors for syntax variants restricted to neutral families so that the transferred syntax carries no competing content. A *syntax variant* keeps the seed’s content in the donor’s syntax, and a *content variant* keeps the seed’s syntax around the donor’s content, 3,020 variants in all, each recording its content source and its syntax source as ground truth. A large language model (claude-opus-4-8, interactive) generated the variants from one system prompt and one template per variant type, reproduced verbatim in Supplementary Note S5, and we checked each to confirm it changed only the intended dimension. We embedded the variants and placed them on the basis exactly as a battery item is placed (§4.3), with nothing refit. For each variant we record its cosine to its content source and to its syntax source, and summarize each variant type by the median of the two and by the gap between them, per encoder, with content variants split by whether the seed’s syntax is neutral or content-bearing. Supplementary Data 2 provides the seed list, the seed-to-donor manifest and all 3,020 variants.

### 4.15. Screening the All of Us cohort

We drew the 151 items from the All of Us Controlled Tier curated data repository by their survey concept identifiers, which gave 30.5 million answer rows and 79,713 participants with a record for every one of the 151. We treat an answer of *Skip*, *Prefer Not To Answer* or *Don’t Know* as missing rather than as a response. We mapped answers to numbers by the survey codebook (Supplementary Data 1, items_full.csv), and where a participant answered an item more than once we kept the latest by response date.

Age and sex at birth come from the participant record. Participants do not complete the battery in one sitting. The median participant spans 504 days across the 151 answers and the 90th percentile 1,798, so we compute age at each participant’s own reference date, the median date of their answers. We removed 401 of the 79,713: 15 whose sex at birth was not reported and 386 recorded as a generalized difference of sex development, a value the binary coding that the residualization (§4.6) requires cannot represent. No participant lacked a date of birth or a participant record. Of the 79,312 who remain, 22,332 answered all 151 items with no *Skip*, *Prefer Not To Answer* or *Don’t Know*, and these are the *N* = 22,332 analyzed here. Their ages at that date run from 18.0 to 87.1 with a mean of 57.6, so none reaches the 89-and-above range that All of Us top-codes. These responses give the matrix *X*, rescaled and reverse-coded as in §4.6. Every participant has a survey weight of 1 in this cohort, so the weighted estimates coincide with the unweighted ones.

Under every encoder the residualized scores form a single unimodal cloud and the Mahalanobis *D*^2^ tracks a *χ*^2^ reference through the bulk with a heavier upper tail, which the elliptical screen assumes (Supplementary Note S6).

To set the threshold we sweep candidate percentiles and measure cross-encoder agreement at each as the mean pairwise Jaccard overlap of the six outlier sets, against the overlap expected if the screens were independent, *f/*(2 *− f*) for a top fraction *f* per encoder (Supplementary Fig. S2). The sweep shows no knee, so we adopt the 95th percentile, and the consensus outliers are the participants above it under all six encoders (*N* = 510).

For validation we operationalized the established criterion of each of the eight instruments that has one (Supplementary Table S17). We evaluate these rules on the codebook coding, on each instrument’s own response scale and in its original direction, and not on the rescaled and reverse-coded *X*, because the validated scoring is defined on that scale. A participant’s breadth is the number of these eight criteria they meet.

Attribution runs as in §4.7, with each component named by the documented constructs of the battery items that core it (§4.5). For the group we report the mean share per component over the outliers, the number of participants each component carries and the number it is largest for, and the cross-encoder agreement of a participant’s largest component against its between-participant floor. The item-level test of Supplementary Note S9 uses the item partition of the same distance.

### 4.16. Response-style robustness checks

To confirm the outliers reflect reported semantic content rather than a fixed answering style, we relate the outlier score (Spearman) to two indices computed from the responses. *Content extremity* is the mean, over the 151 items, of the absolute standardized response (each item *z*-scored across the cohort)—how far a participant’s answers sit from the typical response, regardless of direction. *Endpoint use*, an extreme-response-style index [49], is the fraction of *polytomous* items (those with at least four observed response levels, so binary and three-level items are excluded) answered at either observed extreme. It is a content-blind, polarity-invariant measure of habitual endpoint marking, the pattern a careless responder produces [50]. Finally, to test whether the screen merely re-ranks the most extreme responders, we rank the whole cohort by endpoint use and, separately, by content extremity, and count how many of the top 510 coincide with the consensus set.

### 4.17. Health-records and access-survey linkage

We linked the cohort to its All of Us electronic health records (Observational Medical Outcomes Partnership common data model) and to the program’s Healthcare Access and Utilization survey. A participant is EHR-linked if the records contain at least one visit, which is 20,972 responders (94%). We use the visit record rather than the observation period to mark linkage because every responder has an observation period in this dataset, so it does not distinguish who has health records. The exposure is the continuous outlier score used throughout, and the consensus outliers are the 510 flagged by all six encoders.

From the survey we derived, per participant, binary indicators of any cost barrier, any structural delay (transportation, time off work, childcare), medication rationing to save money, and lack of a usual source of care. Separately, as a mechanism linking to the discrimination of §2.4, we derived care avoidance attributed to a provider’s race or religion. Their union forms the any-access-barrier indicator. We contrast the consensus outliers with the rest of the linked cohort on each.

From the records we take two views of care use. To capture how care is *used* rather than how much, we compute emergency-department reliance, the share of a participant’s visits made to the emergency department, and relate it to the score by rank regression adjusting for the total number of visits, age, and sex. Because it is a ratio, this quantity is independent of how long or how completely a record spans, which is why it is the record-based signal we rely on. To capture how *much* care is used, we count total visits and relate them to the score adjusting for age and sex. The visit count is reported in the supplement with the caveat that it tracks how completely each record is populated rather than care-seeking (Supplementary Note S8). All linkage analyses ran inside the Researcher Workbench, and no record-level data left it.

### 4.18. Is the care association severity alone?

We compare the cross-instrument distance against two simpler measures built from the same 151 responses. *Content extremity* is the mean absolute standardized response over the items (§4.16), a severity score with no cross-instrument structure in it. The *diagonal distance* is the same component scores with every off-diagonal term of the covariance set to zero, which keeps the reference basis and drops the covariance between components. The third is the screen itself. We take each as the participant’s percentile, averaged over the six encoders for the two computed per encoder, then rank-transform and standardize it so a coefficient is a change per standard deviation. We fit the three together by ordinary least squares on the ranks, as in §4.17, adjusting for age, sex at birth, and for the emergency-department outcome the participant’s total visit count (Supplementary Note S25 and Table S23).

### 4.19. Statistical analysis

We report each quantity in the Results as an effect estimate with a two-sided 95% confidence interval rather than a decision against a fixed significance level. The estimates come from three samples—the complete-case cohort (*N* = 22,332), the consensus outliers within it (*N* = 510), and the health-record-linked subset (*N* = 20,972). Proportions, such as the share of outliers meeting each clinical criterion (§2.3) or reporting each access barrier (§2.6), use Wilson score intervals. The outlier-versus-cohort differences in those proportions use Newcombe intervals, and the barrier risk ratios use Katz log-method intervals. Associations of the continuous outlier score with the response-style indices (content extremity and endpoint use) and with other cohort quantities are Spearman rank correlations. We relate emergency-department reliance and total visit count to the score by rank regression, the former adjusting for the total number of visits, age and sex, the latter for age and sex, and report each as a coefficient with its 95% interval and exact two-sided *P* value. We use rank-based methods because the responses and derived scores are bounded, ordinal, and right-skewed. Because these are estimates with intervals and exact *P* values rather than decisions thresholded on significance, the conclusions do not require a multiple-comparison correction. The analysis code produces interval bounds and *P* values for every reported quantity (Code availability).

### 4.20. Code availability

We provide the analysis code—the ross_proj package, the All of Us example pipeline, and the response-level screening notebook—as supplementary material for review and will make it openly available. Analyses use NumPy, SciPy and scikit-learn, and we fit the projection in JAX [51, 52].

## Ethics

This study is a secondary analysis of de-identified data from the All of Us Research Program, conducted within the program’s controlled-tier Researcher Workbench under its governance framework. All of Us participants provided written informed consent for broad secondary research use of their data, and the program operates under institutional review board oversight [14].

## Data availability

All of Us Research Program data are not publicly downloadable: individual-level survey responses are available only to authorized investigators through the program’s controlled-tier Researcher Workbench, a secure cloud environment, after institutional registration, identity verification, completion of responsible-conduct training, and execution of the Data User Code of Conduct and Data Use and Registration Agreement [14]. No individual-level data leave the Workbench. Consistent with the program’s data-dissemination policy, this study reports only aggregate statistics and de-identified item-level structure, and all response-level analyses were run inside the Workbench. The text of the 151 selected survey items and their response coding is documented in the program’s public survey codebook and reproduced in Supplementary Data 1 (items_full.csv), so the text-level analyses (embedding, the concentrated projection, and construct recovery) are fully reproducible without controlled-tier access.

## Acknowledgments

This research was supported by the Intramural Research Program of the National Institute of Diabetes and Digestive and Kidney Diseases (NIDDK) within the National Institutes of Health (NIH). The contributions of the NIH author(s) were made as part of their official duties as NIH federal employees, are in compliance with agency policy requirements, and are considered Works of the United States Government. However, the findings and conclusions presented in this paper are those of the author(s) and do not necessarily reflect the views of the NIH or the U.S. Department of Health and Human Services.

## Author contributions

**M.A.**: Formal analysis, Investigation, Methodology, Software, Visualization, Writing—original draft, Writing—review & editing. **V.P.**: Conceptualization, Formal analysis, Funding acquisition, Investigation, Methodology, Software, Supervision, Writing—original draft, Writing—review & editing.

