## Supplemental notes for "Scoring respondents on a semantic frame of reference identifies a group that is burdened across many questionnaires and reaches care less easily"

The numbered sections below are the **Supplementary Notes**, cited in the main text as Supplementary Note S1, Supplementary Note S2 and so on. Supplementary Figures, Supplementary Tables and Supplementary Data are numbered separately from them and from the main-text display items.

#### S1 Selected items and constructs

**Supplementary Data 1** (`items_full.csv`) provides the 151 selected items, sorted by construct, each with its AoU concept id, variable name, source instrument, assigned construct, response coding, and full prompt text. The construct assignments follow the manuscript taxonomy (Table S2).

Table S1: **The 151 selected items and the instruments they come from.** Mapped from the survey codebook: 17 validated multi-item instruments and 13 standalone single items, spanning six All of Us survey modules. Full item text, instrument, and response coding are in Supplementary Data 1 (`items_full.csv`). The Mental Health Questionnaire (MHQ) and the World Mental Health Composite International Diagnostic Interview (WMH-CIDI) each span two surveys and contribute different items to each, so no item is counted twice. (EDS: 9 of the instrument's 10 items appear in this extract.)

| Instrument | Acronym | AoU survey | Items |
|---|---|---|---|
| Big Five Inventory–2 Extra-Short Form [1] | BFI-2-XS | bhp | 15 |
| Perceived Neighborhood Disorder (adapted) [2] | NDS | SDOH | 13 |
| UK Biobank Mental Health Questionnaire [3] | MHQ | bhp, ehhwb | 12 |
| Adverse Childhood Experiences (BRFSS) [4] | ACE | ehhwb | 10 |
| Perceived Stress Scale-10 [5] | PSS-10 | SDOH | 10 |
| Patient Health Questionnaire-9 [6] | PHQ-9 | ehhwb | 9 |
| WHO World Mental Health CIDI [7] | WMH-CIDI | bhp, ehhwb | 9 |
| Everyday Discrimination Scale [8] | EDS | SDOH | 9 |
| Short-Form UCLA Loneliness Scale [9] | ULS-8 | SDOH | 8 |
| Modified MOS Social Support Survey [10] | mMOS-SS | SDOH | 8 |
| Discrimination in Medical Settings [11] | DMS | SDOH | 7 |
| Generalized Anxiety Disorder-7 [12] | GAD-7 | ehhwb | 7 |
| Adult ADHD Self-Report Scale v1.1 [13] | ASRS | bhp | 6 |
| Daily Spiritual Experiences (BMMRS) [14] | DSES | SDOH | 6 |
| Neighborhood Social Cohesion (adapted) [15] | SCNS | SDOH | 4 |
| PROMIS Global Health [16] | PROMIS | OverallHealth | 3 |
| Hunger Vital Sign [17] | HVS | SDOH | 2 |
| Standalone single items | — | multiple | 13 |
| **Total (17 instruments + 13 single items)** | | 6 surveys | **151** |

Table S2: **The 151 items organized into 30 constructs.** Each item was assigned to one construct from its prompt text and cross-checked against the source instrument's published structure. Counts are items per construct. The full item-level assignment is in Supplementary Data 1 (`items_full.csv`).

| Construct | $n$ | Example item |
|---|---|---|
| Everyday discrimination | 9 | You are treated with less courtesy than other people are |
| Healthcare discrimination | 7 | A doctor or nurse acts as if he or she thinks you are not smart |
| Depression | 10 | Feeling down, depressed, or hopeless |
| Generalized anxiety | 8 | Feeling nervous, anxious, or on edge |
| Panic & phobia | 3 | With this definition in mind, did you ever in your life have a panic... |
| Perceived stress | 10 | In the last month, how often have you felt that you were unable to... |

*Table S2 (continued)*

| Construct | $n$ | Example item |
|---|---|---|
| Suicidality & self-harm | 4 | Did you ever in your life have thoughts of killing yourself? |
| Psychosis | 4 | Did you ever hear voices that other people did not hear like voices... |
| Mania | 2 | Have you ever had a period of time when you were feeling so good,... |
| Body-focused repetitive behavior | 2 | Have you ever been unable to stop pulling out your hair, eyebrows or... |
| Childhood adversity | 10 | During your first 18 years of life, how often did a parent or adult... |
| Interpersonal & sexual violence | 4 | In your life, have you ever had a partner or ex-partner deliberately... |
| Life-threatening events | 2 | In your life, have you ever been diagnosed with a life-threatening... |
| Extraversion | 3 | I am someone who is full of energy |
| Agreeableness | 3 | I am someone who is compassionate, has a soft heart |
| Conscientiousness | 3 | I am someone who is reliable, can always be counted on |
| Negative emotionality | 3 | I am someone who worries a lot |
| Open-mindedness | 3 | I am someone who is original, comes up with new ideas |
| Loneliness | 8 | I lack companionship |
| Perceived social support | 8 | Someone who understands your problems |
| Neighborhood disorder & safety | 14 | There is a lot of crime in my neighborhood |
| Neighborhood social cohesion | 4 | People in my neighborhood can be trusted |
| Substance use | 6 | Have you smoked at least 100 cigarettes in your entire life? (There... |
| Adult ADHD | 6 | How often do you have problems remembering appointments or... |
| Daily spiritual experience | 6 | I feel deep inner peace or harmony |
| Religious participation | 1 | How often do you go to religious meetings or services? |
| Food insecurity | 2 | Within the past 12 months, the food we bought just didn't last and we... |
| Housing instability | 1 | In the past 6 months, have you been worried or concerned about NOT... |
| Subjective wellbeing | 2 | In general, how happy are you? |
| Self-rated health | 3 | In general, how would you rate your mental health, including your... |

#### S2 Construct labels and instrument structure

We assigned each of the 151 items to one construct from its prompt, then checked the assignment against the published structure of its source instrument. Table S3 accounts for every instrument. Thirteen instruments each measure a single attribute and map to one construct. Nine are unidimensional. Four document published sub-factors that we retain as one broader construct (perceived stress, perceived social support, adult ADHD, and self-rated health), because that broader attribute is the unit relevant to the cross-instrument analysis. The two composite screeners, the UK Biobank Mental Health Questionnaire and the WMH-CIDI, are batteries whose items each screen a different topic, so we assign items by topic rather than to a single factor. The PHQ-9 is a depression instrument whose ninth item screens suicidal ideation, which we assign to suicidality. The 13 standalone items belong to no instrument, and we assigned each from its prompt.

Table S3: **Each construct's grounding in its source instrument.** For every source instrument, its published structure and the construct(s) its items are assigned to in this analysis. Construct lists and item counts are taken directly from the item-level assignment (Supplementary Data 1). "Items" is the number of the instrument's items in the 151-item set.

| Instrument | Items | Published structure | Construct(s) in this analysis |
|---|---|---|---|
| *Multidimensional instrument, factor structure recovered* | | | |
| BFI-2-XS | 15 | Five trait factors, three facets each | Agreeableness, Conscientiousness, Extraversion, Negative emotionality, Open-mindedness |
| *Unidimensional instrument, one construct* | | | |
| NDS | 13 | Perceived neighborhood disorder | Neighborhood disorder & safety |
| ACE | 10 | Childhood adversity index | Childhood adversity |
| EDS | 9 | Everyday discrimination | Everyday discrimination |
| ULS-8 | 8 | Loneliness | Loneliness |
| DMS | 7 | Discrimination in medical settings | Healthcare discrimination |
| GAD-7 | 7 | Generalized anxiety | Generalized anxiety |
| DSES | 6 | Daily spiritual experience | Daily spiritual experience |
| SCNS | 4 | Neighborhood social cohesion | Neighborhood social cohesion |
| HVS | 2 | Food insecurity | Food insecurity |
| *Multidimensional instrument kept as one construct* | | | |
| PSS-10 | 10 | Helplessness and self-efficacy factors | Perceived stress |
| mMOS-SS | 8 | Emotional-informational and instrumental support | Perceived social support |
| ASRS | 6 | Inattention and hyperactivity | Adult ADHD |
| PROMIS | 3 | Physical and mental health | Self-rated health |
| *Composite screener, items assigned by topic* | | | |
| MHQ | 12 | Multi-disorder screening battery | Interpersonal & sexual violence, Depression, Mania, Subjective wellbeing, Life-threatening events, Psychosis |
| WMH-CIDI | 9 | Diagnostic screening modules | Panic & phobia, Psychosis, Body-focused repetitive behavior, Life-threatening events |
| PHQ-9 | 9 | Depression severity, ninth item screens suicidal ideation | Depression, Suicidality & self-harm |
| *Standalone single items* | | | |
| — | 13 | No parent instrument | Substance use, Suicidality & self-harm, Generalized anxiety, Housing instability, Neighborhood disorder & safety, Religious participation |

The BFI-2-XS is the one instrument whose full multidimensional factor structure we recover. Its 15 items fall into five validated trait domains, three facets each [1]. From item text alone, our assignment places all 15 items in the correct domain (Table S4), and the three items we assign to each domain span its three published facets, so the content-based labels reproduce the instrument's factor structure (15 of 15).

Table S4: **The content-based constructs recover the BFI-2-XS factor structure.** All 15 Big Five Inventory–2 Extra-Short Form items, grouped by the construct we assigned from the item text. Each construct corresponds exactly to one of the five validated BFI-2-XS trait domains, and the three items per domain span its three published facets [1]. "(R)" marks reverse-keyed items.

| Construct (= BFI-2 domain) | Published facet | Item text |
|---|---|---|
| Extraversion | Sociability | I am someone who tends to be quiet (R) |
| | Assertiveness | I am someone who is dominant, acts as a leader |
| | Energy level | I am someone who is full of energy |
| Agreeableness | Compassion | I am someone who is compassionate, has a soft heart |
| | Respectfulness | I am someone who is sometimes rude to others (R) |
| | Trust | I am someone who assumes the best about people |
| Conscientiousness | Organization | I am someone who tends to be disorganized (R) |
| | Productiveness | I am someone who has difficulty getting started on tasks (R) |
| | Responsibility | I am someone who is reliable, can always be counted on |
| Negative emotionality | Anxiety | I am someone who worries a lot |
| | Depression | I am someone who tends to feel depressed, blue |
| | Emotional volatility | I am someone who is emotionally stable, not easily upset (R) |
| Open-mindedness | Aesthetic sensibility | I am someone who is fascinated by art, music, or literature |
| | Intellectual curiosity | I am someone who has little interest in abstract ideas (R) |
| | Creative imagination | I am someone who is original, comes up with new ideas |

Six constructs draw items from more than one instrument: depression (PHQ-9 and the MHQ), psychosis (the WMH-CIDI and the MHQ), generalized anxiety (GAD-7 and a standalone item), suicidality (PHQ-9 and standalone items), life-threatening events (the MHQ and the WMH-CIDI), and neighborhood disorder and safety (the NDS and a standalone item). Pooling items that different instruments phrase separately is deliberate, and it is the cross-instrument grouping the concentrated components recover in the main-text construct analysis (Section 2.1).

#### S3 Sentence encoders

Table S5: **The six sentence encoders.** Hugging Face model identifier, embedding dimension $D$, and citation. Each pool item's published text is embedded with each encoder, yielding the item-embedding matrix $M \in \mathbb{R}^{577 \times D}$. The number of components each encoder's basis retains, and the size of their cores, are given per encoder in Table S14.

| Encoder | Hugging Face identifier | $D$ | Reference |
|---|---|---|---|
| bge-m3 | `BAAI/bge-m3` | 1024 | [18] |
| bge-large-en-v1.5 | `BAAI/bge-large-en-v1.5` | 1024 | [19] |
| all-mpnet-base-v2 | `sentence-transformers/all-mpnet-base-v2` | 768 | [20, 21] |
| e5-large-v2 | `intfloat/e5-large-v2` | 1024 | [22] |
| gte-large-en-v1.5 | `Alibaba-NLP/gte-large-en-v1.5` | 1024 | [23, 24] |
| qwen3-embedding-0.6b | `Qwen/Qwen3-Embedding-0.6B` | 1024 | [25] |

#### S4 Syntax families

Testing whether the embedding follows semantic content or syntax means holding one fixed and varying the other, so each item's *syntax*, the reusable scaffold with a content slot, must be known separately from its semantic content. We grouped the 151 items into **13 syntax families** (Table S6), labeling a family *content-bearing* when its template itself names a construct (so semantic content and syntax cannot be separated within it) and *neutral* otherwise. Two cross-instrument syntax overlaps that a naive "different-instrument" donor rule would miss collapse here: the Everyday and Medical-Settings discrimination instruments share one syntax, as do PHQ-9 and GAD-7. In the test (§4.14), the syntax variant draws its donors only from neutral families, and we report content variants split by whether the seed's syntax is content-bearing.

Table S6: **The 13 syntax families.** Each of the 151 items belongs to one syntax family. A family is *content-bearing* (CB) when the template itself names a construct, *neutral* otherwise. EDS+DMS and PHQ-9+GAD-7 each collapse to a single shared syntax.

| Frame family | Type | $n$ | Source instruments | Example template |
|---|---|---|---|---|
| discrimination | CB | 16 | DMS+EDS | "You are treated with less ____ / a ____ acts as if ____" |
| childhood | CB | 10 | ACE-BRFSS | "During your first 18 years of life, ____" |
| neighborhood | CB | 18 | NDS+SCNS+single items | "____ in my neighborhood / my neighborhood is ____" |
| social support | CB | 8 | mMOS-SS | "Someone to ____" |
| food scarcity | CB | 2 | HVS | "Within the past 12 months, . . . money to buy/get more" |
| housing | CB | 1 | single items | ". . . worried about NOT having a place to live" |
| trait | neutral | 15 | BFI-2-XS | "I am someone who ____" |
| symptom | neutral | 16 | GAD-7+PHQ-9 | "[bare symptom phrase]" |
| stress frequency | neutral | 10 | PSS-10 | "In the last month, how often have you felt ____" |
| self state | neutral | 14 | BMMRS+ULS-8 | "I feel ____ / I lack ____" |
| frequency behavior | neutral | 7 | ASRS+single items | "How often do you ____" |
| lifetime | neutral | 31 | MHQ+WMH-CIDI+single items | "Have you ever / Did you ever ____" |
| self rating | neutral | 3 | PROMIS | "In general, how would you rate your ____" |

#### S5 Semantic content versus syntax

**Content-versus-syntax results.** Both variant types sit closer to their content source than to their syntax source (Fig. S1). A syntax variant stays close to its seed, the item whose semantic content it kept, and far from the donor whose syntax it took (panel a). A content variant does the reverse and moves to the donor whose semantic content it carries (panel b). Against its two sources, each variant sits far closer to the content source (gte-large median cosine 0.77 versus 0.14 for syntax variants and 0.73 versus 0.21 for content variants). The content−syntax gap stays positive for every one of the six encoders, from +0.62 (qwen3) to +0.44 (e5-large, panel c). Semantic content survives a wholesale change of syntax, and follows when it is swapped in. The one systematic exception is content-bearing syntax—templates that themselves name a construct, such as ". . . in my neighborhood" or "During your first 18 years of life," where semantic content and syntax cannot be separated by design. For content variants built on such syntax the two sources pull closer together (gte-large median cosine 0.66 versus 0.33), an attenuation rather than a collapse. We place variants on the pool reference basis exactly as battery items are placed. The same measurement without the pool-standard-deviation division returns a gap essentially unchanged from the published one, so the result does not depend on that calibration.

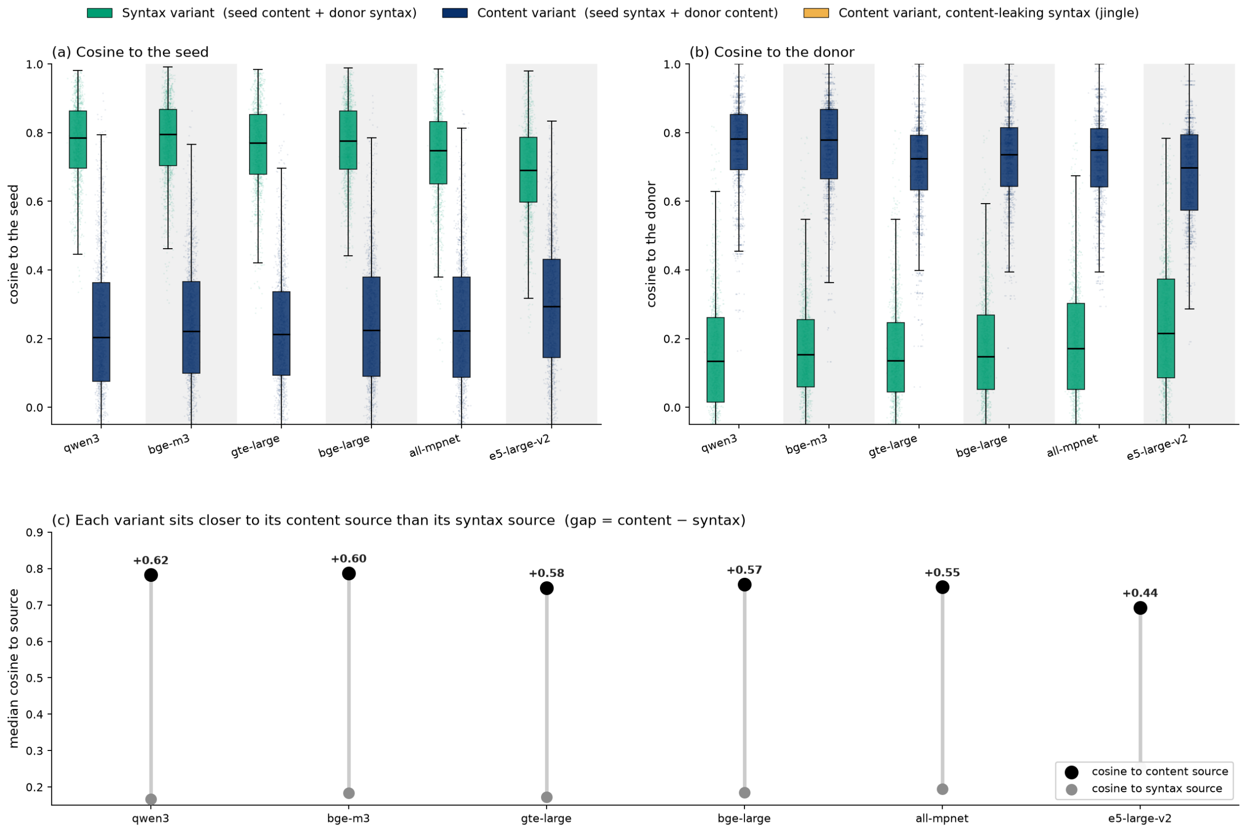


Figure S1: **Semantic content, more than syntax, drives the grouping.** Counterfactual variants recombine the items' own semantic content and syntax: a *syntax variant* keeps the seed's semantic content in a donor's syntax and a *content variant* keeps the seed's syntax with a donor's semantic content, every donor differing from the seed in both. Each variant is projected onto the pool reference basis, exactly as a battery item is and with nothing refit, and compared, by cosine, to its *content source* and its *syntax source*. Box plots span 151 seeds and 3,020 variants across six encoders, ordered by effect size. (a) Cosine to the *seed*: syntax variants (semantic content kept) stay high, content variants (semantic content swapped in) fall away, and content variants on content-bearing syntax (jingle) remain partly tied to the seed. (b) Cosine to the *donor*, the mirror image. (c) Per encoder, the median cosine to each variant's content source (black) and syntax source (gray), pooled over the clean variants (the jingle case excluded). The gap between them—semantic content minus syntax—is positive for every encoder, from +0.62 to +0.44, the margin by which semantic content prevails over syntax.

**Content-versus-syntax generation prompts.** The 3,020 content-versus-syntax variants were generated by `claude-opus-4-8` (interactive) following the rewriting instructions below—one per variant type. Each variant pairs a seed with a donor chosen by the deterministic rule of §S4: the donor differs from the seed in *both* syntax family and construct, and for the syntax variant the donor's syntax is neutral. **Supplementary Data 2** provides the full seed-to-donor manifest, every rewritten prompt, and all 3,020 variants (`content_grammar_variants/`: `manifest.csv`, `generated_items_v2.csv`, and the batch `.jsonl` files). *System prompt:*

```
You rewrite survey questionnaire items with surgical control over wording. You change
ONLY what each task asks you to change and never alter a question's meaning, severity,
timeframe, or specificity unless explicitly told to. You always respond with strictly
valid JSON and nothing else.
```

*Cell A — content preserved, grammatical frame transferred:*

```
TARGET survey item (id: {seed_id}):
  "{seed_prompt}"

STYLE-EXAMPLE items from the same questionnaire battery (different topics):
{donor_block}

Task: rewrite the TARGET item once for EACH style example, so that the rewrite means
EXACTLY the same thing as the TARGET but mimics the grammatical structure / phrasing of
that style example (e.g. question vs statement, the opening stem, sentence form). Change
ONLY the sentence form. Do NOT change the topic, severity, timeframe, or specificity of
the TARGET. If a style would force a meaning change (e.g. a different timeframe), keep
the
TARGET's meaning and approximate the structure.

Respond with JSON: a list of {n} objects {"style_id": <id>, "rewritten": <text>}.
```

*Cell B — frame preserved, content swapped:*

```
TEMPLATE survey item (id: {seed_id}) -- copy its sentence structure exactly:
  "{seed_prompt}"

TOPIC items (the subject matter to ask about instead):
{donor_block}

Task: for EACH topic item, write a new survey item that keeps the TEMPLATE's exact
grammatical structure / phrasing (same stem, same sentence form) but asks about the
TOPIC
item's subject matter instead of the template's. Preserve the template's form; swap in
the
topic's content.

Respond with JSON: a list of {n} objects {"topic_id": <id>, "rewritten": <text>}.
```

#### S6 Cross-encoder outlier agreement

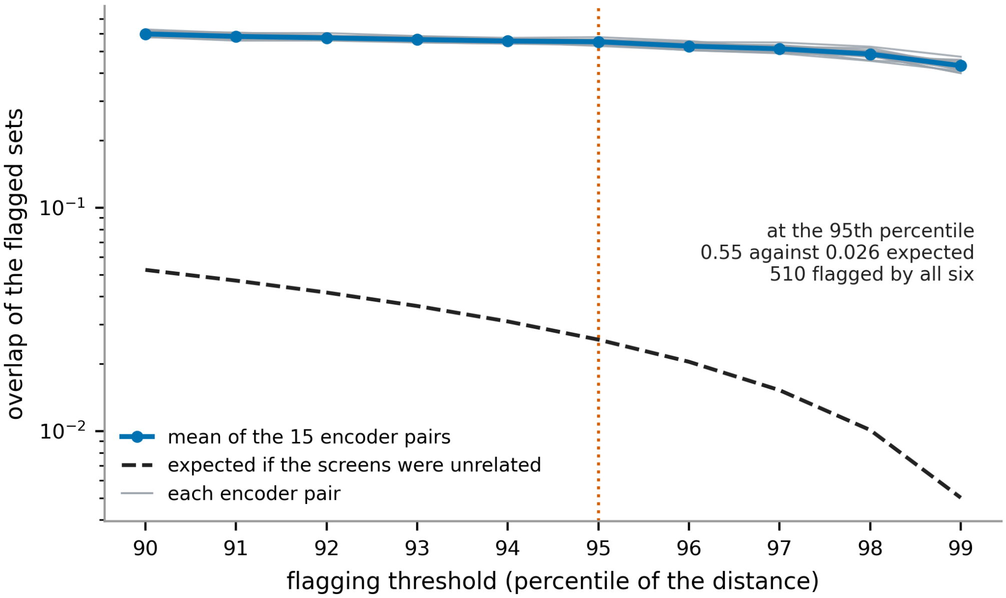


Figure S2: **Cross-encoder outlier agreement versus threshold.** Overlap of the flagged-outlier sets for each of the 15 encoder pairs (gray) and their mean (blue), as the empirical percentile threshold that defines an outlier varies, against the overlap two unrelated screens of the same size would give (dashed). Overlap is 1 for the same set of people and 0 for no one in common. The 15 pairs stay close together and their mean stays far above the independence null across the whole sweep with no knee, declining only gently toward the most extreme cutoffs. We therefore screen at the 95th percentile (top 5%) per encoder as the canonical top tail, marked, and take the consensus across all six for downstream analyses (§2.3).

**The score distribution supports an elliptical screen.** Across all six encoders the residualized component scores form a single unimodal cloud, and Hartigan's dip test does not reject unimodality (dip statistic $< 0.002$) despite the large sample. The Mahalanobis $D^2$ tracks the $\chi^2$ reference through the bulk and departs only in a heavier upper tail (about $1.6\times$ the $\chi^2$ 99th percentile). That is the signature of a skewed adversity distribution rather than of multimodal or non-elliptical structure, and it is why we define outliers by the empirical upper tail rather than by a nominal $\chi^2$ cutoff (Methods §4.15).

#### S7 The outliers' breadth is not a response-style or severity artifact

The breadth of the consensus outliers across instruments (main-text §2.3) could in principle reflect how participants use the response scale rather than genuine burden.

**The outliers report genuine distress, not a fixed answering style.** The "extreme responder" is a well-documented threat to validity: a participant who habitually marks the most extreme option on every question, out of haste or a fixed answering style rather than from what the question asks [26, 27]. To separate genuine distress from this habit we summarized each participant in two ways. *Content extremity* captures *what* a person reported: how far, on average, their answers sit from the typical response (their mean absolute standardized response), so it is high for someone who genuinely endorses more burden than most people do. *Endpoint use*, a standard extreme-response-style index [27], captures *how* a person answered: how often they pick the very end of the scale, whatever the question and in either direction. It is high for a content-blind habit of always choosing the most extreme option and says nothing about what was reported.

We then related each participant's outlier score—their averaged Mahalanobis percentile across the six encoders—to both measures across the cohort. Were the outliers careless extreme responders, the score would climb with endpoint use, but it does not. It correlates strongly with content extremity (Spearman $\rho = 0.77$, 95% CI [0.77, 0.78], with outliers averaging 1.37 vs. cohort 0.79) yet is, if anything, *negatively* related to endpoint use ($\rho = -0.35$, 95% CI [−0.36, −0.34]), on which the outliers (0.49) are indistinguishable from the cohort (0.48). Ranking everyone by endpoint use alone recovers only 7 of the 510, so the screen is not simply re-ranking habitual endpoint markers.

**The screen is not a relabeled severity sum.** The Mahalanobis distance could in principle just rediscover the participants with the highest average burden, so we ranked everyone by that same content extremity and asked how many of the consensus set it recovers: fewer than half (244/510). The screen therefore does not reduce to "who deviates most" on a single flat average of the items. As §S9 shows, it differs by integrating and standardizing the responses component by component, not that it exploits any configuration across components.

#### S8 The under-served group: barriers and the visit-count caveat

Table S7: **Healthcare-access barriers, consensus outliers versus the rest of the EHR-linked cohort.** Among access-survey respondents (outliers $N$ = 452, rest $N$ = 19,858), the percentage reporting each barrier, the difference in percentage points with Newcombe 95% confidence intervals, and the risk ratio with Katz log-method 95% confidence intervals. Every component is more common among the outliers. Brackets give the corresponding confidence interval.

| Barrier | Outliers (%) | Rest (%) | Difference (pp) | Risk ratio |
|---|---|---|---|---|
| Any access barrier | 82.3 | 44.2 | +38.1 [+34, +41] | 1.86 [1.78, 1.95] |
| Cost barrier | 67.3 | 27.9 | +39.3 [+35, +44] | 2.41 [2.25, 2.58] |
| Structural delay (transport, time off work, childcare) | 54.6 | 16.5 | +38.1 [+33, +43] | 3.30 [3.02, 3.61] |
| Medication rationing to save money | 46.0 | 22.8 | +23.3 [+19, +28] | 2.02 [1.82, 2.24] |
| No usual source of care | 10.8 | 3.7 | +7.2 [+5, +10] | 2.96 [2.25, 3.89] |
| Care avoidance, provider race or religion | 31.9 | 8.7 | +23.2 [+19, +28] | 3.68 [3.19, 4.24] |

We rely on two signals, the self-reported access barriers (Table S7) and the emergency-department reliance reported in the main text (§4.17). Raw visit counts tell a consistent-looking but confounded story that we report here for completeness. Median total visits fall from 21 in the lowest-scoring tenth of the cohort to 1 in the highest, and the age- and sex-adjusted rank regression is negative ($\beta = -0.029$, 95% CI $[-0.042, -0.015]$, $p = 3.0 \times 10^{-5}$). This does not survive the two completeness checks. Adjusting additionally for the observation-window length removes it ($\beta = -0.007$, $[-0.016, +0.003]$, $p = 0.17$), and the visits-per-observation-year rate shows no association with the score ($\beta = +0.006$, $[-0.009, +0.021]$, $p = 0.42$). Because a participant's observation window in All of Us is set largely by which sites contributed data and when, the falling counts track how completely each record is populated rather than how much care a person seeks (Fig. S3). Emergency-department reliance, a ratio, is by construction free of this confound, and it rises with the score holding the total number of visits, age, and sex fixed ($\beta = +0.054$, $[+0.045, +0.062]$, $p = 4.0 \times 10^{-36}$).

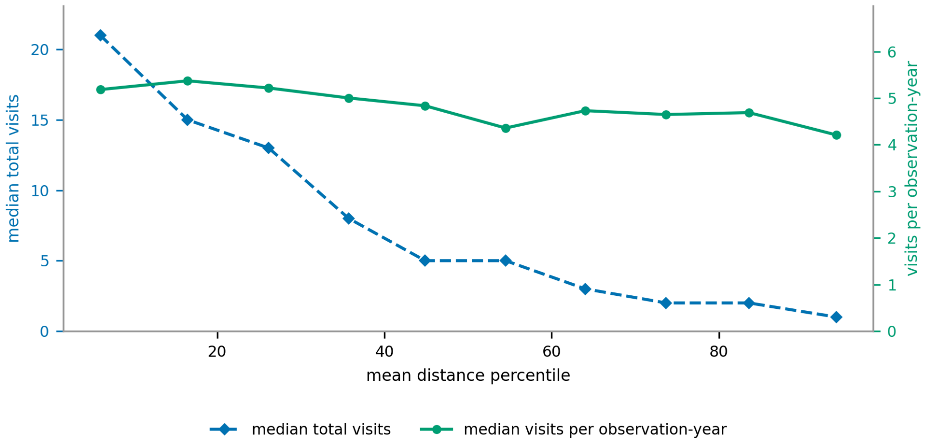


Figure S3: **Raw visit counts fall with the outlier score, but the per-year rate does not.** Participants are grouped into ten equal-size bins by their cross-instrument outlier score, and each marker sits at its bin's mean Mahalanobis percentile on the horizontal axis, from most typical to most outlying (EHR-linked cohort, $N = 20{,}972$). The median total number of electronic-health-record visits falls (left axis), while the median visits per observation-year stays flat (right axis). The falling counts reflect shorter, less complete records rather than reduced care-seeking.

#### S9 The multivariate view integrates the responses rather than detecting otherwise-invisible cases

A natural question is whether the cross-instrument distance flags participants that instrument-by-instrument analysis of the same responses could not. It does not. Standardizing every item across the cohort, not one of the 510 consensus outliers is beyond the reach of both a single item and a single construct, so each would be identifiable from an individual instrument examined on its own.

Two things qualify how demanding that test is. Responses are discrete and bounded, so one category can hold far more than five percent of a cohort and an exact top five percent does not exist. Taking the threshold inclusively, a single item's top band covers a median 12.8% of the cohort, which makes reachability by one item easy to satisfy. Taking it strictly, 509 of the 510 outliers remain reachable by one item while only 48.6% of the cohort is, and an outlier is reached by a median of 13 items at once against a cohort median of none. The outliers are therefore conspicuous on many items rather than marginal on one, under either definition.

The covariance between components does change which participants the screen selects. A diagonal distance computed from the same component scores with every off-diagonal term set to zero ranks participants similarly but not identically, Spearman $\rho = 0.74$ to $0.80$ across the six encoders. Its own top five percent overlaps the full distance's with a Jaccard index of 0.36. Of the 510 consensus outliers, 360 to 381 (71 to 75%) fall in that diagonal tail, so about a quarter of the group would not be flagged without the covariance. The diagonal distance still uses every component the battery reaches, so the contrast is between accounting for the covariance among components and not, rather than between instruments alone and instruments together.

A per-component view points the same way. Each component of a participant's distance is either extreme on its own or extreme only given that participant's other components (Methods §4.7), and across the six encoders 40 to 45% of the distance comes from the latter.

The sharpest form of the question is whether an outlier survives the removal of everything individually conspicuous about them. For each participant we set every item on which they exceed that item's own 95th percentile to the cohort mean, hold the projection, the covariate coefficients, the covariance and the flagging threshold at their values on the unmodified cohort, and recompute the distance. A participant who is still above the threshold afterwards then has no item above its own 95th percentile anywhere in the battery and is nonetheless in the top five percent of the distance. This neutralizes a median of thirteen items per participant. 190 **of the** 510 **remain above the threshold under at least four of the six encoders and** 87 **under all six.** Capping each item at its own 95th percentile instead of setting it to the mean, which removes the extremeness but leaves the elevation, gives 150 and 67.

Two controls bound what that means. Applying the identical operation to 3,000 unflagged participants moves 0.1% of those below the threshold above it, so the operation does not inflate the distance on its own. Neutralizing a random set of items of the same size instead leaves 90.4% of the flagged participants above the threshold, against 41.1% when the individually extreme items are the ones removed, so the identity of the removed items matters, not the number.

The participants who survive are the diffuse ones. Summing the item contributions within constructs, the construct carrying the most of their distance takes a smaller share than it does for those who drop out (18% against 22%), the encoders agree less often on which construct that is (4.5 of six against 5.1), and they had more individually extreme items to begin with (16.5 against 12.8), so the test removed more from them and they remained. An accumulation of many moderate elevations therefore holds them above the threshold, not any particular combination of them. Six participants are in that position with no intervention at all, having two or fewer individually extreme items, and one of the six has none.

These are constructed profiles rather than observed ones. The participants' own answers did include those thirteen items, so they are not undetectable, and the test establishes that being an outlier does not depend on them. Taking the instruments together therefore does not detect otherwise-invisible cases. It integrates 151 items into a single, coherent, small group of the broadly and multiply burdened, who would otherwise be dispersed across many separate instrument-level flags. It attributes each outlier to its driving items, and it surfaces adverse exposures, such as discrimination and interpersonal violence, that no validated criterion covers.

**The five clinically invisible outliers under the same operation.** The five consensus outliers who meet none of the eight validated criteria (main-text §2.4) all fall below the threshold under this operation, under every one of the six encoders. Neutralizing the individually extreme items one at a time, largest first, drops the flag after a median of one to four items per participant, taking each participant's median over the six encoders (the five medians are 1, 2, 3, 3.5 and 4). Taken the other way, as each encoder's median over the five, it is two to three. Generator `all_of_us_codes/five_items_to_drop.py`.

#### S10 Core components: characteristic versus reconstruction

An item's core components (Methods §4.2) come either from its raw projection row, the components it is *reconstructed* from, or from the row standardized to unit variance per component, the components that *distinguish* it. On the pool the two agree on everything we report. The item-defining and component-defining cores nest at 0.96 under both (gte-large exemplar). This nesting is expected rather than independent confirmation, because both cores come from the same projection $P$. An item in a component's core is extreme on that component, so it holds a large standardized weight there, and so it tends to include that component among its own core components. They differ only in a shared background. The raw definition pulls one of the four highest-variance components, which rest on depression, openness, sleep, and physical quality of life, into 64% of item cores, against 23% under standardization. The openness component is the clearest case. The five Overall Anxiety Severity and Impairment Scale (OASIS) anxiety items place 17 to 26% of their squared weight on it, and it is the leading core component for two of them under the raw definition, whereas the standardized definition returns anxiety components alone. We therefore standardize each component to unit variance before taking an item's core components.

#### S11 Where core-component placement follows content over the instrument label

Methods §4.9 defines how we measure whether an item's core components (§S10) land on its own semantic content. On the gte-large exemplar it holds for 87% of all 577 pool items against a label-shuffle chance rate of 40%. Split by whether an item cores a component, it holds for 88% of the items that do, chance 35%, and 84% of the items that do not, chance 53%, so the placement is almost as faithful for the items that define no core as the rest (Fig. S4b).

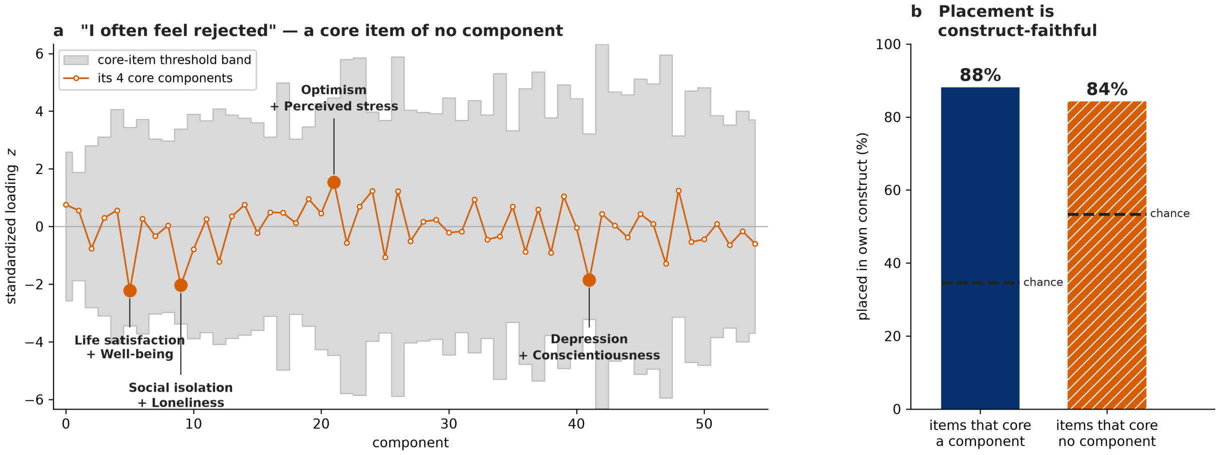


Figure S4: **Core components place every item, including those that define no component. (a)** The standardized loadings of the loneliness item *I often feel rejected* (De Jong Gierveld) on all 55 gte-large components. The item stays inside every component's core-item threshold band (gray), so it is a core item of no component, yet its own *core components*, the four components on which it stands out most, fit what the item asks. Each is named by the leading constructs of its own core items, one of them holding loneliness alongside social isolation. **(b)** For the whole pool, the fraction of items whose own documented construct appears among the constructs of their core components, shown for items that core a component and for the items that core none, against a label-shuffle chance rate (dashed). An item never counts its own membership of a core, so the first group is not favored by construction. The placement is construct-faithful for both groups and nearly as often for the items that core nothing (main-text §2.1, Methods §4.9).

Of the 75 items whose documented construct does not appear among their core components, 26 core no component, and these divide into four kinds. Seven are Big Five personality items, which the pool covers with a single instrument (the 50-item IPIP Big-Five Factor Markers, 28), so they spread onto affect and psychopathology components rather than a personality one, the clearest limitation of the pool. A second group lands on a component whose content matches the item even though its instrument gives it another label, so the placement follows content over label. The CES-D item on poor appetite lands on food insecurity, a PROMIS global-health item that asks about feeling anxious, depressed or irritable lands on anxiety, and the PROMIS Sleep Disturbance item *I felt worried at bedtime* lands on anxiety. A third group is reverse keyed, its positively worded items landing on elevated-mood or self-regard components, as when *I usually come through difficult times with little trouble* lands near optimism and *I have felt active and vigorous* near mania. The remainder are simply diffuse, with generic content that concentrates nowhere, among them four of the six Brief Resilience Scale items and three Perceived Stress Scale items, the instrument that finds no single home elsewhere in this work as well. The first group is a genuine gap and the rest are largely the content-over-label behavior described in the main text. The 84% is therefore a conservative floor. Panic disorder, social phobia and agoraphobia have no dedicated instrument in the pool, so a panic item landing on the anxiety components counts as a disagreement although its content is anxiety. All 75 disagreements are provided as **Supplementary Data 6** (`faithfulness_disagreements.csv`), one row per item with its documented construct, the constructs of its core components, and whether it anchors a core, the property that separates the 26 examined here from the 49 that anchor one.

#### S12 Cross-encoder reproduction of core components

On the component side, Table S8 gives the breakdown behind main-text Fig. 3a, under the reproduction rule of Methods §4.11. Of the 20 components whose core rests on one or two constructs, 18 reproduce in at least four of the six encoders and 14 in all six, against 21 and 13 of the 35 components whose cores span three or more.

Table S8: **Cross-encoder reproduction of the 55 gte-large components, by the number of distinct documented constructs in the component's core.** A component reproduces in an encoder when its matched core there clears the permutation threshold. Cores resting on one or two constructs recur most often and reproduction falls as a core spans more.

| Constructs in the core | Components | Reproduce in $\geq 4$ of 6 | Reproduce in all 6 |
|---|---|---|---|
| 1 | 13 | 11 | 10 |
| 2 | 7 | 7 | 4 |
| 3 | 12 | 9 | 7 |
| 4 | 7 | 4 | 2 |
| 5 or more | 16 | 8 | 4 |
| All | 55 | 39 | 27 |

Which constructs those reproducing components rest on is a separate question from how many reproduce, and **Supplementary Data 5** (`component_reproduction.csv`) answers it. It gives all 55 components, each with the number of encoders it reproduces in, its core items, and the documented constructs of those items with the item count for each, so all 55 are there rather than only the 39 above the four-of-six threshold. The components reproducing in all six rest on depression, sleep, anxiety, social support, social isolation, neighborhood disorder, food insecurity, adverse childhood experiences, trauma exposure, psychosis, suicidality, and everyday, major and healthcare discrimination, so every construct the main text names as socially and clinically central is covered by one of them. Those reproducing in fewer are mostly the components whose cores spread over many constructs at once.

Figure S5 shows all 55 as a grid, on the card layout of main-text Fig. 2a but shaded by how widely the component reproduces, so the two displays can be compared with each other. Figure 2a shows only the 32 components whose cores rest on at most three documented constructs, a presentational cut that keeps the card text legible at the printed figure width, and the ones it leaves out are those whose cores draw on four or more constructs, which are also the least reproducible here. Each component's full core, with the question text of every core item, is provided per encoder in **Supplementary Data 4**.

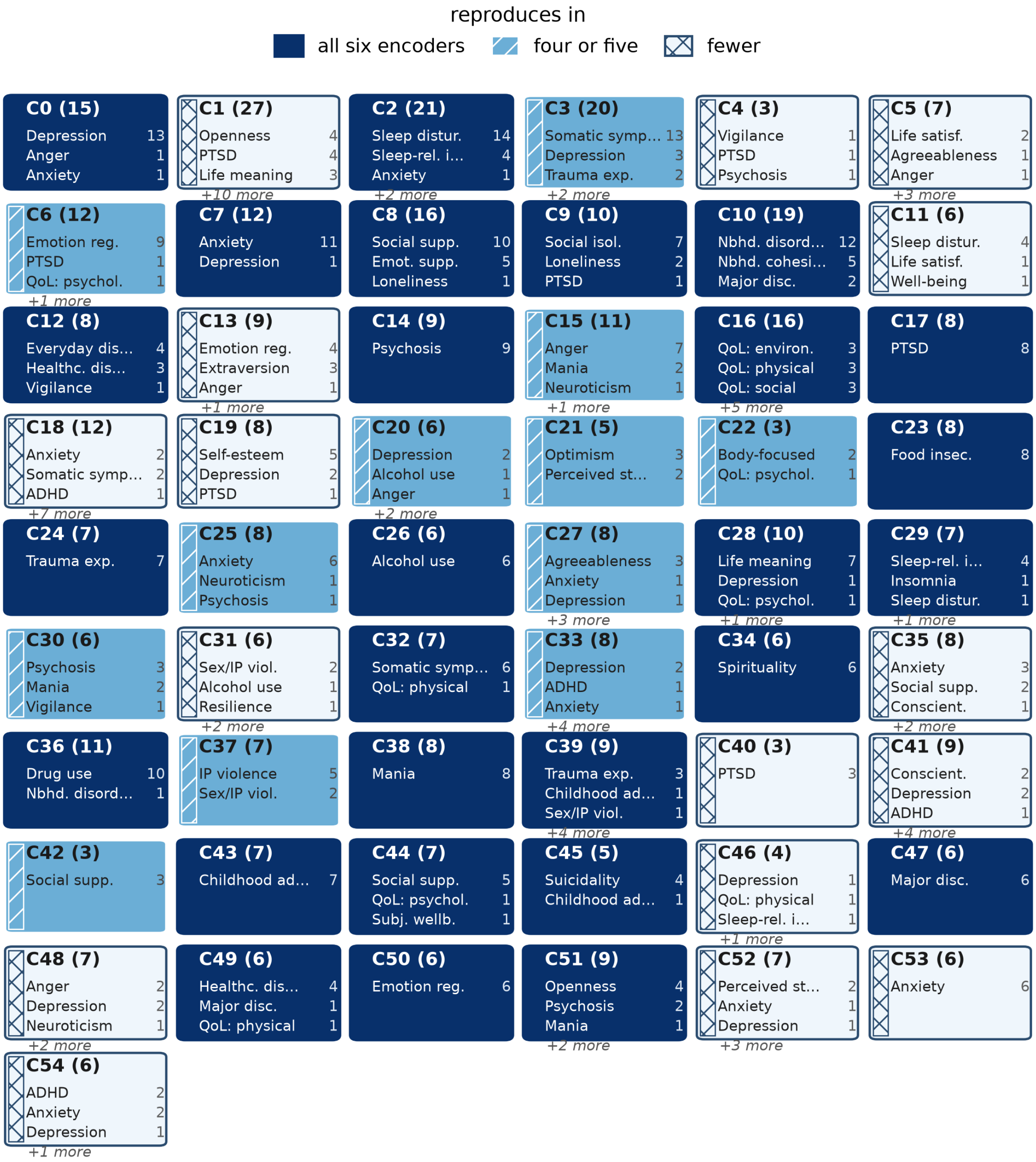


Figure S5: **All 55 components of the reference basis, and which of them reproduce across encoders.** Each card gives the component, its core size in parentheses, and the documented constructs of its core items with the number of core items assigned to each, on the layout of main-text Fig. 2a, which shows only the 32 whose cores rest on at most three constructs. Where a core draws on more than three the two largest are named and the remainder counted. Shading and the texture strip give the number of encoders the component reproduces in, on the same three-way encoding as main-text Fig. 3a. Reproduction is the label-free core-item matching of §S12, and the constructs enter only afterwards.

Reading the projection from the item rather than the component (§S10), we asked how far an item's core components depend on the encoder. For each item we collected the constructs of the core items of each of its core components, giving the item a set of constructs under each of the six encoders, and measured their agreement by the mean pairwise Jaccard across the fifteen encoder pairs, against a null that permutes item identities. Over all 577 items the mean pairwise Jaccard is 0.38 against 0.11 by chance. The agreement tracks how sharply an item is defined. It is 0.41 for items that core some component and 0.27 for items that core none, a narrow gap given that the second group defines no component at all, and it declines monotonically with the number of core components an item has (Table S9). Items with a single core component reproduce at 0.68, so the attribution is stable where an item is concentrated and loosens as an item spreads over many weak components.

Because a component contributes its whole construct profile, the sets compared here are large, a mean of 7.8 constructs, which raises the chance level along with the agreement. Weighting each construct by its share of the profile rather than treating the profile as a flat set gives the same picture with a much lower chance level, an agreement of 0.36 against 0.06, and a steeper gradient with sharpness (Table S9). Neither form depends on designating a single construct for a component, which the main text does not do.

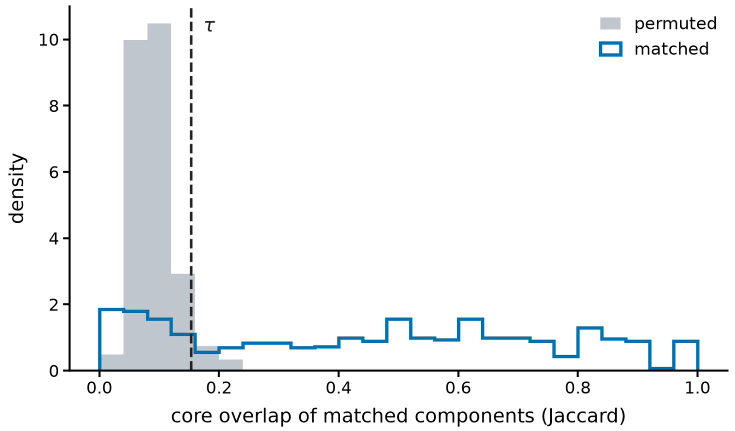


Figure S6: **The threshold that decides whether a component reproduces.** Core overlap between components matched across the fifteen encoder pairs (blue), against a null that permutes item identities before matching and rematches (gray). Matched cores share a median Jaccard of 0.48 against 0.08 under the null, and the two distributions barely meet. The threshold $\tau = 0.15$ is the null's 95th percentile, and a matched pair counts as reproduced above it (main-text Fig. 3a, Methods).

Table S9: **Cross-encoder reproduction of an item's core-component constructs, binned by the item's average number of core components across the six encoders.** Agreement is the mean pairwise Jaccard of the construct sets over the fifteen encoder pairs, and the weighted column is the same comparison with each construct weighted by its share of the profile it comes from. Concentrated items reproduce well and diffuse items much less, under either form. The final row gives the chance level from the permutation null.

| Core components | Items | Mean pairwise Jaccard | Share-weighted |
|---|---|---|---|
| 1 | 78 | 0.68 | 0.73 |
| 2 | 219 | 0.39 | 0.40 |
| 3 to 4 | 259 | 0.29 | 0.23 |
| 5 or more | 21 | 0.27 | 0.18 |
| All items | 577 | 0.38 | 0.36 |
| Chance | 577 | 0.11 | 0.06 |

#### S13 Stability of a response-based structure

The Introduction states that a structure estimated from responses is a property of the cohort that produced it. This section gives the sweep behind that claim, by the resampling procedure of Methods §4.12. A factor model and principal component analysis behave alike. Both agree between two disjoint halves at 0.29 when each half holds one hundred respondents, and reach only ≈0.85 at 11,000 per half, so the grouping a psychometrician would recover depends on how many people answered and on which people they were. The text-derived components do not enter the sweep.

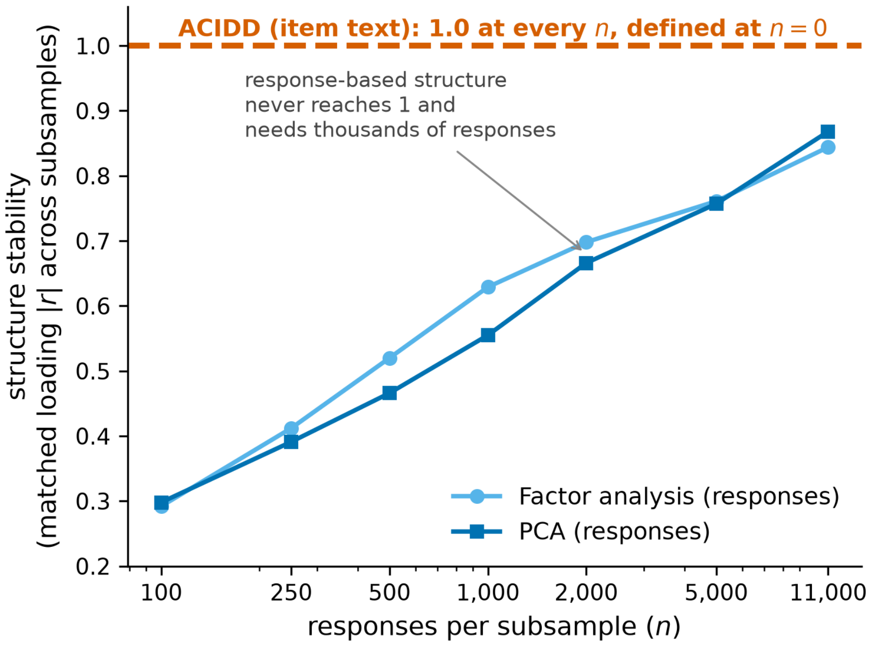


Figure S7: **A response-based structure is not portable and a text-derived one is.** Stability of the recovered item grouping—the mean matched loading correlation between two disjoint subsamples—as the sample grows, for a response factor model and for PCA, against the text-derived structure. The response-based structure is barely reproducible at small samples and reaches only ≈0.85 at 11,000 responses per half. The text-derived structure comes from the item text, so it is identical at every sample size and defined with no responses at all.

#### S14 The HRS battery under all six encoders

A component is kept for a battery when one of its items cores the component or stands out most on it (Methods), and HRS keeps 21 of the 55 components under the exemplar encoder, 18 to 21 of the 49 to 55 across the six. A battery of 35 items therefore occupies a small part of a basis built from 577, and parallel analysis on the projected items says the same: their geometry supports three to five dimensions, against four to twelve for the 151 All of Us items. Every construct HRS asks about is covered by a component it keeps under all six encoders, and none is confined to the components it drops, which are led by content its instruments do not ask about. A battery's set of kept components is itself only moderately stable across encoders. Matching components between encoder pairs by core-item overlap alone and comparing the keep decision, the two agree for 75% of matched components against 54% expected if the decisions were independent (Cohen's $\kappa = 0.46$ over the fifteen pairs), so the kept set is reproducible in the constructs it covers rather than component by component. Figure S8 shows the placement in full, every HRS item against every component under each encoder, with the dropped components marked.

Two labeling decisions are recorded here. HRS documents its eight neighborhood items as two instruments, social cohesion and physical disorder, which the item file we work from collapses under one label, so we split them again by the published item text. The pool documents the same two constructs under shorter strings, neighborhood cohesion and neighborhood disorder, which we rename to the batteries' spelling where the three are compared. It is the only label pairing used anywhere in this work, and every other comparison uses each battery's documented labels as they stand.

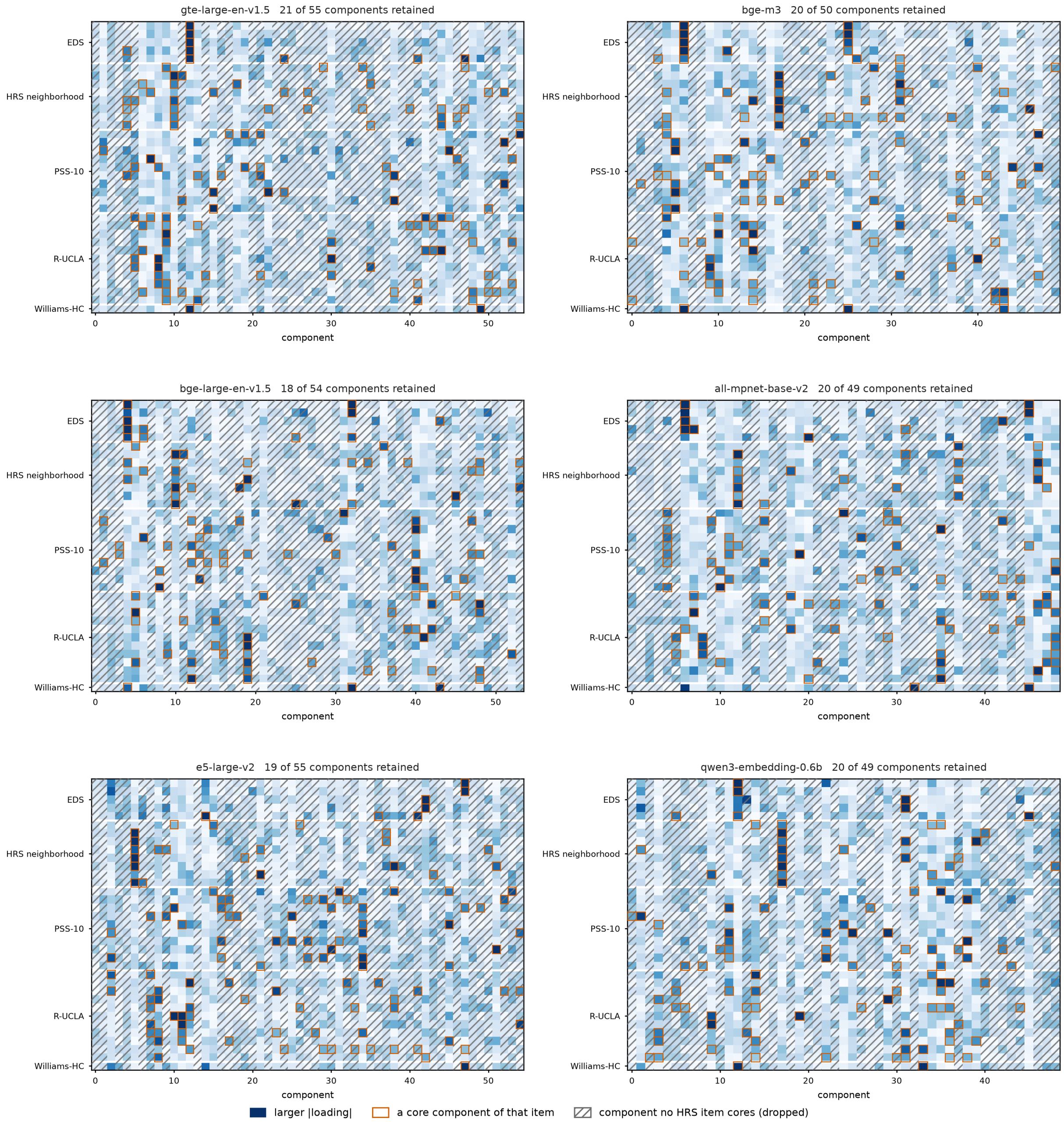


Figure S8: **Every HRS item against every component, under each encoder.** Rows are the 35 HRS items grouped by instrument, columns the components of that encoder's basis in index order, and shade is the item's standardized loading magnitude on the component, the quantity the placement rule uses. A cell is outlined when the component is one of that item's core components. A component is kept for the battery when one of its items cores it or stands out most on it, so the hatched columns, which have no outlined cell, are the components HRS never reaches. The kept count is given above each panel.

This section gives the two-side comparison of Fig. S9 (Methods §4.13) under the five encoders that figure does not show, and the cross-encoder agreement behind the statement that a thin

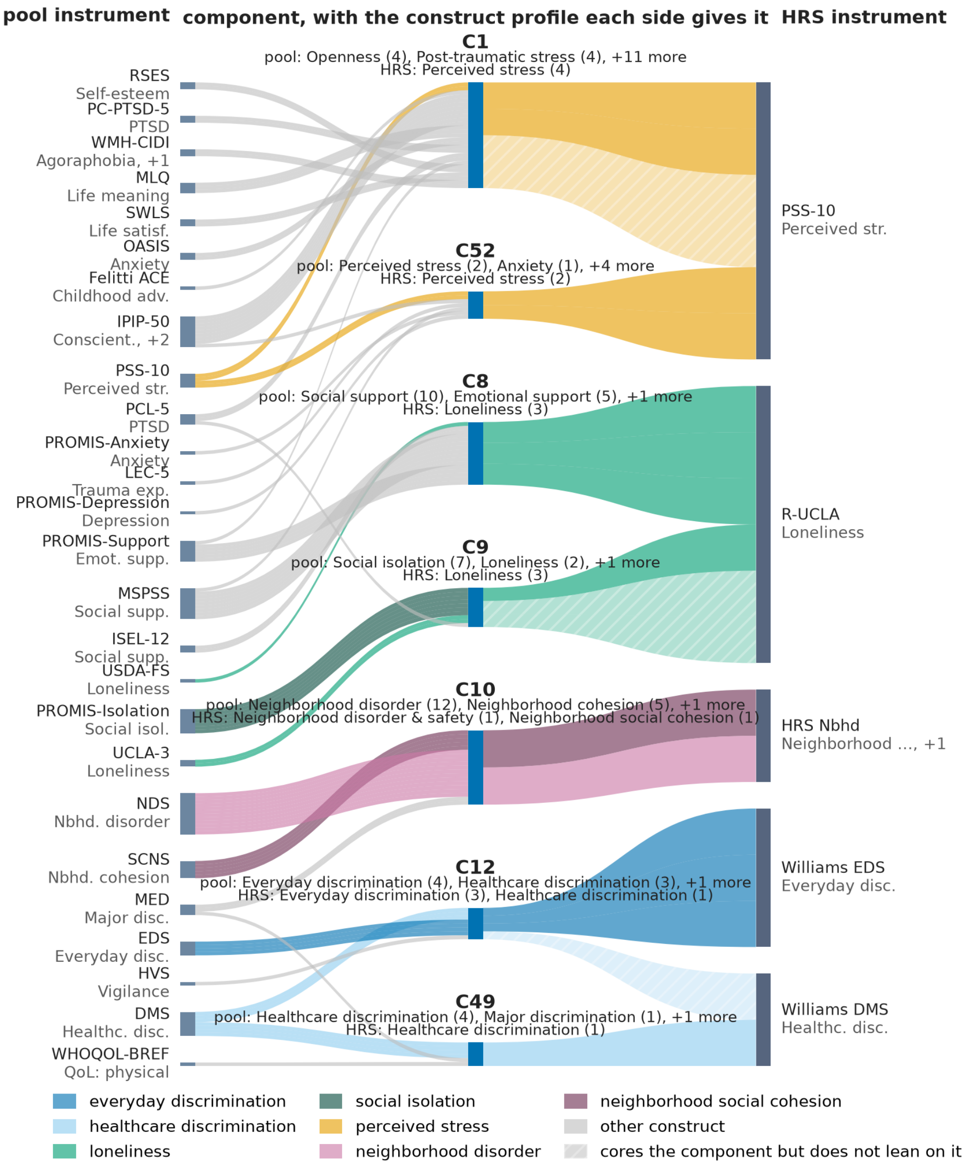


Figure S9: **The basis places a battery outside the pool where its documentation says it belongs.** Laid out as in main-text Fig. 4, with the pool instruments on the left and the Health and Retirement Study (HRS) battery on the right (gte-large exemplar). The components shown are those whose pool-side core includes a construct HRS also asks about, so that the question of whether two instruments core the same component is well posed. Above each component the two sides' construct profiles are printed one under the other. Ribbon color also marks that construct, gray any other, with related constructs sharing a hue at different lightness, as with loneliness and social isolation. Constructs are never grouped. An item that joins a component's core within the battery without itself being one of the items that keep that component is hatched.

battery is placed less reproducibly than a broad one.

A component enters the comparison when its pool-side core includes a documented construct HRS also measures and the component is kept for HRS, which is the condition under which the question of whether two instruments core the same component is well posed. The two sides agree on a component when some HRS item that leans on it has the same documented construct as the pool side, with each side labeled in its own documentation. Across the six encoders 54 components meet the condition and 44 of them agree (Table S10). The count of components varies with the encoder, from 7 to 11, and the constructs behind them do not: everyday discrimination, healthcare discrimination, loneliness and social isolation, the two neighborhood constructs, and perceived stress recur throughout Figs. S10 to S14. The instruments differ sharply in how narrowly they are placed. The discrimination items take 1.9 core components each on average over the six encoders, while the loneliness and perceived stress items take 3.8 and 3.5, so a third of the battery is spread thinly over the basis under every encoder.

Table S10: **The HRS battery placed on the fixed components, under each encoder.** A component is counted when its pool-side core includes a construct HRS also measures and the component is kept for HRS, that is when an HRS item cores it or stands out most on it. The two sides agree when an HRS item that leans on the component has the same documented construct as the pool items that define it.

| Encoder | Columns compared | Sides agree |
|---|---|---|
| gte-large-en-v1.5 (main text) | 7 | 7 |
| bge-m3 | 11 | 10 |
| bge-large-en-v1.5 | 9 | 8 |
| all-mpnet-base-v2 | 9 | 7 |
| e5-large-v2 | 8 | 5 |
| qwen3-embedding-0.6b | 10 | 7 |
| All six | 54 | 44 |

Read from the item rather than the component, by the construct-profile comparison of §S12, the 151 All of Us items reproduce as well as the pool's own items do, 0.38 against a chance level of 0.10, while the 35 HRS items reach 0.28 against 0.14 (Table S11). Both batteries are foreign to the basis in the same sense, neither having been fitted on, so the gap is a property of their breadth rather than of their provenance. The HRS chance level is the higher of the two because a 35-item battery covers few constructs, so even a permuted HRS item shares constructs with the item it is compared against. Correcting each figure for its own chance level leaves 0.31 for All of Us against 0.16 for HRS.

The agreement within HRS tracks how sharply an item is placed, falling from 0.69 for the one item with a single core component and 0.42 for the seven items with two, to 0.23 for the twenty-one items with three or four and 0.21 for the six with more. By construct, the two agree (Table S12). The discrimination items, the most concentrated of the battery, reproduce best at 0.47 for everyday discrimination, neighborhood disorder follows at 0.32, and loneliness, neighborhood social cohesion, and perceived stress trail at 0.25, 0.24, and 0.20. Perceived stress is last on both counts, spread over three and a half components on average and landing on different ones from encoder to encoder, which is the sense in which its items find no single home. The healthcare-discrimination figure rests on a single item and is reported for completeness only.

Table S11: **Cross-encoder agreement of item placement for three batteries on the same fixed basis.** Shown under the construct-profile definition used throughout and under a single-modal-construct alternative. The pool's own items are the control, All of Us and HRS are placed from their text without refitting. The two labelings give the same ordering, so the comparison does not depend on designating a single construct for a component.

| Battery | Items | Agreement | Chance | Chance-corrected |
|---|---|---|---|---|
| Pool (control) | 577 | 0.38 | 0.11 | 0.30 |
| All of Us | 151 | 0.38 | 0.10 | 0.31 |
| HRS | 35 | 0.28 | 0.14 | 0.16 |
| *single-modal-construct labeling, same items and same null* | | | | |
| Pool (control) | 577 | 0.41 | 0.04 | 0.38 |
| All of Us | 151 | 0.40 | 0.05 | 0.37 |
| HRS | 35 | 0.26 | 0.08 | 0.20 |

Table S12: **Cross-encoder agreement of the placement of each HRS item, by the item's documented construct.** Agreement is the mean pairwise Jaccard of the item's core-component construct profile over the fifteen encoder pairs, with the chance level for the battery as a whole at 0.14. HRS documents its eight neighborhood items under two headings, split here by the published item text (Methods §4.13).

| Documented construct | Items | Mean pairwise Jaccard |
|---|---|---|
| Healthcare discrimination | 1 | 0.51 |
| Everyday discrimination | 5 | 0.47 |
| Neighborhood disorder and safety | 4 | 0.32 |
| Loneliness | 11 | 0.25 |
| Neighborhood social cohesion | 4 | 0.24 |
| Perceived stress | 10 | 0.20 |
| All HRS items | 35 | 0.28 |
| Chance | 35 | 0.14 |

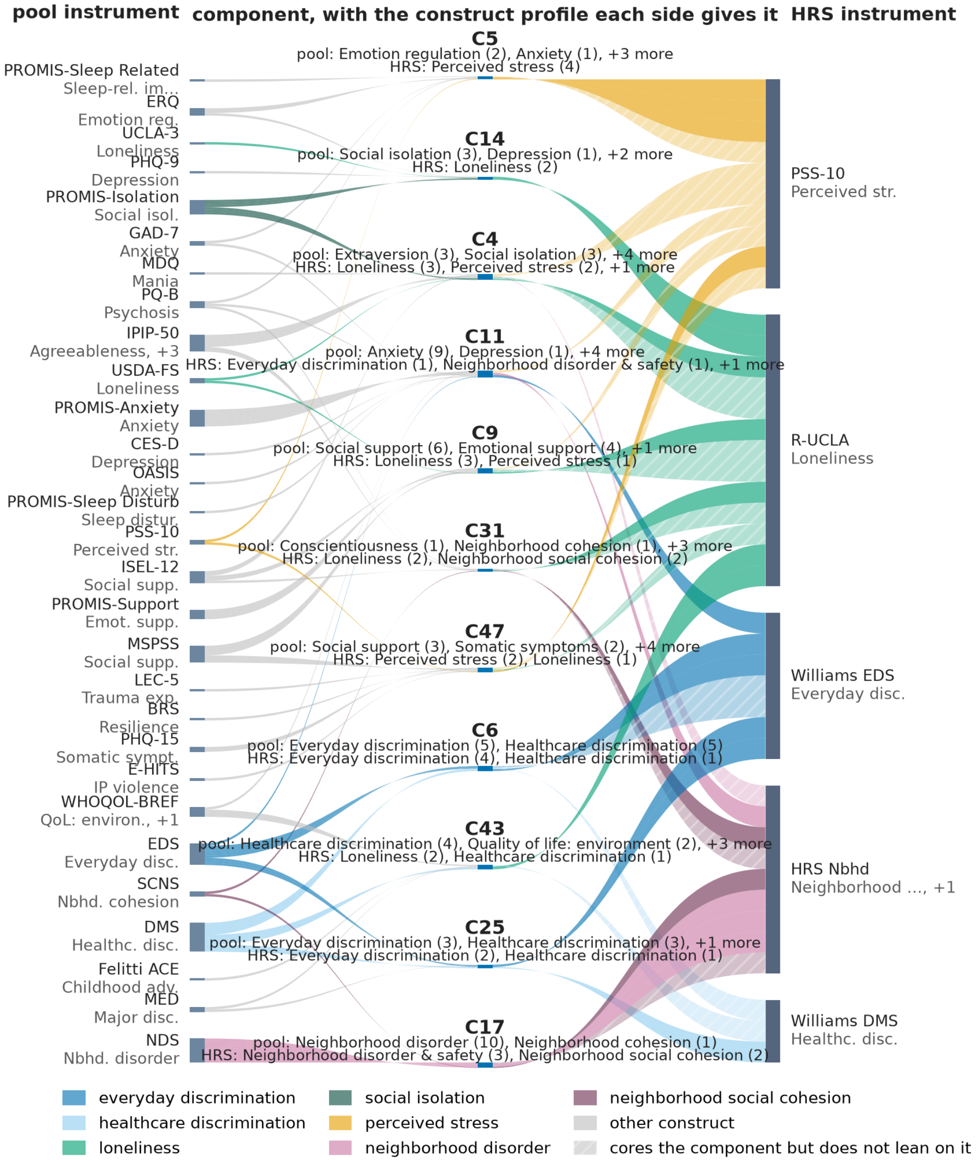


Figure S10: **HRS on the fixed components, bge-m3.** As Fig. S9, which gives the full legend, with the basis fitted on the pool under this encoder.

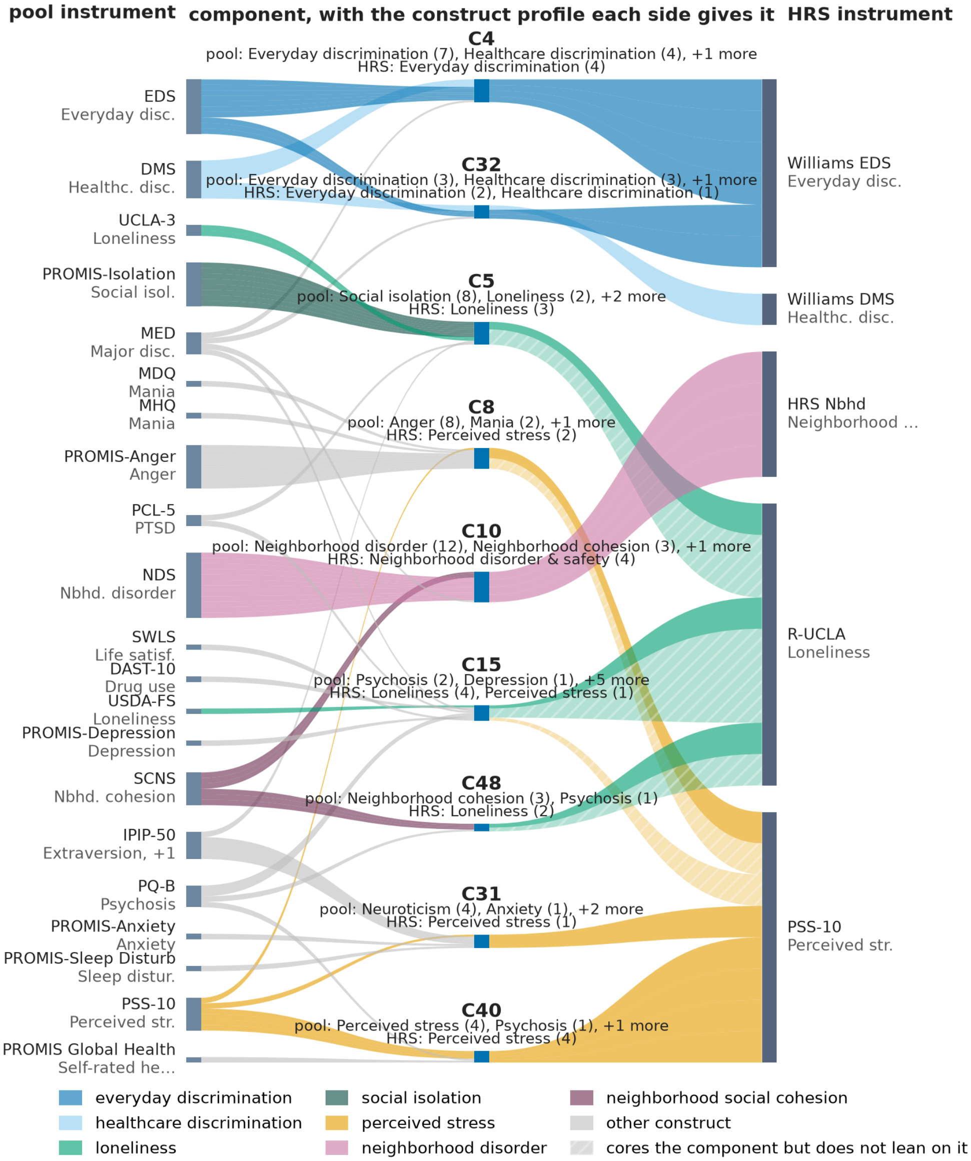


Figure S11: **HRS on the fixed components, bge-large-en-v1.5.** As Fig. S9, which gives the full legend, with the basis fitted on the pool under this encoder.

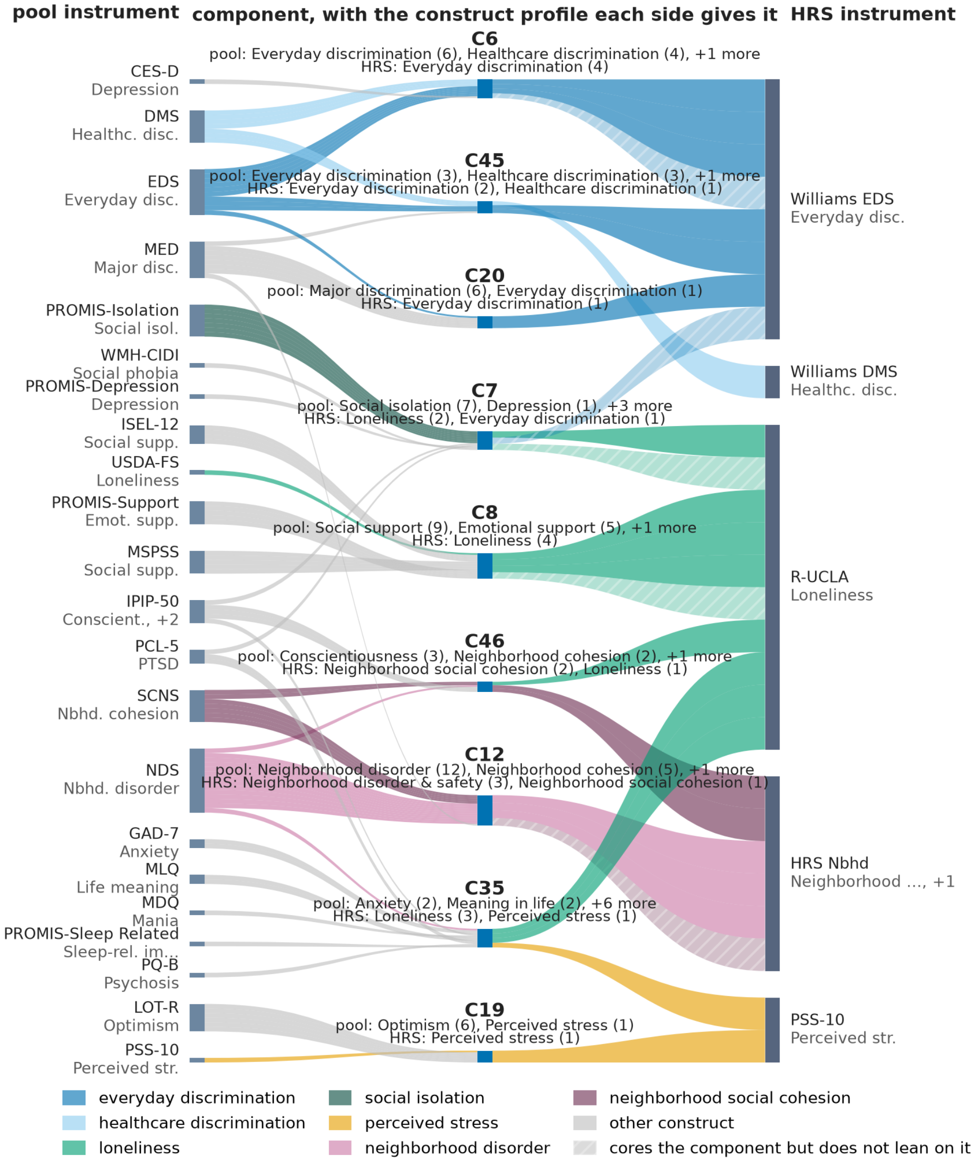


Figure S12: **HRS on the fixed components, all-mpnet-base-v2.** As Fig. S9, which gives the full legend, with the basis fitted on the pool under this encoder.

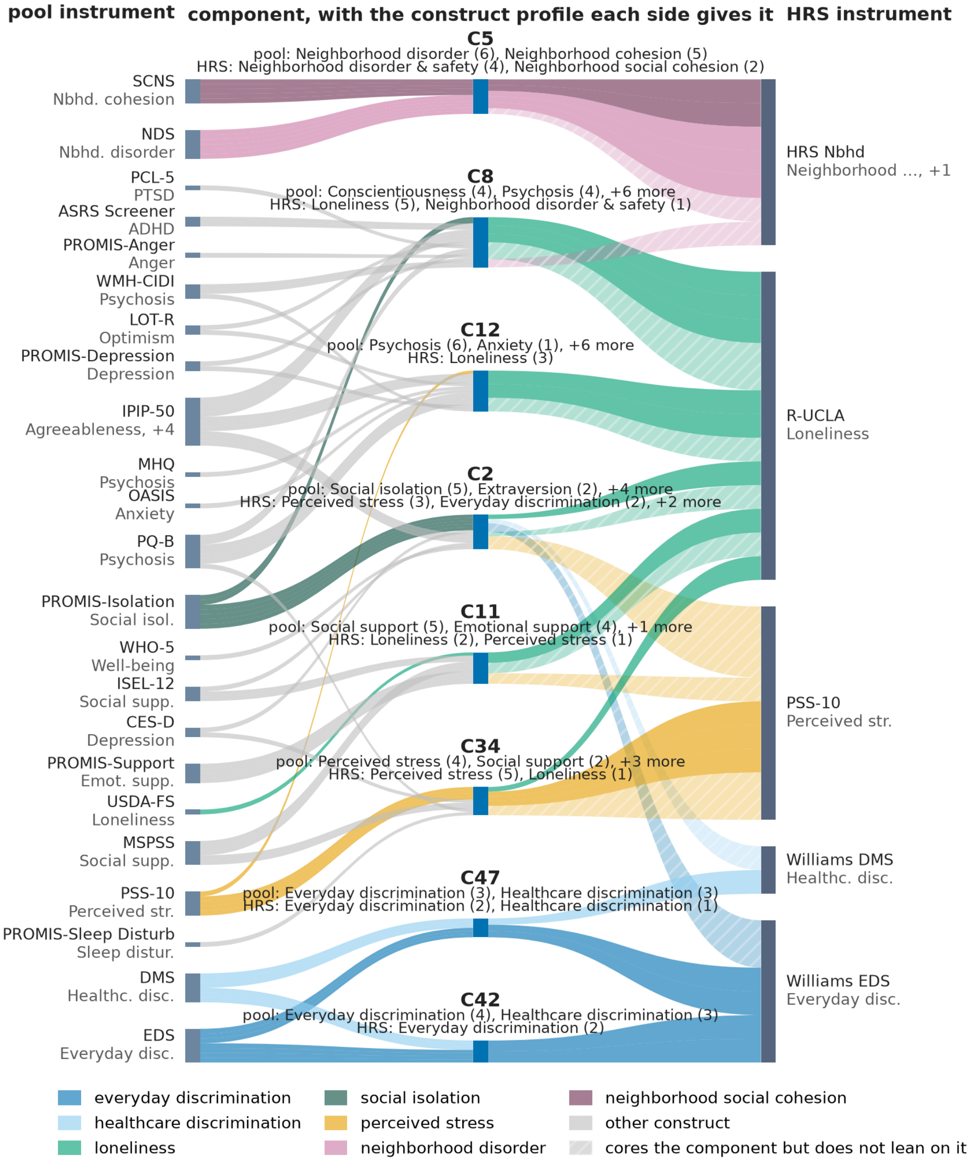


Figure S13: **HRS on the fixed components, e5-large-v2.** As Fig. S9, which gives the full legend, with the basis fitted on the pool under this encoder.

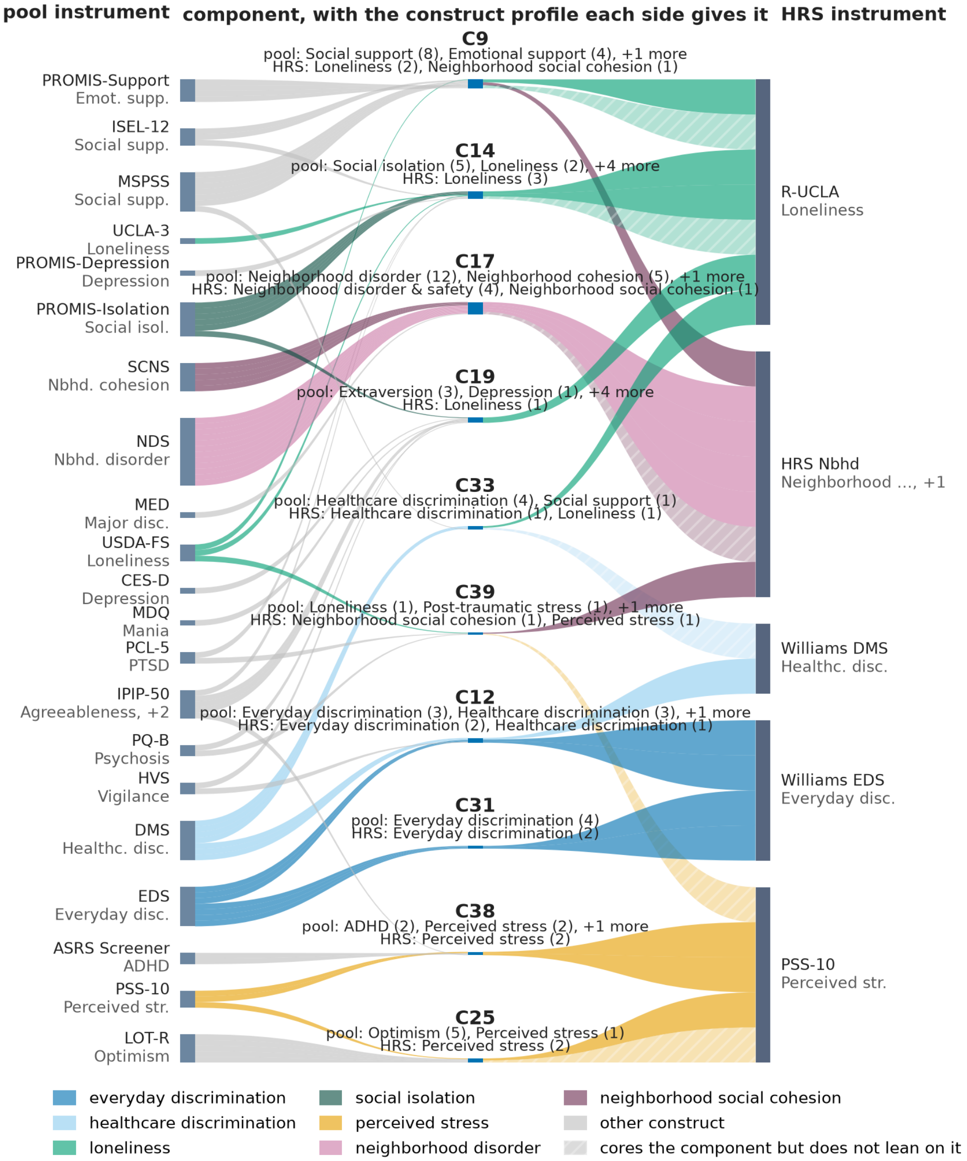


Figure S14: **HRS on the fixed components, qwen3-embedding-0.6b.** As Fig. S9, which gives the full legend, with the basis fitted on the pool under this encoder.

#### S15 Setting the sparsity weight

The sparsity weight $\lambda_{\text{score}}$ is the one free quantity in the basis, and Table S13 gives the sweep behind the value used throughout (Methods §4.2). Each row is an independent refit from the principal components of the same embeddings, so the rows are comparable to one another and the weight is the only thing that varies. At $\lambda_{\text{score}} = 5$ every diagnostic turns at once, the columns of $W$ losing orthonormality, the retained variance falling by more than a third, and the median core growing rather than shrinking, the signature of a collapse rather than of a sparser solution. Below that the median core energy rises with the weight and the retained variance falls slowly, and the two meet at $\lambda_{\text{score}} = 1$.

The faithfulness columns are reported for completeness and took no part in the choice. Their margin over chance is flat from $\lambda_{\text{score}} = 0.05$ to 1 in every encoder, between 0.39 and 0.48, so the documented labels do not distinguish weights across the range where the basis is sound, and the weight is set on the label-free diagnostics alone.

Table S13: **The sparsity-weight sweep.** For each encoder the basis is refit independently from the principal components at each $\lambda_{\text{score}}$, with $\lambda_{\text{ortho}} = 0.1$ and $k$ from parallel analysis. *ortho* is the largest off-diagonal cosine between the columns of $W$, *core* the median core size in items, *energy* the median core energy, and *VE/PCA* the variance the $k$ components retain relative to principal component analysis at the same $k$. These four use the embeddings alone and are what the weight was chosen on. The faithfulness columns, the rate at which an item's documented construct appears among the constructs of its core components and the chance rate from a label shuffle, are reported as a consequence of the weight and took no part in choosing it.

| Encoder | $\lambda_{\text{score}}$ | ortho | core | energy | VE/PCA | faithful | chance |
|---|---|---|---|---|---|---|---|
| mpnet ($k = 49$) | 0.001 | 0.0008 | 10.0 | 0.170 | 1.0000 | 0.842 | 0.614 |
| | 0.005 | 0.0006 | 9.0 | 0.203 | 0.9999 | 0.834 | 0.499 |
| | 0.01 | 0.0004 | 9.0 | 0.221 | 0.9997 | 0.841 | 0.473 |
| | 0.05 | 0.0006 | 8.0 | 0.253 | 0.9983 | 0.832 | 0.436 |
| | 0.1 | 0.0007 | 9.0 | 0.272 | 0.9967 | 0.827 | 0.437 |
| | 0.5 | 0.0046 | 8.0 | 0.283 | 0.9833 | 0.839 | 0.419 |
| | **1** | **0.0098** | **7.0** | **0.284** | **0.9656** | **0.820** | **0.396** |
| | 5 | 0.2232 | 14.0 | 0.202 | 0.5637 | 0.868 | 0.616 |
| bge-large ($k = 54$) | 0.001 | 0.0006 | 11.0 | 0.174 | 1.0001 | 0.856 | 0.629 |
| | 0.005 | 0.0006 | 9.0 | 0.214 | 0.9999 | 0.839 | 0.517 |
| | 0.01 | 0.0003 | 9.0 | 0.240 | 0.9998 | 0.861 | 0.479 |
| | 0.05 | 0.0010 | 8.5 | 0.276 | 0.9986 | 0.873 | 0.435 |
| | 0.1 | 0.0012 | 8.0 | 0.289 | 0.9971 | 0.861 | 0.434 |
| | 0.5 | 0.0055 | 8.0 | 0.293 | 0.9855 | 0.884 | 0.428 |
| | **1** | **0.0066** | **8.0** | **0.298** | **0.9709** | **0.861** | **0.414** |
| | 5 | 0.3487 | 13.0 | 0.228 | 0.6029 | 0.873 | 0.590 |
| bge-m3 ($k = 50$) | 0.001 | 0.0007 | 11.0 | 0.182 | 1.0000 | 0.827 | 0.651 |
| | 0.005 | 0.0008 | 8.0 | 0.215 | 0.9998 | 0.815 | 0.515 |
| | 0.01 | 0.0004 | 7.5 | 0.240 | 0.9996 | 0.815 | 0.471 |
| | 0.05 | 0.0010 | 8.0 | 0.271 | 0.9985 | 0.847 | 0.445 |
| | 0.1 | 0.0009 | 8.0 | 0.271 | 0.9969 | 0.849 | 0.451 |
| | 0.5 | 0.0072 | 7.0 | 0.294 | 0.9839 | 0.854 | 0.440 |
| | **1** | **0.0124** | **7.0** | **0.301** | **0.9678** | **0.820** | **0.424** |
| | 5 | 0.4833 | 16.0 | 0.204 | 0.5788 | 0.893 | 0.638 |
| e5-large ($k = 55$) | 0.001 | 0.0007 | 10.0 | 0.191 | 1.0004 | 0.846 | 0.608 |
| | 0.005 | 0.0009 | 8.0 | 0.227 | 1.0001 | 0.849 | 0.465 |
| | 0.01 | 0.0010 | 8.0 | 0.233 | 1.0000 | 0.844 | 0.432 |
| | 0.05 | 0.0008 | 7.0 | 0.265 | 0.9980 | 0.870 | 0.403 |
| | 0.1 | 0.0010 | 7.0 | 0.260 | 0.9956 | 0.847 | 0.392 |
| | 0.5 | 0.0077 | 7.0 | 0.294 | 0.9779 | 0.821 | 0.369 |
| | **1** | **0.0130** | **7.0** | **0.291** | **0.9560** | **0.839** | **0.376** |
| | 5 | 0.7604 | 17.0 | 0.201 | 0.4031 | 0.872 | 0.695 |
| gte-large ($k = 55$) | 0.001 | 0.0012 | 10.0 | 0.173 | 1.0000 | 0.839 | 0.659 |

*Table S13 (continued)*

| Encoder | $\lambda_{\text{score}}$ | ortho | core | energy | VE/PCA | faithful | chance |
|---|---|---|---|---|---|---|---|
| | 0.005 | 0.0007 | 9.0 | 0.212 | 0.9998 | 0.853 | 0.545 |
| | 0.01 | 0.0005 | 8.0 | 0.232 | 0.9997 | 0.856 | 0.486 |
| | 0.05 | 0.0007 | 8.0 | 0.281 | 0.9986 | 0.877 | 0.406 |
| | 0.1 | 0.0008 | 7.0 | 0.296 | 0.9973 | 0.868 | 0.390 |
| | 0.5 | 0.0044 | 8.0 | 0.310 | 0.9861 | 0.861 | 0.388 |
| | **1** | **0.0088** | **8.0** | **0.316** | **0.9728** | **0.870** | **0.398** |
| | 5 | 0.6396 | 14.0 | 0.197 | 0.5954 | 0.922 | 0.605 |
| qwen3 ($k = 49$) | 0.001 | 0.0007 | 10.0 | 0.179 | 1.0000 | 0.839 | 0.619 |
| | 0.005 | 0.0005 | 9.0 | 0.218 | 0.9999 | 0.854 | 0.519 |
| | 0.01 | 0.0006 | 7.0 | 0.239 | 0.9997 | 0.830 | 0.455 |
| | 0.05 | 0.0005 | 8.0 | 0.299 | 0.9985 | 0.846 | 0.395 |
| | 0.1 | 0.0008 | 7.0 | 0.289 | 0.9969 | 0.849 | 0.391 |
| | 0.5 | 0.0041 | 7.0 | 0.309 | 0.9851 | 0.835 | 0.385 |
| | **1** | **0.0070** | **7.0** | **0.314** | **0.9695** | **0.809** | **0.370** |
| | 5 | 0.4938 | 13.0 | 0.206 | 0.5758 | 0.873 | 0.597 |

#### S16 What a documented construct can and cannot resolve

An instrument documents one construct for all of its items, or one for each validated subscale, so the label does not distinguish the items within an instrument. Agreement with it tests the basis at that resolution, and disagreement shows where the basis separates items that share a construct but differ in semantic content. Both are measured pool-wide.

Items sharing a documented construct land in one component's core 18% of the time and items whose constructs differ under one percent, a ratio of 16 to 24 across the six encoders (Methods §4.8). Of the 27 constructs with at least three core items under all six encoders, the items of 8 fall in more than one component's core under every encoder and those of 5 under none. Where they split, the encoders largely agree on how, at an adjusted Rand index of 0.27 against 0.00 expected by chance (method at the end of this note). The agreement is strongest for depression and anxiety, which have the most items to differentiate and split into a median of four components each at 0.93 and 0.90, and weakest for somatic symptoms at 0.10. A basis that reproduced the codebook would show none of this splitting, and one that placed items arbitrarily would show none of the grouping.

**Method.** A documented construct labels every item of an instrument, or of a validated subscale, alike, so items sharing one label may still differ in content. For each construct with at least three items that core some component under all six encoders, we record how many distinct component cores its items occupy. That number exceeds one when the basis resolves finer than the label. Reproducibility decides whether such a split is signal or noise. For each pair of encoders that both split a construct we compare the two partitions its items induce by the adjusted Rand index, zero at chance agreement and one for identical partitions, and average over the fifteen pairs. The null shuffles items across that construct's components and recovers zero.

#### S17 The external reference pool

We built the reference basis from a pool of published questionnaire item text rather than from any cohort's answers (main-text §2.1, Methods §4.2). The pool holds 577 items from 50 published instruments spanning 54 documented constructs. It is an item bank and not a response dataset. It contains no respondent data of any kind, though it records response coding where the source instrument documents it.

We assembled the pool to span what published psychosocial instruments measure rather than to mirror one cohort's battery, and it is a selection of published instruments rather than a census. It deliberately includes several independently written instruments for the same construct, so a battery can be placed on the basis by content rather than by shared item text. Fourteen of the 50 instruments are *shared*, meaning All of Us administers that same instrument, and they contribute 101 of the 577 items. The remaining 36 are *independent*, contributing 476 items, and they measure the same content through different questions. `pool_instruments.csv` in **Supplementary Data 4** lists all 50 with their item counts, their role, the documented constructs of their items, and the citation each instrument was verified against.

We take every item's construct label from the parent instrument's own codebook or published manual: either the construct the instrument documents or, for an instrument with documented subscales, the subscale that item belongs to. 480 items are labeled at the instrument-construct level and 97 at a documented subscale. The label is never re-derived from the item text. This matters because the components are computed from that same text, so a text-derived label would test the method against its own input rather than against the instrument's documentation. Where an instrument's own manual and a cohort's coarser grouping of it disagree, we use the manual. This affects six All of Us items: three panic and phobia items that the WHO CIDI documents as its separate panic disorder, social phobia, and agoraphobia modules, and three life-threatening event items that the source manuals place under traumatic events. One item represents each of the three CIDI modules and no independent instrument in the pool covers them. We record that as it stands rather than resolve it by folding them into a broader label.

The item table records the provenance of each label. 556 items are labeled from the instrument's own published documentation, 19 from the printed manual where no machine-readable codebook exists, and 2 from the All of Us survey codebook for its DSM-5-aligned CIDI items. At the instrument level, we verified 27 of the 50 against the published source document, 7 are standard public-domain instruments, 10 inherit an earlier verified curation, and 6 come from the All of Us item table. Every instrument row gives the citation the verification used.

We ship two versions of each item's text. `prompt_raw` is the item verbatim. `prompt_clean` is the text we embed, and it differs only in that we remove a leading time-frame phrase, as with *In the past 7 days*, so that items are not grouped by a recall window they happen to share. 162 of the 577 items have such a phrase, and `stem_removed` records the removed text item by item. We keep framing that carries content, as with the childhood-adversity frame *while you were growing up* and the partner reference in the intimate-partner-violence items, because removing it would change what the item asks. All of Us and HRS items keep their own recall windows when they are placed on the basis, so the agreement reported in the main text holds across this difference in syntax rather than depending on matched preprocessing.

Table S14 summarizes the components the pool yields under each encoder. The count is stable across encoders at 49 to 55, cores hold a median of seven or eight items, and 62 to 71 percent of the pool's items are a core item of at least one component. The items that core nothing are still placed, by their core components, at close to the same construct-faithfulness (§S11).

Table S14: **The pool's components under each encoder.** The number of components, the size of their cores, the fraction of the 577 pool items that are a core item of at least one component, and how many distinct documented constructs a core spans. The per-component composition, with the full text of every core item, is Supplementary Data 4 (`cores/`).

| Encoder | Components | Core size median [range] | Coverage | Constructs per core | Single-construct cores |
|---|---|---|---|---|---|
| `gte-large-en-v1.5` | 55 | 8 [3–27] | 0.71 | 3 | 13 |
| `bge-large-en-v1.5` | 54 | 8 [3–35] | 0.70 | 3 | 11 |
| `bge-m3` | 50 | 7 [2–32] | 0.67 | 3 | 11 |
| `e5-large-v2` | 55 | 7 [3–28] | 0.69 | 3 | 18 |
| `all-mpnet-base-v2` | 49 | 7 [2–21] | 0.62 | 3 | 13 |
| `qwen3-embedding-0.6b` | 49 | 7 [3–20] | 0.64 | 2 | 15 |

**Supplementary Data 4** (`supplementary_data/global_pool/`) provides the pool in full. It contains `pool_items.csv`, one row per item with its identifier, parent instrument, role, documented construct, the exact codebook module the label came from, the provenance of that label, both text versions, the removed stem, the response coding, and the source citation. `pool_instruments.csv` gives one row per instrument. `pool_core_summary.csv` gives the per-encoder summary of Table S14. `cores/` holds one file per encoder with every component's core items and their full question text, which is the per-component composition the main text refers to. A `README.md` states the definitions the files use.

#### S18 Placement under a difference in syntax, across encoders

Both batteries ask questions the reference pool also asks, so the effect of a difference in syntax can be measured rather than assumed (Methods §4.14). The main text reports this on the exemplar encoder, and Table S15 gives it under all six. We split matched pairs by whether the two sources word a question identically or differ in syntax, and score each pair by the overlap of the core components the two versions reach on the same fixed basis.

Byte-identical text places identically under every encoder for both batteries, as the procedure requires. For the All of Us battery the questions whose syntax differs, which differ from the pool text by a stem the pool strips or by punctuation, reach a mean overlap of 0.70 to 0.81 across the six. All 35 of them share at least one core component with their pool version under every encoder, against a random-pool baseline of 0.025 to 0.032. For HRS, which administers bare fragments where the pool keeps the interrogative frame, the same quantity is lower and more variable, 0.24 to 0.55, with 12 to 14 of the 14 pairs sharing at least one core component. The wider spread for HRS is expected, given both what is varied and that it has 14 matched questions against 35. The ordering against the baseline holds under every encoder for both.

Table S15: **Placement under a difference in syntax, under each encoder.** For each battery, the questions the reference pool also asks are split by whether the two sources word them identically or differ in syntax, and each pair is scored by the overlap of the core components the two versions reach on the same fixed basis (1 is full overlap and 0 is none). *shares one* counts the differing pairs reaching at least one core component in common and *same set* those reaching exactly the same components. *random* pairs each battery item with a random pool item. Identical text must place identically, so that column is a check on the procedure rather than a result (Methods §4.14).

| Encoder | identical | differing | shares one | same set | random |
|---|---|---|---|---|---|
| *All of Us*, 57 identical and 35 differing in syntax | | | | | |
| gte-large | 1.00 | 0.73 | 35/35 | 15/35 | 0.025 |
| bge-m3 | 1.00 | 0.76 | 35/35 | 17/35 | 0.030 |
| bge-large | 1.00 | 0.81 | 35/35 | 17/35 | 0.027 |
| all-mpnet | 1.00 | 0.70 | 35/35 | 11/35 | 0.032 |
| e5-large | 1.00 | 0.71 | 35/35 | 13/35 | 0.026 |
| qwen3 | 1.00 | 0.73 | 35/35 | 14/35 | 0.028 |
| *HRS*, 4 identical and 14 differing in syntax | | | | | |
| gte-large | 1.00 | 0.49 | 13/14 | 3/14 | 0.026 |
| bge-m3 | 1.00 | 0.55 | 14/14 | 2/14 | 0.033 |
| bge-large | 1.00 | 0.44 | 13/14 | 1/14 | 0.028 |
| all-mpnet | 1.00 | 0.39 | 12/14 | 1/14 | 0.029 |
| e5-large | 1.00 | 0.24 | 13/14 | 0/14 | 0.029 |
| qwen3 | 1.00 | 0.29 | 12/14 | 1/14 | 0.028 |

#### S19 Interpretability requires the sparsity

A basis of a few dozen directions is not interpretable on its own. To separate the reduction in dimension from the requirement that each component rest on a few questions, we fit a dense basis of the same size on the same pool items under each encoder, gave it the same item budget, and scored both bases for faithfulness by the same rule (Methods §4.10). We compare each basis's margin over its own label-shuffle chance rate, never the raw rate, because a basis whose components span more constructs attains a higher raw rate by construction.

The sparse basis places items faithfully 39.5 to 47.2 points above its own chance level and the dense basis 13.2 to 20.4 points, with no overlap between the two across the six encoders (Table S16). The gap in naming is starker. Between 11 and 18 sparse components are named by a single documented construct, against zero or one dense component under every encoder, and a dense component names 5.6 to 6.7 constructs on average against 3.1 to 3.7 for a sparse one. Reducing the pool to a few dozen directions is therefore not what makes a component interpretable. The requirement that each rest on a handful of questions is.

Table S16: **The dense-basis control, under each encoder.** For each encoder, a dense basis of the same size is fit on the same pool items and given the same item budget, and both bases are scored for faithfulness by the same rule (Methods §4.10). *Margin* is the faithfulness rate above that basis's own label-shuffle chance rate, which is the comparable quantity because a basis whose components carry more constructs attains a higher raw rate by construction. *Single-construct* counts the components whose naming items all carry one documented construct, and *constructs per component* averages how many each names.

| | | Margin | | Single-construct | | Constructs/component |
|---|---|---|---|---|---|---|
| Encoder | $k$ | sparse | dense | sparse | dense | sparse vs dense |
| gte-large | 55 | 47.2% | 13.2% | 13 | 0 | 3.56 vs 6.53 |
| bge-m3 | 50 | 39.5% | 14.1% | 11 | 1 | 3.72 vs 6.60 |
| bge-large | 54 | 44.8% | 13.7% | 11 | 0 | 3.48 vs 6.65 |
| all-mpnet | 49 | 42.3% | 20.4% | 13 | 0 | 3.20 vs 5.76 |
| e5-large | 55 | 46.3% | 17.0% | 18 | 1 | 3.16 vs 5.76 |
| qwen3 | 49 | 43.9% | 16.0% | 15 | 0 | 3.12 vs 5.63 |

#### S20 The validated clinical criteria

Main-text §2.3 checks the screen against the eight criteria of Table S17. Neither the basis nor the screen used any of these rules.

Table S17: **The eight validated clinical criteria, and how often each flags the cohort versus the consensus outliers.** Of the selected instruments, eight have an established criterion, and we operationalize each rule against the codebook response coding (Supplementary Data 1, `items_full.csv`). "% cohort" and "% outliers" are the share of the full cohort ($N$ = 22,332) and of the 510 consensus outliers that meet the criterion. The remaining instruments have no established criterion.

| Instrument | Validated flag criterion | % cohort | % outliers |
|---|---|---|---|
| PHQ-9 [6] | sum $\geq$ 10, or any self-harm-item endorsement | 17.7 | 72.0 |
| GAD-7 [12] | sum $\geq$ 10 | 13.8 | 64.1 |
| ACE [4] | $\geq$ 4 exposure categories | 23.1 | 64.1 |
| ASRS [13] | $\geq$ 4 of 6 Part-A items at clinical frequency (sometimes+ for items 1–3, often+ for items 4–6) | 19.5 | 52.7 |
| WMH-CIDI [7] | any of the 3 psychotic-experience items endorsed | 7.4 | 51.2 |
| Suicide screen (standalone) | any of the 3 self-harm/suicide items endorsed | 37.2 | 73.3 |
| HVS [17] | either item "sometimes/often true" | 11.4 | 67.5 |
| PROMIS [16] | self-rated mental health fair or poor | 13.8 | 54.5 |
| **Any of the eight** | meets $\geq$ 1 criterion | **58.4** | **99.0** |

#### S21 How well a battery ports, item by item

Placing a battery on the fixed basis is not all or nothing. Two quantities describe what the placement does for a given item, and neither compares the battery's documentation with the pool's. The first is *coverage*, the share of the item's energy about the pool's center that lies in the span of the components the battery keeps, the same projection as Methods §4.3 computed per item rather than per battery. The second is where the retained part points, the constructs the pool documents for the core items of the item's core components, reported as a description rather than scored. Alongside them we report the *stability* of that description across the six encoders and how concentrated the placement is, in core components and in the item's largest standardized loading.

Coverage on its own does not establish that a placement is right. A placement is right when a battery's items land on components whose documented constructs are a justifiable interpretation of those items, and main-text Fig. 4 and Fig. S9 give that side by side for the two batteries. Coverage says only how much of an item is there to be interpreted at all, so it qualifies that judgment rather than standing in for it.

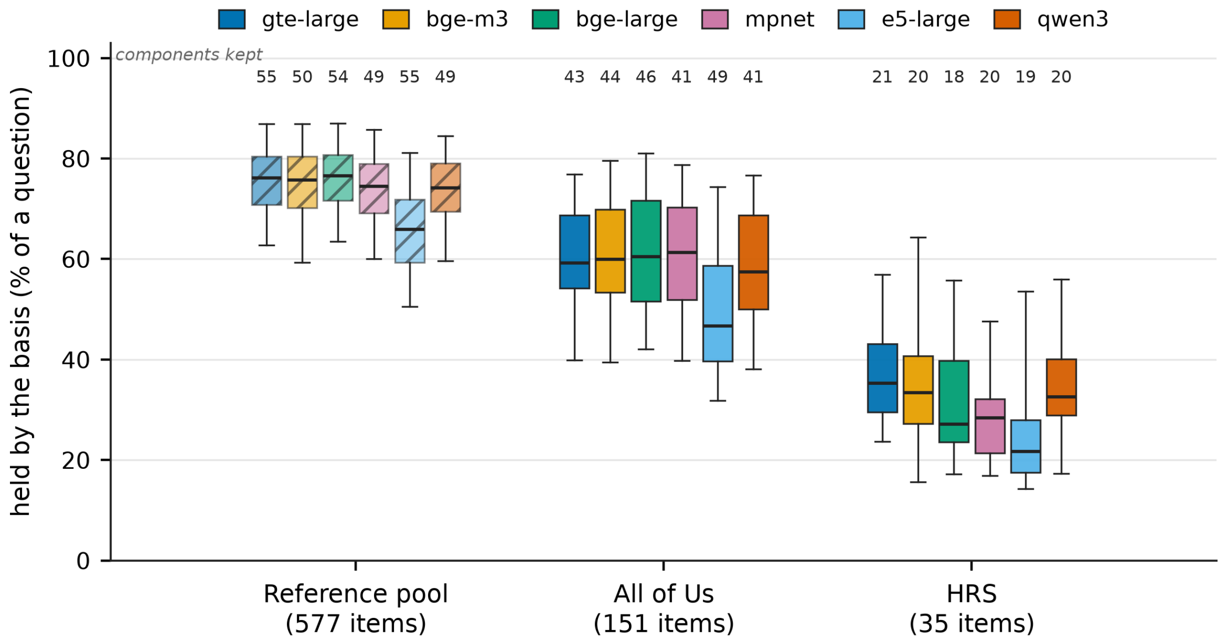


Figure S15: **How much of a question the fixed basis holds.** Per item, the percentage of the item that the components its battery keeps account for, under each of the six encoders. Boxes give the median and the interquartile range, whiskers the 5th and 95th percentiles. Above each box is the number of components that battery keeps under that encoder. The reference pool is the set of items the basis was built from and it keeps every component, so its counts are the size of the basis itself and it is the ceiling rather than a third ported battery, drawn hatched. The kept components differ between the two batteries, and the size of the basis differs between encoders, so the colors are not a ranking of the encoders. The six encoders appear in the same left-to-right order within each group, in the order of the key. The figure indicates how much of an item is placed, not whether the placement is justifiable, which is the two-side comparison of main-text Fig. 4 and Fig. S9.

Coverage predicts the rest in both batteries (Fig. S15, Table S18). The basis holds All of Us items at a median of 57.7% and HRS items at 28.7%, and the difference is that HRS administers bare fragments where the pool keeps a full question. Within each battery the best-covered third sits on half as many components as the worst-covered third, at a markedly higher peak loading, and the six encoders agree far more often on where those items land. A well-held item therefore concentrates on one or two components and every encoder puts it in the same place, while a poorly held one spreads thinly and the encoders disagree.

Coverage is a separate quantity from the number of components a battery reaches, and the two can move in opposite directions. Under e5-large the All of Us battery reaches the most components of any of the six encoders, 49 of that encoder's 55, and holds the least of each item, a median of 46.7% against 57 to 61% under the other five (Fig. S15). That encoder is also the lowest of the six on the pool's own items, so the comparison shows a property of its geometry rather than of porting.

Tables S19 and S20 describe every component each battery keeps, twice over. The pool's description comes from the constructs of the items that core the component within the pool, the battery's from the constructs of its own items that core it there, and the two are printed side by side without either being scored against the other, because the two documentations name the same content differently. All of Us spreads its energy evenly, its largest component carrying 5.4%, while HRS concentrates, its discrimination component alone carrying 13.2% of the battery's ported energy.

Content the pool does not hold behaves as the extreme case (Table S21). The five All of Us tobacco singles are the worst-covered items of the 151, spread over as many as five and a half core components at low peak loadings, and the encoders barely agree on where they land. The alcohol single administered in the same survey is covered nearly twice as well, takes exactly one core component under every encoder at $|z| = 5.6$, and lands on the pool's alcohol-use component every time. The pool fields alcohol through the Alcohol Use Disorders Identification Test (AUDIT) and drug use through the Drug Abuse Screening Test (DAST-10), and contains no item mentioning smoking, tobacco, cigarettes, vaping or nicotine, so the contrast is between content the pool holds and content it does not.

Table S18: **How much of each battery the basis holds, and how sharply it places it.** *Coverage* is the share of an item's energy about the pool's centre that lies in the span of the kept components, averaged over the six encoders. *Stability* is the mean pairwise overlap, over the 15 encoder pairs, of the pool constructs the item's core components carry. *Core components* and $|z|_{\max}$ describe how concentrated the placement is. Items are split into tertiles by coverage.

| Battery | items | $n$ | coverage | range | stability | core comps | $\|z\|_{\max}$ |
|---|---|---|---|---|---|---|---|
| All of Us | all | 151 | 57.7% | 23.1–82.6% | 0.38 | 2.6 | 3.9 |
| | low | 51 | 46.0% | — | 0.30 | 3.4 | 3.1 |
| | mid | 50 | 57.9% | — | 0.33 | 2.8 | 3.7 |
| | high | 50 | 71.0% | — | 0.51 | 1.7 | 5.0 |
| HRS | all | 35 | 28.7% | 21.1–70.1% | 0.28 | 3.3 | 3.2 |
| | low | 12 | 24.1% | — | 0.20 | 3.6 | 2.8 |
| | mid | 11 | 29.4% | — | 0.25 | 3.6 | 2.9 |
| | high | 12 | 43.8% | — | 0.39 | 2.7 | 3.8 |

Table S19: **The components All of Us keeps, described in both documentations.** *Energy* is the share of the battery's ported energy the component carries and $n$ the number of battery items that core it there. The last two columns are the leading constructs of the component's core items in the pool and in the battery, each taken from its own documentation and neither scored against the other (gte-large exemplar).

| Component | energy | $n$ | the pool documents it as | All of Us documents it as |
|---:|---:|---:|---|---|
| C10 | 5.4% | 16 | Neighborhood disorder, Neighborhood cohesion, Major discrimination | Neighborhood disorder & safety, Neighborhood social cohesion |
| C34 | 4.8% | 6 | Spirituality | Daily spiritual experience |
| C12 | 4.6% | 7 | Everyday discrimination, Healthcare discrimination, Heightened vigilance | Everyday discrimination, Healthcare discrimination |
| C43 | 3.8% | 5 | Adverse childhood experiences | Childhood adversity |
| C49 | 3.8% | 5 | Healthcare discrimination, Quality of life: physical, Major discrimination | Healthcare discrimination, Perceived social support |
| C54 | 3.4% | 5 | Anxiety, ADHD, Trauma exposure | Adult ADHD, Generalized anxiety, Life-threatening events |
| C45 | 3.2% | 3 | Suicidality, Adverse childhood experiences | Suicidality & self-harm |
| C39 | 3.0% | 5 | Trauma exposure, Interpersonal violence, Mania | Childhood adversity, Interpersonal & sexual violence |
| C41 | 2.8% | 8 | Depression, Conscientiousness, Self-esteem | Adult ADHD, Healthcare discrimination, Religious participation |
| C22 | 2.7% | 2 | Body-focused repetitive behavior, Quality of life: psychological | Body-focused repetitive behavior |
| C1 | 2.6% | 13 | Openness, Post-traumatic stress, Meaning in life | Perceived stress, Agreeableness, Panic & phobia |
| C14 | 2.6% | 4 | Psychosis | Psychosis |
| C31 | 2.6% | 3 | Interpersonal & sexual violence, Alcohol use, Trauma exposure | Interpersonal & sexual violence |
| C33 | 2.5% | 4 | Depression, ADHD, Quality of life: psychological | Adult ADHD, Depression |

| Component | energy | $n$ | the pool documents it as | All of Us documents it as |
|---|---|---|---|---|
| C44 | 2.4% | 4 | Social support, Quality of life: psychological, Subjective wellbeing | Perceived social support, Subjective wellbeing |
| C25 | 2.4% | 4 | Anxiety, Neuroticism, Psychosis | Generalized anxiety, Negative emotionality, Psychosis |
| C52 | 2.3% | 5 | Perceived stress, Anxiety, Trauma exposure | Perceived stress, Perceived social support, Adult ADHD |
| C40 | 2.3% | 5 | Post-traumatic stress | Depression, Neighborhood disorder & safety |
| C35 | 2.2% | 3 | Anxiety, Social support, Social phobia | Conscientiousness, Panic & phobia, Generalized anxiety |
| C36 | 2.1% | 3 | Drug use, Neighborhood disorder | Substance use, Neighborhood disorder & safety, Childhood adversity |
| C37 | 2.1% | 3 | Interpersonal violence, Interpersonal & sexual violence | Interpersonal & sexual violence |
| C46 | 2.1% | 2 | Quality of life: physical, Somatic symptoms, Depression | Generalized anxiety, Depression |
| C18 | 2.0% | 6 | Somatic symptoms, Anxiety, Sleep disturbance | Adult ADHD, Generalized anxiety, Perceived social support |
| C23 | 1.9% | 2 | Food insecurity | Food insecurity |
| C21 | 1.9% | 3 | Optimism, Perceived stress | Perceived stress, Conscientiousness |
| C53 | 1.9% | 3 | Anxiety | Self-rated health, Perceived stress, Generalized anxiety |
| C15 | 1.9% | 3 | Anger, Mania, Post-traumatic stress | Perceived stress, Mania, Generalized anxiety |
| C8 | 1.9% | 4 | Social support, Emotional support, Loneliness | Perceived social support, Loneliness |
| C16 | 1.8% | 4 | Quality of life: physical, Quality of life: environment, Quality of life: social | Self-rated health, Subjective wellbeing |
| C48 | 1.8% | 6 | Anger, Depression, Neuroticism | Self-rated health, Perceived stress, Adult ADHD |
| C26 | 1.8% | 3 | Alcohol use | Substance use, Neighborhood disorder & safety, Childhood adversity |
| C7 | 1.7% | 6 | Anxiety, Depression | Depression, Perceived stress, Generalized anxiety |
| C51 | 1.7% | 6 | Openness, Psychosis, Sleep disturbance | Open-mindedness, Self-rated health, Everyday discrimination |

| Component | energy | $n$ | the pool documents it as | All of Us documents it as |
|---|---|---|---|---|
| C38 | 1.6% | 3 | Mania | Perceived stress, Loneliness, Mania |
| C3 | 1.6% | 6 | Somatic symptoms, Depression, Trauma exposure | Depression, Generalized anxiety |
| C6 | 1.6% | 6 | Emotion regulation, Post-traumatic stress, Quality of life: psychological | Self-rated health, Loneliness, Negative emotionality |
| C9 | 1.6% | 4 | Social isolation, Loneliness, Post-traumatic stress | Loneliness |
| C5 | 1.5% | 8 | Life satisfaction, Well-being, Conscientiousness | Perceived social support, Agreeableness, Housing instability |
| C13 | 1.3% | 3 | Emotion regulation, Extraversion, Depression | Loneliness, Extraversion, Depression |
| C32 | 1.2% | 2 | Somatic symptoms, Quality of life: physical | Adult ADHD, Generalized anxiety |
| C28 | 1.2% | 2 | Meaning in life, Quality of life: psychological, Subjective wellbeing | Subjective wellbeing, Suicidality & self-harm |
| C20 | 1.1% | 6 | Depression, Post-traumatic stress, Alcohol use | Agreeableness, Loneliness, Adult ADHD |
| C27 | 1.1% | 1 | Agreeableness, Depression, Openness | Open-mindedness |

Table S20: **The components HRS keeps, described in both documentations.** *Energy* is the share of the battery's ported energy the component carries and $n$ the number of battery items that core it there. The last two columns are the leading constructs of the component's core items in the pool and in the battery, each taken from its own documentation and neither scored against the other (gte-large exemplar).

| Component | energy | $n$ | the pool documents it as | HRS documents it as |
|---|---|---|---|---|
| C12 | 13.2% | 4 | Everyday discrimination, Healthcare discrimination, Heightened vigilance | Everyday discrimination, Healthcare discrimination |
| C52 | 7.5% | 2 | Perceived stress, Anxiety, Trauma exposure | Perceived stress |
| C9 | 7.3% | 3 | Social isolation, Loneliness, Post-traumatic stress | Loneliness |
| C44 | 5.6% | 4 | Social support, Quality of life: psychological, Subjective wellbeing | Loneliness, Neighborhood |

| Component | energy | $n$ | the pool documents it as | HRS documents it as |
|---|---|---|---|---|
| C10 | 5.5% | 2 | Neighborhood disorder, Neighborhood cohesion, Major discrimination | Neighborhood |
| C8 | 5.5% | 3 | Social support, Emotional support, Loneliness | Loneliness |
| C49 | 4.4% | 1 | Healthcare discrimination, Quality of life: physical, Major discrimination | Healthcare discrimination |
| C38 | 4.4% | 2 | Mania | Perceived stress |
| C30 | 4.3% | 3 | Psychosis, Mania, Heightened vigilance | Loneliness |
| C45 | 4.2% | 1 | Suicidality, Adverse childhood experiences | Perceived stress |
| C22 | 4.1% | 2 | Body-focused repetitive behavior, Quality of life: psychological | Everyday discrimination, Perceived stress |
| C15 | 4.0% | 3 | Anger, Mania, Post-traumatic stress | Perceived stress, Neighborhood |
| C48 | 4.0% | 7 | Anger, Depression, Neuroticism | Loneliness, Perceived stress, Neighborhood |
| C7 | 3.9% | 2 | Anxiety, Depression | Perceived stress, Neighborhood |
| C54 | 3.8% | 2 | Anxiety, ADHD, Trauma exposure | Perceived stress |
| C13 | 3.6% | 4 | Emotion regulation, Extraversion, Depression | Perceived stress, Loneliness |
| C42 | 3.4% | 3 | Social support | Loneliness, Everyday discrimination |
| C19 | 3.0% | 3 | Self-esteem, Depression, Post-traumatic stress | Perceived stress, Everyday discrimination |
| C18 | 2.9% | 4 | Somatic symptoms, Anxiety, Sleep disturbance | Perceived stress, Neighborhood |
| C1 | 2.7% | 4 | Openness, Post-traumatic stress, Meaning in life | Perceived stress |
| C6 | 2.7% | 4 | Emotion regulation, Post-traumatic stress, Quality of life: psychological | Loneliness, Neighborhood |

Table S21: **Content the pool does not hold, against content it does.** The five All of Us tobacco singles and the alcohol single administered in the same survey. The pool fields alcohol through the AUDIT and drug use through the DAST-10 and contains no item mentioning smoking, tobacco, cigarettes, vaping or nicotine. Columns as in Table S18, with the landing taken under the gte-large exemplar.

| Item | coverage | stability | core comps | $\lvert z\rvert_{\max}$ | lands on |
|---|---|---|---|---|---|
| `electronicsmoking_electricsmokeparticipant` | 23.1% | 0.27 | 2.3 | 2.9 | Alcohol use, Drug use, Neighborhood disorder |
| `smokelesstobacco_smokelesstobaccoparticipant` | 24.5% | 0.17 | 3.7 | 2.6 | Alcohol use, Anxiety, Conscientiousness |
| `smoking_100cigslifetime` | 24.6% | 0.22 | 5.5 | 1.9 | ADHD, Adverse childhood experiences, Alcohol use |
| `cigarsmoking_cigarsmokeparticipant` | 26.1% | 0.21 | 4.2 | 2.1 | ADHD, Alcohol use, Anxiety |
| `hookahsmoking_hookahsmokeparticipant` | 28.3% | 0.25 | 5.0 | 2.2 | Adverse childhood experiences, Alcohol use, Drug use |
| `alcohol_alcoholparticipant` | 47.9% | 1.00 | 1.0 | 5.6 | Alcohol use |

### S22 The five under all six encoders

Main-text §2.4 accounts for the five clinically invisible outliers, the consensus outliers who meet none of the eight validated criteria, on the exemplar encoder. This section gives the same participants under all six.

Each encoder fits its own basis, so component indices have no meaning across encoders. The comparable object is the construct profile, the documented constructs of the All of Us items that core a component within the battery (§2.2), so a participant's account is compared as a set of constructs rather than as a set of components. Table S22 gives, for each participant and encoder, the component carrying the most of their distance, its share, how many components reach half the distance, and the constructs of the component with the largest share.

Under all six encoders a documented discrimination construct carries every one of the five. It is everyday and healthcare discrimination for participants 1 and 2, everyday discrimination for participant 3, and healthcare discrimination for participants 4 and 5. Three of the five also carry content of their own under at least four of the six, and that content distinguishes them from one another: interpersonal and sexual violence for participant 4, depression for participant 2, and substance use, childhood adversity, neighborhood disorder and safety, and perceived social support for participant 5.

Agreement is the mean overlap of those construct sets over the fifteen encoder pairs, 1 for the same constructs and 0 for none in common, which is the per-participant form of the item-side quantity in main-text Fig. 3b and needs no matching of components. It requires a floor, because an overlap has no meaning on its own and because a method that answered the same content for everybody would appear perfectly stable. Two floors are given. Against a different participant drawn from the 510 consensus outliers, whose content is varied, the component with the largest share agrees at 0.47 for the same participant against 0.08, and what carries them at 0.34 against 0.18. Against a different participant drawn from these five, the same quantities are 0.38 and 0.26. The second floor is high because the five were selected for meeting no clinical criterion, which is a criterion that makes them resemble one another, so they are strongly identifiable as a group and only weakly separable within it.

Participants differ in how well the account holds, so it is reported per participant rather than averaged. It runs from 0.73 for participant 1, whose largest component carries everyday and healthcare discrimination under five of the six, to 0.18 for participant 5, whose largest component is cored on childhood adversity, neighborhood disorder and safety and substance use under one encoder, on perceived social support under two, and on healthcare discrimination under two others. For that participant only the statement that healthcare discrimination carries them survives all six.

Table S22: **The five clinically invisible outliers, under each of the six encoders.** For each, the component carrying the most of that participant's distance, the share it carries, the number of components reaching half the distance, and the documented constructs of the All of Us items that core the component with the largest share within the battery. Component indices are local to an encoder and are given only for reference. Participants are numbered as they appear in main-text §2.4.

| Encoder | Component | Share | To half | Constructs of the largest component |
|---|---|---|---|---|
| **Participant 1**, agreement 0.73 | | | | |
| gte-large | C12 | 44.9% | 2 | Everyday and healthcare discrimination |
| bge-m3 | C6 | 57.4% | 1 | Everyday and healthcare discrimination |
| bge-large | C43 | 21.8% | 3 | Healthcare discrimination |
| all-mpnet | C6 | 40.8% | 2 | Everyday and healthcare discrimination |
| e5-large | C42 | 35.3% | 2 | Everyday and healthcare discrimination |
| qwen3 | C33 | 26.3% | 3 | Healthcare discrimination |
| **Participant 2**, agreement 0.37 | | | | |
| gte-large | C51 | 28.5% | 3 | Adult ADHD, everyday discrimination, open-mindedness, self-rated health |
| bge-m3 | C45 | 20.3% | 3 | Adult ADHD, conscientiousness |
| bge-large | C4 | 32.4% | 2 | Everyday and healthcare discrimination |
| all-mpnet | C6 | 38.8% | 2 | Everyday and healthcare discrimination |
| e5-large | C42 | 14.3% | 5 | Everyday and healthcare discrimination |
| qwen3 | C31 | 30.9% | 3 | Everyday discrimination |
| **Participant 3**, agreement 0.67 | | | | |
| gte-large | C12 | 26.0% | 3 | Everyday and healthcare discrimination |
| bge-m3 | C6 | 23.2% | 3 | Everyday and healthcare discrimination |
| bge-large | C32 | 23.5% | 3 | Everyday and healthcare discrimination |
| all-mpnet | C45 | 32.7% | 3 | Everyday and healthcare discrimination |
| e5-large | C47 | 65.2% | 1 | Everyday and healthcare discrimination |
| qwen3 | C15 | 26.7% | 3 | Subjective wellbeing |
| **Participant 4**, agreement 0.42 | | | | |
| gte-large | C49 | 21.5% | 3 | Healthcare discrimination, perceived social support |
| bge-m3 | C29 | 24.6% | 3 | Childhood adversity, interpersonal and sexual violence |
| bge-large | C43 | 24.2% | 3 | Healthcare discrimination |
| all-mpnet | C32 | 30.5% | 3 | Healthcare discrimination |
| e5-large | C14 | 12.7% | 5 | Everyday and healthcare discrimination |
| qwen3 | C33 | 34.1% | 3 | Healthcare discrimination |
| **Participant 5**, agreement 0.18 | | | | |
| gte-large | C26 | 19.1% | 4 | Childhood adversity, neighborhood disorder and safety, substance use |

| Encoder | Component | Share | To half | Constructs of the largest component |
|---|---|---|---|---|
| bge-m3 | C9 | 27.1% | 3 | Perceived social support |
| bge-large | C43 | 18.5% | 3 | Healthcare discrimination |
| all-mpnet | C8 | 30.5% | 2 | Perceived social support |
| e5-large | C9 | 19.5% | 4 | Everyday discrimination, generalized anxiety, healthcare discrimination |
| qwen3 | C33 | 29.1% | 3 | Healthcare discrimination |

#### S23 Negative contributions to a distance

The partition of $D^2$ over components is exact and signed, so a component can carry a negative contribution and move a participant closer to the cohort than their other components on their own would place them (Methods §4.7). This follows from the covariance rather than from any choice we make. A component whose score is close to what its correlated neighbors predict contributes little, and one closer than predicted contributes less than nothing.

Across the 510 outliers under the exemplar encoder, a participant has a median of 16 of the 43 kept components with negative contributions, between 6 and 23, and those sum to a median of −32% of that participant's distance and at most −80%. The positive contributions therefore sum to a median of 132%. Every share reported in the main text is a share of the exact total, negatives included, so the shares over all components sum to one.

Renormalizing over the positive contributions alone is possible and would rescale a participant's shares by a single positive factor, leaving every ordering within that participant unchanged. We do not do it, because shares from different participants would then no longer be shares of the same quantity, and because the signed shares sum to the distance being accounted for.

#### S24 How a participant's distance concentrates, and how far that governs reproducibility

Main-text §2.5 names the content that carries a participant and reports how far that naming reproduces across encoders. This section gives the layer beneath it. A participant's distance concentrates in a handful of components. Under the exemplar encoder the component with the largest share carries a median 23.2% of it, with quartiles of 18.6 and 29.6%, and a median of three components reach half of it, with quartiles of three and four. Every one of the 43 kept components is the largest for at least one participant.

The identity of that content matters at the top of a participant's ordering and not below it (Fig. S16). Among the six components that are the largest for at least twenty participants, the median share at rank 1 runs from 30.9% for the component cored on psychosis down to 20.8% for the one cored on healthcare discrimination and perceived social support. By rank 2 the same six run from 15.7 to 18.0% and by rank 3 from 12.5 to 14.2%, a spread of under two points, and the remaining 37 components pooled sit within the same range. Being the content that carries a participant is therefore not the same as carrying them far, and the difference lives entirely in the largest component.

Reproducibility follows that concentration. Comparing components across encoders by the documented constructs of the items that core them, since component indices are not comparable, the component with the largest share of a participant's distance agrees across the six encoders at 0.21 against 0.08 for a different participant of the 510. That agreement rises with how much of the distance the component carries (Spearman $\rho = 0.37$, 95% CI 0.29 to 0.44) and falls as more components are needed to reach half of it ($-0.28$, $-0.36$ to $-0.21$), with the most concentrated quarter of the group agreeing at 0.31 and the least at 0.13. The whole set of components that carry a participant, rather than its largest member, agrees at 0.26 against a floor of 0.18 and does not vary with concentration (0.07, $-0.02$ to 0.16). Naming the one component that carries most of a participant is therefore reproducible in proportion to how much of their distance it carries, while naming the whole set is not. That is why the main text makes the per-participant claim at the construct layer instead.

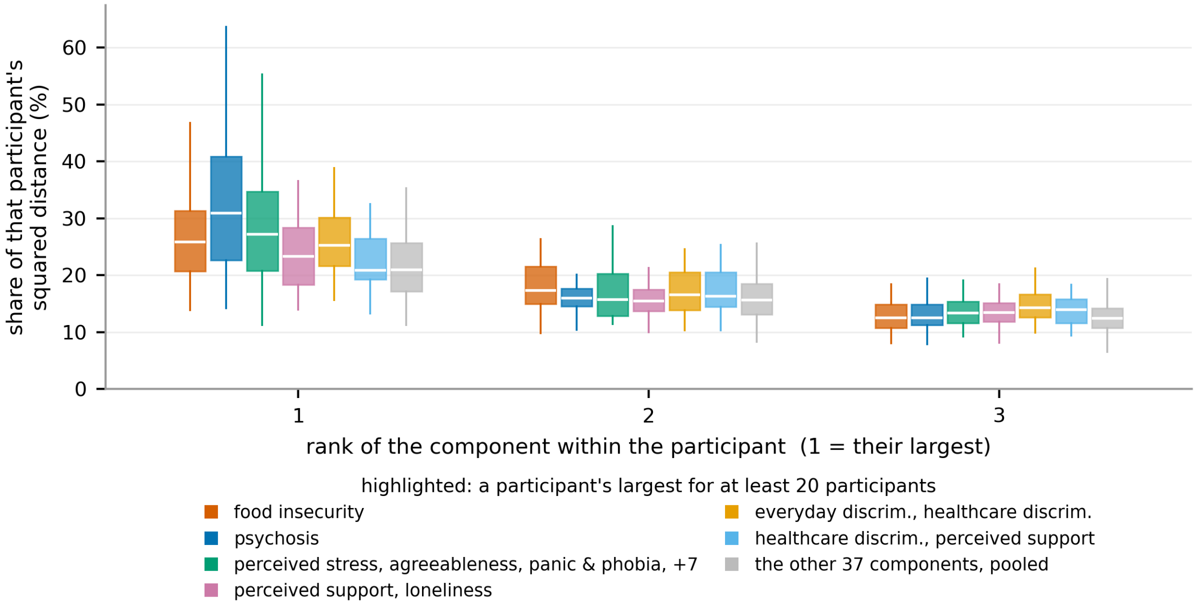


Figure S16: **How much of a participant's distance a component accounts for, by its rank.** Every participant's component shares are sorted largest first, so rank 1 is that participant's largest component whatever it is. Each box is one component at one rank, over the participants for whom it sits at that rank: median, quartiles, and whiskers to 1.5 times the interquartile range. The six components that are the largest for at least twenty participants are drawn separately and the remaining 37 are pooled into the gray box, so all 43 are accounted for. Points beyond the whiskers are not drawn and the vertical axis is scaled to the whiskers, since a few participants place almost all of their distance on one component. A component with fewer than ten participants at a rank has no box there.

#### S25 Is the care association just severity?

Main-text §2.6 reports that the outliers face more barriers to care and have care more concentrated in the emergency department. Almost any measure of how burdened a participant is would show that, so on its own the association says the group is real and says nothing about whether taking the instruments together contributes.

We build three measures from the same 151 responses (Methods §4.18). *Content extremity* is the mean absolute standardized response over the items (Methods §4.16), a severity score with no cross-instrument structure. The *diagonal distance* is the same component scores with every off-diagonal term of the covariance set to zero (§S9), which keeps the reference basis and drops the covariance between components. The *cross-instrument distance* is the screen itself. We take all three as the participant's percentile, averaged over the six encoders for the two computed per encoder, then rank-transform and standardize them so a coefficient is a change per standard deviation. Models are ordinary least squares on the ranks, matching Methods §4.17, adjusted for age, sex at birth, and for the emergency-department outcome the participant's total visit count.

The three overlap substantially, at Spearman 0.77 between the cross-instrument distance and content extremity and 0.81 between it and the diagonal distance, and their most extreme five percent coincide for only 618 participants. Taken one at a time each is associated with each outcome. Taken together (Table S23), the cross-instrument distance carries the emergency-department association, both other measures are indistinguishable from no association, and the same ordering holds for the two self-reported barriers, where content extremity also contributes.

Table S23: **The three measures compared on the same outcomes.** Standardized coefficients from the model containing all three, with 95% confidence intervals, and the $R^2$ each measure adds to the others. $N = 20{,}972$ for the emergency-department share and 20,310 for the survey outcomes.

| | standardized coefficient (95% CI) | | |
|---|---|---|---|
| Outcome | Content extremity | Diagonal distance | Cross-instrument |
| Emergency-department share | +0.009 (−0.011, +0.028) | +0.013 (−0.009, +0.035) | +0.061 (+0.040, +0.082) |
| Any access barrier | +0.038 (+0.028, +0.049) | −0.014 (−0.026, −0.002) | +0.119 (+0.107, +0.131) |
| Cost barrier | +0.064 (+0.054, +0.074) | −0.006 (−0.017, +0.005) | +0.088 (+0.078, +0.099) |
| $R^2$ added by the cross-instrument distance over the other two: +0.0011, +0.0165, +0.0110 | | | |

Three limits are worth stating. The increments are small in absolute terms, from $R^2 = 0.2965$ to 0.2976 for the emergency-department share, a model the visit count dominates, so the finding is that the association attaches to the cross-instrument distance rather than that it predicts emergency-department use well. The two survey outcomes are self-report from the same participants whose answers produced the predictor, so they share method with it and cannot corroborate the records independently. The diagonal distance takes a small negative coefficient for the survey outcomes. That is how a suppressor looks when two predictors correlate at 0.81, not an association in its own right, and we do not interpret it.

### S26 Content that leads many participants and content that leads a few

Main-text §2.5 reports that for 143 of the 510 the leading component is one that is largest for fewer than ten participants each. This section is the split behind that number, and asks what the validated criteria make of each side (`all_of_us_codes/heterogeneity.py`).

We split the group on whether the component carrying the largest share of a participant's distance is one that is largest for many people or for almost nobody, at a threshold of ten. The 143 carried by rare content meet a mean of 4.71 criteria against 5.11 for the 367 carried by common content, and 1.4% of them meet none against 0.8%. The direction is the one rare content would predict and the size is small. That is why the main text does not treat the clinically invisible outliers of §2.4 as the extreme of this tail. Four of those five are carried by components that are largest for more than twenty participants each.

[18] Jianlv Chen, Shitao Xiao, Peitian Zhang, Kun Luo, Defu Lian, and Zheng Liu. M3-Embedding: Multi-linguality, multi-functionality, multi-granularity text embeddings through self-knowledge distillation. *arXiv preprint arXiv:2402.03216*, 2024.

[19] Shitao Xiao, Zheng Liu, Peitian Zhang, Niklas Muennighoff, Defu Lian, and Jian-Yun Nie. C-Pack: Packed resources for general Chinese embeddings. In *Proceedings of the 47th International ACM SIGIR Conference on Research and Development in Information Retrieval*, 2024.

[20] Nils Reimers and Iryna Gurevych. Sentence-BERT: Sentence embeddings using Siamese BERT-networks. In *Proceedings of the 2019 Conference on Empirical Methods in Natural Language Processing (EMNLP-IJCNLP)*, pages 3982–3992, 2019. doi: 10.18653/v1/D19-1410.

[21] Kaitao Song, Xu Tan, Tao Qin, Jianfeng Lu, and Tie-Yan Liu. MPNet: Masked and permuted pre-training for language understanding. In *Advances in Neural Information Processing Systems (NeurIPS)*, volume 33, 2020.

[22] Liang Wang, Nan Yang, Xiaolong Huang, Binxing Jiao, Linjun Yang, Daxin Jiang, Rangan Majumder, and Furu Wei. Text embeddings by weakly-supervised contrastive pre-training. *arXiv preprint arXiv:2212.03533*, 2022.

[23] Zehan Li, Xin Zhang, Yanzhao Zhang, Dingkun Long, Pengjun Xie, and Meishan Zhang. Towards general text embeddings with multi-stage contrastive learning. *arXiv preprint arXiv:2308.03281*, 2023.

[24] Xin Zhang, Yanzhao Zhang, Dingkun Long, Wen Xie, Ziqi Dai, Jialong Tang, Huan Lin, Baosong Yang, Pengjun Xie, Fei Huang, Meishan Zhang, Wenjie Li, and Min Zhang. mGTE: Generalized long-context text representation and reranking models for multilingual text retrieval. *arXiv preprint arXiv:2407.19669*, 2024.

[25] Yanzhao Zhang, Mingxin Li, Dingkun Long, Xin Zhang, Huan Lin, An Yang, et al. Qwen3 embedding: Advancing text embedding and reranking through foundation models. *arXiv preprint arXiv:2506.05176*, 2025.

[26] Adam W. Meade and S. Bartholomew Craig. Identifying careless responses in survey data. *Psychological Methods*, 17(3):437–455, 2012. doi: 10.1037/a0028085.

[27] Yves Van Vaerenbergh and Troy D. Thomas. Response styles in survey research: A literature review of antecedents, consequences, and remedies. *International Journal of Public Opinion Research*, 25(2):195–217, 2013. doi: 10.1093/ijpor/eds021.

[28] Lewis R. Goldberg. A broad-bandwidth, public domain, personality inventory measuring the lower-level facets of several five-factor models. In Ivan Mervielde, Ian Deary, Filip De Fruyt, and Fritz Ostendorf, editors, *Personality Psychology in Europe*, volume 7, pages 7–28. Tilburg University Press, Tilburg, The Netherlands, 1999. 50-item IPIP Big-Five Factor Markers, `https://ipip.ori.org/New_IPIP-50-item-scale.htm`.
