## Supplementary material for "Scoring respondents on a semantic frame of reference identifies a group that is burdened across many questionnaires and reaches care less easily": README.md

# Supplementary Data — Global item pool

A cross-instrument pool of **577 validated psychosocial questionnaire items** drawn from
**50 instruments** and mapped to **54 documented constructs**, assembled from published item text
alone. It is the reference item bank for the portability / meaning-transfer analysis: a meaning
basis is learned from this pool and external instruments (and cohorts) are placed onto it from
wording alone.

**This is an item-text bank — it contains no respondent data.** Response *scales* are given where
documented (see `response_coding` below); the pool itself is not a response dataset.

## Definitions
- **Component** — one of the sparse, text-derived axes fitted to the pool embeddings (49–55 per
  encoder). Each item has a loading on each component.
- **Core** — the subset of a component's items that dominate its loadings (the fixed-point
  participation-ratio set defined in the main-text Methods). A component is summarized and named by
  its core.
- **Construct profile** — a component is read by the *set* of documented constructs of its core
  items, with the item count for each. A component whose core holds six depression items and two
  anxiety items has the profile `Depression (6); Anxiety (2)`. This set is the component's
  interpretation; no single construct is designated for it.
- **Single-construct component** — one whose core items all share one documented construct.
- **Coverage** — the fraction of the 577 pool items that appear in at least one component's core.

## Files

### `pool_items.csv` — one row per item (577)
| column | meaning |
|---|---|
| `item_id` | unique id within the pool |
| `source` | short provenance tag |
| `parent_instrument` | the instrument (scale) the item belongs to |
| `role` | `anchor` (an instrument All of Us also administered) or `independent` (a different instrument for the same construct — the transfer test) |
| `documented_construct` | the construct label used in analysis (**from the instrument's official codebook/manual**, harmonized across instruments measuring the same construct) |
| `documented_module` | the exact section/module heading in the parent instrument's codebook (audit trail; e.g. UK Biobank MHQ "Traumatic Events", WHO CIDI "Panic disorder") |
| `label_level` | `instrument construct` (single-construct scale) or `documented subscale` (a documented subscale of a multi-construct instrument) |
| `provenance` | `documented` / `manual-grounded` / `instrument-codebook (AoU CIDI-5)` |
| `prompt_raw` | verbatim item text |
| `prompt_clean` | item text with the leading time-frame stem removed (what is embedded); see stem note |
| `stem_removed` | the leading time phrase stripped from `prompt_raw`, if any |
| `response_coding` | documented response coding where available (All of Us–sourced items); blank otherwise — those instruments use their standard published response scales (see `source_citation`) |
| `source_citation` | citation / source for the instrument text |

### `pool_instruments.csv` — one row per instrument (50)
Scale-level provenance: role, item count, the documented constructs it contributes, the All of Us
instrument it matches (if any), how the wording was verified, and the citation.

### `pool_core_summary.csv` — one row per encoder (6)
Per-encoder structure of the fitted components: number of components `k`, `median_core` (and
`min_core`/`max_core`), `coverage` (fraction of the 577 items appearing in at least one core),
`median_constructs` (median number of distinct constructs in a core's profile), and
`n_single_construct` (components whose core holds one construct only).

### `cores/<encoder>_component_cores.csv` — every component's core, per encoder (6 files)
One row per (component, core item). Columns: `component`, `component_constructs` (its construct
profile, each construct with its item count), `n_constructs` (how many distinct constructs that
profile holds), `core_size`, `item_id`, `item_construct` (that item's documented construct), and
`item_text` (full prompt). This is the complete listing of which items define each component, for each of the six
encoders — the item-level backing for the component figures and for every co-location claim. The
*core* of a component is the subset of items that dominate its loadings (see the main-text Methods for
the exact fixed-point definition).

## Construct labelling rule (important)
Every construct label is the item's **parent-instrument official codebook/manual** construct or
documented subscale — **not** re-read from the item text (which would be circular with a
text-embedding), and **not** the administering study's re-grouping. For the six All of Us–administered
multi-construct instruments the instrument codebook is authoritative over All of Us's own grouping.

The 21 items from the two broad multi-construct instruments were verified against source documentation:
- **UK Biobank Mental Health Questionnaire** — sections confirmed against the official questionnaire
  (v1.3): Depression, Mania, Unusual and Psychotic Experiences, Intimate Partner Violence, Traumatic
  Events, Subjective Wellbeing. (Cite the 2020 reanalysis; the 2018 original was retracted.)
- **WHO World Mental Health CIDI** — panic / social phobia / agoraphobia are distinct documented
  modules (not one "panic & phobia"); the skin-picking / hair-pulling items are All of Us's
  DSM-5-aligned CIDI-5 (Behavioral Health survey) and correspond to DSM-5 excoriation /
  trichotillomania, labelled Body-focused repetitive behavior.

## Stem processing
`prompt_clean` removes only a leading time-frame / recall-window phrase (e.g. "In the past 7 days")
so items cluster by content rather than recall window; response wrappers ("have you ever …") are
kept so every cleaned item stays a whole sentence. `stem_removed` records exactly what came off
(reversible against `prompt_raw`). 162 of 577 items had a stem removed.

## Anchor vs independent
`anchor` = an instrument All of Us also administered (sanity check); `independent` = a *different*
instrument for the same construct (the genuine transfer test). Landing an independent instrument's
items on the construct-correct component proves the meaning basis generalizes beyond the fitting data.
