## Supplementary material for "Scoring respondents on a semantic frame of reference identifies a group that is burdened across many questionnaires and reaches care less easily": README.md

# Supplementary data

Six data items accompany the manuscript. Construct labels throughout are the documented constructs
of the parent instrument. There is no broader grouping of constructs in any file.

## Supplementary Data 1 — `items_full.csv`
The 151 selected All of Us survey items, one per row, sorted by construct.

| column | meaning |
|---|---|
| `aou_concept_id` | the All of Us concept id for the item |
| `item` | variable name |
| `instrument` | source instrument |
| `construct` | the construct assigned to the item |
| `coding` | response value range from the codebook |
| `prompt` | full item text |

## Supplementary Data 2 — `content_grammar_variants/`
The content-versus-syntax control experiment: 151 seeds × 2 variant types × 10 donors = 3,020 variants.

- `manifest.csv` — seed-to-donor design for every variant: `variant_id`, `cell` (A syntax / B content),
  `seed_item`, `donor_item`, `intended_item`, seed and donor frame and construct, and the seed and
  donor prompt texts.
- `generated_items_v2.csv` — the assembled rewrites: `variant_id`, `cell`, `seed_item`, `donor_item`,
  `intended_item`, `rewritten_prompt`.
- `batch_s*.jsonl` — the raw generations (claude-opus-4-8), one `{variant_id, rewritten}` per line,
  batched by seed range, 3,020 lines in total.

Cell A is a syntax variant, the seed's semantic content in a donor's neutral syntax. Cell B is a
content variant, the seed's syntax carrying a donor's content.

## Supplementary Data 3 — `core_items/`
The All of Us items that form each component of the battery **on the fixed pool basis**, one CSV per
sentence encoder, one row per core item: `component` (the pool component's index), `item`,
`instrument`, `construct`, `loading` (the item's standardized loading on that component, in pool
units), `prompt`. Ordered by component, then by descending |loading|.

A component appears here when it is kept for the battery — at least one battery item clears that
component's core-item band on the pool, or the component is some item's modal one — and its rows are
the fixed point of the participation-ratio pruning down that component's column, the same rule the
pool's own cores use. The counts match main-text §R2: 43 of the 55 components under the exemplar
encoder, and 41 to 49 across the six.

| file | encoder | components kept | core rows |
|---|---|---|---|
| `core_items_gte.csv` | gte-large-en-v1.5 (the exemplar read in the main text) | 43 | 202 |
| `core_items_bge-m3.csv` | bge-m3 | 44 | 209 |
| `core_items_bge-lg.csv` | bge-large-en-v1.5 | 46 | 198 |
| `core_items_qwen.csv` | qwen3-embedding-0.6b | 41 | 209 |
| `core_items_mpnet.csv` | all-mpnet-base-v2 | 41 | 213 |
| `core_items_e5.csv` | e5-large-v2 | 49 | 223 |

Written by `make_core_items_pool.py`, which travels with the submitted code under
`code/examples/allofus/`. It replaces an earlier generator that produced this file from a **retired
151-item basis fitted to the battery itself** (23 to 26 components) — an object the manuscript no
longer contains, and that generator is not part of the submitted code.

## Supplementary Data 4 — `global_pool/`
The external reference pool: 577 published items from 50 instruments carrying 54 documented
constructs, the per-encoder component summary, and every component's core items with their full
question text. See `global_pool/README.md` for the file-by-file description and the definitions the
files use, and Supplementary Note S17 for how the pool was assembled and how its construct labels
were grounded. `pool_instruments.csv` is the instrument-level listing Supplementary Note S17 cites
for all 50 instruments.

## Supplementary Data 5 — `component_reproduction.csv`
All 55 components of the gte-large reference basis, with how widely each reproduces across the six
encoders and what its core carries. Read by Supplementary Note S12 and by the main text's
cross-encoder result. Sorted by descending `reproduces_in_encoders`, then by component.

| column | meaning |
|---|---|
| `component` | component id on the gte-large basis |
| `core_items` | number of items in its participation-ratio fixed-point core |
| `reproduces_in_encoders` | in how many of the six encoders a matching core is found, 1 to 6 |
| `documented_constructs` | documented constructs of the core items, with the item count for each |
| `core_item_ids` | the core items themselves, space separated, keyed to `global_pool/pool_items.csv` |

Reproduction is the label-free core-item matching of Supplementary Note S12, so the constructs are
read only after the matching and play no part in it.

## Supplementary Data 6 — `faithfulness_disagreements.csv`
The 75 pool items whose documented construct is absent from the constructs of their core components
(gte-large exemplar), one per row, sorted by construct. Read by Supplementary Note S11.

| column | meaning |
|---|---|
| `item_id` | item identifier, keyed to `global_pool/pool_items.csv` |
| `item` | full item text |
| `instrument` | source instrument |
| `documented_construct` | the construct the parent instrument documents for the item |
| `anchors_a_core` | 1 if the item is itself a core item of some component, 0 if it cores nothing |
| `constructs_of_core_components` | the constructs its core components carry, most-loaded component first |

The item's own membership of a core is excluded when the constructs of its core components are
collected, so an item that anchors a core is not counted faithful merely for being there. The 26 rows
with `anchors_a_core = 0` are the ones Supplementary Note S11 reads one by one.
