## Supplemental code for "Scoring respondents on a semantic frame of reference identifies a group that is burdened across many questionnaires and reaches care less easily": attribution.py

```
#!/usr/bin/env python3
"""attribution.py -- what a flagged respondent's distance is made of.

STANDALONE (numpy, pandas, sklearn). Recomputes the screen exactly as screen_battery.py does, then
decomposes each flagged respondent's distance. Everything below is an EXACT partition of T^2, not a
ranking or a proxy.

THE DECOMPOSITION

  T^2 = y' Omega y      y = the respondent's residualized component scores, Omega the Ledoit-Wolf
                        precision of the cohort's score covariance

  COMPONENT SHARE   a_j = y_j (Omega y)_j        sums to T^2 exactly, order-free
                    share_j = a_j / T^2. A NEGATIVE share is a suppressor: that component pulls the
                    distance down. This is the contribution GIVEN the others, not a solo score.

  CHARACTER         u_j = y_j^2 / Sigma_jj       extreme on its own
                    c_j = (Omega y)_j^2 / Omega_jj   extreme given the rest
                    the component is JOINT when c_j > u_j. In the uncorrelated limit u = c = a, so
                    the reading self-adapts. "Integration" = the share of T^2 carried by joint ones.

  ITEM SHARE        Because the component scores are linear in the responses, T^2 is a quadratic
                    form in the 151 residualized responses themselves:
                        Xr = responses with the covariates projected out
                        Q  = P_S[:, kept] Omega P_S[:, kept]'          (151 x 151)
                        T^2 = xr' Q xr        and     a_i = xr_i (Q xr)_i    sums to T^2 exactly
                    This layer is the one to prefer for reporting. It is well posed -- 151 items in
                    151 dimensions, no rank deficit -- and every item carries exactly ONE documented
                    construct, so no profile splitting or modal labelling is needed.

  CONSTRUCT SHARE   item shares summed within each item's documented construct. Exact, and in the
                    battery's own vocabulary.

The PAIRWISE layer (Omega_jk y_j y_k) is deliberately NOT reported. On a battery whose kept
components approach its item count it is dominated by cancellation between components the battery
cannot separate. It is a diagnostic, not a result.

WHY PER-PERSON READINGS NEED CARE. Cross-encoder work found the per-person share vector only weakly
reproducible across encoders. So this script reports every subset BOTH ways: the group mean, and the
per-person top constructs under each of the six encoders, so agreement can be seen rather than
assumed. For a handful of people, read the per-person table and not the group mean.

Run:  python3 attribution.py            # all consensus outliers
      python3 attribution.py --invisible   # only those meeting none of the eight cutoffs
"""
import json
import os
import sys
from collections import Counter

import numpy as np
import pandas as pd
from sklearn.covariance import LedoitWolf

HERE = os.path.dirname(os.path.abspath(__file__))
BASIS_DIR     = os.path.join(HERE, "reference_basis")
EMB_DIR       = os.path.join(HERE, "item_embeddings")
EMB_PREFIX    = "core151_items"
RESPONSES_CSV = "/home/jupyter/responses_out/responses_k0.csv"
WEIGHTS_CSV   = "/home/jupyter/responses_out/weights_k0.csv"
SCALES_CSV    = os.path.join(HERE, "scales_prism.csv")
ITEMS_CSV     = os.path.join(HERE, "core_items_151.csv")
CONSTRUCTS_CSV = os.path.join(HERE, "construct_map_151.csv")   # item, construct
OUT_DIR       = os.path.join(HERE, "screen_out")
ID_COL        = "id"
COVARIATES    = ["age", "sex"]
PCTILE        = 95.0
TOP_N         = 12

ENCODERS = ["gte-large-en-v1.5", "bge-m3", "bge-large-en-v1.5",
            "all-mpnet-base-v2", "e5-large-v2", "qwen3-embedding-0.6b"]


def keep_columns(Ztil, band):
    anchored = {c for c in range(Ztil.shape[1]) if (np.abs(Ztil[:, c]) >= band[c]).any()}
    modal = {int(np.argmax(np.abs(Ztil[i]))) for i in range(Ztil.shape[0])}
    return sorted(anchored | modal)


def residualizer(C, w):
    """Return the operator that projects the covariates (and intercept) out of any column block."""
    A = np.column_stack([np.ones(len(w))] + ([C] if C is not None and C.shape[1] else []))
    sw = np.sqrt(w)[:, None]

    def apply(V):
        beta, *_ = np.linalg.lstsq(A * sw, V * sw, rcond=None)
        return V - A @ beta
    return apply


def main():
    invisible_only = "--invisible" in sys.argv
    items = pd.read_csv(ITEMS_CSV)["item"].astype(str).tolist()
    scales = pd.read_csv(SCALES_CSV).set_index("item").reindex(items)
    cmap = pd.read_csv(CONSTRUCTS_CSV).set_index("item")["construct"]
    con = [str(cmap.get(it, "unlabelled")) for it in items]

    R = pd.read_csv(RESPONSES_CSV)
    R[ID_COL] = R[ID_COL].astype(str)
    R = R.set_index(ID_COL)
    raw = R[items].values.astype(float)
    assert not np.isnan(raw).any(), "missing responses -- use the k0 file"

    lo = scales["min"].values.astype(float)
    hi = scales["max"].values.astype(float)
    rev = scales["reverse"].astype(str).str.lower().eq("true").values
    X = np.clip(2.0 * (raw - lo) / (hi - lo) - 1.0, -1.0, 1.0)
    X[:, rev] *= -1.0

    cov = R[COVARIATES].values.astype(float) if COVARIATES else None
    w = (pd.read_csv(WEIGHTS_CSV).iloc[:, 0].values.astype(float)
         if WEIGHTS_CSV and os.path.exists(WEIGHTS_CSV) else np.ones(len(R)))
    resid = residualizer(cov, w)
    Xr = resid(X)                                # responses with covariates projected out

    sel = pd.read_csv(os.path.join(OUT_DIR, "consensus_outliers.csv"))
    sel[ID_COL] = sel[ID_COL].astype(str)
    target = set(sel.loc[sel["flagged_by_all"].astype(bool), ID_COL])
    label = "consensus outliers"
    if invisible_only:
        cf = pd.read_csv(os.path.join(OUT_DIR, "cutoff_flags.csv"))
        cf[ID_COL] = cf[ID_COL].astype(str)
        inv = set(cf.loc[(cf["breadth"] == 0), ID_COL])
        target &= inv
        label = "INVISIBLE outliers (meet none of the eight cutoffs)"
    idx = np.array([i for i, pid in enumerate(R.index) if pid in target])
    ids = R.index[idx]
    print(f"{label}: {len(idx)} of {len(R):,}\n")
    assert len(idx), "no respondents selected"

    item_share, con_share, comp_top, integ, comp_layer = {}, {}, {}, {}, {}
    for enc in ENCODERS:
        b = np.load(os.path.join(BASIS_DIR, f"{enc}.npz"), allow_pickle=True)
        W, mu, sigma, mu_s, band = b["W"], b["mu"], b["sigma"], b["mu_scores"], b["band"]
        e = np.load(os.path.join(EMB_DIR, f"{EMB_PREFIX}_{enc}.npz"), allow_pickle=True)
        pos = {str(x): j for j, x in enumerate(e["items"])}
        E = np.asarray(e["vectors"], float)[[pos[it] for it in items]]
        P = (E - mu) @ W.T
        kept = keep_columns((P - mu_s) / sigma, band)
        Pk = P[:, kept]

        Y = resid(X @ Pk)
        Yc = Y - np.average(Y, axis=0, weights=w)
        lw = LedoitWolf().fit(Yc)
        Om, Sig = lw.precision_, lw.covariance_
        G = Yc @ Om
        T2 = np.einsum("ij,ij->i", Yc, G)

        # component layer, on the selected respondents
        a = (Yc[idx] * G[idx]) / T2[idx, None]
        u = Yc[idx] ** 2 / np.diag(Sig)
        c = G[idx] ** 2 / np.diag(Om)
        integ[enc] = float(((a * (c > u)).sum(1)).mean())
        comp_top[enc] = [kept[j] for j in np.argsort(-a.mean(0))[:5]]

        # the component layer is SAVED, not only summarized: one row per respondent per kept
        # component, so the attribution can be reported at the layer the basis is built on
        # (share sums to 1 within a respondent, and a negative share is a suppressor)
        comp_layer[enc] = pd.DataFrame({
            ID_COL: np.repeat(np.asarray(ids, dtype=object), len(kept)),
            "component": np.tile(np.asarray(kept, dtype=int), len(idx)),
            "share": a.ravel(), "u": u.ravel(), "c": c.ravel(),
            "joint": (c > u).ravel()})

        # item layer: exact, and in the battery's own vocabulary
        Q = Pk @ Om @ Pk.T
        QX = Xr @ Q
        ai = (Xr[idx] * QX[idx])
        t2i = ai.sum(1)
        assert np.allclose(t2i, T2[idx], rtol=1e-6), "item partition does not reproduce T^2"
        si = ai / t2i[:, None]
        item_share[enc] = pd.DataFrame(si, index=ids, columns=items)
        cs = pd.DataFrame(si, index=ids, columns=items).T.groupby(con).sum().T
        con_share[enc] = cs

    constructs = sorted(set().union(*[set(v.columns) for v in con_share.values()]))
    CS = pd.concat([v.reindex(columns=constructs).fillna(0.0) for v in con_share.values()],
                   keys=ENCODERS)
    mean_cs = CS.groupby(level=1).mean().mean(0).sort_values(ascending=False)

    print(f"{'construct':38s} {'mean share of T2':>17s}   (averaged over encoders and people)")
    for k, v in mean_cs.head(TOP_N).items():
        print(f"  {k:36s} {100*v:15.1f}%")
    neg = mean_cs[mean_cs < 0]
    if len(neg):
        print(f"\nsuppressor constructs (mean share < 0): "
              + ", ".join(f"{k} {100*v:.1f}%" for k, v in neg.items()))

    print(f"\nintegration (share of T2 from joint-type components), per encoder:")
    print("  " + "  ".join(f"{e.split('-')[0]} {100*integ[e]:.0f}%" for e in ENCODERS))

    # per-person reading, which is what a small subset needs
    per = []
    for pid in ids:
        tops = []
        for enc in ENCODERS:
            row = con_share[enc].loc[pid]
            tops.append(row.idxmax())
        cnt = Counter(tops)
        lead, n_agree = cnt.most_common(1)[0]
        avg = pd.concat([con_share[e].loc[pid] for e in ENCODERS], axis=1).mean(1)
        top3 = avg.sort_values(ascending=False).head(3)
        per.append({ID_COL: pid, "top_construct": lead, "encoders_agreeing": n_agree,
                    "top3": "; ".join(f"{k} {100*v:.0f}%" for k, v in top3.items())})
    P = pd.DataFrame(per)
    print(f"\nper-person leading construct ({len(P)} people, agreement out of {len(ENCODERS)} encoders)")
    print(P.to_string(index=False))
    print(f"\nleading construct agrees across >= 4 of 6 encoders for "
          f"{int((P['encoders_agreeing'] >= 4).sum())} of {len(P)}")

    tag = "invisible" if invisible_only else "outliers"
    os.makedirs(OUT_DIR, exist_ok=True)
    mean_cs.rename("mean_share").to_frame().to_csv(os.path.join(OUT_DIR, f"attribution_constructs_{tag}.csv"))
    P.to_csv(os.path.join(OUT_DIR, f"attribution_per_person_{tag}.csv"), index=False)
    pd.concat([item_share[e] for e in ENCODERS], keys=ENCODERS).to_csv(
        os.path.join(OUT_DIR, f"attribution_item_shares_{tag}.csv"))
    pd.concat([comp_layer[e] for e in ENCODERS], keys=ENCODERS,
              names=["encoder", None]).reset_index(level=0).to_csv(
        os.path.join(OUT_DIR, f"attribution_component_shares_{tag}.csv"), index=False)
    json.dump({"n": len(idx), "subset": label, "integration": integ,
               "leading_components": {e: comp_top[e] for e in ENCODERS}},
              open(os.path.join(OUT_DIR, f"attribution_summary_{tag}.json"), "w"), indent=2)
    print(f"\nwrote -> {OUT_DIR}")


if __name__ == "__main__":
    main()
```
