## Supplemental code for "Scoring respondents on a semantic frame of reference identifies a group that is burdened across many questionnaires and reaches care less easily": attribution_components.py

```
#!/usr/bin/env python3
"""attribution_components.py -- what a flag is made of, at the component layer.

R4 reports the attribution on the components the basis is built from, never on items, so the flag
comes back in the same objects R1 and R2 interpret. A component is named by its CONSTRUCT PROFILE
(DEFINITIONS D6-A), and the profile of record here is the BATTERY-SIDE one: the documented constructs
of the All of Us items that core that component WITHIN the battery, which is the rule main-text S2.2
states for a placed battery. The pool-side profile, the constructs of the pool items that core the
same component within the pool, is printed alongside it for reference and is never scored against it.
Both are read after everything is computed and play no part in any computation.

Four things are measured, all on one encoder at a time (gte-large by default, the exemplar):

  COMPOSITION   the mean share of the distance each kept component carries across the group
  SUPPRESSORS   how much of the distance sits in NEGATIVE shares, since share_j = a_j / T^2 sums to
                exactly 1 per person only if the negatives are kept. This decides whether they are a
                clause or a finding, and is measured rather than assumed away
  CONCENTRATION how much of one person's distance their largest component carries, and how many
                components it takes to reach half of it
  LEADING vs    a component LEADS a person when it is their largest share and CARRIES them when it is
  CARRYING      in their half-set, the components cumulating to half the distance. Carrying contains
                leading, so the ratio says whether a component dominates whenever it is in play or is
                a broad background. No share threshold: the discrete levels are retired
  CHARACTER     u_j (extreme on its own) against c_j (extreme given the rest), per component. The
                component is JOINT when c_j > u_j. Note |a_j| = sqrt(u_j c_j / (1 - R_j^2)), so a
                component cannot carry much unless BOTH are non-trivial -- a_j says how much is
                carried, u_j against c_j says which of the two ways it got there
  IS CHARACTER  a control on the above. A component with a small core might be joint for a mechanical
  AN ARTIFACT   reason rather than because of its content, so the joint fraction is correlated against
                core size, profile size and mean share, and reported binned by core size. If jointness
                tracked core size, "this content is unexpected given the rest" would be a statement
                about the basis rather than about the respondents

Reads only what the pipeline wrote, and hardcodes no result:
    screen_out/attribution_component_shares_<tag>.csv   (attribution.py) -- encoder,id,component,share,u,c,joint
    reference_basis/<enc>.npz, item_embeddings/, construct_map_151.csv -- for the battery-side profile
    reference_basis/pool_components.csv                 -- the pool-side profile, for reference

Out: prints, and screen_out/component_report_<enc>_<tag>.csv (one row per kept component)
Run: python3 attribution_components.py [--enc gte-large-en-v1.5] [--tag outliers]
"""
import os
import sys
from collections import Counter

import numpy as np
import pandas as pd

HERE = os.path.dirname(os.path.abspath(__file__))
OUT_DIR = os.path.join(HERE, "screen_out")
BASIS_DIR = os.path.join(HERE, "reference_basis")
EMB_DIR = os.path.join(HERE, "item_embeddings")
EMB_PREFIX = "core151_items"
ITEMS_CSV = os.path.join(HERE, "core_items_151.csv")
CONSTRUCTS_CSV = os.path.join(HERE, "construct_map_151.csv")
POOL_COMPONENTS = os.path.join(BASIS_DIR, "pool_components.csv")
ID_COL = "id"
DEFAULT_ENC = "gte-large-en-v1.5"
TOP_CONSTRUCTS = 4          # how many of a profile's constructs to print, by item count


def arg(flag, default):
    return sys.argv[sys.argv.index(flag) + 1] if flag in sys.argv else default


def pr(v):
    s2 = float((v ** 2).sum()); s4 = float((v ** 4).sum())
    return s2 * s2 / s4 if s4 > 0 else 0.0


def fpc(v):
    """Fixed-point participation-ratio core of a vector. Mirrors examples/allofus/_common usage --
    if one changes, change both."""
    idx = np.arange(len(v))
    while True:
        n = max(1, min(int(round(pr(v[idx]))), len(idx)))
        top = np.sort(idx[np.argsort(-np.abs(v[idx]))[:n]])
        if len(top) == len(idx) and (top == idx).all():
            return top
        idx = top


def battery_profiles(enc):
    """component -> (battery-side profile string, battery-side core size).

    The battery-side core of a component is the fixed-point participation-ratio core down that
    component's column WITHIN the 151 items, and its profile is the documented constructs of those
    items. This is the rule main-text S2.2 states for interpreting a placed battery.
    """
    items = pd.read_csv(ITEMS_CSV)["item"].astype(str).tolist()
    cmap = pd.read_csv(CONSTRUCTS_CSV).set_index("item")["construct"]
    con = [str(cmap.get(it, "unlabelled")) for it in items]

    b = np.load(os.path.join(BASIS_DIR, f"{enc}.npz"), allow_pickle=True)
    W, mu, sigma, mu_s = b["W"], b["mu"], b["sigma"], b["mu_scores"]
    e = np.load(os.path.join(EMB_DIR, f"{EMB_PREFIX}_{enc}.npz"), allow_pickle=True)
    pos = {str(x): j for j, x in enumerate(e["items"])}
    E = np.asarray(e["vectors"], float)[[pos[it] for it in items]]
    Z = ((E - mu) @ W.T - mu_s) / sigma

    prof, size = {}, {}
    for c in range(Z.shape[1]):
        core = fpc(Z[:, c])
        counts = Counter(con[j] for j in core)
        prof[c] = "; ".join(f"{k}:{v}" for k, v in counts.most_common())
        size[c] = len(core)
    return prof, size


def pool_profiles(enc):
    """component -> pool-side profile string, from the shipped basis. For reference only."""
    pc = pd.read_csv(POOL_COMPONENTS)
    pc = pc[pc["encoder"] == enc]
    assert len(pc), f"no components for {enc} in pool_components.csv"
    return dict(zip(pc["component"], pc["construct_profile"]))


def main():
    enc = arg("--enc", DEFAULT_ENC)
    tag = arg("--tag", "outliers")
    src = os.path.join(OUT_DIR, f"attribution_component_shares_{tag}.csv")
    D = pd.read_csv(src)
    D = D[D["encoder"] == enc].copy()
    assert len(D), f"no rows for {enc} in {os.path.basename(src)}"
    prof, ncore = battery_profiles(enc)
    pprof = pool_profiles(enc)

    n_people = D[ID_COL].nunique()
    kept = sorted(D["component"].unique())
    print(f"\n{'='*100}\ncomponent-layer attribution, {enc}, tag={tag}")
    print(f"{n_people} respondents, {len(kept)} kept components\n")

    tot = D.groupby(ID_COL)["share"].sum()
    print(f"  partition check: per-person shares sum to {tot.min():.6f}-{tot.max():.6f} (exactly 1 "
          f"if the partition is complete)")

    # ---- suppressors: how much of the distance is carried by negative shares
    neg = D[D["share"] < 0]
    neg_mass = neg.groupby(ID_COL)["share"].sum().reindex(D[ID_COL].unique()).fillna(0.0)
    neg_count = neg.groupby(ID_COL).size().reindex(D[ID_COL].unique()).fillna(0)
    pos_mass = D[D["share"] > 0].groupby(ID_COL)["share"].sum()
    print(f"\n  SUPPRESSORS (negative shares)")
    print(f"    per person the negatives sum to a median {100*neg_mass.median():.1f}% of the "
          f"distance (mean {100*neg_mass.mean():.1f}%, most negative {100*neg_mass.min():.1f}%)")
    print(f"    a person has a median {neg_count.median():.0f} negative components of {len(kept)} "
          f"(range {int(neg_count.min())}-{int(neg_count.max())})")
    print(f"    the positive shares therefore sum to a median {100*pos_mass.median():.1f}% of the "
          f"distance, which is what renormalizing over positives alone would rescale")

    # ---- concentration within a person
    piv = D.pivot_table(index=ID_COL, columns="component", values="share").fillna(0.0)
    top = piv.max(axis=1)
    lead = piv.idxmax(axis=1)

    def n_to_half(row):
        s = np.sort(row.values)[::-1]
        return int(np.searchsorted(np.cumsum(s), 0.5) + 1)
    half = piv.apply(n_to_half, axis=1)

    # LEADING and CARRYING. A component LEADS a person when it is their largest share, so every person
    # has exactly one leader and rank 1 is forced. It CARRIES them when it is in their HALF-SET, the
    # components whose shares cumulate to half the distance -- the same set five_xenc.py compares
    # across encoders and the same one main-text S2.4 means by "two or three components reach half".
    # Carrying contains leading, so n_carries >= n_leads, and the ratio is the reportable contrast:
    # a component that carries many but leads few of them is a broad background, one that leads
    # nearly everyone it carries dominates whenever it is in play. No share threshold is involved --
    # the discrete levels this replaces were retired with fig_construct_ranks.py.
    cols = piv.columns.values
    order = np.argsort(-piv.values, axis=1)
    n_half = (np.cumsum(np.take_along_axis(piv.values, order, axis=1), axis=1) < 0.5).sum(axis=1) + 1
    carries = Counter()
    for i, n in enumerate(n_half):
        carries.update(cols[order[i, :n]])
    n_carries = pd.Series(carries).reindex(kept).fillna(0).astype(int)

    print(f"\n  CONCENTRATION within a person")
    print(f"    the largest component carries a median {100*top.median():.1f}% of the distance "
          f"(quartiles {100*top.quantile(.25):.1f}-{100*top.quantile(.75):.1f}%)")
    print(f"    it takes a median {half.median():.0f} components to reach half the distance "
          f"(quartiles {half.quantile(.25):.0f}-{half.quantile(.75):.0f})")
    print(f"    {lead.nunique()} distinct components lead at least one person, "
          f"{int((n_carries > 0).sum())} carry at least one")

    # ---- character: joint against marginal
    integration = D.assign(j=D["share"] * D["joint"]).groupby(ID_COL)["j"].sum()
    print(f"\n  CHARACTER")
    print(f"    integration, the share of the distance carried by joint components "
          f"(c > u), is a median {100*integration.median():.1f}% per person "
          f"(mean {100*integration.mean():.1f}%)")
    print(f"    {100*D['joint'].mean():.1f}% of all person-component cells are joint")

    # ---- per-component table
    g = D.groupby("component")
    T = pd.DataFrame({
        "mean_share": 100 * g["share"].mean(),
        "pct_of_distance_when_positive": 100 * g["share"].apply(lambda s: s[s > 0].mean()),
        "n_leads": lead.value_counts().reindex(kept).fillna(0).astype(int),
        "n_carries": n_carries,
        "pct_lead_when_carrying": 100 * lead.value_counts().reindex(kept).fillna(0) /
                                  n_carries.replace(0, np.nan),
        "pct_joint": 100 * g["joint"].mean(),
        "median_u": g["u"].median(),
        "median_c": g["c"].median(),
        "battery_core_items": pd.Series({c: ncore.get(c, np.nan) for c in kept}),
        "battery_profile": pd.Series({c: prof.get(c, "(no battery core)") for c in kept}),
        "pool_profile": pd.Series({c: pprof.get(c, "(not in the pool basis)") for c in kept}),
    }).sort_values("mean_share", ascending=False)

    # ---- is character an artifact of core size? ----
    def spearman(a, b):
        ra, rb = pd.Series(a).rank().values, pd.Series(b).rank().values
        return float(np.corrcoef(ra, rb)[0, 1]) if len(a) > 2 else float("nan")

    A = T.copy()
    A["n_constructs"] = [len(str(p).split(";")) for p in A["battery_profile"]]
    ok = A["battery_core_items"].notna()
    print(f"\n  IS CHARACTER AN ARTIFACT OF CORE SIZE? (Spearman over {int(ok.sum())} components)")
    for col, lab in (("battery_core_items", "core items"), ("n_constructs", "constructs in profile"),
                     ("mean_share", "mean share")):
        print(f"    joint% against {lab:22s} {spearman(A.loc[ok, 'pct_joint'], A.loc[ok, col]):+.2f}")
    print(f"    {'core items':>12s} {'n':>4s} {'mean joint%':>12s} {'mean share%':>12s}")
    for lo, hi in ((1, 2), (3, 4), (5, 8), (9, 99)):
        g = A[ok & A["battery_core_items"].between(lo, hi)]
        if len(g):
            print(f"    {f'{lo}-{hi}':>12s} {len(g):4d} {g['pct_joint'].mean():11.1f}% "
                  f"{g['mean_share'].mean():11.2f}%")
    print(f"    a flat column means jointness is a property of the content, not of the core size")

    def short(p, n=TOP_CONSTRUCTS):
        return "; ".join(str(p).split("; ")[:n])

    print(f"\n  the kept components by mean share of the distance, named by the constructs of the")
    print(f"  All of Us items that core them WITHIN the battery (the pool's own naming beneath)\n")
    print(f"    {'comp':>5s} {'mean%':>7s} {'leads':>6s} {'carries':>8s} {'lead|carry':>11s} "
          f"{'joint%':>7s} {'u':>7s} {'c':>7s}  battery-side profile")
    for cid, r in T.iterrows():
        lw = f"{r['pct_lead_when_carrying']:10.1f}%" if pd.notna(r['pct_lead_when_carrying']) else \
             f"{'--':>11s}"
        print(f"    C{int(cid):<4d} {r['mean_share']:6.2f}% {int(r['n_leads']):6d} "
              f"{int(r['n_carries']):8d} {lw} "
              f"{r['pct_joint']:6.1f}% {r['median_u']:7.2f} {r['median_c']:7.2f}  "
              f"{short(r['battery_profile'])[:72]}")
        print(f"    {'':>5s} {'':>7s} {'':>6s} {'':>8s} {'':>11s} {'':>7s} {'':>7s} {'':>7s}  pool: "
              f"{short(r['pool_profile'])[:66]}")

    out = os.path.join(OUT_DIR, f"component_report_{enc}_{tag}.csv")
    T.rename_axis("component").to_csv(out)
    print(f"\n  wrote {out}")


if __name__ == "__main__":
    main()
```
