## Supplemental code for "Scoring respondents on a semantic frame of reference identifies a group that is burdened across many questionnaires and reaches care less easily": construct_concentration.py

```
#!/usr/bin/env python3
"""construct_concentration.py -- breadth and intensity of each construct, with no thresholds.

Replaces the discrete LEVELS reading of construct_distribution.py (n_ge10 / n_ge20 / n_ge30). Those
counts mix two things that should be separate -- how many people a construct touches, and how hard it
hits them -- so a construct that is rare but dominant (interpersonal & sexual violence) is
indistinguishable from one that does nothing. Both show a small mean, few leaders, and small counts
at every cut.

THE DECOMPOSITION. For construct c, with s_p its share of participant p's squared distance, averaged
over the six encoders exactly as construct_distribution.py does:

    M_c = sum_p s_p+                     total mass
    N_c = (sum_p s_p+)^2 / sum_p (s_p+)^2    effective breadth -- the participation ratio, i.e. the
                                             effective number of participants carrying it
    I_c = M_c / N_c                      intensity -- the share a typical carrier gives it

    M_c = N_c * I_c                      exact, by construction, and free of any cutoff

A construct spread evenly over 200 people has N_c near 200. One concentrated in four has N_c near 4.
The participation ratio is the same parameter-free operator the pool cores are defined by (D4/D5), so
this introduces no new concept for a reader.

POSITIVE PART, NEGATIVE MASS REPORTED SEPARATELY (settled 2026-08-21). A share can be negative --
a_i = xr_i (Q xr)_i < 0 is a suppressor, an answer that pulls the distance DOWN given the rest. The
participation ratio needs non-negative input, and a suppressor is not carrying anything, so M/N/I are
computed on s+ = max(s, 0) and the negative mass is reported in its own columns rather than folded in
or discarded silently.

Reads screen_out/attribution_item_shares_<tag>.csv, written by attribution.py. Recomputes nothing.

  M_pct        M_c as a percentage of the total positive mass across constructs -- the diffuse
               composition number, comparable to the group-mean share already reported
  N_eff        effective breadth, in participants
  N_pct        N_eff as a percentage of the group
  intensity    I_c, as a percentage of a carrier's squared distance
  N_range      min-max of N_eff computed per encoder, so breadth is not one encoder's arithmetic
  I_range      the same for intensity
  neg_mass     total negative (suppressor) mass, same units as M_c
  n_neg        participants for whom the construct's mean share is negative
  n_leads      participants for whom it is the largest share (kept from the old table for continuity)

AGGREGATION. By default the per-person shares are averaged over the six encoders and the PR is taken
on that average, mirroring construct_distribution.py. That order SMOOTHS the per-person distribution
and therefore biases N up and intensity down -- for several constructs the pooled intensity falls
below every per-encoder value, which is the signature. `--encoder <name>` computes everything within
one encoder instead, which is free of that bias. The N_range / I_range columns are always the
per-encoder spread, so they can be compared against whichever mode is being read.

  python3 construct_concentration.py
  python3 construct_concentration.py --encoder gte          # one encoder, no averaging bias
  python3 construct_concentration.py --encoder all          # each of the six in turn
  python3 construct_concentration.py --tag invisible
"""
import os
import sys

import numpy as np
import pandas as pd

HERE = os.path.dirname(os.path.abspath(__file__))
OUT_DIR = os.path.join(HERE, "screen_out")
CONSTRUCTS_CSV = os.path.join(HERE, "construct_map_151.csv")
ID_COL = "id"


def breadth_intensity(s):
    """(mass, effective breadth, intensity) of one construct's per-person shares, positive part."""
    p = np.clip(np.asarray(s, float), 0.0, None)
    m = float(p.sum())
    if m <= 0:
        return 0.0, 0.0, 0.0
    n = m * m / float((p ** 2).sum())
    return m, n, m / n


def main():
    tag = "outliers"
    if "--tag" in sys.argv:
        tag = sys.argv[sys.argv.index("--tag") + 1]
    want_enc = sys.argv[sys.argv.index("--encoder") + 1] if "--encoder" in sys.argv else None

    path = os.path.join(OUT_DIR, f"attribution_item_shares_{tag}.csv")
    assert os.path.exists(path), f"{path} not found -- run attribution.py first"
    sh = pd.read_csv(path, index_col=[0, 1])
    sh.index = sh.index.set_names(["encoder", ID_COL])
    sh.index = sh.index.set_levels(sh.index.levels[1].astype(str), level=1)

    cmap = pd.read_csv(CONSTRUCTS_CSV).set_index("item")["construct"]
    con = cmap.reindex(sh.columns).fillna("unlabelled").values
    C = sh.T.groupby(con).sum().T                       # (encoder, id) x construct

    encoders = list(C.index.get_level_values("encoder").unique())
    if want_enc and want_enc != "all":
        hit = [e for e in encoders if e == want_enc or e.startswith(want_enc)]
        assert len(hit) == 1, f"--encoder {want_enc!r} matched {hit or 'nothing'} of {encoders}"
        want_enc = hit[0]
    targets = encoders if want_enc == "all" else [want_enc]

    for which in targets:
        report(C, encoders, which, tag)


def report(C, encoders, which, tag):
    """`which` is None for the encoder-averaged reading, or one encoder name."""
    if which is None:
        M = C.groupby(level=ID_COL).mean()              # ids x constructs, averaged over encoders
        head = f"averaged over {len(encoders)} encoders (smooths the distribution -- see AGGREGATION)"
        suffix = ""
    else:
        M = C.xs(which, level="encoder")
        head = f"encoder {which} only (no averaging bias)"
        suffix = "_" + which
    n_people = len(M)
    leader = M.idxmax(axis=1)

    rows = []
    for c in M.columns:
        s = M[c]
        mass, neff, inten = breadth_intensity(s)
        per_enc = [breadth_intensity(C.xs(e, level="encoder")[c]) for e in encoders]
        neg = abs(float(np.clip(s.values, None, 0.0).sum()))   # abs: avoids printing -0.0
        rows.append({"construct": c,
                     "M": mass, "N_eff": neff, "intensity": inten,
                     "N_lo": min(x[1] for x in per_enc), "N_hi": max(x[1] for x in per_enc),
                     "I_lo": min(x[2] for x in per_enc), "I_hi": max(x[2] for x in per_enc),
                     "neg_mass": neg, "n_neg": int((s < 0).sum()),
                     "n_leads": int((leader == c).sum())})

    T = pd.DataFrame(rows).set_index("construct")
    total_mass = T["M"].sum()
    T["M_pct"] = 100 * T["M"] / total_mass
    T["N_pct"] = 100 * T["N_eff"] / n_people

    print(f"\n{'='*100}")
    print(f"{n_people} respondents, {C.shape[1]} constructs -- {head}")
    print(f"total positive mass {total_mass:.1f}, total suppressor mass {T['neg_mass'].sum():.1f} "
          f"(each person's shares sum to 1 across constructs before clipping)\n")

    def show(title, key, asc):
        print(f"--- {title}\n")
        print(f"{'construct':34s} {'M%':>6s} {'N_eff':>7s} {'N%':>6s} {'intens':>7s} "
              f"{'N range':>13s} {'I range':>13s} {'neg':>6s} {'leads':>6s}")
        for c, r in T.sort_values(key, ascending=asc).iterrows():
            print(f"{str(c)[:34]:34s} {r['M_pct']:5.1f}% {r['N_eff']:7.1f} {r['N_pct']:5.1f}% "
                  f"{100*r['intensity']:6.1f}% {r['N_lo']:5.0f}-{r['N_hi']:<7.0f} "
                  f"{100*r['I_lo']:5.1f}-{100*r['I_hi']:<7.1f} {r['neg_mass']:6.1f} "
                  f"{int(r['n_leads']):6d}")
        print()

    show("by INTENSITY -- what a construct is worth to the people it does carry", "intensity", False)
    show("by MASS -- what a construct is worth to the group as a whole", "M_pct", False)

    print("reading the table")
    print("  high intensity + low N_eff  = rare but dominant, the whole story for a few people")
    print("  low intensity  + high N_eff = broad background, drives nobody")
    print("  high on both                = the constructs that carry the group")
    print("  M = N_eff x intensity exactly, so the two columns decompose the mass with no cutoff.")

    T.to_csv(os.path.join(OUT_DIR, f"construct_concentration_{tag}{suffix}.csv"))
    print(f"\nwrote -> {OUT_DIR}/construct_concentration_{tag}{suffix}.csv")


if __name__ == "__main__":
    main()
```
