## Supplemental code for "Scoring respondents on a semantic frame of reference identifies a group that is burdened across many questionnaires and reaches care less easily": construct_distribution.py

```
#!/usr/bin/env python3
"""construct_distribution.py -- is a construct a diffuse background, or a driving force in a few?

A group mean and a count of who it leads both hide the same thing. A construct carrying 25% of the
distance for 20 people and nothing for the other 490 has a small mean and few leaders, yet it is the
whole story for those 20. This script reports the DISTRIBUTION of each construct's per-person share.

Reads screen_out/attribution_item_shares_<tag>.csv, written by attribution.py. Recomputes nothing.

  mean, median, p90, max      the shape of the per-person share
  n_ge10 / n_ge20 / n_ge30    how many people it carries substantially
  top10pct_mass               of all the mass this construct holds across the group, the fraction
                              held by its top 10% of people. ~0.1 = spread evenly, >=0.4 = a few
                              people carry it. This is the diffuse-versus-driving-force number.
  n_robust20                  people with >= 20% under at least 4 of the 6 encoders, so a large
                              share is not one encoder's arithmetic
  n_leads                     people for whom it is the largest share

  python3 construct_distribution.py
  python3 construct_distribution.py --tag invisible
  python3 construct_distribution.py --mark screen_out/attribution_per_person_invisible.csv
      ... adds a column counting, among the marked ids, how many that construct carries >= 20%
"""
import os
import sys

import numpy as np
import pandas as pd

HERE = os.path.dirname(os.path.abspath(__file__))
OUT_DIR = os.path.join(HERE, "screen_out")
CONSTRUCTS_CSV = os.path.join(HERE, "construct_map_151.csv")
ID_COL = "id"
ROBUST_MIN_ENC = 4
LEVELS = (0.10, 0.20, 0.30)


def main():
    tag = "outliers"
    if "--tag" in sys.argv:
        tag = sys.argv[sys.argv.index("--tag") + 1]
    mark_ids = None
    if "--mark" in sys.argv:
        mp = sys.argv[sys.argv.index("--mark") + 1]
        mark_ids = set(pd.read_csv(mp)[ID_COL].astype(str))

    path = os.path.join(OUT_DIR, f"attribution_item_shares_{tag}.csv")
    assert os.path.exists(path), f"{path} not found -- run attribution.py first"
    sh = pd.read_csv(path, index_col=[0, 1])
    sh.index = sh.index.set_names(["encoder", ID_COL])
    sh.index = sh.index.set_levels(sh.index.levels[1].astype(str), level=1)

    cmap = pd.read_csv(CONSTRUCTS_CSV).set_index("item")["construct"]
    con = cmap.reindex(sh.columns).fillna("unlabelled").values
    C = sh.T.groupby(con).sum().T                       # (encoder, id) x construct

    encoders = list(C.index.get_level_values("encoder").unique())
    per_enc_ge20 = (C >= 0.20).groupby(level=ID_COL).sum()          # ids x constructs, 0..n_enc
    M = C.groupby(level=ID_COL).mean()                              # ids x constructs, mean share
    n = len(M)
    leader = M.idxmax(axis=1)

    rows = []
    for c in M.columns:
        s = M[c]
        pos = s.clip(lower=0)                                       # mass, ignoring suppressors
        k = max(1, int(round(0.10 * n)))
        top_mass = pos.nlargest(k).sum() / pos.sum() if pos.sum() > 0 else np.nan
        r = {"construct": c,
             "mean": round(100 * s.mean(), 1),
             "median": round(100 * s.median(), 1),
             "p90": round(100 * s.quantile(0.90), 1),
             "max": round(100 * s.max(), 1),
             "n_leads": int((leader == c).sum())}
        for lv in LEVELS:
            r[f"n_ge{int(100*lv)}"] = int((s >= lv).sum())
        r["top10pct_mass"] = round(float(top_mass), 2)
        r["n_robust20"] = int((per_enc_ge20[c] >= ROBUST_MIN_ENC).sum())
        if mark_ids is not None:
            hit = s.index.isin(mark_ids) & (s >= 0.20).values
            r["marked_ge20"] = int(hit.sum())
        rows.append(r)

    T = pd.DataFrame(rows).set_index("construct")
    T = T.sort_values("n_ge20", ascending=False)
    print(f"{n} respondents, {len(encoders)} encoders, {C.shape[1]} constructs "
          f"(shares are % of each person's squared distance)\n")
    print(T.to_string())

    print("\nreading the table")
    print("  a construct with a small mean but a high top10pct_mass and a real n_ge20 carries a few")
    print("  people hard. one with a moderate mean and top10pct_mass near 0.1 is a broad background.")
    conc = T[(T["n_ge20"] >= 3)].sort_values("top10pct_mass", ascending=False)
    if len(conc):
        print("\nmost concentrated among the constructs carrying at least 3 people to 20%:")
        for c, r in conc.head(6).iterrows():
            print(f"  {c:34s} mean {r['mean']:5.1f}%  n>=20% {int(r['n_ge20']):4d}  "
                  f"robust {int(r['n_robust20']):4d}  top10%mass {r['top10pct_mass']:.2f}")

    T.to_csv(os.path.join(OUT_DIR, f"construct_distribution_{tag}.csv"))
    print(f"\nwrote -> {OUT_DIR}/construct_distribution_{tag}.csv")


if __name__ == "__main__":
    main()
```
