## Supplemental code for "Scoring respondents on a semantic frame of reference identifies a group that is burdened across many questionnaires and reaches care less easily": cutoffs.py

```
#!/usr/bin/env python3
"""cutoffs.py -- the eight validated clinical cutoffs, and how the screen's outliers sit against them.

STANDALONE (numpy, pandas). Reads the response file built by build_responses_v2 and the consensus
outlier list written by screen_battery.py.

Cutoffs are evaluated on the CODEBOOK CODING, on each instrument's own response scale and in its
original direction -- NOT on the rescaled and reverse-coded matrix the screen uses. The validated
scoring is defined on that scale, so applying it to the rescaled matrix would not be the published
criterion. This is the same convention the manuscript states.

THE EIGHT CRITERIA (Table tab:cutoffs), each traced to the items that carry it

  PHQ-9      sum of the 9 items (0-3) >= 10, OR phq9_9 > 0  (the self-harm item, endorsed at all)
  GAD-7      sum of the 7 items (0-3) >= 10
  ACE        >= 4 of 7 exposure CATEGORIES. The BRFSS module scores categories, not items: the
             three sexual-abuse items collapse to one category, problem drinking and drug use are
             scored together as one household-substance category, and the household-incarceration
             category is not among the 151. So 7 categories come from the 10 items.
             A binary item counts when Yes, a frequency item when Once or More than once.
             The two collapses follow the instrument rather than convenience. Felitti asks household
             substance abuse as ONE question -- problem drinker or alcoholic, OR street drugs -- which
             BRFSS splits into ace_2 and ace_3, and the three sexual-abuse questions likewise score
             one category. The standard eight are emotional abuse, physical abuse, sexual abuse,
             household mental illness, household substance abuse, witnessed domestic violence,
             parental separation or divorce, and household incarceration. Matches
             compare_outliers_v3 Cell 5, which produced the published 23.1% cohort rate.
  ASRS       >= 4 of the 6 Part-A items at clinical frequency: Sometimes or worse (>= 2) for items
             1-3, Often or worse (>= 3) for items 4-6
  WMH-CIDI   any of the three psychotic-experience items endorsed (voices, signs and signals, plot)
  Suicide    any of ss_1 / ss_2 / ss_3 endorsed (self-harm without intent to die, ideation, attempt)
  HVS        either vigilance item Sometimes true or Often true (>= 2)
  PROMIS     self-rated general mental health Fair or Poor (<= 2)

OUTPUT (-> OUT_DIR)
  cutoff_flags.csv     id, one column per instrument, breadth (0-8), any_of_8, is_outlier
  cutoff_summary.csv   per instrument: % cohort and % outliers, with Wilson 95% intervals
  breadth_table.csv    the 0-8 breadth distribution, cohort against outliers

Run:  python3 cutoffs.py
"""
import math
import os

import numpy as np
import pandas as pd

HERE = os.path.dirname(os.path.abspath(__file__))
RESPONSES_CSV = "/home/jupyter/responses_out/responses_k0.csv"
OUTLIERS_CSV  = os.path.join(HERE, "screen_out", "consensus_outliers.csv")
OUT_DIR       = os.path.join(HERE, "screen_out")
ID_COL        = "id"

PHQ = [f"phq9_{i}" for i in range(1, 10)]
GAD = [f"gad7_{i}" for i in range(1, 8)]
ASRS_INATT, ASRS_HYPER = ["asrs_1", "asrs_2", "asrs_3"], ["asrs_4", "asrs_5", "asrs_6"]
CIDI_PSYCH = ["cidi5_21", "cidi5_22", "cidi5_23"]
SUICIDE = ["ss_1", "ss_2", "ss_3"]
HVS = ["hvs_1", "hvs_2"]
SRMH = "overallhealth_generalmentalhealth"
# BRFSS ACE categories: name -> the items that can trigger it (any one suffices)
ACE_CATEGORIES = {
    "household mental illness": ["ace_1"],
    "household substance abuse": ["ace_2", "ace_3"],    # ONE category: Felitti asks alcohol OR drugs
    "parental separation or divorce": ["ace_5"],
    "witnessed domestic violence": ["ace_6"],
    "physical abuse": ["ace_7"],
    "emotional abuse": ["ace_8"],
    "sexual abuse": ["ace_9", "ace_10", "ace_11"],      # ONE category from three questions
}
# Endorsement is >= 0 for every ACE item, which covers all three codings the module uses:
#   binary      -1 No   / 1 Yes                       -> 1 only
#   frequency   -1 Never / 0 Once / 1 More than once   -> Once or more
#   ace_5       -1 No, -1 Parents not married / 1 Yes  -> 1 only
ACE_ENDORSED = lambda v: v >= 0


def wilson(k, n, z=1.959963985):
    if n == 0:
        return float("nan"), float("nan")
    p = k / n
    d = 1 + z * z / n
    c = (p + z * z / (2 * n)) / d
    h = z * math.sqrt(p * (1 - p) / n + z * z / (4 * n * n)) / d
    return 100 * (c - h), 100 * (c + h)


def main():
    R = pd.read_csv(RESPONSES_CSV)
    R[ID_COL] = R[ID_COL].astype(str)
    R = R.set_index(ID_COL)

    need = PHQ + GAD + ASRS_INATT + ASRS_HYPER + CIDI_PSYCH + SUICIDE + HVS + [SRMH] \
        + [i for v in ACE_CATEGORIES.values() for i in v]
    absent = [c for c in need if c not in R.columns]
    assert not absent, f"items absent from the response file: {absent}"

    ace_hits = []
    for cat, its in ACE_CATEGORIES.items():
        hit = np.zeros(len(R), bool)
        for it in its:
            hit |= ACE_ENDORSED(R[it].values.astype(float))
        ace_hits.append(hit)
    ace_count = np.sum(ace_hits, axis=0)

    asrs_count = sum((R[i].values.astype(float) >= 2).astype(int) for i in ASRS_INATT) \
        + sum((R[i].values.astype(float) >= 3).astype(int) for i in ASRS_HYPER)

    F = pd.DataFrame(index=R.index)
    F["PHQ-9"] = (R[PHQ].sum(axis=1).values >= 10) | (R["phq9_9"].values > 0)
    F["GAD-7"] = R[GAD].sum(axis=1).values >= 10
    F["ACE"] = ace_count >= 4
    F["ASRS"] = asrs_count >= 4
    F["WMH-CIDI"] = (R[CIDI_PSYCH].values == 1).any(axis=1)
    F["Suicide screen"] = (R[SUICIDE].values == 1).any(axis=1)
    F["HVS"] = (R[HVS].values >= 2).any(axis=1)
    F["PROMIS mental health"] = R[SRMH].values <= 2

    INSTR = list(F.columns)
    F["breadth"] = F[INSTR].sum(axis=1)
    F["any_of_8"] = F["breadth"] > 0

    o = pd.read_csv(OUTLIERS_CSV)
    o[ID_COL] = o[ID_COL].astype(str)
    out_ids = set(o.loc[o["flagged_by_all"].astype(bool), ID_COL])
    assert out_ids <= set(F.index), "an outlier id is absent from the response file"
    F["is_outlier"] = F.index.isin(out_ids)
    n_out, n_coh = int(F["is_outlier"].sum()), len(F)
    print(f"cohort {n_coh:,} | consensus outliers {n_out:,}\n")

    rows = []
    for name in INSTR + ["any_of_8"]:
        kc, ko = int(F[name].sum()), int(F.loc[F["is_outlier"], name].sum())
        cl, ch = wilson(kc, n_coh)
        ol, oh = wilson(ko, n_out)
        rows.append({"criterion": name, "n_cohort": kc, "pct_cohort": round(100 * kc / n_coh, 1),
                     "cohort_lo": round(cl, 1), "cohort_hi": round(ch, 1),
                     "n_outliers": ko, "pct_outliers": round(100 * ko / n_out, 1),
                     "outliers_lo": round(ol, 1), "outliers_hi": round(oh, 1)})
    S = pd.DataFrame(rows)
    print(S.to_string(index=False))

    bo = F.loc[F["is_outlier"], "breadth"]
    bc = F["breadth"]
    B = pd.DataFrame({"breadth": range(9),
                      "pct_cohort": [round(100 * (bc == k).mean(), 1) for k in range(9)],
                      "pct_outliers": [round(100 * (bo == k).mean(), 1) for k in range(9)],
                      "n_outliers": [int((bo == k).sum()) for k in range(9)]})
    print(f"\nbreadth (number of the eight criteria met)")
    print(B.to_string(index=False))
    print(f"\nmean breadth: outliers {bo.mean():.2f}  cohort {bc.mean():.2f}   mode {int(bo.mode()[0])}")
    for t in (2, 5, 8):
        k = int((bo >= t).sum())
        lo, hi = wilson(k, n_out)
        print(f"  outliers meeting >= {t}: {k} ({100*k/n_out:.0f}%, 95% CI {lo:.0f}-{hi:.0f})")
    inv = F.index[F["is_outlier"] & (F["breadth"] == 0)]
    lo, hi = wilson(len(inv), n_out)
    print(f"\nINVISIBLE outliers, meeting none of the eight: {len(inv)} of {n_out} "
          f"({100*len(inv)/n_out:.1f}%, 95% CI {lo:.1f}-{hi:.1f})")

    os.makedirs(OUT_DIR, exist_ok=True)
    F.rename_axis(ID_COL).reset_index().to_csv(os.path.join(OUT_DIR, "cutoff_flags.csv"), index=False)
    S.to_csv(os.path.join(OUT_DIR, "cutoff_summary.csv"), index=False)
    B.to_csv(os.path.join(OUT_DIR, "breadth_table.csv"), index=False)
    print(f"\nwrote -> {OUT_DIR}")


if __name__ == "__main__":
    main()
```
