## Supplemental code for "Scoring respondents on a semantic frame of reference identifies a group that is burdened across many questionnaires and reaches care less easily": fig_component_ranks.py

```
#!/usr/bin/env python3
"""fig_component_ranks.py -- one line per component: how much it carries at each rank (main-text R5).

THE OBJECT. Every (participant, component) pair gives a (share, rank), where the RANK is that
component's position within THAT PARTICIPANT'S ordering -- rank 1 is the participant's largest
component, rank 2 their second, and so on. Fixing a component and a rank, we take every participant
for whom that component sits at that rank and summarize their shares. That is one point. Sweeping the
rank gives one line per component.

BOXES, NOT LINES. Ranks are three categories rather than a continuum, and no participant travels along
a line from rank 1 to rank 3, so connecting the medians would draw a trajectory nobody has. Each
component gets one box per rank, dodged within the rank, showing the median, the quartiles and
whiskers to 1.5 IQR. Points beyond the whiskers are NOT drawn and the y axis is scaled to the whiskers:
a handful of participants put almost all of their distance on one component, and plotting them stretches
the axis until every box is a sliver. The count omitted is printed by the run, never silently dropped. The shares at a fixed rank are skewed and bounded
below, which is also why a mean with a symmetric standard-deviation band was rejected: at rank 1 it
would reach below zero.

THE COMPONENTS NOT HIGHLIGHTED ARE POOLED into one grey box per rank rather than dropped, so the panel
still accounts for every component and a reader can see whether the highlighted ones sit apart from
the rest or among them.

WHAT IT SEPARATES. Every line falls with rank, since a rank-2 share is smaller than a rank-1 share by
construction. The differences BETWEEN components are the finding:

    at rank 1, a component that dominates whenever it is someone's largest sits high, and one that
    leads only as the largest of several comparable shares sits low
    at higher ranks, a component that still carries a real share when it is only a participant's
    fifth or tenth is a broad background, present and substantial well down the ordering

HOW MANY PARTICIPANTS EACH POINT RESTS ON varies -- a component may be rank 1 for seventy participants
and rank 30 for three. A point computed from fewer than MIN_N participants is not drawn, and the count
behind every point is written to the CSV, so a thin tail is never mistaken for a measured one.

ONE ENCODER, and this is not a shortcut. A documented construct is the same object under every
encoder, but a COMPONENT is not: each encoder fits its own basis, so its component 12 is not another's
and there is no correspondence to average over. Cross-encoder evidence belongs at the construct layer.

HIGHLIGHTING is ONE stated threshold, checkable against the picture, and --hl-by chooses which:

  --hl-by leads   [default] a component is highlighted when it is a participant's largest for at
                  least --hl-leads participants. This asks how much of the GROUP a component
                  accounts for, and by construction it can never admit content that leads a handful
                  of people however hard it carries them
  --hl-by r1      a component is highlighted when its RANK-1 median is at least --hl-median per
                  cent, over at least --min-n participants. This is the rule for the question "when
                  this content is what carries someone, how hard does it carry them?", and it is
                  the only one of the three that a component leading five people can pass
  --hl-by median  at least --hl-median per cent AT EVERY RANK WHERE IT IS MEASURED. KNOWN TO BE
                  UNSATISFIABLE at any selective level, and kept only so the reason is on record:
                  shares fall with rank by construction, so every component sits at 8-12% by rank 5
                  whatever it does at rank 1. A threshold high enough to separate the head is
                  impossible in the tail, and one reachable in the tail admits nearly everything.
                  On the 510 at 15% it admits ZERO of the 43

A rank with fewer than --min-n participants is not drawn and does not count for or against any of
the rules. An earlier version also required a measured median at EVERY shown rank, which silently
disqualified any component that is rarely a participant's largest -- interpersonal and sexual
violence among them -- however much it carried at the ranks where it was measured. That was a second
rule masquerading as eligibility, and it is gone.

THE RULES ANSWER DIFFERENT QUESTIONS and none is the right one by default. A lead count says what the
GROUP is made of and by construction can never admit content that leads a handful of people. A rank-1
median says what a component does FOR THE PEOPLE IT REACHES, which is the only way rare-but-dominant
content is visible at all -- on the 510, IPV leads five to seven participants and takes a rank-1
median of 22.6 to 23.2% of their distance, level with content that leads seventy. Whichever rule is
used, the run prints the components the others would have admitted, and the legend states the rule
in force.

--hl-max caps how many are drawn separately at the size of the palette. Any component that meets the
rule and does not fit is NAMED in the printout rather than silently pooled.

The run prints each component's smallest median across the shown ranks beside the threshold, so moving
the threshold is a judgment made against the numbers rather than against the picture.

Every other component is drawn as thin grey context, so all of them are present and none is dropped.
Components are labeled by their battery-side construct profile, in short forms checked one-to-one
against the documented labels.

Reads screen_out/attribution_component_shares_<tag>.csv (attribution.py) plus the basis, for the
profiles. Recomputes nothing and hardcodes no result.

--view chooses the panel. "ranks" [default] is the box figure described above. "leadshare" draws
one point per component instead: how many participants it is largest for against the median share it
takes there, with the interquartile range. That panel needs no threshold and pools nothing, and it
turns "rare content still carries hard" from a two-group contrast into a slope -- see
lead_share_panel for its labelling rule and why its x axis is log.

Out: screen_out/component_ranks_<enc>_<tag>.csv  (mean, sd and n per component per rank)
     screen_out/fig_component_ranks.png     --view ranks
     screen_out/fig_lead_vs_share.png       --view leadshare
     screen_out/fig_lead_count.png          --view leadcount
     ... copy into manuscript_nhb/figures/ BY HAND
Run: python3 fig_component_ranks.py [--enc gte-large-en-v1.5] [--tag outliers] [--max-rank 3]
                                    [--hl-by leads|r1|median] [--hl-leads 20] [--hl-median 20]
                                    [--hl-max 8] [--min-n 10] [--view ranks|leadshare|leadcount]
"""
import os
import re

import numpy as np
import pandas as pd
import matplotlib
matplotlib.use("Agg")
import matplotlib.pyplot as plt

from attribution import OUT_DIR, ID_COL
from attribution_components import battery_profiles, arg, DEFAULT_ENC

HERE = os.path.dirname(os.path.abspath(__file__))
CONSTRUCTS_CSV = os.path.join(HERE, "construct_map_151.csv")

MIN_N = 10            # a point needs this many participants at that rank, or it is not drawn
HL_LEADS = 20         # --hl-by leads: a participant's largest for at least this many participants
HL_MEDIAN = 15        # --hl-by median: median share (%) at every rank where it is measured
HL_MAX = 8            # at most this many drawn separately, the palette's size. Overflow is printed
LABEL_N = 4           # --view leadshare: labels per stated rule, see lead_share_panel
MIN_BOX = 5           # --view leadstrip: a column needs this many participants for a box
LABEL_CONSTRUCTS = 3  # --view leadstrip: constructs named per component on the x axis
RARE_LEADS = 10       # a component leading fewer than this is counted into the "rare content" tail
FIGW, FIGH = 6.5, 3.6

PALETTE = ["#D55E00", "#0072B2", "#009E73", "#CC79A7", "#E69F00", "#56B4E9", "#000000", "#8B4A9C"]
GREY = "#BBBBBB"

# Short display forms for the 30 documented constructs, so a legend entry can carry a whole profile.
# These shorten by dropping qualifying words, never letters. The map must be ONE-TO-ONE: "Perceived
# social support" must not become "social support" and "Open-mindedness" must not become "openness",
# both being documented constructs in their own right.
SHORT = {
    "Adult ADHD": "ADHD", "Agreeableness": "agreeableness",
    "Body-focused repetitive behavior": "BFRB", "Childhood adversity": "childhood adversity",
    "Conscientiousness": "conscientiousness", "Daily spiritual experience": "spiritual experience",
    "Depression": "depression", "Everyday discrimination": "everyday discrim.",
    "Extraversion": "extraversion", "Food insecurity": "food insecurity",
    "Generalized anxiety": "anxiety", "Healthcare discrimination": "healthcare discrim.",
    "Housing instability": "housing instability", "Interpersonal & sexual violence": "IPV",
    "Life-threatening events": "life-threatening events", "Loneliness": "loneliness",
    "Mania": "mania", "Negative emotionality": "neg. emotionality",
    "Neighborhood disorder & safety": "neighborhood safety",
    "Neighborhood social cohesion": "neighborhood cohesion", "Open-mindedness": "open-mindedness",
    "Panic & phobia": "panic & phobia", "Perceived social support": "perceived support",
    "Perceived stress": "perceived stress", "Psychosis": "psychosis",
    "Religious participation": "religious participation", "Self-rated health": "self-rated health",
    "Subjective wellbeing": "wellbeing", "Substance use": "substance use",
    "Suicidality & self-harm": "suicidality",
}


def check_short_map():
    documented = set(pd.read_csv(CONSTRUCTS_CSV)["construct"].dropna().astype(str))
    missing = sorted(documented - set(SHORT))
    assert not missing, f"no short form for documented constructs: {missing}"
    seen = {}
    for k, v in SHORT.items():
        assert v not in seen, f"short form {v!r} is shared by {seen[v]!r} and {k!r}"
        seen[v] = k


def label(profile, max_constructs=3):
    p = str(profile).strip()
    if not p or p.startswith("("):
        return "no battery core"
    names = [SHORT[x.split(":")[0].strip()] for x in p.split(";") if x.strip()]
    extra = len(names) - max_constructs
    return ", ".join(names[:max_constructs]) + (f", +{extra}" if extra > 0 else "")


def lead_strip_panel(S, R, comps, prof, enc, tag, n_people):
    """Every outlier once, at the component that carries the most of their distance.

    THE OBJECT. For each of the participants, take the component with their largest share and plot
    the pair (that component, that share). Each participant therefore contributes exactly one point,
    so the columns partition the group: a column's height in points is how many people that content
    leads, and the spread within it is how hard it leads them. The lead-count panel shows the first
    of those and the leadshare panel shows a summary of the second. This shows both, and the shape
    of the second.

    GROUPED BY HOW MANY CONSTRUCTS CORE THE COMPONENT, because the question the grouping asks is
    whether content resting on one documented construct behaves differently from content resting on
    several. WITHIN a group the columns run by decreasing median leading share, so the descent is
    readable along each band and two components of the same profile size sit side by side in the
    order that matters. Shaded bands separate the groups and are labelled above the panel.

    A BOX NEEDS MIN_BOX PARTICIPANTS. Quartiles over three people describe those three and nothing
    else, so a column below the threshold is drawn as its points alone. The threshold is stated on
    the panel rather than left for a reader to infer from box widths.

    POINTS ARE JITTERED horizontally within the column and drawn small and semi-transparent, since
    the busiest columns hold seventy of them. Jitter is deterministic, seeded from the component
    index, so the figure is identical on every run.
    """
    lead_j = np.argmax(S, axis=1)                       # each participant's largest component
    lead_s = S[np.arange(S.shape[0]), lead_j]
    nconstr = {}
    for j, c in enumerate(comps):
        p = str(prof.get(int(c), "")).strip()
        nconstr[j] = 0 if (not p or p.startswith("(")) else len([x for x in p.split(";") if x.strip()])
    counts = {j: int((lead_j == j).sum()) for j in range(len(comps))}
    # within a group, DECREASING MEDIAN leading share, so a column's height on the page reads
    # directly against its neighbours of the same profile size. A component that leads nobody has
    # no median and sorts last in its group.
    medlead = {j: (float(np.median(lead_s[lead_j == j])) if counts[j] else -np.inf)
               for j in range(len(comps))}
    order = sorted(range(len(comps)), key=lambda j: (nconstr[j], -medlead[j]))

    print(f"\n  every outlier at its leading component, {enc}, tag={tag}")
    print(f"    {len(lead_s)} participants, {sum(1 for j in order if counts[j])} components lead at "
          f"least one, ordered by construct-profile size")
    for g in sorted(set(nconstr.values())):
        js=[j for j in order if nconstr[j]==g]
        led=sum(counts[j] for j in js)
        v=lead_s[np.isin(lead_j,js)]
        print(f"    {g} construct(s): {len(js):2d} components, {led:3d} participants "
              f"({100*led/n_people:.0f}%), median leading share {np.median(v):.1f}%" if len(v)
              else f"    {g} construct(s): {len(js):2d} components, 0 participants")

    fig, ax = plt.subplots(figsize=(FIGW, 6.6), facecolor="white")
    # shaded bands, one per construct-profile size
    bounds, prev = [], None
    for pos, j in enumerate(order):
        if nconstr[j] != prev: bounds.append([pos, pos, nconstr[j]]); prev = nconstr[j]
        bounds[-1][1] = pos
    # BAND MARKERS GO ABOVE THE PANEL. Below it belongs to the component names, and a marker there
    # would sit among forty-three rotated labels rather than over the band it names.
    for i, (a, b, g) in enumerate(bounds):
        if i % 2 == 0:
            ax.axvspan(a - 0.5, b + 0.5, color="#F2F2F2", lw=0, zorder=0)
        # ONLY THE FIRST band carries the word. Sizing the word to the band width does not work --
        # two adjacent three-column bands both clear any such test and still collide, which is what
        # "4 constructs 5 constructs" did. The first band names the unit and the rest are numerals,
        # which fit in one column however many bands the basis turns out to have.
        txt = f"{g} construct" + ("s" if g != 1 else "") if i == 0 else f"{g}"
        ax.annotate(txt, xy=((a + b) / 2, 1.015), xycoords=("data", "axes fraction"),
                    ha="center", va="bottom", fontsize=6.8, color="#444444",
                    annotation_clip=False)

    # Y SCALED TO THE WHISKERS, as the ranks view does. A few participants put more than half their
    # distance on one component, and drawing them stretches the axis until every box is a sliver in
    # the bottom third of the panel. The points above the limit are NOT drawn and the run prints how
    # many, so nothing is dropped silently.
    top = 0.0
    for j in range(len(comps)):
        v = lead_s[lead_j == j]
        if len(v) >= MIN_BOX:
            q1v, q3v = np.percentile(v, [25, 75])
            top = max(top, q3v + 1.5 * (q3v - q1v))
        elif len(v):
            top = max(top, v.max())
    hidden = int((lead_s > top).sum())

    rng = np.random.RandomState(0)
    for pos, j in enumerate(order):
        v = lead_s[lead_j == j]
        if not len(v): continue
        jit = (rng.rand(len(v)) - 0.5) * 0.62
        ax.scatter(pos + jit, v, s=6.5, color="#0072B2", alpha=0.55, lw=0, zorder=3)
        if len(v) >= MIN_BOX:
            bp = ax.boxplot([v], positions=[pos], widths=0.66, patch_artist=True, showfliers=False,
                            zorder=4, medianprops=dict(color="#D55E00", lw=1.2),
                            whiskerprops=dict(color="#555555", lw=0.7),
                            capprops=dict(color="#555555", lw=0.7),
                            boxprops=dict(facecolor="none", edgecolor="#555555", lw=0.7))
    ax.set_xlim(-0.8, len(order) - 0.2)
    ax.set_ylim(top=top * 1.02)
    print(f"    y axis scaled to the whiskers: {hidden} of {len(lead_s)} points "
          f"({100*hidden/len(lead_s):.1f}%) lie above {top:.0f}% and are not drawn")
    # EVERY component is named on the axis, by up to LABEL_CONSTRUCTS constructs of its profile,
    # comma-joined on ONE line. One line matters: a rotated label runs vertically, so a second line
    # sits BESIDE the first and doubles the column's width, which collides at a sixth of an inch per
    # column. Length instead grows downward, which the bottom margin absorbs. The trailing "+N" is
    # dropped because the band the component sits in already states how many constructs core it.
    ax.set_xticks(range(len(order)))
    ax.set_xticklabels([re.sub(r",\s*\+\d+$", "",
                               label(prof.get(int(comps[j]), ""), max_constructs=LABEL_CONSTRUCTS))
                        for j in order], rotation=90, fontsize=5.2, color="#333333")
    ax.set_ylabel("share of that participant's distance\ncarried by their largest component (%)")
    ax.yaxis.label.set_size(8.5)
    ax.tick_params(axis="y", labelsize=8)
    ax.tick_params(axis="x", length=0, pad=2)
    ax.spines[["top", "right", "bottom"]].set_visible(False)
    ax.grid(axis="y", color="#E8E8E8", lw=0.5, zorder=1); ax.set_axisbelow(False)
    fig.tight_layout()
    png = os.path.join(OUT_DIR, "fig_lead_strip.png")
    fig.savefig(png, dpi=400, facecolor="white")
    print(f"\nwrote -> {png}")


def lead_count_panel(leads, prof, enc, tag, n_people):
    """How many participants each component is the largest share for. One bar per component.

    THE SHAPE IS THE FINDING. A few components lead dozens of people and most lead a handful, so
    what a participant is carried by is common content for about half the group and near-unique
    content for a quarter of it. Sorting by the count and drawing every component says that in one
    picture, without a threshold deciding anything.

    THREE TONES, TWO STATED CUTS. Components leading at least HL_LEADS people, those leading fewer
    than RARE_LEADS, and the ones between. The cuts are the ones main-text R5 already reports, and
    the caption line gives how many PARTICIPANTS each band accounts for, which is the number the
    text uses -- a band's share of the group is not its share of the components.

    LABELS go to the components that lead the most, by count, and stop where the bars become too
    narrow to carry text. The tail is deliberately unlabelled: that it is a tail of many small
    components is the point, and naming twenty-nine of them would say less, not more.

    EVERY component that leads at least one participant is drawn. Components leading nobody are
    counted in the printout and omitted from the axis, since a zero-height bar is not visible.
    """
    s = leads[leads > 0].sort_values(ascending=False)
    n_zero = int((leads == 0).sum())
    x = np.arange(len(s))
    v = s.values.astype(int)

    common = v >= HL_LEADS
    rare = v < RARE_LEADS
    mid = ~common & ~rare
    print(f"\n  how many participants each component leads, {enc}, tag={tag}")
    for m, name in ((common, f"lead {HL_LEADS}+"), (mid, f"lead {RARE_LEADS}-{HL_LEADS-1}"),
                    (rare, f"lead 1-{RARE_LEADS-1}")):
        print(f"    {int(m.sum()):2d} components {name:16s} account for {int(v[m].sum()):3d} "
              f"participants ({100*v[m].sum()/n_people:.0f}%)")
    print(f"    {n_zero} components lead nobody and are not drawn")

    fig, ax = plt.subplots(figsize=(FIGW, 4.0), facecolor="white")
    colr = np.where(common, "#0072B2", np.where(rare, "#C9C9C9", "#7FB2D5"))
    ax.bar(x, v, color=colr, width=0.82, linewidth=0)
    ax.set_xlim(-0.9, len(x) - 0.1)
    ax.set_ylabel("participants it is the largest share for")
    ax.set_xlabel(f"the {len(x)} components, ordered by that count")
    ax.yaxis.label.set_size(8.5)
    ax.xaxis.label.set_size(8.5)
    ax.set_xticks([])
    ax.tick_params(axis="y", labelsize=8)
    ax.spines[["top", "right", "bottom"]].set_visible(False)
    ax.grid(axis="y", color="#E8E8E8", lw=0.5, zorder=0)
    ax.set_axisbelow(True)

    # ONE construct per label, never wrapped and never truncated mid-word. A rotated label runs
    # vertically, so a second line sits BESIDE the first and doubles the label's width -- at roughly
    # a sixth of an inch per bar, two-line labels from neighbouring bars overlap. The leading
    # construct alone is short, one line, and distinct across the six named here.
    # LABEL BOTH DRAWN BANDS, not a fixed count: every component that is the largest for at least
    # RARE_LEADS people is named, which is exactly the two coloured bands. The grey tail is left
    # unlabelled because being a tail of many small components is what it is there to show, and
    # twenty-nine names would bury that.
    for i in np.flatnonzero(~rare):
        ax.text(x[i], v[i] + max(v) * 0.03, label(prof.get(int(s.index[i]), ""), max_constructs=1),
                rotation=90, ha="center", va="bottom", fontsize=6.4, color="#222222")

    band = (f"{int(common.sum())} components are the largest for {HL_LEADS}+ people, "
            f"{100*v[common].sum()/n_people:.0f}% of the group\n"
            f"{int(rare.sum())} are the largest for fewer than {RARE_LEADS}, "
            f"{100*v[rare].sum()/n_people:.0f}% of the group")
    ax.annotate(band, xy=(0.985, 0.95), xycoords="axes fraction", ha="right", va="top",
                fontsize=7, color="#444444")
    ax.set_ylim(0, max(v) * 1.55)                 # headroom for the rotated labels

    fig.tight_layout()
    png = os.path.join(OUT_DIR, "fig_lead_count.png")
    fig.savefig(png, dpi=400, facecolor="white")
    print(f"\nwrote -> {png}")


def lead_share_panel(med, q1, q3, cnt, leads, prof, enc, tag, min_n):
    """How many people a component leads, against how hard it carries them when it does.

    One point per component, all of them, no threshold and nothing pooled. The horizontal axis is
    how many of the group the component is largest for and the vertical axis its median share at
    rank 1, with the interquartile range across those same people. The question the panel answers is
    whether the two are related at all: if the cloud is flat, content that leads a handful carries
    them as hard as content that leads seventy, and being rare is not being weak.

    THE HORIZONTAL AXIS IS LOG because the counts run from 1 to about 70 and most components sit
    below ten. On a linear axis the whole rare tail -- the side the question is about -- collapses
    into the left margin.

    A COMPONENT LEADING FEWER THAN min_n PEOPLE IS STILL DRAWN, in pale grey, because dropping it
    would remove exactly the cases at issue. Its median rests on that many people and no more, which
    the position on the x axis already states, and the pale colour marks it as thin rather than
    absent.

    LABELS follow two stated rules, declared here so the selection cannot be tuned: the LABEL_N
    components leading the most people, and the LABEL_N with the highest rank-1 median among those
    leading fewer than the median number. The second rule is the hypothesis -- rare but dominant --
    written as a selection rather than applied to the picture after the fact.
    """
    from scipy import stats as _st                       # only this panel needs it

    comps = list(med.index)
    x = leads.reindex(comps).astype(float).values
    y = med[1].reindex(comps).astype(float).values
    lo = np.clip(y - q1[1].reindex(comps).values, 0, None)
    hi = np.clip(q3[1].reindex(comps).values - y, 0, None)
    ok = np.isfinite(x) & np.isfinite(y) & (x > 0)
    thin = ok & (x < min_n)

    rho, p = _st.spearmanr(x[ok], y[ok])
    print(f"\n  how many a component leads against how hard it leads them, over the "
          f"{int(ok.sum())} components that lead anyone:")
    print(f"    Spearman rho = {rho:+.3f} (p = {p:.3g}). A rho near zero is rare content carrying "
          f"its people as hard as common content carries anyone")
    med_leads = float(np.median(x[ok]))
    rare, common = ok & (x < med_leads), ok & (x >= med_leads)
    print(f"    median rank-1 share {np.median(y[rare]):.1f}% for the {int(rare.sum())} leading "
          f"fewer than {med_leads:.0f} people, {np.median(y[common]):.1f}% for the "
          f"{int(common.sum())} leading more")

    idx = np.arange(len(comps))
    by_x = list(idx[ok][np.argsort(-x[ok])][:LABEL_N])
    rare_i = idx[rare]
    by_y = list(rare_i[np.argsort(-y[rare])][:LABEL_N])
    lab = list(dict.fromkeys(by_x + by_y))

    fig, ax = plt.subplots(figsize=(FIGW, 4.0), facecolor="white")
    for sel, colr, alpha, ms in ((ok & ~thin, "#0072B2", 0.85, 5.0),
                                 (thin, "#9A9A9A", 0.75, 4.0)):
        if sel.any():
            ax.errorbar(x[sel], y[sel], yerr=np.vstack([lo[sel], hi[sel]]), fmt="o", ms=ms,
                        color=colr, ecolor=colr, elinewidth=0.9, capsize=0, lw=0, alpha=alpha,
                        markeredgecolor="white", markeredgewidth=0.5, zorder=3)
    ax.set_xscale("log")
    ax.set_xlabel("participants the component is largest for")
    ax.set_ylabel("share of their distance it carries there (%),\nmedian and interquartile range")
    ax.xaxis.label.set_size(8.5)
    ax.yaxis.label.set_size(8.5)
    ax.tick_params(labelsize=8)
    ax.spines[["top", "right"]].set_visible(False)
    ax.grid(color="#E4E4E4", lw=0.5, zorder=0)
    ax.set_axisbelow(True)

    for i in lab:
        ax.annotate(label(prof.get(int(comps[i]), ""))[:34], xy=(x[i], y[i]),
                    xytext=(4, 5), textcoords="offset points", fontsize=6.4, color="#222222",
                    ha="left", va="bottom", zorder=5)
    ax.annotate(f"pale: leads fewer than {min_n}, so the median rests on that many people",
                xy=(0.015, 0.03), xycoords="axes fraction", fontsize=6.4, color="#9A9A9A")

    fig.tight_layout()
    png = os.path.join(OUT_DIR, "fig_lead_vs_share.png")
    fig.savefig(png, dpi=400, facecolor="white")
    print(f"\nwrote -> {png}")


def main():
    enc = arg("--enc", DEFAULT_ENC)
    tag = arg("--tag", "outliers")
    max_rank = int(arg("--max-rank", "3"))
    hl_leads = int(arg("--hl-leads", str(HL_LEADS)))
    hl_by = arg("--hl-by", "leads")
    hl_median = float(arg("--hl-median", str(HL_MEDIAN)))
    hl_max = int(arg("--hl-max", str(HL_MAX)))
    view = arg("--view", "ranks")
    MIN_N = int(arg("--min-n", str(globals()["MIN_N"])))
    assert hl_by in ("leads", "median", "r1"), "--hl-by leads|median|r1"
    assert view in ("ranks", "leadshare", "leadcount", "leadstrip"), "--view ranks|leadshare|leadcount|leadstrip"
    check_short_map()

    D = pd.read_csv(os.path.join(OUT_DIR, f"attribution_component_shares_{tag}.csv"))
    D[ID_COL] = D[ID_COL].astype(str)
    D = D[D["encoder"] == enc].copy()
    assert len(D), f"no rows for {enc}"
    prof, _ = battery_profiles(enc)

    piv = D.pivot_table(index=ID_COL, columns="component", values="share").fillna(0.0)
    comps = np.array(piv.columns)
    S = 100 * piv.values
    n, k = S.shape

    # rank of each component WITHIN each participant, 1 = that participant's largest share
    order = np.argsort(-S, axis=1)
    R = np.empty_like(order)
    np.put_along_axis(R, order, np.arange(1, k + 1)[None, :].repeat(n, axis=0), axis=1)

    ranks = np.arange(1, min(max_rank, k) + 1)
    med = pd.DataFrame(index=comps, columns=ranks, dtype=float)
    q1 = pd.DataFrame(index=comps, columns=ranks, dtype=float)
    q3 = pd.DataFrame(index=comps, columns=ranks, dtype=float)
    cnt = pd.DataFrame(index=comps, columns=ranks, dtype=int)
    for j, c in enumerate(comps):
        for r in ranks:
            v = S[R[:, j] == r, j]
            cnt.loc[c, r] = len(v)
            if len(v):
                med.loc[c, r] = np.median(v)
                q1.loc[c, r], q3.loc[c, r] = np.percentile(v, 25), np.percentile(v, 75)
    drawn = med.where(cnt >= MIN_N)

    mass_pct = pd.Series(100 * np.clip(S, 0, None).sum(axis=0) / np.clip(S, 0, None).sum(),
                         index=comps)
    meas = med.where(cnt >= MIN_N)                       # medians only where they are measured
    leads = cnt[1]                                       # participants this component is largest for
    if hl_by == "r1":
        # Shares fall with rank by construction, so a threshold held at EVERY rank is either
        # impossible at rank 5 or trivial at rank 1 -- the "median" rule below admits nothing at any
        # useful level. Rank 1 is where components differ at all (S25: they converge to within two
        # points by rank 3), and it is the rank the question is about: when this content is what
        # carries someone, how hard does it carry them?
        r1 = med[1].where(cnt[1] >= MIN_N)
        hl = list(r1[r1 >= hl_median].sort_values(ascending=False).index)
        if len(hl) > hl_max:
            drop = hl[hl_max:]
            print(f"  {len(drop)} components reach {hl_median:.0f}% at rank 1 but are pooled, over "
                  f"the --hl-max {hl_max} the palette holds: "
                  + ", ".join(f"C{int(c)}" for c in drop))
            hl = hl[:hl_max]
    elif hl_by == "median":
        # the rule this script's header states: at least hl_median per cent at EVERY rank where the
        # component is measured. A rank with too few participants is unmeasured and counts neither
        # way, so content that is rarely anyone's largest -- IPV among it -- can still qualify on
        # the ranks it does reach, which a lead count forecloses.
        seen = meas.notna()
        ok = (meas.ge(hl_median) | ~seen).all(axis=1) & seen.any(axis=1)
        hl = list(med[1].where(ok).dropna().sort_values(ascending=False).index)
        hl += [c for c in ok[ok].index if c not in hl]   # qualified, but unmeasured at rank 1
        if len(hl) > hl_max:
            drop = hl[hl_max:]
            print(f"  {len(drop)} components meet the {hl_median:.0f}% rule but are pooled, over "
                  f"the --hl-max {hl_max} the palette holds: "
                  + ", ".join(f"C{int(c)}" for c in drop))
            hl = hl[:hl_max]
    else:
        hl = list(leads[leads >= hl_leads].sort_values(ascending=False).index)
    rule = {"median": f"median at least {hl_median:.0f}% at every measured rank",
            "r1": f"rank-1 median at least {hl_median:.0f}%",
            "leads": f"a participant's largest for at least {hl_leads} participants"}[hl_by]
    # what the rule NOT in force would have admitted, so the choice is visible rather than assumed
    seen = meas.notna()
    by_med = set(((meas.ge(hl_median) | ~seen).all(axis=1) & seen.any(axis=1)).pipe(
        lambda s: s[s].index))
    by_lead = set(leads[leads >= hl_leads].index)
    other = (by_lead if hl_by == "median" else by_med) - set(hl)
    if other:
        print(f"\n  the other rule would also have drawn "
              + ", ".join(f"C{int(c)} ({label(prof.get(int(c), ''))[:28]}, leads "
                          f"{int(leads[c])}, r1 median "
                          f"{med.loc[c, 1]:.1f}%)" for c in sorted(other)))

    T = pd.DataFrame({"component": comps,
                      "battery_profile": [prof.get(int(c), "") for c in comps],
                      "mass_pct": mass_pct.values})
    for r in ranks:
        T[f"median_r{r}"] = med[r].values
        T[f"q1_r{r}"] = q1[r].values
        T[f"q3_r{r}"] = q3[r].values
        T[f"n_r{r}"] = cnt[r].values
    T = T.set_index("component").sort_values("median_r1", ascending=False)

    print(f"{n} participants, {k} components, {enc}, ranks 1-{ranks[-1]}, min n {MIN_N}")
    print(f"\n{'comp':>5s} {'mass':>6s} " + " ".join(f"{'r'+str(r):>20s}" for r in ranks[:5])
          + "  profile")
    for c, r_ in T.iterrows():
        cells = " ".join(f"{r_['median_r'+str(r)]:5.1f}[{r_['q1_r'+str(r)]:4.1f}-{r_['q3_r'+str(r)]:<4.1f}]"
                         f"({int(r_['n_r'+str(r)]):3d})"
                         if r_[f'n_r{r}'] else f"{'--':>20s}" for r in ranks[:5])
        print(f"C{int(c):<4d} {r_['mass_pct']:5.1f}% {cells}  {label(r_['battery_profile'])[:40]}")
    print(f"\nwho leads whom, and how hard (highlighting: {rule}):")
    for c in leads.sort_values(ascending=False).index:
        if leads[c] == 0:
            continue
        m = f"{med.loc[c, 1]:5.1f}%" if cnt.loc[c, 1] >= MIN_N else f"{med.loc[c, 1]:5.1f}%*"
        print(f" {'*' if c in hl else ' '}C{int(c):<4d} leads {int(leads[c]):3d}  median {m}  "
              f"{label(prof.get(int(c), ''))[:44]}")
    print(f"  ({len(hl)} highlighted of {len(comps)}; * marks a median over fewer than {MIN_N} "
          f"participants, printed but not drawn)")

    # THE TAIL. A participant led by a component that leads almost nobody else is still led at a
    # comparable share -- rare content, not diffuse content. Counted exactly rather than summed off
    # the table by eye, and the two medians are printed side by side rather than characterized.
    rare = leads[(leads > 0) & (leads < RARE_LEADS)]
    rare_ppl = int(rare.sum())
    led_by_hl = int(leads[hl].sum())
    rare_med = float(np.median([med.loc[c, 1] for c in rare.index if pd.notna(med.loc[c, 1])]))
    hl_vals = [med.loc[c, 1] for c in hl if pd.notna(med.loc[c, 1])]
    hl_med = float(np.median(hl_vals)) if hl_vals else float("nan")
    print(f"\n  {led_by_hl} participants ({100*led_by_hl/n:.0f}%) are led by one of the {len(hl)} "
          f"components leading {hl_leads}+ people")
    print(f"  {rare_ppl} ({100*rare_ppl/n:.0f}%) are led by one of the {len(rare)} components that "
          f"lead fewer than {RARE_LEADS} people each,")
    print(f"  median rank-1 share {rare_med:.1f}% for those components against {hl_med:.1f}% for "
          f"the {len(hl)}")

    if view == "leadstrip":
        lead_strip_panel(S, R, comps, prof, enc, tag, n)
        return
    if view == "leadcount":
        lead_count_panel(leads, prof, enc, tag, n)
        return
    if view == "leadshare":
        lead_share_panel(med, q1, q3, cnt, leads, prof, enc, tag, MIN_N)
        return

    fig, ax = plt.subplots(figsize=(FIGW, FIGH), facecolor="white")
    others = [c for c in comps if c not in hl]
    groups = [(c, PALETTE[i % len(PALETTE)], label(prof.get(int(c), ""))) for i, c in enumerate(hl)]
    groups.append((None, GREY, f"the other {len(others)} components, pooled"))

    width = 0.72 / len(groups)
    skipped, top_whisker, n_hidden, n_total = 0, 0.0, 0, 0
    for gi, (c, colr, _) in enumerate(groups):
        dx = (gi - (len(groups) - 1) / 2) * width
        for r in ranks:
            if c is None:
                v = np.concatenate([S[R[:, list(comps).index(o)] == r, list(comps).index(o)]
                                    for o in others])
            else:
                j = list(comps).index(c)
                v = S[R[:, j] == r, j]
                if len(v) < MIN_N:            # too few participants at that rank to summarize
                    skipped += 1
                    continue
            bp = ax.boxplot([v], positions=[r + dx], widths=width * 0.86, patch_artist=True,
                            showcaps=False, showfliers=False, manage_ticks=False,
                            medianprops=dict(color="white", lw=1.0),
                            boxprops=dict(facecolor=colr, edgecolor=colr, lw=0.6, alpha=0.75),
                            whiskerprops=dict(color=colr, lw=0.7))
            hi = max(w.get_ydata()[1] for w in bp["whiskers"])
            top_whisker = max(top_whisker, float(hi))
            n_hidden += int((v > hi).sum())
            n_total += len(v)

    handles = [plt.Line2D([], [], marker="s", ls="none", ms=5, mfc=colr, mec="none", label=lb)
               for _, colr, lb in groups]
    ax.set_xticks(list(ranks))
    ax.set_xticklabels([str(r) for r in ranks])
    ax.set_xlim(ranks[0] - 0.55, ranks[-1] + 0.55)
    ax.set_ylim(0, top_whisker * 1.06)
    ax.set_xlabel("rank of the component within the participant  (1 = their largest)", fontsize=8)
    ax.set_ylabel("share of that participant's\nsquared distance (%)", fontsize=8)
    ax.tick_params(labelsize=7.5)
    ax.grid(True, axis="y", color="#EDEDED", lw=0.6, zorder=0)
    ax.set_axisbelow(True)
    ax.spines[["top", "right"]].set_visible(False)
    for sp in ax.spines.values():
        sp.set_color("#9A9A9A")
    ax.legend(handles=handles, loc="upper center", bbox_to_anchor=(0.5, -0.19), ncol=2,
              frameon=False, fontsize=6.6, handlelength=1.0, columnspacing=1.4,
              title=f"highlighted: {rule}",
              title_fontsize=7.0)
    if skipped:
        print(f"  {skipped} component-rank boxes not drawn, fewer than {MIN_N} participants there")
    print(f"  y axis scaled to the whiskers: {n_hidden} of {n_total} plotted values "
          f"({100*n_hidden/max(n_total,1):.1f}%) lie above and are not drawn")

    fig.subplots_adjust(left=0.115, right=0.985, top=0.955, bottom=0.40)
    # the rule and the rank span are in the name: the published figure is the --hl-by leads default,
    # and an exploratory run must not overwrite it
    stem = ("fig_component_ranks" if (hl_by == "leads" and max_rank == 3)
            else f"fig_component_ranks_{hl_by}_r{max_rank}")
    png = os.path.join(OUT_DIR, f"{stem}.png")
    fig.savefig(png, dpi=400, facecolor="white")
    T.to_csv(os.path.join(OUT_DIR, f"component_ranks_{enc}_{tag}.csv"))
    print(f"\nwrote -> {png}\n         {OUT_DIR}/component_ranks_{enc}_{tag}.csv")


if __name__ == "__main__":
    main()
```
