## Supplemental code for "Scoring respondents on a semantic frame of reference identifies a group that is burdened across many questionnaires and reaches care less easily": fig_construct_ranks.py

```
#!/usr/bin/env python3
"""fig_construct_ranks.py -- each construct's whole per-person share distribution, as a rank curve.

Replaces the discrete levels (n_ge10 / n_ge20 / n_ge30) and the participation-ratio summary. Both
reduced a construct to one number and lost the tail, which is where the question lives: does a
construct matter a little to many people, or a lot to a few?

THE CURVE. For construct c, sort its per-person shares of T^2 in descending order and plot rank
against share. A construct that carries a few people hard starts high at ranks 1-10 and falls off a
cliff. A broad background starts low and decays slowly across hundreds of ranks. The crossings
between curves are the finding, and no threshold appears anywhere.

  x  rank within the construct, 1 .. n, LOG scale. Log is what makes the head visible: on a linear
     axis the first ten ranks occupy under two per cent of the width.
  y  share of that participant's squared distance, per cent.

ENCODER HANDLING. Sorting happens WITHIN each encoder, and the six curves are combined RANK-WISE --
the plotted line is the median across encoders at each rank, with a min-max band for the highlighted
constructs. Averaging shares per person BEFORE sorting would smooth the distribution and pull the
head down, the same bias that showed up in construct_concentration.py, where the pooled intensity
fell below every per-encoder value for eight constructs.

HIGHLIGHTING is by rule, not by hand, so the choice is reproducible. Four clauses, unioned:

  top HL_LEAD  by participants led          the constructs that carry the group
  top HL_HEAD  by head height (ranks 1-5)   tall, whether or not they lead anyone
  top HL_STEEP by r5/r250 ratio             CONCENTRATED -- a lot to a few
  top HL_FLAT  by the inverse of that       BROAD BACKGROUND -- a little to many, the counter-example

The last two are the shape clauses and they are the point of the figure. Height alone does not
separate a construct that carries a few people hard from one that carries many people moderately, and
leadership alone misses a construct that is decisive for a handful and never wins. Both shape clauses
are restricted to constructs holding at least MIN_MASS_PCT of total mass, so the ranking is not won by
something with no presence in the group at all. Everything else is drawn as thin grey context, so all
thirty constructs are present and none is silently dropped.

LEADERSHIP RULE. `n_leads` is the argmax of the ENCODER-AVERAGED construct shares, which is what
outlier_profile.py:54-55 uses and therefore what sec:group publishes (psychosis 138, food insecurity
107, everyday discrimination 76, neighborhood 52, depression 49, healthcare discrimination 21).
attribution.py:186-195 uses a DIFFERENT rule for its own per-person table -- per-encoder argmax, then
the modal answer across encoders -- which gives 137/100/83/51/51/24 on the same data. Do not mix
them, and in particular do not quote attribution.py's ">= 4 of 6" percentage as the published 81%:
the modal leader maximises encoder agreement by construction, so that number is not comparable.

Reads screen_out/attribution_item_shares_<tag>.csv, written by attribution.py. Recomputes nothing.

  python3 fig_construct_ranks.py
  python3 fig_construct_ranks.py --tag invisible

Out: screen_out/construct_ranks_<tag>.csv     shares at selected ranks, plus the leadership columns
     screen_out/fig_construct_ranks_<tag>.png
     Copy the PNG into manuscript_nhb/figures/ BY HAND -- nothing syncs it.
"""
import os
import sys

import numpy as np
import pandas as pd
import matplotlib
matplotlib.use("Agg")
import matplotlib.pyplot as plt

HERE = os.path.dirname(os.path.abspath(__file__))
OUT_DIR = os.path.join(HERE, "screen_out")
CONSTRUCTS_CSV = os.path.join(HERE, "construct_map_151.csv")
ID_COL = "id"

HL_LEAD, HL_HEAD = 4, 3          # highlight: top-N by leadership, top-N by head height
HL_STEEP, HL_FLAT = 2, 1         # ... plus the shape clauses, concentrated and broad
HEAD_RANKS = 5                   # "head" = mean share over ranks 1..5
STEEP_NUM, STEEP_DEN = 5, 250    # steepness = share at rank 5 / share at rank 250
MIN_MASS_PCT = 2.0               # shape clauses only consider constructs holding at least this much
REPORT_RANKS = (1, 5, 10, 25, 50, 100, 250)
FIGW, FIGH = 6.5, 4.4            # print size, per the project's figure rule

# Okabe-Ito, yellow omitted (illegible as a thin line)
PALETTE = ["#D55E00", "#0072B2", "#009E73", "#CC79A7", "#E69F00", "#56B4E9", "#000000"]
GREY = "#BBBBBB"


def main():
    tag = "outliers"
    if "--tag" in sys.argv:
        tag = sys.argv[sys.argv.index("--tag") + 1]

    path = os.path.join(OUT_DIR, f"attribution_item_shares_{tag}.csv")
    assert os.path.exists(path), f"{path} not found -- run attribution.py first"
    sh = pd.read_csv(path, index_col=[0, 1])
    sh.index = sh.index.set_names(["encoder", ID_COL])
    sh.index = sh.index.set_levels(sh.index.levels[1].astype(str), level=1)

    cmap = pd.read_csv(CONSTRUCTS_CSV).set_index("item")["construct"]
    con = cmap.reindex(sh.columns).fillna("unlabelled").values
    C = sh.T.groupby(con).sum().T                        # (encoder, id) x construct
    encoders = list(C.index.get_level_values("encoder").unique())
    constructs = list(C.columns)
    n = C.xs(encoders[0], level="encoder").shape[0]

    # rank curves: sort within each encoder, then combine rank-wise
    curves, lo, hi = {}, {}, {}
    for c in constructs:
        S = np.array([np.sort(C.xs(e, level="encoder")[c].values)[::-1] for e in encoders])
        curves[c] = np.median(S, axis=0)
        lo[c], hi[c] = S.min(axis=0), S.max(axis=0)

    # leadership, by outlier_profile.py's rule (argmax of encoder-averaged shares) -- the one the
    # published counts come from. See the LEADERSHIP RULE note above before changing this.
    lead = C.groupby(level=ID_COL).mean().idxmax(axis=1)
    n_leads = lead.value_counts().reindex(constructs).fillna(0).astype(int)

    head = pd.Series({c: curves[c][:HEAD_RANKS].mean() for c in constructs})
    mass = pd.Series({c: np.clip(curves[c], 0, None).sum() for c in constructs})
    mass_pct = 100 * mass / mass.sum()
    den = min(STEEP_DEN, n)
    steep = pd.Series({c: (curves[c][STEEP_NUM - 1] / curves[c][den - 1]
                           if curves[c][den - 1] > 0 else np.inf) for c in constructs})
    eligible = steep[mass_pct >= MIN_MASS_PCT].replace(np.inf, np.nan).dropna()

    by_lead = list(n_leads.sort_values(ascending=False).index[:HL_LEAD])
    by_head = list(head.sort_values(ascending=False).index[:HL_HEAD])
    by_steep = list(eligible.sort_values(ascending=False).index[:HL_STEEP])
    by_flat = list(eligible.sort_values(ascending=True).index[:HL_FLAT])
    hl = list(dict.fromkeys(by_lead + by_head + by_steep + by_flat))

    # ---------------------------------------------------------------- table
    rows = []
    for c in constructs:
        r = {"construct": c, "n_leads": int(n_leads[c]),
             "head_r1_5": 100 * head[c],
             "steepness_r5_r250": float(steep[c]),
             "total_mass_pct": float(mass_pct[c])}
        for k in REPORT_RANKS:
            r[f"r{k}"] = 100 * curves[c][k - 1] if k <= n else np.nan
        rows.append(r)
    T = pd.DataFrame(rows).set_index("construct").sort_values("head_r1_5", ascending=False)

    print(f"{n} respondents, {len(encoders)} encoders, {len(constructs)} constructs")
    print("share of squared distance at each rank, median across encoders (%)\n")
    cols = " ".join(f"{'r'+str(k):>7s}" for k in REPORT_RANKS)
    print(f"{'construct':34s} {'leads':>6s} {'head':>6s} {cols} {'r5/r250':>8s} {'mass':>6s}")
    for c, r in T.iterrows():
        vals = " ".join(f"{r['r'+str(k)]:7.1f}" for k in REPORT_RANKS)
        print(f"{str(c)[:34]:34s} {int(r['n_leads']):6d} {r['head_r1_5']:6.1f} {vals} "
              f"{r['steepness_r5_r250']:8.1f} {r['total_mass_pct']:5.1f}%")
    print(f"\nhighlighted, by the four clauses (mass floor {MIN_MASS_PCT}% on the shape ones):")
    print(f"  top {HL_LEAD} by leadership   {by_lead}")
    print(f"  top {HL_HEAD} by head         {by_head}")
    print(f"  top {HL_STEEP} most concentrated {by_steep}")
    print(f"  top {HL_FLAT} broadest         {by_flat}")

    # ---------------------------------------------------------------- figure
    fig, ax = plt.subplots(figsize=(FIGW, FIGH))
    x = np.arange(1, n + 1)
    for c in constructs:
        if c not in hl:
            ax.plot(x, 100 * curves[c], color=GREY, linewidth=0.6, alpha=0.65, zorder=1)
    for i, c in enumerate(hl):
        col = PALETTE[i % len(PALETTE)]
        ax.fill_between(x, 100 * lo[c], 100 * hi[c], color=col, alpha=0.16, linewidth=0, zorder=2)
        ax.plot(x, 100 * curves[c], color=col, linewidth=1.7, label=c, zorder=3)

    ax.set_xscale("log")
    ax.set_xlim(1, n)
    ax.set_ylim(0, None)
    ax.set_xlabel(f"participant rank within construct  (1 = carried most, of {n})")
    ax.set_ylabel("share of that participant's\nsquared distance (%)")
    ax.grid(True, color="#E8E8E8", linewidth=0.6, zorder=0)
    ax.set_axisbelow(True)
    for s in ("top", "right"):
        ax.spines[s].set_visible(False)
    # legend BELOW rather than to the right: at 6.5 in a right-hand legend of construct names takes
    # nearly half the canvas and squeezes the data panel to about three inches.
    ax.legend(loc="upper center", bbox_to_anchor=(0.5, -0.16), ncol=2, frameon=False,
              fontsize=7.5, handlelength=1.8, columnspacing=1.6,
              title="highlighted: most participants led, tallest head, "
                    "most concentrated, or broadest",
              title_fontsize=7.5)

    fig.tight_layout()
    png = os.path.join(OUT_DIR, f"fig_construct_ranks_{tag}.png")
    fig.savefig(png, dpi=400, bbox_inches="tight")
    T.to_csv(os.path.join(OUT_DIR, f"construct_ranks_{tag}.csv"))
    print(f"\nwrote -> {png}\n         {OUT_DIR}/construct_ranks_{tag}.csv")


if __name__ == "__main__":
    main()
```
