## Supplemental code for "Scoring respondents on a semantic frame of reference identifies a group that is burdened across many questionnaires and reaches care less easily": fig_jaccard_threshold.py

```
#!/usr/bin/env python3
"""fig_jaccard_threshold.py -- Supplementary Figure S2: cross-encoder outlier agreement vs threshold.

The six encoders each flag the top (100-t)% of their own distance. This draws how much those six
flagged sets overlap as t sweeps from 90 to 99, for all 15 encoder pairs, against the overlap two
unrelated screens of the same size would give, f/(2-f) for a top fraction f.

The per-pair values are NOT in screen_summary.csv, which stores only their mean, so they are
recomputed here from the saved per-encoder distances. Nothing is refit and no threshold is re-chosen:
this reads the same scores_<enc>.csv the screen itself wrote, so the operating point (95th percentile,
consensus of all six) and every downstream number are untouched.

Reads only what the pipeline wrote, and hardcodes no result:
    screen_out/scores_<enc>.csv      (screen_battery.py) -- id, distance, pctile, flagged

Out: screen_out/figS_jaccard_threshold.png   -- copy into manuscript_nhb/figures/
     screen_out/jaccard_sweep_pairs.csv      -- the per-pair sweep, so the caption can quote it
Run: python3 fig_jaccard_threshold.py
"""
import itertools
import os

import numpy as np
import pandas as pd
import matplotlib
matplotlib.use("Agg")
import matplotlib.pyplot as plt

HERE = os.path.dirname(os.path.abspath(__file__))
OUT_DIR = os.path.join(HERE, "screen_out")
OUT_PNG = os.path.join(OUT_DIR, "figS_jaccard_threshold.png")
OUT_CSV = os.path.join(OUT_DIR, "jaccard_sweep_pairs.csv")

ENCODERS = ["gte-large-en-v1.5", "bge-m3", "bge-large-en-v1.5",
            "all-mpnet-base-v2", "e5-large-v2", "qwen3-embedding-0.6b"]
SHORT = {"gte-large-en-v1.5": "gte-large", "bge-m3": "bge-m3",
         "bge-large-en-v1.5": "bge-large", "all-mpnet-base-v2": "mpnet",
         "e5-large-v2": "e5-large", "qwen3-embedding-0.6b": "qwen3"}
SWEEP = range(90, 100)
OPERATING = 95
BLUE, VERM, GREY, INK = "#0072B2", "#D55E00", "#9AA3AB", "#222222"


def load_distances():
    """id -> distance, one column per encoder, in a common row order."""
    cols = {}
    for enc in ENCODERS:
        s = pd.read_csv(os.path.join(OUT_DIR, f"scores_{enc}.csv")).set_index("id")
        cols[enc] = s["distance"]
    D = pd.DataFrame(cols)
    assert not D.isna().any().any(), "encoders disagree on the respondent set"
    return D


def sweep(D):
    rows = []
    for t in SWEEP:
        thr = D.quantile(t / 100.0)
        sets = {m: set(D.index[D[m] > thr[m]]) for m in ENCODERS}
        f = (100 - t) / 100.0
        null = f / (2 - f)
        row = {"pctile": t, "null_jaccard": null,
               "consensus_N": len(set.intersection(*sets.values()))}
        for a, b in itertools.combinations(ENCODERS, 2):
            row[f"{SHORT[a]} / {SHORT[b]}"] = len(sets[a] & sets[b]) / len(sets[a] | sets[b])
        pairs = [v for k, v in row.items() if " / " in k]
        row["mean_jaccard"] = float(np.mean(pairs))
        row["x_null"] = row["mean_jaccard"] / null
        rows.append(row)
    return pd.DataFrame(rows)


def draw(S, out):
    pair_cols = [c for c in S.columns if " / " in c]
    fig, ax = plt.subplots(figsize=(5.4, 3.3), facecolor="white")

    for c in pair_cols:
        ax.plot(S["pctile"], S[c], "-", lw=0.7, color=GREY, alpha=0.85, zorder=2)
    ax.plot(S["pctile"], S["mean_jaccard"], "-o", ms=3.4, lw=1.8, color=BLUE, zorder=4,
            label="mean of the 15 encoder pairs")
    ax.plot(S["pctile"], S["null_jaccard"], "--", lw=1.4, color=INK, zorder=3,
            label="expected if the screens were unrelated")
    ax.plot([], [], "-", lw=0.7, color=GREY, label="each encoder pair")

    ax.axvline(OPERATING, color=VERM, lw=1.1, ls=":", zorder=1)
    r = S.loc[S["pctile"] == OPERATING].iloc[0]
    ax.annotate(f"at the {OPERATING}th percentile\n"
                f"{r['mean_jaccard']:.2f} against {r['null_jaccard']:.3f} expected\n"
                f"{int(r['consensus_N'])} flagged by all six",
                xy=(0.97, 0.52), xycoords="axes fraction", ha="right", va="center",
                fontsize=7.0, color=INK)

    ax.set_yscale("log")
    ax.set_xlabel("flagging threshold (percentile of the distance)", fontsize=8.5)
    ax.set_ylabel("overlap of the flagged sets", fontsize=8.5)
    ax.set_xticks(list(SWEEP))
    ax.tick_params(labelsize=7.5)
    ax.legend(fontsize=7.0, frameon=False, loc="lower left", handlelength=1.6)
    ax.spines[["top", "right"]].set_visible(False)
    for sp in ax.spines.values():
        sp.set_color("#9A9A9A")

    fig.tight_layout()
    fig.savefig(out, dpi=400, facecolor="white")
    print(f"  wrote {out}")


def main():
    D = load_distances()
    S = sweep(D)
    S.to_csv(OUT_CSV, index=False)

    pair_cols = [c for c in S.columns if " / " in c]
    print(f"\ncross-encoder agreement of the flagged sets, {len(D):,} respondents, "
          f"{len(pair_cols)} encoder pairs\n")
    print(f"  {'pctile':>7s} {'mean':>7s} {'null':>7s} {'x null':>7s} {'min pair':>9s} "
          f"{'max pair':>9s} {'consensus':>10s}")
    for _, r in S.iterrows():
        p = r[pair_cols].astype(float)
        print(f"  {int(r['pctile']):7d} {r['mean_jaccard']:7.3f} {r['null_jaccard']:7.4f} "
              f"{r['x_null']:7.1f} {p.min():9.3f} {p.max():9.3f} {int(r['consensus_N']):10d}")
    o = S.loc[S["pctile"] == OPERATING].iloc[0]
    print(f"\n  caption numbers at the {OPERATING}th percentile: mean {o['mean_jaccard']:.2f}, "
          f"null {o['null_jaccard']:.3f}, {o['x_null']:.0f}x the null, "
          f"{int(o['consensus_N'])} flagged by all six")
    print(f"  across the sweep the mean runs {S['mean_jaccard'].min():.2f} to "
          f"{S['mean_jaccard'].max():.2f}, never below {S['x_null'].min():.0f}x the null")

    draw(S, OUT_PNG)


if __name__ == "__main__":
    main()
```
