## Supplemental code for "Scoring respondents on a semantic frame of reference identifies a group that is burdened across many questionnaires and reaches care less easily": fig_screen.py

```
#!/usr/bin/env python3
"""fig_screen.py -- clinical breadth of the consensus outliers, as one panel.

  For each participant, how many of the eight cutoff-bearing instruments they meet the validated
  criterion for, cohort against consensus outliers.

The cross-encoder agreement sweep that used to be panel (a) is Supplementary Figure S2 (all 15
encoder pairs, generated by make_supp_tables.py), so it is no longer drawn here.

Reads only what the pipeline wrote, and hardcodes no result:
    screen_out/breadth_table.csv     (cutoffs.py)
    screen_out/run_config.json       (screen_battery.py, for the cohort N)

Out: screen_out/fig_screen.png   -- copy into manuscript_nhb/figures/
Run: python3 fig_screen.py
"""
import json
import os

import numpy as np
import pandas as pd
import matplotlib
matplotlib.use("Agg")
import matplotlib.pyplot as plt

HERE = os.path.dirname(os.path.abspath(__file__))
OUT_DIR = os.path.join(HERE, "screen_out")
BREADTH_CSV = os.path.join(OUT_DIR, "breadth_table.csv")
CFG_JSON = os.path.join(OUT_DIR, "run_config.json")
OUT_PNG = os.path.join(OUT_DIR, "fig_screen.png")
GREY, VERM, BLUE, INK = "#B4BAC0", "#D55E00", "#0072B2", "#222222"


def main():
    B = pd.read_csv(BREADTH_CSV)
    n_out = int(B["n_outliers"].sum())
    n_coh = json.load(open(CFG_JSON))["n_cohort"] if os.path.exists(CFG_JSON) else None

    fig, axB = plt.subplots(1, 1, figsize=(3.6, 2.9), facecolor="white")

    # ---- (b) clinical breadth ----
    x, w = B["breadth"].values, 0.40
    axB.bar(x - w / 2, B["pct_cohort"], w, color=GREY, lw=0,
            label=f"cohort ({n_coh:,})" if n_coh else "cohort")
    axB.bar(x + w / 2, B["pct_outliers"], w, color=VERM, lw=0,
            label=f"consensus outliers ({n_out:,})")
    mo = float((B["pct_outliers"] * x).sum() / 100.0)
    mc = float((B["pct_cohort"] * x).sum() / 100.0)
    axB.annotate(f"mean {mo:.1f} of 8\nvs {mc:.1f}", xy=(0.97, 0.72), xycoords="axes fraction",
                 ha="right", fontsize=7.5, color=INK)
    axB.set_xticks(x)
    axB.set_xlabel("validated criteria met, of the eight", fontsize=8)
    axB.set_ylabel("% of group", fontsize=8)
    axB.tick_params(labelsize=7.5)
    axB.legend(fontsize=7, frameon=False, loc="upper right")
    axB.set_title("outliers meet many criteria at once", fontsize=8.5, loc="left")

    axB.spines[["top", "right"]].set_visible(False)
    for sp in axB.spines.values():
        sp.set_color("#9A9A9A")
    fig.subplots_adjust(left=0.20, right=0.975, top=0.88, bottom=0.19)
    fig.savefig(OUT_PNG, dpi=300, facecolor="white")
    print(f"wrote {OUT_PNG}")
    print(f"  {n_out} outliers, mean breadth {mo:.2f} of 8 against cohort {mc:.2f}")


if __name__ == "__main__":
    main()
```
