## Supplemental code for "Scoring respondents on a semantic frame of reference identifies a group that is burdened across many questionnaires and reaches care less easily": five_items_to_drop.py

```
#!/usr/bin/env python3
"""five_items_to_drop.py -- how many individually-extreme items it takes to unflag each of the five.

Settles one sentence in main-text R4: "Each falls below the threshold once we set the items on which
they are individually extreme to the cohort mean, a median of one to four items." The manuscript
cites Supplementary Note S9 for that range, but the note reports the cohort-wide neutralization test
and not this quantity, and "a median of one to four" has two readings that are different numbers:

  PER PARTICIPANT   each of the five has a median over the six encoders, and those five medians
                    span some range. This is what joint_only.csv's median_items_to_drop holds.
  PER ENCODER       each encoder has a median over the five, and those six medians span some range.

Both are printed. Whichever the manuscript means, the sentence should name it.

The script REFITS NOTHING and re-chooses no threshold. It reads what the pipeline already wrote and
recomputes only medians, so running it cannot change a published number.

VERDICT LINES. Two claims in that sentence are checked explicitly rather than assumed:
  1. the group is exactly the five consensus outliers meeting none of the eight validated criteria
  2. "each falls below the threshold" -- true only if no encoder leaves one of them above it, which
     is survives_mean_n_enc == 0 for all five

In   screen_out/cutoff_flags.csv             (cutoffs.py)     id, is_outlier, breadth
     screen_out/joint_only.csv               (joint_only.py)  id, n_extreme_items,
                                                              survives_mean_n_enc, median_items_to_drop
     screen_out/joint_only_needed_mean.csv   (joint_only.py)  id x encoder, items needed to drop
                                                              the flag; 0 = never dropped
Out  screen_out/five_items_to_drop.csv       one row per participant, plus printed medians
Run  python3 five_items_to_drop.py
"""
import os
import sys

import numpy as np
import pandas as pd

HERE = os.path.dirname(os.path.abspath(__file__))
OUT_DIR = os.path.join(HERE, "screen_out")
ID_COL = "id"

CUTOFFS = os.path.join(OUT_DIR, "cutoff_flags.csv")
JOINT = os.path.join(OUT_DIR, "joint_only.csv")
NEEDED = os.path.join(OUT_DIR, "joint_only_needed_mean.csv")


def require(path, produced_by):
    if not os.path.exists(path):
        sys.exit(f"missing {path}\n  run {produced_by} first, against the same screen that "
                 f"produced the consensus outliers")
    return pd.read_csv(path)


def main():
    cut = require(CUTOFFS, "cutoffs.py")
    jo = require(JOINT, "joint_only.py")

    five = cut.loc[cut["is_outlier"].astype(bool) & (cut["breadth"] == 0), ID_COL]
    print(f"consensus outliers meeting none of the eight validated criteria: {len(five)}")
    if len(five) != 5:
        print("  NOTE: the manuscript says five. This run says "
              f"{len(five)}, so the main text is describing a different screen.")

    f = jo[jo[ID_COL].isin(five)].copy()
    missing = set(five) - set(f[ID_COL])
    if missing:
        print(f"  {len(missing)} of them are absent from joint_only.csv: {sorted(missing)}")

    print("\nper participant, from joint_only.csv")
    print(f[[ID_COL, "n_extreme_items", "survives_mean_n_enc",
             "median_items_to_drop"]].to_string(index=False))

    med_pp = f["median_items_to_drop"].dropna()
    if len(med_pp):
        print(f"\n  PER PARTICIPANT medians: {sorted(med_pp)}"
              f"\n  range {med_pp.min():g} to {med_pp.max():g}")

    # claim 2: does every one of them actually drop below the threshold, under every encoder?
    all_drop = bool((f["survives_mean_n_enc"] == 0).all()) and not missing
    print(f"\n  CLAIM 'each falls below the threshold': {'holds' if all_drop else 'DOES NOT HOLD'}"
          f"  (survives_mean_n_enc must be 0 for all; observed "
          f"{sorted(f['survives_mean_n_enc'].tolist())})")

    if not os.path.exists(NEEDED):
        print(f"\nno {os.path.basename(NEEDED)} -- the per-encoder reading needs it. It is written by"
              f"\nthe current joint_only.py, so rerun that script and then rerun this one.")
        f.to_csv(os.path.join(OUT_DIR, "five_items_to_drop.csv"), index=False)
        print(f"\nwrote -> {os.path.join(OUT_DIR, 'five_items_to_drop.csv')}")
        return

    n = pd.read_csv(NEEDED).set_index(ID_COL)
    n = n.loc[n.index.intersection(five)]
    # 0 marks a person-encoder pair where the flag never dropped. Counting those as zero items
    # would drag both medians toward zero, so they are missing, exactly as _rowmedian treats them.
    n = n.replace(0, np.nan)

    print("\nitems needed per participant and encoder (blank = the flag never dropped)")
    print(n.to_string(na_rep="-"))

    per_enc = n.median(axis=0)
    per_part = n.median(axis=1)
    print("\n  PER ENCODER medians over the five:")
    print(per_enc.round(2).to_string())
    print(f"  range {per_enc.min():g} to {per_enc.max():g}")
    print("\n  PER PARTICIPANT medians over the encoders:")
    print(per_part.round(2).to_string())
    print(f"  range {per_part.min():g} to {per_part.max():g}")

    agree = np.allclose(per_part.reindex(f[ID_COL]).values.astype(float),
                        f["median_items_to_drop"].values.astype(float), equal_nan=True)
    print(f"\n  per-participant medians match joint_only.csv: {agree}")

    out = f.merge(per_part.rename("median_over_encoders"), left_on=ID_COL, right_index=True,
                  how="left")
    out.to_csv(os.path.join(OUT_DIR, "five_items_to_drop.csv"), index=False)
    print(f"\nwrote -> {os.path.join(OUT_DIR, 'five_items_to_drop.csv')}")
    print("\nreport back: the two range lines above, and the CLAIM line.")


if __name__ == "__main__":
    main()
```
