## Supplemental code for "Scoring respondents on a semantic frame of reference identifies a group that is burdened across many questionnaires and reaches care less easily": five_report.py

```
#!/usr/bin/env python3
"""five_report.py -- the whole account of a handful of participants, one encoder, nothing aggregated.

Written for the five consensus outliers who meet NONE of the eight validated criteria, but it takes
any small set (--tag). For each participant it gives, on ONE encoder:

  WHERE THEY SIT     their distance and where it falls in the cohort

  WHAT CARRIES IT    share_c = a_c / D^2 per component, an exact partition summing to 1. A negative
                     share is a suppressor: that component pulls the distance down. Components are
                     named by their BATTERY-SIDE construct profile, the documented constructs of the
                     All of Us items that core them within the battery.

  BY WHICH ROUTE     |z|_marg = sqrt(u) = how far they sit from the COHORT on that component alone
                     |z|_cond = sqrt(c) = how far they sit from what their OTHER components predict
                     Both are z-scores, so read them in SD. The component is JOINT when c > u: more
                     surprising given the rest than on its own. Note a_c = z_marg * z_cond /
                     sqrt(1 - R_c^2), so a component carries weight only if BOTH are non-trivial, and
                     a suppressor is the two disagreeing in sign.

  DOES IT SURVIVE    the component-level counterpart of the item-level neutralization in Supplementary
                     S10. Every component on which the participant exceeds that component's own 95th
                     percentile in the cohort is set to the cohort mean, the projection, covariance
                     and threshold are all held at their unmodified values, and the distance is
                     recomputed. A participant still above the threshold is flagged by the
                     CONFIGURATION of ordinary components rather than by any conspicuous one.
                     Reported two ways: all at once, and one at a time largest-first until they drop.

                     The SAME operation is run over all 510 consensus outliers FIRST, so a
                     participant's result is quoted against the rate in the group they sit inside --
                     surviving means nothing until you know how often survival happens. It is also
                     run over a sample of never-flagged participants as a specificity check, because
                     zeroing coordinates is NOT monotone on D^2: the remaining coordinates become
                     more discordant with their conditional expectations once their neighbours are
                     set to average, so the operation can raise a distance rather than lower it.

                     This is a counterfactual on the coordinates, not on the answers -- a component is
                     a combination of items and cannot be edited on its own. The item-level version,
                     which IS a real intervention, is joint_only.py, and its verdict for the same
                     participants is read from joint_only.csv when that file is present. The two
                     answer different questions and their numbers must not be pooled.

Reads only what the pipeline wrote and recomputes the screen exactly as attribution.py does:
    responses_k0.csv, weights_k0.csv, reference_basis/, item_embeddings/, scales_prism.csv
    screen_out/consensus_outliers.csv, screen_out/cutoff_flags.csv
    screen_out/joint_only.csv                      (optional, for the item-level verdict)

Out: prints, and screen_out/five_report_<enc>_<tag>.csv (one row per participant per component)
Run: python3 five_report.py [--enc gte-large-en-v1.5] [--tag invisible] [--top 12]
     add --no-population to skip the group run and report the handful alone
"""
import os
import sys

import numpy as np
import pandas as pd
from sklearn.covariance import LedoitWolf

from attribution import (BASIS_DIR, EMB_DIR, EMB_PREFIX, RESPONSES_CSV, WEIGHTS_CSV, SCALES_CSV,
                         ITEMS_CSV, OUT_DIR, ID_COL, COVARIATES, PCTILE,
                         keep_columns, residualizer)
from attribution_components import battery_profiles, arg, DEFAULT_ENC

RULE_PCT = 95.0          # a component counts as individually extreme above its own cohort percentile


def load_cohort():
    items = pd.read_csv(ITEMS_CSV)["item"].astype(str).tolist()
    scales = pd.read_csv(SCALES_CSV).set_index("item").reindex(items)
    R = pd.read_csv(RESPONSES_CSV)
    R[ID_COL] = R[ID_COL].astype(str)
    R = R.set_index(ID_COL)
    raw = R[items].values.astype(float)
    assert not np.isnan(raw).any(), "missing responses -- use the k0 file"

    lo = scales["min"].values.astype(float)
    hi = scales["max"].values.astype(float)
    rev = scales["reverse"].astype(str).str.lower().eq("true").values
    X = np.clip(2.0 * (raw - lo) / (hi - lo) - 1.0, -1.0, 1.0)
    X[:, rev] *= -1.0

    cov = R[COVARIATES].values.astype(float) if COVARIATES else None
    w = (pd.read_csv(WEIGHTS_CSV).iloc[:, 0].values.astype(float)
         if WEIGHTS_CSV and os.path.exists(WEIGHTS_CSV) else np.ones(len(R)))
    return R, items, X, cov, w


def select(R, tag):
    sel = pd.read_csv(os.path.join(OUT_DIR, "consensus_outliers.csv"))
    sel[ID_COL] = sel[ID_COL].astype(str)
    target = set(sel.loc[sel["flagged_by_all"].astype(bool), ID_COL])
    if tag == "invisible":
        cf = pd.read_csv(os.path.join(OUT_DIR, "cutoff_flags.csv"))
        cf[ID_COL] = cf[ID_COL].astype(str)
        target &= set(cf.loc[cf["breadth"] == 0, ID_COL])
    return [pid for pid in R.index if pid in target]


def short(profile, n=3):
    return "; ".join(str(profile).split("; ")[:n])


def partners(y, Om, sd, c_idx):
    """What the participant's OTHER components predict for component c, partner by partner.

    Splitting the diagonal out of g_c = (Omega y)_c,

        g_c = Omega_cc y_c  +  sum_{k != c} Omega_ck y_k,   sum_{k != c} Omega_ck y_k = -Omega_cc E[y_c | y_-c]

    so the own term IS the observed score and the partner terms ARE the prediction, signed. Dividing
    by -Omega_cc puts each partner's contribution in the same units as the score, and they sum
    EXACTLY to the predicted value. A component the participant is unremarkable on (y_k ~ 0)
    contributes ~0 and drops out on its own, so ignoring quiet partners is exact.

    Everything is returned in MARGINAL SD of component c, which is what the observed score is read
    in. The residual y_c - E is reported separately in CONDITIONAL SD (that is |z|_cond = sqrt(c_c)),
    a tighter yardstick, which is why the two do not subtract.

    CANCELLATION. The prediction can be a near-cancellation of large opposing partner terms, in which
    case no small set of components accounts for it. That is a fact about the participant, not a
    defect, and it is reported per person rather than used to suppress the layer. sum|contrib| against
    |prediction| measures it.

    This is NOT the quadratic pairwise layer A_cd = Omega_cd y_c y_d, which Methods declines to report
    because on a battery whose kept components approach its item count it is dominated by cancellation
    (HRS: diagonal 6049% of T^2, off-diagonal -5949%). This one sums to the prediction, an
    interpretable quantity in its own units.
    """
    contrib = -(Om[c_idx] * y) / Om[c_idx, c_idx] / sd[c_idx]   # each partner's share of E[y_c|rest]
    contrib[c_idx] = 0.0                                        # the own term is the observed score
    pred = float(contrib.sum())
    obs = float(y[c_idx] / sd[c_idx])
    cancel = float(np.abs(contrib).sum() / max(abs(pred), 1e-9))
    return contrib, obs, pred, cancel


def neutralize(y, Om, sd, zcut, thr):
    """Zero every component above its own cohort percentile, on a FIXED yardstick.

    Returns (n_neutralized, D2 after zeroing them all at once, how many are needed one at a time
    largest-first before the distance falls below the threshold, -1 if it never does).
    """
    ex = np.where(np.abs(y) / sd > zcut)[0]
    y0 = y.copy()
    y0[ex] = 0.0
    batch = float(y0 @ Om @ y0)
    need, y1 = -1, y.copy()
    for n, j in enumerate(ex[np.argsort(-np.abs(y[ex]) / sd[ex])], start=1):
        y1[j] = 0.0
        if float(y1 @ Om @ y1) <= thr:
            need = n
            break
    return len(ex), batch, need


def population_survival(Yc, Om, sd, zcut, thr, T2, flagged, label, rng_seed=0, n_control=2000):
    """The base rate the five are read against, plus the specificity check for the supplement.

    Every flagged participant is put through the same operation, so 'four of the five survive' can be
    compared with how often survival happens across the group. The control asks the opposite question:
    does the operation push participants who were NEVER flagged over the threshold? Zeroing
    coordinates is not monotone on D^2 -- the remaining ones become more discordant with their
    conditional expectations once their neighbours are set to average -- so this cannot be assumed.
    """
    def run(rows):
        n_ex, batch, need = [], [], []
        for i in rows:
            a, b, c = neutralize(Yc[i], Om, sd, zcut, thr)
            n_ex.append(a); batch.append(b); need.append(c)
        return np.array(n_ex), np.array(batch), np.array(need)

    idx = np.where(flagged)[0]
    n_ex, batch, need = run(idx)
    surv = batch > thr
    rose = batch > T2[idx]
    print(f"\n{'='*104}\ncomponent-level neutralization across the {len(idx)} {label}\n")
    print(f"  a participant has a median {np.median(n_ex):.0f} components above their own "
          f"{RULE_PCT:g}th percentile (range {n_ex.min()}-{n_ex.max()})")
    print(f"  STILL FLAGGED after zeroing all of them: {int(surv.sum())} of {len(idx)} "
          f"({100*surv.mean():.1f}%)")
    print(f"  the distance ROSE rather than fell for {int(rose.sum())} of {len(idx)} "
          f"({100*rose.mean():.1f}%) -- zeroing coordinates is not monotone on D^2")
    drops = need[need > 0]
    print(f"  among those that do drop, a median {np.median(drops):.0f} components are needed "
          f"(range {drops.min()}-{drops.max()})" if len(drops) else "  none drop")

    rng = np.random.default_rng(rng_seed)
    pool = np.where((~flagged) & (T2 <= thr))[0]
    ctrl = rng.choice(pool, size=min(n_control, len(pool)), replace=False)
    _, cbatch, _ = run(ctrl)
    crossed = cbatch > thr
    print(f"\n  CONTROL, {len(ctrl):,} participants who were never flagged: the same operation puts "
          f"{int(crossed.sum())} of them ({100*crossed.mean():.2f}%) above the threshold")
    return surv, batch


def main():
    enc = arg("--enc", DEFAULT_ENC)
    tag = arg("--tag", "invisible")
    top = int(arg("--top", "12"))

    R, items, X, cov, w = load_cohort()
    resid = residualizer(cov, w)
    prof, _ = battery_profiles(enc)

    b = np.load(os.path.join(BASIS_DIR, f"{enc}.npz"), allow_pickle=True)
    W, mu, sigma, mu_s, band = b["W"], b["mu"], b["sigma"], b["mu_scores"], b["band"]
    e = np.load(os.path.join(EMB_DIR, f"{EMB_PREFIX}_{enc}.npz"), allow_pickle=True)
    pos = {str(x): j for j, x in enumerate(e["items"])}
    E = np.asarray(e["vectors"], float)[[pos[it] for it in items]]
    P = (E - mu) @ W.T
    kept = keep_columns((P - mu_s) / sigma, band)

    Y = resid(X @ P[:, kept])
    Yc = Y - np.average(Y, axis=0, weights=w)
    lw = LedoitWolf().fit(Yc)
    Om, Sig = lw.precision_, lw.covariance_
    G = Yc @ Om
    T2 = np.einsum("ij,ij->i", Yc, G)
    thr = np.percentile(T2, PCTILE)                       # the flagging threshold, held fixed
    sd = np.sqrt(np.diag(Sig))
    zmarg_all = np.abs(Yc) / sd                           # cohort-wide, for the extremity rule
    zcut = np.percentile(zmarg_all, RULE_PCT, axis=0)     # per component, its own 95th percentile

    row_of = {pid: i for i, pid in enumerate(R.index)}
    # the population run first, so the handful below is quoted against the group it sits inside
    grp_surv = None
    if "--no-population" not in sys.argv:
        all_flagged = np.zeros(len(R), dtype=bool)
        for pid in select(R, "outliers"):
            all_flagged[row_of[pid]] = True
        grp_surv, _ = population_survival(Yc, Om, sd, zcut, thr, T2, all_flagged,
                                          "consensus outliers")

    ids = select(R, tag)
    assert ids, f"no participants for tag={tag}"
    jo = None
    jo_path = os.path.join(OUT_DIR, "joint_only.csv")
    if os.path.exists(jo_path):
        jo = pd.read_csv(jo_path)
        jo[ID_COL] = jo[ID_COL].astype(str)
        jo = jo.set_index(ID_COL)

    print(f"\n{'='*104}\n{len(ids)} participants, tag={tag}, encoder {enc}")
    print(f"cohort {len(R):,}, {len(kept)} kept components, flagging threshold "
          f"D^2 = {thr:.1f} (the {PCTILE:g}th percentile, held fixed throughout)\n")

    out = []
    for pid in ids:
        i = row_of[pid]
        y, g = Yc[i], G[i]
        a = y * g
        share = a / T2[i]
        u = y ** 2 / np.diag(Sig)
        c = g ** 2 / np.diag(Om)
        pct = 100.0 * (T2 < T2[i]).mean()

        print(f"{'-'*104}\nparticipant {pid}   D^2 = {T2[i]:.1f}, above "
              f"{pct:.1f}% of the cohort ({T2[i]/thr:.2f}x the threshold)")

        order = np.argsort(-np.abs(share))[:top]
        print(f"  {'comp':>5s} {'share':>7s} {'|z|marg':>8s} {'|z|cond':>8s} {'route':>9s}  "
              f"battery-side profile")
        for j in order:
            route = "joint" if c[j] > u[j] else "marginal"
            mark = "  (suppressor)" if share[j] < 0 else ""
            print(f"  C{kept[j]:<4d} {100*share[j]:6.1f}% {np.sqrt(u[j]):8.2f} "
                  f"{np.sqrt(c[j]):8.2f} {route:>9s}  {short(prof.get(kept[j], ''))[:52]}{mark}")
        pos_n = int((share > 0).sum())
        print(f"  {pos_n} of {len(kept)} components carry, {len(kept)-pos_n} suppress; "
              f"the top {int(np.searchsorted(np.cumsum(np.sort(share)[::-1]), 0.5) + 1)} reach half "
              f"the distance; integration {100*share[c > u].sum():.1f}%")

        # ---- component-level neutralization, the fixed-yardstick counterfactual
        n_ex, t2_batch, need = neutralize(y, Om, sd, zcut, thr)
        extreme = np.where(np.abs(y) / sd > zcut)[0]
        survives = t2_batch > thr
        print(f"  neutralizing the {n_ex} components above their own {RULE_PCT:g}th "
              f"percentile: D^2 {T2[i]:.1f} -> {t2_batch:.1f}, "
              f"{'STILL FLAGGED' if survives else 'drops below threshold'}"
              + (f", against {100*grp_surv.mean():.0f}% of the group" if grp_surv is not None else ""))
        print(f"    one at a time, largest first: "
              + (f"{need} needed to drop below" if need > 0 else "never drops"))

        # ---- what do their OTHER components predict for the one that carries the most
        lead = int(np.argmax(share))
        contrib, obs, pred, cancel = partners(y, Om, sd, lead)
        print(f"  on C{kept[lead]} they scored {obs:+.2f} SD and their other components predict "
              f"{pred:+.2f} SD, a gap of {np.sqrt(c[lead]):.2f} conditional SD")
        if cancel < 4:
            print(f"    that prediction is carried by:")
            print(f"    {'comp':>5s} {'contrib':>8s} {'% of pred':>10s} {'own |z|':>8s}  "
                  f"battery-side profile")
            for j in np.argsort(-np.abs(contrib))[:5]:
                print(f"    C{kept[j]:<4d} {contrib[j]:+8.2f} {100*contrib[j]/pred:+9.0f}% "
                      f"{np.abs(y[j])/sd[j]:8.2f}  {short(prof.get(kept[j], ''))[:44]}")
            print(f"    contributions sum to the prediction exactly; sum of |contributions| "
                  f"{np.abs(contrib).sum():.2f} against |prediction| {abs(pred):.2f} "
                  f"(ratio {cancel:.1f}, a few components account for it)")
        else:
            big = np.argsort(-np.abs(contrib))[:3]
            print(f"    NO SMALL SET ACCOUNTS FOR THAT PREDICTION: the contributions are large and "
                  f"opposing, summing to {pred:+.2f} SD from {np.abs(contrib).sum():.2f} SD of "
                  f"absolute size (ratio {cancel:.1f})")
            print(f"    largest, for the record: "
                  + ", ".join(f"C{kept[j]} {contrib[j]:+.2f}" for j in big))
        if jo is not None and pid in jo.index:
            r = jo.loc[pid]
            print(f"    item-level test (Supplementary S10, a different question): "
                  f"{int(r['n_extreme_items'])} items individually extreme, survives their removal "
                  f"under {int(r['survives_mean_n_enc'])} of 6 encoders "
                  f"({int(r['survives_winsor_n_enc'])} of 6 when capped rather than zeroed), "
                  f"median {r['median_items_to_drop']} items needed to drop")

        for j in range(len(kept)):
            out.append(dict(id=pid, component=kept[j], share=share[j], z_marg=np.sqrt(u[j]),
                            z_cond=np.sqrt(c[j]), joint=bool(c[j] > u[j]),
                            neutralized=bool(j in set(extreme)),
                            battery_profile=prof.get(kept[j], "")))

    path = os.path.join(OUT_DIR, f"five_report_{enc}_{tag}.csv")
    pd.DataFrame(out).to_csv(path, index=False)
    print(f"{'-'*104}\n  wrote {path}")


if __name__ == "__main__":
    main()
```
