## Supplemental code for "Scoring respondents on a semantic frame of reference identifies a group that is burdened across many questionnaires and reaches care less easily": five_xenc.py

```
#!/usr/bin/env python3
"""five_xenc.py -- the same handful of participants under all six encoders (supplement).

The main text reports these participants on one encoder. This is the check that the account does not
depend on that choice.

WHAT IS COMPARED. Component indices are NOT comparable across encoders -- each encoder fits its own
basis, so its C12 is not another's C12. The comparable object is the CONSTRUCT PROFILE: the documented
constructs of the All of Us items that core a component within the battery. So for each participant
this reports, under each encoder:

    the component carrying the most of their distance, its share, and its battery-side profile
    the union of profiles over the components that reach half their distance -- "what carries them"

and then how far the six agree, as the mean pairwise overlap of those construct sets over the 15
encoder pairs (1 for identical sets, 0 for sets sharing nothing). This is the per-participant version
of the item-side quantity in main-text Figure 2b, and it needs no component matching at all.

CHANCE LEVEL, and why one is needed. An overlap of 0.47 means nothing on its own: Jaccard between two
construct sets depends on how many constructs are in circulation and how common each is. More to the
point, "the same participant gets the same content under six independently trained encoders" is
evidence that the method identifies THAT PARTICIPANT only if a DIFFERENT participant would have got
different content. A method that answered "discrimination" for everybody would look perfectly stable.

Two floors are reported, because they answer different questions:
  WITHIN the reported set  -- different participants drawn from the handful being reported. Harsh when
                             that handful is content-homogeneous, as the five invisible outliers are:
                             the floor is high because they resemble each other, not because the
                             method is unstable.
  ACROSS the 510           -- different participants drawn from all consensus outliers, whose content
                             is far more varied. This is the floor the capability claim needs, and it
                             requires attribution_component_shares_outliers.csv to be present.

Reads only what the pipeline wrote, and hardcodes no result:
    screen_out/attribution_component_shares_<tag>.csv   (attribution.py, all six encoders)
    reference_basis/<enc>.npz, item_embeddings/, construct_map_151.csv, core_items_151.csv

--summary suppresses the per-participant blocks and prints their DISTRIBUTION instead, which is what
makes `--tag outliers` runnable: over all 510 the per-participant listing is 510 blocks long, and the
reportable object is the spread of the per-participant agreements, not any one of them. With
--tag outliers the within-set floor IS the 510 floor, so only one floor is printed.

Out: prints, and screen_out/five_xenc_<tag>.csv (one row per participant per encoder,
     plus agree_lead / agree_half per participant)
Run: python3 five_xenc.py [--tag invisible] [--half 0.5] [--summary]
"""
import itertools
import os
import sys

import numpy as np
import pandas as pd

from attribution import OUT_DIR, ID_COL, ENCODERS
from attribution_components import battery_profiles, arg

SHORT = {"gte-large-en-v1.5": "gte-large", "bge-m3": "bge-m3", "bge-large-en-v1.5": "bge-large",
         "all-mpnet-base-v2": "mpnet", "e5-large-v2": "e5-large", "qwen3-embedding-0.6b": "qwen3"}


def constructs(profile):
    """The SET of documented constructs in a battery-side profile string 'A:3; B:1'."""
    p = str(profile).strip()
    if not p or p.startswith("("):
        return set()
    return {s.split(":")[0].strip() for s in p.split(";") if s.strip()}


def jac(a, b):
    return len(a & b) / len(a | b) if (a | b) else 0.0


def spearman(a, b):
    """Rank correlation, without a scipy dependency (the pipeline uses numpy and pandas only)."""
    ra = pd.Series(a).rank().values - (len(a) + 1) / 2
    rb = pd.Series(b).rank().values - (len(b) + 1) / 2
    d = np.sqrt((ra ** 2).sum() * (rb ** 2).sum())
    return float((ra * rb).sum() / d) if d > 0 else float("nan")


def boot_spearman(a, b, n=2000, seed=0):
    """Percentile interval over participants."""
    rng = np.random.default_rng(seed)
    r = [spearman(a[i], b[i]) for i in (rng.integers(len(a), size=len(a)) for _ in range(n))]
    return float(np.nanpercentile(r, 2.5)), float(np.nanpercentile(r, 97.5))


def profile_sets(D, prof, half):
    """(participant, encoder) -> leading-component constructs, and constructs of what carries them."""
    rows, lead_sets, half_sets = [], {}, {}
    for pid, gp in D.groupby(ID_COL):
        for enc in ENCODERS:
            g = gp[gp["encoder"] == enc].sort_values("share", ascending=False)
            if not len(g):
                continue
            lead = g.iloc[0]
            n_half = int(np.searchsorted(np.cumsum(g["share"].values), half) + 1)
            carrying = g.head(n_half)
            lset = constructs(prof[enc].get(int(lead["component"]), ""))
            hset = set().union(*[constructs(prof[enc].get(int(c), ""))
                                 for c in carrying["component"]]) if n_half else set()
            lead_sets[(pid, enc)] = lset
            half_sets[(pid, enc)] = hset
            rows.append(dict(id=pid, encoder=enc, lead_component=int(lead["component"]),
                             lead_share=float(lead["share"]), n_to_half=n_half,
                             lead_profile=prof[enc].get(int(lead["component"]), ""),
                             half_constructs="; ".join(sorted(hset))))
    return pd.DataFrame(rows), lead_sets, half_sets


def cross_person_null(lead_sets, half_sets, n_pairs=40000, seed=0):
    """Overlap between DIFFERENT participants under DIFFERENT encoders, sampled."""
    keys = list(lead_sets)
    rng = np.random.default_rng(seed)
    nl, nh = [], []
    while len(nl) < n_pairs:
        (p1, e1) = keys[rng.integers(len(keys))]
        (p2, e2) = keys[rng.integers(len(keys))]
        if p1 == p2 or e1 == e2:
            continue
        nl.append(jac(lead_sets[(p1, e1)], lead_sets[(p2, e2)]))
        nh.append(jac(half_sets[(p1, e1)], half_sets[(p2, e2)]))
    return float(np.mean(nl)), float(np.mean(nh)), len(nl)


def main():
    tag = arg("--tag", "invisible")
    half = float(arg("--half", "0.5"))
    summary = "--summary" in sys.argv

    D = pd.read_csv(os.path.join(OUT_DIR, f"attribution_component_shares_{tag}.csv"))
    D[ID_COL] = D[ID_COL].astype(str)
    prof = {e: battery_profiles(e)[0] for e in ENCODERS}
    T, lead_sets, half_sets = profile_sets(D, prof, half)

    ids = list(dict.fromkeys(T[ID_COL]))
    print(f"\n{'='*112}\n{len(ids)} participants under {len(ENCODERS)} encoders, tag={tag}")
    print(f"components are named by the constructs of the All of Us items that core them within the "
          f"battery,\nwhich is the comparable object across encoders -- component indices are not\n")

    agr_l, agr_h = {}, {}
    for pid in ids:
        lv = [jac(lead_sets[(pid, a)], lead_sets[(pid, b)])
              for a, b in itertools.combinations(ENCODERS, 2)]
        hv = [jac(half_sets[(pid, a)], half_sets[(pid, b)])
              for a, b in itertools.combinations(ENCODERS, 2)]
        agr_l[pid], agr_h[pid] = float(np.mean(lv)), float(np.mean(hv))
        if summary:
            continue
        g = T[T[ID_COL] == pid]
        print(f"{'-'*112}\nparticipant {pid}")
        print(f"  {'encoder':12s} {'lead':>5s} {'share':>7s} {'to half':>8s}  "
              f"constructs of the leading component")
        for _, r in g.iterrows():
            print(f"  {SHORT.get(r['encoder'], r['encoder']):12s} C{r['lead_component']:<4d} "
                  f"{100*r['lead_share']:6.1f}% {r['n_to_half']:8d}  "
                  f"{', '.join(sorted(constructs(r['lead_profile'])))[:62]}")
        shared = set.intersection(*[half_sets[(pid, e)] for e in ENCODERS])
        recur = {c for c in set().union(*[half_sets[(pid, e)] for e in ENCODERS])
                 if sum(c in half_sets[(pid, e)] for e in ENCODERS) >= 4}
        print(f"  agreement over the 15 encoder pairs: leading component {agr_l[pid]:.2f}, "
              f"what carries them {agr_h[pid]:.2f}")
        print(f"  constructs carrying them under ALL six: "
              f"{', '.join(sorted(shared)) if shared else 'none'}")
        print(f"  under at least four:                   "
              f"{', '.join(sorted(recur)) if recur else 'none'}")

    T["agree_lead"] = T[ID_COL].map(agr_l)
    T["agree_half"] = T[ID_COL].map(agr_h)
    obs_l = float(np.mean([agr_l[p] for p in ids]))
    obs_h = float(np.mean([agr_h[p] for p in ids]))
    within_l, within_h, n_within = cross_person_null(lead_sets, half_sets)

    if summary:
        L = pd.Series(agr_l), pd.Series(agr_h)
        print(f"{'-'*112}\nper-participant agreement over the 15 encoder pairs, {len(ids)} "
              f"participants\n")
        print(f"  {'':22s} {'median':>8s} {'quartiles':>16s} {'min':>7s} {'max':>7s}")
        for name, s in zip(("leading component", "what carries them"), L):
            print(f"  {name:22s} {s.median():8.2f} "
                  f"{s.quantile(.25):7.2f}-{s.quantile(.75):<8.2f} {s.min():7.2f} {s.max():7.2f}")
        n_half = T.groupby(ID_COL)["n_to_half"].median()
        print(f"\n  the leading component carries a median {100*T['lead_share'].median():.1f}% of a "
              f"participant's distance,\n  and a median {n_half.median():.0f} components reach half "
              f"of it")

        # DOES REPRODUCIBILITY TRACK CONCENTRATION? A participant whose distance piles onto one
        # component gives the six encoders one thing to agree about, and a participant whose distance
        # is spread over many gives them a choice among near-ties. This is the same relation S10 finds
        # at the construct layer (the survivors of item-level neutralization are the diffuse ones, and
        # the encoders agree less often on them), tested here on components. Concentration is a
        # participant's MEDIAN leading share over the six encoders, so it does not depend on the
        # exemplar, and the correlation is Spearman with a bootstrap interval over participants.
        conc = T.groupby(ID_COL)["lead_share"].median()
        A = pd.DataFrame({"conc": conc, "n_half": n_half,
                          "agree_lead": pd.Series(agr_l), "agree_half": pd.Series(agr_h)}).dropna()
        print(f"\n  CONCENTRATION AGAINST REPRODUCIBILITY, {len(A)} participants (Spearman, "
              f"95% bootstrap)")
        for x, y in (("conc", "agree_lead"), ("conc", "agree_half"),
                     ("n_half", "agree_lead"), ("n_half", "agree_half")):
            r = spearman(A[x], A[y])
            lo, hi = boot_spearman(A[x].values, A[y].values)
            print(f"    {x:7s} vs {y:11s} rho {r:+.2f}  ({lo:+.2f} to {hi:+.2f})")
        q = A["conc"].quantile([.25, .75])
        lowq, highq = A[A["conc"] <= q[.25]], A[A["conc"] >= q[.75]]
        print(f"    the most concentrated quartile (leading share >= {100*q[.75]:.1f}%) agrees at "
              f"{highq['agree_lead'].mean():.2f} on the leading\n    component against "
              f"{lowq['agree_lead'].mean():.2f} for the least concentrated quartile "
              f"(<= {100*q[.25]:.1f}%)")

    print(f"{'-'*112}\nacross all {len(ids)} participants, mean over the 15 encoder pairs")
    print(f"  {'':40s} {'leading component':>18s} {'what carries them':>18s}")
    print(f"  {'same participant, different encoder':40s} {obs_l:18.2f} {obs_h:18.2f}")
    print(f"  {'different participant, within this set':40s} {within_l:18.2f} {within_h:18.2f}")

    # the floor the capability claim needs: a different participant drawn from ALL consensus outliers,
    # whose content is far more varied than a handful selected for meeting no clinical criterion
    grp_path = os.path.join(OUT_DIR, "attribution_component_shares_outliers.csv")
    if os.path.exists(grp_path) and tag != "outliers":
        G = pd.read_csv(grp_path)
        G[ID_COL] = G[ID_COL].astype(str)
        _, g_lead, g_half = profile_sets(G, prof, half)
        n_grp = len({p for p, _ in g_lead})
        mix_l, mix_h, n_mix = cross_person_null({**lead_sets, **g_lead}, {**half_sets, **g_half})
        print(f"  {f'different participant, of the {n_grp}':40s} {mix_l:18.2f} {mix_h:18.2f}")
        print(f"\n  the two floors differ because the {len(ids)} reported here were selected for "
              f"meeting no clinical\n  criterion, which makes them resemble one another -- the "
              f"{n_grp}-outlier floor is the one the\n  per-participant claim has to clear "
              f"({n_mix:,} sampled pairs, {n_within:,} within-set)")
    elif tag == "outliers":
        print(f"\n  the reported set IS the flagged group, so the within-set floor above is the "
              f"floor the\n  per-participant claim has to clear ({n_within:,} sampled pairs) -- "
              f"there is no separate one")
    else:
        print(f"\n  (attribution_component_shares_outliers.csv absent -- the {n_within:,}-pair "
              f"within-set floor is all that could be computed)")

    path = os.path.join(OUT_DIR, f"five_xenc_{tag}.csv")
    T.to_csv(path, index=False)
    print(f"\n  wrote {path}")


if __name__ == "__main__":
    main()
```
