## Supplemental code for "Scoring respondents on a semantic frame of reference identifies a group that is burdened across many questionnaires and reaches care less easily": heterogeneity.py

```
#!/usr/bin/env python3
"""heterogeneity.py -- do participants with the same total meet it with the same content? (R5 P1)

THE QUESTION. A validated screen reduces a participant to a count: how many of the eight cutoffs they
meet. If that count told you what carries them, attribution would add nothing over the count. This
asks the opposite way round -- hold the count fixed and look at how varied the content is.

WHAT IS COMPARED. A participant's LEADING CONSTRUCT, the documented construct carrying the most of
their squared distance, summing item contributions within constructs (Methods, the attribution
section). Constructs rather than components, because the construct is the object all six encoders
share, and the leading construct is what attribution.py already publishes per participant.

WHAT IS REPORTED, for each breadth stratum and for the group:

  distinct        how many different constructs lead the participants at that breadth
  largest share   the biggest single construct's share of that stratum. If the count determined the
                  content this would be near 1
  Cramer's V      association between breadth (0-8) and leading construct over the whole group, 0 when
                  the count says nothing at all about the content and 1 when it fixes it. Reported
                  with the number of strata and constructs it is computed over, since V is bounded
                  away from 0 in small tables and should not be read as a correlation

RARE CONTENT. The second half asks a different question of the same participants. Components differ
enormously in how many people they are largest for -- a few are largest for dozens, most for a handful
-- so a participant is either carried by content shared with much of the group or by content almost
specific to them. This splits the group on that and compares what the validated criteria make of each
side. If the participants carried by rare content meet fewer criteria, the tail is not merely
heterogeneity: it is where reading the instruments together adds what scoring them one at a time
cannot.

NO NULL IS COMPUTED. The comparison is against the count itself, not against chance: the claim is that
two participants with the same count are carried by different content, and the stratum tables show it
directly.

Reads only what the pipeline wrote, and hardcodes no result:
    screen_out/attribution_per_person_<tag>.csv        (attribution.py) -- id, top_construct
    screen_out/cutoff_flags.csv                        (cutoffs.py)     -- id, breadth, is_outlier
    screen_out/attribution_component_shares_<tag>.csv  (attribution.py) -- for the rare-content split
    reference_basis/, item_embeddings/                 -- to name the components

Out: prints, screen_out/heterogeneity_<tag>.csv (one row per breadth stratum) and
     screen_out/rare_content_<tag>.csv (one row per component: how many it is largest for, and the
     mean breadth of those participants)
Run: python3 heterogeneity.py [--tag outliers] [--enc gte-large-en-v1.5] [--rare-leads 10]
"""
import os

import numpy as np
import pandas as pd

from attribution import OUT_DIR, ID_COL
from attribution_components import arg, battery_profiles, DEFAULT_ENC


def cramers_v(tab):
    """Association between two categoricals from their contingency table, without scipy."""
    n = tab.values.sum()
    if n == 0:
        return float("nan")
    exp = np.outer(tab.sum(axis=1), tab.sum(axis=0)) / n
    with np.errstate(divide="ignore", invalid="ignore"):
        chi2 = np.nansum(np.where(exp > 0, (tab.values - exp) ** 2 / exp, 0.0))
    r, k = tab.shape
    denom = n * (min(r, k) - 1)
    return float(np.sqrt(chi2 / denom)) if denom > 0 else float("nan")


def short(profile, n=3):
    p = str(profile).strip()
    if not p or p.startswith("("):
        return "no battery core"
    names = [x.split(":")[0].strip().lower() for x in p.split(";") if x.strip()]
    return ", ".join(names[:n]) + (f", +{len(names) - n}" if len(names) > n else "")


def rare_content(F, tag, enc, rare_leads):
    """Split the group by whether the component carrying the most of a participant's distance is one
    that carries most for many participants or for almost none, then compare the validated criteria."""
    path = os.path.join(OUT_DIR, f"attribution_component_shares_{tag}.csv")
    if not os.path.exists(path):
        print(f"\n  ({os.path.basename(path)} absent -- rare-content split skipped)")
        return
    D = pd.read_csv(path)
    D[ID_COL] = D[ID_COL].astype(str)
    D = D[D["encoder"] == enc]
    assert len(D), f"no rows for {enc}"
    piv = D.pivot_table(index=ID_COL, columns="component", values="share").fillna(0.0)
    largest = piv.idxmax(axis=1).rename("largest_component")
    n_largest = largest.value_counts()

    prof, _ = battery_profiles(enc)
    G = pd.DataFrame(largest).join(F.set_index(ID_COL)["breadth"], how="inner")
    G["n_for_that_component"] = G["largest_component"].map(n_largest)
    G["rare"] = G["n_for_that_component"] < rare_leads

    R = pd.DataFrame({"n_largest_for": n_largest})
    R["mean_breadth"] = G.groupby("largest_component")["breadth"].mean()
    R["profile"] = [short(prof.get(int(c), "")) for c in R.index]
    R = R.sort_values("n_largest_for", ascending=False)

    print(f"\n{'='*100}\nRARE CONTENT: is the largest component one that leads many, or almost "
          f"nobody? (threshold {rare_leads})\n")
    print(f"  {'':22s} {'n':>5s} {'mean criteria':>14s} {'meet none':>10s} {'meet 5+':>8s}")
    for lab, sel in (("carried by rare content", G["rare"]), ("by common content", ~G["rare"])):
        g = G[sel]
        print(f"  {lab:22s} {len(g):5d} {g['breadth'].mean():14.2f} "
              f"{100*(g['breadth'] == 0).mean():9.1f}% {100*(g['breadth'] >= 5).mean():7.1f}%")
    print(f"\n  the {int(G['rare'].sum())} carried by rare content are spread over "
          f"{int(G.loc[G['rare'], 'largest_component'].nunique())} components:")
    for c, r in R[R["n_largest_for"] < rare_leads].iterrows():
        print(f"    largest for {int(r['n_largest_for']):2d}, mean criteria "
              f"{r['mean_breadth']:.1f}   {r['profile'][:52]}")
    out = os.path.join(OUT_DIR, f"rare_content_{tag}.csv")
    R.rename_axis("component").to_csv(out)
    print(f"\n  wrote {out}")


def main():
    tag = arg("--tag", "outliers")
    enc = arg("--enc", DEFAULT_ENC)
    rare_leads = int(arg("--rare-leads", "10"))

    P = pd.read_csv(os.path.join(OUT_DIR, f"attribution_per_person_{tag}.csv"))
    P[ID_COL] = P[ID_COL].astype(str)
    F = pd.read_csv(os.path.join(OUT_DIR, "cutoff_flags.csv"))
    F[ID_COL] = F[ID_COL].astype(str)

    D = P.merge(F[[ID_COL, "breadth"]], on=ID_COL, how="inner")
    assert len(D) == len(P), (f"{len(P) - len(D)} of {len(P)} participants have no breadth row -- "
                              f"cutoff_flags.csv and attribution_per_person_{tag}.csv disagree")

    print(f"\n{len(D)} participants, {D['top_construct'].nunique()} distinct leading constructs, "
          f"breadth {int(D['breadth'].min())}-{int(D['breadth'].max())}\n")

    rows = []
    print(f"  {'criteria met':>12s} {'n':>5s} {'distinct':>9s} {'largest':>8s}  most common leading "
          f"constructs")
    for b, g in D.groupby("breadth"):
        vc = g["top_construct"].value_counts()
        rows.append(dict(breadth=int(b), n=len(g), distinct=int(vc.size),
                         largest_share=float(vc.iloc[0] / len(g)),
                         largest_construct=str(vc.index[0])))
        top = ", ".join(f"{c} ({n})" for c, n in vc.head(3).items())
        print(f"  {int(b):12d} {len(g):5d} {vc.size:9d} {100*vc.iloc[0]/len(g):7.0f}%  {top[:58]}")

    T = pd.DataFrame(rows).set_index("breadth")
    tab = pd.crosstab(D["breadth"], D["top_construct"])
    v = cramers_v(tab)
    big = T[T["n"] >= 20]
    print(f"\n  over the whole group, breadth against leading construct: Cramer's V = {v:.2f} "
          f"({tab.shape[0]} strata x {tab.shape[1]} constructs)")
    if len(big):
        print(f"  in the {len(big)} strata holding at least 20 participants, a stratum spans "
              f"{int(big['distinct'].min())} to {int(big['distinct'].max())} distinct leading "
              f"constructs,\n  and the most common accounts for "
              f"{100*big['largest_share'].min():.0f} to {100*big['largest_share'].max():.0f}% of it")

    path = os.path.join(OUT_DIR, f"heterogeneity_{tag}.csv")
    T.to_csv(path)
    print(f"\n  wrote {path}")

    rare_content(F, tag, enc, rare_leads)


if __name__ == "__main__":
    main()
```
