## Supplemental code for "Scoring respondents on a semantic frame of reference identifies a group that is burdened across many questionnaires and reaches care less easily": joint_only.py

```
#!/usr/bin/env python3
"""joint_only.py -- can a flag be accounted for by the items the participant is individually extreme on?

The reachability test in the supplement asks whether SOME single item puts a participant in its own
upper tail. Nearly everyone passes that, so it separates no one. This asks the sharper question:

    neutralize every item on which the participant is individually extreme, and see whether the
    distance still exceeds the flagging threshold.

A participant who survives that is a genuinely JOINT case -- their flag is not accounted for by any
item that a single-instrument review would have stopped on. A participant who drops out was, to that
extent, findable one item at a time.

PROCEDURE, per participant and encoder

  1. Item z-scores are computed against the cohort, so an item counts as INDIVIDUALLY EXTREME when
     z exceeds that item's own 95th percentile, strictly. Strict is deliberate: under the inclusive
     rule a median of ~64 of the 151 items qualify, which guts the profile and tells us nothing.
  2. Neutralize those items one at a time, largest z first, recomputing the distance each time and
     stopping as soon as the participant falls below threshold. The count of items needed is a graded
     measure of how single-item-driven the flag is. Marginal z-scores do not change as items are
     neutralized, so the candidate set is fixed at step 1 and the order only affects the count.
  3. Also report the batch result: the distance with ALL individually-extreme items neutralized.

  NEUTRALIZE = "mean"       set the item to the cohort mean, removing the elevation entirely
             = "winsorize"  cap the item at its own 95th percentile, removing only the extremeness
  The mean rule is the stronger test. Both are reported so the answer does not hinge on the choice.

  Neutralizing can INCREASE the distance -- setting an item to average can leave a more discordant
  profile than the original. Those participants never drop out, which is the signature being hunted,
  and they are counted separately rather than treated as a failure.

FIXED YARDSTICK. The projection, the covariate coefficients, the cohort score mean, the Ledoit-Wolf
precision and the flagging threshold are all estimated once on the unmodified cohort and held fixed.

Run:  python3 joint_only.py
"""
import json
import os

import numpy as np
import pandas as pd
from sklearn.covariance import LedoitWolf

HERE = os.path.dirname(os.path.abspath(__file__))
BASIS_DIR = os.path.join(HERE, "reference_basis")
EMB_DIR = os.path.join(HERE, "item_embeddings")
EMB_PREFIX = "core151_items"
RESPONSES_CSV = "/home/jupyter/responses_out/responses_k0.csv"
WEIGHTS_CSV = "/home/jupyter/responses_out/weights_k0.csv"
SCALES_CSV = os.path.join(HERE, "scales_prism.csv")
ITEMS_CSV = os.path.join(HERE, "core_items_151.csv")
CONSTRUCTS_CSV = os.path.join(HERE, "construct_map_151.csv")
OUT_DIR = os.path.join(HERE, "screen_out")
ID_COL = "id"
COVARIATES = ["age", "sex"]
PCTILE = 95.0
CONSENSUS_MIN_ENC = 4          # "still an outlier" = above threshold under at least this many encoders
CTRL_A_SAMPLE = 3000           # unflagged participants used for the distance-inflation control
CTRL_B_REPEATS = 5             # random-item draws per participant for the same-size control

ENCODERS = ["gte-large-en-v1.5", "bge-m3", "bge-large-en-v1.5",
            "all-mpnet-base-v2", "e5-large-v2", "qwen3-embedding-0.6b"]


def _rowmedian(needed):
    """Median items-to-drop per participant, NaN where the flag never dropped under any encoder."""
    m = np.where(needed > 0, needed, np.nan).astype(float)
    out = np.full(m.shape[0], np.nan)
    ok = np.isfinite(m).any(1)
    out[ok] = np.nanmedian(m[ok], axis=1)
    return out


def keep_columns(Z, band):
    anchored = {c for c in range(Z.shape[1]) if (np.abs(Z[:, c]) >= band[c]).any()}
    modal = {int(np.argmax(np.abs(Z[i]))) for i in range(Z.shape[0])}
    return sorted(anchored | modal)


def main():
    items = pd.read_csv(ITEMS_CSV)["item"].astype(str).tolist()
    scales = pd.read_csv(SCALES_CSV).set_index("item").reindex(items)
    cmap = pd.read_csv(CONSTRUCTS_CSV).set_index("item")["construct"]

    R = pd.read_csv(RESPONSES_CSV)
    R[ID_COL] = R[ID_COL].astype(str)
    R = R.set_index(ID_COL)
    raw = R[items].values.astype(float)
    lo, hi = scales["min"].values.astype(float), scales["max"].values.astype(float)
    rev = scales["reverse"].astype(str).str.lower().eq("true").values
    X = np.clip(2.0 * (raw - lo) / (hi - lo) - 1.0, -1.0, 1.0)
    X[:, rev] *= -1.0

    w = (pd.read_csv(WEIGHTS_CSV).iloc[:, 0].values.astype(float)
         if WEIGHTS_CSV and os.path.exists(WEIGHTS_CSV) else np.ones(len(R)))
    A = np.column_stack([np.ones(len(R))] + ([R[COVARIATES].values.astype(float)] if COVARIATES else []))

    sel = pd.read_csv(os.path.join(OUT_DIR, "consensus_outliers.csv"))
    sel[ID_COL] = sel[ID_COL].astype(str)
    out_ids = [i for i in R.index if i in set(sel.loc[sel["flagged_by_all"].astype(bool), ID_COL])]
    pos = {pid: j for j, pid in enumerate(R.index)}
    oidx = np.array([pos[i] for i in out_ids])
    n_out = len(oidx)

    # which items is each participant individually extreme on (strict, against the item's own p95)
    Z = (X - X.mean(0)) / X.std(0, ddof=0)
    thr_item = np.percentile(Z, PCTILE, axis=0)
    X_mean = X.mean(0)
    X_cap = np.percentile(X, PCTILE, axis=0)
    print(f"cohort {len(R):,} | consensus outliers {n_out}")
    ext = Z[oidx] > thr_item
    print(f"individually extreme items per outlier: median {int(np.median(ext.sum(1)))}, "
          f"max {int(ext.sum(1).max())}, none for {int((ext.sum(1) == 0).sum())} of them\n")

    rng0 = np.random.default_rng(0)
    non_out = np.setdiff1d(np.arange(len(R)), oidx)
    ctrl_idx = rng0.choice(non_out, size=min(CTRL_A_SAMPLE, len(non_out)), replace=False)
    ctrlA, ctrlB = {"mean": [], "winsorize": []}, {"mean": [], "winsorize": []}

    results = {}
    for mode in ("mean", "winsorize"):
        repl = X_mean if mode == "mean" else X_cap
        survive = np.zeros((n_out, len(ENCODERS)), bool)
        needed = np.full((n_out, len(ENCODERS)), -1, int)      # -1 = never dropped below threshold
        rose = np.zeros((n_out, len(ENCODERS)), bool)
        for e, enc in enumerate(ENCODERS):
            b = np.load(os.path.join(BASIS_DIR, f"{enc}.npz"), allow_pickle=True)
            W, mu, sigma, mu_s, band = b["W"], b["mu"], b["sigma"], b["mu_scores"], b["band"]
            em = np.load(os.path.join(EMB_DIR, f"{EMB_PREFIX}_{enc}.npz"), allow_pickle=True)
            p = {str(x): j for j, x in enumerate(em["items"])}
            E = np.asarray(em["vectors"], float)[[p[it] for it in items]]
            P = (E - mu) @ W.T
            Pk = P[:, keep_columns((P - mu_s) / sigma, band)]

            Y = X @ Pk
            sw = np.sqrt(w)[:, None]
            beta, *_ = np.linalg.lstsq(A * sw, Y * sw, rcond=None)   # fixed covariate fit
            Yr = Y - A @ beta
            ybar = np.average(Yr, axis=0, weights=w)
            lw = LedoitWolf().fit(Yr - ybar)
            Om = lw.precision_
            d2 = np.einsum("ij,ij->i", Yr - ybar, (Yr - ybar) @ Om)
            thr = np.percentile(d2, PCTILE)

            for r, i in enumerate(oidx):
                order = np.argsort(-Z[i])
                cand = [j for j in order if Z[i, j] > thr_item[j]]
                xi = X[i].copy()
                # run every candidate: the distance is NOT monotone in the number neutralized, so we
                # record the FIRST step it drops below threshold but still evaluate survival on the
                # fully neutralized profile
                for step, j in enumerate(cand, start=1):
                    xi[j] = repl[j]
                    y = xi @ Pk - A[i] @ beta - ybar
                    if needed[r, e] < 0 and float(y @ Om @ y) <= thr:
                        needed[r, e] = step
                y = xi @ Pk - A[i] @ beta - ybar
                dd = float(y @ Om @ y)
                survive[r, e] = dd > thr                      # after ALL of them are neutralized
                rose[r, e] = dd > d2[i] * (1.0 + 1e-9)   # tolerance: no items neutralized must read as no change

            def batch_d2(Xm, rows):
                y = Xm @ Pk - A[rows] @ beta - ybar
                return np.einsum("ij,ij->i", y, y @ Om)

            # ---- CONTROL A: does the operation inflate distance on its own? ----
            # Apply the identical neutralization to participants who were NOT flagged. Setting an
            # item to z = 0 puts the profile somewhere no real respondent sits, and an off-manifold
            # point can be far from the centroid for that reason alone. If the operation were
            # distance-neutral almost no unflagged participant would cross the threshold.
            Xa = X[ctrl_idx].copy()
            ea = Z[ctrl_idx] > thr_item
            Xa[ea] = np.broadcast_to(repl, Xa.shape)[ea]
            da = batch_d2(Xa, ctrl_idx)
            below = d2[ctrl_idx] <= thr          # a non-consensus participant can still clear ONE
            crossed = below & (da > thr)         # encoder's threshold, so count only genuine crossings
            ctrlA[mode].append((float(crossed.sum() / max(below.sum(), 1)),
                                float((da > d2[ctrl_idx]).mean())))

            # ---- CONTROL B: is it THESE items, or just removing this many items? ----
            # Neutralize a random set of the same size drawn from all items. Survival here is the
            # baseline the extreme-item result has to beat.
            surv_b = []
            for rep in range(CTRL_B_REPEATS):
                rng = np.random.default_rng(1000 * rep + e)
                Xb = X[oidx].copy()
                for r in range(n_out):
                    k = int(ext[r].sum())
                    if k:
                        pick = rng.choice(X.shape[1], size=k, replace=False)
                        Xb[r, pick] = repl[pick]
                db = batch_d2(Xb, oidx)
                surv_b.append(float((db > thr).mean()))
            ctrlB[mode].append(float(np.mean(surv_b)))
        n_joint = (survive.sum(1) >= CONSENSUS_MIN_ENC)
        results[mode] = {"survive": survive, "needed": needed, "rose": rose, "n_joint": n_joint}

        print(f"=== neutralize by setting the item to the cohort {'mean' if mode=='mean' else '95th pctile'} ===")
        print(f"  still above threshold after ALL individually-extreme items are neutralized:")
        print(f"    under >= {CONSENSUS_MIN_ENC} of {len(ENCODERS)} encoders : "
              f"{int(n_joint.sum())} of {n_out}")
        print(f"    under all {len(ENCODERS)}                  : "
              f"{int((survive.sum(1) == len(ENCODERS)).sum())} of {n_out}")
        print(f"    under none                        : {int((survive.sum(1) == 0).sum())} of {n_out}")
        med = np.where(needed > 0, needed, np.nan)
        if np.isfinite(med).any():
            print(f"  items that had to be neutralized before the flag dropped: median "
                  f"{np.nanmedian(med):.1f} (over person-encoder pairs where it dropped at all)")
        else:
            print("  no participant-encoder pair dropped below threshold at any step")
        print(f"  distance INCREASED after neutralization for "
              f"{int(rose.any(1).sum())} of {n_out} under at least one encoder")
        ca = np.array(ctrlA[mode])
        print(f"  CONTROL A, same operation on {len(ctrl_idx):,} UNFLAGGED participants: "
              f"{100*ca[:, 0].mean():.1f}% of those below threshold CROSS it "
              f"(range {100*ca[:, 0].min():.1f}-{100*ca[:, 0].max():.1f}% across encoders), "
              f"{100*ca[:, 1].mean():.0f}% move further out")
        cb = np.mean(ctrlB[mode])
        print(f"  CONTROL B, neutralizing the same NUMBER of items chosen at random: "
              f"{100*cb:.1f}% of outliers stay above threshold, against "
              f"{100*survive.mean():.1f}% when the individually-extreme items are the ones removed\n")

    # who they are, under the stronger rule
    sur = results["mean"]["survive"]
    jo = results["mean"]["n_joint"]
    if jo.sum():
        print(f"the {int(jo.sum())} joint cases under the mean rule (id, encoders surviving, "
              f"individually-extreme items):")
        for r in np.where(jo)[0]:
            print(f"  {out_ids[r]:>10s}  {int(sur[r].sum())}/{len(ENCODERS)}  {int(ext[r].sum())}")

    os.makedirs(OUT_DIR, exist_ok=True)
    pd.DataFrame({ID_COL: out_ids,
                  "n_extreme_items": ext.sum(1),
                  "survives_mean_n_enc": results["mean"]["survive"].sum(1),
                  "survives_winsor_n_enc": results["winsorize"]["survive"].sum(1),
                  "joint_mean": results["mean"]["n_joint"],
                  "joint_winsor": results["winsorize"]["n_joint"],
                  "median_items_to_drop": _rowmedian(results["mean"]["needed"]),
                  }).to_csv(os.path.join(OUT_DIR, "joint_only.csv"), index=False)
    # the full person x encoder matrix behind median_items_to_drop, saved because a median of
    # medians cannot be re-derived from the row summary and the two directions of the median
    # (over encoders, over participants) are different numbers. 0 = never dropped, not zero items.
    pd.DataFrame(results["mean"]["needed"], index=pd.Index(out_ids, name=ID_COL),
                 columns=ENCODERS).reset_index().to_csv(
        os.path.join(OUT_DIR, "joint_only_needed_mean.csv"), index=False)
    json.dump({"n_outliers": n_out,
               "joint_mean_rule": int(results["mean"]["n_joint"].sum()),
               "joint_winsorize_rule": int(results["winsorize"]["n_joint"].sum()),
               "consensus_min_encoders": CONSENSUS_MIN_ENC},
              open(os.path.join(OUT_DIR, "joint_only_summary.json"), "w"), indent=2)
    print(f"\nwrote -> {OUT_DIR}/joint_only.csv")


if __name__ == "__main__":
    main()
```
