## Supplemental code for "Scoring respondents on a semantic frame of reference identifies a group that is burdened across many questionnaires and reaches care less easily": joint_profile.py

```
#!/usr/bin/env python3
"""joint_profile.py -- are the joint survivors the same people the cutoffs under-catch?

Merges what the earlier scripts wrote. Recomputes nothing.

  joint_only.csv                    who stays above threshold once every item they are individually
                                    extreme on is neutralized  (joint_only.py)
  cutoff_flags.csv                  the eight validated criteria and each participant's breadth
                                    (cutoffs.py)
  outlier_leaders_outliers.csv      each participant's leading construct and its share
                                    (outlier_profile.py)

If the joint survivors carry lower clinical breadth and are led by the constructs with no validated
cutoff, then the joint result and the breadth gradient are one finding rather than two. If they look
the same as everyone else on those measures, they are a separate property of the distance and should
be reported separately.

Run:  python3 joint_profile.py
"""
import os

import numpy as np
import pandas as pd

HERE = os.path.dirname(os.path.abspath(__file__))
OUT_DIR = os.path.join(HERE, "screen_out")
ID_COL = "id"
MIN_LEADS = 8            # constructs leading fewer people than this are pooled out of the breakdown
CUTOFF_CONSTRUCTS = {    # constructs an instrument in this battery screens with a validated cutoff
    "Depression", "Generalized anxiety", "Psychosis", "Adult ADHD",
    "Suicidality & self-harm", "Childhood adversity", "Self-rated health",
}


def wilson(k, n, z=1.959963985):
    if n == 0:
        return float("nan"), float("nan")
    p, d = k / n, 1 + z * z / n
    c = (p + z * z / (2 * n)) / d
    h = z * np.sqrt(p * (1 - p) / n + z * z / (4 * n * n)) / d
    return 100 * (c - h), 100 * (c + h)


def main():
    J = pd.read_csv(os.path.join(OUT_DIR, "joint_only.csv"))
    C = pd.read_csv(os.path.join(OUT_DIR, "cutoff_flags.csv"))
    L = pd.read_csv(os.path.join(OUT_DIR, "outlier_leaders_outliers.csv"))
    for d in (J, C, L):
        d[ID_COL] = d[ID_COL].astype(str)

    M = J.merge(C[[ID_COL, "breadth"]], on=ID_COL).merge(
        L[[ID_COL, "leader", "lead_share", "agree"]], on=ID_COL, how="left")
    assert len(M) == len(J), "merge lost rows -- id types or a missing input"
    M["joint"] = M["joint_mean"].astype(bool)
    n_j, n_o = int(M["joint"].sum()), int((~M["joint"]).sum())
    print(f"{len(M)} consensus outliers | joint survivors {n_j} | the rest {n_o}\n")

    # ---- clinical breadth ----
    print("clinical breadth (number of the eight validated criteria met)")
    for name, sub in (("joint survivors", M[M["joint"]]), ("the rest", M[~M["joint"]])):
        b = sub["breadth"]
        k0, k5 = int((b == 0).sum()), int((b >= 5).sum())
        lo0, hi0 = wilson(k0, len(b))
        lo5, hi5 = wilson(k5, len(b))
        print(f"  {name:18s} n={len(b):3d}  mean {b.mean():.2f}  meeting none {k0:3d} "
              f"({100*k0/len(b):4.1f}%, CI {lo0:.1f}-{hi0:.1f})  meeting >=5 {k5:3d} "
              f"({100*k5/len(b):4.1f}%, CI {lo5:.1f}-{hi5:.1f})")
    diff = M[M["joint"]]["breadth"].mean() - M[~M["joint"]]["breadth"].mean()
    print(f"  difference in mean breadth: {diff:+.2f}")

    # ---- how concentrated the flag is, and how many items were neutralized ----
    print("\nother measures")
    for col, lab in (("n_extreme_items", "individually-extreme items"),
                     ("lead_share", "share carried by the leading construct"),
                     ("agree", "encoders agreeing on the leader")):
        a, b = M[M["joint"]][col], M[~M["joint"]][col]
        sc = 100 if col == "lead_share" else 1
        print(f"  {lab:38s} joint {sc*a.mean():6.2f}   rest {sc*b.mean():6.2f}")

    # ---- leading construct ----
    print("\nleading construct, and how often it survives neutralization")
    g = M.groupby("leader")
    T = pd.DataFrame({
        "n": g.size(),
        "n_joint": g["joint"].sum(),
        "pct_joint": (100 * g["joint"].mean()).round(1),
        "mean_breadth": g["breadth"].mean().round(2),
        "has_cutoff": [c in CUTOFF_CONSTRUCTS for c in g.size().index],
    }).sort_values("n", ascending=False)
    print(T[T["n"] >= MIN_LEADS].to_string())
    small = T[T["n"] < MIN_LEADS]
    if len(small):
        print(f"  ({int(small['n'].sum())} further participants across {len(small)} constructs "
              f"leading fewer than {MIN_LEADS} each)")

    big = T[T["n"] >= MIN_LEADS]
    for flag, lab in ((True, "constructs WITH a validated cutoff"),
                      (False, "constructs WITHOUT one")):
        s = big[big["has_cutoff"] == flag]
        if len(s):
            n, j = int(s["n"].sum()), int(s["n_joint"].sum())
            print(f"  {lab:36s} {j:3d} of {n:3d} joint ({100*j/n:.1f}%), "
                  f"mean breadth {np.average(s['mean_breadth'], weights=s['n']):.2f}")

    M.to_csv(os.path.join(OUT_DIR, "joint_profile.csv"), index=False)
    print(f"\nwrote -> {OUT_DIR}/joint_profile.csv")


if __name__ == "__main__":
    main()
```
