## Supplemental code for "Scoring respondents on a semantic frame of reference identifies a group that is burdened across many questionnaires and reaches care less easily": outlier_profile.py

```
#!/usr/bin/env python3
"""outlier_profile.py -- who is led by what, and how that relates to clinical breadth.

Reads what attribution.py and cutoffs.py already wrote. Nothing is recomputed from the responses,
and no construct is grouped into a domain or family -- every row is a documented construct.

Two questions the group mean cannot answer:

  1. HOW MANY PEOPLE does each construct LEAD? A construct can top the group mean by carrying a
     moderate share of many people, or a large share of a few. These are different findings.

  2. Do the people led by a construct meet FEWER validated cutoffs? Breadth is the number of the
     eight criteria a person meets. If the constructs with no cutoff of their own lead people with
     low breadth, that is the mechanism behind the screen surfacing what instruments do not.

  python3 outlier_profile.py                 # the consensus outliers
  python3 outlier_profile.py --tag invisible # whichever subset attribution.py was last run on
"""
import os
import sys

import numpy as np
import pandas as pd

HERE = os.path.dirname(os.path.abspath(__file__))
OUT_DIR = os.path.join(HERE, "screen_out")
CONSTRUCTS_CSV = os.path.join(HERE, "construct_map_151.csv")
ID_COL = "id"
MIN_LEADS = 3          # constructs leading fewer people than this are pooled into a footer line


def main():
    tag = "outliers"
    if "--tag" in sys.argv:
        tag = sys.argv[sys.argv.index("--tag") + 1]

    shares_path = os.path.join(OUT_DIR, f"attribution_item_shares_{tag}.csv")
    assert os.path.exists(shares_path), f"{shares_path} not found -- run attribution.py first"
    sh = pd.read_csv(shares_path, index_col=[0, 1])
    sh.index = sh.index.set_names(["encoder", ID_COL])
    sh.index = sh.index.set_levels(sh.index.levels[1].astype(str), level=1)

    cmap = pd.read_csv(CONSTRUCTS_CSV).set_index("item")["construct"]
    con = cmap.reindex(sh.columns).fillna("unlabelled").values
    C = sh.T.groupby(con).sum().T                       # (encoder, id) x construct

    encoders = list(C.index.get_level_values("encoder").unique())
    ids = list(C.index.get_level_values(ID_COL).unique())
    n = len(ids)

    # per person: the leading construct under each encoder, and the mean share of the winner
    lead_by_enc = C.groupby(level=ID_COL).apply(lambda g: g.idxmax(axis=1))
    per_enc = C.reset_index().pivot_table(index=ID_COL, columns="encoder", values=C.columns.tolist())
    mean_share = C.groupby(level=ID_COL).mean()          # ids x constructs
    leader = mean_share.idxmax(axis=1)
    lead_share = mean_share.max(axis=1)
    agree = pd.Series(
        {pid: (C.xs(pid, level=ID_COL).idxmax(axis=1) == leader[pid]).sum() for pid in ids})

    P = pd.DataFrame({"leader": leader, "lead_share": lead_share, "agree": agree})

    cut_path = os.path.join(OUT_DIR, "cutoff_flags.csv")
    have_cut = os.path.exists(cut_path)
    if have_cut:
        cf = pd.read_csv(cut_path)
        cf[ID_COL] = cf[ID_COL].astype(str)
        P = P.join(cf.set_index(ID_COL)["breadth"])
    else:
        print("(cutoff_flags.csv not found -- breadth columns omitted; run cutoffs.py for them)\n")
        P["breadth"] = np.nan

    print(f"{n} respondents, {len(encoders)} encoders, {C.shape[1]} documented constructs\n")
    print(f"leading construct agrees across >= 4 of {len(encoders)} encoders for "
          f"{int((P['agree'] >= 4).sum())} of {n} ({100*(P['agree'] >= 4).mean():.0f}%)")
    print(f"mean share carried by a person's leading construct: {100*P['lead_share'].mean():.1f}%\n")

    g = P.groupby("leader")
    T = pd.DataFrame({
        "n_leads": g.size(),
        "pct_of_group": (100 * g.size() / n).round(1),
        "mean_agree": g["agree"].mean().round(1),
        "mean_lead_share": (100 * g["lead_share"].mean()).round(1),
        "group_mean_share": (100 * mean_share.mean()).reindex(g.size().index).round(1),
        "mean_breadth": g["breadth"].mean().round(2),
        "pct_breadth_0": (100 * g["breadth"].apply(lambda s: (s == 0).mean())).round(1),
        "pct_breadth_ge5": (100 * g["breadth"].apply(lambda s: (s >= 5).mean())).round(1),
    }).sort_values("n_leads", ascending=False)

    big = T[T["n_leads"] >= MIN_LEADS]
    print("Leading construct: how many people, and what they meet clinically")
    print(big.to_string())
    small = T[T["n_leads"] < MIN_LEADS]
    if len(small):
        print(f"\n  {int(small['n_leads'].sum())} further people are led by {len(small)} constructs "
              f"leading fewer than {MIN_LEADS} each: " + ", ".join(small.index))

    if have_cut and P["breadth"].notna().any():
        overall = P["breadth"].mean()
        print(f"\nbreadth over all {n}: mean {overall:.2f}")
        lo = big.sort_values("mean_breadth").head(5)
        hi = big.sort_values("mean_breadth", ascending=False).head(5)
        print(f"  lowest  mean breadth: " + ", ".join(f"{i} {r.mean_breadth:.2f}" for i, r in lo.iterrows()))
        print(f"  highest mean breadth: " + ", ".join(f"{i} {r.mean_breadth:.2f}" for i, r in hi.iterrows()))
        rho = P[["lead_share", "breadth"]].corr(method="spearman").iloc[0, 1]
        print(f"  Spearman(leading-construct share, breadth) = {rho:+.2f}")

    T.to_csv(os.path.join(OUT_DIR, f"outlier_profile_{tag}.csv"))
    P.rename_axis(ID_COL).reset_index().to_csv(
        os.path.join(OUT_DIR, f"outlier_leaders_{tag}.csv"), index=False)
    print(f"\nwrote -> {OUT_DIR}/outlier_profile_{tag}.csv and outlier_leaders_{tag}.csv")


if __name__ == "__main__":
    main()
```
