## Supplemental code for "Scoring respondents on a semantic frame of reference identifies a group that is burdened across many questionnaires and reaches care less easily": person_report.py

```
#!/usr/bin/env python3
"""person_report.py -- one respondent's attribution, encoder by encoder.

Reads the item shares attribution.py already wrote, so nothing is recomputed. Answers the question
a group table cannot: is a construct carrying this person under EVERY encoder, or under one or two?

  python3 person_report.py 1078710
  python3 person_report.py 1078710 --tag outliers      # if the run was not --invisible
  python3 person_report.py 1078710 --construct "Interpersonal & sexual violence"

Shares are a_i / T^2 summed within each item's documented construct, exact and signed. A negative
share is a suppressor -- that content pulls the person's distance down.
"""
import os
import sys

import pandas as pd

HERE = os.path.dirname(os.path.abspath(__file__))
OUT_DIR = os.path.join(HERE, "screen_out")
CONSTRUCTS_CSV = os.path.join(HERE, "construct_map_151.csv")
TOP_CONSTRUCTS = 8
TOP_ITEMS = 10


def main():
    args = [a for a in sys.argv[1:] if not a.startswith("--")]
    assert args, "give a respondent id, e.g. python3 person_report.py 1078710"
    pid = str(args[0])
    tag = "invisible"
    if "--tag" in sys.argv:
        tag = sys.argv[sys.argv.index("--tag") + 1]
    focus = None
    if "--construct" in sys.argv:
        focus = sys.argv[sys.argv.index("--construct") + 1]

    path = os.path.join(OUT_DIR, f"attribution_item_shares_{tag}.csv")
    assert os.path.exists(path), f"{path} not found -- run attribution.py first"
    sh = pd.read_csv(path, index_col=[0, 1])
    sh.index = sh.index.set_names(["encoder", "id"])
    sh.index = sh.index.set_levels(sh.index.levels[1].astype(str), level=1)
    assert pid in set(sh.index.get_level_values("id")), f"{pid} is not in {os.path.basename(path)}"

    cmap = pd.read_csv(CONSTRUCTS_CSV).set_index("item")["construct"]
    rows = sh.xs(pid, level="id")                       # encoders x items
    con = cmap.reindex(rows.columns).fillna("unlabelled").values
    C = rows.T.groupby(con).sum().T * 100.0             # encoders x constructs, per cent of T^2

    order = C.mean(0).sort_values(ascending=False)
    keep = list(order.index[:TOP_CONSTRUCTS])
    if focus and focus not in keep:
        keep.append(focus)

    print(f"respondent {pid}   ({len(rows)} encoders, shares as % of that encoder's T^2)\n")
    tbl = C[keep].round(1)
    tbl.index = [i.split("-")[0] for i in tbl.index]
    print(tbl.to_string())

    print(f"\n{'construct':34s} {'mean':>6s} {'min':>6s} {'max':>6s} {'rank across encoders':>22s}")
    ranks = C.rank(axis=1, ascending=False)
    for c in keep:
        r = ranks[c].astype(int).tolist()
        print(f"  {c:32s} {C[c].mean():6.1f} {C[c].min():6.1f} {C[c].max():6.1f} "
              f"{str(r):>22s}")
    print("\n  rank 1 = the construct carrying the most of that encoder's distance for this person.")

    if focus:
        r = ranks[focus].astype(int)
        print(f"\n{focus}: leads under {int((r == 1).sum())} of {len(r)} encoders, "
              f"top-3 under {int((r <= 3).sum())} of {len(r)}, "
              f"share {C[focus].min():.1f}%-{C[focus].max():.1f}%")

    it = rows.mean(0).sort_values(ascending=False).head(TOP_ITEMS) * 100.0
    print(f"\ntop {TOP_ITEMS} items for this respondent (mean over encoders)")
    for name, v in it.items():
        print(f"  {v:6.1f}%  {name:34s} {cmap.get(name, '')}")


if __name__ == "__main__":
    main()
```
