## Supplemental code for "Scoring respondents on a semantic frame of reference identifies a group that is burdened across many questionnaires and reaches care less easily": portability_sweep.py

```
#!/usr/bin/env python
"""How stable is a response-based grouping, as a function of how many people answered?

WORKBENCH ONLY -- needs the participant response matrix, which cannot leave the Researcher Workbench.

This replaces the transcribed values that were hardcoded in `examples/allofus/fig_portability.py`
(a sweep run in chat, never saved). It writes a CSV that the figure generator reads, so the number the
manuscript quotes has a saved artefact behind it.

Procedure (matches Methods, "Response-based baseline and portability"):
  for each subsample size n
    repeat REPEATS times
      draw two DISJOINT random subsamples of n respondents
      fit the structure independently on each  (PCA, and factor analysis with a varimax rotation)
      match the two fits' components one-to-one to maximize total |loading correlation| (Hungarian)
      record the mean matched absolute loading correlation
  report the mean over repeats

Responses are prepared exactly as the screen prepares them (screen_battery.py): rescaled to [-1, 1] on
each item's own published range, then reverse-coded where the scale file marks the item reverse-keyed.
The text-derived basis does not enter the sweep -- it is identical at every n and defined at n = 0.

Out: screen_out/portability_sweep.csv  (columns: method, n, repeat, matched_abs_corr)
Run on the Workbench:
    python portability_sweep.py
"""
import os
import numpy as np
import pandas as pd
from scipy.optimize import linear_sum_assignment
from sklearn.decomposition import PCA, FactorAnalysis

HERE = os.path.dirname(os.path.abspath(__file__))
OUT_DIR = os.path.join(HERE, "screen_out")
ITEMS_CSV = os.path.join(HERE, "core_items_151.csv")
SCALES_CSV = os.path.join(HERE, "scales_prism.csv")
RESPONSES_CSV = "/home/jupyter/responses_out/responses_k0.csv"
ID_COL = "person_id"

SIZES = [100, 250, 500, 1000, 2000, 5000, 11000]
REPEATS = 3
K = 26                 # components per fit, held fixed across the sweep
SEED = 0


def load_X():
    items = pd.read_csv(ITEMS_CSV)["item"].astype(str).tolist()
    scales = pd.read_csv(SCALES_CSV).set_index("item").reindex(items)
    resp = pd.read_csv(RESPONSES_CSV).set_index(ID_COL)
    raw = resp[items].values.astype(float)
    assert not np.isnan(raw).any(), "missing response cells: use the k0 (complete) file"
    lo, hi = scales["min"].values.astype(float), scales["max"].values.astype(float)
    rev = scales["reverse"].astype(str).str.lower().eq("true").values
    X = np.clip(2.0 * (raw - lo) / (hi - lo) - 1.0, -1.0, 1.0)
    X[:, rev] *= -1.0
    return X


def loadings(model, Xs, method):
    if method == "pca":
        return PCA(n_components=K, random_state=SEED).fit(Xs).components_.T   # items x k
    fa = FactorAnalysis(n_components=K, rotation="varimax", random_state=SEED).fit(Xs)
    return fa.components_.T


def matched_abs_corr(A, B):
    """Mean |correlation| of loading vectors after the best one-to-one component matching."""
    A = (A - A.mean(0)) / (A.std(0) + 1e-12)
    B = (B - B.mean(0)) / (B.std(0) + 1e-12)
    C = np.abs(A.T @ B) / A.shape[0]
    r, c = linear_sum_assignment(-C)
    return float(C[r, c].mean())


def main():
    os.makedirs(OUT_DIR, exist_ok=True)
    X = load_X()
    N = X.shape[0]
    rng = np.random.default_rng(SEED)
    rows = []
    for n in SIZES:
        if 2 * n > N:
            print(f"skipping n={n}: needs {2*n} respondents, cohort has {N}")
            continue
        for rep in range(REPEATS):
            idx = rng.permutation(N)
            a, b = idx[:n], idx[n:2 * n]
            for method in ("pca", "fa"):
                r = matched_abs_corr(loadings(None, X[a], method), loadings(None, X[b], method))
                rows.append(dict(method=method, n=n, repeat=rep, matched_abs_corr=r))
                print(f"{method:3s} n={n:6d} rep={rep} matched |r| = {r:.4f}", flush=True)

    df = pd.DataFrame(rows)
    p = os.path.join(OUT_DIR, "portability_sweep.csv")
    df.to_csv(p, index=False)
    print("\nmean over repeats:")
    print(df.groupby(["method", "n"])["matched_abs_corr"].mean().unstack(0).to_string(
        float_format=lambda v: f"{v:.3f}"))
    print(f"\nwrote {p}")


if __name__ == "__main__":
    main()
```
