## Supplemental code for "Scoring respondents on a semantic frame of reference identifies a group that is burdened across many questionnaires and reaches care less easily": README.md

# All of Us screening on the fixed reference basis

Self-contained pipeline for placing the All of Us battery on the pool reference basis, screening the
cohort, and producing every number in the manuscript's screening sections. Everything here is
standalone — numpy, pandas, scikit-learn, scipy, statsmodels, and BigQuery only for the EHR step. No
imports from the `ross_proj` repo, so the folder can be uploaded to the Workbench as it is.

Built 2026-08-20. Supersedes the `outputs_dense/` pipeline and the N=584 screen it produced.

## Run order

| # | Script | Where | Needs | Writes |
|---|---|---|---|---|
| 0 | `build_responses_v2.ipynb` | Workbench | the CDR | `/home/jupyter/responses_out/responses_{k0,k3,k5,all}.csv`, `weights_*.csv` |
| 1 | `screen_battery.py` | Workbench | `responses_k0.csv`, `reference_basis/`, `item_embeddings/` | `screen_out/scores_<enc>.csv`, `kept_<enc>.csv`, `consensus_outliers.csv`, `screen_summary.csv`, `run_config.json` |
| 2 | `cutoffs.py` | Workbench | 1 | `cutoff_flags.csv`, `cutoff_summary.csv`, `breadth_table.csv` |
| 3 | `attribution.py` | Workbench | 1 | `attribution_{constructs,per_person,item_shares}_<tag>.csv`, `attribution_summary_<tag>.json` |
| 4 | `outlier_profile.py` | Workbench | 2, 3 | `outlier_profile_<tag>.csv`, `outlier_leaders_<tag>.csv` |
| 5 | `construct_distribution.py` | Workbench | 3 | `construct_distribution_<tag>.csv` |
| 5b | `construct_concentration.py` | Workbench | 3 | `construct_concentration_<tag>.csv` |
| 5c | `fig_construct_ranks.py` | Workbench | 3 | `construct_ranks_<tag>.csv`, `fig_construct_ranks_<tag>.png` |
| 6 | `separability.py` | Workbench | 1 | `separability_{reach,diagonal}.csv`, `separability_summary.json` |
| 7 | `joint_only.py` | Workbench | 1 | `joint_only.csv`, `joint_only_summary.json` |
| 8 | `joint_profile.py` | Workbench | 2, 4, 7 | `joint_profile.csv` |
| 9 | `response_style.py` | Workbench | 1 | `response_style.json` |
| 10 | `underserved.py` | Workbench | 1 + BigQuery | `underserved_stats.json`, `fig_ehr_gradient.png`, `fig_ehr_visits_supp.png` |
| 11 | `fig_screen.py` | Workbench | 1, 2 | `fig_screen.png` |
| 12 | `portability_sweep.py` | Workbench | responses only | `portability_sweep.csv` |
| — | `person_report.py <id>` | anywhere | 3 | prints one respondent's attribution, encoder by encoder |

Everything lands in `screen_out/`. **Copy figures into `manuscript_nhb/figures/` by hand** — nothing
syncs them, and stale copies there have caused wrong figures to ship twice.

`reference_basis/` and `item_embeddings/` are built by
`../examples/allofus/build_reference_basis_package.py` and shipped with the folder, so the Workbench
needs no encoder installed and no access to this repo.

## The keep rule

A pool component is scored for a battery when **either** at least one battery item's standardized
loading on it clears that component's own pool core-item band (`band` in `reference_basis/<enc>.npz`),
**or** it is the modal component of some battery item. Both conditions are per item against
quantities fixed on the pool, so the kept set is battery-independent and monotone.

This replaced the union of item fixed-point cores, which had no rank discipline: on the 35-item HRS
battery it kept 34–43 columns, the score matrix came out rank-deficient, and the screen collapsed onto
plain Mahalanobis on the raw responses (r 0.989–0.998). All of Us keeps 43/44/46/41/49/41 of 55/50/54/49/55/49.

Implementation of record is `_common.keep_columns` in `../examples/allofus/`, mirrored in
`screen_battery.py` so this folder stays standalone. **If one changes, change both.**

## Results, as of 2026-08-20

- **Screen** — 510 consensus outliers of 22,332. Mean pairwise Jaccard of the six flagged sets 0.55
  against 0.026 expected, holding from the 90th to the 99th percentile.
- **Cutoffs** — cohort rates reproduce the published table exactly (17.7 / 13.8 / 23.1 / 19.5 / 7.4 /
  37.2 / 11.4 / 13.8, any-of-eight 58.4). Outliers 99.0% meet at least one, mean breadth 4.99 against
  1.44, 62% meet five or more. **Five meet none**, all carried by discrimination.
  ACE is **≥4 of 7 categories** — the score counts categories, not questions, so alcohol and drugs are
  one category and the three sexual-abuse items are one. Do not "fix" this to 8.
- **Attribution** — composition diffuse, nothing above 10%. Integration 40–45%. The leading construct
  agrees across ≥4 of 6 encoders for **82%** of the group (419 of 510), so per-person attribution is supported here
  (it is not on the 35-item HRS battery).
- **Leading ≠ carrying** — food insecurity leads 107 but reaches a fifth of the distance for 27,
  psychosis leads 138 and reaches a fifth for 106. Childhood adversity reaches a fifth for nobody.
- **Separability** — 0 of 510 are beyond the reach of both a single item and a single construct. The
  diagonal distance ranks differently though: ρ 0.74–0.80, top-5% Jaccard 0.36, 71–75% of the outliers
  in the diagonal tail.
- **Joint-only** — neutralizing every individually-extreme item leaves **190 of 510** above threshold
  under ≥4 encoders, 87 under all six. Controls pass: the operation moves 0.1% of unflagged
  participants across, and random same-size removal leaves 90.4% against 41.1%.
- **Under-served** — barriers 82% vs 44%, cost 67% vs 28%, care avoidance 32% vs 9%. ED share 2.4% vs
  1.2%, β +0.054 holding total visits fixed. Raw visit counts fall but collapse under window
  adjustment (p 0.17) and per observation-year (p 0.42), so they stay in the supplement.
- **Response style** — content extremity ρ 0.77 but recovers only 244 of 510 by itself, endpoint use
  ρ −0.35 and recovers 7. The screen is not a severity sum and not a careless-responder detector.

## Breadth and intensity, replacing the discrete levels (added 2026-08-21, NOT YET RUN)

`construct_concentration.py` reports each construct as `M_c = N_c x I_c` — total mass, effective
breadth (the participation ratio over per-person shares), and the intensity a typical carrier gives
it. No thresholds. The point is that `n_ge10 / n_ge20 / n_ge30` mixes how many people a construct
touches with how hard it hits them, so a rare-but-dominant construct (interpersonal & sexual
violence, dominant in four) looks the same as one that does nothing.

The positive part carries `M`, `N` and `I`, and suppressor mass is reported in its own columns
(`neg_mass`, `n_neg`) rather than folded in or dropped. Settled 2026-08-21.

**The reporting form is the rank curve, not the summary** (settled 2026-08-21). `fig_construct_ranks.py`
sorts each construct's per-person shares descending and plots rank against share, log x. A construct
that carries a few people hard starts high and falls off a cliff, a broad background starts low and
decays slowly, and the crossings are the finding. Nothing is thresholded and all thirty constructs
appear, the un-highlighted ones as grey context.

`construct_concentration.py` is kept for its diagnostic value, not as the reporting form. It is what
established that the participation ratio is a bulk statistic and cannot surface a construct whose
whole story is a handful of participants, and that averaging shares across encoders BEFORE the ratio
smooths the distribution — pooled intensity fell below every per-encoder value for eight constructs,
which `--encoder gte` then corrected. Sorting in `fig_construct_ranks.py` therefore happens WITHIN
each encoder and the curves are combined rank-wise, never person-wise.

**If either is adopted for reporting, it supersedes the `n_ge20` sentences currently in `sec:group`** —
"food insecurity leads 107 but reaches a fifth of the distance for only 27", "psychosis reaches a
fifth for 106 and a third for 57", "childhood adversity reaches a tenth for 81 and a fifth for none".
Those numbers come from `construct_distribution.py`. Do not change them without deciding to.

## ★ TRAP — two different "leading construct" rules live in this folder

They give different answers on the same data. Checked 2026-08-21.

| rule | where | psychosis / food insec. / everyday discr. / neighborhood / depression / healthcare discr. |
|---|---|---|
| argmax of **encoder-averaged** shares | `outlier_profile.py:54-55` | 138 / 107 / 76 / 52 / 49 / 21 |
| per-encoder argmax, then **modal** | `attribution.py:186-195` | 137 / 100 / 83 / 51 / 51 / 24 |

**The published numbers in `sec:group` are the first rule**, and so is the 81 % agreement figure
(`outlier_profile.py:73-74`, where `agree` counts encoders whose own argmax matches the *averaged*
leader). The two are consistent with each other because they come from the same script.

`attribution.py` prints its own ">= 4 of 6" line from the modal rule. **Do not quote it as the 81 %.**
The modal leader maximises encoder agreement by construction, so that percentage is not comparable
and will read as inflated.

Anything new that needs leader counts should use `outlier_profile.py`'s rule.
`fig_construct_ranks.py` does.

## Two threads that must not be mixed

**Clinical** — do the flagged meet validated criteria, and what content do the cutoff-bearing
instruments fail to cover. That is `cutoffs.py`, the breadth gradient, and the five.

**Statistical** — is a participant extreme in the joint distribution when no element is extreme alone.
That is `separability.py`, `joint_only.py`, and integration. It would mean the same on a battery with
no validated cutoffs anywhere.

`joint_profile.py` cross-tabulates the two deliberately, to check whether the joint survivors are the
under-caught group. **They are not** — survivors have *higher* breadth (5.33 vs 4.79) and none of them
meets zero criteria. Keep the two findings in separate sections.

## Retired

`fig_breadth.py` and `fig_breadth.png` were superseded by `fig_screen.py` and removed; it covers the same
breadth panel plus the threshold sweep. Delete them if they reappear from a sync.
