## Supplemental code for "Scoring respondents on a semantic frame of reference identifies a group that is burdened across many questionnaires and reaches care less easily": response_style.py

```
#!/usr/bin/env python3
"""response_style.py -- is the screen just finding extreme or careless responders?

STANDALONE (numpy, pandas, scipy). Two competing explanations for the flagged group, both computed
from the responses alone and neither using the projection.

  CONTENT EXTREMITY   the mean |z| of a participant's answers over the 151 items, each item
                      standardized across the cohort. High if someone reports a lot of everything.
                      The screen SHOULD track this to some degree -- the flagged group is burdened --
                      so the question is whether it reduces to it.

  ENDPOINT USE        the fraction of polytomous items (four or more distinct answers) a participant
                      answers at that item's lowest or highest option. This is the careless- or
                      extreme-responding style: someone who picks the ends regardless of content.
                      If the screen were picking up response style, the score would climb with it.

  DIRECTIONAL SKEW    among a participant's items beyond |z| = 2, the fraction on the adverse side.
                      Distinguishes "reports a lot of adversity" from "uses the ends in both
                      directions".

For each measure: its correlation with the continuous outlier score, and how many of the flagged
group you would recover by ranking the whole cohort on that measure alone and taking the same number
of people. A measure that recovered most of them would mean the screen is a relabelling of it.

The 151 SURVEY ITEMS ONLY. Age and sex are covariates that the screen residualizes out, and sweeping
them into the mean |z| was a bug in an earlier version of this check.

INPUTS   the k0 response file, and screen_out/consensus_outliers.csv (screen_battery.py)
OUTPUT   screen_out/response_style.json
Run:     python3 response_style.py
"""
import json
import math
import os

import numpy as np
import pandas as pd
from scipy.stats import spearmanr

HERE = os.path.dirname(os.path.abspath(__file__))
RESPONSES_CSV = "/home/jupyter/responses_out/responses_k0.csv"
ITEMS_CSV = os.path.join(HERE, "core_items_151.csv")
OUT_DIR = os.path.join(HERE, "screen_out")
OUTLIERS_CSV = os.path.join(OUT_DIR, "consensus_outliers.csv")
ID_COL = "id"
Z95 = 1.959963985
POLYTOMOUS_MIN = 4          # distinct answers for an item to count as a true Likert scale
EXTREME_Z = 2.0


def spearman_ci(x, y):
    """Spearman with a Fisher-z interval, Bonett-Wright standard error."""
    x, y = np.asarray(x, float), np.asarray(y, float)
    m = np.isfinite(x) & np.isfinite(y)
    x, y = x[m], y[m]
    rho, p = spearmanr(x, y)
    se = 1.03 / math.sqrt(len(x) - 3)
    z = math.atanh(rho)
    return rho, math.tanh(z - Z95 * se), math.tanh(z + Z95 * se), p, len(x)


def main():
    items = pd.read_csv(ITEMS_CSV)["item"].astype(str).tolist()
    R = pd.read_csv(RESPONSES_CSV)
    R[ID_COL] = R[ID_COL].astype(str)
    R = R.set_index(ID_COL)
    X = R[items].apply(pd.to_numeric, errors="coerce")
    keep = [c for c in items if X[c].notna().any()]
    X = X[keep]

    sel = pd.read_csv(OUTLIERS_CSV)
    sel[ID_COL] = sel[ID_COL].astype(str)
    sel = sel.set_index(ID_COL).reindex(X.index)
    score = sel["outlier_score"]
    is_out = sel["flagged_by_all"].astype(bool).values
    out_ids = set(X.index[is_out])
    n_out = int(is_out.sum())
    print(f"cohort {len(X):,} | {len(keep)} items | consensus outliers {n_out}\n")

    Z = (X - X.mean()) / X.std(ddof=0).replace(0, np.nan)
    content_ext = Z.abs().mean(axis=1)

    poly = [c for c in keep if X[c].nunique() >= POLYTOMOUS_MIN]
    lo, hi = X[poly].min(), X[poly].max()
    ers = (X[poly].eq(lo) | X[poly].eq(hi)).mean(axis=1)

    n_hi = (Z >= EXTREME_Z).sum(axis=1)
    n_lo = (Z <= -EXTREME_Z).sum(axis=1)
    dir_skew = n_hi / (n_hi + n_lo).replace(0, np.nan)

    print(f"{len(poly)} of {len(keep)} items are polytomous ({POLYTOMOUS_MIN}+ distinct answers)\n")
    res = {}
    for name, ser in (("content extremity", content_ext), ("endpoint use", ers)):
        rho, lo_ci, hi_ci, p, n = spearman_ci(score.values, ser.values)
        top = set(ser.sort_values(ascending=False).head(n_out).index)
        rec = len(top & out_ids)
        res[name] = {"outlier_mean": float(ser[is_out].mean()), "cohort_mean": float(ser.mean()),
                     "rho": float(rho), "ci": [float(lo_ci), float(hi_ci)], "p": float(p),
                     "recovered": rec, "of": n_out}
        print(f"{name}")
        print(f"  outliers {ser[is_out].mean():.2f} vs cohort {ser.mean():.2f}")
        print(f"  rho with the outlier score {rho:+.2f}  95% CI [{lo_ci:+.2f}, {hi_ci:+.2f}]  p={p:.1e}")
        print(f"  ranking the cohort on this alone recovers {rec} of the {n_out} "
              f"({100*rec/n_out:.0f}%)\n")

    n_def = int(dir_skew.notna().sum())
    adverse = float(dir_skew[is_out].mean()) if dir_skew[is_out].notna().any() else None
    adverse_coh = float(dir_skew.mean()) if n_def else None
    res["directional_skew"] = {"outlier_mean": adverse, "cohort_mean": adverse_coh,
                               "n_defined": n_def}
    print(f"directional skew, the adverse-side share of a participant's items beyond |z|={EXTREME_Z:.0f}")
    if n_def:
        print(f"  outliers {adverse:.2f} vs cohort {adverse_coh:.2f}   "
              f"(defined for {n_def:,} participants with at least one such item)")
    else:
        print("  undefined -- no participant has an item beyond that threshold")
    print("  near 1 means the extremes are on the adverse side rather than the ends of the scale\n")

    print("reading this: a high correlation with content extremity is expected and not a problem, "
          "since\nthe flagged group is burdened. The test is whether ranking on it alone recovers "
          "the group,\nand whether the score climbs with endpoint use, which is what a careless "
          "responder produces.")

    os.makedirs(OUT_DIR, exist_ok=True)
    res["n_cohort"] = int(len(X))
    res["n_outliers"] = n_out
    res["n_items"] = len(keep)
    res["n_polytomous"] = len(poly)
    with open(os.path.join(OUT_DIR, "response_style.json"), "w") as fh:
        json.dump(res, fh, indent=2)
    print(f"\nwrote -> {OUT_DIR}/response_style.json")


if __name__ == "__main__":
    main()
```
