## Supplemental code for "Scoring respondents on a semantic frame of reference identifies a group that is burdened across many questionnaires and reaches care less easily": screen_battery.py

```
#!/usr/bin/env python3
"""screen_battery.py -- place a battery on the fixed reference basis and screen its respondents.

STANDALONE. Needs only numpy, pandas and scikit-learn. Nothing is refitted: the reference basis
is computed once on the 577-item pool and shipped in reference_basis/, and this script only
projects onto it. That makes the script a template -- point it at another battery's items,
responses and scale file and it runs unchanged.

WHAT IT DOES, per encoder

  1. PLACE the battery on the basis
        P_S = (E_S - mu) @ W.T                     items x k        E_S = battery item embeddings
        Z_S = (P_S - mu_scores) / sigma            items x k        in the pool's own SD units
     A component is KEPT when at least one battery item ANCHORS it -- clears that component's pool
     core-item band -- or when it is the MODAL component of some item, the one it stands out on most.
     Both conditions are per item against quantities fixed on the pool, so the kept set does not
     depend on what else is in the battery, and it stays below the battery's item count. That last
     property is what the screen needs: a scored subspace whose dimension reaches the item count
     spans the responses themselves, and the distance is then the one a response-only screen gives.

  2. SCORE the respondents
        X    = responses rescaled to [-1, 1] per the scale file, reverse-coded where it says so
        Y    = X @ P_S[:, kept]                    respondents x |kept|
        Yr   = Y residualized on the covariates (age, sex) by weighted least squares
        T2   = Mahalanobis distance^2 of Yr under a Ledoit-Wolf precision
     Responses are bounded and discrete, so the flag is the empirical upper tail of the cohort's own
     distance distribution, not a chi-square cutoff.

  3. CONSENSUS across encoders: a respondent is a consensus outlier when flagged under every encoder.
     Reports the p90-p99 threshold sweep, pairwise Jaccard against the independence null f/(2-f),
     and the cross-encoder outlier score (mean distance percentile).

INPUTS
  reference_basis/<enc>.npz          W, mu, sigma, mu_scores, band          (built once, shipped)
  item_embeddings/<prefix>_<enc>.npz items, vectors                         (the battery's items)
  RESPONSES_CSV                      id, age, sex, then one column per item (build_responses_v2)
  SCALES_CSV                         item, min, max, reverse
  WEIGHTS_CSV                        one weight per respondent, positional  (optional)

OUTPUTS (-> OUT_DIR)
  scores_<enc>.csv        id, distance, distance2, pctile, flagged
  kept_<enc>.csv          component, band, n_battery_items, battery items and their constructs
  consensus_outliers.csv  id, n_encoders_flagging, outlier_score, flagged_by_all
  screen_summary.csv      the p90-p99 sweep
  run_config.json         every setting and count, for the record

Run:  python screen_battery.py
"""
import json
import os
import sys

import numpy as np
import pandas as pd
from sklearn.covariance import LedoitWolf

# ----------------------------------------------------------------------------- CONFIG
HERE = os.path.dirname(os.path.abspath(__file__))
BASIS_DIR     = os.path.join(HERE, "reference_basis")
EMB_DIR       = os.path.join(HERE, "item_embeddings")
EMB_PREFIX    = "core151_items"                       # item_embeddings/<prefix>_<enc>.npz
RESPONSES_CSV = "/home/jupyter/responses_out/responses_k0.csv"
WEIGHTS_CSV   = "/home/jupyter/responses_out/weights_k0.csv"   # None to use unit weights
SCALES_CSV    = os.path.join(HERE, "scales_prism.csv")
ITEMS_CSV     = os.path.join(HERE, "core_items_151.csv")       # canonical item order
CONSTRUCTS_CSV = None            # optional: item,construct -- labels the kept components
OUT_DIR       = os.path.join(HERE, "screen_out")

ID_COL     = "id"
COVARIATES = ["age", "sex"]      # residualized out of the component scores. [] to skip.
PCTILE     = 95.0                # per-encoder flag threshold, empirical
SWEEP      = range(90, 100)

ENCODERS = ["gte-large-en-v1.5", "bge-m3", "bge-large-en-v1.5",
            "all-mpnet-base-v2", "e5-large-v2", "qwen3-embedding-0.6b"]


# ----------------------------------------------------------------------------- the keep rule
def keep_columns(Ztil, band):
    """Components the battery reaches: it anchors them, or they are some item's modal component."""
    anchored = {c for c in range(Ztil.shape[1]) if (np.abs(Ztil[:, c]) >= band[c]).any()}
    modal = {int(np.argmax(np.abs(Ztil[i]))) for i in range(Ztil.shape[0])}
    return sorted(anchored | modal)


def wls_residualize(Y, C, w):
    """Residualize each column of Y on the covariate matrix C (intercept added) by WLS."""
    if C is None or C.shape[1] == 0:
        return Y - np.average(Y, axis=0, weights=w)
    A = np.column_stack([np.ones(len(Y)), C])
    sw = np.sqrt(w)[:, None]
    beta, *_ = np.linalg.lstsq(A * sw, Y * sw, rcond=None)
    return Y - A @ beta


def mahalanobis2(Yr, w):
    """Squared Mahalanobis distance under a Ledoit-Wolf precision of the weighted covariance."""
    ctr = Yr - np.average(Yr, axis=0, weights=w)
    lw = LedoitWolf().fit(ctr)
    return np.einsum("ij,ij->i", ctr, ctr @ lw.precision_), lw


# ----------------------------------------------------------------------------- main
def main():
    os.makedirs(OUT_DIR, exist_ok=True)

    items = pd.read_csv(ITEMS_CSV)["item"].astype(str).tolist()
    scales = pd.read_csv(SCALES_CSV).set_index("item").reindex(items)
    assert scales["min"].notna().all(), "scale file does not cover every item"

    resp = pd.read_csv(RESPONSES_CSV)
    resp[ID_COL] = resp[ID_COL].astype(str)
    resp = resp.set_index(ID_COL)
    missing = [c for c in items if c not in resp.columns]
    assert not missing, f"{len(missing)} item columns absent from the responses: {missing[:5]}"
    raw = resp[items].values.astype(float)
    n_missing = int(np.isnan(raw).sum())
    assert n_missing == 0, (f"{n_missing} missing response cells. This screen has no imputation "
                            f"path -- use the k0 (complete) response file or add one deliberately.")

    # rescale to [-1, 1] on each item's own published range, then reverse-code where marked
    lo = scales["min"].values.astype(float)
    hi = scales["max"].values.astype(float)
    rev = scales["reverse"].astype(str).str.lower().eq("true").values
    X = np.clip(2.0 * (raw - lo) / (hi - lo) - 1.0, -1.0, 1.0)
    X[:, rev] *= -1.0                                     # +1 = the instrument's keyed direction

    cov = None
    if COVARIATES:
        absent = [c for c in COVARIATES if c not in resp.columns]
        assert not absent, f"covariates absent from the responses file: {absent}"
        cov = resp[COVARIATES].values.astype(float)
        assert not np.isnan(cov).any(), "a covariate is missing for some respondent"

    if WEIGHTS_CSV and os.path.exists(WEIGHTS_CSV):
        wdf = pd.read_csv(WEIGHTS_CSV)
        w = wdf.iloc[:, 0].values.astype(float)
        assert len(w) == len(resp), f"weights length {len(w)} != respondents {len(resp)}"
    else:
        w = np.ones(len(resp))

    con = None
    if CONSTRUCTS_CSV and os.path.exists(CONSTRUCTS_CSV):
        cdf = pd.read_csv(CONSTRUCTS_CSV)
        con = dict(zip(cdf.iloc[:, 0].astype(str), cdf.iloc[:, 1].astype(str)))

    print(f"cohort {len(resp):,} respondents x {len(items)} items | "
          f"covariates {COVARIATES or 'none'} | weights "
          f"{'unit' if np.allclose(w, 1) else 'supplied'}")

    dist, flags, kept_n = {}, {}, {}
    for enc in ENCODERS:
        b = np.load(os.path.join(BASIS_DIR, f"{enc}.npz"), allow_pickle=True)
        W, mu, sigma, mu_s, band = b["W"], b["mu"], b["sigma"], b["mu_scores"], b["band"]

        e = np.load(os.path.join(EMB_DIR, f"{EMB_PREFIX}_{enc}.npz"), allow_pickle=True)
        order = [str(x) for x in e["items"]]
        pos = {it: j for j, it in enumerate(order)}
        absent = [it for it in items if it not in pos]
        assert not absent, f"{enc}: {len(absent)} items have no embedding: {absent[:5]}"
        E = np.asarray(e["vectors"], float)[[pos[it] for it in items]]   # battery row order

        P = (E - mu) @ W.T                                # items x k
        Z = (P - mu_s) / sigma                            # in pool SD units
        kept = keep_columns(Z, band)
        modal_of = [int(np.argmax(np.abs(Z[i]))) for i in range(len(items))]

        Y = X @ P[:, kept]                                # respondents x |kept|
        Yr = wls_residualize(Y, cov, w)
        T2, _ = mahalanobis2(Yr, w)
        d = np.sqrt(np.maximum(T2, 0.0))
        pct = pd.Series(d).rank(pct=True).values * 100.0
        thr = np.percentile(d, PCTILE)
        flag = d > thr

        dist[enc] = pd.Series(d, index=resp.index)
        flags[enc] = pd.Series(flag, index=resp.index)
        kept_n[enc] = len(kept)

        pd.DataFrame({ID_COL: resp.index, "distance": d, "distance2": T2,
                      "pctile": pct, "flagged": flag}).to_csv(
            os.path.join(OUT_DIR, f"scores_{enc}.csv"), index=False)

        rows = []
        for c in kept:
            who = [items[i] for i in range(len(items))
                   if modal_of[i] == c or abs(Z[i, c]) >= band[c]]
            rows.append({"component": c, "band": round(float(band[c]), 4),
                         "n_battery_items": len(who),
                         "anchors": int((np.abs(Z[:, c]) >= band[c]).sum()),
                         "battery_items": " ".join(who),
                         "constructs": ("; ".join(f"{k}:{v}" for k, v in
                                        pd.Series([con[i] for i in who if i in con])
                                        .value_counts().items()) if con else "")})
        pd.DataFrame(rows).to_csv(os.path.join(OUT_DIR, f"kept_{enc}.csv"), index=False)
        print(f"  {enc:22s} k={W.shape[0]:3d}  kept={len(kept):3d}  flagged={int(flag.sum()):5d}")

    D = pd.DataFrame(dist)
    F = pd.DataFrame(flags)
    cons = F.all(axis=1)
    score = (D.rank(pct=True) * 100).mean(axis=1)
    pd.DataFrame({ID_COL: D.index, "n_encoders_flagging": F.sum(axis=1).values,
                  "outlier_score": score.values, "flagged_by_all": cons.values}).to_csv(
        os.path.join(OUT_DIR, "consensus_outliers.csv"), index=False)

    # threshold sweep, with the independence null for the pairwise overlap
    sweep = []
    for t in SWEEP:
        thr = D.quantile(t / 100.0)
        sets = {m: set(D.index[D[m] > thr[m]]) for m in ENCODERS}
        js = [len(sets[a] & sets[b]) / len(sets[a] | sets[b])
              for i, a in enumerate(ENCODERS) for b in ENCODERS[i + 1:]]
        f = (100 - t) / 100.0
        null = f / (2 - f)
        sweep.append({"pctile": t, "mean_flagged_per_encoder": int(np.mean([len(s) for s in sets.values()])),
                      "consensus_N": len(set.intersection(*sets.values())),
                      "union_N": len(set.union(*sets.values())),
                      "mean_jaccard": round(float(np.mean(js)), 3),
                      "null_jaccard": round(null, 4),
                      "x_null": round(float(np.mean(js)) / null, 1)})
    sw = pd.DataFrame(sweep)
    sw.to_csv(os.path.join(OUT_DIR, "screen_summary.csv"), index=False)

    print(f"\nconsensus outliers at p{PCTILE:.0f} (flagged by all {len(ENCODERS)}): "
          f"{int(cons.sum()):,} of {len(D):,}")
    print(sw.to_string(index=False))

    json.dump({"n_cohort": int(len(resp)), "n_items": len(items), "encoders": ENCODERS,
               "kept_per_encoder": kept_n, "pctile": PCTILE, "covariates": COVARIATES,
               "responses_csv": RESPONSES_CSV, "weights_csv": WEIGHTS_CSV,
               "unit_weights": bool(np.allclose(w, 1)),
               "n_consensus_outliers": int(cons.sum())},
              open(os.path.join(OUT_DIR, "run_config.json"), "w"), indent=2)
    print(f"\nwrote -> {OUT_DIR}")


if __name__ == "__main__":
    main()
```
