## Supplemental code for "Scoring respondents on a semantic frame of reference identifies a group that is burdened across many questionnaires and reaches care less easily": separability.py

```
#!/usr/bin/env python3
"""separability.py -- could each flagged participant have been found without reading the battery together?

Reruns, on the CURRENT screen, the two checks that decide whether the cross-instrument distance is
doing something a simpler reading of the same responses would not. No earlier result is assumed.

  CHECK 1  single-item and single-construct reach.
           Standardize every item across the cohort in its keyed direction. A participant is
           ITEM-FINDABLE if some single item puts them in the cohort's top 5% on that item, and
           CONSTRUCT-FINDABLE if some construct score (the mean of its items' z) does. A participant
           findable by NEITHER would have been reachable only by combining instruments.

  CHECK 2  does the cross-component covariance change who is flagged?
           The diagonal distance sum_j y_j^2 / Sigma_jj ignores every off-diagonal term of the score
           covariance. If it ranks participants the same way the full Mahalanobis distance does, the
           joint reading is not what produces the flags. Reported as Spearman over the cohort and as
           the overlap of the two top-5% sets, per encoder.

Both checks describe DETECTION. They say nothing about attribution, which is the separate claim that
a flag can be traced to the items behind it.

Run:  python3 separability.py
"""
import json
import os

import numpy as np
import pandas as pd
from scipy.stats import spearmanr
from sklearn.covariance import LedoitWolf

HERE = os.path.dirname(os.path.abspath(__file__))
BASIS_DIR = os.path.join(HERE, "reference_basis")
EMB_DIR = os.path.join(HERE, "item_embeddings")
EMB_PREFIX = "core151_items"
RESPONSES_CSV = "/home/jupyter/responses_out/responses_k0.csv"
WEIGHTS_CSV = "/home/jupyter/responses_out/weights_k0.csv"
SCALES_CSV = os.path.join(HERE, "scales_prism.csv")
ITEMS_CSV = os.path.join(HERE, "core_items_151.csv")
CONSTRUCTS_CSV = os.path.join(HERE, "construct_map_151.csv")
OUT_DIR = os.path.join(HERE, "screen_out")
ID_COL = "id"
COVARIATES = ["age", "sex"]
PCTILE = 95.0

ENCODERS = ["gte-large-en-v1.5", "bge-m3", "bge-large-en-v1.5",
            "all-mpnet-base-v2", "e5-large-v2", "qwen3-embedding-0.6b"]


def keep_columns(Z, band):
    anchored = {c for c in range(Z.shape[1]) if (np.abs(Z[:, c]) >= band[c]).any()}
    modal = {int(np.argmax(np.abs(Z[i]))) for i in range(Z.shape[0])}
    return sorted(anchored | modal)


def residualize(V, C, w):
    A = np.column_stack([np.ones(len(w))] + ([C] if C is not None and C.shape[1] else []))
    sw = np.sqrt(w)[:, None]
    beta, *_ = np.linalg.lstsq(A * sw, V * sw, rcond=None)
    return V - A @ beta


def main():
    items = pd.read_csv(ITEMS_CSV)["item"].astype(str).tolist()
    scales = pd.read_csv(SCALES_CSV).set_index("item").reindex(items)
    cmap = pd.read_csv(CONSTRUCTS_CSV).set_index("item")["construct"]

    R = pd.read_csv(RESPONSES_CSV)
    R[ID_COL] = R[ID_COL].astype(str)
    R = R.set_index(ID_COL)
    raw = R[items].values.astype(float)
    lo, hi = scales["min"].values.astype(float), scales["max"].values.astype(float)
    rev = scales["reverse"].astype(str).str.lower().eq("true").values
    X = np.clip(2.0 * (raw - lo) / (hi - lo) - 1.0, -1.0, 1.0)
    X[:, rev] *= -1.0                                   # +1 = the instrument's keyed direction

    cov = R[COVARIATES].values.astype(float) if COVARIATES else None
    w = (pd.read_csv(WEIGHTS_CSV).iloc[:, 0].values.astype(float)
         if WEIGHTS_CSV and os.path.exists(WEIGHTS_CSV) else np.ones(len(R)))

    sel = pd.read_csv(os.path.join(OUT_DIR, "consensus_outliers.csv"))
    sel[ID_COL] = sel[ID_COL].astype(str)
    out_ids = set(sel.loc[sel["flagged_by_all"].astype(bool), ID_COL])
    is_out = R.index.isin(out_ids).astype(bool)
    n_out = int(is_out.sum())
    print(f"cohort {len(R):,} | consensus outliers {n_out}\n")

    # ---------------- CHECK 1 ----------------
    # Items are discrete and bounded, so a single response category can hold far more than 5% of the
    # cohort and an exact top-5% does not exist. The threshold often IS the top category, where a
    # strict > excludes everyone. We report the INCLUSIVE rule (at or above the 95th percentile) as
    # primary -- it is the reading that makes a participant easiest to reach by one item, so it is
    # conservative against the screen -- and the strict rule beside it so the tie handling is visible.
    Z = (X - X.mean(0)) / X.std(0, ddof=0)
    item_thr = np.percentile(Z, PCTILE, axis=0)
    cvals = cmap.reindex(items).fillna("unlabelled").values
    CS = pd.DataFrame(Z, index=R.index, columns=items).T.groupby(cvals).mean().T
    con_thr = np.percentile(CS.values, PCTILE, axis=0)

    print("CHECK 1  reach of a single item or a single construct, at the top-5% convention")
    reach = {}
    for rule, op in (("inclusive (>=)", np.greater_equal), ("strict (>)", np.greater)):
        item_hit = op(Z, item_thr).sum(1)
        con_hit = op(CS.values, con_thr).sum(1)
        item_f, con_f = item_hit > 0, con_hit > 0
        neither = ~(item_f | con_f)
        cover = op(Z, item_thr).mean(0)                 # what share of the cohort each item reaches
        print(f"  --- {rule}")
        print(f"      outliers findable by >= 1 item      : {int(item_f[is_out].sum())} of {n_out}"
              f"  ({100*item_f[is_out].mean():.1f}%)")
        print(f"      outliers findable by >= 1 construct : {int(con_f[is_out].sum())} of {n_out}"
              f"  ({100*con_f[is_out].mean():.1f}%)")
        print(f"      outliers findable by NEITHER        : {int(neither[is_out].sum())} of {n_out}"
              f"  ({100*neither[is_out].mean():.1f}%)")
        print(f"      median items reaching an outlier    : {int(np.median(item_hit[is_out]))}"
              f"   (cohort median {int(np.median(item_hit))})")
        print(f"      a single item's 'top 5%' actually covers a median "
              f"{100*np.median(cover):.1f}% of the cohort under this rule")
        print(f"      cohort findable by >= 1 item        : {100*item_f.mean():.1f}%"
              f"   <- the bar this test sets")
        if rule.startswith("inclusive"):
            reach = {"item_hit": item_hit, "con_hit": con_hit,
                     "item_f": item_f, "con_f": con_f, "neither": neither}
    item_hit, con_hit = reach["item_hit"], reach["con_hit"]
    item_findable, con_findable, neither = reach["item_f"], reach["con_f"], reach["neither"]
    print()

    # ---------------- CHECK 2 ----------------
    print("CHECK 2  full Mahalanobis versus the diagonal distance that ignores cross-covariance")
    rows = []
    for enc in ENCODERS:
        b = np.load(os.path.join(BASIS_DIR, f"{enc}.npz"), allow_pickle=True)
        W, mu, sigma, mu_s, band = b["W"], b["mu"], b["sigma"], b["mu_scores"], b["band"]
        e = np.load(os.path.join(EMB_DIR, f"{EMB_PREFIX}_{enc}.npz"), allow_pickle=True)
        pos = {str(x): j for j, x in enumerate(e["items"])}
        E = np.asarray(e["vectors"], float)[[pos[it] for it in items]]
        P = (E - mu) @ W.T
        kept = keep_columns((P - mu_s) / sigma, band)

        Y = residualize(X @ P[:, kept], cov, w)
        Yc = Y - np.average(Y, axis=0, weights=w)
        lw = LedoitWolf().fit(Yc)
        full = np.einsum("ij,ij->i", Yc, Yc @ lw.precision_)
        diag = (Yc ** 2 / np.diag(lw.covariance_)).sum(1)
        rho = spearmanr(full, diag).statistic
        tf, td = np.percentile(full, PCTILE), np.percentile(diag, PCTILE)
        sf, sd = set(np.where(full > tf)[0]), set(np.where(diag > td)[0])
        ov = len(sf & sd) / len(sf | sd)
        keep_out = np.array([i for i, f in enumerate(is_out) if f])
        rows.append({"encoder": enc, "kept": len(kept), "spearman_full_vs_diag": round(float(rho), 3),
                     "top5_jaccard": round(ov, 3),
                     "outliers_in_diag_top5": int(np.isin(keep_out, list(sd)).sum())})
    D = pd.DataFrame(rows)
    print(D.to_string(index=False))
    print(f"\n  a high Spearman with a high overlap means the off-diagonal terms do not decide who is")
    print(f"  flagged. a lower overlap means they do. 'outliers_in_diag_top5' counts how many of the")
    print(f"  {n_out} consensus outliers the diagonal screen also reaches under that encoder.")

    os.makedirs(OUT_DIR, exist_ok=True)
    pd.DataFrame({ID_COL: R.index, "is_outlier": is_out, "n_items_top5": item_hit,
                  "n_constructs_top5": con_hit}).to_csv(
        os.path.join(OUT_DIR, "separability_reach.csv"), index=False)
    D.to_csv(os.path.join(OUT_DIR, "separability_diagonal.csv"), index=False)
    json.dump({"n_outliers": n_out,
               "item_findable": int(item_findable[is_out].sum()),
               "construct_findable": int(con_findable[is_out].sum()),
               "neither": int(neither[is_out].sum())},
              open(os.path.join(OUT_DIR, "separability_summary.json"), "w"), indent=2)
    print(f"\nwrote -> {OUT_DIR}")


if __name__ == "__main__":
    main()
```
