## Supplemental code for "Scoring respondents on a semantic frame of reference identifies a group that is burdened across many questionnaires and reaches care less easily": severity_baseline.py

```
#!/usr/bin/env python3
"""severity_baseline.py -- does the cross-instrument screen add anything over plain severity?

THE QUESTION R6 CANNOT ANSWER ON ITS OWN. That the flagged group reports more barriers and uses the
emergency department more is an association, and almost any severity measure on the same items would
produce it: people broadly burdened report more barriers. So the association says the group is real,
and says nothing about whether reading the instruments together adds anything. This asks the question
that would: against the same outcomes, does the cross-instrument distance carry information a plain
severity score does not?

TWO BASELINES.

(1) CONTENT EXTREMITY, the mean absolute standardized response over the 151 items, defined
in Methods and computed exactly as response_style.py computes it. Same items, same responses, none of
the cross-instrument structure -- "who deviates most" with no basis, no covariance and no components.
It is the right comparator precisely because it is what a reviewer would reach for first.

(2) THE DIAGONAL DISTANCE, the same component scores with every off-diagonal term of the covariance
set to zero, exactly as Supplementary S10 defines it. This separates two explanations that content
extremity cannot: the screen might beat a plain severity score simply because a Mahalanobis distance
weights by covariance, with the semantic basis doing none of the work. The diagonal distance uses the
basis and drops the covariance, so if it carries the association as well as the full distance does,
the reference structure is not what matters. Averaged over the six encoders as a percentile, the way
the outlier score is.

THE MODELS, nested, mirroring underserved.py's own specifications rather than inventing new ones. All
predictors are rank-transformed as that script does, then standardized, so a coefficient is per SD:

    M0   covariates only          (age, sex, and for ED reliance the visit count)
    M1   + content extremity      severity alone
    M2   + outlier percentile     the screen alone
    M3   + both                   what each adds over the other
    M4   + all three              the full distance against severity AND the diagonal distance

Reported per outcome: each coefficient in M3 with its interval, the R^2 increments M1->M3 (what the
screen adds over severity) and M2->M3 (what severity adds over the screen), and the correlation
between the two predictors.

HOW A NULL IS TO BE READ, agreed before running. The two predictors correlate around rho = 0.77 and
agree on only 244 of the 510 flagged, so they are far from the same measure, and at N ~ 21,000 the
incremental test is well powered. A null therefore means what it says rather than reflecting a lack of
power: if the screen adds nothing over content extremity, the main text says so and R6 shrinks to a
descriptive paragraph.

OUTCOMES, in the order of how much they are worth. ED share of visits comes from the records, a stream
that never touched the screen. The barriers are self-report from the same participants who produced
the responses the screen was computed on, so they share method with the predictor and are secondary.

INPUTS   responses_k0.csv, core_items_151.csv          (paths imported from attribution.py)
         screen_out/consensus_outliers.csv             (outlier_score, outlier percentile)
         the CDR via BigQuery, exactly as underserved.py pulls it
OUTPUT   screen_out/severity_baseline.json             every coefficient and interval

Run:  python3 severity_baseline.py
"""
import json
import os

import numpy as np
import pandas as pd
import statsmodels.formula.api as smf
from google.cloud import bigquery

# paths come from attribution.py rather than being redefined here: responses_k0.csv lives in
# /home/jupyter/responses_out/, not beside the scripts, and one copy of that fact is enough
from attribution import RESPONSES_CSV, ITEMS_CSV, OUT_DIR, ID_COL

OUTLIERS_CSV = os.path.join(OUT_DIR, "consensus_outliers.csv")

# the survey concept codes, IMPORTED from underserved.py rather than restated, so the outcomes here
# are the same objects that script reports and cannot drift from them
from underserved import USUAL, COST, DELAYED, COPING, CDR
from separability import (BASIS_DIR, EMB_DIR, EMB_PREFIX, SCALES_CSV, WEIGHTS_CSV, COVARIATES,
                          ENCODERS, keep_columns, residualize)
from sklearn.covariance import LedoitWolf


def Q(sql):
    return bigquery.Client().query(sql).result().to_dataframe()


def content_extremity():
    """Mean absolute standardized response over the 151 items. Mirrors response_style.py -- if one
    changes, change both."""
    items = pd.read_csv(ITEMS_CSV)["item"].astype(str).tolist()
    R = pd.read_csv(RESPONSES_CSV)
    R[ID_COL] = R[ID_COL].astype(str)
    X = R.set_index(ID_COL)[items].apply(pd.to_numeric, errors="coerce")
    X = X[[c for c in items if X[c].notna().any()]]
    Z = (X - X.mean()) / X.std(ddof=0).replace(0, np.nan)
    return Z.abs().mean(axis=1).rename("extremity")


def diagonal_percentile():
    """The diagonal distance per participant, averaged over encoders as a percentile.

    Mirrors separability.py's CHECK 2 exactly: same keyed rescaling, same keep rule, same covariate
    residualisation and weights, same Ledoit-Wolf fit -- then sum(y^2 / Sigma_jj) instead of the full
    quadratic form. If one changes, change both."""
    items = pd.read_csv(ITEMS_CSV)["item"].astype(str).tolist()
    scales = pd.read_csv(SCALES_CSV).set_index("item").reindex(items)
    R = pd.read_csv(RESPONSES_CSV)
    R[ID_COL] = R[ID_COL].astype(str)
    R = R.set_index(ID_COL)
    raw = R[items].values.astype(float)
    lo, hi = scales["min"].values.astype(float), scales["max"].values.astype(float)
    rev = scales["reverse"].astype(str).str.lower().eq("true").values
    X = np.clip(2.0 * (raw - lo) / (hi - lo) - 1.0, -1.0, 1.0)
    X[:, rev] *= -1.0
    cov = R[COVARIATES].values.astype(float) if COVARIATES else None
    w = (pd.read_csv(WEIGHTS_CSV).iloc[:, 0].values.astype(float)
         if WEIGHTS_CSV and os.path.exists(WEIGHTS_CSV) else np.ones(len(R)))

    pct = np.zeros(len(R))
    for enc in ENCODERS:
        b = np.load(os.path.join(BASIS_DIR, f"{enc}.npz"), allow_pickle=True)
        W, mu, sigma, mu_s, band = b["W"], b["mu"], b["sigma"], b["mu_scores"], b["band"]
        e = np.load(os.path.join(EMB_DIR, f"{EMB_PREFIX}_{enc}.npz"), allow_pickle=True)
        pos = {str(x): j for j, x in enumerate(e["items"])}
        E = np.asarray(e["vectors"], float)[[pos[it] for it in items]]
        P = (E - mu) @ W.T
        kept = keep_columns((P - mu_s) / sigma, band)
        Y = residualize(X @ P[:, kept], cov, w)
        Yc = Y - np.average(Y, axis=0, weights=w)
        lw = LedoitWolf().fit(Yc)
        diag = (Yc ** 2 / np.diag(lw.covariance_)).sum(1)
        pct += pd.Series(diag).rank(pct=True).values
    return pd.Series(pct / len(ENCODERS), index=R.index, name="diag_pctile")


def fit(df, outcome, predictors, covars):
    """Rank-transformed OLS, standardized coefficients. Returns (R^2, {name: (beta, lo, hi)})."""
    cols = [outcome] + predictors + covars
    m = df[cols].dropna().copy()
    for c in cols:
        if m[c].nunique() > 2:                      # leave binary covariates (sex) alone
            r = m[c].rank(pct=True)
            m[c] = (r - r.mean()) / r.std(ddof=0)
    f = smf.ols(f"{outcome} ~ " + " + ".join(predictors + covars), m).fit()
    out = {}
    for p in predictors:
        ci = f.conf_int().loc[p]
        out[p] = (float(f.params[p]), float(ci[0]), float(ci[1]), float(f.pvalues[p]))
    return float(f.rsquared), out, int(f.nobs)


def main():
    ext = content_extremity()
    dia = diagonal_percentile()
    sel = pd.read_csv(OUTLIERS_CSV)
    sel[ID_COL] = sel[ID_COL].astype(str)
    D = sel.set_index(ID_COL).join(ext, how="inner").join(dia, how="inner")
    D["outlier_pctile"] = D["outlier_score"].rank(pct=True)
    rho = float(D["outlier_pctile"].corr(D["extremity"].rank(pct=True), method="pearson"))
    print(f"{len(D):,} participants | Spearman(outlier score, content extremity) = {rho:.2f}")
    top_o = D["outlier_pctile"] >= 0.95
    top_e = D["extremity"].rank(pct=True) >= 0.95
    print(f"  top 5% by each: {int(top_o.sum()):,} and {int(top_e.sum()):,}, "
          f"overlapping on {int((top_o & top_e).sum()):,}")
    print(f"  Spearman(outlier score, diagonal distance) = "
          f"{D['outlier_pctile'].corr(D['diag_pctile'].rank(pct=True)):.2f}")

    # ---- EHR, exactly the linkage and outcome underserved.py uses ----
    ehr = Q(f"""SELECT co.person_id, COUNT(DISTINCT co.visit_occurrence_id) AS n_visits,
                SUM(CASE WHEN LOWER(c.concept_name) LIKE '%emergency%' THEN 1 ELSE 0 END) AS n_ed
                FROM `{CDR}.visit_occurrence` co
                JOIN `{CDR}.concept` c ON co.visit_concept_id = c.concept_id
                GROUP BY co.person_id""")
    demo = Q(f"""SELECT p.person_id, 2024 - EXTRACT(YEAR FROM p.birth_datetime) AS age,
                 LOWER(cs.concept_name) AS sex FROM `{CDR}.person` p
                 LEFT JOIN `{CDR}.concept` cs ON p.sex_at_birth_concept_id = cs.concept_id""")
    E = ehr.merge(demo, on="person_id", how="left")
    E["person_id"] = E.person_id.astype(str)
    E["female"] = (E.sex == "female").astype(int)
    E["ed_reliance"] = E.n_ed / E.n_visits.replace(0, np.nan)
    M = D.reset_index().rename(columns={ID_COL: "person_id"}).merge(E, on="person_id", how="inner")
    print(f"  EHR-linked: {len(M):,}")

    results, covar_sets = {}, {"ed_reliance": ["n_visits", "age", "female"]}

    # ---- self-reported barriers, same definitions ----
    codes = ",".join("'" + x + "'" for x in USUAL + COST + DELAYED + COPING)
    s = Q(f"""SELECT s.person_id, c.concept_code, s.answer FROM `{CDR}.ds_survey` s
              JOIN `{CDR}.concept` c ON s.question_concept_id = c.concept_id
              WHERE c.concept_code IN ({codes})""")
    s["person_id"] = s.person_id.astype(str)
    s["val"] = s.answer.astype(str).str.lower().str.rsplit(": ", n=1).str[-1].str.strip()
    yes = lambda cc: set(s[s.concept_code.isin(cc) & (s.val == "yes")].person_id)
    B = pd.DataFrame({"person_id": sorted(set(s.person_id))})
    B["any_cost_barrier"] = B.person_id.isin(yes(COST)).astype(int)
    B["any_access_barrier"] = B.person_id.isin(
        yes(COST) | yes(DELAYED) | yes(COPING)
        | set(s[(s.concept_code == USUAL[0]) & (s.val == "no")].person_id)).astype(int)
    M = M.merge(B, on="person_id", how="left")
    matched = int(M["any_access_barrier"].notna().sum())
    assert matched > 0, ("the barrier survey joined to nobody -- check the concept codes and that "
                         "person_id types match")
    print(f"  barrier survey answered by {matched:,} of the {len(M):,} linked")
    covar_sets["any_access_barrier"] = ["age", "female"]
    covar_sets["any_cost_barrier"] = ["age", "female"]

    for outcome, covars in covar_sets.items():
        r0, _, n = fit(M, outcome, [], covars)
        r1, c1, _ = fit(M, outcome, ["extremity"], covars)
        r2, c2, _ = fit(M, outcome, ["outlier_pctile"], covars)
        r3, c3, _ = fit(M, outcome, ["extremity", "outlier_pctile"], covars)
        r4, c4, _ = fit(M, outcome, ["extremity", "diag_pctile", "outlier_pctile"], covars)
        rd, cd, _ = fit(M, outcome, ["diag_pctile"], covars)
        b_e, le, he, pe = c3["extremity"]
        b_o, lo, ho, po = c3["outlier_pctile"]
        print(f"\n{outcome}  (n={n:,}, covariates {', '.join(covars)})")
        print(f"  severity alone            R2 {r1:.4f}   (+{r1-r0:.4f} over covariates)")
        print(f"  screen alone              R2 {r2:.4f}   (+{r2-r0:.4f} over covariates)")
        print(f"  both                      R2 {r3:.4f}")
        print(f"  diagonal distance alone   R2 {rd:.4f}   (+{rd-r0:.4f} over covariates)")
        print(f"  all three                 R2 {r4:.4f}")
        print(f"    screen ADDS over severity           +{r3-r1:.4f}")
        print(f"    severity ADDS over screen           +{r3-r2:.4f}")
        print(f"    screen ADDS over severity+diagonal  +{r4-fit(M, outcome, ['extremity', 'diag_pctile'], covars)[0]:.4f}")
        print(f"  in the joint model, per SD:")
        print(f"    content extremity   beta {b_e:+.3f} [{le:+.3f},{he:+.3f}]  p={pe:.1e}")
        print(f"    outlier percentile  beta {b_o:+.3f} [{lo:+.3f},{ho:+.3f}]  p={po:.1e}")
        print(f"  with the diagonal distance added, per SD:")
        for nm in ("extremity", "diag_pctile", "outlier_pctile"):
            b, l_, h_, pp = c4[nm]
            print(f"    {nm:18s} beta {b:+.3f} [{l_:+.3f},{h_:+.3f}]  p={pp:.1e}")
        results[outcome] = dict(n=n, r2_covars=r0, r2_severity=r1, r2_screen=r2, r2_both=r3,
                                screen_adds=r3 - r1, severity_adds=r3 - r2,
                                r2_diagonal=rd, r2_all_three=r4,
                                joint_extremity=[b_e, le, he, pe],
                                joint_screen=[b_o, lo, ho, po],
                                triple={k: list(v) for k, v in c4.items()}, covariates=covars)

    path = os.path.join(OUT_DIR, "severity_baseline.json")
    with open(path, "w") as fh:
        json.dump({"rho_predictors": rho, "outcomes": results}, fh, indent=2)
    print(f"\nwrote {path}")
    print("\nread the ED share first: it is the only outcome the screen never touched. The barriers "
          "are\nself-report from the same participants, so they share method with the predictor.")


if __name__ == "__main__":
    main()
```
