## Supplemental code for "Scoring respondents on a semantic frame of reference identifies a group that is burdened across many questionnaires and reaches care less easily": table_five.py

```
#!/usr/bin/env python3
"""table_five.py -- the main-text table of the handful of participants who meet no clinical criterion.

Replaces the prose enumeration that ran through R4, which gave a leading component and a share for
each participant in a sentence apiece and reported the routes in an unordered set ("the remaining
three fall between, at 44, 66 and 78%"). A table has one cell per participant per quantity, so every
one of them has to be filled from the pipeline rather than from the prose.

WHAT EACH COLUMN IS, all on ONE encoder (the exemplar), all read from five_report.py's own output:

  leading component   the component carrying the largest share of that participant's squared distance,
                      named by its BATTERY-SIDE construct profile -- the documented constructs of the
                      All of Us items that core it WITHIN the battery (DEFINITIONS D6-A). Component
                      indices are not printed: they are encoder-specific and mean nothing to a reader
  share               that component's share of the distance
  next largest        the same two things for the second-largest share, which is what makes "two or
                      three components reach half" concrete
  |z| alone           sqrt(u) for the leading component, how far the participant sits from the COHORT
                      on it with the other components ignored
  |z| given the rest  sqrt(c), how far they sit from what their OTHER components predict. These are
                      the two halves of a component's CHARACTER as Methods defines it, and they are
                      reported rather than the implied prediction, which needs R_c^2 as well and is
                      not in this file
  integration         the share of the participant's distance carried by components that are joint
                      (c > u), the quantity R4 reports per participant

PARTICIPANT ORDER is DESCENDING by the share of the leading component, so the most concentrated
participant is row 1. The file's own row order is not used: it is ascending by that share, which is an
artifact of how five_report.py iterates, and the prose this table replaces used a third order again.
Supplementary Table S22 is numbered to match this one. IDs are deliberately not printed anywhere.

Reads only what the pipeline wrote, and hardcodes no result:
    screen_out/five_report_<enc>_<tag>.csv   (five_report.py) -- id, component, share, z_marg, z_cond,
                                                joint, neutralized, battery_profile

Out: five_table.tex, to be \\input from main_nhb.tex, plus a plain-text preview on stdout
Run: python3 table_five.py [--enc gte-large-en-v1.5] [--tag invisible] [--out five_table.tex]
"""
import os

import pandas as pd

from attribution import OUT_DIR, ID_COL
from attribution_components import arg, DEFAULT_ENC

MAX_CONSTRUCTS = 4          # constructs listed per component, most items first, before "and others"


def constructs(profile):
    """'Everyday discrimination:4; Healthcare discrimination:3' -> 'everyday discrimination,
    healthcare discrimination'. The counts are dropped: they are core-item counts, not shares, and a
    reader would take them for weights."""
    p = str(profile).strip()
    if not p or p.startswith("("):
        return "---"
    names = [s.split(":")[0].strip() for s in p.split(";") if s.strip()]
    names = [n[0].lower() + n[1:] if n[:2].upper() != n[:2] else n for n in names]
    shown = names[:MAX_CONSTRUCTS]
    tail = " and others" if len(names) > MAX_CONSTRUCTS else ""
    return ", ".join(shown) + tail


def tex_escape(s):
    return s.replace("&", "\\&").replace("%", "\\%").replace("_", "\\_")


def main():
    enc = arg("--enc", DEFAULT_ENC)
    tag = arg("--tag", "invisible")
    out = arg("--out", "five_table.tex")

    src = os.path.join(OUT_DIR, f"five_report_{enc}_{tag}.csv")
    D = pd.read_csv(src)
    D[ID_COL] = D[ID_COL].astype(str)
    lead_share = D.groupby(ID_COL)["share"].max()
    ids = list(lead_share.sort_values(ascending=False).index)   # most concentrated first

    rows = []
    for n, pid in enumerate(ids, start=1):
        g = D[D[ID_COL] == pid].sort_values("share", ascending=False)
        lead, second = g.iloc[0], g.iloc[1]
        integration = float(g.loc[g["joint"].astype(bool), "share"].sum())
        rows.append(dict(n=n,
                         lead=constructs(lead["battery_profile"]),
                         lead_share=100 * float(lead["share"]),
                         second=constructs(second["battery_profile"]),
                         second_share=100 * float(second["share"]),
                         z_marg=float(lead["z_marg"]), z_cond=float(lead["z_cond"]),
                         integration=100 * integration))

    hdr = (f"% generated by all_of_us_codes/table_five.py on {enc}, tag={tag} -- do not edit\n")
    cap = ("\\caption{\\textbf{The five clinically invisible outliers, who meet none of the eight "
           "validated criteria.} "
           "For each, the component carrying the largest share of their squared distance and the "
           "next largest, each named by the documented constructs of the All of Us items that core it "
           "within the battery, under the exemplar encoder. $|z|$ alone is how far the participant "
           "sits from the cohort on that component with the others ignored, and $|z|$ given "
           "the rest is how far they sit from what their other components predict. Integration is the "
           "share of the distance carried by components that are extreme given the rest "
           "(Methods~\\S\\ref{sec:methods_stage3}). Participants are ordered by their "
           "largest share and numbered as in Supplementary Table~S22. The same five under all "
           "six encoders are Supplementary "
           "Note~S22.}\\label{tab:five}\n")

    # The leading component's four columns are kept adjacent so one spanning header covers them, and
    # the two construct columns wrap rather than run off the text block.
    lines = [hdr, "\\begin{table}[H]", "\\centering", cap, "\\small",
             "\\setlength{\\tabcolsep}{4.5pt}",
             "\\begin{tabular}{@{}c p{3.9cm} r r r p{2.9cm} r r@{}}", "\\toprule",
             "& \\multicolumn{4}{c}{Largest share} & \\multicolumn{2}{c}{Next largest} & \\\\",
             "\\cmidrule(lr){2-5}\\cmidrule(lr){6-7}",
             "& Constructs of its core items & Share & $|z|$ alone & $|z|$ given rest & Constructs & "
             "Share & Integration \\\\", "\\midrule"]
    for r in rows:
        lines.append(f"{r['n']} & {tex_escape(r['lead'])} & ${r['lead_share']:.1f}\\%$ & "
                     f"${r['z_marg']:.1f}$ & ${r['z_cond']:.1f}$ & "
                     f"{tex_escape(r['second'])} & ${r['second_share']:.1f}\\%$ & "
                     f"${r['integration']:.0f}\\%$ \\\\")
    lines += ["\\bottomrule", "\\end{tabular}", "\\end{table}", ""]

    with open(out, "w") as f:
        f.write("\n".join(lines))

    print(f"\n{len(rows)} participants, {enc}, tag={tag}\n")
    print(f"  {'#':>2s} {'share':>7s} {'2nd':>7s} {'|z|':>6s} {'|z|cond':>8s} {'integ':>6s}  leading")
    for r in rows:
        print(f"  {r['n']:2d} {r['lead_share']:6.1f}% {r['second_share']:6.1f}% {r['z_marg']:6.2f} "
              f"{r['z_cond']:8.2f} {r['integration']:5.0f}%  {r['lead'][:52]}")
    print(f"\n  wrote {os.path.abspath(out)}")


if __name__ == "__main__":
    main()
```
