## Supplemental code for "Scoring respondents on a semantic frame of reference identifies a group that is burdened across many questionnaires and reaches care less easily": underserved.py

```
#!/usr/bin/env python3
"""underserved.py -- is the flagged group under-served by the health system?

STANDALONE Workbench script. A port of results_3_4_4_EHR_new_v2.ipynb with one substantive change:
the flagged group comes from screen_battery.py (the reference-basis screen) rather than from the
retired dense per-encoder runs. The BigQuery pulls, the barrier definitions and every statistic are
carried over unchanged.

FRAMING (read before editing prose). The flagged group is derived from one thing, the meaning of the
survey items and the responses to them. Electronic health records are a separate stream that never
touched the screen. If that stream shows the group is under-served, that is a gap in CARE, not a
validation of the screen. Do not frame it as evidence the outliers are real.

WHAT IS REPORTED WHERE (decision carried over from the notebook)
  main, self-report      access and cost barriers, and care avoidance for a provider's race or religion
  main, records          ED share of visits. A ratio, so free of the observation-window confound
  supplement, records    raw visit counts, WITH the caveat that the gradient collapses under window
                         adjustment and per observation-year, so it is record completeness rather
                         than per-time under-engagement
  dropped                physical multimorbidity, same completeness confound, kept only as a printed
                         diagnostic

That reporting split was decided on the previous flagged group. The robustness rows are printed again
here so it can be re-decided rather than inherited.

INPUTS   screen_out/consensus_outliers.csv     (screen_battery.py)
         the CDR, via BigQuery
OUTPUTS  screen_out/underserved_stats.json     every number and interval
         screen_out/fig_ehr_gradient.png       main: barriers + ED share, by score decile
         screen_out/fig_ehr_visits_supp.png    supplement: raw visits against the per-year rate

Run:  python3 underserved.py
"""
import json
import math
import os

import numpy as np
import pandas as pd
from scipy.stats import spearmanr
import statsmodels.formula.api as smf
import matplotlib
matplotlib.use("Agg")
import matplotlib.pyplot as plt
from google.cloud import bigquery

HERE = os.path.dirname(os.path.abspath(__file__))
OUT_DIR = os.path.join(HERE, "screen_out")
OUTLIERS_CSV = os.path.join(OUT_DIR, "consensus_outliers.csv")
PCTILE = 95.0
# workspace-specific. Blank it before syncing anywhere else, so it falls back to WORKSPACE_CDR.
CDR_OVERRIDE = "wb-affable-acorn-7941.R2024Q3R8"
CDR = os.environ.get("WORKSPACE_CDR") or CDR_OVERRIDE
OKABE = {"red": "#D55E00", "orange": "#E69F00", "purple": "#CC79A7",
         "blue": "#0072B2", "green": "#009E73", "ink": "#222222"}

USUAL = ["HealthAdvice_PlaceforHealthAdvice"]
COST = ["CantAffordCare_HealthcareProvider", "CantAffordCare_PrescriptionMedicines",
        "CantAffordCare_Specialist", "CantAffordCare_FollowupCare", "CantAffordCare_EmergencyCare",
        "CantAffordCare_DentalCare", "CantAffordCare_Eyeglasses",
        "DelayedMedicalCare_CantAffordCoPay", "DelayedMedicalCare_DeductibleTooHigh",
        "DelayedMedicalCare_HadToPayOutOfPocket"]
DELAYED = ["DelayedMedicalCare_ChildCare", "DelayedMedicalCare_ElderlyCare",
           "DelayedMedicalCare_RuralArea", "DelayedMedicalCare_TimeOffWork",
           "DelayedMedicalCare_Transportation"]
COPING = ["CantAffordCare_DelayedFillingRxToSaveMoney", "CantAffordCare_LowerCostRxToSaveMoney",
          "CantAffordCare_SkippedMedToSaveMoney", "CantAffordCare_TookLessMedToSaveMoney"]
MECH = ["HealthProviderRaceReligion_DelayedOrNoCare"]


def wilson(k, n, z=1.959963985):
    p = k / n; d = 1 + z * z / n
    ctr = (p + z * z / (2 * n)) / d
    h = z * math.sqrt(p * (1 - p) / n + z * z / (4 * n * n)) / d
    return ctr - h, ctr + h


def newcombe(k1, n1, k2, n2):
    p1, p2 = k1 / n1, k2 / n2
    l1, u1 = wilson(k1, n1); l2, u2 = wilson(k2, n2)
    return ((p1 - p2) - math.sqrt((p1 - l1) ** 2 + (u2 - p2) ** 2),
            (p1 - p2) + math.sqrt((u1 - p1) ** 2 + (p2 - l2) ** 2))


def rr_katz(ko, no, kr, nr, z=1.959963985):
    if min(ko, kr, no, nr) == 0:
        return float("nan"), float("nan"), float("nan")
    rr = (ko / no) / (kr / nr)
    se = math.sqrt(1.0 / ko - 1.0 / no + 1.0 / kr - 1.0 / nr)
    return rr, rr * math.exp(-z * se), rr * math.exp(z * se)


def main():
    if not CDR:
        raise SystemExit("no CDR dataset. print(os.environ['WORKSPACE_CDR']) and set CDR_OVERRIDE.")
    client = bigquery.Client()
    def Q(sql):
        return client.query(sql).to_dataframe()

    # ---- the flagged group, from the reference-basis screen ----
    sel = pd.read_csv(OUTLIERS_CSV)
    sel["id"] = sel["id"].astype(str)
    sel = sel.set_index("id")
    outlier_score = sel["outlier_score"]                 # mean distance percentile across encoders
    OUTLIERS = sel.index[sel["flagged_by_all"].astype(bool)]
    N_OUT = len(OUTLIERS)
    local = pd.DataFrame({"outlier_pctile": outlier_score,
                          "is_consensus_outlier": outlier_score.index.isin(OUTLIERS)})
    local.index.name = "person_id"
    print(f"cohort {len(local):,} | consensus outliers {N_OUT} | CDR {CDR}")

    # ---- EHR linkage is >= 1 visit. observation_period exists for every responder, so it does
    #      not mark linkage. obs_years is used only for the window adjustment and the per-year rate.
    obs = Q(f"""SELECT person_id,
                DATE_DIFF(MAX(observation_period_end_date), MIN(observation_period_start_date), DAY)/365.25 AS obs_years
                FROM `{CDR}.observation_period` GROUP BY person_id""")
    util = Q(f"""SELECT person_id, COUNT(DISTINCT visit_occurrence_id) AS n_visits,
                 COUNTIF(visit_concept_id IN (9203, 262)) AS n_ed_visits
                 FROM `{CDR}.visit_occurrence` GROUP BY person_id""")
    multimorb = Q(f"""
      WITH psych AS (
        SELECT DISTINCT ca.descendant_concept_id AS cid
        FROM `{CDR}.concept_ancestor` ca
        JOIN `{CDR}.concept` c ON ca.ancestor_concept_id = c.concept_id
        WHERE c.vocabulary_id='SNOMED' AND c.standard_concept='S'
          AND c.concept_name IN ('Mental disorder','Substance-related disorder',
                                 'Disorder due to psychoactive substance'))
      SELECT co.person_id, COUNT(DISTINCT co.condition_concept_id) AS n_conditions
      FROM `{CDR}.condition_occurrence` co
      WHERE co.condition_concept_id != 0 AND co.condition_concept_id NOT IN (SELECT cid FROM psych)
      GROUP BY co.person_id""")
    demo = Q(f"""SELECT p.person_id, 2024 - EXTRACT(YEAR FROM p.birth_datetime) AS age,
                 LOWER(cs.concept_name) AS sex
                 FROM `{CDR}.person` p
                 LEFT JOIN `{CDR}.concept` cs ON p.sex_at_birth_concept_id = cs.concept_id""")
    for df in (obs, util, multimorb, demo):
        df["person_id"] = df.person_id.astype(str)

    ehr = (local.reset_index()
           .merge(util, on="person_id", how="inner")      # inner = EHR-linked
           .merge(obs, on="person_id", how="left")
           .merge(multimorb, on="person_id", how="left")
           .merge(demo, on="person_id", how="left"))
    ehr["n_conditions"] = ehr["n_conditions"].fillna(0)
    ehr["female"] = (ehr.sex == "female").astype(int)
    ehr["ed_reliance"] = ehr.n_ed_visits / ehr.n_visits.replace(0, np.nan)
    ehr["visit_rate"] = ehr.n_visits / ehr.obs_years.replace(0, np.nan)
    n_link = len(ehr)
    n_out_link = int(ehr.is_consensus_outlier.sum())
    print(f"EHR-linked (>=1 visit): {n_link:,} of {len(local):,} "
          f"({100*n_link/len(local):.0f}%) | outliers linked {n_out_link} of {N_OUT} "
          f"({100*n_out_link/N_OUT:.0f}%)")

    # ---- self-reported access barriers ----
    codes = ",".join("'" + x + "'" for x in USUAL + COST + DELAYED + COPING + MECH)
    s = Q(f"""SELECT s.person_id, c.concept_code, s.answer
              FROM `{CDR}.ds_survey` s JOIN `{CDR}.concept` c ON s.question_concept_id = c.concept_id
              WHERE c.concept_code IN ({codes})""")
    s["person_id"] = s.person_id.astype(str)
    s["val"] = s.answer.astype(str).str.lower().str.rsplit(": ", n=1).str[-1].str.strip()
    def yes(cc):
        return set(s[s.concept_code.isin(cc) & (s.val == "yes")].person_id)

    F = pd.DataFrame({"person_id": sorted(set(s.person_id))})
    F["no_usual_source"] = F.person_id.isin(set(
        s[(s.concept_code == USUAL[0]) & (s.val == "no")].person_id)).astype(int)
    F["any_cost_barrier"] = F.person_id.isin(yes(COST)).astype(int)
    F["any_delayed_care"] = F.person_id.isin(yes(DELAYED)).astype(int)
    F["any_cost_coping"] = F.person_id.isin(yes(COPING)).astype(int)
    F["any_access_barrier"] = F[["no_usual_source", "any_cost_barrier",
                                 "any_delayed_care", "any_cost_coping"]].max(axis=1)
    F["care_avoid_provider"] = F.person_id.isin(set(
        s[(s.concept_code == MECH[0])
          & (s.val.isin({"some of the time", "most of the time", "always"}))].person_id)).astype(int)

    bar = ehr.merge(F, on="person_id", how="inner")
    oc = bar.is_consensus_outlier.values
    n_o, n_r = int(oc.sum()), int((~oc).sum())
    print(f"\naccess barriers (survey answerers: outliers {n_o} vs rest {n_r:,})")
    bar_rows = []
    for c, lab in [("any_access_barrier", "any access barrier"),
                   ("any_cost_barrier", "cost barrier"),
                   ("any_delayed_care", "delayed care, structural"),
                   ("any_cost_coping", "medication rationing to save money"),
                   ("no_usual_source", "no usual source of care"),
                   ("care_avoid_provider", "care avoidance, provider race or religion")]:
        ko, kr = int(bar[c][oc].sum()), int(bar[c][~oc].sum())
        op, rp = 100 * ko / n_o, 100 * kr / n_r
        dlo, dhi = newcombe(ko, n_o, kr, n_r)
        rr, rlo, rhi = rr_katz(ko, n_o, kr, n_r)
        bar_rows.append({"barrier": lab, "outlier_pct": op, "rest_pct": rp, "diff_pp": op - rp,
                         "diff_lo": 100 * dlo, "diff_hi": 100 * dhi,
                         "rr": rr, "rr_lo": rlo, "rr_hi": rhi})
        print(f"  {lab:44s} {op:5.1f}% vs {rp:5.1f}%  diff {op-rp:+5.1f}pp "
              f"[{100*dlo:+.0f},{100*dhi:+.0f}]   RR {rr:4.2f} [{rlo:.2f},{rhi:.2f}]")

    # ---- engagement and ED reliance, rank-OLS ----
    for c in ["outlier_pctile", "n_visits", "visit_rate", "age", "obs_years"]:
        ehr[c + "_r"] = ehr[c].rank(pct=True)

    def scoef(formula, cols):
        m = ehr.dropna(subset=cols).copy()
        fit = smf.ols(formula, m).fit()
        b = fit.params["outlier_pctile_r"]
        lo, hi = fit.conf_int().loc["outlier_pctile_r"]
        return b, lo, hi, fit.pvalues["outlier_pctile_r"], len(m)

    b0, l0, h0, p0, nm = scoef("n_visits_r ~ outlier_pctile_r + age_r + female",
                               ["n_visits_r", "outlier_pctile_r", "age_r", "female"])
    b1, l1, h1, p1, _ = scoef("n_visits_r ~ outlier_pctile_r + age_r + female + obs_years_r",
                              ["n_visits_r", "outlier_pctile_r", "age_r", "female", "obs_years_r"])
    b2, l2, h2, p2, _ = scoef("visit_rate_r ~ outlier_pctile_r + age_r + female",
                              ["visit_rate_r", "outlier_pctile_r", "age_r", "female"])
    print(f"\nengagement, rank-OLS (n={nm:,})")
    print(f"  visits, age+sex adjusted      beta={b0:+.3f} [{l0:+.3f},{h0:+.3f}]  p={p0:.1e}")
    print(f"  visits, + observation window  beta={b1:+.3f} [{l1:+.3f},{h1:+.3f}]  p={p1:.1e}")
    print(f"  visits per observation-year   beta={b2:+.3f} [{l2:+.3f},{h2:+.3f}]  p={p2:.1e}")
    print("  the last two decide whether the raw gradient is record completeness or under-engagement")

    oce = ehr.is_consensus_outlier.values
    rho_ed, p_ed = spearmanr(ehr.outlier_pctile, ehr.ed_reliance)
    edshare_o = ehr.ed_reliance[oce].mean() * 100
    edshare_r = ehr.ed_reliance[~oce].mean() * 100
    pool_o = 100 * ehr.n_ed_visits[oce].sum() / ehr.n_visits[oce].sum()
    pool_r = 100 * ehr.n_ed_visits[~oce].sum() / ehr.n_visits[~oce].sum()
    ehr["ed_reliance_r"] = ehr.ed_reliance.rank(pct=True)
    be, le, he, pe, _ = scoef("ed_reliance_r ~ outlier_pctile_r + n_visits_r + age_r + female",
                              ["ed_reliance_r", "outlier_pctile_r", "n_visits_r", "age_r", "female"])
    print(f"\nED reliance (ED visits / total visits, a ratio so window-free)")
    print(f"  rho with the score        {rho_ed:+.3f}  p={p_ed:.1e}")
    print(f"  mean per-person ED share  outliers {edshare_o:.1f}% vs rest {edshare_r:.1f}%")
    print(f"  pooled ED share           outliers {pool_o:.1f}% vs rest {pool_r:.1f}%")
    print(f"  ~ score + visits/age/sex  beta={be:+.3f} [{le:+.3f},{he:+.3f}]  p={pe:.1e}")

    rho_mm, p_mm = spearmanr(ehr.outlier_pctile, ehr.n_conditions)
    med_mm = ehr.groupby("is_consensus_outlier").n_conditions.median()
    print(f"\n[diagnostic, not reported] multimorbidity rho={rho_mm:+.3f} p={p_mm:.1e}, median "
          f"conditions outliers {med_mm.get(True, float('nan')):.0f} vs rest "
          f"{med_mm.get(False, float('nan')):.0f}")

    # ---- decile gradient ----
    d = ehr.merge(F, on="person_id", how="left")
    d["decile"] = pd.qcut(d.outlier_pctile, 10, labels=False) + 1
    g = d.groupby("decile").agg(
        n=("person_id", "size"), mean_pctile=("outlier_pctile", "mean"),
        access_barrier_pct=("any_access_barrier", lambda x: 100 * np.nanmean(x)),
        cost_barrier_pct=("any_cost_barrier", lambda x: 100 * np.nanmean(x)),
        care_avoid_pct=("care_avoid_provider", lambda x: 100 * np.nanmean(x)),
        median_visits=("n_visits", "median"), median_rate=("visit_rate", "median"),
        ed_reliance_pct=("ed_reliance", lambda x: 100 * np.nanmean(x)),
        median_conditions=("n_conditions", "median"))
    print("\nscore-decile gradient")
    print(g.round(2).to_string())

    # ---- figures ----
    os.makedirs(OUT_DIR, exist_ok=True)
    fig, axL = plt.subplots(figsize=(6.9, 3.4), facecolor="white")
    axL.plot(g.mean_pctile, g.access_barrier_pct, "-o", ms=3.5, color=OKABE["red"],
             label="any access barrier")
    axL.plot(g.mean_pctile, g.cost_barrier_pct, "-s", ms=3.5, color=OKABE["orange"],
             label="cost barrier")
    axL.plot(g.mean_pctile, g.care_avoid_pct, "-^", ms=3.5, color=OKABE["purple"],
             label="care avoidance, provider race or religion")
    axL.set_xlabel("mean distance percentile", fontsize=8)
    axL.set_ylabel("% reporting (survey)", fontsize=8)
    axR = axL.twinx()
    axR.plot(g.mean_pctile, g.ed_reliance_pct, "--D", ms=3.5, color=OKABE["blue"],
             label="ED share of visits (records)")
    axR.set_ylabel("ED share of visits (%)", fontsize=8, color=OKABE["blue"])
    axR.tick_params(axis="y", colors=OKABE["blue"], labelsize=7.5)
    axL.tick_params(labelsize=7.5)
    hl, ll = axL.get_legend_handles_labels()
    hr, lr = axR.get_legend_handles_labels()
    axL.legend(hl + hr, ll + lr, loc="upper center", bbox_to_anchor=(0.5, -0.24),
               ncol=2, frameon=False, fontsize=7.5)
    for sp in list(axL.spines.values()) + list(axR.spines.values()):
        sp.set_color("#9A9A9A")
    axL.spines["top"].set_visible(False); axR.spines["top"].set_visible(False)
    fig.subplots_adjust(left=0.09, right=0.90, top=0.96, bottom=0.36)
    fig.savefig(os.path.join(OUT_DIR, "fig_ehr_gradient.png"), dpi=300, facecolor="white")
    plt.close(fig)

    figs, axv = plt.subplots(figsize=(6.9, 3.4), facecolor="white")
    axv.plot(g.mean_pctile, g.median_visits, "--D", ms=3.5, color=OKABE["blue"],
             label="median total visits")
    axv.set_xlabel("mean distance percentile", fontsize=8)
    axv.set_ylabel("median total visits", fontsize=8, color=OKABE["blue"])
    axv.tick_params(axis="y", colors=OKABE["blue"], labelsize=7.5)
    axv.set_ylim(0, max(g.median_visits) * 1.1)
    axr = axv.twinx()
    axr.plot(g.mean_pctile, g.median_rate, "-o", ms=3.5, color=OKABE["green"],
             label="median visits per observation-year")
    axr.set_ylabel("visits per observation-year", fontsize=8, color=OKABE["green"])
    axr.tick_params(axis="y", colors=OKABE["green"], labelsize=7.5)
    axr.set_ylim(0, max(g.median_rate) * 1.3)
    hv, lv = axv.get_legend_handles_labels()
    hr2, lr2 = axr.get_legend_handles_labels()
    axv.legend(hv + hr2, lv + lr2, loc="upper center", bbox_to_anchor=(0.5, -0.24),
               ncol=2, frameon=False, fontsize=7.5)
    axv.tick_params(axis="x", labelsize=7.5)
    for sp in list(axv.spines.values()) + list(axr.spines.values()):
        sp.set_color("#9A9A9A")
    axv.spines["top"].set_visible(False); axr.spines["top"].set_visible(False)
    figs.subplots_adjust(left=0.09, right=0.90, top=0.96, bottom=0.30)
    figs.savefig(os.path.join(OUT_DIR, "fig_ehr_visits_supp.png"), dpi=300, facecolor="white")
    plt.close(figs)

    out = {"pctile": PCTILE, "n_cohort": int(len(local)), "n_consensus": int(N_OUT),
           "n_linked": int(n_link), "n_outliers_linked": n_out_link,
           "n_survey_outlier": n_o, "n_survey_rest": n_r, "barriers": bar_rows,
           "engagement": {"headline_beta": b0, "headline_ci": [l0, h0], "headline_p": p0,
                          "window_adj_beta": b1, "window_adj_ci": [l1, h1], "window_adj_p": p1,
                          "rate_beta": b2, "rate_ci": [l2, h2], "rate_p": p2},
           "ed_reliance": {"rho": rho_ed, "rho_p": p_ed,
                           "mean_share_outlier": edshare_o, "mean_share_rest": edshare_r,
                           "pooled_share_outlier": pool_o, "pooled_share_rest": pool_r,
                           "adj_beta": be, "adj_ci": [le, he], "adj_p": pe},
           "multimorbidity_rho": rho_mm, "multimorbidity_p": p_mm,
           "gradient": json.loads(g.reset_index().to_json(orient="records"))}
    with open(os.path.join(OUT_DIR, "underserved_stats.json"), "w") as fh:
        json.dump(out, fh, indent=2)
    print(f"\nwrote -> {OUT_DIR}/underserved_stats.json, fig_ehr_gradient.png, "
          f"fig_ehr_visits_supp.png")


if __name__ == "__main__":
    main()
```
