## Supplemental code for "Scoring respondents on a semantic frame of reference identifies a group that is burdened across many questionnaires and reaches care less easily": underserved_setup.py

```
#!/usr/bin/env python3
"""underserved_setup.py -- drop-in replacement for Cell 0 of results_3_4_4_EHR_new_v2.ipynb.

That notebook derived the flagged group from the dense per-encoder runs in outputs_dense/. The
screen now runs on the fixed reference basis instead, so the group is the 510 written by
screen_battery.py. Everything downstream of Cell 0 in that notebook uses only `local` and `N_OUT`,
so nothing else has to change.

USE, in the notebook:

    %run /home/jupyter/all_of_us_codes/underserved_setup.py

    placed where the dense loader was -- that is, REPLACE the block that runs from

        # ---- load 6 dense encoders -> distance matrix D ----
    through
        local.index = local.index.astype(str); local.index.name = "person_id"

    including the `assert 560 <= N_OUT <= 610` line, which refers to the retired 584.
    Keep the config block above it (PCTILE, CDR, the CI helpers) as it is.

DEFINES, with the same names and meanings the notebook expects:

    outlier_score   continuous exposure, the mean of a participant's distance percentile across the
                    six encoders. Identical in construction to the notebook's
                    (D.rank(pct=True) * 100).mean(axis=1) -- screen_battery.py computes it the same way.
    OUTLIERS        index of the consensus outliers, flagged above the per-encoder threshold by all six
    N_OUT           how many
    local           person_id-indexed frame with outlier_pctile and is_consensus_outlier
"""
import os

import pandas as pd

_HERE = os.path.dirname(os.path.abspath(__file__))
_SRC = os.path.join(_HERE, "screen_out", "consensus_outliers.csv")

_s = pd.read_csv(_SRC)
_s["id"] = _s["id"].astype(str)
_s = _s.set_index("id")

outlier_score = _s["outlier_score"]
OUTLIERS = _s.index[_s["flagged_by_all"].astype(bool)]
N_OUT = len(OUTLIERS)

local = pd.DataFrame({"outlier_pctile": outlier_score,
                      "is_consensus_outlier": outlier_score.index.isin(OUTLIERS)})
local.index = local.index.astype(str)
local.index.name = "person_id"

print(f"[setup] {os.path.relpath(_SRC, _HERE)}")
print(f"[setup] cohort {len(local):,} | consensus outliers N = {N_OUT} "
      f"(flagged above the per-encoder threshold by all six)")
print(f"[setup] exposure = mean distance percentile across encoders, "
      f"range {outlier_score.min():.1f} to {outlier_score.max():.1f}")
assert N_OUT > 0, "no consensus outliers -- run screen_battery.py first"
assert local.index.is_unique, "duplicate person ids in consensus_outliers.csv"
```
