## Supplemental code for "Scoring respondents on a semantic frame of reference identifies a group that is burdened across many questionnaires and reaches care less easily": zero_breadth.py

```
#!/usr/bin/env python3
"""zero_breadth.py -- the consensus outliers that meet none of the eight validated criteria.

Main-text R3 reports that 505 of the 510 meet at least one validated cutoff. This describes the
remainder: who they are in the attribution, so the main text can say what carries their flag instead
of asserting it. It also gives the leading-construct composition by breadth, which is the check that
the remainder is not simply the rest of the group in miniature.

Reads only what the pipeline wrote, and hardcodes no result:
    screen_out/outlier_leaders_<tag>.csv   (outlier_profile.py) -- id, leader, lead_share, agree, breadth
    screen_out/cutoff_flags.csv            (cutoffs.py)         -- per-instrument flags and breadth

Out: prints only. Nothing is refit and no threshold is re-chosen.
Run: python3 zero_breadth.py [--tag outliers]
"""
import os
import sys

import pandas as pd

HERE = os.path.dirname(os.path.abspath(__file__))
OUT_DIR = os.path.join(HERE, "screen_out")
ID_COL = "id"


def main():
    tag = "outliers"
    if "--tag" in sys.argv:
        tag = sys.argv[sys.argv.index("--tag") + 1]

    L = pd.read_csv(os.path.join(OUT_DIR, f"outlier_leaders_{tag}.csv"))
    L[ID_COL] = L[ID_COL].astype(str)
    n = len(L)
    print(f"\n{n} consensus outliers, tag={tag}\n")

    print("  breadth (validated criteria met, of the eight)")
    for b, g in L.groupby("breadth"):
        print(f"    {int(b)}: {len(g):4d}")

    zero = L[L["breadth"] == 0].sort_values("lead_share", ascending=False)
    print(f"\n  the {len(zero)} that meet NONE of the eight, and what carries them\n")
    print(f"    {'leader construct':34s} {'lead share':>11s} {'encoders agreeing':>18s}")
    for _, r in zero.iterrows():
        print(f"    {str(r['leader'])[:34]:34s} {100*r['lead_share']:10.1f}% {int(r['agree']):18d}")
    print(f"\n    leading constructs among them: "
          f"{', '.join(f'{k} ({v})' for k, v in zero['leader'].value_counts().items())}")

    print(f"\n  the same construct as leader in the rest of the group, for comparison")
    rest = L[L["breadth"] > 0]["leader"].value_counts()
    for k, v in rest.head(8).items():
        print(f"    {str(k)[:34]:34s} {v:4d}  ({100*v/len(L[L['breadth']>0]):.1f}% of the {len(L)-len(zero)})")

    # which cutoff instruments could have caught this content at all
    cf_path = os.path.join(OUT_DIR, "cutoff_flags.csv")
    if os.path.exists(cf_path):
        cf = pd.read_csv(cf_path)
        instr = [c for c in cf.columns if c not in (ID_COL, "breadth")]
        print(f"\n  the eight criteria cover these instruments: {', '.join(instr)}")


if __name__ == "__main__":
    main()
```
