## Supplemental code for "Scoring respondents on a semantic frame of reference identifies a group that is burdened across many questionnaires and reaches care less easily": ANALYSIS_SCRIPTS.md

# Analysis scripts behind the NHB results

**The step-by-step is `../README.md`** — environment, the four stages in order, and what runs where.
This file is the index the other way round: from a result in the manuscript to the script that
produces it. `FIGURES.md` is the same index for figures and generated tables.

> Rewritten 2026-08-31. The previous version described the **retired 151-item own basis** — k of 23
> to 26, outputs under `outputs_core151/` — which no longer appears anywhere in the manuscript. The
> live object is the 577-item reference pool with k of 49 to 55. Scripts from the retired line are in
> `../archive/superseded_20260831/`.

**Where each runs**

- **LOCAL** — published question text only, no responses. Project env, CPU:
  `CUDA_VISIBLE_DEVICES="" /data_gpu5/ROSS_Proj/env/bin/python <script>.py` from `../examples/allofus/`.
  Set `ROSS_NPZ_PREFIX=pool_items` and `ROSS_OUT_DIR=.../outputs_pool` to run the pipeline on the pool.
- **WORKBENCH** — needs individual-level All of Us responses, which cannot leave the Researcher
  Workbench. Scripts in `../all_of_us_codes/`, run there against `reference_basis/` from stage A8.

---

## R1 — the basis groups items by content

| Result | Script | Where |
|---|---|---|
| The basis itself: k per encoder, the sparse fit, the fixed-point cores | `1_select_k.py`, `2_fit.py`, `pool_fixedpoint_cores.py` | LOCAL |
| Penalty weight chosen once (Note S15, Table S11) | `lambda_sweep_pool.py`, `table_lambda_sweep.py` | LOCAL |
| Same-construct items share a core 18x chance (Note S16) | `core_separation.py` | LOCAL |
| Constructs split across components, reproducibly (Note S16) | `construct_splitting.py` | LOCAL |
| Documented construct among an item's core components, 87 v 40 per cent (Note S11) | `table_faithfulness_disagreements.py` | LOCAL |
| Cross-encoder reproduction, 39 of 55 and 27 of 55 (Note S12) | `fig_xenc.py`, `pool_xenc_hungarian.py` | LOCAL |
| Item-side profile agreement, 0.38 v 0.11 (Table S7) | `pool_xenc_itemside_profiles.py` | LOCAL |
| Sparsity, not dimension reduction, is what interprets (Note S19, Table S14) | `dense_control.py`, `make_dense_basis_from_npz.py`, `table_dense_control.py` | LOCAL |
| Response-based structure is sample-dependent, 0.85 falling to 0.29 (Note S13) | `portability_sweep.py`, `fig_portability.py` | WORKBENCH then LOCAL |

## R2 — the basis places instruments outside the pool

| Result | Script | Where |
|---|---|---|
| The All of Us battery placed, 43 of 55 components kept | `build_aou_poolw_projection.py` | LOCAL |
| Cores read from both sides, and the three external instruments | `make_core_items_pool.py`, `fig_aou_twoside_cores.py` | LOCAL |
| HRS placed, 21 of 55 kept, every construct covered (Notes S14, S18) | `hrs_kept_components.py`, `hrs_xenc_itemside_profiles.py` | LOCAL |
| Identical versus reworded phrasing, 0.73 and 0.49 (Table S13) | `table_phrasing_xenc.py` | LOCAL |
| Content versus syntax, 3,020 variants (Note S5, Fig S1) | `content_grammar_manifest.py`, `generate_variants.py`, `content_grammar_poolbasis.py`, `fig_contentgrammar_3panel.py` | LOCAL |
| How much of a question the basis holds (Note S21, Fig S15) | `fig_coverage.py`, `porting_quality.py` | LOCAL |

## R3 — scoring the cohort

| Result | Script | Where |
|---|---|---|
| The screen: distances, the 95th percentile, the 510 consensus outliers | `screen_battery.py` | WORKBENCH |
| Cross-encoder agreement of the flagged sets, 0.55 over the fifteen pairs (Note S6, Fig S2) | `fig_jaccard_threshold.py` | WORKBENCH |
| The eight validated criteria, 505 of 510 and a mean of 5.0 (Table S15, Fig 5) | `cutoffs.py`, `fig_screen.py` | WORKBENCH |
| Breadth is not response style or severity (Note S7) | `response_style.py`, `severity_baseline.py` | WORKBENCH |

## R4 and R5 — attribution

| Result | Script | Where |
|---|---|---|
| The distance partitioned over components, joint versus marginal, integration | `attribution.py`, `attribution_components.py` | WORKBENCH |
| The five who meet no criterion, and Table 1 | `zero_breadth.py`, `table_five.py`, `five_report.py` | WORKBENCH |
| The five under all six encoders (Note S22, Table S20) | `five_xenc.py` | WORKBENCH |
| Neutralizing individually extreme items (Note S9) | `joint_only.py` | WORKBENCH |
| How many items unflag each of the five | `five_items_to_drop.py` | WORKBENCH |
| What carries each outlier, and the rare tail (Note S26, Fig 6, Fig S16) | `outlier_profile.py`, `fig_component_ranks.py`, `heterogeneity.py` | WORKBENCH |
| Construct-level naming agrees across encoders for 82 per cent (Note S24) | `construct_distribution.py`, `construct_concentration.py` | WORKBENCH |
| Reachability by a single item or construct (Note S9) | `separability.py` | WORKBENCH |

## R6 — reaching care

| Result | Script | Where |
|---|---|---|
| Health-record and access-survey linkage, barriers and emergency-department share | `underserved_setup.py`, `underserved.py`, `outlier_ehr_underserved.py` | WORKBENCH |
| The visit-count caveat (Note S8, Fig S3) | `underserved.py` | WORKBENCH |
| Severity and covariance-free comparisons (Note S25, Table S21) | `severity_baseline.py` | WORKBENCH |
