## Supplemental code for "Scoring respondents on a semantic frame of reference identifies a group that is burdened across many questionnaires and reaches care less easily": environment.yml

```
# ──────────────────────────────────────────────────────────────────────
# Conda environment for ROSS-Proj (sparse-projection PCA).
#
# GPU target: NVIDIA + CUDA 12. For a CPU-only machine, replace
# "jax[cuda12]" with "jax" in the pip section.
#
# Create (run from the repo root so `-e .` resolves):
#     conda env create -f environment.yml
#     conda activate ross-proj
# ──────────────────────────────────────────────────────────────────────
name: ross-proj
channels:
  - conda-forge
dependencies:
  - python=3.11
  - numpy
  - scipy
  - scikit-learn
  - tqdm
  # Optional: .h5ad data loading for cpp_z on real data.
  - anndata
  - scanpy
  - pip
  - pip:
      - "jax[cuda12]"
      - optax
      - pytest
      - -e .
```
