## Supplemental code for "Scoring respondents on a semantic frame of reference identifies a group that is burdened across many questionnaires and reaches care less easily": 0_embed.py

```
#!/usr/bin/env python
"""Stage 0 — generate item embeddings from the prompts (one model per run).

Reads `core_items_151.csv` (`item,prompt`), encodes the prompts with a
**local** sentence-transformers model, and writes
`data/core151_items_<model>.npz` in the exact format the rest of the pipeline
consumes: `items` (151 labels), `vectors` (L2-normalized, 151 × D), `meta`
([backend, model_name, slug]). Mirrors the original survey_semantics embedding:
`SentenceTransformer(path, local_files_only=True).encode(texts, normalize_embeddings=True)`.

This runs in a **torch + sentence-transformers** env with local model weights —
NOT the JAX `ross_proj` env. See this folder's README for creating the envs and
downloading the models from scratch. To reuse the existing setup:

    SF=/data_gpu5/semantic_framework_vipul
    # 5 models in the transformers-5.x env:
    $SF/env/bin/python examples/allofus/0_embed.py --model bge-m3 --models-dir $SF/models
    # gte needs the transformers-4.x env + its custom remote code:
    $SF/env_gte_tf4/bin/python examples/allofus/0_embed.py \
        --model gte-large-en-v1.5 --models-dir $SF/models --trust-remote-code

(or run all six via 0_embed_all.sh).
"""
import argparse
import csv
import os

import numpy as np

HERE = os.path.dirname(os.path.abspath(__file__))
DATA_DIR = os.path.join(HERE, "data")

# model -> (local subdir under --models-dir, output suffix, needs trust_remote_code)
MODELS = {
    "bge-m3":               ("bge-m3",               "",                     False),
    "bge-large-en-v1.5":    ("bge-large-en-v1.5",    "bge-large-en-v1.5",    False),
    "all-mpnet-base-v2":    ("all-mpnet-base-v2",    "all-mpnet-base-v2",    False),
    "e5-large-v2":          ("e5-large-v2",          "e5-large-v2",          False),
    "gte-large-en-v1.5":    ("gte-large-en-v1.5",    "gte-large-en-v1.5",    True),
    "qwen3-embedding-0.6b": ("qwen3-embedding-0.6b", "qwen3-embedding-0.6b", False),
}


def main():
    ap = argparse.ArgumentParser(description=__doc__,
                                 formatter_class=argparse.RawDescriptionHelpFormatter)
    ap.add_argument("--model", required=True, choices=list(MODELS))
    ap.add_argument("--models-dir", default="/data_gpu5/semantic_framework_vipul/models",
                    help="directory holding the local model subfolders")
    ap.add_argument("--prompts", default=os.path.join(HERE, "core_items_151.csv"))
    ap.add_argument("--out", default=None, help="output .npz (default: data/core151_items_<model>.npz)")
    ap.add_argument("--trust-remote-code", action="store_true",
                    help="run a model's bundled custom code (gte needs this)")
    args = ap.parse_args()

    subdir, _suffix, trust_default = MODELS[args.model]
    trust = args.trust_remote_code or trust_default

    # prompts: item,prompt — preserve file order
    items, texts = [], []
    with open(args.prompts, newline="") as f:
        for row in csv.DictReader(f):
            items.append(row["item"])
            texts.append(row["prompt"])

    # local-only / offline, mirroring survey_semantics' policy
    for k, v in {"HF_HUB_OFFLINE": "1", "TRANSFORMERS_OFFLINE": "1",
                 "TOKENIZERS_PARALLELISM": "false"}.items():
        os.environ.setdefault(k, v)

    model_path = os.path.join(args.models_dir, subdir)
    from sentence_transformers import SentenceTransformer
    model = SentenceTransformer(model_path, local_files_only=True, trust_remote_code=trust)
    vectors = np.asarray(model.encode(texts, normalize_embeddings=True), dtype=float)

    model_name = f"models/{subdir}"
    slug = "sentence-transformers__" + model_name.replace("/", "_")
    out = args.out or os.path.join(DATA_DIR, f"core151_items_{args.model}.npz")
    np.savez(out, items=np.array(items),
             vectors=vectors,
             meta=np.array(["sentence-transformers", model_name, slug]))
    print(f"{args.model}: {vectors.shape[0]}×{vectors.shape[1]} (trust_remote_code={trust}) -> {out}")


if __name__ == "__main__":
    main()
```
