## Supplemental code for "Scoring respondents on a semantic frame of reference identifies a group that is burdened across many questionnaires and reaches care less easily": 1_select_k.py

```
#!/usr/bin/env python
"""Stage 1 — choose k per embedder by permutation Parallel Analysis.

Calls `ross_proj.select_k` on each embedder's item matrix (items × dims) and
writes, under outputs/k_selection/<embedder>/, the KResult JSON + a scree plot;
plus a combined outputs/k_selection/summary.csv. Stage 2 reads k from here.

Run from the repo root:
    python examples/allofus/1_select_k.py                 # all 6, B=200
    python examples/allofus/1_select_k.py --only bge-m3 --n-perm 500
"""
import argparse
import csv
import os

from ross_proj import select_k
from ross_proj.parallel_analysis import save_scree
from ross_proj.io import save_kresult

from _common import EMBEDDERS, OUT_DIR, load_embedding


def main():
    ap = argparse.ArgumentParser()
    ap.add_argument("--only", nargs="*", choices=list(EMBEDDERS))
    ap.add_argument("--n-perm", type=int, default=200)
    ap.add_argument("--percentile", type=float, default=95.0)
    ap.add_argument("--seed", type=int, default=0)
    args = ap.parse_args()

    names = args.only or list(EMBEDDERS)
    out_root = os.path.join(OUT_DIR, "k_selection")
    os.makedirs(out_root, exist_ok=True)
    rows = []
    for name in names:
        items, X = load_embedding(name)
        kres = select_k(X, n_perm=args.n_perm, percentile=args.percentile,
                        seed=args.seed)
        d = os.path.join(out_root, name)
        save_kresult(kres, d)
        save_scree(kres, os.path.join(d, "scree.png"), title=f"{name} (k={kres.k})")
        rows.append({"embedder": name, "n_items": X.shape[0], "embed_dim": X.shape[1],
                     "k": kres.k, "k_count": kres.k_count, "k_mean": kres.k_mean,
                     "d80": kres.d80, "d90": kres.d90})
        print(f"{name:22s} D={X.shape[1]:5d}  k={kres.k:3d}  "
              f"(count {kres.k_count}, mean {kres.k_mean}, d80 {kres.d80})", flush=True)

    with open(os.path.join(out_root, "summary.csv"), "w", newline="") as f:
        w = csv.DictWriter(f, fieldnames=list(rows[0].keys()))
        w.writeheader(); w.writerows(rows)
    print(f"\n-> {os.path.join(out_root, 'summary.csv')}")


if __name__ == "__main__":
    main()
```
