## Supplemental code for "Scoring respondents on a semantic frame of reference identifies a group that is burdened across many questionnaires and reaches care less easily": 2_fit.py

```
#!/usr/bin/env python
"""Stage 2 — fit ROSS-Proj per embedder × lambda_score.

Calls `ross_proj.fit_projection` and saves each fit (mean, loadings W, scores Z,
model) under outputs/fits/<embedder>__k{k}_ls{lambda_score}/ via `save_fit`,
plus a small config.json so stage 3 knows which embedding to reload.

k comes from stage 1 (outputs/k_selection/<embedder>/parallel_analysis.json) by
default; override with --k <int> to use a fixed k for all.

Run from the repo root (run 1_select_k.py first if using k=auto):
    python examples/allofus/2_fit.py                              # all 6, lambda_score=1.0
    python examples/allofus/2_fit.py --lambda-score 0.5 1.0 2.0   # sweep
    python examples/allofus/2_fit.py --only bge-m3 --k 39
"""
import argparse
import json
import os

from ross_proj import fit_projection
from ross_proj.io import save_fit

from _common import EMBEDDERS, OUT_DIR, load_embedding


def resolve_k(name, k_arg):
    if k_arg != "auto":
        return int(k_arg)
    path = os.path.join(OUT_DIR, "k_selection", name, "parallel_analysis.json")
    if not os.path.exists(path):
        raise SystemExit(f"missing {path} — run 1_select_k.py first, or pass --k <int>")
    return int(json.load(open(path))["k"])


def main():
    ap = argparse.ArgumentParser()
    ap.add_argument("--only", nargs="*", choices=list(EMBEDDERS))
    ap.add_argument("--lambda-score", nargs="+", type=float, default=[1.0])
    ap.add_argument("--lambda-ortho", type=float, default=0.1)
    ap.add_argument("--k", default="auto",
                    help="'auto' (read stage-1 k) or an integer for all embedders")
    ap.add_argument("--total-steps", type=int, default=5000)
    ap.add_argument("--seed", type=int, default=42)
    args = ap.parse_args()

    names = args.only or list(EMBEDDERS)
    fits_root = os.path.join(OUT_DIR, "fits")
    for name in names:
        items, X = load_embedding(name)
        n = X.shape[0]
        k = resolve_k(name, args.k)
        for ls in args.lambda_score:
            res = fit_projection(
                X, n_components=k, lambda_score=ls, lambda_ortho=args.lambda_ortho,
                batch_size=n, subsample_fraction=1.0, total_steps=args.total_steps,
                random_state=args.seed, verbose=False, log_interval=0)
            tag = f"{name}__k{k}_ls{ls}"
            d = os.path.join(fits_root, tag)
            save_fit(res, d)
            json.dump({"embedder": name, "n_components": k, "lambda_score": ls,
                       "lambda_ortho": args.lambda_ortho},
                      open(os.path.join(d, "config.json"), "w"), indent=2)
            print(f"{tag:42s} fit {n}×{X.shape[1]} -> {d}", flush=True)


if __name__ == "__main__":
    main()
```
