## Supplemental code for "Scoring respondents on a semantic frame of reference identifies a group that is burdened across many questionnaires and reaches care less easily": _common.py

```
"""Shared config for the core-151 example (paths + embedder list + loader).

151 survey "core" items (`core_items_151.csv`: `item,prompt`), each embedded by
several sentence-transformer models. Each `data/core151_items_<model>.npz` holds
`items` (151 labels), `vectors` (151 × D), `meta`.

The dataset is overridable without editing this file:
    ROSS_NPZ_PREFIX=<prefix>   ->  load data/<prefix>_<model>.npz   (default: core151_items)
    ROSS_OUT_DIR=<dir>         ->  write outputs there              (default: ./outputs_core151)
"""
import os

import numpy as np

HERE = os.path.dirname(os.path.abspath(__file__))
DATA_DIR = os.path.join(HERE, "data")
OUT_DIR = os.environ.get("ROSS_OUT_DIR") or os.path.join(HERE, "outputs_core151")

# The six models, in canonical order.
_MODELS = ["bge-m3", "bge-large-en-v1.5", "all-mpnet-base-v2",
           "e5-large-v2", "gte-large-en-v1.5", "qwen3-embedding-0.6b"]

# Each embedder loads data/<prefix>_<model>.npz (uniform naming, incl. bge-m3).
_PREFIX = os.environ.get("ROSS_NPZ_PREFIX", "core151_items")
EMBEDDERS = {m: f"{_PREFIX}_{m}.npz" for m in _MODELS}


def load_embedding(name):
    """Return (items, X) for an embedder.

    X = `vectors` exactly as stored: (151 items × D dims), **items as samples**
    (the orientation ROSS-Proj needs so that sparse scores → a few items per
    component). Raw vectors, no normalization.
    """
    d = np.load(os.path.join(DATA_DIR, EMBEDDERS[name]), allow_pickle=True)
    return np.asarray(d["items"]).astype(str), np.asarray(d["vectors"], dtype=np.float64)


# ──────────────────────────────────────────────────────────────────────────────
# Which columns of the pool reference basis a transferred battery is scored on.
# Definition of record for hrs_screen_poolw.py, hrs_attribution_poolw.py and
# build_aou_poolw_projection.py -- they must not carry their own copies.
# ──────────────────────────────────────────────────────────────────────────────

def pool_bands(enc, Zpool, pool_ids):
    """Per component, the pool's core-item threshold band.

    band[c] = the smallest standardized loading among the items that anchor component c's
    core on the pool itself, i.e. the edge of the grey band in the core-item figure. An
    item of a new battery counts as anchoring c when its own loading clears band[c].
    """
    import csv
    cores = {int(r["component"]): str(r["items"]).split() for r in csv.DictReader(
        open(os.path.join(HERE, "outputs_pool", "fixedpoint_cores", f"{enc}_cores.csv")))}
    pos = {it: j for j, it in enumerate(pool_ids)}
    return np.array([min(abs(Zpool[pos[h], c]) for h in cores[c])
                     for c in range(Zpool.shape[1])])


def keep_columns(Ztil, band):
    """The keep rule for scoring a battery on the fixed pool basis (settled 2026-08-19).

    A pool component is kept when

      (1) at least one battery item's standardized loading on it clears that component's
          own pool core-item band, so the battery ANCHORS it by the pool's standard, OR
      (2) it is the MODAL component of at least one battery item, the one component that
          item stands out on most.

    `Ztil` is the battery's item projections in pool SD units (items × k). Condition 2
    carries the rule for a small battery -- on the 35-item HRS battery only 5 of 55
    components satisfy (1), since a battery that small scored on a 577-item basis rarely
    produces a pool-grade core item -- and (1) is the safeguard for a component the
    battery anchors without any single item peaking there.

    This replaced the union of item fixed-point cores, which had no rank discipline: its
    size grows with the item count, so on 35 items it kept 34-43 columns. The score matrix
    was then rank-deficient, Omega became a near pseudo-inverse, and the screen collapsed
    onto plain Mahalanobis on the raw responses (r 0.989-0.998 across encoders, 151 of 152
    consensus outliers also flagged by the response-only screen). Horn's parallel analysis
    on the undropped projections supports 3-5 dimensions for HRS and 4-12 for All of Us.
    """
    anchored = {c for c in range(Ztil.shape[1]) if (np.abs(Ztil[:, c]) >= band[c]).any()}
    modal = {int(np.argmax(np.abs(Ztil[i]))) for i in range(Ztil.shape[0])}
    return sorted(anchored | modal)
```
