## Supplemental code for "Scoring respondents on a semantic frame of reference identifies a group that is burdened across many questionnaires and reaches care less easily": build_aou_poolw_projection.py

```
#!/usr/bin/env python
"""Build the pool-W projection package for the AoU screen (LOCAL, non-sensitive).

For each encoder, project the 151 AoU items onto the FIXED pool basis and keep the
coverage-honest subspace, mirroring the HRS screen. Outputs small matrices to upload to
the Workbench, where the screen (X_keyed @ P_AoU[:, kept]) runs on the response matrix
that never leaves.

  P_AoU = (E_aou - mu_pool) @ W_pool^T          # 151 x k, rows in canonical item order
  Ztil  = (P_AoU - mu_scores) / sd_scores
  kept  = _common.keep_columns(Ztil, band)      # anchors the component OR is its modal item

The keep rule (settled 2026-08-19, _common.keep_columns) applies to both batteries. It
replaced the union of item fixed-point cores, which has no rank discipline -- harmless on
151 items but degenerate on the 35-item HRS battery, where it collapsed the screen onto
plain Mahalanobis on the raw responses. Using one rule for the cohort and another for the
transfer would not be defensible, so All of Us moves too.

Writes to outputs_pool/aou_screen_poolw/:
  P_AoU_<enc>.npy      (151 x |kept|, canonical item order)
  kept_cols_<enc>.npy  (pool component ids kept, int)
  item_order.csv       (151 item ids + construct, the shared row order)
  column_labels.csv    (per enc/kept component: n items that put it there + its construct profile,
                        reported whole -- no modal label and no purity, both retired by DEFINITIONS.md)

Run: CUDA_VISIBLE_DEVICES="" /data_gpu5/ROSS_Proj/env/bin/python build_aou_poolw_projection.py
"""
import os, glob, numpy as np, pandas as pd
from collections import Counter

from _common import keep_columns, pool_bands

HERE = os.path.dirname(os.path.abspath(__file__))
FITS = os.path.join(HERE, "outputs_pool", "fits")
GP = os.path.join(HERE, "..", "global_pool")
POOL_IDS = pd.read_csv(os.path.join(GP, "pool_processed.csv"))["item_id"].tolist()
OUT = os.path.join(HERE, "outputs_pool", "aou_screen_poolw")
os.makedirs(OUT, exist_ok=True)

ENCS = ["gte-large-en-v1.5", "bge-m3", "bge-large-en-v1.5",
        "all-mpnet-base-v2", "e5-large-v2", "qwen3-embedding-0.6b"]

cm = pd.read_csv(os.path.join(HERE, "construct_map_151.csv"))
canon = cm["item"].astype(str).tolist()                 # canonical 151-item order
i2con = dict(zip(cm["item"].astype(str), cm["construct"]))
cm[["item", "construct", "instrument"]].to_csv(os.path.join(OUT, "item_order.csv"), index=False)


def fit_dir(enc):
    d = glob.glob(os.path.join(FITS, f"{enc}__k*_ls1.0"))
    assert len(d) == 1, (enc, d)
    return d[0]


label_rows = []
print(f"{'encoder':22s} {'k':>3s} {'kept':>4s} {'empty':>5s}  empty components")
for enc in ENCS:
    fd = fit_dir(enc)
    W = np.load(os.path.join(fd, "loadings.npy"))
    mu = np.load(os.path.join(fd, "mean.npy"))
    Zp = np.load(os.path.join(fd, "scores.npy"))
    mu_s, sd = Zp.mean(0), Zp.std(0)

    a = np.load(os.path.join(HERE, "data", f"core151_items_{enc}.npz"), allow_pickle=True)
    aitems = [str(x) for x in a["items"]]
    pos = {it: j for j, it in enumerate(aitems)}
    E = a["vectors"][[pos[it] for it in canon]]         # reorder to canonical

    P = (E - mu) @ W.T                                  # 151 x k
    Ztil = (P - mu_s) / sd
    band = pool_bands(enc, (Zp - mu_s) / sd, POOL_IDS)
    kept = keep_columns(Ztil, band)
    empty = [c for c in range(W.shape[0]) if c not in set(kept)]
    modal_of = [int(np.argmax(np.abs(Ztil[i]))) for i in range(len(canon))]
    # a component is described by the items that put it in the kept set, the same rule
    sel = {c: [i for i in range(len(canon)) if modal_of[i] == c or abs(Ztil[i, c]) >= band[c]]
           for c in kept}

    np.save(os.path.join(OUT, f"P_AoU_{enc}.npy"), P[:, kept].astype(np.float32))
    np.save(os.path.join(OUT, f"kept_cols_{enc}.npy"), np.array(kept, dtype=int))

    for c in kept:
        cnt = Counter(i2con[canon[i]] for i in sel[c])
        label_rows.append({"encoder": enc, "component": c, "n_items": len(sel[c]),
                           "construct_profile": "; ".join(f"{k}:{v}" for k, v in cnt.most_common())})
    print(f"{enc:22s} {W.shape[0]:3d} {len(kept):4d} {len(empty):5d}  "
          + ", ".join(f"C{c}" for c in empty))

pd.DataFrame(label_rows).to_csv(os.path.join(OUT, "column_labels.csv"), index=False)
print("\nwrote package ->", OUT)
print("files:", sorted(os.listdir(OUT)))
```
