## Supplemental code for "Scoring respondents on a semantic frame of reference identifies a group that is burdened across many questionnaires and reaches care less easily": build_construct_map.py

```
#!/usr/bin/env python
"""Build the a-priori construct taxonomy for the 151 selected All of Us items.

Each item is assigned, by reading its prompt text, to exactly one *construct* (the specific
attribute the item asks about), and each construct rolls up into a broader *domain*. Assignments
were made from item content and cross-checked against each instrument's published (sub)scale
structure (e.g. the BFI-2-XS items recover its five validated trait factors). See main-text
\S3.1 and Methods.

Outputs (examples/allofus/):
  construct_map_151.csv     item, instrument, construct, domain, prompt   (151 rows; authoritative)

Also prints (to stdout):
  - a coverage check (151 items, no gaps, no duplicates),
  - the condensed main-text LaTeX table (domain -> construct -> n -> example item),
  - the domain/construct summary.

Run from examples/allofus/:
  python build_construct_map.py
"""
import csv
import os
from collections import OrderedDict

HERE = os.path.dirname(os.path.abspath(__file__))


def load():
    enc = list(csv.DictReader(open(os.path.join(HERE, "encoding_map_all151.csv"))))
    instr = {}
    for r in enc:
        instr.setdefault(r["concept_id"], r["instrument"])
    rows = list(csv.DictReader(open(os.path.join(HERE, "core_items_151.csv"))))
    i2instr = {r["item"]: instr.get(r["concept_id"], "?") for r in rows}
    prompt = {r["item"]: r["prompt"] for r in rows}
    return i2instr, prompt


# ---- the taxonomy: construct -> (domain, [items]) -------------------------------------------
# Order is the manuscript display order (domains grouped; constructs within a domain).
TAXONOMY = OrderedDict()


def add(construct, domain, items):
    TAXONOMY[construct] = (domain, items)


add("Everyday discrimination", "Discrimination & unfair treatment",
    [f"sdoh_eds_{i}" for i in range(1, 10)])
add("Healthcare discrimination", "Discrimination & unfair treatment",
    [f"sdoh_dms_{i}" for i in range(1, 8)])

add("Depression", "Depression & anxiety",
    [f"phq9_{i}" for i in range(1, 9)] + ["mhqukb_5", "mhqukb_6"])   # phq9_9 -> suicidality
add("Generalized anxiety", "Depression & anxiety",
    [f"gad7_{i}" for i in range(1, 8)] + ["worryanxiety"])
add("Panic & phobia", "Depression & anxiety",
    ["cidi5_16", "cidi5_27", "cidi5_30"])
add("Perceived stress", "Depression & anxiety",
    [f"sdoh_cpss_{i}" for i in range(1, 11)])

add("Suicidality & self-harm", "Suicidality & self-harm",
    ["ss_1", "ss_2", "ss_3", "phq9_9"])

add("Psychosis", "Severe mental illness",
    ["cidi5_21", "cidi5_22", "cidi5_23", "mhqukb_49"])
add("Mania", "Severe mental illness",
    ["mhqukb_43", "mhqukb_44"])

add("Body-focused repetitive behavior", "Compulsive behaviors",
    ["cidi5_25", "cidi5_26"])

add("Childhood adversity", "Trauma & adversity",
    ["ace_1", "ace_2", "ace_3", "ace_5", "ace_6", "ace_7", "ace_8", "ace_9", "ace_10", "ace_11"])
add("Interpersonal & sexual violence", "Trauma & adversity",
    ["mhqukb_34", "mhqukb_35", "mhqukb_36", "mhqukb_37"])
add("Life-threatening events", "Trauma & adversity",
    ["mhqukb_41", "cidi5_5"])

add("Extraversion", "Personality (Big Five)", ["bfi2xs_1", "bfi2xs_11", "bfi2xs_6"])
add("Agreeableness", "Personality (Big Five)", ["bfi2xs_12", "bfi2xs_2", "bfi2xs_7"])
add("Conscientiousness", "Personality (Big Five)", ["bfi2xs_3", "bfi2xs_13", "bfi2xs_8"])
add("Negative emotionality", "Personality (Big Five)", ["bfi2xs_4", "bfi2xs_14", "bfi2xs_9"])
add("Open-mindedness", "Personality (Big Five)", ["bfi2xs_10", "bfi2xs_5", "bfi2xs_15"])

add("Loneliness", "Social connection",
    ["sdoh_ucla_ls8_11", "sdoh_ucla_ls8_14", "sdoh_ucla_ls8_15", "sdoh_ucla_ls8_17",
     "sdoh_ucla_ls8_18", "sdoh_ucla_ls8_2", "sdoh_ucla_ls8_3", "sdoh_ucla_ls8_9"])
add("Perceived social support", "Social connection",
    [f"sdoh_mos_ss_{i}" for i in range(1, 9)])

add("Neighborhood disorder & safety", "Neighborhood & physical environment",
    [f"nds_{i}" for i in range(1, 14)] + ["ips_16"])
add("Neighborhood social cohesion", "Neighborhood & physical environment",
    [f"scns_{i}" for i in range(1, 5)])

add("Substance use", "Substance use",
    ["alcohol_alcoholparticipant", "cigarsmoking_cigarsmokeparticipant",
     "electronicsmoking_electricsmokeparticipant", "hookahsmoking_hookahsmokeparticipant",
     "smokelesstobacco_smokelesstobaccoparticipant", "smoking_100cigslifetime"])

add("Adult ADHD", "Attention / ADHD", [f"asrs_{i}" for i in range(1, 7)])

add("Daily spiritual experience", "Spirituality & religion", [f"bmmrs_{i}" for i in range(1, 7)])
add("Religious participation", "Spirituality & religion", ["nhs_xx"])

add("Food insecurity", "Material hardship", ["hvs_1", "hvs_2"])
add("Housing instability", "Material hardship", ["livingsituation_stablehouseconcern"])

add("Subjective wellbeing", "General health & wellbeing", ["mhqukb_57", "mhqukb_58"])
add("Self-rated health", "General health & wellbeing",
    ["overallhealth_generalmentalhealth", "overallhealth_emotionalproblem7days",
     "overallhealth_socialsatisfaction"])


# instrument-code -> short display name (matches main-text Table 1 / part1_main_figures DISPLAY)
DISPLAY = {
    "UCLA-V3": "ULS-8", "CIDI-5": "WMH-CIDI", "MOS-SS": "mMOS-SS", "ASRS-v1.1": "ASRS",
    "BMMRS-DSE": "DSES", "nds (adapted)": "NDS", "scns (adapted)": "SCNS", "ACE-BRFSS": "ACE",
    "PROMIS-Global": "PROMIS", "single item": "single item",
}


def disp(code):
    return DISPLAY.get(code, code)


# a short, readable example item per construct (for the main-text table); falls back to shortest.
EXAMPLE = {
    "Everyday discrimination": "sdoh_eds_1",
    "Healthcare discrimination": "sdoh_dms_4",
    "Depression": "phq9_2",
    "Generalized anxiety": "gad7_1",
    "Panic & phobia": "cidi5_16",
    "Perceived stress": "sdoh_cpss_2",
    "Suicidality & self-harm": "ss_2",
    "Psychosis": "cidi5_21",
    "Mania": "mhqukb_43",
    "Body-focused repetitive behavior": "cidi5_26",
    "Childhood adversity": "ace_8",
    "Interpersonal & sexual violence": "mhqukb_34",
    "Life-threatening events": "mhqukb_41",
    "Extraversion": "bfi2xs_11",
    "Agreeableness": "bfi2xs_2",
    "Conscientiousness": "bfi2xs_13",
    "Negative emotionality": "bfi2xs_4",
    "Open-mindedness": "bfi2xs_15",
    "Loneliness": "sdoh_ucla_ls8_2",
    "Perceived social support": "sdoh_mos_ss_7",
    "Neighborhood disorder & safety": "nds_8",
    "Neighborhood social cohesion": "scns_3",
    "Substance use": "smoking_100cigslifetime",
    "Adult ADHD": "asrs_3",
    "Daily spiritual experience": "bmmrs_3",
    "Religious participation": "nhs_xx",
    "Food insecurity": "hvs_2",
    "Housing instability": "livingsituation_stablehouseconcern",
    "Subjective wellbeing": "mhqukb_57",
    "Self-rated health": "overallhealth_generalmentalhealth",
}

_SPECIAL = [("\\", r"\textbackslash{}"), ("&", r"\&"), ("%", r"\%"), ("$", r"\$"),
            ("#", r"\#"), ("_", r"\_"), ("{", r"\{"), ("}", r"\}"), ("~", r"\textasciitilde{}"),
            ("^", r"\textasciicircum{}")]


def tex(s):
    s = str(s)
    for a, b in _SPECIAL:
        s = s.replace(a, b)
    return s


def shorten(p, w=70):
    """Return (tex-escaped, possibly-truncated) cell text with a trailing \\dots if cut."""
    p = p.replace("\n", " ").strip().rstrip(".")
    if len(p) <= w:
        return tex(p)
    return tex(p[:w].rsplit(" ", 1)[0]) + r"\dots"


def main():
    i2instr, prompt = load()

    # coverage check
    assigned = [it for (_, items) in TAXONOMY.values() for it in items]
    assert len(assigned) == len(set(assigned)) == 151, (
        f"coverage error: {len(assigned)} assigned, {len(set(assigned))} distinct")
    assert set(assigned) == set(prompt), "items missing or extra vs core_items_151"

    # ---- write authoritative CSV ----
    out_csv = os.path.join(HERE, "construct_map_151.csv")
    with open(out_csv, "w", newline="") as f:
        w = csv.writer(f)
        w.writerow(["item", "instrument", "construct", "domain", "prompt"])
        for construct, (domain, items) in TAXONOMY.items():
            for it in items:
                w.writerow([it, i2instr[it], construct, domain, prompt[it]])

    # ---- domain -> constructs (display order, preserving first appearance) ----
    by_domain = OrderedDict()
    for construct, (domain, items) in TAXONOMY.items():
        by_domain.setdefault(domain, []).append((construct, items))

    # ---- emit condensed main-text LaTeX table ----
    # longtable, not table: 30 rows over 14 domains is taller than one page, so a
    # non-breaking float would be clipped. Needs \usepackage{longtable} + \usepackage{array}
    # and \newcolumntype{L}[1]{>{\raggedright\arraybackslash}p{#1}} in the preamble.
    print("% ---- main-text construct table (paste into main_npj.tex, \S3.2) ----")
    print(r"{\small")
    print(r"\setlength{\LTcapwidth}{\textwidth}")
    print(r"\begin{longtable}{@{}L{3.1cm} L{3.3cm} r L{7.0cm}@{}}")
    print(r"\caption{\textbf{The 151 items organized into 30 constructs and 14 domains.} "
          r"Each item was assigned to one construct from its prompt text and cross-checked "
          r"against the source instrument's published structure, and constructs roll up into "
          r"domains. Counts are items per construct. The full item-level assignment is in "
          r"Supplementary Data~1 (\texttt{items\_full.csv}).}")
    print(r"\label{tab:constructs}\\")
    print(r"\toprule")
    print(r"Domain & Construct & $n$ & Example item \\")
    print(r"\midrule")
    print(r"\endfirsthead")
    print(r"\multicolumn{4}{@{}l}{\textit{Table \thetable\ (continued)}}\\")
    print(r"\toprule")
    print(r"Domain & Construct & $n$ & Example item \\")
    print(r"\midrule")
    print(r"\endhead")
    print(r"\bottomrule")
    print(r"\endlastfoot")
    for di, (domain, cs) in enumerate(by_domain.items()):
        if di:
            print(r"\midrule")                       # horizontal rule between domains
        for j, (construct, items) in enumerate(cs):
            dcell = tex(domain) if j == 0 else ""
            ex = shorten(prompt[EXAMPLE[construct]])
            print(f"{dcell} & {tex(construct)} & {len(items)} & \\footnotesize {ex} \\\\")
    print(r"\end{longtable}}")

    print(f"\n% wrote {out_csv}  ({len(assigned)} items, "
          f"{len(TAXONOMY)} constructs, {len(by_domain)} domains)")


if __name__ == "__main__":
    main()
```
