## Supplemental code for "Scoring respondents on a semantic frame of reference identifies a group that is burdened across many questionnaires and reaches care less easily": build_reference_basis_package.py

```
#!/usr/bin/env python
"""Package the fixed pool reference basis so any battery can be placed on it elsewhere.

The reference basis is the portable object: it is computed once on the 577-item pool and never
refitted for a new battery. This script copies everything a downstream screen needs into a single
self-contained folder, so a Workbench (or any other machine) needs no access to this repository.

Per encoder it writes ``reference_basis/<enc>.npz`` holding

    W          (k, D)   component loadings of the pool fit
    mu         (D,)     pool embedding mean, the centering for the projection
    sigma      (k,)     pool per-component score standard deviations, the calibration
    mu_scores  (k,)     pool per-component score means (zero by construction, kept for exactness)
    band       (k,)     pool core-item band per component: the smallest standardized loading among
                        the pool items that anchor that component's core. This is what the keep
                        rule needs and it cannot be recovered from W, mu and sigma alone.
    k, dim, encoder, fit_dir, lambda_score, n_pool_items

and one shared ``pool_components.csv`` describing each component in the pool's own vocabulary
(core items and their construct profile), for interpretation on the pool side.

Placing a battery S on this basis, which is all a downstream script has to do:

    P_S = (E_S - mu) @ W.T                 # items x k, E_S = the battery's item embeddings
    Z_S = (P_S - mu_scores) / sigma        # in pool SD units
    kept = keep_columns(Z_S, band)         # anchors a component, or is its modal component

Run: CUDA_VISIBLE_DEVICES="" /data_gpu5/ROSS_Proj/env/bin/python build_reference_basis_package.py
"""
import csv
import glob
import json
import os
import re
import shutil
from collections import Counter

import numpy as np
import pandas as pd

HERE = os.path.dirname(os.path.abspath(__file__))
REPO = os.path.abspath(os.path.join(HERE, "..", ".."))
GP = os.path.join(HERE, "..", "global_pool")
OUT = os.path.join(REPO, "all_of_us_codes", "reference_basis")
EMB_OUT = os.path.join(REPO, "all_of_us_codes", "item_embeddings")

ENCS = ["gte-large-en-v1.5", "bge-m3", "bge-large-en-v1.5",
        "all-mpnet-base-v2", "e5-large-v2", "qwen3-embedding-0.6b"]


def main():
    os.makedirs(OUT, exist_ok=True)
    os.makedirs(EMB_OUT, exist_ok=True)
    pool = pd.read_csv(os.path.join(GP, "pool_processed.csv"))
    pool_ids = pool["item_id"].tolist()
    pcon = dict(zip(pool.item_id, pool.construct))
    pos = {it: j for j, it in enumerate(pool_ids)}

    rows, manifest = [], []
    for enc in ENCS:
        fit = glob.glob(os.path.join(HERE, "outputs_pool", "fits", f"{enc}__k*_ls1.0"))[0]
        W = np.load(os.path.join(fit, "loadings.npy"))
        mu = np.load(os.path.join(fit, "mean.npy"))
        S = np.load(os.path.join(fit, "scores.npy"))
        mu_s, sigma = S.mean(0), S.std(0)
        Zpool = (S - mu_s) / sigma

        cores = {int(r["component"]): str(r["items"]).split() for r in csv.DictReader(
            open(os.path.join(HERE, "outputs_pool", "fixedpoint_cores", f"{enc}_cores.csv")))}
        band = np.array([min(abs(Zpool[pos[h], c]) for h in cores[c]) for c in range(W.shape[0])])

        np.savez_compressed(
            os.path.join(OUT, f"{enc}.npz"),
            W=W.astype(np.float64), mu=mu.astype(np.float64),
            sigma=sigma.astype(np.float64), mu_scores=mu_s.astype(np.float64),
            band=band.astype(np.float64),
            k=W.shape[0], dim=W.shape[1], encoder=enc,
            fit_dir=os.path.basename(fit), lambda_score=1.0, n_pool_items=len(pool_ids))

        for c in range(W.shape[0]):
            prof = Counter(pcon[i] for i in cores[c])
            rows.append({"encoder": enc, "component": c, "n_core_items": len(cores[c]),
                         "band": round(float(band[c]), 4),
                         "core_items": " ".join(cores[c]),
                         "construct_profile": "; ".join(f"{k}:{v}" for k, v in prof.most_common())})

        # the battery's item embeddings, so a downstream host needs no encoder installed
        src = os.path.join(HERE, "data", f"core151_items_{enc}.npz")
        shutil.copy(src, os.path.join(EMB_OUT, f"core151_items_{enc}.npz"))
        d = np.load(src, allow_pickle=True)
        manifest.append({"encoder": enc, "k": int(W.shape[0]), "embedding_dim": int(W.shape[1]),
                         "pool_items": len(pool_ids), "battery_items": int(len(d["items"])),
                         "band_min": round(float(band.min()), 3),
                         "band_median": round(float(np.median(band)), 3),
                         "band_max": round(float(band.max()), 3)})

    pd.DataFrame(rows).to_csv(os.path.join(OUT, "pool_components.csv"), index=False)
    pd.DataFrame(manifest).to_csv(os.path.join(OUT, "manifest.csv"), index=False)
    print(pd.DataFrame(manifest).to_string(index=False))
    tot = sum(os.path.getsize(os.path.join(OUT, f)) for f in os.listdir(OUT))
    emb = sum(os.path.getsize(os.path.join(EMB_OUT, f)) for f in os.listdir(EMB_OUT))
    print(f"\nwrote {OUT}  ({tot/1e6:.1f} MB)")
    print(f"wrote {EMB_OUT}  ({emb/1e6:.1f} MB)")


if __name__ == "__main__":
    main()
```
