## Supplemental code for "Scoring respondents on a semantic frame of reference identifies a group that is burdened across many questionnaires and reaches care less easily": construct_splitting.py

```
#!/usr/bin/env python
"""Do items sharing a documented construct split across components, and is the split reproducible?

A documented construct is a coarse yardstick: one label per instrument or validated subscale, blind to
variation between the items inside it. Agreement with it above chance shows the basis tracks meaning at
the resolution the label has (core_separation.py). This script measures the other half -- where the
basis resolves finer than the label -- and, crucially, whether that finer structure is reproducible
rather than noise.

For each documented construct with at least MIN_ITEMS core-anchoring items:

    n_components  how many distinct component cores its items occupy under one encoder
    split         n_components > 1

Reproducibility is the question that decides whether a split is meaning or noise. Two encoders agree
on a construct when both split it or both do not. We report:

    split_rate        share of encoders that split the construct
    unanimous         all six agree (split or not)
    partition_agree   for constructs split under both encoders, the Rand index between the two
                      partitions its items induce (adjusted for chance), averaged over the 15 encoder
                      pairs, against a null that shuffles items across that construct's components

A construct whose items split the SAME WAY under six independently trained encoders is being resolved
by something in the item text. One that splits differently every time is not.

Reads saved artefacts only:
    outputs_pool/fixedpoint_cores/<enc>_cores.csv     core items per component
    ../global_pool/pool_processed.csv                 documented construct per item

Out: outputs_pool/construct_splitting.csv
Run: CUDA_VISIBLE_DEVICES="" /data_gpu5/ROSS_Proj/env/bin/python construct_splitting.py
"""
import csv
import itertools
import os
from collections import defaultdict

import numpy as np
import pandas as pd
from sklearn.metrics import adjusted_rand_score

HERE = os.path.dirname(os.path.abspath(__file__))
POOL = os.path.join(HERE, "outputs_pool")
GP = os.path.join(HERE, "..", "global_pool")
ENCODERS = ["bge-m3", "bge-large-en-v1.5", "all-mpnet-base-v2", "e5-large-v2",
            "gte-large-en-v1.5", "qwen3-embedding-0.6b"]
MIN_ITEMS = 3
NPERM = 200
SEED = 0


def item_components(enc):
    """item -> set of components whose core contains it."""
    m = defaultdict(set)
    with open(os.path.join(POOL, "fixedpoint_cores", f"{enc}_cores.csv")) as f:
        for r in csv.DictReader(f):
            for it in str(r["items"]).split():
                m[it].add(int(r["component"]))
    return m


def ari(a, b):
    """Adjusted Rand index: the Rand index corrected for chance agreement (0 = chance, 1 = identical)."""
    if len(a) < 2:
        return np.nan
    return float(adjusted_rand_score(a, b))


def main():
    pp = pd.read_csv(os.path.join(GP, "pool_processed.csv"))
    con = dict(zip(pp.item_id.astype(str), pp.construct.astype(str)))
    by_con = defaultdict(list)
    for h, c in con.items():
        by_con[c].append(h)

    maps = {e: item_components(e) for e in ENCODERS}

    rows = []
    for cname, items in sorted(by_con.items()):
        # restrict to items that anchor some core under every encoder, so the comparison is like for like
        anchored = [h for h in items if all(maps[e].get(h) for e in ENCODERS)]
        if len(anchored) < MIN_ITEMS:
            continue
        # one label per item per encoder: its lowest-numbered core component (items core at most a few)
        lab = {e: [min(maps[e][h]) for h in anchored] for e in ENCODERS}
        ncomp = {e: len(set(lab[e])) for e in ENCODERS}
        splits = {e: ncomp[e] > 1 for e in ENCODERS}
        both = [(a, b) for a, b in itertools.combinations(ENCODERS, 2) if splits[a] and splits[b]]
        ri = [ari(lab[a], lab[b]) for a, b in both]
        rng = np.random.RandomState(SEED)
        null = []
        for a, b in both:
            for _ in range(NPERM // max(1, len(both))):
                null.append(ari(lab[a], list(rng.permutation(lab[b]))))
        rows.append(dict(construct=cname, n_items=len(anchored),
                         split_rate=float(np.mean(list(splits.values()))),
                         unanimous=bool(all(splits.values()) or not any(splits.values())),
                         median_components=float(np.median(list(ncomp.values()))),
                         ari=float(np.mean(ri)) if ri else np.nan,
                         ari_null=float(np.mean(null)) if null else np.nan))

    df = pd.DataFrame(rows).sort_values(["split_rate", "n_items"], ascending=[False, False])
    p = os.path.join(POOL, "construct_splitting.csv")
    df.to_csv(p, index=False)

    n = len(df)
    always = int((df.split_rate == 1.0).sum())
    never = int((df.split_rate == 0.0).sum())
    print(f"{n} documented constructs with >= {MIN_ITEMS} core-anchoring items under all six encoders\n")
    print(f"  split under every encoder : {always}")
    print(f"  split under none          : {never}")
    print(f"  mixed                     : {n - always - never}")
    print(f"  unanimous either way      : {int(df.unanimous.sum())} of {n}")
    sub = df[df['ari'].notna()]
    print(f"\nfor constructs split under two encoders, how alike are the two splits (adjusted Rand):")
    print(f"  observed {sub['ari'].mean():.3f}   shuffled null {sub['ari_null'].mean():.3f}   "
          f"n = {len(sub)} constructs")
    print("\nmost-split constructs:")
    print(df.head(10).to_string(index=False, float_format=lambda v: f"{v:6.3f}"))
    print(f"\nwrote {p}")


if __name__ == "__main__":
    main()
```
