## Supplemental code for "Scoring respondents on a semantic frame of reference identifies a group that is burdened across many questionnaires and reaches care less easily": content_grammar_manifest.py

```
#!/usr/bin/env python
"""Reproducibility manifest for the §3.3.2 content-vs-grammar sub-study.

Writes an explicit, human-readable record of the experiment's two random choices —
the 30 stratified SEED items and the deterministic seed->donor SWAPS — and verifies
that the recomputed swaps match the donors actually used in the released variants.

The only stochastic step in the whole sub-study is donor selection, and it is fully
deterministic: for each seed, `generate_variants.py` draws `--n-variants` donor items
(without replacement) from the items of a *different* instrument, using a per-item RNG
`random.Random(seed + item_index)` over the fixed 151-item order of core_items_151.csv.
The same donor set supplies the transferred frames (Cell A) and the substituted topics
(Cell B). Re-running this script reproduces the swaps byte-for-byte; it then cross-checks
them against the donor_item column of the released generated_items_strat.csv.

Run from examples/allofus/:
  /data_gpu5/ROSS_Proj/env/bin/python content_grammar_manifest.py
Writes outputs_core151/content_grammar/seeds_and_swaps.csv  and prints a PASS/FAIL check.
"""
import csv
import os
import random

from generate_variants import load_items, pick_donors

CG = "outputs_core151/content_grammar"
SEEDS_FILE = os.path.join(CG, "seeds_stratified.txt")
VARIANTS_CSV = os.path.join(CG, "generated_items_strat.csv")
OUT = os.path.join(CG, "seeds_and_swaps.csv")
SEED = 0           # global RNG seed used for the released run (rng_seed column == 0)
N_VARIANTS = 10    # donors per seed (== variants per cell)


def recompute_swaps():
    """Reproduce the deterministic per-seed donor lists exactly as generate_variants.py did."""
    order, prompts, inst = load_items()
    # donors are pre-drawn per item over the FULL 151-item order, RNG = Random(SEED + index)
    donors = {s: pick_donors(s, order, inst, N_VARIANTS, random.Random(SEED + i))
              for i, s in enumerate(order)}
    seeds = [l.strip() for l in open(SEEDS_FILE) if l.strip()]
    seeds = [s for s in order if s in set(seeds)]          # keep canonical order
    return seeds, donors, inst, prompts


def verify(seeds, donors):
    """Check recomputed donors match the donor_item column actually used per seed×cell."""
    rows = list(csv.DictReader(open(VARIANTS_CSV)))
    used = {}                                              # (seed, cell) -> [donor per variant]
    for r in rows:
        used.setdefault((r["seed_item"], r["cell"]), []).append(r["donor_item"])
    ok = True
    for s in seeds:
        for cell in ("A", "B"):
            got = used.get((s, cell), [])
            exp = donors[s][:len(got)]
            if got != exp:
                ok = False
                print(f"  MISMATCH {s} cell {cell}: used={got} expected={exp}")
    return ok


def main():
    seeds, donors, inst, prompts = recompute_swaps()
    with open(OUT, "w", newline="") as f:
        w = csv.writer(f)
        w.writerow(["seed_item", "seed_instrument", "donor_rank", "donor_item",
                    "donor_instrument", "used_as_frame_cellA_and_topic_cellB"])
        for s in seeds:
            for rank, d in enumerate(donors[s]):
                w.writerow([s, inst[s], rank, d, inst[d], "yes"])
    n_pairs = sum(len(donors[s]) for s in seeds)
    print(f"-> {OUT}  ({len(seeds)} seeds x {N_VARIANTS} donors = {n_pairs} swaps; "
          f"global RNG seed={SEED})")
    ok = verify(seeds, donors)
    print("VERIFY recomputed swaps == donors used in generated_items_strat.csv:",
          "PASS" if ok else "FAIL")


if __name__ == "__main__":
    main()
```
