## Supplemental code for "Scoring respondents on a semantic frame of reference identifies a group that is burdened across many questionnaires and reaches care less easily": content_grammar_poolbasis.py

```
#!/usr/bin/env python
"""The content-versus-syntax test, re-run on the POOL reference basis.

The original run (crossencoder_content_grammar.py) projected the 151 items and their 3,020
counterfactual variants onto the retired own basis selected by random restarts. The paper no longer
uses that basis, so this script repeats the measurement on the fixed pool basis, which is the object
the manuscript actually describes.

An item or a variant is placed exactly as a battery item is placed (Methods, "Placing a battery on the
basis"): embed with the encoder, subtract the pool mean, project on the pool components, divide
column-wise by the pool standard deviations. The row is then L2-normalized so the comparison is a
cosine, as in the original test. `--raw` skips the sigma division, as a check that the result is not an
artefact of the calibration.

For each variant we record the cosine to its CONTENT source and to its SYNTAX source:
    cell A (syntax variant)  content = seed,  syntax = donor
    cell B (content variant) content = donor, syntax = seed
and report per encoder the median of each and the content-minus-syntax gap, overall and split by
whether the seed's syntax family is content-bearing (the jingle case).

Out: outputs_pool/content_grammar_poolbasis[_raw].csv
Run: CUDA_VISIBLE_DEVICES="" /data_gpu5/ROSS_Proj/env/bin/python content_grammar_poolbasis.py
"""
import argparse, csv, glob, os
import numpy as np
import pandas as pd

HERE = os.path.dirname(os.path.abspath(__file__))
POOL = os.path.join(HERE, "outputs_pool")
DATA = os.path.join(HERE, "data")
VARCSV = os.path.join(HERE, "outputs_core151", "content_grammar_v2", "generated_items_v2.csv")
VARDIR = os.path.join(HERE, "data", "variants_strat_v2")
EMBEDDERS = ["bge-m3", "bge-large-en-v1.5", "all-mpnet-base-v2", "e5-large-v2",
             "gte-large-en-v1.5", "qwen3-embedding-0.6b"]


def pool_basis(enc):
    fit = glob.glob(os.path.join(POOL, "fits", f"{enc}__k*_ls1.0"))[0]
    mean = np.load(os.path.join(fit, "mean.npy"))
    W = np.load(os.path.join(fit, "loadings.npy"))       # (k, D)
    sigma = np.load(os.path.join(fit, "scores.npy")).std(0)   # pool column sigmas
    return mean, W, sigma


def place(E, mean, W, sigma, raw=False):
    P = (np.asarray(E, float) - mean) @ W.T
    if not raw:
        P = P / sigma
    return P / (np.linalg.norm(P, axis=1, keepdims=True) + 1e-12)


def main():
    ap = argparse.ArgumentParser()
    ap.add_argument("--raw", action="store_true", help="skip the pool-sigma division")
    args = ap.parse_args()

    cb = {r["item"]: bool(int(r["content_bearing"]))
          for r in csv.DictReader(open(os.path.join(HERE, "frame_map_151.csv")))}
    rows = list(csv.DictReader(open(VARCSV)))
    by_id = {r["variant_id"]: r for r in rows}

    out = []
    for enc in EMBEDDERS:
        mean, W, sigma = pool_basis(enc)
        orig = np.load(os.path.join(DATA, f"core151_items_{enc}.npz"), allow_pickle=True)
        oitems = list(orig["items"].astype(str))
        Po = place(orig["vectors"], mean, W, sigma, args.raw)
        idx = {it: i for i, it in enumerate(oitems)}
        var = np.load(os.path.join(VARDIR, f"{enc}.npz"), allow_pickle=True)
        vids = list(var["items"].astype(str))
        Pv = place(var["vectors"], mean, W, sigma, args.raw)

        for vi, vid in enumerate(vids):
            r = by_id.get(vid)
            if r is None:
                continue
            seed, donor, cell = r["seed_item"], r["donor_item"], r["cell"]
            if seed not in idx or donor not in idx:
                continue
            cs, cd = float(Pv[vi] @ Po[idx[seed]]), float(Pv[vi] @ Po[idx[donor]])
            content, syntax = (cs, cd) if cell != "B" else (cd, cs)
            out.append(dict(variant_id=vid, cell=cell, seed_item=seed, donor_item=donor,
                            seed_content_bearing=cb.get(seed, False), encoder=enc,
                            cos_content=content, cos_syntax=syntax,
                            margin=content - syntax, win=int(content > syntax)))

    df = pd.DataFrame(out)
    tag = "_raw" if args.raw else ""
    df = df[["variant_id", "cell", "seed_item", "donor_item", "seed_content_bearing",
             "encoder", "cos_content", "cos_syntax", "margin", "win"]]
    p = os.path.join(POOL, f"content_grammar_poolbasis{tag}.csv")
    df.to_csv(p, index=False)

    def block(sub, label):
        g = sub.groupby("encoder").agg(n=("margin", "size"), cos_content=("cos_content", "median"),
                                       cos_syntax=("cos_syntax", "median"),
                                       gap=("margin", "median"), win=("win", "mean"))
        print(f"\n{label}")
        print(g.to_string(float_format=lambda v: f"{v:7.3f}"))

    print(f"{len(df)} variant-encoder placements, {df.variant_id.nunique()} variants"
          + (" (raw, no sigma division)" if args.raw else ""))
    block(df[df.cell != "B"], "SYNTAX VARIANTS (cell A: seed's content in a donor's syntax)")
    block(df[df.cell == "B"], "CONTENT VARIANTS (cell B: donor's content in the seed's syntax)")
    block(df[(df.cell == "B") & (~df.seed_content_bearing.astype(bool))], "  cell B, neutral seed syntax")
    block(df[(df.cell == "B") & (df.seed_content_bearing.astype(bool))], "  cell B, content-bearing syntax (jingle)")
    print(f"\nwrote {p}")


if __name__ == "__main__":
    main()
```
