## Supplemental code for "Scoring respondents on a semantic frame of reference identifies a group that is burdened across many questionnaires and reaches care less easily": core_separation.py

```
#!/usr/bin/env python
"""Does the basis SEPARATE items that mean different things, not only group items that mean the same?

Grouping is shown by instruments written for one construct converging on one component. Separation is
the other half: two items that land in the same component's core should be items that mean the same
thing, and items meaning different things should end up on different components.

The measure is pairwise and pool-wide, so it says nothing about any single component:

    p_same    = P(two items share a documented construct | they share a core component)
    chance    = P(two items share a documented construct | drawn at random from the pool)
    ratio     = p_same / chance

and the reverse conditional, which is the one that states SEPARATION directly:

    p_core_same = P(two items share a core component | they share a documented construct)
    p_core_diff = P(two items share a core component | their documented constructs differ)
    sep_ratio   = p_core_same / p_core_diff

Chance is the same-construct rate over all unordered item pairs, which is fixed by the construct-size
distribution alone. A permutation null (constructs shuffled over items, cores held fixed) is reported
as well, and reproduces `chance` up to sampling error -- it is there to confirm nothing in the core
structure is doing the work.

NOT COMPONENT PURITY. Purity summarised one component with one number and was retired. This is a
single pool-wide statistic over item pairs, and it is not computed or reported per component.

Reads saved artefacts only:
    outputs_pool/fixedpoint_cores/<enc>_cores.csv     core items per component
    ../global_pool/pool_processed.csv                 documented construct per item

Out: outputs_pool/core_separation.csv
Run: CUDA_VISIBLE_DEVICES="" /data_gpu5/ROSS_Proj/env/bin/python core_separation.py
"""
import csv
import itertools
import os

import numpy as np
import pandas as pd

HERE = os.path.dirname(os.path.abspath(__file__))
POOL = os.path.join(HERE, "outputs_pool")
GP = os.path.join(HERE, "..", "global_pool")
ENCODERS = ["bge-m3", "bge-large-en-v1.5", "all-mpnet-base-v2", "e5-large-v2",
            "gte-large-en-v1.5", "qwen3-embedding-0.6b"]
NPERM = 200
SEED = 0


def core_pairs(cores):
    """Unordered item pairs that share at least one component core."""
    pairs = set()
    for items in cores.values():
        for a, b in itertools.combinations(sorted(set(items)), 2):
            pairs.add((a, b))
    return sorted(pairs)


def main():
    pp = pd.read_csv(os.path.join(GP, "pool_processed.csv"))
    con = dict(zip(pp.item_id.astype(str), pp.construct.astype(str)))
    ids = [str(h) for h in pp.item_id]

    # chance: same-construct rate over ALL unordered pairs of pool items
    sizes = pd.Series([con[h] for h in ids]).value_counts().values
    n = len(ids)
    all_pairs = n * (n - 1) / 2
    same_pairs = float(sum(s * (s - 1) / 2 for s in sizes))
    chance = same_pairs / all_pairs

    rows = []
    for enc in ENCODERS:
        cores = {int(r["component"]): str(r["items"]).split()
                 for r in csv.DictReader(open(os.path.join(POOL, "fixedpoint_cores", f"{enc}_cores.csv")))}
        pairs = core_pairs(cores)
        same = [con[a] == con[b] for a, b in pairs]
        p_same = float(np.mean(same))

        rng = np.random.RandomState(SEED)
        vals = [con[h] for h in ids]
        null = []
        for _ in range(NPERM):
            lab = dict(zip(ids, rng.permutation(vals)))
            null.append(np.mean([lab[a] == lab[b] for a, b in pairs]))
        perm = float(np.mean(null))

        share = set(pairs) | {(b, a) for a, b in pairs}
        ns = nd = cs = cd = 0
        for a, b in itertools.combinations(ids, 2):
            sh = (a, b) in share
            if con[a] == con[b]:
                ns += 1; cs += sh
            else:
                nd += 1; cd += sh
        p_core_same, p_core_diff = cs / ns, cd / nd

        rows.append(dict(encoder=enc, n_components=len(cores), n_core_pairs=len(pairs),
                         p_same=p_same, chance=chance, perm_null=perm,
                         ratio=p_same / chance,
                         p_core_same=p_core_same, p_core_diff=p_core_diff,
                         sep_ratio=p_core_same / p_core_diff))
        print(f"{enc:24s} {len(cores):3d} components  {len(pairs):5d} core pairs   "
              f"same-construct {100*p_same:5.1f}%   chance {100*chance:4.1f}%   "
              f"permuted {100*perm:4.1f}%   ratio {p_same/chance:5.1f}x | "
              f"P(core|same) {100*p_core_same:5.1f}%  P(core|diff) {100*p_core_diff:5.2f}%  "
              f"sep {p_core_same/p_core_diff:4.1f}x", flush=True)

    df = pd.DataFrame(rows)
    p = os.path.join(POOL, "core_separation.csv")
    df.to_csv(p, index=False)
    print(f"\nacross encoders: same-construct {100*df.p_same.min():.1f}-{100*df.p_same.max():.1f}%, "
          f"chance {100*chance:.1f}%, ratio {df.ratio.min():.1f}-{df.ratio.max():.1f}x")
    print(f"separation: P(core|same) {100*df.p_core_same.min():.1f}-{100*df.p_core_same.max():.1f}%, "
          f"P(core|diff) {100*df.p_core_diff.min():.2f}-{100*df.p_core_diff.max():.2f}%, "
          f"{df.sep_ratio.min():.0f}-{df.sep_ratio.max():.0f}x")
    print(f"wrote {p}")


if __name__ == "__main__":
    main()
```
