## Supplemental code for "Scoring respondents on a semantic frame of reference identifies a group that is burdened across many questionnaires and reaches care less easily": dense_control.py

```
#!/usr/bin/env python
"""Control: does sparsity buy the interpretability, or would a dense basis do as well?

The claim R1 makes is that compressing 577 published questions to ~55 directions AND forcing each
direction to rest on a handful of questions still leaves something every component can be named from
published documentation. This script tests the second half of that claim by removing it: it fits a
DENSE basis (plain PCA, same encoder, same k, same items) and scores it by the same rule.

Design (settled 2026-08-24, before running):

  matched-size anchors   A dense component has no core, so it is given the same anchor budget as the
                         sparse basis: its top-m items by absolute standardized loading, with m the
                         mean sparse core size. Applying the sparse band rule to dense loadings would
                         select most of the pool and stack the deck.

  margin over own null   Faithfulness is "the item's documented construct appears among the constructs
                         of its core components". A basis with larger, more heterogeneous profiles
                         scores a HIGHER raw rate for free, so the raw rate cannot be compared across
                         bases. Each basis is scored against its own label-shuffle null and what is
                         compared is the margin.

Everything is read from saved artefacts, nothing is hardcoded:
    data/pool_items_<enc>.npz                          pool item embeddings
    outputs_pool/fits/<enc>__k*_ls1.0/scores.npy       the sparse pool projection
    outputs_pool/fixedpoint_cores/<enc>_cores.csv      sparse core items per component
    ../global_pool/pool_processed.csv                  item text and documented construct

Out: outputs_pool/dense_control_<enc>.json  (and a printed table)
Run: CUDA_VISIBLE_DEVICES="" /data_gpu5/ROSS_Proj/env/bin/python dense_control.py
"""
import os, glob, csv, json, sys
import numpy as np
import pandas as pd
from sklearn.decomposition import PCA

HERE = os.path.dirname(os.path.abspath(__file__))
GP = os.path.join(HERE, "..", "global_pool")

ENC = sys.argv[1] if len(sys.argv) > 1 else "gte-large-en-v1.5"
NPERM = 200
SEED = 0


def pr(v):
    s2 = float((v ** 2).sum()); s4 = float((v ** 4).sum())
    return s2 * s2 / s4 if s4 > 0 else 0.0


def fpc(v):
    """An item's core components: the fixed point of the participation ratio on its loadings."""
    idx = np.arange(len(v))
    while True:
        n = max(1, min(int(round(pr(v[idx]))), len(idx)))
        top = np.sort(idx[np.argsort(-np.abs(v[idx]))[:n]])
        if len(top) == len(idx) and (top == idx).all():
            return top
        idx = top


def load():
    pp = pd.read_csv(os.path.join(GP, "pool_processed.csv"))
    fit = glob.glob(os.path.join(HERE, "outputs_pool", "fits", f"{ENC}__k*_ls1.0"))[0]
    Zs = np.load(os.path.join(fit, "scores.npy"))
    rows = list(csv.DictReader(open(os.path.join(HERE, "outputs_pool", "fixedpoint_cores",
                                                 f"{ENC}_cores.csv"))))
    core = {int(r["component"]): str(r["items"]).split() for r in rows}
    d = np.load(os.path.join(HERE, "data", f"pool_items_{ENC}.npz"), allow_pickle=True)
    # align the embedding rows to pool_processed order by item id rather than trusting file order
    pos = {str(h): j for j, h in enumerate(d["items"])}
    X = np.asarray(d["vectors"], dtype=np.float64)[[pos[str(h)] for h in pp.item_id]]
    return pp, Zs, core, X


def constructs_of(cores, con, comps, drop=None):
    out = set()
    for c in comps:
        items = [h for h in cores[c] if h != drop]
        out.update(con[h] for h in items)
    return out


def score(ids, cores, F, con, rng, label):
    """Leave-one-out faithfulness and its label-shuffle null, for one basis."""
    anchored = {h for v in cores.values() for h in v}

    def rate(sub, lab):
        if not sub:
            return float("nan")
        return float(np.mean([lab[g] in constructs_of(cores, lab, F[g], drop=g) for g in sub]))

    out = {"labelling": label, "n_anchored": len(anchored)}
    groups = [("all", ids),
              ("anchors_a_core", [g for g in ids if g in anchored]),
              ("anchors_none", [g for g in ids if g not in anchored])]
    for name, sub in groups:
        real = rate(sub, con)
        null = float(np.mean([rate(sub, dict(zip(ids, rng.permutation(list(con.values())))))
                              for _ in range(NPERM)]))
        out[name] = {"n": len(sub), "rate": real, "chance": null, "margin": real - null}
    out["mean_core_components_per_item"] = float(np.mean([len(F[g]) for g in ids]))
    out["constructs_per_component"] = {
        "mean": float(np.mean([len({con[h] for h in v}) for v in cores.values()])),
        "median": float(np.median([len({con[h] for h in v}) for v in cores.values()])),
        "single_construct": int(sum(len({con[h] for h in v}) == 1 for v in cores.values())),
        "n_components": len(cores),
    }
    return out


def main():
    pp, Zs, sparse_core, X = load()
    con = dict(zip(pp.item_id, pp.construct))
    ids = list(pp.item_id)
    k = Zs.shape[1]
    assert X.shape[0] == len(ids), f"embedding rows {X.shape[0]} != pool items {len(ids)}"

    sizes = [len(v) for v in sparse_core.values()]
    m = int(round(float(np.mean(sizes))))

    # ---- dense basis: plain PCA, same encoder, same k, same items ----
    Zd = PCA(n_components=k, random_state=SEED).fit_transform(X)

    Zst = Zs / Zs.std(0)
    Zdt = Zd / Zd.std(0)
    dense_core = {c: [ids[j] for j in np.argsort(-np.abs(Zdt[:, c]))[:m]] for c in range(k)}

    Fs = {g: [int(c) for c in fpc(Zst[j])] for j, g in enumerate(ids)}
    Fd = {g: [int(c) for c in fpc(Zdt[j])] for j, g in enumerate(ids)}

    res = {
        "encoder": ENC, "k": k, "n_items": len(ids), "n_perm": NPERM,
        "anchor_budget": {"sparse_core_sizes_mean": float(np.mean(sizes)),
                          "sparse_core_sizes_median": float(np.median(sizes)),
                          "sparse_total_anchor_slots": int(sum(sizes)),
                          "dense_m_per_component": m,
                          "dense_total_anchor_slots": int(m * k)},
        "sparse": score(ids, sparse_core, Fs, con, np.random.RandomState(SEED), "sparse (fixed-point cores)"),
        "dense": score(ids, dense_core, Fd, con, np.random.RandomState(SEED), f"dense PCA (top-{m} anchors)"),
    }

    out = os.path.join(HERE, "outputs_pool", f"dense_control_{ENC}.json")
    json.dump(res, open(out, "w"), indent=2)

    print(f"\n{ENC}   k={k}   {len(ids)} items   anchors: sparse {sum(sizes)} slots, dense {m*k} slots (m={m})\n")
    print(f"{'':22s} {'rate':>7s} {'chance':>8s} {'margin':>8s}   n")
    for side in ("sparse", "dense"):
        print(f"  {res[side]['labelling']}")
        for g in ("all", "anchors_a_core", "anchors_none"):
            r = res[side][g]
            print(f"    {g:18s} {100*r['rate']:6.1f}% {100*r['chance']:7.1f}% {100*r['margin']:7.1f}%  {r['n']:4d}")
    print()
    for side in ("sparse", "dense"):
        cc = res[side]["constructs_per_component"]
        print(f"  {side:6s}  constructs per component: mean {cc['mean']:.2f}, median {cc['median']:.0f}, "
              f"single-construct {cc['single_construct']}/{cc['n_components']}   "
              f"core components per item {res[side]['mean_core_components_per_item']:.2f}")
    print(f"\nwrote {out}")


if __name__ == "__main__":
    main()
```
