## Supplemental code for "Scoring respondents on a semantic frame of reference identifies a group that is burdened across many questionnaires and reaches care less easily": fig1_panels.py

```
#!/usr/bin/env python
"""Figure 1, emitted as THREE INDEPENDENT EDITABLE PANELS instead of one composed image.

Why this exists. The composed `fig1_pool.py` fixes the arrangement in matplotlib, which makes hand
editing impossible: the only way to move a label was to change code and re-render. This writes each
panel as its own SVG with the text left as TEXT (`svg.fonttype = "none"`, so labels are editable
strings and not outlines). They open in Inkscape, Illustrator or Affinity, and PowerPoint imports
SVG and will convert it to editable shapes.

  panels/fig1a_overview.svg   11.62 x 2.35 in   what a reference basis is
  panels/fig1b_apply.svg      11.62 x 3.30 in   what it is for: place, score, flag, attribute
  panels/fig1c_cards.svg      11.62 x 5.20 in   the 32 cards with cores on <= 3 constructs
  panels/fig1d_ribbon.svg     10.50 x 6.60 in   instrument -> construct -> component flow
  panels/figS_cards_all.svg    7.20 x 9.60 in   all 55 cards (Supplementary Figure S17)

SCALE, which is the thing to keep in mind while editing. Each panel is placed in the manuscript at
`width=\\textwidth`, i.e. 6.5 in, so a panel drawn 11.62 in wide is reduced by 0.559 and its 11.5 pt
label prints at 6.4 pt. NHB's figure guidance sets a 5 pt floor, so on these three panels do not take
any type below:

  fig1a / fig1b   9.0 pt drawn  ->  5.0 pt printed
  fig1c          8.1 pt drawn  ->  5.0 pt printed   (placed at 0.619)
  figS_cards_all  5.6 pt drawn  ->  5.0 pt printed   (placed at 0.903)

The composed PNG is still built by `fig1_pool.py` and is what the manuscript currently includes.
Once the edited panels come back they are arranged in TeX instead, and this file's SVGs become the
source of record for the arrangement.

Run:  /data_gpu5/ROSS_Proj/env/bin/python fig1_panels.py
"""
import os
import matplotlib
matplotlib.use("Agg")
import matplotlib.pyplot as plt

# importing the composed generator gives us its drawing functions and its loaded pool data, so the
# panels and the composite cannot drift apart. It re-renders the composite as a side effect.
import fig1_pool as F

OUT = os.path.join(F.OUT, "panels")
os.makedirs(OUT, exist_ok=True)
SVG = {"svg.fonttype": "none"}          # keep labels as editable text, not outlines


def save(fig, name):
    with matplotlib.rc_context(SVG):
        p = os.path.join(OUT, name)
        fig.savefig(p, bbox_inches="tight", facecolor="white")
    plt.close(fig)
    print("wrote", p)


# ---- panel a: the schematic, at the size it has inside the composed figure ----
# the axes patch is hidden on both schematic panels: it is opaque white, and bbox_inches="tight"
# would otherwise crop to the axes rather than to the artwork, leaving a wide empty margin
figA = plt.figure(figsize=(11.62, 2.35), facecolor="white")
axA = figA.add_axes([0, 0, 1, 1]); axA.patch.set_visible(False)
F.draw_basis(axA)
save(figA, "fig1a_overview.svg")

# ---- panel b: what the fixed basis is for -- a battery placed, its respondents scored, flagged,
#      and the flag returned to the components. Its own panel because (a) answers "what is the
#      basis" and (b) answers "what is it for", and a reader can take them one at a time.
figAp = plt.figure(figsize=(11.62, 3.30), facecolor="white")
axAp = figAp.add_axes([0, 0, 1, 1]); axAp.patch.set_visible(False)
F.draw_apply(axAp)
save(figAp, "fig1b_apply.svg")

# ---- panel b: the 32 cards whose cores rest on at most three constructs ----
figB = plt.figure(figsize=(11.62, 5.20), facecolor="white")
axB = figB.add_axes([0, 0.02, 1, 0.935])
# top_pad is the dead band the legend sits in, in row units. The default 0.75 leaves a visible gap
# between the legend and the first row of cards at this panel size. The legend sits ABOVE the axes
# (bbox_to_anchor y=1.005), so this band is pure white space and 0.10 is enough to keep them apart.
F.draw_cards(axB, F.MAIN_CARDS, F.MAIN_NCOL, fs_hdr=13.0, fs_con=11.5, fs_leg=12.5,
             nc_levels=[1, 2, 3], top_pad=0.10)
save(figB, "fig1c_cards.svg")

# ---- supplementary: all 55 cards, drawn near print size already ----
figS = plt.figure(figsize=(7.2, 9.6), facecolor="white")
axS = figS.add_axes([0, 0.01, 1, 0.94])
F.draw_cards(axS, F.cards, 5, fs_hdr=6.2, fs_con=5.6, fs_leg=6.4, nc_levels=[1, 2, 3, 4, 5],
             first_long=False)
save(figS, "figS_cards_all.svg")

# ---- panel d: written as vector by its own generator, copied in beside the others ----
import shutil
src = os.path.join(F.SRC, "fig_pool_ribbon.svg")
if os.path.exists(src):
    shutil.copy(src, os.path.join(OUT, "fig1d_ribbon.svg"))
    print("wrote", os.path.join(OUT, "fig1d_ribbon.svg"))
else:
    print("panel c: run fig_pool_ribbon.py first to write fig_pool_ribbon.svg")
```
