## Supplemental code for "Scoring respondents on a semantic frame of reference identifies a group that is burdened across many questionnaires and reaches care less easily": fig1_pool.py

```
#!/usr/bin/env python
"""MAIN Figure 1 (NHB, global pool): three stacked full-width panels.

  (a) what a reference basis is -- published question text, embedded, projected onto a small set of
      fixed COMPONENTS drawn as orthogonal directions from one origin, each dominated by a few core
      items that sit at the far end of it while the items it does not rest on sit near the origin.
      Schematic, but the three featured item texts are read from the pool.
  (b) the 32 component cards whose cores rest on at most three constructs, six columns in
      component-index order. Each card: component id + core size (header), then up to three
      constructs with in-core counts, all INSIDE the card. Background coloured by the NUMBER of
      distinct constructs in the core, with a legend. All 55 go to figS_pool_cards_all.png
      (Supplementary Figure S17) and, as data, to Supplementary Data 4.
  (c) lean pool ribbon (fig_pool_ribbon.png, 5 merge families) -- imported image, white margin cropped

  Panel order follows the order the main text cites them: the overview says what the basis is, the
  cards carry the component count and the single-construct count, the ribbon then shows instruments
  merging onto a shared component.

LAYOUT RULE (2026-08-28). Three stacked FULL-WIDTH rows, never b and c side by side. Both lower
panels are limited by how much page width their content gets -- the cards by the width of one card,
the ribbon by the distance between its three label columns -- and sharing a row left each about
2.1 in on the page, where neither can carry the 5 pt type NHB's figure guidance sets as the floor.
Panel c's height is pinned to the ribbon image's own aspect, because imshow letterboxes otherwise
and the ribbon would then use only part of the width its type size depends on. Panel b shows 32 of
the 55 cards for the same reason: 55 cards in half the figure width forced 5.9 pt labels, which
land near 2.6 pt on the page.

The EDITABLE per-panel SVGs are written by fig1_panels.py, which imports the drawing functions from
here so the two cannot drift apart.

Run (regenerate the ribbon panel first):
  /data_gpu5/ROSS_Proj/env/bin/python fig_pool_ribbon.py
  /data_gpu5/ROSS_Proj/env/bin/python fig1_pool.py     # -> manuscript_nhb/figures/fig1_pool.png
"""
import os
import textwrap
from collections import Counter
import numpy as np, pandas as pd
import matplotlib; matplotlib.use("Agg")
import matplotlib.pyplot as plt
import matplotlib.image as mpimg
from matplotlib.patches import FancyBboxPatch, Patch, Rectangle

HERE = os.path.dirname(os.path.abspath(__file__))
REPO = os.path.abspath(os.path.join(HERE, "..", ".."))
SRC = os.path.join(REPO, "examples", "global_pool", "figures")
OUT = os.path.join(REPO, "manuscript_nhb", "figures"); os.makedirs(OUT, exist_ok=True)
ENC = "gte-large-en-v1.5"
CORES = os.path.join(HERE, "outputs_pool", "fixedpoint_cores", f"{ENC}_cores.csv")
POOL = os.path.join(HERE, "..", "global_pool", "pool_processed.csv")

from pool_card_labels import short          # shared with fig_xenc.py

# colour by number of core constructs on a single warm gradient: FEWER = brighter (pure = lightest).
# Single-hue lightness ramp is colourblind-safe. Pure (1) additionally gets a thick black border.
NCCOL = {1: "#FFF3E0", 2: "#FDCB8E", 3: "#F79A45", 4: "#DE6E12", 5: "#9C4A02"}
nccol = lambda n: NCCOL.get(n if n < 5 else 5)
ncedge = {1: "#000000", 2: "#C9973F", 3: "#B96C1E", 4: "#8F4708", 5: "#5E2C01"}
ncedg = lambda n: ncedge.get(n if n < 5 else 5)
# the ramp spans L* 96 to 41, so the card label ink has to follow it: dark text on the light cards,
# white on the dark ones, otherwise the busiest cards are unreadable
NCINK = {1: ("0.10", "0.12", "0.45"), 2: ("0.10", "0.12", "0.45"), 3: ("0.10", "0.12", "0.38"),
         4: ("white", "white", "#F3E2D2"), 5: ("white", "white", "#EBD8C6")}
ncink = lambda n: NCINK.get(n if n < 5 else 5)

pp = pd.read_csv(POOL); CON = dict(zip(pp.item_id, pp.construct))
cards = []
for _, r in pd.read_csv(CORES).iterrows():
    items = str(r["items"]).split()
    comp = Counter(CON.get(i, "?") for i in items).most_common()
    cards.append(dict(idx=int(r["component"]), name=f"C{r['component']}", k=len(items),
                      comp=comp, nc=len(comp)))
cards.sort(key=lambda c: c["idx"])                         # component-index order
NCOL, NROW = 5, 11

# ---- ribbon panel: crop white margin ----
rib = mpimg.imread(os.path.join(SRC, "fig_pool_ribbon.png"))
m = (rib[:, :, :3] < 0.97).any(axis=2)
ys, xs = np.where(m.any(axis=1))[0], np.where(m.any(axis=0))[0]
rib = rib[ys.min():ys.max() + 1, xs.min():xs.max() + 1]

# ---- overview panel: schematic, but the featured item texts come from the pool ----
# Presentation choices only: which three items to show, and that two of them share a construct.
FEATURED = ["CESD_6", "PROMIS_DEP_4", "MSPSS_6"]
# the pool's provenance tag for the PROMIS banks is the collection, not the bank, so shorten it to
# the name the ribbon panel uses for the same items
SRCNAME = {i: ("PROMIS" if s == "PROMIS-CDE" else s) for i, s in zip(pp.item_id, pp.source)}
TEXT = dict(zip(pp.item_id, pp.prompt_clean))
INK, RULE = "#222222", "#8A8A8A"
STAGE = "#08306B"                                       # stage headings, L* 21
MARK = {"Depression": "#08306B", "Social support": "#D55E00"}   # the two featured constructs


def arrow_l(ax, x0, x1, y):
    """A right-to-left arrow. The scoring row runs backwards along the panel, continuing the
    direction of the placement arrow above it, so it needs its own helper rather than arrow()."""
    ax.annotate("", xy=(x1, y), xytext=(x0, y),
                arrowprops=dict(arrowstyle="-|>", lw=1.4, color=RULE, mutation_scale=16), zorder=2)


def chip(ax, x, y, w, h, label, fc="white", ec=RULE, lw=0.9, fs=8.5, color=INK, style="normal"):
    ax.add_patch(FancyBboxPatch((x, y), w, h, boxstyle="round,pad=0.004,rounding_size=0.012",
                                fc=fc, ec=ec, lw=lw, transform=ax.transData, zorder=3))
    ax.text(x + w / 2, y + h / 2, label, ha="center", va="center", fontsize=fs, color=color,
            style=style, zorder=4)


def arrow(ax, x0, x1, y, label):
    ax.annotate("", xy=(x1, y), xytext=(x0, y),
                arrowprops=dict(arrowstyle="-|>", lw=1.4, color=RULE, mutation_scale=16), zorder=2)
    ax.text((x0 + x1) / 2, y + 0.038, label, ha="center", va="bottom", fontsize=12.5,
            style="italic", color=INK)


def draw_basis(ax):
    ax.set_xlim(0, 1); ax.set_ylim(0, 1); ax.axis("off")

    # 1. published question text, three real items from three instruments
    ax.text(0.092, 0.90, "Published question text", ha="center", fontsize=13.5,
            fontweight="bold", color=STAGE)
    for i, iid in enumerate(FEATURED):
        y = 0.70 - 0.200 * i
        con = CON[iid]
        # wrapped, because at print size the chip text has to carry a readable point size and the
        # longest featured item does not fit the chip width on one line
        label = "\n".join(textwrap.wrap(f"“{str(TEXT[iid]).rstrip('.')}”", 20))
        chip(ax, 0.005, y, 0.174, 0.150, label, fs=11.0)
        ax.text(0.092, y - 0.033, SRCNAME[iid], ha="center", va="center", fontsize=11.0,
                color=MARK.get(con, INK), fontweight="bold")
    ax.text(0.092, 0.200, "577 items · 50 instruments\n54 documented constructs",
            ha="center", va="top", fontsize=11.5, color=INK, linespacing=1.35)

    arrow(ax, 0.195, 0.262, 0.50, "semantic\nencoder")

    # 2. the embedding: featured items sit near items of their own construct
    ax.text(0.340, 0.90, "Semantic space", ha="center", fontsize=13.5, fontweight="bold", color=STAGE)
    rng = np.random.RandomState(3)
    ax.scatter(0.278 + 0.125 * rng.rand(70), 0.26 + 0.54 * rng.rand(70), s=13, color="#C8C8C8",
               zorder=2)
    for iid, (dx, dy) in zip(FEATURED, [(0.045, 0.655), (0.060, 0.600), (0.098, 0.360)]):
        ax.scatter(0.278 + dx, dy, s=70, color=MARK.get(CON[iid], INK), zorder=4,
                   edgecolor="white", linewidth=0.9)
    ax.text(0.340, 0.200, "each item is a point,\nplaced by what it asks", ha="center", va="top",
            fontsize=11.5, color=INK, linespacing=1.35)

    arrow(ax, 0.432, 0.500, 0.50, "sparse projection")

    # 3. the basis: fixed components drawn as a set of mutually orthogonal directions from a common
    #    origin. Position along a direction is the item's score on that component, so an item that
    #    dominates the component sits at the far end and the items it does not sit near the origin.
    ax.text(0.632, 0.90, "Reference basis:\ninterpretable components", ha="center", fontsize=13.5,
            fontweight="bold", color=STAGE)
    # the panel is far wider than it is tall, so a direction drawn in data units would be sheared.
    # Convert an inch-length and a true angle into data units to keep the right angles right.
    bb = ax.get_position(); FW, FH = ax.figure.get_size_inches()
    W_in, H_in = FW * bb.width, FH * bb.height

    def ray(x0, y0, deg, L):
        t = np.deg2rad(deg)
        return x0 + L * np.cos(t) / W_in, y0 + L * np.sin(t) / H_in

    # the basis is one object that is built once and then not touched, so it is drawn as one
    # object: everything a battery is placed onto sits inside this box
    ax.add_patch(FancyBboxPatch((0.487, 0.020), 0.318, 0.835,
                               boxstyle="round,pad=0.004,rounding_size=0.012",
                               fc="#FAFAFA", ec=RULE, lw=1.0, ls="-", zorder=0))
    OX, OY, LEN = 0.632, 0.400, 0.72
    # three of the 55, drawn at 120 degrees apart: mutually orthogonal directions seen in projection
    COMPS = [("Depression", 90, "center", "bottom", 0.10),
             ("Anxiety", 210, "right", "top", 0.14),
             ("Social support", 330, "left", "top", 0.14)]
    for name, deg, ha, va, pad in COMPS:
        xt, yt = ray(OX, OY, deg, LEN)
        ax.annotate("", xy=(xt, yt), xytext=(OX, OY),
                    arrowprops=dict(arrowstyle="-|>", lw=1.2, color=RULE, mutation_scale=15),
                    zorder=2)
        # items the component does not rest on: small scores, so near the origin
        for f in (0.10, 0.17, 0.24, 0.31, 0.38, 0.45):
            x, y = ray(OX, OY, deg, LEN * f)
            ax.scatter(x, y, s=15, color="#C8C8C8", zorder=3)
        # core items: the few that dominate the component, so out at the extreme
        for f in (0.62, 0.74, 0.86):
            x, y = ray(OX, OY, deg, LEN * f)
            ax.scatter(x, y, s=66, color=MARK.get(name, "#6E6E6E"), zorder=4,
                       edgecolor="white", linewidth=0.8)
        lx, ly = ray(OX, OY, deg, LEN + pad)
        ax.text(lx, ly, name, ha=ha, va=va, fontsize=12.5, color=MARK.get(name, INK),
                fontweight="bold")
    ax.scatter([OX], [OY], s=22, color=INK, zorder=5)
    ax.annotate("its core items",
                xy=ray(OX, OY, 90, LEN * 0.74), xytext=(0.726, 0.660),
                fontsize=11.5, color=INK, ha="left", va="center", linespacing=1.35,
                arrowprops=dict(arrowstyle="-", lw=0.9, color=RULE,
                                connectionstyle="angle3,angleA=0,angleB=70"))
    ax.text(0.632, 0.100, "built once, then held fixed", ha="center", va="top", fontsize=11.0,
            fontweight="bold", color=INK)


def draw_apply(ax):
    """Panel b: what the fixed basis is for.

    Two inputs meet at the circled plus, which is the step the linear version could not show: the
    BASIS arrives from above and the ANSWERS from the left, and only the two together give a
    respondent a position. Everything downstream of that node is per person -- a position, a
    distance, and a flag for the far tail.

    The dashed arrow returns the flag TO THE COMPONENTS, which is the layer the attribution is
    reported at (Methods, attributing an outlier). It is deliberately not returned to individual
    items: the distance does partition over items, but that partition is used only for the
    item-level test in the supplement. No method is named anywhere in the panel.
    """
    ax.set_xlim(0, 1); ax.set_ylim(0, 1); ax.axis("off")
    FLAG = "#D55E00"
    rng = np.random.RandomState(11)
    TOP, LOW = 0.775, 0.275            # the two rows: placement above, respondents below
    BX, PLUS = 0.318, 0.318            # basis column, and the node beneath it

    # ---- upper row: a battery is placed on the basis, and the basis does not change ----
    ax.text(0.088, TOP + 0.185, "A new battery", ha="center", va="bottom", fontsize=12.5,
            fontweight="bold", color=STAGE)
    chip(ax, 0.020, TOP - 0.072, 0.136, 0.148, "All of Us, 151 items", fs=11.0, fc="#F4F4F4")
    ax.text(0.088, TOP - 0.098, "the basis was not built from it", ha="center", va="top",
            fontsize=10.5, style="italic", color=INK)

    arrow(ax, 0.162, 0.216, TOP, "placed")

    ax.add_patch(FancyBboxPatch((0.228, TOP - 0.215), 0.180, 0.430,
                                boxstyle="round,pad=0.004,rounding_size=0.012",
                                fc="#FAFAFA", ec=RULE, lw=1.0, zorder=0))
    ax.text(BX, TOP + 0.245, "Reference basis", ha="center", va="bottom", fontsize=12.5,
            fontweight="bold", color=STAGE)
    bb = ax.get_position(); FW, FH = ax.figure.get_size_inches()
    W_in, H_in = FW * bb.width, FH * bb.height
    for deg, col in ((90, "#08306B"), (210, "#6E6E6E"), (330, "#D55E00")):
        tr = np.deg2rad(deg)
        xt, yt = BX + 0.40 * np.cos(tr) / W_in, TOP + 0.40 * np.sin(tr) / H_in
        ax.annotate("", xy=(xt, yt), xytext=(BX, TOP),
                    arrowprops=dict(arrowstyle="-|>", lw=1.1, color=RULE, mutation_scale=13),
                    zorder=2)
        for f in (0.66, 0.84):
            ax.scatter(BX + 0.40 * f * np.cos(tr) / W_in, TOP + 0.40 * f * np.sin(tr) / H_in,
                       s=30, color=col, zorder=4, edgecolor="white", linewidth=0.6)
    # ---- the node: the basis comes down, the answers come in from the left ----
    ax.annotate("", xy=(PLUS, LOW + 0.075), xytext=(BX, TOP - 0.220),
                arrowprops=dict(arrowstyle="-|>", lw=1.4, color=RULE, mutation_scale=16), zorder=2)
    ax.scatter([PLUS], [LOW], s=430, facecolor="white", edgecolor=STAGE, linewidth=1.5, zorder=5)
    ax.plot([PLUS - 0.017, PLUS + 0.017], [LOW, LOW], color=STAGE, lw=1.5, zorder=6)
    ax.plot([PLUS, PLUS], [LOW - 0.048, LOW + 0.048], color=STAGE, lw=1.5, zorder=6)

    ax.text(0.088, LOW + 0.152, "Responses to the\nAll of Us battery", ha="center", va="bottom",
            fontsize=12.5, fontweight="bold", color=STAGE, linespacing=1.25)
    for r in range(5):
        for c in range(9):
            ax.add_patch(Rectangle((0.026 + 0.0140 * c, LOW + 0.115 - 0.052 * r), 0.0124, 0.044,
                                   fc=plt.cm.Blues(0.20 + 0.55 * rng.rand()), ec="white", lw=0.4,
                                   zorder=3))
    ax.text(0.088, LOW - 0.155, "every respondent \u00d7 every question", ha="center", va="top",
            fontsize=10.5, style="italic", color=INK)
    ax.annotate("", xy=(PLUS - 0.026, LOW), xytext=(0.166, LOW),
                arrowprops=dict(arrowstyle="-|>", lw=1.4, color=RULE, mutation_scale=16), zorder=2)

    # ---- downstream of the node, everything is per person ----
    ax.annotate("", xy=(0.470, LOW), xytext=(PLUS + 0.030, LOW),
                arrowprops=dict(arrowstyle="-|>", lw=1.4, color=RULE, mutation_scale=16), zorder=2)
    ax.text(0.585, LOW + 0.185, "Each person is a point", ha="center", va="bottom", fontsize=12.5,
            fontweight="bold", color=STAGE)
    pts = rng.randn(60, 2) * np.array([0.028, 0.055])
    ax.scatter(0.585 + pts[:, 0], LOW + pts[:, 1], s=10, color="#C8C8C8", zorder=3)

    arrow_l_none = None
    ax.annotate("", xy=(0.752, LOW), xytext=(0.700, LOW),
                arrowprops=dict(arrowstyle="-|>", lw=1.4, color=RULE, mutation_scale=16), zorder=2)
    ax.text(0.868, LOW + 0.185, "One distance each", ha="center", va="bottom", fontsize=12.5,
            fontweight="bold", color=STAGE)
    H = [0.10, 0.32, 0.66, 0.90, 1.00, 0.88, 0.66, 0.46, 0.30, 0.19, 0.12, 0.08, 0.06, 0.05]
    for i, h in enumerate(H):
        ax.add_patch(Rectangle((0.790 + 0.0114 * i, LOW - 0.105), 0.0100, 0.30 * h,
                               fc=FLAG if i >= len(H) - 3 else "#BFBFBF", ec="none", zorder=3))
    ax.text(0.868, LOW - 0.155, "the most distant five percent are flagged", ha="center", va="top",
            fontsize=10.5, style="italic", color=FLAG)

    # ---- and the flag comes apart into the components, not into single items ----
    # the return runs UP the right edge and then LEFT into the basis, mirroring the forward path
    # (basis down, answers right): the two together enclose the respondent stages between them.
    ax.annotate("", xy=(0.965, 0.630), xytext=(0.965, 0.300),
                arrowprops=dict(arrowstyle="-", lw=1.5, color=FLAG,
                                linestyle=(0, (5, 2.6))), zorder=6)
    ax.annotate("", xy=(0.412, 0.630), xytext=(0.965, 0.630),
                arrowprops=dict(arrowstyle="-|>", lw=1.5, color=FLAG, mutation_scale=17,
                                linestyle=(0, (5, 2.6))), zorder=6)
    ax.text(0.800, 0.668, "every flag divides\namong the components", ha="center", va="bottom",
            fontsize=11.5, color=FLAG, fontweight="bold", linespacing=1.35)


def draw_cards(ax, subset, ncol, fs_hdr, fs_con, fs_leg, nc_levels, first_long=True, top_pad=0.75):
    """The card grid. Card text size is set by the card's own width, not chosen freely: the longest
    short() label is 14 characters, so a label must fit the ~0.78-of-a-cell text column.

    top_pad is the band above the first row, in row units, that the legend sits in. It is dead space
    inside the axes, so bbox_inches="tight" cannot trim it, and on a figure with few rows it is a
    visible strip of white."""
    nrow = int(np.ceil(len(subset) / ncol))
    ax.set_xlim(0, ncol); ax.set_ylim(-top_pad, nrow); ax.axis("off"); ax.invert_yaxis()
    for pos, c in enumerate(subset):
        col, row = pos % ncol, pos // ncol
        lw = 2.0 if c["nc"] == 1 else 0.9
        ax.add_patch(FancyBboxPatch((col + 0.04, row + 0.07), 0.92, 0.86,
                     boxstyle="round,pad=0.006,rounding_size=0.04", ec=ncedg(c["nc"]),
                     fc=nccol(c["nc"]), lw=lw))
        hdr = (f"{c['name']} ({c['k']} items in core)" if (pos == 0 and first_long)
               else f"{c['name']} ({c['k']})")
        hdr_c, con_c, num_c = ncink(c["nc"])
        ax.text(col + 0.08, row + 0.235, hdr,
                fontsize=(fs_hdr * 0.78 if (pos == 0 and first_long) else fs_hdr),
                fontweight="bold", va="center", color=hdr_c)
        # at most three text lines, all INSIDE the card: a fourth line lands on the next row's card
        shown = c["comp"][:3] if c["nc"] <= 3 else c["comp"][:2]
        yy = row + 0.45
        for con, n in shown:
            ax.text(col + 0.08, yy, short(con), fontsize=fs_con, va="center", color=con_c)
            ax.text(col + 0.93, yy, f"{n}", fontsize=fs_con, va="center", ha="right", color=num_c)
            yy += 0.205
        if c["nc"] > 3:
            ax.text(col + 0.08, yy, f"+{c['nc'] - 2} more constructs", fontsize=fs_con * 0.92,
                    va="center", color=con_c, style="italic")
    leg = [Patch(fc=nccol(n), ec=ncedg(n), lw=(1.8 if n == 1 else 0.9),
                 label=f"{n if n < 5 else '5+'} construct" + ("s" if n != 1 else ""))
           for n in nc_levels]
    ax.legend(handles=leg, loc="lower center", bbox_to_anchor=(0.5, 1.005), ncol=len(nc_levels),
              frameon=False, fontsize=fs_leg, handlelength=1.3, handleheight=1.3,
              columnspacing=1.3, title="core constructs", title_fontsize=fs_leg * 1.05)


# ---------------------------------------------------------------- main figure
# Panel b carries only the components whose cores rest on at most three constructs. All 55 are in
# the supplementary card figure and, as data, in Supplementary Data 4. The cut is what makes the
# card type legible at \textwidth: 55 cards in the narrow half of the figure forced 5.9 pt labels,
# which land near 2.6 pt on the page, under the 5 pt floor NHB's figure guidance sets.
MAIN_CARDS = [c for c in cards if c["nc"] <= 3]
MAIN_NCOL = 6

# Three stacked full-width rows rather than b and c side by side. Both lower panels are limited by
# how much page width their content gets: the cards by the width of one card, the ribbon by the
# distance between its three label columns. Sharing a row left each with about 2.1 in on the page,
# where neither can carry 5 pt type.
# Panel c's height is set by the ribbon image's own aspect: imshow preserves it, so an axes wider
# than the image would letterbox and the ribbon would use only part of the page width, which is the
# one thing its type size depends on.
RIB_ASPECT = rib.shape[1] / rib.shape[0]
PANEL_W = 11.62
H_A, H_B = 2.35, 5.20
H_C = PANEL_W / RIB_ASPECT
fig = plt.figure(figsize=(15.0, (H_A + H_B + H_C) * 1.055), facecolor="white")
gs = fig.add_gridspec(3, 1, height_ratios=[H_A, H_B, H_C], hspace=0.10)

axOver = fig.add_subplot(gs[0])
axOver.text(-0.012, 1.03, "a", transform=axOver.transAxes, fontsize=18, fontweight="bold",
            va="top", ha="left")
draw_basis(axOver)

axCards = fig.add_subplot(gs[1])
axCards.text(-0.012, 1.0, "b", transform=axCards.transAxes, fontsize=18, fontweight="bold",
             va="top", ha="left")
draw_cards(axCards, MAIN_CARDS, MAIN_NCOL, fs_hdr=13.0, fs_con=11.5, fs_leg=12.5,
           nc_levels=[1, 2, 3])

axRibbon = fig.add_subplot(gs[2]); axRibbon.imshow(rib); axRibbon.axis("off")
axRibbon.text(-0.012, 1.0, "c", transform=axRibbon.transAxes, fontsize=18, fontweight="bold",
              va="top", ha="left")

out = os.path.join(OUT, "fig1_pool.png")
fig.savefig(out, dpi=230, bbox_inches="tight", facecolor="white")
print("wrote", out, f"| {len(MAIN_CARDS)} of {len(cards)} cards | ribbon crop {rib.shape[1]}x{rib.shape[0]}")

# ------------------------------------------------- supplementary: all 55 cards
# Built near print size (7.2 in against a 6.5 in \textwidth), so the point sizes here are close to
# what the page shows and no scaling correction is needed.
# SIX columns, not five. At five the grid needed 11 rows and the figure was 9.6 in tall, which at
# \textwidth prints 8.97 in against a 9 in text height, leaving nowhere for the caption: the page
# ran 78 pt over. Six columns take 10 rows, and the height comes down with them. The width stays
# 7.2 in, because that is what holds the card type at 5.1 pt on the page, above NHB's 5 pt floor --
# scaling the five-column version down to fit would have put it at 4.6 pt instead. The narrower
# card still holds the longest short() label: 14 characters at 5.6 pt is 0.65 in against the
# 0.77-of-a-cell text column, which is 0.92 in at six columns.
figS = plt.figure(figsize=(7.2, 7.7), facecolor="white")
axS = figS.add_subplot(111)
draw_cards(axS, cards, 6, fs_hdr=6.2, fs_con=5.6, fs_leg=6.4, nc_levels=[1, 2, 3, 4, 5],
           first_long=False, top_pad=0.32)
outS = os.path.join(OUT, "figS_pool_cards_all.png")
figS.savefig(outS, dpi=400, bbox_inches="tight", facecolor="white")
print("wrote", outS, f"| all {len(cards)} cards")
```
