## Supplemental code for "Scoring respondents on a semantic frame of reference identifies a group that is burdened across many questionnaires and reaches care less easily": fig_aou_twoside_cores.py

```
#!/usr/bin/env python
"""AoU two-sided core-item ribbon: do the matching instruments core the same pool column?

Same construction as fig_hrs_twoside_cores.py, under the settled definitions (DEFINITIONS.md D1-D7).
Both sides are CORE ITEMS of the same fixed column, nothing else:
  LEFT   pool instruments whose items form E_i^G
  RIGHT  All of Us instruments whose items form E_i^S
Only columns All of Us reaches are eligible (A_i^S non-empty, D6 as amended 2026-08-19): an item
leans on a column when it clears that column's pool core-item band or when the column is the one the
item stands out on most. Both conditions are per item against quantities fixed on the pool, so
relevance stays battery-independent.

FEATURED columns, chosen to answer two questions at once:
  * pure external transfer   C43 (ACE-BRFSS), C27 (BFI-2-XS)
  * external sharing a core with an All of Us instrument that IS in the pool
                             C39 (ACE-BRFSS + MHQ), C44 (MOS-SS + MHQ),
                             C8  (MOS-SS + UCLA-V3), C51 (BFI-2-XS + others)
  * same-instrument anchors, for contrast
                             C49 (DMS both sides), C25 (GAD-7 both sides)
The three All of Us instruments absent from the pool are drawn in red on the right axis.

Out: examples/global_pool/figures/fig_aou_twoside_cores.png
Run: CUDA_VISIBLE_DEVICES="" /data_gpu5/ROSS_Proj/env/bin/python fig_aou_twoside_cores.py
"""
import os, glob, csv, re
import numpy as np, pandas as pd
import matplotlib; matplotlib.use("Agg")
import matplotlib.pyplot as plt
from matplotlib.path import Path
from matplotlib.patches import PathPatch, Rectangle, Patch

from _common import keep_columns, pool_bands
from pool_card_labels import short   # the margin labels must fit the print-size margin

HERE = os.path.dirname(os.path.abspath(__file__))
POOL_IDS = pd.read_csv(os.path.join(HERE, "..", "global_pool", "pool_processed.csv"))["item_id"].tolist()
REPO = os.path.abspath(os.path.join(HERE, "..", ".."))
OUT = os.path.join(REPO, "examples", "global_pool", "figures"); os.makedirs(OUT, exist_ok=True)
ENC = "gte-large-en-v1.5"
NOT_IN_POOL = {"ACE-BRFSS", "MOS-SS", "BFI-2-XS"}

# Which components to feature is the one presentation choice here. Everything a component is said to
# be about is computed: a component is read by the SET of documented constructs of its core items, its
# construct profile (DEFINITIONS.md D6-A). No construct is designated for a component and constructs
# are never grouped into families.
FEATURED = [43, 39, 44, 8, 51, 27, 49, 25]
NPROF = 2                    # constructs listed per profile before "+N more"


def profile(items, con):
    """The construct profile of a set of core items: documented constructs with their item counts.

    Ties are broken alphabetically so the figure is byte-reproducible -- `value_counts` alone
    breaks them in hash-table order, which moves with Python's string-hash randomization.
    """
    return (pd.Series([con[i] for i in items]).value_counts()
            .sort_index().sort_values(ascending=False, kind="mergesort"))


def proftext(prof, n=NPROF):
    head = ", ".join(f"{k} ({v})" for k, v in list(prof.items())[:n])
    return head + (f", +{len(prof) - n} more" if len(prof) > n else "")
FCOL = {43: "#009E73", 39: "#D55E00", 44: "#0072B2", 8: "#56B4E9", 51: "#E69F00",
        27: "#F0E442", 49: "#CC79A7", 25: "#8C6BB1"}
OTHER, INK = "#C4CBD2", "#222222"


# Short display names, matched on the full scale name. Keying on the trailing parenthetical is not
# safe: several instruments share one, and "(6-item)" alone maps both the USDA food-security module
# and the De Jong Gierveld loneliness scale onto the same label.
POOLSHORT = {
    "Adverse Childhood Experiences Questionnaire": "Felitti-ACE",
    "De Jong Gierveld Loneliness Scale (6-item)": "De Jong Gierveld",
    "Discrimination in Medical Settings": "DMS",
    "Extended Hurt Insult Threaten Scream (E-HITS)": "E-HITS",
    "Generalized Anxiety Disorder-7": "GAD-7",
    "IPIP Big-Five Factor Markers (50-item)": "IPIP-50",
    "Interpersonal Support Evaluation List (12-item)": "ISEL-12",
    "Life Events Checklist for DSM-5 (LEC-5)": "LEC-5",
    "Major Experiences of Discrimination": "Major-Disc",
    "Mood Disorder Questionnaire (MDQ)": "MDQ",
    "Multidimensional Scale of Perceived Social Support (MSPSS)": "MSPSS",
    "PROMIS Item Bank v1.0 - Emotional Distress - Anxiety": "PROMIS-Anxiety",
    "PROMIS Item Bank v1.0 - Emotional Distress - Depression": "PROMIS-Depression",
    "PROMIS Item Bank v1.0 - Sleep Disturbance": "PROMIS-Sleep",
    "PROMIS Item Bank v2.0 - Emotional Support (SF 6a)": "PROMIS-EmSupport",
    "PROMIS Item Bank v2.0 - Social Isolation": "PROMIS-Isolation",
    "Patient Health Questionnaire-15 (PHQ-15)": "PHQ-15",
    "Perceived Neighborhood Disorder (AoU adapted)": "NDS",
    "Primary Care PTSD Screen for DSM-5 (PC-PTSD-5)": "PC-PTSD-5",
    "Prodromal Questionnaire-Brief (PQ-B)": "PQ-B",
    "Three-Item UCLA Loneliness Scale": "UCLA-3",
    "UK Biobank Mental Health Questionnaire": "UKB-MHQ",
    "USDA Household Food Security Survey Module (6-item Short Form)": "USDA-FS",
    "WHOQOL-BREF": "WHOQOL-BREF",
    "Center for Epidemiologic Studies Depression Scale": "CES-D",
    "Everyday Discrimination Scale": "EDS",
    "Heightened Vigilance Scale": "HVS",
    "Hunger Vital Sign": "Hunger Vital Sign",
    "Neighborhood Social Cohesion": "Nbhd. cohesion",
    "PROMIS Global Health": "PROMIS-Global",
    "PROMIS Item Bank v1.0 - Sleep Related Impairment": "PROMIS-SleepImpair",
    "PROMIS Item Bank v1.1 - Anger": "PROMIS-Anger",
    "Patient Health Questionnaire-9": "PHQ-9",
    "Rosenberg Self-Esteem Scale": "Rosenberg-SE",
    "WHO World Mental Health CIDI": "WMH-CIDI",
    "WHO-5 Well-Being Index": "WHO-5",
}


def poolshort(s):
    if s in POOLSHORT:
        return POOLSHORT[s]
    m = re.findall(r"\(([^)]+)\)", s)          # unmapped instruments keep their own abbreviation
    return m[-1] if m else s


def pr(v):
    s2 = float((v**2).sum()); s4 = float((v**4).sum()); return s2*s2/s4 if s4 > 0 else 0.0


def fpc(v):
    idx = np.arange(len(v))
    while True:
        n = max(1, min(int(round(pr(v[idx]))), len(idx)))
        top = np.sort(idx[np.argsort(-np.abs(v[idx]))[:n]])
        if len(top) == len(idx) and (top == idx).all():
            return top
        idx = top


def ribbon(ax, x0, ya0, ya1, x1, yb0, yb1, color, alpha, hatch=None):
    cx = (x0 + x1) / 2.0
    v = [(x0, ya0), (cx, ya0), (cx, yb0), (x1, yb0), (x1, yb1), (cx, yb1), (cx, ya1), (x0, ya1), (x0, ya0)]
    c = [Path.MOVETO, Path.CURVE4, Path.CURVE4, Path.CURVE4, Path.LINETO,
         Path.CURVE4, Path.CURVE4, Path.CURVE4, Path.CLOSEPOLY]
    ax.add_patch(PathPatch(Path(v, c), facecolor=color, edgecolor=("white" if hatch else "none"),
                           hatch=hatch, lw=0.0, alpha=alpha, zorder=1))


def main():
    fit = glob.glob(os.path.join(HERE, "outputs_pool", "fits", f"{ENC}__k*_ls1.0"))[0]
    W = np.load(os.path.join(fit, "loadings.npy")); mu = np.load(os.path.join(fit, "mean.npy"))
    Mg = np.load(os.path.join(HERE, "data", f"pool_items_{ENC}.npz"), allow_pickle=True)["vectors"].astype(float)
    sigma = ((Mg - mu) @ W.T).std(0)
    pp = pd.read_csv(os.path.join(HERE, "..", "global_pool", "pool_processed.csv"))
    pcon = dict(zip(pp.item_id, pp.construct)); pscale = dict(zip(pp.item_id, pp.scale))
    pcores = {int(r["component"]): str(r["items"]).split() for r in
              csv.DictReader(open(os.path.join(HERE, "outputs_pool", "fixedpoint_cores", f"{ENC}_cores.csv")))}
    d = np.load(os.path.join(HERE, "data", f"core151_items_{ENC}.npz"), allow_pickle=True)
    aid = [str(x) for x in d["items"]]
    Z = ((np.asarray(d["vectors"], float) - mu) @ W.T) / sigma
    cm = pd.read_csv(os.path.join(HERE, "construct_map_151.csv"))
    ains = dict(zip(cm.item, cm.instrument)); acon = dict(zip(cm.item, cm.construct))
    # an item leans on a component when it anchors it (clears the pool core-item band) or when the
    # component is that item's modal one -- the keep rule of _common.keep_columns (D6, amended)
    band = pool_bands(ENC, ((Mg - mu) @ W.T) / sigma, POOL_IDS)
    kept = set(keep_columns(Z, band))
    modal_of = [int(np.argmax(np.abs(Z[i]))) for i in range(len(aid))]

    cols, Lcon, Rcon = [], {}, {}
    for c in FEATURED:
        lean = {g for g in range(len(aid)) if modal_of[g] == c or abs(Z[g, c]) >= band[c]}
        assert c in kept and lean, f"C{c} is not kept for AoU under the settled rule"
        acore = [int(g) for g in fpc(Z[:, c])]
        cols.append(dict(c=c,
                         pprof=profile(pcores[c], pcon),
                         aprof=profile([aid[g] for g in acore], acon),
                         left=[(poolshort(pscale[i]), True) for i in pcores[c]],
                         right=[(ains[aid[g]], g in lean) for g in acore]))
        for i in pcores[c]:
            Lcon.setdefault(poolshort(pscale[i]), set()).add(pcon[i])
        for g in acore:
            Rcon.setdefault(ains[aid[g]], set()).add(acon[aid[g]])

    H = 100.0
    gap = 0.105 * H

    def names(key):
        out, seen = [], set()
        for x in cols:
            for t in x[key]:
                if t[0] not in seen:
                    seen.add(t[0]); out.append(t[0])
        return out

    def arrange(cols, rounds=8):
        """Barycentre ordering: place every node at the mean position of the nodes it connects to.

        Sweeping the three columns a few times settles into a layout with far fewer ribbon crossings
        than first-appearance order, and changes nothing about what is drawn.
        """
        Lord, Rord = names("left"), names("right")
        for _ in range(rounds):
            cpos = {x["c"]: i for i, x in enumerate(cols)}
            Lord = sorted(Lord, key=lambda n: np.mean([cpos[x["c"]] for x in cols
                                                       for t in x["left"] if t[0] == n]))
            Rord = sorted(Rord, key=lambda n: np.mean([cpos[x["c"]] for x in cols
                                                       for t in x["right"] if t[0] == n]))
            lp = {n: i / max(1, len(Lord) - 1) for i, n in enumerate(Lord)}
            rp = {n: i / max(1, len(Rord) - 1) for i, n in enumerate(Rord)}
            cols = sorted(cols, key=lambda x: np.mean([lp[t[0]] for t in x["left"]]
                                                      + [rp[t[0]] for t in x["right"]]))
        return cols, Lord, Rord

    cols, Lo, Ro = arrange(cols)

    def side(key, order):
        sz = {i: sum(1 for x in cols for t in x[key] if t[0] == i) for i in order}
        g2 = 0.042 * H; av = H - (len(order) - 1) * g2
        pos = {}; yy = H
        for i in order:
            h = av * sz[i] / sum(sz.values()); pos[i] = (yy - h, yy); yy -= h + g2
        return sz, pos

    Lsz, Lpos = side("left", Lo); Rsz, Rpos = side("right", Ro)

    tot = sum(max(len(x["left"]), len(x["right"])) for x in cols)
    avail = H - (len(cols) - 1) * gap
    Cpos = {}; y = H
    for x in cols:
        h = avail * max(len(x["left"]), len(x["right"])) / tot
        Cpos[x["c"]] = (y - h, y); y -= h + gap

    fig, ax = plt.subplots(# authored at print size: 7.2 in wide against a 6.5 in \textwidth, so a label drawn at 6.3 pt
    # prints at 5.7 pt. At the old 12.0 in it printed at 3.4 pt, under the 5 pt floor.
    figsize=(7.2, 7.2))
    xL, xC, xR, bw = 0.0, 1.35, 2.7, 0.032
    outL = {i: Lpos[i][1] for i in Lo}; outR = {i: Rpos[i][1] for i in Ro}
    inLC = {x["c"]: Cpos[x["c"]][1] for x in cols}; inRC = {x["c"]: Cpos[x["c"]][1] for x in cols}
    for x in cols:
        c = x["c"]; hC = Cpos[c][1] - Cpos[c][0]; col = FCOL[c]
        for ins, hit in sorted(x["left"], key=lambda t: (not t[1], t[0])):
            h = hC / len(x["left"]); hl = (Lpos[ins][1] - Lpos[ins][0]) / Lsz[ins]
            ribbon(ax, xL + bw, outL[ins], outL[ins] - hl, xC - bw, inLC[c], inLC[c] - h,
                   col if hit else OTHER, 0.60 if hit else 0.45)
            outL[ins] -= hl; inLC[c] -= h
        for ins, lean in sorted(x["right"], key=lambda t: (not t[1], ins not in NOT_IN_POOL, t[0])):
            h = hC / len(x["right"]); hr = (Rpos[ins][1] - Rpos[ins][0]) / Rsz[ins]
            ribbon(ax, xC + bw, inRC[c], inRC[c] - h, xR - bw, outR[ins], outR[ins] - hr,
                   col if lean else "#9AA3AB", 0.60 if lean else 0.28, None if lean else "////")
            outR[ins] -= hr; inRC[c] -= h

    # the documented construct of the items each instrument contributes here, in its own column beside
    # the instrument name rather than under it, since the thin nodes have no room for a second line
    def context(cons, n=1):
        c = sorted(short(x) for x in cons)
        return ", ".join(c[:n]) + (f", +{len(c) - n}" if len(c) > n else "")

    # LABEL PAIRING. An instrument and the construct its items are documented under are ONE label in
    # two lines. The gap inside the pair must be visibly smaller than the gap between pairs, or the
    # construct reads as belonging to the neighbouring instrument -- which it did at +-0.008/0.009H,
    # where the two gaps were the same size. The pair is now set tight around its node's midpoint.
    IN = 0.0015                       # half-gap inside a pair, in figure units
    for i in Lo:
        lo, hi = Lpos[i]; mid = (lo + hi) / 2
        ax.add_patch(Rectangle((xL - bw, lo), 2 * bw, hi - lo, fc="#6C86A0", ec="none", zorder=3))
        ax.text(xL - 2.2 * bw, mid + IN * H, i, ha="right", va="bottom", fontsize=6.9, color=INK)
        ax.text(xL - 2.2 * bw, mid - IN * H, context(Lcon[i]), ha="right", va="top",
                fontsize=6.9, color="#5A5A5A")
    for i in Ro:
        lo, hi = Rpos[i]; mid = (lo + hi) / 2; ext = i in NOT_IN_POOL
        ax.add_patch(Rectangle((xR - bw, lo), 2 * bw, hi - lo, fc="#C0392B" if ext else "#56657E",
                               ec="none", zorder=3))
        ax.text(xR + 2.2 * bw, mid + IN * H, i, ha="left", va="bottom", fontsize=6.9,
                color="#C0392B" if ext else INK, fontweight="bold" if ext else "normal")
        ax.text(xR + 2.2 * bw, mid - IN * H, context(Rcon[i]), ha="left", va="top",
                fontsize=6.9, color="#5A5A5A")
    for x in cols:
        lo, hi = Cpos[x["c"]]
        ax.add_patch(Rectangle((xC - bw, lo), 2 * bw, hi - lo, fc=FCOL[x["c"]], ec="none", zorder=3))
        ax.text(xC, hi + 0.030 * H, f"C{x['c']}", ha="center", va="bottom",
                fontsize=8.0, color=INK, fontweight="bold")
        ax.text(xC, hi + 0.017 * H, f"pool: {proftext(x['pprof'])}", ha="center", va="bottom",
                fontsize=6.3, color=INK)
        ax.text(xC, hi + 0.004 * H, f"All of Us: {proftext(x['aprof'])}", ha="center", va="bottom",
                fontsize=6.3, color=INK)

    ax.text(xL - 2.2 * bw, H + 0.085 * H, "pool instrument", ha="right",
            va="bottom", fontsize=7.6, fontweight="bold", color=INK, linespacing=1.3)
    ax.text(xC, H + 0.085 * H, "component, with the construct profile each side gives it",
            ha="center", va="bottom", fontsize=7.6, fontweight="bold", color=INK)
    ax.text(xR + 2.2 * bw, H + 0.085 * H, "All of Us instrument", ha="left",
            va="bottom", fontsize=7.6, fontweight="bold", color=INK, linespacing=1.3)
    leg = [Patch(fc="#9AA3AB", ec="white", hatch="////", alpha=0.60,
                 label="cores the component but does not lean on it"),
           Patch(fc="#C0392B", ec="none", label="All of Us instrument absent from the pool")]
    ax.legend(handles=leg, loc="lower center", bbox_to_anchor=(0.5, -0.055), ncol=2, frameon=False,
              fontsize=7.8, handlelength=1.4, handleheight=1.0, columnspacing=1.8)
    ax.set_xlim(-1.38, 4.42); ax.set_ylim(-0.02 * H, H + 0.19 * H); ax.axis("off")
    fig.subplots_adjust(left=0.005, right=0.995, top=0.985, bottom=0.09)
    out = os.path.join(OUT, "fig_aou_twoside_cores.png")
    fig.savefig(out, dpi=300, facecolor="white")
    print("wrote", out, f"| {len(cols)} featured components")


if __name__ == "__main__":
    main()
```
