## Supplemental code for "Scoring respondents on a semantic frame of reference identifies a group that is burdened across many questionnaires and reaches care less easily": fig_contentgrammar_3panel.py

```
#!/usr/bin/env python
"""§3.3.1 content-vs-syntax, THREE-panel cross-encoder figure (seed/donor notation, 151 seeds).

Notation (retires Cell A/B): every variant is a (seed, donor) pair.
  Syntax variant  = seed content + donor syntax   (content kept, syntax swapped)  -> near SEED
  Content variant = seed syntax  + donor content   (content swapped in)            -> near DONOR
The content-leakage split lives on the CONTENT variant: when the kept (seed) frame is itself
content-bearing, that frame leaks content and the content variant stays anchored to the seed -- the
"jingle" near-tie. (In this dataset every syntax-variant donor frame is neutral by construction, so
the syntax variant carries no leakage and is a single clean group.)

Groups (panels a, b):
  SYN   Syntax variant   (seed content + donor syntax)                    green
  CON_N Content variant  (seed syntax + donor content), neutral syntax    blue
  CON_L Content variant  (seed syntax + donor content), content-leaking   vermillion

Panels (box plots over all six encoders, ordered by clean content-vs-syntax gap):
  (a) cosine of each variant to the SEED
  (b) cosine of each variant to the DONOR   (mirror of a)
  (c) dumbbell -- per encoder, median cosine to the content source vs the syntax source, pooled over
      clean variants (excludes the content-leaking jingle case). The content-source point pools the
      syntax variants' cosine-to-seed (green in a) with the content variants' cosine-to-donor (blue in
      b); the syntax-source point pools the mirror pair. Neutral colours keep (c) distinct from a/b.

  env/bin/python examples/allofus/fig_contentgrammar_3panel.py \
     --long outputs_core151/content_grammar_v2/crossencoder_long.csv \
     --out  outputs_core151/content_grammar_v2/fig_contentgrammar_3panel.png
"""
import argparse
import csv
from collections import defaultdict

import numpy as np
import matplotlib
matplotlib.use("Agg")
import matplotlib.pyplot as plt
from matplotlib.patches import Patch

SYN, CON_N, CON_L = "SYN", "CON_N", "CON_L"
GROUPS = [SYN, CON_N, CON_L]
LABEL = {SYN:   "Syntax variant  (seed content + donor syntax)",
         CON_N: "Content variant  (seed syntax + donor content)",
         CON_L: "Content variant, content-leaking syntax (jingle)"}
# Colour-blind safe AND greyscale safe: the three sit at L* 58 / 21 / 77, so they separate on
# lightness as well as hue (the Okabe-Ito green and vermillion alone were 3.6 L* apart).
COL = {SYN: "#009E73", CON_N: "#08306B", CON_L: "#F0B24B"}
SRC_CONTENT, SRC_SYNTAX = "#000000", "#8c8c8c"   # panel-c source colours, kept off the a/b palette


def group_of(r):
    if r["cell"] == "A":
        return SYN
    return CON_L if r["seed_content_bearing"] == "1" else CON_N


def cos_to_seed(r, g):
    # content source of a syntax variant is the seed; of a content variant it is the donor
    return float(r["cos_content"]) if g == SYN else float(r["cos_syntax"])


def cos_to_donor(r, g):
    return float(r["cos_syntax"]) if g == SYN else float(r["cos_content"])


def short(e):
    return e.replace("-en-v1.5", "").replace("-embedding-0.6b", "").replace("-base-v2", "")


def box_panel(ax, order, data, title, ylabel):
    rng = np.random.default_rng(0)
    offs = np.array([-0.26, 0.0, 0.26]); bw = 0.19
    for i in range(len(order)):
        if i % 2 == 1:
            ax.axvspan(i - 0.5, i + 0.5, color="0.94", zorder=0)
    for i, e in enumerate(order):
        for j, g in enumerate(GROUPS):
            vals = np.asarray(data[e][g]); xc = i + offs[j]
            if not len(vals):
                continue
            ax.scatter(xc + rng.normal(0, 0.03, len(vals)), vals, s=1.5, color=COL[g],
                       alpha=0.10, edgecolors="none", zorder=1, rasterized=True)
            ax.boxplot([vals], positions=[xc], widths=bw, showfliers=False, patch_artist=True,
                       zorder=3, medianprops=dict(color="black", lw=1.3),
                       boxprops=dict(facecolor=COL[g], alpha=0.85, edgecolor="black", lw=0.7),
                       whiskerprops=dict(color="black", lw=0.8), capprops=dict(color="black", lw=0.8))
    ax.set_xticks(range(len(order)))
    ax.set_xticklabels([short(e) for e in order], rotation=18, ha="right", fontsize=10)
    ax.set_xlim(-0.55, len(order) - 0.45); ax.set_ylim(-0.05, 1.0)
    ax.set_ylabel(ylabel, fontsize=11)
    ax.set_title(title, fontsize=12, loc="left")
    ax.tick_params(labelsize=9.5)
    ax.spines[["top", "right"]].set_visible(False)


def main():
    ap = argparse.ArgumentParser()
    ap.add_argument("--long", required=True)
    ap.add_argument("--out", required=True)
    ap.add_argument("--preview", default="/data_gpu5/ROSS_Proj/figure_preview.png")
    ap.add_argument("--dpi", type=int, default=170)
    args = ap.parse_args()

    rows = list(csv.DictReader(open(args.long)))
    to_seed = defaultdict(lambda: defaultdict(list))
    to_donor = defaultdict(lambda: defaultdict(list))
    clean_cc = defaultdict(list)   # clean-variant cosine to content source
    clean_cs = defaultdict(list)   # clean-variant cosine to syntax source
    for r in rows:
        g = group_of(r); e = r["encoder"]
        to_seed[e][g].append(cos_to_seed(r, g))
        to_donor[e][g].append(cos_to_donor(r, g))
        if g != CON_L:                                   # clean = excludes the jingle case
            clean_cc[e].append(float(r["cos_content"]))
            clean_cs[e].append(float(r["cos_syntax"]))

    encoders = list(to_seed)
    order = sorted(encoders, key=lambda e: -(np.median(clean_cc[e]) - np.median(clean_cs[e])))

    fig = plt.figure(figsize=(15, 10.5))
    gs = fig.add_gridspec(2, 2, height_ratios=[1.32, 1.0], hspace=0.34, wspace=0.16,
                          top=0.88, bottom=0.06, left=0.065, right=0.985)
    axA = fig.add_subplot(gs[0, 0]); axB = fig.add_subplot(gs[0, 1]); axC = fig.add_subplot(gs[1, :])

    box_panel(axA, order, to_seed, "(a) Cosine to the seed", "cosine to the seed")
    box_panel(axB, order, to_donor, "(b) Cosine to the donor", "cosine to the donor")

    # shared group legend, outside at the top
    fig.legend(handles=[Patch(facecolor=COL[g], edgecolor="black", lw=0.5, label=LABEL[g])
                        for g in GROUPS],
               loc="upper center", bbox_to_anchor=(0.5, 0.955), ncol=3,
               frameon=False, fontsize=10.5)

    # ---- (c) dumbbell: content source vs syntax source per encoder (clean variants) ----
    x = np.arange(len(order))
    print("\nencoder            content-src  syntax-src   gap")
    for i, e in enumerate(order):
        mc, ms = np.median(clean_cc[e]), np.median(clean_cs[e])
        axC.plot([x[i], x[i]], [ms, mc], "-", color="#cccccc", lw=3, zorder=1)
        axC.scatter([x[i]], [mc], color=SRC_CONTENT, s=110, zorder=3)
        axC.scatter([x[i]], [ms], color=SRC_SYNTAX, s=75, zorder=3)
        axC.annotate(f"{mc - ms:+.2f}", (x[i], mc), xytext=(0, 9), textcoords="offset points",
                     ha="center", fontsize=9.5, color="#222", fontweight="bold")
        print(f"{short(e):18s}  {mc:8.3f}   {ms:8.3f}   {mc - ms:+.3f}")
    axC.scatter([], [], color=SRC_CONTENT, s=110, label="cosine to content source")
    axC.scatter([], [], color=SRC_SYNTAX, s=75, label="cosine to syntax source")
    axC.set_xticks(x); axC.set_xticklabels([short(e) for e in order], fontsize=10.5)
    axC.set_xlim(-0.5, len(order) - 0.5); axC.set_ylim(0.15, 0.9)
    axC.set_ylabel("median cosine to source", fontsize=11)
    axC.set_title("(c) Each variant sits closer to its content source than its syntax source  "
                  "(gap = content − syntax)", fontsize=12, loc="left")
    axC.legend(fontsize=9.5, loc="lower right", frameon=True, framealpha=0.95, edgecolor="0.8")
    axC.tick_params(labelsize=9.5)
    axC.spines[["top", "right"]].set_visible(False)

    fig.savefig(args.out, dpi=args.dpi, bbox_inches="tight")
    print(f"\n-> {args.out}")
    if args.preview:
        fig.savefig(args.preview, dpi=args.dpi, bbox_inches="tight")
        print(f"-> {args.preview}")


if __name__ == "__main__":
    main()
```
