## Supplemental code for "Scoring respondents on a semantic frame of reference identifies a group that is burdened across many questionnaires and reaches care less easily": fig_corecomp.py

```
#!/usr/bin/env python
"""Figure: core components place every item, including those that define no component.

(a) The standardized loadings of one exemplar item on all pool components, against each component's
    core-item threshold band, with the item's own core components marked and named.
(b) How often an item's own construct appears among the constructs of its core components, for items
    that core some component and for items that core none, against a label-shuffle null.

Everything is read from saved artefacts - no numbers, labels or component ids are hardcoded:
    outputs_pool/fits/<enc>__k*_ls1.0/scores.npy      the pool projection P_G
    outputs_pool/fixedpoint_cores/<enc>_cores.csv     core items per component
    ../global_pool/pool_processed.csv                 item text and documented construct

The only configuration is which item to feature and how panel (b) is scored.

LABELLING (panel b and the panel-a annotations)
    "profile"  a component contributes the set of constructs of its core items          (D6-A)
    "modal"    a component contributes the single most common construct of its core     (retired)
Panel (b) is always leave-one-out: an item never counts its own membership of a core, which would
otherwise make the core-item group 100 percent by construction.

Out: manuscript_nhb/figures/fig_corecomp.png
Run: CUDA_VISIBLE_DEVICES="" /data_gpu5/ROSS_Proj/env/bin/python fig_corecomp.py
"""
import os, glob, csv
import numpy as np
import pandas as pd
import matplotlib
matplotlib.use("Agg")
import matplotlib.pyplot as plt

HERE = os.path.dirname(os.path.abspath(__file__))
REPO = os.path.abspath(os.path.join(HERE, "..", ".."))
GP = os.path.join(HERE, "..", "global_pool")
OUT = os.path.join(REPO, "manuscript_nhb", "figures")

ENC = "gte-large-en-v1.5"
ITEM = "DJG_3"                 # the featured item, keyed into pool_processed.csv
LABELLING = "profile"          # "profile" (D6-A) or "modal" (retired, reproduces the published panel)
NPERM = 200
# Okabe-Ito blue/orange are colour-blind safe but sit at similar lightness, so the two groups are
# also separated by texture and by their axis labels, never by hue alone.
BLUE, ORANGE, GREY, INK = "#08306B", "#D55E00", "#D9D9D9", "#222222"
BARHATCH = ["", "///"]


def pr(v):
    s2 = float((v ** 2).sum()); s4 = float((v ** 4).sum())
    return s2 * s2 / s4 if s4 > 0 else 0.0


def fpc(v):
    idx = np.arange(len(v))
    while True:
        n = max(1, min(int(round(pr(v[idx]))), len(idx)))
        top = np.sort(idx[np.argsort(-np.abs(v[idx]))[:n]])
        if len(top) == len(idx) and (top == idx).all():
            return top
        idx = top


def load():
    pp = pd.read_csv(os.path.join(GP, "pool_processed.csv"))
    fit = glob.glob(os.path.join(HERE, "outputs_pool", "fits", f"{ENC}__k*_ls1.0"))[0]
    Z = np.load(os.path.join(fit, "scores.npy"))
    Zt = Z / Z.std(0)                                   # pool sigma, columns already zero-mean
    rows = list(csv.DictReader(open(os.path.join(HERE, "outputs_pool", "fixedpoint_cores",
                                                 f"{ENC}_cores.csv"))))
    core = {int(r["component"]): str(r["items"]).split() for r in rows}
    return pp, Zt, core


def component_constructs(core, con, comps, drop=None):
    """The constructs a set of components contributes, under the configured labelling."""
    out = set()
    for c in comps:
        items = [h for h in core[c] if h != drop]
        if not items:
            continue
        if LABELLING == "modal":
            out.add(pd.Series([con[h] for h in items]).value_counts().idxmax())
        else:
            out.update(con[h] for h in items)
    return out


def main():
    pp, Zt, core = load()
    con = dict(zip(pp.item_id, pp.construct))
    text = dict(zip(pp.item_id, pp.prompt_raw))
    ids = list(pp.item_id)
    anchored = {h for v in core.values() for h in v}

    F = {g: [int(c) for c in fpc(Zt[j])] for j, g in enumerate(ids)}

    # ---------- panel (b): own construct among the core components' constructs ----------
    def rate(sub, lab):
        return np.mean([lab[g] in component_constructs(core, lab, F[g], drop=g) for g in sub])

    groups = [("items that core\na component", [g for g in ids if g in anchored], BLUE),
              ("items that core\nno component", [g for g in ids if g not in anchored], ORANGE)]
    plt.rcParams["hatch.linewidth"] = 0.8
    rng = np.random.RandomState(0)
    bars = []
    for name, sub, colour in groups:
        real = rate(sub, con)
        null = np.mean([rate(sub, dict(zip(ids, rng.permutation(list(con.values())))))
                        for _ in range(NPERM)])
        bars.append((name, colour, 100 * real, 100 * null, len(sub)))
        print(f"{name.replace(chr(10), ' '):28s} n={len(sub):3d}  {100*real:5.1f}%  chance {100*null:5.1f}%")

    # ---------- panel (a): the featured item against every component's threshold band ----------
    j = ids.index(ITEM)
    z = Zt[j]
    thresh = np.array([min(abs(Zt[ids.index(h), c]) for h in core[c]) for c in range(Zt.shape[1])])
    mine = sorted(F[ITEM], key=lambda c: -abs(z[c]))
    names = {}
    for c in mine:
        cs = pd.Series([con[h] for h in core[c]]).value_counts()
        names[c] = cs.index[0] if LABELLING == "modal" or len(cs) == 1 else \
            f"{cs.index[0]}\n+ {cs.index[1]}"

    fig, (ax, bx) = plt.subplots(1, 2, figsize=(13.0, 5.2), gridspec_kw=dict(width_ratios=[2.35, 1]))
    x = np.arange(len(z))
    ax.fill_between(x, -thresh, thresh, step="mid", color=GREY, lw=0.8, edgecolor="#B9B9B9",
                    label="core-item threshold band", zorder=1)
    ax.plot(x, z, "-o", color=ORANGE, ms=3.4, lw=1.2, mfc="white", mew=1.0, zorder=3,
            label=f"its {len(mine)} core components")
    ax.plot([c for c in mine], [z[c] for c in mine], "o", color=ORANGE, ms=9, zorder=4)
    ax.axhline(0, color="#AAAAAA", lw=0.8, zorder=2)
    lim = 1.12 * max(np.abs(z).max(), float(np.percentile(thresh, 92)))
    # stagger the annotations: two rows on each side, and nudge apart components that sit close
    rows = {True: [0.62, 0.86], False: [0.58, 0.84]}
    used = {True: 0, False: 0}
    placed = []
    for c in mine:
        up = bool(z[c] > 0)
        y = lim * rows[up][used[up] % 2] * (1 if up else -1)
        used[up] += 1
        xt = c
        while any(abs(xt - px) < 7 and abs(y - py) < 0.2 * lim for px, py in placed):
            xt += 6
        placed.append((xt, y))
        ax.annotate(names[c], xy=(c, z[c]), xytext=(xt, y),
                    ha="center", va="bottom" if up else "top", fontsize=9, fontweight="bold",
                    color=INK, linespacing=1.25,
                    arrowprops=dict(arrowstyle="-", color=INK, lw=0.8, shrinkA=2, shrinkB=6))
    ax.set_xlabel("component"); ax.set_ylabel("standardized loading  $z$")
    ax.set_ylim(-lim, lim); ax.set_xlim(-1, len(z))
    ax.set_title(f'a   "{str(text[ITEM]).rstrip(".")}" — a core item of no component',
                 loc="left", fontsize=12, fontweight="bold", color=INK)
    ax.legend(loc="upper left", fontsize=9, framealpha=0.95)
    for s in ("top", "right"):
        ax.spines[s].set_visible(False)

    for i, (name, colour, real, null, n) in enumerate(bars):
        bx.bar(i, real, width=0.62, color=colour, zorder=2, hatch=BARHATCH[i],
               edgecolor="white", linewidth=0.0)
        bx.text(i, real + 1.8, f"{real:.0f}%", ha="center", fontsize=13, fontweight="bold", color=INK)
        bx.hlines(null, i - 0.31, i + 0.31, color=INK, lw=2.2, linestyle="--", zorder=3)
        bx.text(i + 0.34, null, "chance", ha="left", va="center", fontsize=8.5, color=INK)
    bx.set_xticks(range(len(bars))); bx.set_xticklabels([b[0] for b in bars], fontsize=10)
    bx.set_xlim(-0.55, len(bars) - 0.15)
    bx.set_ylim(0, 100); bx.set_ylabel("placed in own construct (%)")
    bx.set_title("b   Placement is\n     construct-faithful", loc="left", fontsize=12,
                 fontweight="bold", color=INK)
    for s in ("top", "right"):
        bx.spines[s].set_visible(False)

    fig.subplots_adjust(left=0.055, right=0.985, top=0.87, bottom=0.14, wspace=0.28)
    out = os.path.join(OUT, "fig_corecomp.png")
    fig.savefig(out, dpi=300, facecolor="white")
    print("wrote", out, f"| {LABELLING} labelling, item {ITEM}, {len(mine)} core components")


if __name__ == "__main__":
    main()
```
