## Supplemental code for "Scoring respondents on a semantic frame of reference identifies a group that is burdened across many questionnaires and reaches care less easily": fig_coverage.py

```
#!/usr/bin/env python
"""Figure: how much of each question the fixed basis holds, for the pool and the two ported batteries.

Placing a battery on the reference basis is not all or nothing. For every item this measures
COVERAGE, the fraction of the item's embedding about the pool's centre that lies in the span of the
components the battery keeps,

    coverage_g = || V^T (x_g - mu_pool) ||^2 / || x_g - mu_pool ||^2,

with V an orthonormal basis of the row space of the KEPT rows of the pool loading matrix. This is the
same quantity as porting_quality.py reports per item, kept per encoder here rather than averaged, so
that the six encoders can be drawn as separate boxes.

Three groups on the x axis:
    Reference pool   the items the basis was built from. Every component is kept, so this is the
                     IN-SAMPLE CEILING and not a peer of the other two -- drawn hatched and labelled
                     as such in the caption.
    All of Us        the 151 items, of which the pool holds no text for three instruments.
    HRS              the 35 Leave-Behind social items.

The kept set differs by battery (HRS reaches fewer components than All of Us) and k itself differs by
encoder, so the encoder hues are NOT an encoder ranking -- part of any spread between them is the
number of components available. Said in the caption.

Reads saved artefacts only -- no result or count is hardcoded:
    outputs_pool/fits/<enc>__k*_ls1.0/{loadings,mean,scores}.npy   the pool basis per encoder
    outputs_pool/fixedpoint_cores/<enc>_cores.csv                  pool core items per component
    data/pool_items_<enc>.npz                                      pool item embeddings
    data/core151_items_<enc>.npz                                   All of Us item embeddings
    ../randhrsdata/acidd_input/hrs_lb2022_<enc>.npz                HRS item embeddings
    ../global_pool/pool_processed.csv                              the pool item list

Out: manuscript_nhb/figures/fig_coverage.png
Run: CUDA_VISIBLE_DEVICES="" /data_gpu5/ROSS_Proj/env/bin/python fig_coverage.py
"""
import glob
import os

import numpy as np
import pandas as pd
import matplotlib
matplotlib.use("Agg")
import matplotlib.pyplot as plt
from matplotlib.patches import Patch

from _common import keep_columns, pool_bands
from hrs_kept_components import ENCS, HERE, GP, HRSD

REPO = os.path.abspath(os.path.join(HERE, "..", ".."))
OUT = os.path.join(REPO, "manuscript_nhb", "figures")

# Okabe-Ito, six of the eight (yellow and black dropped: yellow is too light against white boxes).
ENC_COLOR = {"gte-large-en-v1.5": "#0072B2", "bge-m3": "#E69F00",
             "bge-large-en-v1.5": "#009E73", "all-mpnet-base-v2": "#CC79A7",
             "e5-large-v2": "#56B4E9", "qwen3-embedding-0.6b": "#D55E00"}
ENC_SHORT = {"gte-large-en-v1.5": "gte-large", "bge-m3": "bge-m3",
             "bge-large-en-v1.5": "bge-large", "all-mpnet-base-v2": "mpnet",
             "e5-large-v2": "e5-large", "qwen3-embedding-0.6b": "qwen3"}
INK = "#222222"

BATTERIES = [("Reference pool", os.path.join(HERE, "data", "pool_items_{enc}.npz")),
             ("All of Us", os.path.join(HERE, "data", "core151_items_{enc}.npz")),
             ("HRS", os.path.join(HRSD, "hrs_lb2022_{enc}.npz"))]


def coverage(enc, path_tmpl, pool_ids):
    """Per-item coverage of one battery under one encoder, plus how many components it keeps."""
    fit = glob.glob(os.path.join(HERE, "outputs_pool", "fits", f"{enc}__k*_ls1.0"))[0]
    W = np.load(os.path.join(fit, "loadings.npy"))
    mu = np.load(os.path.join(fit, "mean.npy"))
    P = np.load(os.path.join(fit, "scores.npy"))
    sigma = P.std(0)
    Zpool = (P - P.mean(0)) / sigma

    d = np.load(path_tmpl.format(enc=enc), allow_pickle=True)
    X = np.asarray(d["vectors"], float) - mu
    Z = (X @ W.T) / sigma

    kept = sorted(set(keep_columns(Z, pool_bands(enc, Zpool, pool_ids))))
    V = np.linalg.svd(W[kept], full_matrices=False)[2]
    cov = ((X @ V.T) ** 2).sum(1) / (X ** 2).sum(1)
    return 100.0 * cov, len(kept), W.shape[0]


def collect():
    pool_ids = pd.read_csv(os.path.join(GP, "pool_processed.csv"))["item_id"].tolist()
    data, kept = {}, {}
    for name, tmpl in BATTERIES:
        for enc in ENCS:
            c, nk, k = coverage(enc, tmpl, pool_ids)
            data[(name, enc)] = c
            kept[(name, enc)] = (nk, k)
    return data, kept


def report(data, kept):
    print(f"\n{'='*84}\ncoverage, per battery per encoder (% of an item held by the kept components)\n")
    print(f"  {'battery':16s} {'encoder':12s} {'n':>4s} {'kept/k':>8s} {'median':>8s} "
          f"{'IQR':>14s} {'min':>7s} {'max':>7s}")
    for name, _ in BATTERIES:
        for enc in ENCS:
            c = data[(name, enc)]
            nk, k = kept[(name, enc)]
            q1, q3 = np.percentile(c, [25, 75])
            print(f"  {name:16s} {ENC_SHORT[enc]:12s} {len(c):4d} {f'{nk}/{k}':>8s} "
                  f"{np.median(c):7.1f}% {f'{q1:.1f}-{q3:.1f}':>14s} {c.min():6.1f}% {c.max():6.1f}%")
        pooled = np.concatenate([data[(name, e)] for e in ENCS])
        print(f"  {'':16s} {'-- all six':12s} {'':4s} {'':>8s} {np.median(pooled):7.1f}%\n")


def draw(data, kept, out):
    fig, ax = plt.subplots(figsize=(6.8, 3.2))
    width, gap = 0.115, 0.30
    KEPT_ROW = 96.0          # the per-encoder kept-column counts sit in the panel's empty top
    centers = []
    for gi, (name, _) in enumerate(BATTERIES):
        c0 = gi * (6 * width + gap)
        centers.append(c0 + 2.5 * width)
        for ei, enc in enumerate(ENCS):
            pos = c0 + ei * width
            bp = ax.boxplot([data[(name, enc)]], positions=[pos], widths=width * 0.82,
                            patch_artist=True, showfliers=False, whis=(5, 95), zorder=3)
            face = ENC_COLOR[enc]
            for b in bp["boxes"]:
                b.set(facecolor=face, edgecolor=INK, linewidth=0.6, alpha=0.95)
                if name == "Reference pool":
                    b.set(hatch="///", alpha=0.55)
            for part in ("whiskers", "caps"):
                for a in bp[part]:
                    a.set(color=INK, linewidth=0.6)
            for m in bp["medians"]:
                m.set(color=INK, linewidth=1.1)
            ax.text(pos, KEPT_ROW, f"{kept[(name, enc)][0]}", ha="center", va="center",
                    fontsize=5.8, color=INK)

    ax.set_xticks(centers)
    ax.set_xticklabels([f"{n}\n({len(data[(n, ENCS[0])])} items)" for n, _ in BATTERIES], fontsize=8.5)
    ax.set_ylabel("held by the basis (% of a question)", fontsize=8.5)
    ax.set_ylim(0, 103)
    ax.text(0.004, (KEPT_ROW + 4.4) / 103, "components kept", transform=ax.transAxes,
            ha="left", va="center", fontsize=5.8, style="italic", color="#666666")
    ax.tick_params(axis="y", labelsize=8)
    ax.grid(axis="y", color="#E6E6E6", linewidth=0.6, zorder=0)
    ax.set_axisbelow(True)
    for s in ("top", "right"):
        ax.spines[s].set_visible(False)
    for s in ("left", "bottom"):
        ax.spines[s].set(color=INK, linewidth=0.7)

    leg = [Patch(facecolor=ENC_COLOR[e], edgecolor=INK, linewidth=0.6, label=ENC_SHORT[e])
           for e in ENCS]
    ax.legend(handles=leg, loc="lower center", bbox_to_anchor=(0.5, 1.005), ncol=6,
              frameon=False, fontsize=7.4, handlelength=1.2, handleheight=0.9,
              columnspacing=1.1, borderaxespad=0.0)

    fig.savefig(out, dpi=400, bbox_inches="tight", facecolor="white")
    print(f"  wrote {out}")


if __name__ == "__main__":
    data, kept = collect()
    report(data, kept)
    os.makedirs(OUT, exist_ok=True)
    draw(data, kept, os.path.join(OUT, "fig_coverage.png"))
```
