## Supplemental code for "Scoring respondents on a semantic frame of reference identifies a group that is burdened across many questionnaires and reaches care less easily": fig_hrs_loadings.py

```
#!/usr/bin/env python
"""Supplement: every HRS item against every component of the reference basis, one panel per encoder.

Cell shade is the item's standardized loading magnitude on that component, |Z_S|, the same quantity the
placement rule reads. A cell is outlined when that component is one of the item's CORE COMPONENTS, so
the retain rule is visible directly: a component is retained for the battery when at least one item
cores it, and the columns with no outlined cell -- the ones HRS never reaches -- are hatched over.

Rows are the 35 HRS items grouped by instrument, columns are the components in index order, so the
column count differs per encoder (49 to 55).

Reads saved artefacts only:
    outputs_pool/fits/<enc>__k*_ls1.0/{loadings,mean}.npy   the pool basis per encoder
    data/pool_items_<enc>.npz                               pool embeddings, for the column scale
    ../randhrsdata/acidd_input/hrs_lb2022_<enc>.npz         HRS item embeddings
    ../randhrsdata/acidd_input/hrs_lb2022_items.csv         HRS instrument and construct per item

Out: manuscript_nhb/figures/figS_hrs_loadings.png
Run: CUDA_VISIBLE_DEVICES="" /data_gpu5/ROSS_Proj/env/bin/python fig_hrs_loadings.py
"""
import csv
import glob
import os

import numpy as np
import pandas as pd
import matplotlib
matplotlib.use("Agg")
import matplotlib.pyplot as plt
from matplotlib.patches import Rectangle, Patch

from _common import keep_columns, pool_bands

HERE = os.path.dirname(os.path.abspath(__file__))
REPO = os.path.abspath(os.path.join(HERE, "..", ".."))
HRSD = os.path.join(HERE, "..", "randhrsdata", "acidd_input")
POOL_IDS = pd.read_csv(os.path.join(HERE, "..", "global_pool", "pool_processed.csv"))["item_id"].tolist()
OUT = os.path.join(REPO, "manuscript_nhb", "figures")

ENCS = ["gte-large-en-v1.5", "bge-m3", "bge-large-en-v1.5", "all-mpnet-base-v2",
        "e5-large-v2", "qwen3-embedding-0.6b"]
SHORT = {"UCLA Loneliness (R-UCLA)": "R-UCLA", "PSS-10": "PSS-10",
         "HRS Neighborhood (bipolar)": "HRS neighborhood",
         "Williams Everyday Discrimination": "EDS", "Williams Discrimination (healthcare)": "Williams-HC"}
INK = "#222222"


def pr(v):
    s2 = float((v ** 2).sum()); s4 = float((v ** 4).sum())
    return s2 * s2 / s4 if s4 > 0 else 0.0


def fpc(v):
    idx = np.arange(len(v))
    while True:
        n = max(1, min(int(round(pr(v[idx]))), len(idx)))
        top = np.sort(idx[np.argsort(-np.abs(v[idx]))[:n]])
        if len(top) == len(idx) and (top == idx).all():
            return top
        idx = top


def load(enc):
    fit = glob.glob(os.path.join(HERE, "outputs_pool", "fits", f"{enc}__k*_ls1.0"))[0]
    W = np.load(os.path.join(fit, "loadings.npy")); mu = np.load(os.path.join(fit, "mean.npy"))
    Mg = np.load(os.path.join(HERE, "data", f"pool_items_{enc}.npz"),
                 allow_pickle=True)["vectors"].astype(float)
    sigma = ((Mg - mu) @ W.T).std(0)
    d = np.load(os.path.join(HRSD, f"hrs_lb2022_{enc}.npz"), allow_pickle=True)
    ids = [str(x) for x in d["items"]]
    Z = ((np.asarray(d["vectors"], float) - mu) @ W.T) / sigma
    Zpool = ((Mg - mu) @ W.T) / sigma
    return ids, Z, pool_bands(enc, Zpool, POOL_IDS)


def main():
    hrs = pd.read_csv(os.path.join(HRSD, "hrs_lb2022_items.csv"))
    inst = dict(zip(hrs.item, hrs.instrument))

    ids0, _, _ = load(ENCS[0])
    order = sorted(range(len(ids0)), key=lambda i: (SHORT.get(inst[ids0[i]], "?"), ids0[i]))
    rows = [ids0[i] for i in order]
    blocks, start = [], 0
    for r in range(1, len(rows) + 1):
        if r == len(rows) or inst[rows[r]] != inst[rows[start]]:
            blocks.append((start, r, SHORT.get(inst[rows[start]], inst[rows[start]])))
            start = r

    fig, axes = plt.subplots(3, 2, figsize=(11.4, 12.0), facecolor="white")
    for ax, enc in zip(axes.ravel(), ENCS):
        ids, Z, band = load(enc)
        pos = {g: i for i, g in enumerate(ids)}
        M = np.abs(np.array([Z[pos[r]] for r in rows]))
        core = [set(int(c) for c in fpc(Z[pos[r]])) for r in rows]
        # dropped = not kept by the rule of _common.keep_columns, which is not the same as
        # being in no item's core: a component in several items' cores is dropped unless one
        # of them anchors it or peaks on it.
        retained = keep_columns(np.array([Z[pos[r]] for r in rows]), band)
        dropped = [c for c in range(M.shape[1]) if c not in set(retained)]

        ax.imshow(M, aspect="auto", cmap="Blues", vmin=0, vmax=float(np.percentile(M, 99)),
                  interpolation="nearest")
        for i, cs in enumerate(core):
            for c in cs:
                ax.add_patch(Rectangle((c - 0.5, i - 0.5), 1, 1, fc="none", ec="#D55E00", lw=0.7,
                                       zorder=4))
        for c in dropped:
            ax.add_patch(Rectangle((c - 0.5, -0.5), 1, M.shape[0], fc="none", ec="#6E6E6E",
                                   hatch="////", lw=0.0, alpha=0.85, zorder=3))
        for s, e, _ in blocks[:-1]:
            ax.axhline(e - 0.5, color="white", lw=1.6)
        ax.set_yticks([(s + e - 1) / 2 for s, e, _ in blocks])
        ax.set_yticklabels([lab for _, _, lab in blocks], fontsize=6.4)
        ax.set_xticks(np.arange(0, M.shape[1], 10))
        ax.tick_params(labelsize=6.2, length=2)
        ax.set_xlabel("component", fontsize=7)
        ax.set_title(f"{enc}   {len(retained)} of {M.shape[1]} components retained",
                     fontsize=7.8, color=INK)

    leg = [Patch(fc="#08306B", ec="none", label="larger |loading|"),
           Patch(fc="none", ec="#D55E00", lw=0.9, label="a core component of that item"),
           Patch(fc="none", ec="#6E6E6E", hatch="////", label="component no HRS item cores (dropped)")]
    fig.legend(handles=leg, loc="lower center", ncol=3, frameon=False, fontsize=7.6,
               handlelength=1.6, handleheight=1.1, bbox_to_anchor=(0.5, 0.005))
    fig.subplots_adjust(left=0.10, right=0.985, top=0.965, bottom=0.055, hspace=0.30, wspace=0.24)
    out = os.path.join(OUT, "figS_hrs_loadings.png")
    fig.savefig(out, dpi=300, facecolor="white")
    print("wrote", out, f"| {len(rows)} HRS items x {len(ENCS)} encoders")


if __name__ == "__main__":
    main()
```
