## Supplemental code for "Scoring respondents on a semantic frame of reference identifies a group that is burdened across many questionnaires and reaches care less easily": fig_hrs_twoside_cores.py

```
#!/usr/bin/env python
"""Two-sided core-item ribbon: do the matching instruments core the SAME pool columns?

Both sides show CORE ITEMS of the same fixed column (D4), nothing else:
  LEFT   pool instruments whose items form E_i^G  (which pool instruments define the axis)
  RIGHT  HRS instruments whose items form E_i^S   (which HRS items core it)
Scope = the pool columns whose own core carries a construct HRS also measures, so the question
"does the matching instrument core the same column?" is well posed. Colour = the shared construct
documented construct, grey = any other construct, so agreement is a colour match across the centre.
Constructs are never grouped: loneliness and social isolation are two colours, as are neighborhood
disorder and neighborhood social cohesion. Related constructs share a hue and differ in lightness.

Right-hand items that core the column WITHOUT leaning on it (column not among their core
components, D5) are drawn hatched: these are the passengers that a 35-item field admits.

Out: examples/global_pool/figures/fig_hrs_twoside_cores.png          (gte-large, main text)
     examples/global_pool/figures/fig_hrs_twoside_cores_<enc>.png    (the other five, Supplementary S18)
Run: CUDA_VISIBLE_DEVICES="" /data_gpu5/ROSS_Proj/env/bin/python fig_hrs_twoside_cores.py [--all]
"""
import os, sys, glob, csv, re
from collections import Counter
import numpy as np, pandas as pd
import matplotlib; matplotlib.use("Agg")
import matplotlib.pyplot as plt
from matplotlib.path import Path
from matplotlib.patches import PathPatch, Rectangle, Patch

from _common import keep_columns, pool_bands
from pool_card_labels import short   # the margin labels must fit the print-size margin

HERE = os.path.dirname(os.path.abspath(__file__))
REPO = os.path.abspath(os.path.join(HERE, "..", ".."))
HRSD = os.path.join(HERE, "..", "randhrsdata", "acidd_input")
POOL_IDS = pd.read_csv(os.path.join(HERE, "..", "global_pool", "pool_processed.csv"))["item_id"].tolist()
OUT = os.path.join(REPO, "examples", "global_pool", "figures"); os.makedirs(OUT, exist_ok=True)
EXEMPLAR = "gte-large-en-v1.5"          # main text
ENCS = [EXEMPLAR, "bge-m3", "bge-large-en-v1.5", "all-mpnet-base-v2",
        "e5-large-v2", "qwen3-embedding-0.6b"]

# one key per DOCUMENTED construct, no grouping. Pool labels are renamed to the batteries' spelling
# for the two neighborhood constructs (the single clerical pairing, Methods sec:methods_hrs).
FAM = {"Everyday discrimination": "EDS", "Healthcare discrimination": "DMS",
       "Loneliness": "LON", "Social isolation": "ISO", "Perceived stress": "PSS",
       "Neighborhood disorder": "NBD", "Neighborhood cohesion": "NBC"}
HFAM = {"Everyday discrimination": "EDS", "Healthcare discrimination": "DMS", "Loneliness": "LON",
        "Perceived stress": "PSS", "Neighborhood disorder & safety": "NBD",
        "Neighborhood social cohesion": "NBC"}
# paired hues, split on lightness so each pair stays separable in greyscale
FCOL = {"EDS": "#0072B2", "DMS": "#8FCDF0", "LON": "#009E73", "ISO": "#084C3F", "PSS": "#E69F00",
        "NBD": "#CC79A7", "NBC": "#6E2F52", "other": "#BFBFBF"}
FLAB = {"EDS": "everyday discrimination", "DMS": "healthcare discrimination", "LON": "loneliness",
        "ISO": "social isolation", "PSS": "perceived stress", "NBD": "neighborhood disorder",
        "NBC": "neighborhood social cohesion", "other": "other construct"}
# HRS documents its eight neighborhood items as two scales, split here by the published item text
HRS_COHESION = ("really feel part of this area", "can be trusted", "are friendly", "were in trouble")
HRS_DISORDER = ("vandalism and graffiti", "safe walking alone", "kept very clean", "vacant houses")


def counts_by_construct(labels):
    """Documented constructs with their item counts, most common first.

    Ties are broken alphabetically. `value_counts` alone breaks them in hash-table order, which
    moves with Python's string-hash randomization, so two runs of the same generator produced
    cards and CSV rows that listed equal-count constructs in a different order. No count or
    threshold ever changed, but the figures were not byte-reproducible.
    """
    return (pd.Series(list(labels)).value_counts()
            .sort_index().sort_values(ascending=False, kind="mergesort"))


def hrs_construct(label, prompt):
    if label != "Neighborhood":
        return label
    t = str(prompt).lower()
    if any(k in t for k in HRS_COHESION):
        return "Neighborhood social cohesion"
    if any(k in t for k in HRS_DISORDER):
        return "Neighborhood disorder & safety"
    return label
HSHORT = {"Williams Everyday Discrimination": "Williams EDS", "Williams Discrimination (healthcare)": "Williams DMS",
          "UCLA Loneliness (R-UCLA)": "R-UCLA", "PSS-10": "PSS-10", "HRS Neighborhood (bipolar)": "HRS Nbhd"}
INK = "#222222"


# presentation only: the published short name of each pool instrument, so the axis is readable
POOLSHORT = {"Adverse Childhood Experiences Questionnaire": "Felitti ACE",
             "Center for Epidemiologic Studies Depression Scale": "CES-D",
             "Discrimination in Medical Settings": "DMS",
             "Everyday Discrimination Scale": "EDS",
             "Generalized Anxiety Disorder-7": "GAD-7",
             "Heightened Vigilance Scale": "HVS",
             "Major Experiences of Discrimination": "MED",
             "Neighborhood Social Cohesion": "SCNS",
             "Patient Health Questionnaire-9": "PHQ-9",
             "Rosenberg Self-Esteem Scale": "RSES",
             "Three-Item UCLA Loneliness Scale": "UCLA-3",
             "UK Biobank Mental Health Questionnaire": "MHQ",
             "WHO World Mental Health CIDI": "WMH-CIDI",
             "WHO-5 Well-Being Index": "WHO-5"}
PAREN = {"12-item": "ISEL-12", "AoU adapted": "NDS", "50-item": "IPIP-50", "6-item": "USDA-FS",
         "R-UCLA": "R-UCLA", "3-item": "UCLA-3", "10-item": "PSS-10",
         "6-item Short Form": "USDA-FS", "SF 6a": "PROMIS-Support"}


def poolshort(s):
    if s in POOLSHORT:
        return POOLSHORT[s]
    m = re.findall(r"\(([^)]+)\)", s)
    if m:
        return PAREN.get(m[-1], m[-1])
    s = re.sub(r"^PROMIS Item Bank v[0-9.]+ - (Emotional Distress - )?", "PROMIS-", s)
    return s.replace("PROMIS-Social Isolation", "PROMIS-Isolation")[:20]


def pr(v):
    s2 = float((v**2).sum()); s4 = float((v**4).sum()); return s2*s2/s4 if s4 > 0 else 0.0


def fpc(v):
    idx = np.arange(len(v))
    while True:
        n = max(1, min(int(round(pr(v[idx]))), len(idx)))
        top = np.sort(idx[np.argsort(-np.abs(v[idx]))[:n]])
        if len(top) == len(idx) and (top == idx).all():
            return top
        idx = top


def ribbon(ax, x0, ya0, ya1, x1, yb0, yb1, color, alpha, hatch=None):
    cx = (x0 + x1) / 2.0
    v = [(x0, ya0), (cx, ya0), (cx, yb0), (x1, yb0), (x1, yb1), (cx, yb1), (cx, ya1), (x0, ya1), (x0, ya0)]
    c = [Path.MOVETO, Path.CURVE4, Path.CURVE4, Path.CURVE4, Path.LINETO,
         Path.CURVE4, Path.CURVE4, Path.CURVE4, Path.CLOSEPOLY]
    ax.add_patch(PathPatch(Path(v, c), facecolor=color, edgecolor=("white" if hatch else "none"),
                           hatch=hatch, lw=0.0, alpha=alpha, zorder=1))


def main(ENC=EXEMPLAR):
    fit = glob.glob(os.path.join(HERE, "outputs_pool", "fits", f"{ENC}__k*_ls1.0"))[0]
    W = np.load(os.path.join(fit, "loadings.npy")); mu = np.load(os.path.join(fit, "mean.npy"))
    S = np.load(os.path.join(fit, "scores.npy"))
    mu_s, sigma = S.mean(0), S.std(0)
    pp = pd.read_csv(os.path.join(HERE, "..", "global_pool", "pool_processed.csv"))
    pcon = dict(zip(pp.item_id, pp.construct)); pscale = dict(zip(pp.item_id, pp.scale))
    pcores = {int(r["component"]): str(r["items"]).split() for r in
              csv.DictReader(open(os.path.join(HERE, "outputs_pool", "fixedpoint_cores", f"{ENC}_cores.csv")))}
    d = np.load(os.path.join(HRSD, f"hrs_lb2022_{ENC}.npz"), allow_pickle=True)
    hid = [str(x) for x in d["items"]]
    Z = ((np.asarray(d["vectors"], float) - mu) @ W.T - mu_s) / sigma
    it = pd.read_csv(os.path.join(HRSD, "hrs_lb2022_items.csv"))
    hcon = {r.item: hrs_construct(r.construct, r.prompt) for r in it.itertuples()}
    hins = dict(zip(it.item, it.instrument))
    # the relevance gate is the keep rule (D5/D6, _common.keep_columns): an item leans on a
    # component when it anchors it (clears the pool core-item band) or when it is that item's
    # modal component.
    band = pool_bands(ENC, (S - mu_s) / sigma, POOL_IDS)
    kept = set(keep_columns(Z, band))
    modal_of = [int(np.argmax(np.abs(Z[i]))) for i in range(len(hid))]

    cols, Lcon, Rcon = [], {}, {}
    for c in sorted(pcores):
        fams = {FAM[pcon[i]] for i in pcores[c] if pcon[i] in FAM}
        if not fams:
            continue
        if c not in kept:
            continue
        lean = {g for g in range(len(hid)) if modal_of[g] == c or abs(Z[g, c]) >= band[c]}
        hcore = [int(g) for g in fpc(Z[:, c])]
        left = [(poolshort(pscale[i]), FAM.get(pcon[i], "other")) for i in pcores[c]]
        right = [(HSHORT[hins[hid[g]]], HFAM[hcon[hid[g]]], g in lean) for g in hcore]
        cols.append(dict(c=c, fams=sorted(fams), left=left, right=right,
                         pprof=counts_by_construct(pcon[i] for i in pcores[c]),
                         hprof=counts_by_construct(hcon[hid[g]] for g in hcore)))
        for i in pcores[c]:
            Lcon.setdefault(poolshort(pscale[i]), set()).add(pcon[i])
        for g in hcore:
            Rcon.setdefault(HSHORT[hins[hid[g]]], set()).add(hcon[hid[g]])

    H = 100.0
    gap = 0.105 * H

    def names(key):
        out, seen = [], set()
        for x in cols:
            for tup in x[key]:
                if tup[0] not in seen:
                    seen.add(tup[0]); out.append(tup[0])
        return out

    def arrange(cols, rounds=8):
        """Barycentre ordering: place every node at the mean position of the nodes it connects to.

        Sweeping the three columns a few times settles into a layout with far fewer ribbon crossings
        than first-appearance order, and changes nothing about what is drawn.
        """
        Lord, Rord = names("left"), names("right")
        for _ in range(rounds):
            cpos = {x["c"]: i for i, x in enumerate(cols)}
            Lord = sorted(Lord, key=lambda n: np.mean([cpos[x["c"]] for x in cols
                                                       for t in x["left"] if t[0] == n]))
            Rord = sorted(Rord, key=lambda n: np.mean([cpos[x["c"]] for x in cols
                                                       for t in x["right"] if t[0] == n]))
            lp = {n: i / max(1, len(Lord) - 1) for i, n in enumerate(Lord)}
            rp = {n: i / max(1, len(Rord) - 1) for i, n in enumerate(Rord)}
            cols = sorted(cols, key=lambda x: np.mean([lp[t[0]] for t in x["left"]]
                                                      + [rp[t[0]] for t in x["right"]]))
        return cols, Lord, Rord

    cols, Lo, Ro = arrange(cols)

    def side(key, order):
        sz = {ins: sum(1 for x in cols for t in x[key] if t[0] == ins) for ins in order}
        g2 = 0.027 * H; av = H - (len(order) - 1) * g2
        pos = {}; yy = H
        for ins in order:
            h = av * sz[ins] / sum(sz.values()); pos[ins] = (yy - h, yy); yy -= h + g2
        return sz, pos

    Lsz, Lpos = side("left", Lo); Rsz, Rpos = side("right", Ro)

    tot = sum(max(len(x["left"]), len(x["right"])) for x in cols)
    avail = H - (len(cols) - 1) * gap
    Cpos = {}; y = H
    for x in cols:
        h = avail * max(len(x["left"]), len(x["right"])) / tot
        Cpos[x["c"]] = (y - h, y); y -= h + gap

    fig, ax = plt.subplots(# authored at print size, as in fig_aou_twoside_cores.py: 6.3 pt drawn prints at 5.7 pt
    figsize=(7.2, 7.6))
    xL, xC, xR, bw = 0.0, 1.35, 2.7, 0.035
    outL = {i: Lpos[i][1] for i in Lo}; outR = {i: Rpos[i][1] for i in Ro}
    inLC = {x["c"]: Cpos[x["c"]][1] for x in cols}; inRC = {x["c"]: Cpos[x["c"]][1] for x in cols}
    for x in cols:
        c = x["c"]; hC = Cpos[c][1] - Cpos[c][0]
        for ins, fam in sorted(x["left"], key=lambda t: (t[1] == "other", t[1], t[0])):
            h = hC / len(x["left"]); hl = (Lpos[ins][1] - Lpos[ins][0]) / Lsz[ins]
            ribbon(ax, xL + bw, outL[ins], outL[ins] - hl, xC - bw, inLC[c], inLC[c] - h,
                   FCOL[fam], 0.62)
            outL[ins] -= hl; inLC[c] -= h
        for ins, fam, lean in sorted(x["right"], key=lambda t: (not t[2], t[1], t[0])):
            h = hC / len(x["right"]); hr = (Rpos[ins][1] - Rpos[ins][0]) / Rsz[ins]
            ribbon(ax, xC + bw, inRC[c], inRC[c] - h, xR - bw, outR[ins], outR[ins] - hr,
                   FCOL[fam], 0.62 if lean else 0.30, None if lean else "////")
            outR[ins] -= hr; inRC[c] -= h

    def context(cons, n=1):
        cc = sorted(short(x) for x in cons)
        return ", ".join(cc[:n]) + (f", +{len(cc) - n}" if len(cc) > n else "")

    def proftext(prof, n=2):
        head = ", ".join(f"{k} ({v})" for k, v in list(prof.items())[:n])
        return head + (f", +{len(prof) - n} more" if len(prof) > n else "")

    # an instrument and its documented construct are ONE label in two lines: the gap inside the
    # pair must stay visibly smaller than the gap between pairs, or the construct reads as the
    # neighbour's (the same defect fixed in fig_aou_twoside_cores.py on 2026-08-31)
    for ins in Lo:
        lo, hi = Lpos[ins]; mid = (lo + hi) / 2
        ax.add_patch(Rectangle((xL - bw, lo), 2 * bw, hi - lo, fc="#6C86A0", ec="none", zorder=3))
        ax.text(xL - 2.4 * bw, mid + 0.0012 * H, ins, ha="right", va="bottom", fontsize=6.4, color=INK)
        ax.text(xL - 2.4 * bw, mid - 0.0012 * H, context(Lcon[ins]), ha="right", va="top",
                fontsize=6.4, color="#5A5A5A")
    for ins in Ro:
        lo, hi = Rpos[ins]; mid = (lo + hi) / 2
        ax.add_patch(Rectangle((xR - bw, lo), 2 * bw, hi - lo, fc="#56657E", ec="none", zorder=3))
        ax.text(xR + 2.4 * bw, mid + 0.0012 * H, ins, ha="left", va="bottom", fontsize=6.4, color=INK)
        ax.text(xR + 2.4 * bw, mid - 0.0012 * H, context(Rcon[ins]), ha="left", va="top",
                fontsize=6.4, color="#5A5A5A")
    for x in cols:
        c = x["c"]; lo, hi = Cpos[c]
        ax.add_patch(Rectangle((xC - bw, lo), 2 * bw, hi - lo, fc="#0072B2", ec="none", zorder=3))
        ax.text(xC, hi + 0.030 * H, f"C{c}", ha="center", va="bottom", fontsize=8.0,
                color=INK, fontweight="bold")
        ax.text(xC, hi + 0.017 * H, f"pool: {proftext(x['pprof'])}", ha="center", va="bottom",
                fontsize=6.3, color=INK)
        ax.text(xC, hi + 0.004 * H, f"HRS: {proftext(x['hprof'])}", ha="center", va="bottom",
                fontsize=6.3, color=INK)

    ax.text(xL - 2.4 * bw, H + 0.055 * H, "pool instrument", ha="right", va="bottom",
            fontsize=7.6, fontweight="bold", color=INK, linespacing=1.3)
    ax.text(xC, H + 0.055 * H, "component, with the construct profile each side gives it", ha="center", va="bottom",
            fontsize=7.6, fontweight="bold", color=INK)
    ax.text(xR + 2.4 * bw, H + 0.055 * H, "HRS instrument", ha="left", va="bottom",
            fontsize=7.6, fontweight="bold", color=INK, linespacing=1.3)
    leg = [Patch(fc=FCOL[f], ec="none", alpha=0.62, label=FLAB[f]) for f in
           ["EDS", "DMS", "LON", "ISO", "PSS", "NBD", "NBC", "other"]]
    leg.append(Patch(fc="#888888", ec="white", hatch="////", alpha=0.30,
                     label="cores the component but does not lean on it"))
    ax.legend(handles=leg, loc="lower center", bbox_to_anchor=(0.5, -0.075), ncol=3, frameon=False,
              fontsize=6.8, handlelength=1.4, handleheight=1.0, columnspacing=1.4)
    ax.set_xlim(-1.40, 4.40); ax.set_ylim(-0.02 * H, H + 0.20 * H); ax.axis("off")
    fig.subplots_adjust(left=0.005, right=0.995, top=0.985, bottom=0.075)
    sfx = "" if ENC == EXEMPLAR else f"_{ENC}"
    out = os.path.join(OUT, f"fig_hrs_twoside_cores{sfx}.png")
    fig.savefig(out, dpi=300, facecolor="white"); plt.close(fig)
    print("wrote", out, f"| {len(cols)} components")
    for x in cols:
        agree = {f for _, f, l in x["right"] if l} & set(x["fams"])
        print(f"  C{x['c']:<3d} pool {'+'.join(x['fams']):8s} HRS-leaners "
              f"{sorted({f for _, f, l in x['right'] if l})}  -> {'AGREE' if agree else 'no'}")
    return cols


if __name__ == "__main__":
    for e in (ENCS if "--all" in sys.argv else [EXEMPLAR]):
        main(e)
```
