## Supplemental code for "Scoring respondents on a semantic frame of reference identifies a group that is burdened across many questionnaires and reaches care less easily": fig_pool_ribbon.py

```
#!/usr/bin/env python
"""Global-pool ribbon (portability figure, NHB). Adapts fig_recarve.py to the POOL, but
item->component is FIXED-POINT PR CORE membership (outputs_pool/fixedpoint_cores), NOT argmax.

Tripartite alluvial:  Instrument -> Construct (meaning) -> Pool component (gte-large exemplar).
Featured = cross-instrument MERGE families: different independent instruments whose items share
one component core. Cores are an overlapping cover, so an item can feed >1 component; items in no
core do not appear.

Grey  = item flow instrument->construct (fan-in).
Blue  = construct->component (the cross-instrument merge into a shared component).

Outputs examples/global_pool/figures/fig_pool_ribbon.png (+ a preview copy).
Run: /data_gpu5/ROSS_Proj/env/bin/python fig_pool_ribbon.py
"""
import os, re
import numpy as np, pandas as pd
import matplotlib; matplotlib.use("Agg")
import matplotlib.pyplot as plt
from matplotlib.path import Path
from matplotlib.patches import PathPatch, Patch

HERE = os.path.dirname(os.path.abspath(__file__))
REPO = os.path.abspath(os.path.join(HERE, "..", ".."))
ENC = "gte-large-en-v1.5"
CORES = os.path.join(HERE, "outputs_pool", "fixedpoint_cores", f"{ENC}_cores.csv")
POOL = os.path.join(HERE, "..", "global_pool", "pool_processed.csv")
OUTDIR = os.path.join(REPO, "examples", "global_pool", "figures")
os.makedirs(OUTDIR, exist_ok=True)

GREY, GREY_E, MERGE_C, INK = "#B8C4D0", "#8798A8", "#0072B2", "#222222"

# featured merge components (data-verified: >=2 independent instruments share the core)
FOCUS = ["C0", "C7", "C2", "C8", "C9"]
COMP_LABEL = {"C0": "Depression", "C7": "Anxiety", "C2": "Sleep",
              "C8": "Social support", "C9": "Social isolation"}
# keep only same-family constructs per component (drops cross-family single-item strays)
ALLOWED = {"C0": {"Depression"},
           "C7": {"Anxiety", "Panic disorder", "Social phobia", "Agoraphobia"},
           "C2": {"Sleep disturbance", "Sleep-related impairment", "Insomnia"},
           "C8": {"Social support", "Emotional support"},
           "C9": {"Social isolation", "Loneliness"}}


NAMEMAP = {"Three-Item UCLA Loneliness Scale": "UCLA-3",
           "De Jong Gierveld Loneliness Scale (6-item)": "DJG-6"}


def shortscale(s):
    if s in NAMEMAP:
        return NAMEMAP[s]
    m = re.findall(r"\(([^)]+)\)", s)
    if m:
        return {"6-item Short Form": "USDA-6", "SF 6a": "PROMIS-Support", "12-item": "ISEL-12",
                "50-item": "IPIP-50"}.get(m[-1], m[-1])
    return (s.replace("PROMIS Item Bank v1.0 - Emotional Distress - ", "PROMIS-")
             .replace("PROMIS Item Bank v2.0 - Social Isolation", "PROMIS-Isolation")
             .replace("PROMIS Item Bank v2.0 - Emotional Support", "PROMIS-Support")
             .replace("PROMIS Item Bank v1.0 - Sleep Related Impairment", "PROMIS-SleepImpair")
             .replace("PROMIS Item Bank v1.0 - Sleep Disturbance", "PROMIS-Sleep")
             .replace("PROMIS Item Bank v1.1 - Anger", "PROMIS-Anger")
             .replace("Center for Epidemiologic Studies Depression Scale", "CES-D")
             .replace("Multidimensional Scale of Perceived Social Support", "MSPSS"))[:20]


def ribbon(ax, x0, y0a, y0b, x1, y1a, y1b, color, alpha, z=1):
    cx = (x0 + x1) / 2.0
    verts = [(x0, y0a), (cx, y0a), (cx, y1a), (x1, y1a), (x1, y1b),
             (cx, y1b), (cx, y0b), (x0, y0b), (x0, y0a)]
    codes = [Path.MOVETO, Path.CURVE4, Path.CURVE4, Path.CURVE4, Path.LINETO,
             Path.CURVE4, Path.CURVE4, Path.CURVE4, Path.CLOSEPOLY]
    ax.add_patch(PathPatch(Path(verts, codes), facecolor=color, edgecolor="none",
                           lw=0.4, alpha=alpha, zorder=z))


def main():
    pp = pd.read_csv(POOL)
    scale = dict(zip(pp.item_id, pp.scale)); con = dict(zip(pp.item_id, pp.construct))
    cores = pd.read_csv(CORES)
    rows = []
    for _, r in cores.iterrows():
        comp = f"C{r['component']}"
        if comp not in FOCUS:
            continue
        for it in str(r["items"]).split():
            c = con.get(it, "?")
            if c not in ALLOWED[comp]:
                continue
            rows.append(dict(item=it, comp=comp, instr=shortscale(scale.get(it, "?")), construct=c))
    df = pd.DataFrame(rows)

    ic = df.groupby(["instr", "construct"]).size().reset_index(name="n")
    cc = df.groupby(["construct", "comp"]).size().reset_index(name="n")

    # ---- orders (by featured-component order, first-appearance) ----
    comp_order = [c for c in FOCUS if c in df.comp.values]
    constr_order, instr_order = [], []
    for comp in comp_order:
        sub = df[df.comp == comp]
        for c in sub.construct.value_counts().index:
            if c not in constr_order:
                constr_order.append(c)
    for c in constr_order:
        sub = df[df.construct == c]
        for i in sub.instr.value_counts().index:
            if i not in instr_order:
                instr_order.append(i)

    instr_sz = {i: int((df.instr == i).sum()) for i in instr_order}
    constr_sz = {c: int((df.construct == c).sum()) for c in constr_order}
    comp_sz = {c: int((df.comp == c).sum()) for c in comp_order}

    H = 45.0
    def layout(order, sizes, gap_frac=0.05):
        tot = sum(sizes[o] for o in order); gap = gap_frac * H
        avail = H - (len(order) - 1) * gap; pos = {}; y = H
        for o in order:
            h = avail * sizes[o] / tot; pos[o] = (y - h, y); y -= h + gap
        return pos
    Lpos, Mpos, Rpos = layout(instr_order, instr_sz), layout(constr_order, constr_sz), layout(comp_order, comp_sz)
    x_i, x_c, x_p = 0.0, 1.2, 2.55; bw = 0.05

    fig, ax = plt.subplots(figsize=(10.5, 6.6))

    out_cur = {i: Lpos[i][1] for i in instr_order}; in_cur = {c: Mpos[c][1] for c in constr_order}
    for c in constr_order:
        for i in instr_order:
            row = ic[(ic.instr == i) & (ic.construct == c)]
            if row.empty: continue
            n = int(row.n.iloc[0])
            hL = (Lpos[i][1] - Lpos[i][0]) * n / instr_sz[i]
            hM = (Mpos[c][1] - Mpos[c][0]) * n / constr_sz[c]
            ribbon(ax, x_i + bw, out_cur[i], out_cur[i] - hL, x_c - bw, in_cur[c], in_cur[c] - hM, GREY, 0.55)
            out_cur[i] -= hL; in_cur[c] -= hM

    outM = {c: Mpos[c][1] for c in constr_order}; inR = {c: Rpos[c][1] for c in comp_order}
    for comp in comp_order:
        for c in constr_order:
            row = cc[(cc.construct == c) & (cc.comp == comp)]
            if row.empty: continue
            n = int(row.n.iloc[0])
            hM = (Mpos[c][1] - Mpos[c][0]) * n / constr_sz[c]
            hR = (Rpos[comp][1] - Rpos[comp][0]) * n / comp_sz[comp]
            ribbon(ax, x_c + bw, outM[c], outM[c] - hM, x_p - bw, inR[comp], inR[comp] - hR, MERGE_C, 0.45, z=2)
            outM[c] -= hM; inR[comp] -= hR

    # nodes
    for i in instr_order:
        b, t = Lpos[i]
        ax.add_patch(plt.Rectangle((x_i - bw, b), 2 * bw, t - b, facecolor=INK, edgecolor="white", lw=0.8, zorder=5))
        cy = (b + t) / 2
        ax.text(x_i - bw - 0.06, cy, i, ha="right", va="center", fontsize=10.5, fontweight="bold", color=INK)
    for c in constr_order:
        b, t = Mpos[c]
        ax.add_patch(plt.Rectangle((x_c - bw, b), 2 * bw, t - b, facecolor=GREY_E, edgecolor="white", lw=0.8, zorder=5))
        ax.text(x_c, t + 0.55, c, ha="center", va="bottom", fontsize=12.5, color=INK)
    for comp in comp_order:
        b, t = Rpos[comp]
        ax.add_patch(plt.Rectangle((x_p - bw, b), 2 * bw, t - b, facecolor=MERGE_C, edgecolor="white", lw=0.8, zorder=5))
        ax.text(x_p + bw + 0.05, (b + t) / 2 + 0.6, comp, ha="left", va="center", fontsize=11.8, fontweight="bold", color=MERGE_C)
        ax.text(x_p + bw + 0.05, (b + t) / 2 - 0.9, COMP_LABEL.get(comp, ""), ha="left", va="center", fontsize=11.8, color=GREY_E)

    ax.text(x_i, H + 3.2, "Instrument", ha="center", fontsize=13.0, fontweight="bold")
    ax.text(x_c, H + 3.2, "Construct", ha="center", fontsize=13.0, fontweight="bold")
    ax.text(x_p, H + 3.2, "Pool component", ha="center", fontsize=13.0, fontweight="bold")
    leg = [Patch(facecolor=MERGE_C, alpha=0.6, label="Cross-instrument merge (shared component core)"),
           Patch(facecolor=GREY, label="Item flow (instrument → construct)")]
    ax.legend(handles=leg, loc="lower center", bbox_to_anchor=(0.5, -0.055), ncol=1, frameon=False, fontsize=10.5, handlelength=1.4)
    ax.set_xlim(-1.05, x_p + 1.35); ax.set_ylim(-2, H + 4.3); ax.axis("off")
    plt.subplots_adjust(left=0.03, right=0.97, top=0.95, bottom=0.08)

    out = os.path.join(OUTDIR, "fig_pool_ribbon.png")
    fig.savefig(out, dpi=200, bbox_inches="tight", facecolor="white")
    # editable vector twin of the same panel: text stays as text (svg.fonttype "none"), so the
    # labels can be retyped in Inkscape / Illustrator / PowerPoint rather than being outlines
    import matplotlib as _mpl
    with _mpl.rc_context({"svg.fonttype": "none"}):
        fig.savefig(os.path.join(OUTDIR, "fig_pool_ribbon.svg"), bbox_inches="tight",
                    facecolor="white")
    print("wrote", out, "| instruments:", len(instr_order), "constructs:", len(constr_order), "comps:", comp_order)


if __name__ == "__main__":
    main()
```
