## Supplemental code for "Scoring respondents on a semantic frame of reference identifies a group that is burdened across many questionnaires and reaches care less easily": fig_portability.py

```
#!/usr/bin/env python
"""Supplementary figure: a response-based structure is sample-dependent, the text-derived one is not.

Loading stability (mean matched |corr| between two disjoint subsamples of size n) of the two
response-based structures a psychometrician would estimate from the same responses, PCA and factor
analysis on the 151 items, against the text-derived basis, which comes from item text and is identical
at every n including n = 0.

READS SAVED DATA. The sweep is run on the Workbench by `all_of_us_codes/portability_sweep.py`, which
writes `portability_sweep.csv`; copy it next to this script or pass --sweep. Earlier versions of this
figure hardcoded values transcribed from a Workbench run with no saved artefact, which is how the
manuscript came to quote 0.29 as "a few hundred respondents" when it is the value at one hundred.

Out: manuscript_nhb/figures/fig_portability.png
Run: CUDA_VISIBLE_DEVICES="" /data_gpu5/ROSS_Proj/env/bin/python fig_portability.py --sweep <csv>
"""
import os
import numpy as np
import matplotlib
matplotlib.use("Agg")
import matplotlib.pyplot as plt

HERE = os.path.dirname(os.path.abspath(__file__))
REPO = os.path.abspath(os.path.join(HERE, "..", ".."))

import argparse
import pandas as pd

ap = argparse.ArgumentParser()
ap.add_argument("--sweep", default=os.path.join(HERE, "portability_sweep.csv"),
                help="CSV from all_of_us_codes/portability_sweep.py (method,n,repeat,matched_abs_corr)")
args = ap.parse_args()

if not os.path.exists(args.sweep):
    raise SystemExit(
        f"missing {args.sweep}\n"
        "Run all_of_us_codes/portability_sweep.py on the Workbench and copy the CSV here.\n"
        "This generator no longer carries transcribed values.")

sweep = pd.read_csv(args.sweep).groupby(["method", "n"])["matched_abs_corr"].mean().unstack(0)
n = sweep.index.values
pca = sweep["pca"].values
fa = sweep["fa"].values

PCA_C, FA_C, ACIDD_C = "#0072B2", "#56B4E9", "#D55E00"

fig, ax = plt.subplots(figsize=(5.2, 3.9))
ax.axhline(1.0, color=ACIDD_C, lw=2.4, ls="--", zorder=2)
ax.text(11000, 1.005, "ACIDD (item text): 1.0 at every $n$, defined at $n=0$",
        ha="right", va="bottom", fontsize=9.5, color=ACIDD_C, fontweight="bold")
ax.plot(n, fa, "-o", color=FA_C, lw=1.8, ms=5, label="Factor analysis (responses)")
ax.plot(n, pca, "-s", color=PCA_C, lw=1.8, ms=5, label="PCA (responses)")

ax.set_xscale("log")
ax.set_xticks(n)
ax.set_xticklabels([f"{x:,}" for x in n], fontsize=8.5, rotation=0)
ax.set_xlabel("responses per subsample ($n$)", fontsize=10.5)
ax.set_ylabel("structure stability\n(matched loading $|r|$ across subsamples)", fontsize=10.5)
ax.set_ylim(0.2, 1.06)
ax.tick_params(labelsize=9)
ax.legend(loc="lower right", frameon=False, fontsize=9.5)
ax.spines[["top", "right"]].set_visible(False)
ax.annotate("response-based structure\nnever reaches 1 and\nneeds thousands of responses",
            xy=(2000, 0.68), xytext=(180, 0.86), fontsize=8.3, color="#444444",
            arrowprops=dict(arrowstyle="->", color="#888888", lw=0.8))

plt.tight_layout()
out1 = os.path.join(REPO, "manuscript_nhb", "figures", "fig_portability.png")
out2 = os.path.join(REPO, "figure_preview.png")
fig.savefig(out1, dpi=300, bbox_inches="tight", facecolor="white")
fig.savefig(out2, dpi=150, bbox_inches="tight", facecolor="white")
print("wrote", out1)
```
