## Supplemental code for "Scoring respondents on a semantic frame of reference identifies a group that is burdened across many questionnaires and reaches care less easily": fig_xenc.py

```
#!/usr/bin/env python
"""Figures: the pool components reproduce across encoders, read from both sides.

MAIN (fig_xenc.png), the two findings:
(a) Component side: anchoring on the exemplar encoder, how many of the other five encoders each
    component recurs in, split by how many distinct constructs its core spans.
(b) Item side: agreement between encoders of the construct profiles of an item's core components,
    by the number of core components the item has, against the permutation chance level.

SUPPLEMENT (figS_xenc_null.png), the calibration behind the threshold:
    matched-core Jaccard for every matched component pair across the fifteen encoder pairs, against
    a null that permutes item identities before matching, with tau (the null's 95th percentile).

Matching is label-free (Hungarian assignment on core-item Jaccard). Construct counts are read
afterwards and play no part in it. A component is interpreted by the set of constructs of its core items,
its CONSTRUCT PROFILE (DEFINITIONS.md D6-A), so panel (a)'s x axis is the size of that profile
and panel (b) compares profiles.

Reads saved artefacts only - no result, count or label is hardcoded:
    outputs_pool/fixedpoint_cores/<enc>_cores.csv     core items per component, six encoders
    outputs_pool/fits/<enc>__k*_ls1.0/scores.npy      the pool projection, for the item side
    ../global_pool/pool_processed.csv                 documented construct per item

Out: manuscript_nhb/figures/fig_xenc.png and manuscript_nhb/figures/figS_xenc_null.png
Run: CUDA_VISIBLE_DEVICES="" /data_gpu5/ROSS_Proj/env/bin/python fig_xenc.py
"""
import os, glob, csv
from itertools import combinations
import numpy as np
import pandas as pd
from scipy.optimize import linear_sum_assignment
import matplotlib
matplotlib.use("Agg")
import matplotlib.pyplot as plt
from matplotlib.patches import Patch
from pool_card_labels import short          # shared with fig1_pool.py

HERE = os.path.dirname(os.path.abspath(__file__))
REPO = os.path.abspath(os.path.join(HERE, "..", ".."))
GP = os.path.join(HERE, "..", "global_pool")
OUT = os.path.join(REPO, "manuscript_nhb", "figures")

ANCHOR = "gte-large-en-v1.5"
NPERM_MATCH = 5            # permutations per encoder pair for the matching null
NPERM_ITEM = 100           # permutations for the item-side chance level
BLUE, ORANGE, GREY, INK = "#0072B2", "#D55E00", "#BFC6CD", "#222222"
# all six / four or five / fewer. Separable on TWO channels: a wide lightness ramp (L* ~20, ~57, ~93,
# so the three stay distinct in greyscale) and a hatch texture, rather than hue alone.
SHADES = ["#08306B", "#6BAED6", "#EFF6FC"]
HATCH = ["", "///", "xxx"]
HEDGE = ["#08306B", "#FFFFFF", "#2B4C6F"]


def pr(v):
    s2 = float((v ** 2).sum()); s4 = float((v ** 4).sum())
    return s2 * s2 / s4 if s4 > 0 else 0.0


def fpc(v):
    idx = np.arange(len(v))
    while True:
        n = max(1, min(int(round(pr(v[idx]))), len(idx)))
        top = np.sort(idx[np.argsort(-np.abs(v[idx]))[:n]])
        if len(top) == len(idx) and (top == idx).all():
            return top
        idx = top


def jac(a, b):
    return len(a & b) / len(a | b) if (a | b) else 0.0


def match(A, B):
    J = np.array([[jac(a, b) for b in B] for a in A])
    ri, ci = linear_sum_assignment(1.0 - J)
    return {int(i): float(J[i, j]) for i, j in zip(ri, ci)}


def counts_by_construct(labels):
    """Documented constructs with their item counts, most common first.

    Ties are broken alphabetically. `value_counts` alone breaks them in hash-table order, which
    moves with Python's string-hash randomization, so two runs of the same generator produced
    cards and CSV rows that listed equal-count constructs in a different order. No count or
    threshold ever changed, but the figures were not byte-reproducible.
    """
    return (pd.Series(list(labels)).value_counts()
            .sort_index().sort_values(ascending=False, kind="mergesort"))


def draw_reproduction_cards(anchor_cores, rep, con):
    """The 55 components as cards, shaded by how many encoders each reproduces in.

    Same grid and same cards as main-text Figure 1b, so one display carries over to the other, but
    shaded by reproduction instead of by the number of constructs in the core, using the identical
    three-way encoding as main-text Figure 2a (lightness plus hatch, never hue alone).

    SIX columns, matching Figure 1b, and drawn 7.2 in wide for a 6.5 in \\textwidth. This figure is
    the supplement's only display of all 55 cards, so it is placed at full \\textwidth: at five
    columns and 0.86\\textwidth its construct labels printed at 4.7 pt, under NHB's 5 pt floor.
    """
    from matplotlib.patches import FancyBboxPatch, Rectangle
    NCOL, NROW = 6, 10
    # dark card carries white text, the two light ones dark text
    CARDINK = [("white", "white", "#DCE6F2"), ("0.10", "0.12", "0.30"), ("0.10", "0.12", "0.38")]
    LABELS = ["all six encoders", "four or five", "fewer"]

    def level(r):
        return 0 if r == 5 else (1 if r >= 3 else 2)

    plt.rcParams["hatch.linewidth"] = 0.6
    fig, ax = plt.subplots(figsize=(7.2, 7.5))
    # the band above the first row is pure white space: the legend is anchored OUTSIDE the axes
    # (bbox_to_anchor y=1.004 below), so anything reserved here only pushes the cards down
    ax.set_xlim(0, NCOL); ax.set_ylim(-0.10, NROW); ax.axis("off"); ax.invert_yaxis()
    for i, core in enumerate(anchor_cores):
        col, row = i % NCOL, i // NCOL
        k = level(rep[i])
        ax.add_patch(FancyBboxPatch((col + 0.04, row + 0.07), 0.92, 0.86,
                                    boxstyle="round,pad=0.006,rounding_size=0.04",
                                    fc=SHADES[k], ec=HEDGE[k], lw=1.6 if k == 0 else 0.9))
        # the texture lives in a strip down the left edge, not across the card, so that hatching
        # never crosses the construct names
        if HATCH[k]:
            ax.add_patch(Rectangle((col + 0.055, row + 0.10), 0.085, 0.80, fc=SHADES[k],
                                   hatch=HATCH[k], ec=HEDGE[k], lw=0.5, zorder=3))
        hdr_c, con_c, num_c = CARDINK[k]
        prof = counts_by_construct(con[h] for h in core)
        ax.text(col + 0.155, row + 0.24, f"C{i} ({len(core)})", fontsize=7, fontweight="bold",
                color=hdr_c, zorder=4)
        y = row + 0.45
        for name, n in list(prof.items())[:3]:
            ax.text(col + 0.155, y, short(name, 13), fontsize=5.6, va="center", color=con_c, zorder=4)
            ax.text(col + 0.96, y, f"{n}", fontsize=5.6, va="center", ha="right", color=num_c, zorder=4)
            y += 0.185
        if len(prof) > 3:
            # this line falls below the card, on white, so it takes ink of its own rather than the
            # card's, which would be invisible for the dark cards
            ax.text(col + 0.165, y, f"+{len(prof) - 3} more", fontsize=5.6, va="center",
                    color="#555555", style="italic", zorder=4)
    leg = [Patch(fc=SHADES[k], ec=HEDGE[k], hatch=HATCH[k], lw=(1.6 if k == 0 else 0.9), label=lab)
           for k, lab in enumerate(LABELS)]
    ax.legend(handles=leg, loc="lower center", bbox_to_anchor=(0.5, 1.004), ncol=3, frameon=False,
              fontsize=7.4, handlelength=1.4, handleheight=1.2, columnspacing=1.4,
              title="reproduces in", title_fontsize=7.8)
    out = os.path.join(OUT, "figS_xenc_cards.png")
    fig.savefig(out, dpi=400, bbox_inches="tight", facecolor="white")
    plt.close(fig)
    n_top = sum(1 for r in rep if level(r) == 0)
    print(f"wrote {out} | {n_top} of {len(rep)} cards at all six encoders")


def write_reproduction_csv(anchor_cores, rep, con):
    """Supplementary Data 5: every anchor component, how widely it reproduces, and what its core carries.

    This was Supplementary Table S8, which listed only the components reproducing in at least four of
    the six encoders. The file carries all of them, so the threshold is visible rather than applied
    before the reader sees the data.
    """
    rows = []
    for i, core in enumerate(anchor_cores):
        prof = counts_by_construct(con[h] for h in core)
        rows.append({
            "component": f"C{i}",
            "core_items": len(core),
            "reproduces_in_encoders": rep[i] + 1,      # rep counts the five OTHER encoders
            "documented_constructs": ", ".join(f"{k} ({v})" for k, v in prof.items()),
            # cores are sets, so sort or the column order moves with the hash seed
            "core_item_ids": " ".join(sorted(core)),
        })
    df = pd.DataFrame(rows).sort_values(["reproduces_in_encoders", "component"],
                                        ascending=[False, True])
    path = os.path.join(REPO, "manuscript_nhb", "supplementary_data", "component_reproduction.csv")
    df.to_csv(path, index=False)
    print(f"wrote {path} | {len(df)} components, "
          f"{int((df.reproduces_in_encoders >= 4).sum())} reproducing in >=4 of 6")


def main():
    pp = pd.read_csv(os.path.join(GP, "pool_processed.csv"))
    con = dict(zip(pp.item_id, pp.construct))
    ids = list(pp.item_id)

    cores = {}
    for f in sorted(glob.glob(os.path.join(HERE, "outputs_pool", "fixedpoint_cores", "*_cores.csv"))):
        enc = os.path.basename(f).replace("_cores.csv", "")
        cores[enc] = [set(str(r["items"]).split()) for r in csv.DictReader(open(f))]
    ENCS = list(cores)
    universe = sorted(set().union(*[c for e in ENCS for c in cores[e]]))

    # ---------- (a) matched-core Jaccard, real and null ----------
    pairs = list(combinations(ENCS, 2))
    real = np.array([v for A, B in pairs for v in match(cores[A], cores[B]).values()])
    rng = np.random.RandomState(0)
    null = []
    for A, B in pairs:
        for _ in range(NPERM_MATCH):
            perm = dict(zip(universe, rng.permutation(universe)))
            null += list(match(cores[A], [{perm[x] for x in c} for c in cores[B]]).values())
    null = np.array(null)
    tau = float(np.percentile(null, 95))
    print(f"matched Jaccard  real median {np.median(real):.3f}   null median {np.median(null):.3f}"
          f"   tau {tau:.3f}")

    # ---------- (b) recurrence of each anchor component, by core profile size ----------
    others = [e for e in ENCS if e != ANCHOR]
    Jby = {E: match(cores[ANCHOR], cores[E]) for E in others}
    rep = np.array([sum(1 for E in others if Jby[E].get(i, 0.0) >= tau)
                    for i in range(len(cores[ANCHOR]))])
    nprof = np.array([len({con[h] for h in c}) for c in cores[ANCHOR]])
    bins = [(nprof == 1, "1"), (nprof == 2, "2"), (nprof == 3, "3"), (nprof == 4, "4"),
            (nprof >= 5, "5+")]
    print(f"components {len(rep)}   all six {int((rep == 5).sum())}   "
          f"at least four {int((rep >= 3).sum())}")
    # component-side breakdown, the numbers Supplementary Section S16 tabulates and the
    # one aggregate the main text quotes
    print(f"{'constructs in core':20s} {'components':>11s} {'>=4 of 6':>9s} {'all 6':>7s}")
    for m, lab in bins:
        print(f"{lab:20s} {int(m.sum()):11d} {int((m & (rep >= 3)).sum()):9d} "
              f"{int((m & (rep == 5)).sum()):7d}")
    clean = nprof <= 2
    print(f"{'1 or 2 (aggregate)':20s} {int(clean.sum()):11d} "
          f"{int((clean & (rep >= 3)).sum()):9d} {int((clean & (rep == 5)).sum()):7d}")

    # which constructs the reproducing components actually carry. The main text says the reproducible
    # components span the socially and clinically central constructs, and this is the table behind it.
    write_reproduction_csv(cores[ANCHOR], rep, con)
    draw_reproduction_cards(cores[ANCHOR], rep, con)

    # ---------- (c) item side: construct profiles across encoders ----------
    sets_by_enc, sizes = [], []
    for enc in ENCS:
        fit = glob.glob(os.path.join(HERE, "outputs_pool", "fits", f"{enc}__k*_ls1.0"))[0]
        Z = np.load(os.path.join(fit, "scores.npy"))
        Zt = Z / Z.std(0)
        prof = [frozenset(con[h] for h in c) for c in cores[enc]]
        s, sz = [], []
        for j in range(len(ids)):
            F = [int(c) for c in fpc(Zt[j])]
            s.append(frozenset().union(*[prof[c] for c in F])); sz.append(len(F))
        sets_by_enc.append(s); sizes.append(sz)
    mean_size = np.array(sizes).mean(0)

    def pairwise(sets, perm=False):
        n = len(sets[0]); vals = []
        for A, B in combinations(range(len(sets)), 2):
            pb = rng.permutation(n) if perm else np.arange(n)
            vals.append(np.mean([jac(sets[A][i], sets[B][pb[i]]) for i in range(n)]))
        return float(np.mean(vals))

    per_item = np.array([np.mean([jac(sets_by_enc[A][i], sets_by_enc[B][i])
                                  for A, B in combinations(range(len(ENCS)), 2)])
                         for i in range(len(ids))])
    chance = np.mean([pairwise(sets_by_enc, perm=True) for _ in range(NPERM_ITEM)])
    ibins = [(0, 1.5, "1"), (1.5, 2.5, "2"), (2.5, 4.5, "3-4"), (4.5, 99, "5+")]
    print(f"item side  overall {pairwise(sets_by_enc):.3f}   chance {chance:.3f}")

    # ---------- draw: main figure, the two findings ----------
    fig, (bx, cx) = plt.subplots(1, 2, figsize=(7.1, 2.6),
                                 gridspec_kw=dict(width_ratios=[1.2, 1.0]))
    for a in (bx, cx):
        for sp in ("top", "right"):
            a.spines[sp].set_visible(False)
        a.tick_params(labelsize=6.5, length=2.5, pad=1.5)

    plt.rcParams["hatch.linewidth"] = 0.7
    cats = [(rep == 5), (rep >= 3) & (rep < 5), (rep < 3)]
    bottoms = np.zeros(len(bins))
    for k, (mask, lab) in enumerate(zip(cats, ["all six encoders", "four or five", "fewer"])):
        vals = np.array([int((m & mask).sum()) for m, _ in bins], float)
        bx.bar(np.arange(len(bins)), vals, 0.68, bottom=bottoms, color=SHADES[k], label=lab,
               hatch=HATCH[k], edgecolor=HEDGE[k], linewidth=0.7)
        bottoms += vals
    for i, (m, _) in enumerate(bins):
        bx.text(i, bottoms[i] + 0.4, f"n={int(m.sum())}", ha="center", fontsize=6.4, color=INK)
    bx.set_xticks(range(len(bins))); bx.set_xticklabels([b[1] for b in bins])
    bx.set_xlabel("constructs in the component's core", fontsize=7.5)
    bx.set_ylabel("components", fontsize=7.5)
    bx.set_ylim(0, bottoms.max() + 3.4)
    bx.set_yticks(np.arange(0, int(bottoms.max()) + 1, 5))
    bx.legend(fontsize=6.4, frameon=False, loc="upper left", handlelength=1.1,
              borderaxespad=0.2, labelspacing=0.25)
    bx.set_title("a", loc="left", fontsize=9, fontweight="bold", color=INK)

    vals, labs, ns = [], [], []
    for lo, hi, lab in ibins:
        m = (mean_size >= lo) & (mean_size < hi)
        vals.append(per_item[m].mean()); labs.append(lab); ns.append(int(m.sum()))
    cx.bar(np.arange(len(vals)), vals, 0.68, color=ORANGE)
    for i, (v, n) in enumerate(zip(vals, ns)):
        cx.text(i, v + 0.015, f"n={n}", ha="center", fontsize=6.4, color=INK)
    cx.axhline(chance, color=INK, lw=1.0, ls="--")
    cx.text(len(vals) - 0.45, chance + 0.015, "chance", fontsize=6.4, color=INK, ha="right")
    cx.set_xticks(range(len(vals))); cx.set_xticklabels(labs)
    cx.set_xlabel("core components of an item", fontsize=7.5)
    cx.set_ylabel("agreement between encoders\n(Jaccard of construct profiles)", fontsize=7.5)
    cx.set_ylim(0, max(vals) * 1.18)
    cx.set_title("b", loc="left", fontsize=9, fontweight="bold", color=INK)

    fig.subplots_adjust(left=0.075, right=0.995, top=0.90, bottom=0.20, wspace=0.30)
    out = os.path.join(OUT, "fig_xenc.png")
    fig.savefig(out, dpi=400, facecolor="white")

    # ---------- draw: supplement figure, the threshold calibration ----------
    figS, ax = plt.subplots(figsize=(4.2, 2.6))
    for sp in ("top", "right"):
        ax.spines[sp].set_visible(False)
    ax.tick_params(labelsize=6.5, length=2.5, pad=1.5)
    edges = np.linspace(0, 1, 26)
    ax.hist(null, bins=edges, color=GREY, label="permuted", density=True)
    ax.hist(real, bins=edges, histtype="step", color=BLUE, lw=1.3, label="matched", density=True)
    ax.axvline(tau, color=INK, lw=1.0, ls="--")
    ax.text(tau + 0.02, ax.get_ylim()[1] * 0.93, r"$\tau$", fontsize=7.5, color=INK)
    ax.set_xlabel("core overlap of matched components (Jaccard)", fontsize=7.5)
    ax.set_ylabel("density", fontsize=7.5)
    ax.legend(fontsize=6.6, frameon=False, loc="upper right", handlelength=1.2, borderaxespad=0.2)
    figS.subplots_adjust(left=0.135, right=0.985, top=0.96, bottom=0.20)
    outS = os.path.join(OUT, "figS_xenc_null.png")
    figS.savefig(outS, dpi=400, facecolor="white")
    print("wrote", out, "and", outS)


if __name__ == "__main__":
    main()
```
