## Supplemental code for "Scoring respondents on a semantic frame of reference identifies a group that is burdened across many questionnaires and reaches care less easily": generate_variants.py

````
#!/usr/bin/env python
"""Generate item variants for the §3.3 content-vs-grammar test (Claude API).

The test is a counterfactual recombination of the battery's own content and grammar:

  Cell A  content preserved, FRAME transferred  -- rewrite a seed item to mean the SAME
          thing but in the grammatical style of OTHER battery items. If grouping is
          content-driven, every Cell-A variant should land in the seed's component.

  Cell B  FRAME preserved, content swapped       -- keep the seed item's exact sentence
          structure but ask about OTHER items' topics. If grammar does NOT bind, each
          Cell-B variant should land in the TOPIC-donor's component, not the seed's.

  Cell N  paraphrase null (optional)             -- same meaning AND structure, light
          reword; calibrates how much a variant moves for non-grammatical reasons.

Frames and topics are drawn from the 151 items themselves (donors from a *different*
instrument than the seed), so every generated item stays in the battery's register --
this is the register-shift control. Each generated item carries its provenance and its
INTENDED construct (the item whose component it should fall into), giving labeled ground
truth for the projection-side evaluation (validate_content_vs_grammar.py).

One Claude call per (seed, cell): 151 seeds x {A,B} = 302 calls (+151 with --null).

Run where an ANTHROPIC_API_KEY is set and `anthropic` is installed:
  pip install anthropic
  export ANTHROPIC_API_KEY=...
  python generate_variants.py --model claude-sonnet-4-6 --n-variants 10
Preview the exact prompts without spending anything:
  python generate_variants.py --dry-run
Output: outputs_core151/content_grammar/generated_items.csv
"""
import argparse
import csv
import json
import os
import random
import re
import sys

ITEMS_CSV = "core_items_151.csv"
ENCODING_CSV = "encoding_map_all151.csv"
OUT_DIR = "outputs_core151/content_grammar"

# ---- prompt templates (the scientific instrument; kept explicit for transparency) ----
SYS = ("You rewrite survey questionnaire items with surgical control over wording. "
       "You change ONLY what each task asks you to change and never alter a question's "
       "meaning, severity, timeframe, or specificity unless explicitly told to. "
       "You always respond with strictly valid JSON and nothing else.")

CELL_A = """\
TARGET survey item (id: {seed_id}):
  "{seed_prompt}"

STYLE-EXAMPLE items from the same questionnaire battery (different topics):
{donor_block}

Task: rewrite the TARGET item once for EACH style example, so that the rewrite means
EXACTLY the same thing as the TARGET but mimics the grammatical structure / phrasing of
that style example (e.g. question vs statement, the opening stem, sentence form). Change
ONLY the sentence form. Do NOT change the topic, severity, timeframe, or specificity of
the TARGET. If a style would force a meaning change (e.g. a different timeframe), keep the
TARGET's meaning and approximate the structure.

Respond with JSON: a list of {n} objects {{"style_id": <id>, "rewritten": <text>}}."""

CELL_B = """\
TEMPLATE survey item (id: {seed_id}) -- copy its sentence structure exactly:
  "{seed_prompt}"

TOPIC items (the subject matter to ask about instead):
{donor_block}

Task: for EACH topic item, write a new survey item that keeps the TEMPLATE's exact
grammatical structure / phrasing (same stem, same sentence form) but asks about the TOPIC
item's subject matter instead of the template's. Preserve the template's form; swap in the
topic's content.

Respond with JSON: a list of {n} objects {{"topic_id": <id>, "rewritten": <text>}}."""

CELL_N = """\
TARGET survey item (id: {seed_id}):
  "{seed_prompt}"

Task: reword the TARGET item {n} different ways. Each reword must preserve BOTH its exact
meaning AND its grammatical structure (light synonym-level paraphrase only -- same stem,
same sentence form, same timeframe and specificity).

Respond with JSON: a list of {n} objects {{"i": <index>, "rewritten": <text>}}."""


def load_items():
    prompts, order = {}, []
    with open(ITEMS_CSV) as f:
        for r in csv.DictReader(f):
            prompts[r["item"]] = r["prompt"]
            order.append(r["item"])
    inst = {}
    with open(ENCODING_CSV) as f:
        for r in csv.DictReader(f):
            v = r["instrument"]
            inst[r["item"]] = r["item"] if v == "single item" else v  # single -> own instrument
    return order, prompts, inst


def donor_block(donor_ids, prompts):
    return "\n".join(f'  [{d}] "{prompts[d]}"' for d in donor_ids)


def build_prompt(cell, seed, prompts, donors, n):
    if cell == "N":
        return CELL_N.format(seed_id=seed, seed_prompt=prompts[seed], n=n)
    tmpl = CELL_A if cell == "A" else CELL_B
    return tmpl.format(seed_id=seed, seed_prompt=prompts[seed],
                       donor_block=donor_block(donors, prompts), n=n)


def pick_donors(seed, order, inst, n, rng):
    pool = [it for it in order if inst[it] != inst[seed]]
    rng.shuffle(pool)
    return pool[:n]


def extract_json(text):
    text = re.sub(r"^```(?:json)?|```$", "", text.strip(), flags=re.MULTILINE).strip()
    m = re.search(r"\[.*\]", text, flags=re.DOTALL)
    return json.loads(m.group(0) if m else text)


def call_claude(client, model, temperature, prompt):
    msg = client.messages.create(
        model=model, max_tokens=4096, temperature=temperature, system=SYS,
        messages=[{"role": "user", "content": prompt}])
    return extract_json(msg.content[0].text)


def main():
    ap = argparse.ArgumentParser()
    ap.add_argument("--model", default="claude-sonnet-4-6")
    ap.add_argument("--n-variants", type=int, default=10)
    ap.add_argument("--temperature", type=float, default=0.7)
    ap.add_argument("--seed", type=int, default=0)
    ap.add_argument("--limit", type=int, default=0, help="only first N seeds (testing)")
    ap.add_argument("--null", action="store_true", help="also generate Cell N (paraphrase null)")
    ap.add_argument("--dry-run", action="store_true", help="print sample prompts, no API calls")
    ap.add_argument("--seeds-file", default="", help="file with one seed item per line (subset)")
    ap.add_argument("--emit-prompts", default="", help="write per-(seed,cell) prompts JSONL; no API")
    ap.add_argument("--ingest", default="", help="prompts JSONL to assemble from (with --responses)")
    ap.add_argument("--responses", default="", help="responses JSONL {key, variants:[...]} for --ingest")
    ap.add_argument("--gen-model", default="claude-opus-4-8 (interactive)",
                    help="model label recorded by --ingest")
    ap.add_argument("--out", default=os.path.join(OUT_DIR, "generated_items.csv"))
    args = ap.parse_args()

    order, prompts, inst = load_items()
    seeds = order[: args.limit] if args.limit else order
    if args.seeds_file:
        wanted = {l.strip() for l in open(args.seeds_file) if l.strip()}
        seeds = [s for s in order if s in wanted]
    cells = ["A", "B"] + (["N"] if args.null else [])
    rng = random.Random(args.seed)
    # pre-draw donors per seed so dry-run and real run match (deterministic)
    donors = {s: pick_donors(s, order, inst, args.n_variants, random.Random(args.seed + i))
              for i, s in enumerate(order)}

    if args.dry_run:
        s0 = seeds[0]
        print(f"PLAN: {len(seeds)} seeds x {len(cells)} cells = {len(seeds)*len(cells)} API "
              f"calls; {args.n_variants} variants/call; model={args.model} temp={args.temperature}\n")
        print(f"system:\n  {SYS}\n")
        for c in cells:
            print("=" * 78, f"\nCELL {c}  (seed {s0}, instrument {inst[s0]})\n" + "=" * 78)
            print(build_prompt(c, s0, prompts, donors[s0], args.n_variants), "\n")
        print("(dry run -- no API calls, no file written)")
        return

    # ---- offline mode: emit the exact prompts (for generation by any Claude, no API) ----
    if args.emit_prompts:
        os.makedirs(os.path.dirname(args.emit_prompts) or ".", exist_ok=True)
        with open(args.emit_prompts, "w") as f:
            for s in seeds:
                for c in cells:
                    rec = {"key": f"{s}|{c}", "seed": s, "cell": c, "instrument": inst[s],
                           "n": args.n_variants, "donors": donors[s],
                           "prompt": build_prompt(c, s, prompts, donors[s], args.n_variants)}
                    f.write(json.dumps(rec) + "\n")
        print(f"-> {args.emit_prompts}  ({len(seeds)*len(cells)} prompts; system prompt in SYS)")
        return

    # ---- assemble generated_items.csv from emitted prompts + responses (no API) ----
    if args.ingest:
        meta = {}
        with open(args.ingest) as f:
            for line in f:
                r = json.loads(line); meta[r["key"]] = r
        rows, vid = [], 0
        with open(args.responses) as f:
            for line in f:
                if not line.strip():
                    continue
                resp = json.loads(line); m = meta[resp["key"]]
                seed, c, dlist = m["seed"], m["cell"], m["donors"]
                for k, o in enumerate(resp["variants"]):
                    text = o if isinstance(o, str) else o.get("rewritten", "")
                    donor = dlist[k] if k < len(dlist) else ""
                    intended = donor if c == "B" else seed
                    if c == "N":
                        donor = ""
                    rows.append({
                        "variant_id": f"v{vid:05d}", "cell": c, "seed_item": seed,
                        "seed_prompt": prompts[seed], "donor_item": donor,
                        "intended_item": intended, "rewritten_prompt": text.strip(),
                        "model": args.gen_model, "temperature": "NA", "rng_seed": args.seed})
                    vid += 1
        os.makedirs(os.path.dirname(args.out) or ".", exist_ok=True)
        with open(args.out, "w", newline="") as f:
            w = csv.DictWriter(f, fieldnames=list(rows[0].keys()))
            w.writeheader(); w.writerows(rows)
        print(f"-> {args.out}  ({len(rows)} variants assembled from {args.responses})")
        return

    try:
        import anthropic
    except ImportError:
        sys.exit("anthropic not installed: `pip install anthropic` (and set ANTHROPIC_API_KEY)")
    client = anthropic.Anthropic()

    os.makedirs(OUT_DIR, exist_ok=True)
    rows, vid = [], 0
    for si, seed in enumerate(seeds):
        for c in cells:
            prompt = build_prompt(c, seed, prompts, donors[seed], args.n_variants)
            try:
                out = call_claude(client, args.model, args.temperature, prompt)
            except Exception as e:                                   # noqa: BLE001
                print(f"  [warn] seed {seed} cell {c}: {e}", file=sys.stderr); continue
            dlist = donors[seed]
            for k, o in enumerate(out):
                # intended construct: Cell A/N -> the seed; Cell B -> the topic donor
                if c == "B":
                    donor = o.get("topic_id") or (dlist[k] if k < len(dlist) else "")
                    intended = donor
                else:
                    donor = o.get("style_id", "") if c == "A" else ""
                    intended = seed
                rows.append({
                    "variant_id": f"v{vid:05d}", "cell": c, "seed_item": seed,
                    "seed_prompt": prompts[seed], "donor_item": donor,
                    "intended_item": intended, "rewritten_prompt": o.get("rewritten", "").strip(),
                    "model": args.model, "temperature": args.temperature, "rng_seed": args.seed,
                })
                vid += 1
        print(f"  [{si+1}/{len(seeds)}] {seed}: {len(rows)} variants so far", file=sys.stderr)

    with open(args.out, "w", newline="") as f:
        w = csv.DictWriter(f, fieldnames=list(rows[0].keys()))
        w.writeheader(); w.writerows(rows)
    print(f"-> {args.out}  ({len(rows)} variants from {len(seeds)} seeds)")


if __name__ == "__main__":
    main()
````
