## Supplemental code for "Scoring respondents on a semantic frame of reference identifies a group that is burdened across many questionnaires and reaches care less easily": hrs_kept_components.py

```
#!/usr/bin/env python
"""The components the HRS battery reaches on the fixed pool basis, and how stable that is.

Produces the numbers R2 quotes for HRS coverage and for robustness to phrasing:
  (a) components kept per encoder -- the keep rule of _common.keep_columns: a component is kept when
      an HRS item anchors it (clears its pool core-item band) or when it is an item's modal component
  (b) whether any construct HRS fields is confined to the components it does not reach
  (c) agreement of the keep decision between encoders, over components matched by core-item overlap
      alone, against the agreement expected if the two decisions were independent
  (d) the phrasing check for BOTH batteries: items the pool also asks, split by whether the two
      versions are worded identically, comparing the core components each version reaches on the same
      basis. Byte-identical text must place identically, so that group is the determinism check.

Reads saved artefacts only:
    outputs_pool/fits/<enc>__k*_ls1.0/{loadings,mean}.npy   the pool basis per encoder
    outputs_pool/fixedpoint_cores/<enc>_cores.csv           pool core items per component
    data/pool_items_<enc>.npz                               pool embeddings, for the column scale
    ../randhrsdata/acidd_input/hrs_lb2022_<enc>.npz         HRS item embeddings
    ../randhrsdata/acidd_input/hrs_lb2022_items.csv         HRS instrument and construct per item
    ../global_pool/pool_processed.csv                       documented construct per pool item

Run: CUDA_VISIBLE_DEVICES="" /data_gpu5/ROSS_Proj/env/bin/python hrs_kept_components.py
"""
import csv
import glob
import os
import re
from itertools import combinations

import numpy as np
import pandas as pd
from scipy.optimize import linear_sum_assignment

from _common import keep_columns, pool_bands

HERE = os.path.dirname(os.path.abspath(__file__))
HRSD = os.path.join(HERE, "..", "randhrsdata", "acidd_input")
GP = os.path.join(HERE, "..", "global_pool")
POOL_IDS = pd.read_csv(os.path.join(GP, "pool_processed.csv"))["item_id"].tolist()
ENCS = ["gte-large-en-v1.5", "bge-m3", "bge-large-en-v1.5", "all-mpnet-base-v2",
        "e5-large-v2", "qwen3-embedding-0.6b"]
TAU = 0.154          # the matching threshold calibrated in fig_xenc.py


def pr(v):
    s2 = float((v ** 2).sum()); s4 = float((v ** 4).sum())
    return s2 * s2 / s4 if s4 > 0 else 0.0


def fpc(v):
    idx = np.arange(len(v))
    while True:
        n = max(1, min(int(round(pr(v[idx]))), len(idx)))
        top = np.sort(idx[np.argsort(-np.abs(v[idx]))[:n]])
        if len(top) == len(idx) and (top == idx).all():
            return top
        idx = top


def jac(a, b):
    return len(a & b) / len(a | b) if (a | b) else 0.0


def main():
    pp = pd.read_csv(os.path.join(GP, "pool_processed.csv"))
    pcon = dict(zip(pp.item_id, pp.construct))
    hrs = pd.read_csv(os.path.join(HRSD, "hrs_lb2022_items.csv"))
    # the pool's labels for what HRS fields. HRS documents loneliness as one construct and the pool
    # splits the same content across loneliness and social isolation, so both count as fielded.
    hrs_constructs = {"Loneliness", "Social isolation", "Perceived stress", "Everyday discrimination",
                      "Healthcare discrimination", "Neighborhood disorder", "Neighborhood cohesion"}

    cores, kept = {}, {}
    for enc in ENCS:
        fit = glob.glob(os.path.join(HERE, "outputs_pool", "fits", f"{enc}__k*_ls1.0"))[0]
        W = np.load(os.path.join(fit, "loadings.npy")); mu = np.load(os.path.join(fit, "mean.npy"))
        S = np.load(os.path.join(fit, "scores.npy"))
        Zpool = (S - S.mean(0)) / S.std(0)
        hv = np.load(os.path.join(HRSD, f"hrs_lb2022_{enc}.npz"), allow_pickle=True)["vectors"]
        Z = ((np.asarray(hv, float) - mu) @ W.T - S.mean(0)) / S.std(0)
        cores[enc] = [set(str(r["items"]).split()) for r in csv.DictReader(
            open(os.path.join(HERE, "outputs_pool", "fixedpoint_cores", f"{enc}_cores.csv")))]
        kept[enc] = set(keep_columns(Z, pool_bands(enc, Zpool, POOL_IDS)))

    print(f"{'encoder':24s} {'k':>3s} {'kept':>5s} {'dropped':>8s}  HRS constructs confined to dropped")
    for enc in ENCS:
        drop = [c for c in range(len(cores[enc])) if c not in kept[enc]]
        kc = {pcon[x] for c in kept[enc] for x in cores[enc][c]}
        dc = {pcon[x] for c in drop for x in cores[enc][c]}
        confined = sorted((dc - kc) & hrs_constructs)
        print(f"{enc:24s} {len(cores[enc]):3d} {len(kept[enc]):5d} {len(drop):8d}  "
              f"{confined if confined else 'none'}")

    obs, exp = [], []
    for A, B in combinations(ENCS, 2):
        J = np.array([[jac(a, b) for b in cores[B]] for a in cores[A]])
        ri, ci = linear_sum_assignment(1.0 - J)
        pairs = [(i, j) for i, j in zip(ri, ci) if J[i, j] >= TAU]
        if not pairs:
            continue
        pa = np.mean([i in kept[A] for i, _ in pairs])
        pb = np.mean([j in kept[B] for _, j in pairs])
        obs.append(np.mean([(i in kept[A]) == (j in kept[B]) for i, j in pairs]))
        exp.append(pa * pb + (1 - pa) * (1 - pb))
    o, e = float(np.mean(obs)), float(np.mean(exp))
    print(f"\nkeep decision agrees on {100 * o:.0f}% of matched components, {100 * e:.0f}% expected "
          f"if independent (kappa {(o - e) / (1 - e):.2f}, {len(obs)} encoder pairs)")
    phrasing(hrs)
    phrasing_aou()


def phrasing(hrs, enc="gte-large-en-v1.5"):
    """Items HRS and the pool both ask, placed on the same basis, worded the same or differently.

    The pool embeds its item text with the leading time phrase removed and HRS administers bare
    fragments, so the same question often reaches the basis in two wordings. Matching is on the tail of
    the normalized text, which is what the two versions share.
    """
    def tail(t):
        return re.sub(r"[^a-z]", "", str(t).lower())[-45:]

    def norm(t):
        return re.sub(r"[^a-z ]", "", str(t).lower()).strip()

    def jac(a, b):
        return len(a & b) / len(a | b) if (a | b) else 0.0

    fit = glob.glob(os.path.join(HERE, "outputs_pool", "fits", f"{enc}__k*_ls1.0"))[0]
    W = np.load(os.path.join(fit, "loadings.npy")); mu = np.load(os.path.join(fit, "mean.npy"))
    Zp = np.load(os.path.join(fit, "scores.npy")); sigma = Zp.std(0); Zp = Zp / sigma
    pp = pd.read_csv(os.path.join(GP, "pool_processed.csv")); ids = list(pp.item_id)
    hv = np.load(os.path.join(HRSD, f"hrs_lb2022_{enc}.npz"), allow_pickle=True)
    hid = [str(x) for x in hv["items"]]
    Zh = ((np.asarray(hv["vectors"], float) - mu) @ W.T) / sigma
    htxt = dict(zip(hrs.item, hrs.prompt))
    pool_by_tail = {tail(t): (i, t) for i, t in zip(ids, pp.prompt_clean)}

    same, diff, shape = [], [], []
    for i, it in enumerate(hid):
        k = tail(htxt[it])
        m = next((pool_by_tail[q] for q in pool_by_tail if q.endswith(k) or k.endswith(q)), None)
        if m is None:
            continue
        A = {int(c) for c in fpc(Zh[i])}
        B = {int(c) for c in fpc(Zp[ids.index(m[0])])}
        v = jac(A, B)
        (same if norm(htxt[it]) == norm(m[1]) else diff).append(v)
        if norm(htxt[it]) != norm(m[1]):
            shape.append((len(A & B) > 0, A == B))
    rng = np.random.RandomState(0)
    null = float(np.mean([jac({int(c) for c in fpc(Zh[i])},
                              {int(c) for c in fpc(Zp[rng.randint(len(ids))])})
                          for i in range(len(hid)) for _ in range(200)]))
    print(f"\n{len(same) + len(diff)} of {len(hid)} HRS items are also asked by the pool ({enc})")
    print(f"  identical wording  n={len(same):2d}  mean Jaccard of core components {np.mean(same):.2f}")
    print(f"  reworded           n={len(diff):2d}  mean Jaccard of core components {np.mean(diff):.2f}")
    print(f"    of those: {sum(a for a, _ in shape)} share at least one core component, "
          f"{sum(b for _, b in shape)} reach exactly the same set")
    print(f"  random pool item baseline                                    {null:.3f}")


def phrasing_aou(enc="gte-large-en-v1.5"):
    """The same comparison for the All of Us battery, which shares far more of its text with the pool."""
    def tail(t):
        return re.sub(r"[^a-z]", "", str(t).lower())[-45:]

    def jac(a, b):
        return len(a & b) / len(a | b) if (a | b) else 0.0

    fit = glob.glob(os.path.join(HERE, "outputs_pool", "fits", f"{enc}__k*_ls1.0"))[0]
    W = np.load(os.path.join(fit, "loadings.npy")); mu = np.load(os.path.join(fit, "mean.npy"))
    Zp = np.load(os.path.join(fit, "scores.npy")); sigma = Zp.std(0); Zp = Zp / sigma
    pp = pd.read_csv(os.path.join(GP, "pool_processed.csv")); ids = list(pp.item_id)
    d = np.load(os.path.join(HERE, "data", f"core151_items_{enc}.npz"), allow_pickle=True)
    aid = [str(x) for x in d["items"]]
    Za = ((np.asarray(d["vectors"], float) - mu) @ W.T) / sigma
    ai = pd.read_csv(os.path.join(HERE, "core_items_151.csv"))
    atxt = dict(zip(ai.item, ai.prompt))
    pool_by_tail = {tail(t): (i, t) for i, t in zip(ids, pp.prompt_clean)}

    exact, differ, shape = [], [], []
    for i, it in enumerate(aid):
        k = tail(atxt.get(it, ""))
        if len(k) < 20:
            continue
        m = next((pool_by_tail[q] for q in pool_by_tail if q.endswith(k) or k.endswith(q)), None)
        if m is None:
            continue
        A = {int(c) for c in fpc(Za[i])}
        B = {int(c) for c in fpc(Zp[ids.index(m[0])])}
        v = jac(A, B)
        (exact if str(atxt[it]).strip() == str(m[1]).strip() else differ).append(v)
        if str(atxt[it]).strip() != str(m[1]).strip():
            shape.append((len(A & B) > 0, A == B))
    rng = np.random.RandomState(0)
    null = float(np.mean([jac({int(c) for c in fpc(Za[i])},
                              {int(c) for c in fpc(Zp[rng.randint(len(ids))])})
                          for i in range(len(aid)) for _ in range(100)]))
    print(f"\n{len(exact) + len(differ)} of {len(aid)} All of Us items are also asked by the pool ({enc})")
    print(f"  identical text     n={len(exact):3d}  mean Jaccard {np.mean(exact):.3f} "
          f"(min {min(exact):.2f})")
    print(f"  differing text     n={len(differ):3d}  mean Jaccard {np.mean(differ):.2f}")
    print(f"    of those: {sum(a for a, _ in shape)} share at least one core component, "
          f"{sum(b for _, b in shape)} reach exactly the same set")
    print(f"  random pool item baseline                                    {null:.3f}")


if __name__ == "__main__":
    main()
```
