## Supplemental code for "Scoring respondents on a semantic frame of reference identifies a group that is burdened across many questionnaires and reaches care less easily": hrs_xenc_itemside_profiles.py

```
#!/usr/bin/env python
"""Item-side cross-encoder agreement for the HRS battery, on construct PROFILES (D6-A).

The settled definitions retire modal labelling: a column is read by the SET of documented
constructs of its core items. So for an HRS item g, under encoder e,

    S_g^e  =  union over c in F_g^e  of  { C_h : h in E_c^e }

    F_g^e  core components of the HRS item  (fixed-point PR down its row of Z_S, D5)
    E_c^e  core items of pool column c      (fixed-point PR down that column of Z_G, D4)
    C_h    documented construct of pool item h (pool_processed.csv)

The columns are read by the POOL's own items on both sides, which is the point: the basis is fixed
before HRS is seen. Agreement = mean pairwise Jaccard of S_g over the 15 encoder pairs, against a
null that permutes item identities (the null of sec:methods_xenc). The pool's own 577 items are run
through the identical code as a control, and the retired modal form is reported alongside, so any
change is attributable to the labelling and nothing else.

Placement follows D7 exactly: P_S = (M_S - mu_G) W_G^T, Z_S = P_S / sigma_G, no refitting.

Run: CUDA_VISIBLE_DEVICES="" /data_gpu5/ROSS_Proj/env/bin/python hrs_xenc_itemside_profiles.py
"""
import os, glob, csv
from itertools import combinations
import numpy as np
import pandas as pd

HERE = os.path.dirname(os.path.abspath(__file__))
GP = os.path.join(HERE, "..", "global_pool")
HRSD = os.path.join(HERE, "..", "randhrsdata", "acidd_input")
ENCS = ["gte-large-en-v1.5", "bge-m3", "bge-large-en-v1.5", "all-mpnet-base-v2",
        "e5-large-v2", "qwen3-embedding-0.6b"]
RNG = np.random.RandomState(0)
NPERM = 200

# HRS documents its eight neighborhood items as two scales, split by the published item text
# (the same clerical split as fig_hrs_twoside_cores.py and Methods sec:methods_hrs).
HRS_COHESION = ("really feel part of this area", "can be trusted", "are friendly", "were in trouble")
HRS_DISORDER = ("vandalism and graffiti", "safe walking alone", "kept very clean", "vacant houses")


def hrs_construct(label, prompt):
    if label != "Neighborhood":
        return label
    t = str(prompt).lower()
    if any(k in t for k in HRS_COHESION):
        return "Neighborhood social cohesion"
    if any(k in t for k in HRS_DISORDER):
        return "Neighborhood disorder & safety"
    return label


def pr(v):
    s2 = float((v**2).sum()); s4 = float((v**4).sum())
    return s2 * s2 / s4 if s4 > 0 else 0.0


def fpc(v):
    idx = np.arange(len(v))
    while True:
        n = max(1, min(int(round(pr(v[idx]))), len(idx)))
        top = np.sort(idx[np.argsort(-np.abs(v[idx]))[:n]])
        if len(top) == len(idx) and (top == idx).all():
            return top
        idx = top


def jac(a, b):
    return len(a & b) / len(a | b) if (a | b) else 1.0


def mean_pairwise(sets_by_enc, perm=False):
    n = len(sets_by_enc[0]); vals = []
    for A, B in combinations(range(len(sets_by_enc)), 2):
        pb = RNG.permutation(n) if perm else np.arange(n)
        vals.append(np.mean([jac(sets_by_enc[A][i], sets_by_enc[B][pb[i]]) for i in range(n)]))
    return float(np.mean(vals))


def per_item(sets_by_enc):
    n = len(sets_by_enc[0])
    return np.array([np.mean([jac(sets_by_enc[A][i], sets_by_enc[B][i])
                              for A, B in combinations(range(len(sets_by_enc)), 2)]) for i in range(n)])


pp = pd.read_csv(os.path.join(GP, "pool_processed.csv"))
con = dict(zip(pp.item_id, pp.construct))
pool_ids = list(pp.item_id)

hitems = pd.read_csv(os.path.join(HRSD, "hrs_lb2022_items.csv"))
hcon = {r.item: hrs_construct(r.construct, r.prompt) for r in hitems.itertuples()}

pool_prof, pool_modal = [], []
hrs_prof, hrs_modal, hrs_sizes = [], [], []
aou_prof, aou_modal = [], []
hids = aids = None
for enc in ENCS:
    fit = glob.glob(os.path.join(HERE, "outputs_pool", "fits", f"{enc}__k*_ls1.0"))[0]
    W = np.load(os.path.join(fit, "loadings.npy"))
    mu = np.load(os.path.join(fit, "mean.npy"))
    Zg = np.load(os.path.join(fit, "scores.npy"))
    sigma = Zg.std(0)                                   # D2, fixed on the pool
    rows = list(csv.DictReader(open(os.path.join(HERE, "outputs_pool", "fixedpoint_cores",
                                                 f"{enc}_cores.csv"))))
    core_items = {int(r["component"]): str(r["items"]).split() for r in rows}
    modal = {int(r["component"]): r["modal_construct"] for r in rows}
    profile = {c: frozenset(con[i] for i in v) for c, v in core_items.items()}

    Zt_pool = Zg / sigma
    pp_sets, pm_sets = [], []
    for j in range(len(pool_ids)):
        F = [int(c) for c in fpc(Zt_pool[j])]
        pp_sets.append(frozenset().union(*[profile[c] for c in F]))
        pm_sets.append(frozenset(modal[c] for c in F))
    pool_prof.append(pp_sets); pool_modal.append(pm_sets)

    d = np.load(os.path.join(HRSD, f"hrs_lb2022_{enc}.npz"), allow_pickle=True)
    if hids is None:
        hids = [str(x) for x in d["items"]]
    assert [str(x) for x in d["items"]] == hids, f"HRS item order differs under {enc}"
    Zt_hrs = ((np.asarray(d["vectors"], float) - mu) @ W.T) / sigma       # D7
    hp_sets, hm_sets, sz = [], [], []
    for j in range(len(hids)):
        F = [int(c) for c in fpc(Zt_hrs[j])]
        hp_sets.append(frozenset().union(*[profile[c] for c in F]))
        hm_sets.append(frozenset(modal[c] for c in F))
        sz.append(len(F))
    hrs_prof.append(hp_sets); hrs_modal.append(hm_sets); hrs_sizes.append(sz)

    # the broad battery placed the same way, so "reproduces less well than All of Us" is like for like
    a = np.load(os.path.join(HERE, "data", f"core151_items_{enc}.npz"), allow_pickle=True)
    if aids is None:
        aids = [str(x) for x in a["items"]]
    assert [str(x) for x in a["items"]] == aids, f"All of Us item order differs under {enc}"
    Zt_aou = ((np.asarray(a["vectors"], float) - mu) @ W.T) / sigma       # D7
    ap_sets, am_sets = [], []
    for j in range(len(aids)):
        F = [int(c) for c in fpc(Zt_aou[j])]
        ap_sets.append(frozenset().union(*[profile[c] for c in F]))
        am_sets.append(frozenset(modal[c] for c in F))
    aou_prof.append(ap_sets); aou_modal.append(am_sets)

hrs_sizes = np.array(hrs_sizes)

print(f"{len(hids)} HRS items, {len(pool_ids)} pool items, {len(ENCS)} encoders, {NPERM} permutations\n")
print(f"{'battery / labelling':38s} {'mean |set|':>10s} {'real':>7s} {'null':>7s} {'chance-corr.':>13s}")
for name, sets in [("pool 577, construct profile (control)", pool_prof),
                   ("pool 577, modal construct (retired)", pool_modal),
                   (f"All of Us {len(aids)}, construct profile", aou_prof),
                   (f"All of Us {len(aids)}, modal construct", aou_modal),
                   ("HRS 35, construct profile", hrs_prof),
                   ("HRS 35, modal construct (retired)", hrs_modal)]:
    real = mean_pairwise(sets)
    null = np.mean([mean_pairwise(sets, perm=True) for _ in range(NPERM)])
    msz = np.mean([len(s) for e in sets for s in e])
    print(f"{name:38s} {msz:10.2f} {real:7.3f} {null:7.3f} {(real-null)/(1-null):13.3f}")

pi = per_item(hrs_prof); pim = per_item(hrs_modal)
mean_size = hrs_sizes.mean(0)
print(f"\nHRS core components per item: median {np.median(hrs_sizes):.1f} over all encoders and items")
print("  per encoder (median):",
      ", ".join(f"{e} {np.median(hrs_sizes[k]):.0f}" for k, e in enumerate(ENCS)))

print("\nHRS item-side agreement by the item's average number of core components:")
print(f"{'core components':16s} {'items':>6s} {'profile':>9s} {'modal':>8s}")
for lo, hi, tag in [(0, 1.5, "1"), (1.5, 2.5, "2"), (2.5, 4.5, "3 to 4"), (4.5, 99, "5 or more")]:
    m = (mean_size >= lo) & (mean_size < hi)
    if m.sum():
        print(f"{tag:16s} {int(m.sum()):6d} {pi[m].mean():9.3f} {pim[m].mean():8.3f}")

print("\nHRS item-side agreement by documented HRS construct:")
print(f"{'construct':34s} {'items':>6s} {'profile':>9s} {'modal':>8s}")
order = ["Everyday discrimination", "Healthcare discrimination", "Loneliness",
         "Neighborhood social cohesion", "Neighborhood disorder & safety", "Perceived stress"]
seen = [c for c in order if any(hcon[h] == c for h in hids)]
seen += sorted({hcon[h] for h in hids} - set(seen))
for c in seen:
    idx = [i for i, h in enumerate(hids) if hcon[h] == c]
    print(f"{c:34s} {len(idx):6d} {pi[idx].mean():9.3f} {pim[idx].mean():8.3f}")
```
