## Supplemental code for "Scoring respondents on a semantic frame of reference identifies a group that is burdened across many questionnaires and reaches care less easily": lambda_sweep_pool.py

```
#!/usr/bin/env python
"""Lambda sweep for the pool reference basis, scored on label-free diagnostics only.

Fits the pool basis at each lambda_score from an independent PCA initialization (the estimator's
default init), holding everything else at the production settings: k from parallel analysis,
lambda_ortho = 0.1, seed 42, full-batch, 5000 steps.

DIAGNOSTICS ARE LABEL-FREE BY DESIGN. Orthogonality, core size, core energy and explained variance
use only the embeddings and the fit. Faithfulness is deliberately NOT computed here: the manuscript
claims that documented labels "arrive only once the components exist, which lets them serve as a test
rather than an input", and choosing lambda by faithfulness would make them an input and void the test.
Report faithfulness afterwards as a consequence of the chosen lambda, never as its criterion.

  orthogonality   max |off-diagonal| of the Gram matrix of the row-normalized components W (0 = exactly
                  orthonormal). The sparsity penalty acts on the item projection, so W can drift.
  core size       per component, the fixed point of the participation-ratio pruning on its column of
                  the item projection P. Reported as the median over components.
  core energy     per component, the share of its squared weight carried by its core. Median over
                  components.
  explained var   sum of the item projection's column variances over the total variance of the centered
                  embeddings, and its ratio to the PCA value at the same k (1.0 = no variance given up).

Out: outputs_pool/lambda_sweep.csv, and figures/lambda_sweep.png via --plot
Run: CUDA_VISIBLE_DEVICES="" /data_gpu5/ROSS_Proj/env/bin/python lambda_sweep_pool.py [--only ENC ...]
"""
import argparse, json, os, time
import numpy as np
import pandas as pd
from sklearn.decomposition import PCA

from ross_proj import fit_projection
from _common import EMBEDDERS, load_embedding

HERE = os.path.dirname(os.path.abspath(__file__))
OUT = os.path.join(HERE, "outputs_pool")
LAMBDAS = [0.001, 0.005, 0.01, 0.05, 0.1, 0.5, 1.0, 5.0]


def pr(v):
    s2 = float((v ** 2).sum()); s4 = float((v ** 4).sum())
    return s2 * s2 / s4 if s4 > 0 else 0.0


def fpc(v):
    idx = np.arange(len(v))
    while True:
        n = max(1, min(int(round(pr(v[idx]))), len(idx)))
        top = np.sort(idx[np.argsort(-np.abs(v[idx]))[:n]])
        if len(top) == len(idx) and (top == idx).all():
            return top
        idx = top


def diagnostics(W, P, total_var, pca_var):
    Wn = W / np.linalg.norm(W, axis=1, keepdims=True)
    G = Wn @ Wn.T
    ortho = float(np.abs(G - np.eye(G.shape[0])).max())
    sizes, energies = [], []
    for c in range(P.shape[1]):
        v = P[:, c]
        core = fpc(v)
        sizes.append(len(core))
        tot = float((v ** 2).sum())
        energies.append(float((v[core] ** 2).sum()) / tot if tot > 0 else np.nan)
    ve = float(P.var(0, ddof=0).sum() / total_var)
    return dict(ortho_max_offdiag=ortho,
                core_size_median=float(np.median(sizes)),
                core_size_min=int(np.min(sizes)), core_size_max=int(np.max(sizes)),
                core_energy_median=float(np.median(energies)),
                explained_var=ve, explained_var_ratio_to_pca=ve / pca_var)


def faithfulness(P, items, con, nperm=200, seed=0):
    """Leave-one-out rate that an item's documented construct appears among the constructs of the
    core items of its core components, and its label-shuffle null. REPORTED, NEVER A CRITERION."""
    Pt = P / P.std(0)
    core = {c: [items[j] for j in fpc(Pt[:, c])] for c in range(P.shape[1])}
    F = {g: [int(c) for c in fpc(Pt[j])] for j, g in enumerate(items)}

    def rate(lab):
        ok = 0
        for g in items:
            s = set()
            for c in F[g]:
                s.update(lab[h] for h in core[c] if h != g)
            ok += lab[g] in s
        return ok / len(items)

    rng = np.random.RandomState(seed)
    vals = list(con.values())
    real = rate(con)
    null = float(np.mean([rate(dict(zip(items, rng.permutation(vals)))) for _ in range(nperm)]))
    return real, null, real - null


def main():
    ap = argparse.ArgumentParser()
    ap.add_argument("--only", nargs="*", choices=list(EMBEDDERS))
    ap.add_argument("--lambdas", nargs="*", type=float, default=LAMBDAS)
    ap.add_argument("--plot", action="store_true")
    ap.add_argument("--faithfulness", action="store_true",
                    help="also report faithfulness and its null (a READOUT, not a selection criterion)")
    args = ap.parse_args()

    con_all = None
    if args.faithfulness:
        pp = pd.read_csv(os.path.join(HERE, "..", "global_pool", "pool_processed.csv"))
        con_all = dict(zip(pp.item_id, pp.construct))

    rows = []
    for name in (args.only or list(EMBEDDERS)):
        items, X = load_embedding(name)
        k = int(json.load(open(os.path.join(OUT, "k_selection", name,
                                            "parallel_analysis.json")))["k"])
        Xc = X - X.mean(0)
        total_var = float(Xc.var(0, ddof=0).sum())
        pca_var = float(PCA(n_components=k, random_state=42).fit_transform(X).var(0, ddof=0).sum()
                        / total_var)
        for ls in args.lambdas:
            t0 = time.time()
            res = fit_projection(X, n_components=k, lambda_score=ls, lambda_ortho=0.1,
                                 batch_size=X.shape[0], subsample_fraction=1.0,
                                 total_steps=5000, random_state=42, verbose=False, log_interval=0)
            d = diagnostics(np.asarray(res.W), np.asarray(res.Z), total_var, pca_var)
            if args.faithfulness:
                ids = [str(h) for h in items]
                r, n_, m = faithfulness(np.asarray(res.Z), ids, {h: con_all[h] for h in ids})
                d.update(faith_rate=r, faith_chance=n_, faith_margin=m)
            d.update(encoder=name, k=k, lambda_score=ls, seconds=round(time.time() - t0, 1))
            rows.append(d)
            print(f"{name:24s} k={k:3d} lam={ls:<6g} ortho={d['ortho_max_offdiag']:.4f} "
                  f"core={d['core_size_median']:5.1f} energy={d['core_energy_median']:.3f} "
                  f"VE/PCA={d['explained_var_ratio_to_pca']:.4f}"
                  + (f" faith={d['faith_rate']:.3f}/{d['faith_chance']:.3f} margin={d['faith_margin']:.3f}"
                     if args.faithfulness else "")
                  + f"  [{d['seconds']}s]", flush=True)

    cols = ["encoder", "k", "lambda_score", "ortho_max_offdiag",
            "core_size_median", "core_size_min", "core_size_max",
            "core_energy_median", "explained_var", "explained_var_ratio_to_pca"]
    if args.faithfulness:
        cols += ["faith_rate", "faith_chance", "faith_margin"]
    df = pd.DataFrame(rows)[cols + ["seconds"]]
    tag = f"_{args.only[0]}" if args.only and len(args.only) == 1 else ""
    out = os.path.join(OUT, f"lambda_sweep{tag}.csv")
    df.to_csv(out, index=False)
    print(f"\nwrote {out}")

    if args.plot:
        import matplotlib
        matplotlib.use("Agg")
        import matplotlib.pyplot as plt
        panels = [("ortho_max_offdiag", "max |off-diagonal| of $W W^\\top$"),
                  ("core_size_median", "median core size (items)"),
                  ("core_energy_median", "median core energy"),
                  ("explained_var_ratio_to_pca", "explained variance / PCA")]
        colours = ["#0072B2", "#D55E00", "#009E73", "#CC79A7", "#E69F00", "#56B4E9"]
        fig, axes = plt.subplots(1, 4, figsize=(15.5, 3.6))
        for ax, (col, lab) in zip(axes, panels):
            for i, (enc, g) in enumerate(df.groupby("encoder")):
                g = g.sort_values("lambda_score")
                ax.plot(g.lambda_score, g[col], "-o", ms=3.5, lw=1.3,
                        color=colours[i % len(colours)], label=enc)
            ax.set_xscale("log"); ax.set_xlabel("$\\lambda_{\\mathrm{score}}$")
            ax.set_ylabel(lab); ax.grid(alpha=0.25, lw=0.6)
        axes[-1].legend(fontsize=7, frameon=False, bbox_to_anchor=(1.02, 1), loc="upper left")
        fig.tight_layout()
        p = os.path.join(HERE, "..", "..", "manuscript_nhb", "figures", "lambda_sweep.png")
        fig.savefig(p, dpi=200, bbox_inches="tight")
        print(f"wrote {p}")


if __name__ == "__main__":
    main()
```
