## Supplemental code for "Scoring respondents on a semantic frame of reference identifies a group that is burdened across many questionnaires and reaches care less easily": make_core_items_pool.py

```
#!/usr/bin/env python3
"""Supplementary Data 3 -- the battery items that form each component, on the POOL basis.

WHAT THIS REPLACES. `make_core_items.py` wrote Supplementary Data 3 from the retired 151-item OWN
basis (k of 23 to 26, `outputs_core151/random_starts/selection_best.json`) into the npj-era
`manuscript/supplementary_data/`. That object is not in the paper any more: the manuscript places
the All of Us battery on the fixed 577-item pool basis and keeps 43 of 55 components under the
exemplar encoder, and main-text R2 cites Supplementary Data 3 for exactly that -- "the items that
form every component of this battery under each encoder".

WHAT IT COMPUTES, per encoder:
  1. the pool fit (W, mu) and the pool's per-column standard deviations sigma;
  2. the 151 All of Us items projected onto it and expressed in pool units,
         Z = ((X_aou - mu) @ W.T) / sigma;
  3. the components KEPT for the battery, by the settled rule in `_common.keep_columns` -- a
     component is kept when at least one battery item clears its pool core-item band, or when it is
     some item's modal component;
  4. for each kept component, its core WITHIN THE BATTERY: the fixed point of the
     participation-ratio pruning down that column of Z, the same rule the pool cores use.

The loading written is the standardized loading on the pool component, so it is comparable across
components and across encoders. Nothing is refitted here and no construct label enters.

In     outputs_pool/fits/<encoder>__k*_ls1.0/{loadings,mean}.npy   the pool fit
       data/pool_items_<encoder>.npz, data/core151_items_<encoder>.npz
       ../global_pool/pool_processed.csv, construct_map_151.csv
       ../../manuscript_nhb/supplementary_data/items_full.csv        (prompts)
Out    ../../manuscript_nhb/supplementary_data/core_items/core_items_<short>.csv
Run    CUDA_VISIBLE_DEVICES="" /data_gpu5/ROSS_Proj/env/bin/python make_core_items_pool.py
"""
import csv
import glob
import os

import numpy as np
import pandas as pd

from _common import keep_columns, pool_bands

HERE = os.path.dirname(os.path.abspath(__file__))
SUPP = os.path.join(HERE, "..", "..", "manuscript_nhb", "supplementary_data")
OUTDIR = os.path.join(SUPP, "core_items")
POOL_IDS = pd.read_csv(os.path.join(HERE, "..", "global_pool", "pool_processed.csv"))["item_id"].tolist()

ENCODERS = {"bge-m3": "bge-m3", "bge-large-en-v1.5": "bge-lg", "all-mpnet-base-v2": "mpnet",
            "e5-large-v2": "e5", "gte-large-en-v1.5": "gte", "qwen3-embedding-0.6b": "qwen"}


def fixed_point_core(v):
    """Keep top-round(PR) by |weight|, recompute PR on the survivors, iterate to convergence.

    The same rule as `pool_fixedpoint_cores.py`. It terminates because PR(S) <= |S| for any set S,
    so the size is non-increasing.
    """
    idx = np.arange(len(v))
    while True:
        w = v[idx]
        s2, s4 = float((w ** 2).sum()), float((w ** 4).sum())
        pr = (s2 * s2) / s4 if s4 > 0 else 0.0
        n = max(1, min(int(round(pr)), len(idx)))
        top = np.sort(idx[np.argsort(-np.abs(w))[:n]])
        if len(top) == len(idx) and (top == idx).all():
            return top
        idx = top


def main():
    items = {r["item"]: r for r in csv.DictReader(open(os.path.join(SUPP, "items_full.csv")))}
    cm = pd.read_csv(os.path.join(HERE, "construct_map_151.csv"))
    ins, con = dict(zip(cm.item, cm.instrument)), dict(zip(cm.item, cm.construct))
    os.makedirs(OUTDIR, exist_ok=True)

    for enc, short in ENCODERS.items():
        fit = glob.glob(os.path.join(HERE, "outputs_pool", "fits", f"{enc}__k*_ls1.0"))
        if not fit:
            raise SystemExit(f"no pool fit for {enc} -- run the stage A pipeline first (README)")
        W = np.load(os.path.join(fit[0], "loadings.npy"))
        mu = np.load(os.path.join(fit[0], "mean.npy"))
        Mg = np.load(os.path.join(HERE, "data", f"pool_items_{enc}.npz"),
                     allow_pickle=True)["vectors"].astype(float)
        Zpool = ((Mg - mu) @ W.T)
        sigma = Zpool.std(0)

        d = np.load(os.path.join(HERE, "data", f"core151_items_{enc}.npz"), allow_pickle=True)
        aid = [str(x) for x in d["items"]]
        Z = ((np.asarray(d["vectors"], float) - mu) @ W.T) / sigma
        assert Z.shape[0] == 151, (enc, Z.shape)

        band = pool_bands(enc, Zpool / sigma, POOL_IDS)
        kept = sorted(keep_columns(Z, band))

        rows = []
        for c in kept:
            core = fixed_point_core(Z[:, c])
            for i in sorted(core, key=lambda i: -abs(Z[i, c])):
                it = items.get(aid[i], {})
                rows.append({"component": f"C{c}", "item": aid[i],
                             "instrument": ins.get(aid[i], it.get("instrument", "")),
                             "construct": con.get(aid[i], it.get("construct", "")),
                             "loading": f"{Z[i, c]:.4f}", "prompt": it.get("prompt", "")})
        path = os.path.join(OUTDIR, f"core_items_{short}.csv")
        with open(path, "w", newline="") as f:
            w = csv.DictWriter(f, fieldnames=["component", "item", "instrument",
                                              "construct", "loading", "prompt"])
            w.writeheader()
            w.writerows(rows)
        print(f"-> core_items_{short}.csv  ({len(kept)} kept components, {len(rows)} core rows)")


if __name__ == "__main__":
    main()
```
