## Supplemental code for "Scoring respondents on a semantic frame of reference identifies a group that is burdened across many questionnaires and reaches care less easily": make_dense_basis_from_npz.py

```
#!/usr/bin/env python
"""Build the CONCENTRATED, UN-PRUNED ACIDD basis files from the existing
`scores_pr_pruned/*.npz` (no ross_proj, no fits, no GPU needed).

Each `<model>__k*_ls0.01__pr_pruned.npz` already stores `scores_dense` = the
un-pruned lambda=0.01 concentrated projection Z (items x k). This script wraps
that Z into survey-semantics' SemanticBasis schema (the same schema
`make_survey_semantics_basis.py` writes), so the detector scores on the
concentrated projection instead of the pruned one.

Pure numpy — copy this file plus the 6 `*_pr_pruned.npz` anywhere and run:
    python make_dense_basis_from_npz.py \
        --in-dir  outputs_core151/scores_pr_pruned \
        --out-dir outputs_core151/ss_basis_dense

Sanity: each printed `sparsity` should be ~0.000 (vs ~0.86 for the pruned basis).
Then copy out-dir/*.npz to the Workbench and point run_survey_semantics_dense.sh
BASIS_DIR at it.
"""
import argparse
import glob
import os

import numpy as np


def write_basis(path, items, Z, model, ls="0.01"):
    """Write Z (items x k) as a survey-semantics SemanticBasis .npz (schema-exact)."""
    Z = np.ascontiguousarray(Z, dtype=float)
    n_items, k = Z.shape
    col_energy = (Z ** 2).sum(0)
    total = float(col_energy.sum())
    explained = (col_energy / total) if total > 0 else np.full(k, 1.0 / k)
    slug = "rossdense_{}_ls{}".format(model, ls)
    np.savez(
        path,
        items=np.asarray(items).astype(str),
        components=Z,                                            # (items x k) == dense concentrated Z
        eigenvalues=col_energy.astype(float),                   # filler (report only)
        explained_variance_ratio=explained.astype(float),       # filler
        cumulative_variance=np.cumsum(explained).astype(float), # filler
        parallel_null_mean=np.zeros(k, dtype=float),
        parallel_null_percentile=np.zeros(k, dtype=float),
        parallel_significant=np.ones(k, dtype=bool),
        parallel_selected_d=np.array([float(k)], dtype=float),  # D = all k columns
        meta=np.array(["ross-proj-dense", model, slug]),
        params=np.array([float(k), 0.0, 95.0, 42.0], dtype=float),
    )
    return n_items, k


def main():
    ap = argparse.ArgumentParser()
    ap.add_argument("--in-dir", default="outputs_core151/scores_pr_pruned")
    ap.add_argument("--out-dir", default="outputs_core151/ss_basis_dense")
    args = ap.parse_args()
    os.makedirs(args.out_dir, exist_ok=True)

    npzs = sorted(glob.glob(os.path.join(args.in_dir, "*__pr_pruned.npz")))
    if not npzs:
        raise SystemExit("no *__pr_pruned.npz in {}".format(args.in_dir))

    for f in npzs:
        d = np.load(f, allow_pickle=True)
        if "scores_dense" not in d:
            raise SystemExit("{} has no 'scores_dense' field".format(f))
        items = np.asarray(d["items"]).astype(str)
        Z = np.asarray(d["scores_dense"], float)
        tag = os.path.basename(f).replace("__pr_pruned.npz", "")     # <model>__k*_ls0.01
        model = tag.split("__", 1)[0]
        out = os.path.join(args.out_dir, tag + "__ss_basis.npz")
        n_items, k = write_basis(out, items, Z, model)
        sparsity = float((Z == 0).mean())
        print("{:34s} {}x{}  D=all({})  dense sparsity {:.3f}  -> {}".format(
            tag, n_items, k, k, sparsity, os.path.basename(out)))

    print("\n-> {}/".format(args.out_dir))


if __name__ == "__main__":
    main()
```
