## Supplemental code for "Scoring respondents on a semantic frame of reference identifies a group that is burdened across many questionnaires and reaches care less easily": make_poolw_basis.py

```
#!/usr/bin/env python
"""Write pool-W AoU screen basis files in survey-semantics `--basis-file` schema.

Drop-in replacement for the AoU-own basis (outputs_core151/ss_basis_dense_selected/):
same schema, same per-encoder item order, but `components` = the AoU items projected
onto the FIXED POOL basis and restricted to the coverage-honest subspace (pool
components that are a core component of >=1 AoU item). Re-running the existing
survey-semantics pipeline with these files screens the 22,332 AoU participants in
pool-component space, keying/residualize/Mahalanobis/consensus all unchanged.

  components = P_AoU[:, kept],  P_AoU = (E_aou - mu_pool) @ W_pool^T
  kept       = union over AoU items of fixed_point_core((P_AoU - mu_s)/sd)   # core of >=1 item

Run: CUDA_VISIBLE_DEVICES="" /data_gpu5/ROSS_Proj/env/bin/python make_poolw_basis.py
"""
import os, glob, numpy as np

HERE = os.path.dirname(os.path.abspath(__file__))
FITS = os.path.join(HERE, "outputs_pool", "fits")
REFDIR = os.path.join(HERE, "outputs_core151", "ss_basis_dense_selected")
OUT = os.path.join(HERE, "outputs_pool", "ss_basis_poolw")
os.makedirs(OUT, exist_ok=True)

ENCS = ["gte-large-en-v1.5", "bge-m3", "bge-large-en-v1.5",
        "all-mpnet-base-v2", "e5-large-v2", "qwen3-embedding-0.6b"]


def fit_dir(enc):
    d = glob.glob(os.path.join(FITS, f"{enc}__k*_ls1.0")); assert len(d) == 1, (enc, d); return d[0]


def refbasis(enc):
    d = glob.glob(os.path.join(REFDIR, f"{enc}__*__ss_basis.npz")); assert len(d) == 1, (enc, d); return d[0]


def pr(v):
    s2 = float((v**2).sum()); s4 = float((v**4).sum()); return s2*s2/s4 if s4 > 0 else 0.0


def fpc(v):
    ix = np.arange(len(v))
    while True:
        n = max(1, min(int(round(pr(v[ix]))), len(ix)))
        top = np.sort(ix[np.argsort(-np.abs(v[ix]))[:n]])
        if len(top) == len(ix) and (top == ix).all():
            return top
        ix = top


def write_basis(path, items, Z):
    Z = np.ascontiguousarray(Z, dtype=float); k = Z.shape[1]
    ce = (Z**2).sum(0); tot = float(ce.sum()); ev = (ce/tot) if tot > 0 else np.full(k, 1.0/k)
    np.savez(path, items=np.asarray(items).astype(str), components=Z,
             eigenvalues=ce.astype(float), explained_variance_ratio=ev.astype(float),
             cumulative_variance=np.cumsum(ev).astype(float),
             parallel_null_mean=np.zeros(k), parallel_null_percentile=np.zeros(k),
             parallel_significant=np.ones(k, dtype=bool),
             parallel_selected_d=np.array([float(k)]),
             meta=np.array(["poolw-aou-screen", "components=P_AoU[:,kept]"]),
             params=np.array([float(k), 0.0, 95.0, 42.0]))


print(f"{'encoder':22s} {'poolk':>5s} {'kept':>4s}  basis file")
for enc in ENCS:
    fd = fit_dir(enc)
    W = np.load(os.path.join(fd, "loadings.npy")); mu = np.load(os.path.join(fd, "mean.npy"))
    Zp = np.load(os.path.join(fd, "scores.npy")); mu_s, sd = Zp.mean(0), Zp.std(0)

    ref = np.load(refbasis(enc), allow_pickle=True)
    items_ref = [str(x) for x in ref["items"]]                # target order (drop-in)

    a = np.load(os.path.join(HERE, "data", f"core151_items_{enc}.npz"), allow_pickle=True)
    pos = {str(it): j for j, it in enumerate(a["items"])}
    E = a["vectors"][[pos[it] for it in items_ref]]           # reorder to ref order

    P = (E - mu) @ W.T                                         # 151 x poolk, items_ref order
    Ztil = (P - mu_s) / sd
    kept = sorted(set().union(*[set(fpc(Ztil[i]).tolist()) for i in range(len(items_ref))]))

    k = W.shape[0]
    out = os.path.join(OUT, f"{enc}__poolw_k{k}__ss_basis.npz")
    write_basis(out, items_ref, P[:, kept])
    # verify round-trip
    back = np.load(out, allow_pickle=True)
    assert back["components"].shape == (151, len(kept))
    assert [str(x) for x in back["items"]] == items_ref
    print(f"{enc:22s} {k:5d} {len(kept):4d}  {os.path.basename(out)}")

print(f"\n-> {OUT}/  ({len(ENCS)} pool-W basis files, drop-in for survey-semantics analyze-file)")
```
