## Supplemental code for "Scoring respondents on a semantic frame of reference identifies a group that is burdened across many questionnaires and reaches care less easily": make_supp_tables.py

```
#!/usr/bin/env python
"""Generate the manuscript supplemental tables into manuscript/supplement.tex.

  items_full.csv (Supplementary Data 1) -- the full 151 selected All of Us survey items: AoU concept
               id, item name, source instrument, the assigned construct and domain, response coding
               (numeric value range), and full prompt text. Sorted by domain then construct. Shipped
               as a CSV, not a PDF table; the supplement's S1 section is a short pointer to it.
  Sec./Table S2 -- recovery of validated subscale structure: all 15 BFI-2-XS items, our assigned
               construct (= trait domain), and the published facet (Soto & John 2017).
  Table S3  -- the six sentence encoders: Hugging Face identifier, embedding dimension D,
               number of components k (parallel analysis), and citation.
  Table S4  -- the 600 content-versus-syntax variants.

Inputs (examples/allofus/):
  core_items_151.csv        item, prompt, survey, concept_id  (151 rows)
  encoding_map_all151.csv   concept_id, item, survey, instrument, rule, response, numeric_value
  outputs_core151/pca_baseline.csv   k per encoder (column `k`)

Output: ../../manuscript/supplement.tex, and supplementary data in
        ../../manuscript/supplementary_data/ (items_full.csv + content_grammar_variants/).

Run from examples/allofus/:
  /data_gpu5/ROSS_Proj/env/bin/python make_supp_tables.py
"""
import csv
import glob
import json
import os
import shutil

HERE = os.path.dirname(os.path.abspath(__file__))
OUT = os.path.join(HERE, "..", "..", "manuscript", "supplement.tex")
PCA_BASELINE = os.path.join(HERE, "outputs_core151", "pca_baseline.csv")
VARIANTS_CSV = os.path.join(HERE, "outputs_core151", "content_grammar_v2",
                            "generated_items_v2.csv")
FRAME_CSV = os.path.join(HERE, "frame_map_151.csv")
KSEL_CSV = os.path.join(HERE, "outputs_core151", "projection_geometry", "k_selection.csv")
PSELECT_JSON = os.path.join(HERE, "outputs_core151", "random_starts", "selection_best.json")

# Supplementary data files shipped inside manuscript/ (zipped into manuscript.zip).
SUPP = os.path.join(HERE, "..", "..", "manuscript", "supplementary_data")
CGV_SRC = os.path.join(HERE, "outputs_core151", "content_grammar_v2")   # content-vs-syntax variants
CGV_DST = os.path.join(SUPP, "content_grammar_variants")

SUPP_README = """\
# Supplementary data

Supplementary Data 1 -- items_full.csv
  The 151 selected All of Us survey items, one per row: aou_concept_id, item (variable name),
  instrument, construct, domain, coding (response value range), prompt (full item text).
  Sorted by domain then construct.

Supplementary Data 2 -- content_grammar_variants/
  The content-vs-syntax control experiment: 151 seeds x 2 variant types x 10 donors = 3,020 variants.
    manifest.csv            seed-to-donor design for every variant: variant_id, cell (A syntax /
                            B content), seed_item, donor_item, intended_item, seed/donor frame and
                            construct, and the seed and donor prompt texts.
    generated_items_v2.csv  the assembled rewrites: variant_id, cell, seed_item, donor_item,
                            intended_item, rewritten_prompt.
    batch_s*.jsonl          the raw generations (claude-opus-4-8), one {variant_id, rewritten} per
                            line, batched by seed range; 3,020 lines in total.
  Cell A = syntax variant (seed content in a donor's neutral syntax).
  Cell B = content variant (seed syntax with a donor's content).

Supplementary Data 3 -- core_items/
  The items carrying each component, one CSV per sentence encoder, one row per core item:
  component, item (variable name), instrument, construct, loading (the item's weight on that
  component), prompt (full item text). Ordered by component, then by descending |loading|.
    core_items_bge-m3.csv   bge-m3                 (25 components)
    core_items_bge-lg.csv   bge-large-en-v1.5      (25)
    core_items_mpnet.csv    all-mpnet-base-v2      (24)
    core_items_e5.csv       e5-large-v2            (23)
    core_items_gte.csv      gte-large-en-v1.5      (26, the exemplar read in the main text)
    core_items_qwen.csv     qwen3-embedding-0.6b   (25)
  Written by make_core_items_pool.py, which projects the battery onto the fixed pool basis, keeps
  the components the battery reaches, and takes the fixed-point core of each kept column,
  crossencoder_cores/construct_consensus.csv, and projection_geometry/construct_cores_gte.csv.
"""

# The 13 syntax families (build_frame_map.py): family -> (content_bearing, template,
# [source instruments]). Read from frame_map_151.csv so it cannot drift from the data.
def _load_frames():
    fam_items, fam_cb = {}, {}
    inst_of = {}
    enc = list(csv.DictReader(open(os.path.join(HERE, "encoding_map_all151.csv"))))
    cid_inst = {}
    for r in enc:
        cid_inst.setdefault(r["concept_id"], r["instrument"])
    i2c = {r["item"]: r["concept_id"] for r in csv.DictReader(open(os.path.join(HERE, "core_items_151.csv")))}
    for r in csv.DictReader(open(FRAME_CSV)):
        fam_items.setdefault(r["frame_family"], []).append(r["item"])
        fam_cb[r["frame_family"]] = bool(int(r["content_bearing"]))
        inst_of[r["item"]] = cid_inst.get(i2c.get(r["item"]), "?")
    return fam_items, fam_cb, inst_of


FRAME_ITEMS, FRAME_CB, FRAME_INST = _load_frames()
# content-bearing seed = a seed whose syntax family is content-bearing (syntax taxonomy, not a list)
CONTENT_BEARING = {it for fam, its in FRAME_ITEMS.items() if FRAME_CB[fam] for it in its}

# short, human template per syntax family (display only) + display order (content-bearing first)
FRAME_TEMPLATE = {
    "discrimination": r"``You are treated with less \_\_\_ / a \_\_\_ acts as if \_\_\_''",
    "childhood": r"``During your first 18 years of life, \_\_\_''",
    "neighborhood": r"``\_\_\_ in my neighborhood / my neighborhood is \_\_\_''",
    "social_support": r"``Someone to \_\_\_''",
    "food_scarcity": r"``Within the past 12 months, \dots money to buy/get more''",
    "housing": r"``\dots worried about NOT having a place to live''",
    "trait": r"``I am someone who \_\_\_''",
    "symptom": r"``[bare symptom phrase]''",
    "stress_frequency": r"``In the last month, how often have you felt \_\_\_''",
    "self_state": r"``I feel \_\_\_ / I lack \_\_\_''",
    "frequency_behavior": r"``How often do you \_\_\_''",
    "lifetime": r"``Have you ever / Did you ever \_\_\_''",
    "self_rating": r"``In general, how would you rate your \_\_\_''",
}
FRAME_ORDER = ["discrimination", "childhood", "neighborhood", "social_support", "food_scarcity",
               "housing", "trait", "symptom", "stress_frequency", "self_state",
               "frequency_behavior", "lifetime", "self_rating"]

# encoding_map instrument code -> canonical name used in main-text Table 1.
CANON = {
    "BFI-2-XS":      "Big Five Inventory--2 Extra-Short Form",
    "nds (adapted)": "Perceived Neighborhood Disorder (adapted)",
    "MHQ":           "UK Biobank Mental Health Questionnaire",
    "ACE-BRFSS":     "Adverse Childhood Experiences (BRFSS)",
    "PSS-10":        "Perceived Stress Scale-10",
    "PHQ-9":         "Patient Health Questionnaire-9",
    "CIDI-5":        "WHO World Mental Health CIDI",
    "EDS":           "Everyday Discrimination Scale",
    "UCLA-V3":       "Short-Form UCLA Loneliness Scale (ULS-8)",
    "MOS-SS":        "Modified MOS Social Support Survey",
    "DMS":           "Discrimination in Medical Settings",
    "GAD-7":         "Generalized Anxiety Disorder-7",
    "ASRS-v1.1":     "Adult ADHD Self-Report Scale v1.1",
    "BMMRS-DSE":     "Daily Spiritual Experiences (BMMRS)",
    "scns (adapted)": "Neighborhood Social Cohesion (adapted)",
    "PROMIS-Global": "PROMIS Global Health",
    "HVS":           "Hunger Vital Sign",
    "single item":   "Standalone single item",
}
# main-text Table 1 ordering (scales by descending item count, then single items last).
ORDER = ["BFI-2-XS", "nds (adapted)", "MHQ", "ACE-BRFSS", "PSS-10", "PHQ-9", "CIDI-5",
         "EDS", "UCLA-V3", "MOS-SS", "DMS", "GAD-7", "ASRS-v1.1", "BMMRS-DSE",
         "scns (adapted)", "PROMIS-Global", "HVS", "single item"]

# construct/domain display order = the manuscript taxonomy (examples/allofus/build_construct_map.py).
import build_construct_map as _bcm                                   # noqa: E402
CONSTRUCT_ORDER = list(_bcm.TAXONOMY.keys())
DOMAIN_ORDER = []
for _c in CONSTRUCT_ORDER:
    _d = _bcm.TAXONOMY[_c][0]
    if _d not in DOMAIN_ORDER:
        DOMAIN_ORDER.append(_d)

# The six encoders (canonical order); k filled from pca_baseline.csv.
ENCODERS = [
    ("bge-m3",               "BAAI/bge-m3",                          1024, "bgem3_chen2024"),
    ("bge-large-en-v1.5",    "BAAI/bge-large-en-v1.5",               1024, "bge_xiao2023"),
    ("all-mpnet-base-v2",    "sentence-transformers/all-mpnet-base-v2", 768, "reimers2019sentencebert,mpnet_song2020"),
    ("e5-large-v2",          "intfloat/e5-large-v2",                 1024, "e5_wang2022"),
    ("gte-large-en-v1.5",    "Alibaba-NLP/gte-large-en-v1.5",        1024, "gte_li2023,mgte_zhang2024"),
    ("qwen3-embedding-0.6b", "Qwen/Qwen3-Embedding-0.6B",            1024, "qwen3emb_zhang2025"),
]

_SPECIAL = [("\\", r"\textbackslash{}"), ("&", r"\&"), ("%", r"\%"), ("$", r"\$"),
            ("#", r"\#"), ("_", r"\_"), ("{", r"\{"), ("}", r"\}"),
            ("~", r"\textasciitilde{}"), ("^", r"\textasciicircum{}")]


def tex(s):
    s = str(s)
    for a, b in _SPECIAL:
        s = s.replace(a, b)
    return s


def coding_str(vals):
    """Compact response coding from the set of numeric values for an item."""
    v = sorted({int(round(float(x))) for x in vals})
    if len(v) <= 1:
        return str(v[0]) if v else "--"
    if len(v) == 2:
        return f"{v[0]} / {v[1]}"
    if v == list(range(v[0], v[-1] + 1)):          # contiguous integer scale
        return f"{v[0]}--{v[-1]}"
    return ", ".join(map(str, v))


def load_data():
    prompts, survey, cid = {}, {}, {}
    with open(os.path.join(HERE, "core_items_151.csv")) as f:
        for r in csv.DictReader(f):
            prompts[r["item"]] = r["prompt"]
            survey[r["item"]] = r["survey"]
            cid[r["item"]] = r["concept_id"]
    inst, vals = {}, {}
    with open(os.path.join(HERE, "encoding_map_all151.csv")) as f:
        for r in csv.DictReader(f):
            inst[r["item"]] = r["instrument"]
            vals.setdefault(r["item"], []).append(r["numeric_value"])
    k_of = {}
    with open(PCA_BASELINE) as f:
        for r in csv.DictReader(f):
            k_of[r["embedder"]] = int(r["k"])
    constr, domain = {}, {}
    with open(os.path.join(HERE, "construct_map_151.csv")) as f:
        for r in csv.DictReader(f):
            constr[r["item"]] = r["construct"]
            domain[r["item"]] = r["domain"]
    return prompts, survey, cid, inst, vals, k_of, constr, domain


def table_s1(prompts, cid, inst, vals, constr, domain):
    # display order: follow the manuscript taxonomy (domain, then construct), then instrument, item
    dord = {d: i for i, d in enumerate(DOMAIN_ORDER)}
    cord = {c: i for i, c in enumerate(CONSTRUCT_ORDER)}
    items = sorted(prompts, key=lambda it: (dord.get(domain[it], 99), cord.get(constr[it], 99),
                                            ORDER.index(inst[it]), it))
    head = r"AoU id & Item & Instrument & Construct & Domain & Coding & Prompt \\"
    lines = [
        r"\begin{small}",
        r"\begin{longtable}{@{}r p{1.7cm} p{1.3cm} p{2.0cm} p{1.9cm} c p{4.5cm}@{}}",
        r"\caption{\textbf{The 151 selected All of Us survey items, with construct and domain.} "
        r"AoU concept id, item variable name, source instrument (by acronym, with full names in "
        r"main-text Table~1), the assigned construct and domain (main-text "
        r"Table~\ref{tab:constructs}), response coding (numeric value range), and full prompt "
        r"text. Sorted by domain, then construct.}\label{tab:items_full}\\",
        r"\toprule",
        head,
        r"\midrule",
        r"\endfirsthead",
        r"\multicolumn{7}{l}{\small\itshape Table S1 continued}\\",
        r"\toprule " + head + r" \midrule",
        r"\endhead",
        r"\midrule \multicolumn{7}{r}{\small\itshape continued on next page}\\",
        r"\endfoot",
        r"\bottomrule",
        r"\endlastfoot",
    ]
    prev = None
    for it in items:
        if domain[it] != prev:                     # rule between domain blocks
            if prev is not None:
                lines.append(r"\addlinespace[2pt]")
            prev = domain[it]
        row = (f"{tex(cid[it])} & \\texttt{{\\footnotesize {tex(it)}}} & "
               f"\\footnotesize {tex(_bcm.disp(inst[it]))} & \\footnotesize {tex(constr[it])} & "
               f"\\footnotesize {tex(domain[it])} & {coding_str(vals[it])} & "
               f"\\footnotesize {tex(prompts[it])} \\\\")
        lines.append(row)
    lines += [r"\end{longtable}", r"\end{small}", ""]
    return "\n".join(lines)


def write_items_csv(prompts, cid, inst, vals, constr, domain):
    """Ship the full 151-item Table S1 as Supplementary Data 1 (items_full.csv), not a PDF table."""
    dord = {d: i for i, d in enumerate(DOMAIN_ORDER)}
    cord = {c: i for i, c in enumerate(CONSTRUCT_ORDER)}
    items = sorted(prompts, key=lambda it: (dord.get(domain[it], 99), cord.get(constr[it], 99),
                                            ORDER.index(inst[it]), it))
    os.makedirs(SUPP, exist_ok=True)
    path = os.path.join(SUPP, "items_full.csv")
    with open(path, "w", newline="") as f:
        w = csv.writer(f)
        w.writerow(["aou_concept_id", "item", "instrument", "construct", "domain", "coding", "prompt"])
        for it in items:
            w.writerow([cid[it], it, _bcm.disp(inst[it]), constr[it], domain[it],
                        coding_str(vals[it]).replace("--", "–"), prompts[it]])
    return path


def copy_variant_data():
    """Copy the content-vs-syntax variants (Supplementary Data 2) into manuscript/ so they ship in
    manuscript.zip: the seed-to-donor manifest, the assembled rewrites, and the raw batch .jsonl."""
    os.makedirs(CGV_DST, exist_ok=True)
    files = [os.path.join(CGV_SRC, "manifest.csv"),
             os.path.join(CGV_SRC, "generated_items_v2.csv")]
    files += sorted(glob.glob(os.path.join(CGV_SRC, "batch_s*.jsonl")))
    for src in files:
        shutil.copy2(src, os.path.join(CGV_DST, os.path.basename(src)))
    with open(os.path.join(SUPP, "README.md"), "w") as f:
        f.write(SUPP_README)
    return len(files)


def items_pointer():
    """Short supplement stub in place of Table S1 -- the full item table now ships as items_full.csv."""
    return ("The 151 selected items, each with its AoU concept id, variable name, source instrument, "
            "assigned construct and domain, response coding, and full prompt text, are provided as "
            r"\textbf{Supplementary Data 1} (\texttt{items\_full.csv}), sorted by domain then construct. "
            r"The construct and domain assignments follow the manuscript taxonomy (main-text "
            r"Table~\ref{tab:constructs}).")


def robustness_section():
    """Response-style and severity robustness checks moved out of main-text section 3.4.2."""
    return "\n".join([
        r"The breadth of the consensus outliers across instruments (main-text \S\ref{sec:breadth}) "
        r"could in principle reflect how participants use the response scale rather than genuine "
        r"burden. We checked this in two ways.",
        r"",
        r"\paragraph{The outliers report genuine distress, not a fixed answering style.} A "
        r"well-documented threat to validity is the ``extreme responder''---a participant who "
        r"habitually marks the most extreme option on every question, out of haste or a fixed answering "
        r"style rather than from what the question asks "
        r"\citep{meade2012careless,vanvaerenbergh2013response}. To separate genuine distress from this "
        r"habit we summarized each participant in two ways. \emph{Content extremity} captures "
        r"\emph{what} a person reported: how far, on average, their answers sit from the typical "
        r"response (their mean absolute standardized response), so it is high for someone who genuinely "
        r"endorses more burden than most people do. \emph{Endpoint use}, a standard "
        r"extreme-response-style index \citep{vanvaerenbergh2013response}, captures \emph{how} a person "
        r"answered: how often they pick the very end of the scale---whatever the question, and in either "
        r"direction---so it is high for a content-blind habit of always choosing the most extreme option "
        r"and says nothing about what was actually reported.",
        r"",
        r"We then related each participant's outlier score---their averaged Mahalanobis percentile "
        r"across the six encoders---to both measures across the cohort. Were the outliers careless "
        r"extreme responders, the score would climb with endpoint use, but it does not. It correlates "
        r"strongly with content extremity (Spearman $\rho=0.75$, 95\% CI $[0.74,\,0.76]$, with outliers "
        r"averaging $1.34$ vs.\ cohort $0.79$) yet is, if anything, \emph{negatively} related to "
        r"endpoint use ($\rho=-0.34$, 95\% CI $[-0.35,\,-0.33]$), on which the outliers ($0.48$) are "
        r"indistinguishable from the cohort ($0.48$). Ranking everyone by endpoint use alone recovers "
        r"only $10$ of the $584$, so the screen is not simply re-ranking habitual endpoint markers.",
        r"",
        r"\paragraph{The screen is not a relabeled severity sum.} The Mahalanobis "
        r"distance could in principle just rediscover the participants with the highest average burden, "
        r"so we ranked everyone by that same content extremity and asked how many of the consensus set "
        r"it recovers: fewer than half ($280/584$). The screen therefore does not reduce to ``who "
        r"deviates most'' on a single flat average of the items. As \S\ref{sec:supp_configural} shows, "
        r"what sets it apart is that it integrates and standardizes the responses component by "
        r"component, not that it exploits any configuration across components.",
        r"",
    ])


# BFI-2-XS: our construct (= validated trait domain), the item's published facet, and reverse-key.
# Domains/facets follow the BFI-2-XS structure (Soto & John 2017, cite bfi2xs_soto2017).
BFI_RECOVERY = [
    ("Extraversion",          "Sociability",            "bfi2xs_1",  True),
    ("Extraversion",          "Assertiveness",          "bfi2xs_6",  False),
    ("Extraversion",          "Energy level",           "bfi2xs_11", False),
    ("Agreeableness",         "Compassion",             "bfi2xs_2",  False),
    ("Agreeableness",         "Respectfulness",         "bfi2xs_7",  True),
    ("Agreeableness",         "Trust",                  "bfi2xs_12", False),
    ("Conscientiousness",     "Organization",           "bfi2xs_3",  True),
    ("Conscientiousness",     "Productiveness",         "bfi2xs_8",  True),
    ("Conscientiousness",     "Responsibility",         "bfi2xs_13", False),
    ("Negative emotionality", "Anxiety",                "bfi2xs_4",  False),
    ("Negative emotionality", "Depression",             "bfi2xs_9",  False),
    ("Negative emotionality", "Emotional volatility",   "bfi2xs_14", True),
    ("Open-mindedness",       "Aesthetic sensibility",  "bfi2xs_5",  False),
    ("Open-mindedness",       "Intellectual curiosity", "bfi2xs_10", True),
    ("Open-mindedness",       "Creative imagination",   "bfi2xs_15", False),
]


# --- per-instrument construct accounting (Supplement S2) -------------------------------
# Each source instrument's published structure and the construct(s) we assign its items to.
# Construct lists and item counts are derived from construct_map_151.csv so they cannot drift;
# the published-structure text and category are editorial.
INSTRUMENT_ACCOUNTING = [
    # csv instrument key, display name, published structure, category
    ("BFI-2-XS",       "BFI-2-XS", "Five trait factors, three facets each",              "recovered"),
    ("nds (adapted)",  "NDS",      "Perceived neighborhood disorder",                    "unidim"),
    ("ACE-BRFSS",      "ACE",      "Childhood adversity index",                          "unidim"),
    ("EDS",            "EDS",      "Everyday discrimination",                            "unidim"),
    ("UCLA-V3",        "ULS-8",    "Loneliness",                                         "unidim"),
    ("DMS",            "DMS",      "Discrimination in medical settings",                 "unidim"),
    ("GAD-7",          "GAD-7",    "Generalized anxiety",                                "unidim"),
    ("BMMRS-DSE",      "DSES",     "Daily spiritual experience",                         "unidim"),
    ("scns (adapted)", "SCNS",     "Neighborhood social cohesion",                       "unidim"),
    ("HVS",            "HVS",      "Food insecurity",                                    "unidim"),
    ("PSS-10",         "PSS-10",   "Helplessness and self-efficacy factors",             "retained"),
    ("MOS-SS",         "mMOS-SS",  "Emotional-informational and instrumental support",   "retained"),
    ("ASRS-v1.1",      "ASRS",     "Inattention and hyperactivity",                      "retained"),
    ("PROMIS-Global",  "PROMIS",   "Physical and mental health",                         "retained"),
    ("MHQ",            "MHQ",      "Multi-disorder screening battery",                   "composite"),
    ("CIDI-5",         "WMH-CIDI", "Diagnostic screening modules",                       "composite"),
    ("PHQ-9",          "PHQ-9",    "Depression severity, ninth item screens suicidal ideation", "composite"),
    ("single item",    "---",      "No parent scale",                                    "single"),
]
_ACCOUNTING_CATS = [
    ("recovered", "Multidimensional scale, factor structure recovered"),
    ("unidim",    "Unidimensional scale, one construct"),
    ("retained",  "Multidimensional scale retained as one construct"),
    ("composite", "Composite screener, items assigned by topic"),
    ("single",    "Standalone single items"),
]


def _constructs_by_instrument():
    """instrument key -> (n_items, [constructs ordered by count desc]) from construct_map_151.csv."""
    counts = {}
    with open(os.path.join(HERE, "construct_map_151.csv")) as f:
        for r in csv.DictReader(f):
            counts.setdefault(r["instrument"], []).append(r["construct"])
    out = {}
    for inst, cons in counts.items():
        order = sorted(set(cons), key=lambda c: (-cons.count(c), c))
        out[inst] = (len(cons), order)
    return out


def table_construct_accounting():
    """Supplemental table + text: how each source instrument's published structure maps to the
    constructs used here. The section heading is emitted by main()."""
    by_inst = _constructs_by_instrument()
    lines = [
        r"Each of the 151 items was assigned to one construct by reading its prompt, then checked "
        r"against the published structure of its source instrument. Table~\ref{tab:construct_accounting} "
        r"accounts for every instrument. The BFI-2-XS is genuinely multidimensional, and our "
        r"content-based labels recover its five-factor structure (below). Thirteen scales each measure a "
        r"single attribute and map to one construct---nine unidimensional scales directly, and four "
        r"scales that carry published sub-factors which we retain as one broader construct (perceived "
        r"stress, perceived social support, adult ADHD, and self-rated health), because that broader "
        r"attribute is the unit relevant to the cross-instrument analysis. The two composite screeners, "
        r"the UK Biobank Mental Health Questionnaire and the WMH-CIDI, are batteries whose items each "
        r"screen a different topic, so we assign items by topic rather than to a single factor. The "
        r"PHQ-9 is a depression scale whose ninth item screens suicidal ideation, which we assign to "
        r"suicidality. The 13 standalone items belong to no scale and were assigned individually from "
        r"their prompts.",
        r"",
        r"\begin{table}[H]",
        r"\centering",
        r"\caption{\textbf{Each construct's grounding in its source instrument.} For every source "
        r"instrument, its published structure and the construct(s) its items are assigned to in this "
        r"analysis. Construct lists and item counts are taken directly from the item-level assignment "
        r"(Supplementary Data~1). ``Items'' is the number of the instrument's items in the 151-item "
        r"set.}\label{tab:construct_accounting}",
        r"\small",
        r"\begin{tabular}{@{}l c p{4.1cm} p{5.4cm}@{}}",
        r"\toprule",
        r"Instrument & Items & Published structure & Construct(s) in this analysis \\",
        r"\midrule",
    ]
    first = True
    for cat, catlabel in _ACCOUNTING_CATS:
        if not first:
            lines.append(r"\midrule")
        first = False
        lines.append(r"\multicolumn{4}{@{}l}{\textit{%s}} \\[1pt]" % catlabel)
        for key, disp, struct, c in INSTRUMENT_ACCOUNTING:
            if c != cat:
                continue
            n, cons = by_inst[key]
            cons_str = ", ".join(tex(x) for x in cons)
            lines.append(f"{tex(disp)} & {n} & {tex(struct)} & {cons_str} \\\\")
    lines += [r"\bottomrule", r"\end{tabular}", r"\end{table}", ""]
    return "\n".join(lines)


def crossinstrument_note():
    """Closing paragraph for S2: the constructs that pool items across instruments."""
    return "\n".join([
        r"Six constructs draw items from more than one instrument: depression (PHQ-9 and the MHQ), "
        r"psychosis (the WMH-CIDI and the MHQ), generalized anxiety (GAD-7 and a standalone item), "
        r"suicidality (PHQ-9 and standalone items), life-threatening events (the MHQ and the WMH-CIDI), "
        r"and neighborhood disorder and safety (the NDS and a standalone item). Pooling items that "
        r"different instruments phrase separately is deliberate, and it is the cross-instrument grouping "
        r"the concentrated components recover in the main-text construct analysis "
        r"(Section~\ref{sec:meaningful}).",
        r"",
    ])


def table_bfi_recovery(prompts):
    """Supplemental table + text: the content-based constructs recover the BFI-2-XS factors.
    The section heading is emitted by main()."""
    lines = [
        r"The BFI-2-XS is the one instrument whose full multidimensional factor structure we recover. "
        r"Its 15 items are organized into five validated trait domains, three facets each "
        r"\citep{bfi2xs_soto2017}. Reading item text alone, our assignment places all 15 items in the "
        r"correct domain (Table~\ref{tab:bfi_recovery}), and the three items we assign to each domain "
        r"span its three distinct published facets, so the content-based labels reproduce the "
        r"instrument's factor structure (15 of 15).",
        r"",
        r"\begin{table}[H]",
        r"\centering",
        r"\caption{\textbf{The content-based constructs recover the BFI-2-XS factor structure.} "
        r"All 15 Big Five Inventory--2 Extra-Short Form items, grouped by the construct we assigned "
        r"from the item text. Each construct corresponds exactly to one of the five validated "
        r"BFI-2-XS trait domains, and the three items per domain span its three published facets "
        r"\citep{bfi2xs_soto2017}. ``(R)'' marks reverse-keyed items.}\label{tab:bfi_recovery}",
        r"\small",
        r"\begin{tabular}{@{}l l p{8.2cm}@{}}",
        r"\toprule",
        r"Construct (= BFI-2 domain) & Published facet & Item text \\",
        r"\midrule",
    ]
    prev = None
    for construct, facet, it, rev in BFI_RECOVERY:
        if construct != prev:
            if prev is not None:
                lines.append(r"\midrule")
            prev, ccell = construct, tex(construct)
        else:
            ccell = ""
        txt = tex(prompts[it].rstrip(".")) + (r"~(R)" if rev else "")
        lines.append(f"{ccell} & {tex(facet)} & \\footnotesize {txt} \\\\")
    lines += [r"\bottomrule", r"\end{tabular}", r"\end{table}", ""]
    return "\n".join(lines)


def table_s2(k_of):
    lines = [
        r"\begin{table}[H]",
        r"\centering",
        r"\caption{\textbf{The six sentence encoders.} Hugging Face model identifier, embedding "
        r"dimension $D$, number of retained components $k$ (parallel analysis), and citation. "
        r"Each item's prompt text is embedded with each encoder, yielding the item-embedding "
        r"matrix $M\in\mathbb{R}^{151\times D}$.}\label{tab:encoders}",
        r"\small",
        r"\begin{tabular}{llccl}",
        r"\toprule",
        r"Encoder & Hugging Face identifier & $D$ & $k$ & Reference \\",
        r"\midrule",
    ]
    for name, hf, dim, key in ENCODERS:
        cite = r"\citep{" + key + "}"
        lines.append(f"{tex(name)} & \\texttt{{\\small {tex(hf)}}} & {dim} & {k_of[name]} & {cite} \\\\")
    lines += [r"\bottomrule", r"\end{tabular}", r"\end{table}", ""]
    return "\n".join(lines)


def table_frames():
    """Section + table: the 13 syntax families used in the content-vs-syntax test."""
    DISPLAY = {"UCLA-V3": "ULS-8", "CIDI-5": "WMH-CIDI", "MOS-SS": "mMOS-SS", "ASRS-v1.1": "ASRS",
               "BMMRS-DSE": "BMMRS", "nds (adapted)": "NDS", "scns (adapted)": "SCNS",
               "PROMIS-Global": "PROMIS", "single item": "single items"}
    lines = [
        r"Whether the embedding follows semantic content or syntax can only be tested by holding one fixed "
        r"and varying the other, which requires knowing each item's \emph{syntax}---the "
        r"reusable scaffold with a content slot---separately from its semantic content. We grouped the 151 "
        r"items into \textbf{13 syntax families} (Table~\ref{tab:frames}), labeling a family "
        r"\emph{content-bearing} when its template itself names a construct (so semantic content and syntax "
        r"cannot be separated within it) and \emph{neutral} otherwise. Two cross-instrument syntax "
        r"overlaps that a naive ``different-instrument'' donor rule would miss are collapsed here: "
        r"the Everyday and Medical-Settings discrimination scales share one syntax, as do PHQ-9 and "
        r"GAD-7. In the test (\S\ref{sec:contentgrammar}), the syntax variant draws its donors only "
        r"from neutral families, and content variants are reported split by whether the seed's syntax "
        r"is content-bearing.",
        r"",
        r"\begin{table}[H]",
        r"\centering",
        r"\caption{\textbf{The 13 syntax families.} Each of the 151 items belongs to one "
        r"syntax family. A family is \emph{content-bearing} (CB) when the template itself names a "
        r"construct, \emph{neutral} otherwise. EDS+DMS and PHQ-9+GAD-7 each collapse to a single "
        r"shared syntax.}\label{tab:frames}",
        r"\small",
        r"\begin{tabular}{@{}l l r p{4.6cm} p{4.0cm}@{}}",
        r"\toprule",
        r"Frame family & Type & $n$ & Source instruments & Example template \\",
        r"\midrule",
    ]
    prev_cb = None
    for fam in FRAME_ORDER:
        cb = FRAME_CB[fam]
        if prev_cb is not None and cb != prev_cb:
            lines.append(r"\midrule")
        prev_cb = cb
        items = FRAME_ITEMS[fam]
        insts = "+".join(sorted({DISPLAY.get(FRAME_INST[it], FRAME_INST[it]) for it in items}))
        lines.append(f"{tex(fam.replace('_', ' '))} & {'CB' if cb else 'neutral'} & {len(items)} & "
                     f"\\footnotesize {tex(insts)} & \\footnotesize {FRAME_TEMPLATE[fam]} \\\\")
    lines += [r"\bottomrule", r"\end{tabular}", r"\end{table}", ""]
    return "\n".join(lines)


def table_s3():
    """Table S3: the 600 content-vs-syntax variants + a-priori syntax classification."""
    with open(VARIANTS_CSV) as f:
        rows = list(csv.DictReader(f))
    # group each seed's 20 variants together (10 Cell A, 10 Cell B); seeds ordered by name.
    rows.sort(key=lambda r: (r["seed_item"], r["cell"], r["variant_id"]))
    n_cb = len({r["seed_item"] for r in rows if r["seed_item"] in CONTENT_BEARING})
    lines = [
        r"\begin{small}",
        r"\begin{longtable}{@{}l c p{2.5cm} p{2.3cm} c p{5.2cm}@{}}",
        r"\caption{\textbf{The 600 content-versus-syntax variants} (\S\ref{sec:contentgrammar}). "
        r"Each of 30 seed items (stratified across the 13 syntax families, Table~\ref{tab:frames}) "
        r"was rewritten into 10 \emph{Cell A} variants---same semantic content, a different item's "
        r"syntax---and 10 \emph{Cell B} variants---same syntax, a different item's "
        r"semantic content. \emph{Donor} is the item supplying the transferred syntax (Cell A) or semantic content "
        r"(Cell B). By construction every donor differs from its seed in both syntax family and "
        r"construct, and Cell A donors are restricted to neutral syntax. \emph{CB} marks the "
        r"content-bearing-syntax seeds, reported separately in the Cell B test. Variants are "
        r"synthetic test items generated by \texttt{claude-opus-4-8} (interactive) and were never "
        r"deployed. Some Cell B variants place semantic content into an unrelated syntax as an artifact of "
        r"the recombination.}\label{tab:variants}\\",
        r"\toprule",
        r"ID & Cell & Seed item & Donor & CB & Rewritten prompt \\",
        r"\midrule",
        r"\endfirsthead",
        r"\multicolumn{6}{l}{\small\itshape Table S3 continued}\\",
        r"\toprule ID & Cell & Seed item & Donor & CB & Rewritten prompt \\ \midrule",
        r"\endhead",
        r"\midrule \multicolumn{6}{r}{\small\itshape continued on next page}\\",
        r"\endfoot",
        r"\bottomrule",
        r"\endlastfoot",
    ]
    prev = None
    for r in rows:
        if r["seed_item"] != prev:                 # rule between seed blocks
            if prev is not None:
                lines.append(r"\addlinespace[2pt]")
            prev = r["seed_item"]
        cb = r"$\bullet$" if r["seed_item"] in CONTENT_BEARING else ""
        lines.append(
            f"\\texttt{{\\footnotesize {tex(r['variant_id'])}}} & {tex(r['cell'])} & "
            f"\\texttt{{\\scriptsize {tex(r['seed_item'])}}} & "
            f"\\texttt{{\\scriptsize {tex(r['donor_item'])}}} & {cb} & "
            f"\\footnotesize {tex(r['rewritten_prompt'])} \\\\")
    lines += [r"\end{longtable}", r"\end{small}", ""]
    return "\n".join(lines), len(rows), n_cb


def gen_prompts_box():
    """Verbatim generation templates for the content-vs-syntax sub-study (reproducibility).

    Imported from generate_variants.py so the published prompts cannot drift from the code.
    """
    import textwrap
    from generate_variants import SYS, CELL_A, CELL_B
    clean = lambda t: t.replace("{{", "{").replace("}}", "}")   # show literal JSON braces

    def wrap(t, w=88):                                          # verbatim does not wrap; fold long lines
        out = []
        for line in clean(t).split("\n"):
            out.extend(textwrap.wrap(line, w) if len(line) > w else [line])
        return "\n".join(out)
    parts = [
        r"\paragraph{Content-versus-syntax generation prompts.}",
        r"The 3{,}020 content-versus-syntax variants were generated by \texttt{claude-opus-4-8} "
        r"(interactive) following the rewriting instructions below---one per variant type. Each "
        r"variant pairs a seed with a donor chosen by the deterministic rule of \S\ref{sec:supp_frames}: "
        r"the donor differs from the seed in \emph{both} syntax family and construct, and for the "
        r"syntax variant the donor's syntax is neutral. The full seed-to-donor manifest, every "
        r"rewritten prompt, and all 3{,}020 variants are provided as \textbf{Supplementary Data 2} "
        r"(\texttt{content\_grammar\_variants/}: \texttt{manifest.csv}, \texttt{generated\_items\_v2.csv}, "
        r"and the batch \texttt{.jsonl} files).",
        r"\begin{footnotesize}",
        r"\textit{System prompt:}",
        r"\begin{verbatim}",
        wrap(SYS),
        r"\end{verbatim}",
        r"\textit{Cell A --- content preserved, grammatical frame transferred:}",
        r"\begin{verbatim}",
        wrap(CELL_A),
        r"\end{verbatim}",
        r"\textit{Cell B --- frame preserved, content swapped:}",
        r"\begin{verbatim}",
        wrap(CELL_B),
        r"\end{verbatim}",
        r"\end{footnotesize}",
        "",
    ]
    return "\n".join(parts)


def table_pselect():
    """Per-encoder selected projection from the random-restart selection (selection_best.json)."""
    sel = json.load(open(PSELECT_JSON))
    lines = [
        r"\begin{table}[H]",
        r"\centering",
        r"\caption{\textbf{The selected projection, by encoder.} For each encoder we fit the "
        r"concentrated projection from 100 random orthonormal initializations at each "
        r"$\lambda_{\mathrm{score}}\in\{0.01,0.05,0.1,0.5,1\}$ and retain the fit that maximizes "
        r"median core energy subject to $W$ orthonormal (largest off-diagonal $|\cos|\le0.01$ to three decimals) and "
        r"explained variance within $1\%$ of the PCA maximum (Methods). For each encoder we list the "
        r"selected penalty weight $\lambda_{\mathrm{score}}$ and initialization seed, and the selected "
        r"projection's median core energy, mean core size, and VE, each against the dense PCA "
        r"baseline ($\lambda_{\mathrm{score}}=0$). Selection concentrates each component onto a "
        r"smaller core carrying more of its weight, at explained variance within $1\%$ of the "
        r"PCA maximum.}\label{tab:pselect}",
        r"\small",
        r"\begin{tabular}{lcccccc}",
        r"\toprule",
        r"Encoder & $k$ & $\lambda_{\mathrm{score}}$ & seed & Core energy & Mean core & VE \\",
        r" & & & & (sel.\,/\,dense) & (sel.\,/\,dense) & (sel.\,/\,dense) \\",
        r"\midrule",
    ]
    for name, _hf, _dim, _key in ENCODERS:
        s = sel[name]
        lines.append(
            f"{tex(name)} & {s['k']} & {s['ls']:g} & {s['seed']} & "
            f"{s['coreE']:.3f}\\,/\\,{s['dense_coreE']:.3f} & "
            f"{s['mean_core']:.2f}\\,/\\,{s['dense_mean_core']:.2f} & "
            f"{s['ve']:.3f}\\,/\\,{s['dense_ve']:.3f} \\\\")
    lines += [r"\bottomrule", r"\end{tabular}", r"\end{table}", ""]
    return "\n".join(lines)


def figure_lambda_selection():
    """Supplemental Figure: selection + concentration achieved (moved from the main text)."""
    return "\n".join([
        r"\begin{figure}[H]",
        r"  \centering",
        r"  \includegraphics[width=\textwidth]{figures/fig_lambda_selection.png}",
        r"  \caption{\textbf{Selecting the concentrated projection, and the concentration it "
        r"achieves} (all six encoders). (a) For each encoder, the median core energy of the $100$ "
        r"random-initialization fits at its selected penalty weight $\lambda_{\mathrm{score}}$ "
        r"(dots). We retain the fit with the largest median core energy (star), well above the "
        r"dense-PCA floor (grey bar). (b) The basis $W$ stays orthonormal across the swept window "
        r"(green, maximum off-diagonal $\lvert\cos\rvert\le0.01$ to three decimals, dashed line) and "
        r"tilts away from orthogonality only for much larger $\lambda$ (collapse, red), so "
        r"concentration cannot be pushed without bound (the full four-panel diagnostic sweep is "
        r"Figure~\ref{fig:sweep_full}). (c) Per-component core energy---the fraction of a "
        r"component's weight on its core---rises from a median of $\approx0.30$ under dense PCA to "
        r"$\approx0.56$ under the selected projection, pooled over encoders. (d) The "
        r"participation-ratio core (Methods)---the self-consistent set of items a component rests "
        r"on and that the participant-level audit traces back to---shrinks from a median of "
        r"$\approx8$ items under dense PCA to $\approx5$.}",
        r"  \label{fig:lambda}",
        r"\end{figure}",
        "",
    ])


def figure_s1():
    """Supplemental Figure S1: the full lambda sweep (made by result2_figures.py)."""
    return "\n".join([
        r"\begin{figure}[H]",
        r"  \centering",
        r"  \includegraphics[width=\textwidth]{figures/figS_lambda_sweep.png}",
        r"  \caption{\textbf{The full sparsity-weight sweep} (all six encoders). "
        r"Figure~\ref{fig:lambda}b shows the orthogonality curve (C) over the swept window. "
        r"We fit the projection across $\lambda$ and select within the swept window "
        r"$\lambda_{\mathrm{score}}\in\{0.01,\dots,1\}$ (green); past it into collapse (red, "
        r"$\lambda\ge5$) the orthonormality term is overwhelmed. "
        r"(A) the subspace variance VE@$\theta_0$ holds flat then collapses. (B) "
        r"$\|WW^\top-I\|_F$ rises gently in the window (component norms drift to $\approx0.98$) "
        r"before blowing up. (C) the directions stay orthogonal (max off-diagonal $|\cos|"
        r"\approx0.01$) until collapse. (D) the median participation ratio falls to its "
        r"minimum in the window and rebounds (degenerately) in collapse. The genuine "
        r"scikit-learn PCA reference is starred in panels A and D (and is exactly $0$ in B and C).}",
        r"  \label{fig:sweep_full}",
        r"\end{figure}",
        "",
    ])


def figure_s_jaccard():
    """Supplemental Figure: pairwise outlier-set Jaccard vs percentile threshold."""
    return "\n".join([
        r"\begin{figure}[H]",
        r"  \centering",
        r"  \includegraphics[width=0.85\textwidth]{figures/figS_jaccard_threshold.png}",
        r"  \caption{\textbf{Cross-encoder outlier agreement versus threshold.} Pairwise Jaccard "
        r"overlap of the flagged-outlier sets for each of the 15 encoder pairs (coloured lines) and "
        r"their mean (dashed), as the empirical percentile threshold that defines an outlier varies. "
        r"Mean agreement stays far above the independence null across the whole sweep with no knee "
        r"(mean pairwise Jaccard ${\approx}0.61$, about $23\times$ the null) and declines only "
        r"gently toward the most extreme cutoffs. We therefore screen at the 95th percentile (top "
        r"$5\%$) per encoder as the canonical top tail, and take the consensus across all six for "
        r"downstream analyses (\S\ref{sec:detection}).}",
        r"  \label{fig:jaccard_threshold}",
        r"\end{figure}",
        "",
    ])


def figure_upset_supp():
    """Supplemental Figure: the construct combinations of four or more constructs, the tail the
    main-text UpSet (Figure~\\ref{fig:attribution}b) leaves out. Produced on the All of Us Workbench
    by examples/allofus/outlier_construct_upset.py as fig_outlier_upset_supp.png.
    PLACEHOLDER IMAGE -- replace figures/fig_outlier_upset_supp.png with the Workbench export."""
    return "\n".join([
        r"\begin{figure}[H]",
        r"  \centering",
        r"  \includegraphics[width=0.85\textwidth]{figures/fig_outlier_upset_supp.png}",
        r"  \caption{\textbf{Construct combinations of four or more constructs.} The consensus "
        r"outliers carried by four or more constructs, each bar one combination with height the "
        r"number of outliers who form it, continuing the single, pair, and triple combinations of "
        r"main-text Figure~\ref{fig:attribution}b. Constructs carry a participant on the criterion "
        r"of \S\ref{sec:nocutoff}. These combinations are the long tail of the attribution, so most "
        r"outliers are accounted for by the smaller combinations shown in the main text.}",
        r"  \label{fig:upset_supp}",
        r"\end{figure}",
        "",
    ])


def table_kselection():
    """Table: silhouette-selected number of item clusters, method x encoder."""
    order = ["bge-m3__k25_ls0", "bge-large-en-v1", "all-mpnet-base-v2__k24_ls0",
             "e5-large-v2__k23_ls0", "gte-large-en-v1", "qwen3-embedding-0"]
    disp = ["bge-m3", "bge-large", "all-mpnet", "e5-large", "gte-large", "qwen3"]
    mlabel = {"average": "Average linkage", "complete": "Complete linkage",
              "ward": "Ward", "spectral": "Spectral"}
    K = {}
    for r in csv.DictReader(open(KSEL_CSV)):
        K[(r["method"], r["encoder"])] = int(r["k_silhouette"])
    rows = []
    for m in ["average", "complete", "ward", "spectral"]:
        ks = [K[(m, e)] for e in order]
        srt = sorted(ks)
        med = srt[len(srt) // 2] if len(srt) % 2 else (srt[len(srt) // 2 - 1] + srt[len(srt) // 2]) // 2
        cells = " & ".join(str(k) for k in ks)
        rows.append(f"{mlabel[m]} & {cells} & {max(ks) - min(ks)} & {med} \\\\")
    return "\n".join([
        r"\begin{table}[H]", r"\centering",
        r"\caption{\textbf{Silhouette-selected number of item clusters, by method and encoder.} "
        r"Number of clusters $k$ maximizing the mean silhouette (cosine distance) when the 151 "
        r"items are clustered by the cosine geometry of the concentrated projection. Spectral "
        r"clustering gives the most consistent $k$ across encoders (range~5, versus 15--19 for the "
        r"others), motivating its use in the main text. All methods place $k$ in the 18--38 range, "
        r"near the $\sim$30 a-priori constructs (main-text Table~\ref{tab:constructs}).}"
        r"\label{tab:kselection}",
        r"\begin{tabular}{lcccccc|cc}", r"\hline",
        r"Method & " + " & ".join(disp) + r" & range & median \\", r"\hline",
        "\n".join(rows), r"\hline", r"\end{tabular}", r"\end{table}"])


def content_results_para():
    """Content-versus-syntax results narrative + numbers (moved from main text 3.3.2)."""
    return "\n".join([
        r"\paragraph{Content-versus-syntax results.}",
        r"Both variant types sit with their content source, not their syntax source "
        r"(Figure~\ref{fig:contentgrammar}). A syntax variant stays close to its seed---the item whose "
        r"semantic content it kept---and far from the donor whose syntax it took (panel a), and a "
        r"content variant does the reverse, moving to the donor whose semantic content it carries "
        r"(panel b). Reading each variant against its two sources, the content source wins by a wide "
        r"margin (gte-large median cosine $0.75$ versus $0.26$ for syntax variants and $0.72$ versus "
        r"$0.26$ for content variants), and the content$-$syntax gap stays positive for every one of the "
        r"six encoders, from $+0.56$ (all-mpnet) to $+0.20$ (e5-large, panel c). Semantic content "
        r"survives a wholesale change of syntax, and follows when it is swapped in. The one systematic "
        r"exception is content-bearing syntax---templates that themselves name a construct, such as "
        r"``\dots in my neighborhood'' or ``During your first 18 years of life,'' where semantic content "
        r"and syntax cannot be separated by design. For content variants built on such syntax the two "
        r"sources pull almost equally (median cosine $0.61$ versus $0.48$), a graceful near-tie in which "
        r"semantic content still scores marginally higher rather than a collapse.",
        "",
    ])


def figure_content_grammar():
    """Supplemental Figure: content-versus-syntax cosine plot (moved from the main text)."""
    return "\n".join([
        r"\begin{figure}[H]",
        r"  \centering",
        r"  \includegraphics[width=\textwidth]{figures/fig_content_grammar.png}",
        r"  \caption{\textbf{Semantic content, more than syntax, drives the grouping.} Counterfactual "
        r"variants recombine the items' own semantic content and syntax: a \emph{syntax variant} keeps "
        r"the seed's semantic content in a donor's syntax and a \emph{content variant} keeps the seed's "
        r"syntax with a donor's semantic content, every donor differing from the seed in both. Each "
        r"variant is projected onto the fixed basis (the selected operating basis, fit on the 151 real "
        r"items, never refit) and compared, by cosine, to its \emph{content source} and its "
        r"\emph{syntax source}. Box plots span 151 seeds and 3{,}020 variants across six encoders, "
        r"ordered by effect size. (a) Cosine to the \emph{seed}: syntax variants (semantic content "
        r"kept) stay high, content variants (semantic content swapped in) fall away, and content "
        r"variants on content-bearing syntax (jingle) remain partly tied to the seed. (b) Cosine to "
        r"the \emph{donor}, the mirror image. (c) Per encoder, the median cosine to each variant's "
        r"content source (black) and syntax source (grey), pooled over the clean variants (the jingle "
        r"case excluded). The gap between them---semantic content minus syntax---is positive for every "
        r"encoder, from $+0.56$ to $+0.20$, the margin by which semantic content prevails over syntax.}",
        r"  \label{fig:contentgrammar}",
        r"\end{figure}",
        "",
    ])


def table_core_examples():
    """Two contrasting cores, item by item (gte-large): C4, a broad disposition axis, and C3,
    the one core whose items come from six different constructs. Read from Supplementary Data 3
    (core_items_gte.csv, written by make_core_items_pool.py) so it cannot drift from the data."""
    path = os.path.join(SUPP, "core_items", "core_items_gte.csv")
    rows = [r for r in csv.DictReader(open(path)) if r["component"] in ("C3", "C4")]
    out = [
        r"\begin{table}[h]", r"\centering",
        r"\caption{\textbf{The core items of two contrasting components} (gte-large). "
        r"\textbf{C4} is a broad disposition axis: eight of its twelve items are personality "
        r"items, and the loneliness item it draws in (``I am an outgoing person'') asks about "
        r"extraversion, so the core groups items by what they ask rather than by the instrument "
        r"they belong to. \textbf{C3} is the one core whose items come from six different "
        r"constructs. Loading is the item's weight on the component, and sign is arbitrary up to "
        r"the direction of the axis. Every component's core is listed in \textbf{Supplementary "
        r"Data 3} (\texttt{core\_items\_gte.csv}).}",
        r"\label{tab:core_examples}", r"\small",
        r"\begin{tabular}{@{}llr p{7.4cm}@{}}", r"\toprule",
        r"Component & Construct & Loading & Item \\", r"\midrule",
    ]
    for comp in ("C4", "C3"):
        sub = [r for r in rows if r["component"] == comp]
        for j, r in enumerate(sub):
            lead = comp if j == 0 else ""
            prompt = (r["prompt"].replace("&", r"\&").replace("%", r"\%")
                      .replace("_", r"\_").replace("#", r"\#"))
            # straight double quotes -> LaTeX paired quotes (``opening'' / closing'')
            parts, opening = prompt.split('"'), True
            prompt = parts[0]
            for seg in parts[1:]:
                prompt += ("``" if opening else "''") + seg
                opening = not opening
            out.append(f"{lead} & {r['construct']} & {r['loading']} & \\footnotesize {prompt} \\\\")
        if comp == "C4":
            out.append(r"\midrule")
    out += [r"\bottomrule", r"\end{tabular}", r"\end{table}", ""]
    return "\n".join(out)


def table_barriers():
    """Table tab:barriers: all six healthcare-access barrier components, consensus outliers vs rest.
    Numbers from examples/allofus/outlier_ehr_underserved.py (p95/584, EHR-linked >=1-visit cohort;
    survey respondents outliers n=511, rest n=19,799)."""
    rows = [
        ("Any access barrier", "81.4", "44.1", "+37.3", "+34, +40", "1.84", "1.76, 1.93"),
        ("Cost barrier", "67.1", "27.8", "+39.3", "+35, +43", "2.41", "2.26, 2.57"),
        ("Structural delay (transport, time off work, childcare)", "51.9", "16.5", "+35.4", "+31, +40", "3.14", "2.87, 3.44"),
        ("Medication rationing to save money", "46.6", "22.7", "+23.9", "+20, +28", "2.05", "1.87, 2.26"),
        ("No usual source of care", "9.6", "3.7", "+5.9", "+4, +9", "2.61", "1.98, 3.44"),
        ("Care avoidance, provider race or religion", "31.3", "8.6", "+22.7", "+19, +27", "3.64", "3.18, 4.17"),
    ]
    lines = [
        r"\begin{table}[h]",
        r"\centering",
        r"\caption{\textbf{Healthcare-access barriers, consensus outliers versus the rest of the "
        r"EHR-linked cohort.} Among access-survey respondents (outliers $N=511$, rest $N=19{,}799$), the "
        r"percentage reporting each barrier, the difference in percentage points with Newcombe $95\%$ "
        r"confidence intervals, and the risk ratio with Katz log-method $95\%$ confidence intervals. "
        r"Every component is more common among the outliers. Brackets give the corresponding "
        r"confidence interval.}\label{tab:barriers}",
        r"\small",
        r"\begin{tabular}{@{}p{5.0cm}rrll@{}}",
        r"\toprule",
        r"Barrier & Outliers (\%) & Rest (\%) & Difference (pp) & Risk ratio \\",
        r"\midrule",
    ]
    for name, o, rest, d, dci, rr, rrci in rows:
        lines.append(f"{tex(name)} & {o} & {rest} & {d} [{dci}] & {rr} [{rrci}] \\\\")
    lines += [r"\bottomrule", r"\end{tabular}", r"\end{table}", ""]
    return "\n".join(lines)


def underserved_visits_section():
    """The visit-count completeness caveat + the supplement figure fig_ehr_visits_supp.png. Numbers
    from outlier_ehr_underserved.py (p95/584, EHR-linked >=1-visit cohort, N=20,972)."""
    return "\n".join([
        r"The self-reported access barriers (Table~\ref{tab:barriers}) and the emergency-department "
        r"reliance reported in the main text (\S\ref{sec:methods_ehr}) are the two signals we rely on. "
        r"Raw visit counts tell a consistent-looking but confounded story that we report here for "
        r"completeness. Median total visits fall across the outlier-score deciles, from $21$ in the "
        r"lowest to $1$ in the highest, and the age- and sex-adjusted rank regression is negative "
        r"($\beta=-0.026$, $95\%$ CI $[-0.040, -0.013]$, $p=1.5\times10^{-4}$). This does not survive "
        r"the two completeness checks. Adjusting additionally for the observation-window length removes "
        r"it ($\beta=-0.007$, $[-0.016, +0.003]$, $p=0.17$), and the visits-per-observation-year rate "
        r"shows no association with the score ($\beta=+0.004$, $[-0.011, +0.018]$, $p=0.64$). Because a "
        r"participant's observation window in All of Us is set largely by which sites contributed data "
        r"and when, the falling counts track how completely each record is populated rather than how "
        r"much care a person seeks (Figure~\ref{fig:ehr_visits}). Emergency-department reliance, a ratio, "
        r"is by construction free of this confound, and it rises with the score holding the total number "
        r"of visits, age, and sex fixed ($\beta=+0.052$, $[+0.044, +0.061]$, $p=3.7\times10^{-34}$).",
        r"",
        r"\begin{figure}[h]",
        r"  \centering",
        r"  \includegraphics[width=0.8\textwidth]{figures/fig_ehr_visits_supp.png}",
        r"  \caption{\textbf{Raw visit counts fall with the outlier score, but the per-year rate does "
        r"not.} Across deciles of the cross-instrument outlier score (EHR-linked cohort, $N=20{,}972$), "
        r"the median total number of electronic-health-record visits falls (left axis), while the median "
        r"visits per observation-year stays flat (right axis). The falling counts reflect shorter, less "
        r"complete records rather than reduced care-seeking.}",
        r"  \label{fig:ehr_visits}",
        r"\end{figure}",
        r"",
    ])


def configural_null_section():
    """Supplement: the multivariate screen integrates rather than detects (configural null).
    Numbers from examples/allofus/configural_test.py (p95/584, 6 encoders)."""
    return "\n".join([
        r"\section{The multivariate view integrates the responses rather than detecting "
        r"otherwise-invisible cases}\label{sec:supp_configural}",
        r"A natural question is whether the cross-instrument distance flags participants that "
        r"scale-by-scale analysis of the same responses could not. It does not, and we report this "
        r"directly. Standardizing every item across the cohort, every one of the $584$ consensus "
        r"outliers falls in the top $5\%$ of at least one single item and at least one single "
        r"construct, so each would be identifiable from an individual scale examined on its own. "
        r"Consistently, the full Mahalanobis distance ranks participants almost identically to a "
        r"diagonal distance that ignores cross-component covariance (Spearman $\rho = 0.84$ to "
        r"$0.89$ across the six encoders), and the outliers sit at the same extreme percentile on "
        r"both. The value of reading the instruments together is therefore not the detection of "
        r"otherwise-invisible cases but the integration of $151$ items into a single, coherent, "
        r"small group of the broadly and multiply burdened---who would otherwise be dispersed across "
        r"many separate scale-level flags---together with the attribution of each flag to its "
        r"driving items and the surfacing of adverse exposures, such as discrimination and "
        r"interpersonal violence, that carry no clinical cutoff.",
        r"",
    ])


def main():
    prompts, survey, cid, inst, vals, k_of, constr, domain = load_data()
    body = [
        r"% ============================================================================",
        r"%  Supplemental materials -- AUTO-GENERATED by examples/allofus/make_supp_tables.py",
        r"%  Do not edit by hand. Re-run the script to regenerate.",
        r"%  This is a FRAGMENT: \input by the standalone wrapper manuscript/supplement_main.tex,",
        r"%  which provides the documentclass, preamble, title, and bibliography.",
        r"% ============================================================================",
        r"\setcounter{section}{0}",
        r"\renewcommand{\thesection}{S\arabic{section}}",
        r"\setcounter{table}{0}",
        r"\renewcommand{\thetable}{S\arabic{table}}",
        r"\setcounter{figure}{0}",
        r"\renewcommand{\thefigure}{S\arabic{figure}}",
        r"",
        r"\section{Selected items, constructs, and domains}\label{sec:supp_items}",
        items_pointer(),                      # 151-item table shipped as items_full.csv (Supp. Data 1)
        r"\section{Construct labels and instrument structure}\label{sec:subscale_recovery}",
        table_construct_accounting(),         # Table S2: per-instrument construct accounting
        table_bfi_recovery(prompts),          # Table S3: BFI-2-XS factor recovery
        crossinstrument_note(),               # constructs pooled across instruments
        r"\section{Sentence encoders}\label{sec:supp_encoders}",
        table_s2(k_of),                       # Table S3: encoders
        r"\section{Selecting the projection}\label{sec:supp_sweep}",
        table_pselect(),                      # Table: per-encoder selected projection
        figure_lambda_selection(),            # Figure: selection + concentration (moved from main text)
        figure_s1(),                          # Figure: concentration vs lambda sweep
        r"\section{The items carrying each component}\label{sec:supp_cores}",
        table_core_examples(),                # Table: C4 (disposition axis) vs C3 (six constructs)
        r"\section{Syntax families}\label{sec:supp_frames}",
        table_frames(),                       # Table S4: the 13 syntax families
        r"\section{Semantic content versus syntax}\label{sec:supp_variants}",
        content_results_para(),               # results narrative + numbers (moved from main 3.3.2)
        figure_content_grammar(),             # Figure: content-vs-syntax cosine plot (moved from main)
        gen_prompts_box(),                    # generation templates (reproducibility); the full
                                              # 3,020 variants are released as jsonl, not tabulated

        r"\section{Cross-encoder outlier agreement}\label{sec:supp_jaccard}",
        figure_s_jaccard(),                   # Figure: pairwise Jaccard vs threshold
        r"\section{The outliers' breadth is not a response-style or severity artifact}"
        r"\label{sec:supp_robust}",
        robustness_section(),                 # response-style + severity checks moved from 3.4.2
        r"\section{Construct combinations of four or more constructs}\label{sec:supp_upset}",
        figure_upset_supp(),                  # Figure: the 4+ tail of the main-text UpSet (3.4.3)
        r"\section{The under-served group: barriers and the visit-count caveat}\label{sec:supp_visits}",
        table_barriers(),                     # Table: six access-barrier components, outliers vs rest
        underserved_visits_section(),         # visit-count completeness caveat + supplement figure
        configural_null_section(),            # multivariate integrates, does not detect the invisible
    ]
    with open(OUT, "w") as f:
        f.write("\n".join(body))
    csv_path = write_items_csv(prompts, cid, inst, vals, constr, domain)
    n_var = copy_variant_data()
    print(f"-> {os.path.normpath(OUT)}")
    print(f"-> {os.path.normpath(csv_path)}  (Supplementary Data 1)")
    print(f"-> {os.path.normpath(CGV_DST)}  (Supplementary Data 2: {n_var} variant files)")
    print(f"   ({len(prompts)} items; {len(ENCODERS)} encoders)")


if __name__ == "__main__":
    main()
```
