## Supplemental code for "Scoring respondents on a semantic frame of reference identifies a group that is burdened across many questionnaires and reaches care less easily": outlier_ehr_underserved.py

```
"""
Section 3.4.4  --  The flagged group is under-served by the health system.

STANDALONE Workbench script (runs on /home/jupyter, where the AoU CDR + the dense
ACIDD runs live).  The EHR tables queried here (visit_occurrence, condition_occurrence,
observation_period, ds_survey, person) are present in both Registered and Controlled
tiers -- confirm which tier this workspace uses (WORKSPACE_CDR: R... = Registered,
C... = Controlled) and match the manuscript wording to it.  Mirrors the standalone
style of the Section 3.4.3 scripts (outlier_construct_*.py); supersedes the Cell 4/4b
block in section3_4_results.py.

PURPOSE (framing -- read before editing prose)
----------------------------------------------
The outliers are derived from ONE thing: the embedding of survey-item responses.
Electronic health records (visits, conditions, ED encounters) are a SEPARATE data
stream that never touched the screen.  If that independent stream shows the flagged
group is under-served, that is a real gap in CARE -- a system-level insight, not a
validation of our method.  We do NOT frame this as "the outliers are real / not an
artifact"; we frame it as "an independent modality reveals a modifiable gap".

The finding: the flagged group reports far more barriers to care, and independently, when they
DO reach the system a larger share of their visits is the ED (crisis door, not routine door).
Raw routine-visit counts also fall with the score, but that tracks EHR record completeness
(window-adjusted and per observation-year the gap is null) -> reported in the supplement with
that caveat, NOT as a per-time engagement claim.

Evidence hierarchy (decision 2026-07-21, option 3):
  MAIN, self-report (mechanism)  access/cost barriers + care avoidance for provider race/religion
                                 (-> S3.4.3 discrimination).
  MAIN, orthogonal EHR (clean)   ED share of visits rises -- a ratio, so independent of record window.
  SUPPLEMENT, orthogonal EHR     raw visit counts fall, but this is record-completeness (caveated).
  DROPPED                        multimorbidity (fewer conditions = same completeness confound).

Reporting altitude (user directive 2026-07-21): the manuscript body states the
headline and explains only the numbers it keeps; the *how* and the detailed
numbers/CIs live in Methods / Supplement.  This script prints everything; the
manuscript picks the headline from it.

RUN NOTE (2026-07-21, clean >=1-visit cohort N=20,972; 538/584 outliers linked)
-------------------------------------------------------------------------------
observation_period exists for ALL 22,332 responders -> NOT an EHR-linkage marker; EHR linkage is
>=1 visit (== the manuscript's 20,972 / 94%).  Result (option 3 = DEMOTE visits to supplement):
 - Barriers (self-report) reproduce the pre-registered RRs exactly: any 1.84, cost 2.41,
   care-avoid 3.64.  MAIN-TEXT pillar.
 - Engagement beta=-0.026 (p=1.5e-4) reproduces the manuscript's -0.03 BUT collapses under window
   adjustment (p=0.17) and per observation-year (p=0.64) -> the raw "visits fall 21->1" is record
   completeness, not per-time under-engagement.  -> SUPPLEMENT, with the caveat.
 - ED reliance (ED/total visits, a ratio -> window-free) is the clean orthogonal EHR signal.  MAIN.
 - Multimorbidity goes the wrong way (fewer conditions, completeness confound) -> DROPPED.

EMITS
-----
  outputs_dense/underserved_stats.json   every number + CI
  fig_ehr_gradient.png                   MAIN: barriers + ED share of visits, vs mean Mahalanobis pctile
  fig_ehr_visits_supp.png                SUPPLEMENT: raw visits fall vs flat per-year rate (caveat)
"""

# %% Cell 0 -- setup: dense scores -> distance matrix D -> consensus + continuous score
import glob, os, math, json
import numpy as np, pandas as pd
from scipy.stats import spearmanr
import statsmodels.formula.api as smf
import matplotlib.pyplot as plt
from google.cloud import bigquery

RUNS_ROOT = "outputs_dense"        # dense run dirs k0_<model>/
PATTERN   = "k0_*"
ID_COL    = "id"
DIST_COL  = "Mahalanobis_Dist"
EXCLUDE   = {"parallel"}           # k0_parallel = dense bge-m3 baseline, not a 7th embedder
PCTILE    = 95.0                   # p95 -> N=584 consensus (matches section3_4_results after migration)

# CDR dataset.  AoU normally sets WORKSPACE_CDR, but a kernel that booted before the
# environment was injected won't have it (a fresh Terminal will -- `echo $WORKSPACE_CDR`).
# CDR_OVERRIDE is this workspace's CDR; BLANK IT before syncing to reproduce_allofus (it is
# workspace-specific), where it should fall back to WORKSPACE_CDR like every other AoU script.
CDR_OVERRIDE = "wb-affable-acorn-7941.R2024Q3R8"
CDR = os.environ.get("WORKSPACE_CDR") or CDR_OVERRIDE
if not CDR:
    raise RuntimeError(
        "No CDR dataset string. From a standard AoU notebook run print(os.environ['WORKSPACE_CDR']), "
        "then set CDR_OVERRIDE above and re-run.")
client = bigquery.Client()
def Q(sql): return client.query(sql).to_dataframe()

# ---- proportion-CI helpers (identical to section3_4_results.py) ----
def _wilson(k, n, z=1.959963985):
    p = k / n; d = 1 + z*z/n; ctr = (p + z*z/(2*n)) / d
    h = z * math.sqrt(p*(1-p)/n + z*z/(4*n*n)) / d
    return ctr - h, ctr + h

def newcombe(k1, n1, k2, n2):                       # 95% CI for p1 - p2 (Newcombe method 10)
    p1, p2 = k1/n1, k2/n2; l1, u1 = _wilson(k1, n1); l2, u2 = _wilson(k2, n2)
    return ((p1-p2) - math.sqrt((p1-l1)**2 + (u2-p2)**2),
            (p1-p2) + math.sqrt((u1-p1)**2 + (p2-l2)**2))

def rr_katz(ko, no, kr, nr, z=1.959963985):         # risk ratio (outlier vs rest) + Katz log 95% CI
    if min(ko, kr, no, nr) == 0:
        return (float("nan"), float("nan"), float("nan"))
    rr = (ko/no) / (kr/nr)
    se = math.sqrt(1.0/ko - 1.0/no + 1.0/kr - 1.0/nr)
    return (rr, rr*math.exp(-z*se), rr*math.exp(z*se))

# ---- load 6 dense encoders -> distance matrix D ----
_label = lambda d: (os.path.basename(d).split("_", 1)[1] if "_" in os.path.basename(d) else os.path.basename(d))
run_dirs = [d for d in sorted(glob.glob(os.path.join(RUNS_ROOT, PATTERN)))
            if os.path.isdir(d) and _label(d) not in EXCLUDE]
assert run_dirs, f"no run dirs matching {PATTERN!r} under {RUNS_ROOT!r}"
dist = {}
for d in run_dirs:
    sc = sorted(glob.glob(os.path.join(d, "*_scores.csv")))
    if not sc:
        print(f"!! no *_scores.csv in {d}; skipping"); continue
    s = pd.read_csv(sc[0]); s[ID_COL] = s[ID_COL].astype(str)
    dist[_label(d)] = s.set_index(ID_COL)[DIST_COL]
D = pd.DataFrame(dist)
MODELS = list(D.columns)
assert len(MODELS) == 6, f"expected 6 encoders, got {len(MODELS)}: {MODELS}"

# consensus outliers: above the per-encoder p95 in ALL 6 encoders  (== section3_4_results.py rule)
thresh   = D.quantile(PCTILE / 100.0)
OUTLIERS = D.index[D.gt(thresh, axis=1).all(axis=1)]
outlier_score = (D.rank(pct=True) * 100).mean(axis=1)          # continuous exposure (dose)
N_OUT = len(OUTLIERS)
print(f"encoders ({len(MODELS)}): {MODELS}")
print(f"consensus outliers @ p{PCTILE:.0f} (all 6): N = {N_OUT}  (expect 584)")
assert 560 <= N_OUT <= 610, f"consensus N={N_OUT} off the expected ~584 -- check dense scores / PCTILE"

local = pd.DataFrame({"outlier_pctile": outlier_score,
                      "is_consensus_outlier": outlier_score.index.isin(OUTLIERS)})
local.index = local.index.astype(str); local.index.name = "person_id"


# %% Cell 1 -- EHR-linked cohort (linkage = >=1 visit_occurrence)
# RUN NOTE: observation_period exists for ALL 22,332 responders (AoU derives it from survey
# enrolment too) -> it is NOT an EHR-linkage marker.  The manuscript's "20,972 (94%) linked" is
# exactly the >=1-visit count, so >=1 visit IS EHR linkage here.  The "keep zero-visit people"
# idea therefore dissolves: the 1,360 zero-visit responders are survey-only (not in the EHR), not
# under-served patients -> excluded by the inner join on util.  obs_years (from observation_period)
# is still used to (a) build a visits-per-year rate and (b) window-adjust the engagement beta.
obs = Q(f"""
  SELECT person_id,
         DATE_DIFF(MAX(observation_period_end_date), MIN(observation_period_start_date), DAY)/365.25 AS obs_years
  FROM `{CDR}.observation_period` GROUP BY person_id""")
obs["person_id"] = obs.person_id.astype(str)

# utilization: inner-joined below -> EHR-linked = has >=1 visit (== the manuscript's 20,972)
util = Q(f"""
  SELECT person_id,
         COUNT(DISTINCT visit_occurrence_id) AS n_visits,
         COUNTIF(visit_concept_id IN (9203, 262)) AS n_ed_visits
  FROM `{CDR}.visit_occurrence` GROUP BY person_id""")
util["person_id"] = util.person_id.astype(str)

# physical multimorbidity: # distinct condition concepts, psych/SUD (SNOMED descendants) removed
# server-side so the burden is NOT circular with the distress screen.  Reported only if it sharpens.
multimorb = Q(f"""
  WITH psych AS (
    SELECT DISTINCT ca.descendant_concept_id AS cid
    FROM `{CDR}.concept_ancestor` ca
    JOIN `{CDR}.concept` c ON ca.ancestor_concept_id = c.concept_id
    WHERE c.vocabulary_id='SNOMED' AND c.standard_concept='S'
      AND c.concept_name IN ('Mental disorder','Substance-related disorder','Disorder due to psychoactive substance')
  )
  SELECT co.person_id, COUNT(DISTINCT co.condition_concept_id) AS n_conditions
  FROM `{CDR}.condition_occurrence` co
  WHERE co.condition_concept_id != 0 AND co.condition_concept_id NOT IN (SELECT cid FROM psych)
  GROUP BY co.person_id""")
multimorb["person_id"] = multimorb.person_id.astype(str)

demo = Q(f"""
  SELECT p.person_id, 2024 - EXTRACT(YEAR FROM p.birth_datetime) AS age, LOWER(cs.concept_name) AS sex
  FROM `{CDR}.person` p LEFT JOIN `{CDR}.concept` cs ON p.sex_at_birth_concept_id = cs.concept_id""")
demo["person_id"] = demo.person_id.astype(str)

# assemble: responders WITH EHR linkage (inner on util = >=1 visit) <- obs_years / conditions LEFT <- demo
ehr = (local.reset_index()
       .merge(util,   on="person_id", how="inner")
       .merge(obs,    on="person_id", how="left")
       .merge(multimorb, on="person_id", how="left")
       .merge(demo,   on="person_id", how="left"))
ehr["n_conditions"] = ehr["n_conditions"].fillna(0)          # a linked person can have 0 coded conditions
ehr["female"]      = (ehr.sex == "female").astype(int)
ehr["ed_reliance"] = ehr.n_ed_visits / ehr.n_visits.replace(0, np.nan)      # n_visits >= 1 by construction
ehr["visit_rate"]  = ehr.n_visits / ehr.obs_years.replace(0, np.nan)        # visits per observation-year

n_link = len(ehr)
print(f"\nEHR-linked responders (>=1 visit): {n_link}  (expect ~20,972)")
print(f"  consensus outliers linked: {int(ehr.is_consensus_outlier.sum())} / {N_OUT}")


# %% Cell 2 -- MECHANISM: self-reported access barriers (Healthcare Access & Utilization survey)
# concept_code = codebook variable name; s.val = the answer text after the last ': '
USUAL   = ["HealthAdvice_PlaceforHealthAdvice"]
COST    = ["CantAffordCare_HealthcareProvider", "CantAffordCare_PrescriptionMedicines", "CantAffordCare_Specialist",
           "CantAffordCare_FollowupCare", "CantAffordCare_EmergencyCare", "CantAffordCare_DentalCare",
           "CantAffordCare_Eyeglasses", "DelayedMedicalCare_CantAffordCoPay", "DelayedMedicalCare_DeductibleTooHigh",
           "DelayedMedicalCare_HadToPayOutOfPocket"]
DELAYED = ["DelayedMedicalCare_ChildCare", "DelayedMedicalCare_ElderlyCare", "DelayedMedicalCare_RuralArea",
           "DelayedMedicalCare_TimeOffWork", "DelayedMedicalCare_Transportation"]
COPING  = ["CantAffordCare_DelayedFillingRxToSaveMoney", "CantAffordCare_LowerCostRxToSaveMoney",
           "CantAffordCare_SkippedMedToSaveMoney", "CantAffordCare_TookLessMedToSaveMoney"]
MECH    = ["HealthProviderRaceReligion_DelayedOrNoCare"]

s = Q(f"""SELECT s.person_id, c.concept_code, s.answer
  FROM `{CDR}.ds_survey` s JOIN `{CDR}.concept` c ON s.question_concept_id = c.concept_id
  WHERE c.concept_code IN ({",".join(chr(39)+x+chr(39) for x in USUAL+COST+DELAYED+COPING+MECH)})""")
s["person_id"] = s.person_id.astype(str)
s["val"] = s.answer.astype(str).str.lower().str.rsplit(": ", n=1).str[-1].str.strip()
yes = lambda codes: set(s[s.concept_code.isin(codes) & (s.val == "yes")].person_id)

F = pd.DataFrame({"person_id": sorted(set(s.person_id))})
F["no_usual_source"]   = F.person_id.isin(set(s[(s.concept_code == "HealthAdvice_PlaceforHealthAdvice") & (s.val == "no")].person_id)).astype(int)
F["any_cost_barrier"]  = F.person_id.isin(yes(COST)).astype(int)
F["any_delayed_care"]  = F.person_id.isin(yes(DELAYED)).astype(int)
F["any_cost_coping"]   = F.person_id.isin(yes(COPING)).astype(int)
F["any_access_barrier"]= F[["no_usual_source", "any_cost_barrier", "any_delayed_care", "any_cost_coping"]].max(axis=1)
F["care_avoid_provider"]= F.person_id.isin(set(s[(s.concept_code == "HealthProviderRaceReligion_DelayedOrNoCare")
                          & (s.val.isin({"some of the time", "most of the time", "always"}))].person_id)).astype(int)

bar = ehr.merge(F, on="person_id", how="inner")            # survey answerers (their own, narrower denominator)
oc  = bar.is_consensus_outlier.values
n_o, n_r = int(oc.sum()), int((~oc).sum())
# Report EVERY barrier component (not just the headline three) so every prose sub-claim has a
# contrast behind it.  Main text: any + cost + care-avoidance verbally; the "delay care for
# structural reasons / ration medications / no usual source" sentence is backed by the middle
# three.  Full table -> supplement.
print(f"\n=== 3.4.4 MECHANISM  access barriers (survey answerers: outliers {n_o} vs rest {n_r}) ===")
bar_rows = []
for c, lab in [("any_access_barrier", "any access barrier"),
               ("any_cost_barrier",   "cost barrier"),
               ("any_delayed_care",   "delayed care, structural (transport/childcare/time-off)"),
               ("any_cost_coping",    "medication rationing to save money"),
               ("no_usual_source",    "no usual source of care"),
               ("care_avoid_provider","care avoidance, provider race/religion")]:
    ko, kr = int(bar[c][oc].sum()), int(bar[c][~oc].sum())
    op, rp = 100*ko/n_o, 100*kr/n_r
    dlo, dhi = newcombe(ko, n_o, kr, n_r)
    rr, rlo, rhi = rr_katz(ko, n_o, kr, n_r)
    bar_rows.append({"barrier": lab, "outlier_pct": op, "rest_pct": rp, "diff_pp": op-rp,
                     "diff_lo": 100*dlo, "diff_hi": 100*dhi, "rr": rr, "rr_lo": rlo, "rr_hi": rhi})
    print(f"  {lab:54s} {op:5.1f}% vs {rp:5.1f}%  diff {op-rp:+5.1f}pp [{100*dlo:+.0f},{100*dhi:+.0f}]"
          f"   RR {rr:4.2f} [{rlo:.2f},{rhi:.2f}]")


# %% Cell 3 -- engagement (routine visits) as outcome + ED reliance + multimorbidity direction
# CAVEAT (see RUN NOTE): the headline age+sex beta is the TOTAL effect, but it is confounded by the
# observation window.  The two robustness rows are the real test -- if the window-adjusted beta and
# the visits-per-year beta go to ~0, the raw "fewer visits" is data-completeness, not per-time
# under-engagement.  rank-OLS so it is scale-free and robust to the heavy visit-count tail.  Rank
# once over the linked cohort (NaN excluded from ranking); each model then uses its own complete cases.
for c in ["outlier_pctile", "n_visits", "visit_rate", "age", "obs_years"]:
    ehr[c + "_r"] = ehr[c].rank(pct=True)

def scoef(formula, cols):
    m = ehr.dropna(subset=cols).copy()
    fit = smf.ols(formula, m).fit()
    b = fit.params["outlier_pctile_r"]; lo, hi = fit.conf_int().loc["outlier_pctile_r"]
    return b, lo, hi, fit.pvalues["outlier_pctile_r"], len(m)

# headline: age + sex only (obs_years withheld -- it is partly a mediator of disengagement)
b0, l0, h0, p0, nm = scoef("n_visits_r ~ outlier_pctile_r + age_r + female",
                           ["n_visits_r", "outlier_pctile_r", "age_r", "female"])
# robustness 1: window-adjusted (is it purely a data-completeness artifact?)
b1, l1, h1, p1, _ = scoef("n_visits_r ~ outlier_pctile_r + age_r + female + obs_years_r",
                          ["n_visits_r", "outlier_pctile_r", "age_r", "female", "obs_years_r"])
# robustness 2: visits-per-observation-year rate (normalizes window structurally, not via covariate)
b2, l2, h2, p2, _ = scoef("visit_rate_r ~ outlier_pctile_r + age_r + female",
                          ["visit_rate_r", "outlier_pctile_r", "age_r", "female"])
print(f"\n=== 3.4.4 PRIMARY  engagement ~ score (rank-OLS) ===  (n={nm})")
print(f"  visits, age+sex adj          beta={b0:+.3f} [{l0:+.3f},{h0:+.3f}]  p={p0:.1e}   <- HEADLINE")
print(f"  visits, +obs_years (window)  beta={b1:+.3f} [{l1:+.3f},{h1:+.3f}]  p={p1:.1e}   (robustness)")
print(f"  visit rate/yr, age+sex adj   beta={b2:+.3f} [{l2:+.3f},{h2:+.3f}]  p={p2:.1e}   (robustness)")

# ED reliance (crisis door) -- now a MAIN-TEXT pillar, so give it legible window/volume-free numbers.
# ed_reliance = ED / total visits is a RATIO -> independent of the observation window that confounds
# raw visit counts.  All rows have >=1 visit, so it is always defined.
oce = ehr.is_consensus_outlier.values
rho_ed, p_ed = spearmanr(ehr.outlier_pctile, ehr.ed_reliance)
edshare_o = ehr.ed_reliance[oce].mean()*100;  edshare_r = ehr.ed_reliance[~oce].mean()*100
pool_o = 100*ehr.n_ed_visits[oce].sum()/ehr.n_visits[oce].sum()
pool_r = 100*ehr.n_ed_visits[~oce].sum()/ehr.n_visits[~oce].sum()
ehr["ed_reliance_r"] = ehr.ed_reliance.rank(pct=True)
# volume-adjusted: does the score predict a higher ED share holding TOTAL visits + age + sex?
be, le, he, pe, _ = scoef("ed_reliance_r ~ outlier_pctile_r + n_visits_r + age_r + female",
                          ["ed_reliance_r", "outlier_pctile_r", "n_visits_r", "age_r", "female"])
print(f"\n  ED reliance (ED / total visits):")
print(f"    rho(score)                = {rho_ed:+.3f}  p={p_ed:.1e}")
print(f"    mean per-person ED share  = outlier {edshare_o:.1f}% vs rest {edshare_r:.1f}%")
print(f"    pooled ED share of visits = outlier {pool_o:.1f}% vs rest {pool_r:.1f}%")
print(f"    ~ score (+visits,age,sex) = beta={be:+.3f} [{le:+.3f},{he:+.3f}]  p={pe:.1e}  <- volume/window-free")

# multimorbidity -- DROPPED from prose (fewer conditions is the same completeness confound), kept as a diagnostic
rho_mm, p_mm = spearmanr(ehr.outlier_pctile, ehr.n_conditions)
med_mm = ehr.groupby("is_consensus_outlier").n_conditions.median()
print(f"\n  [diagnostic, not reported] physical multimorbidity rho(score) = {rho_mm:+.3f}  p={p_mm:.1e}   "
      f"median conditions outlier {med_mm.get(True, float('nan')):.0f} vs rest {med_mm.get(False, float('nan')):.0f}")


# %% Cell 4 -- score-decile gradient (deciles over the EHR-linked cohort)
d = ehr.merge(F, on="person_id", how="left")
d["decile"] = pd.qcut(d.outlier_pctile, 10, labels=False) + 1
g = d.groupby("decile").agg(
    n=("person_id", "size"),
    mean_pctile=("outlier_pctile", "mean"),   # x-position: the group's mean Mahalanobis percentile (0-100)
    access_barrier_pct=("any_access_barrier", lambda x: 100*np.nanmean(x)),
    cost_barrier_pct=("any_cost_barrier",   lambda x: 100*np.nanmean(x)),
    care_avoid_pct=("care_avoid_provider",  lambda x: 100*np.nanmean(x)),
    median_visits=("n_visits", "median"),
    median_rate=("visit_rate", "median"),
    ed_reliance_pct=("ed_reliance", lambda x: 100*np.nanmean(x)),
    median_conditions=("n_conditions", "median"))
print("\n=== 3.4.4  score-decile gradient (barriers vs routine care) ===")
print(g.round(2).to_string())


# %% Cell 5 -- MAIN figure: barriers (left) + ED share of visits (right); routine-visit gradient -> supplement
fig, axL = plt.subplots(figsize=(7.5, 4.5), constrained_layout=True)
axL.plot(g.mean_pctile, g.access_barrier_pct, "-o", color="#A11526", label="any access barrier")
axL.plot(g.mean_pctile, g.cost_barrier_pct,   "-s", color="#E8A33D", label="cost barrier")
axL.plot(g.mean_pctile, g.care_avoid_pct,     "-^", color="#CC79A7", label="care avoidance (provider race/religion)")
axL.set_xlabel("mean Mahalanobis percentile"); axL.set_ylabel("% reporting barrier (survey)")
axL.legend(loc="upper left", fontsize=8, frameon=False)
axR = axL.twinx()
axR.plot(g.mean_pctile, g.ed_reliance_pct, "--D", color="#2166ac", label="ED share of visits (EHR)")
axR.set_ylabel("ED share of visits (%)", color="#2166ac"); axR.tick_params(axis="y", colors="#2166ac")
axR.legend(loc="upper right", fontsize=8, frameon=False)
fig.savefig("fig_ehr_gradient.png", dpi=300, bbox_inches="tight")
print("\nsaved fig_ehr_gradient.png"); plt.close(fig)


# %% Cell 5b -- SUPPLEMENT figure: raw visits fall but per observation-year the rate is flat (the caveat)
figs, axv = plt.subplots(figsize=(7.5, 4.5), constrained_layout=True)
axv.plot(g.mean_pctile, g.median_visits, "--D", color="#2166ac", label="median total visits")
axv.set_xlabel("mean Mahalanobis percentile")
axv.set_ylabel("median total EHR visits", color="#2166ac"); axv.tick_params(axis="y", colors="#2166ac")
axv.set_ylim(0, max(g.median_visits)*1.1)
axv.legend(loc="upper right", fontsize=8, frameon=False)
axr = axv.twinx()
axr.plot(g.mean_pctile, g.median_rate, "-o", color="#1a9850", label="median visits per observation-year")
axr.set_ylabel("median visits per observation-year", color="#1a9850"); axr.tick_params(axis="y", colors="#1a9850")
axr.set_ylim(0, max(g.median_rate)*1.3); axr.legend(loc="lower left", fontsize=8, frameon=False)
figs.savefig("fig_ehr_visits_supp.png", dpi=300, bbox_inches="tight")
print("saved fig_ehr_visits_supp.png"); plt.close(figs)


# %% Cell 6 -- PARKED (decision 3, 2026-07-21): does the score add over the 8 standard flags?
# Incremental-beyond-standard-screens rank-OLS (score coeff with n_standard_flags in the model)
# would show the under-served group is one that scale-by-scale screening would MISS.  Held for a
# reporting decision -- build n_standard_flags from responses_k0 (PHQ9/GAD7/ACE/ASRS/CIDI/Suicide/
# HVS/PROMIS) and add `+ n_standard_flags_r` to the Cell 3 models.  NOT run yet.


# %% Cell 7 -- persist every number for the manuscript / traceability
out = {
    "pctile": PCTILE, "n_consensus": int(N_OUT), "n_linked": int(n_link),
    "n_survey_outlier": n_o, "n_survey_rest": n_r,
    "barriers": bar_rows,
    "engagement": {"headline_beta": b0, "headline_ci": [l0, h0], "headline_p": p0,
                   "window_adj_beta": b1, "window_adj_ci": [l1, h1], "window_adj_p": p1,
                   "rate_beta": b2, "rate_ci": [l2, h2], "rate_p": p2},
    "ed_reliance": {"rho": rho_ed, "rho_p": p_ed,
                    "mean_share_outlier": edshare_o, "mean_share_rest": edshare_r,
                    "pooled_share_outlier": pool_o, "pooled_share_rest": pool_r,
                    "adj_beta": be, "adj_ci": [le, he], "adj_p": pe},
    "multimorbidity_rho": rho_mm, "multimorbidity_p": p_mm,   # diagnostic, not reported
    "gradient": json.loads(g.reset_index().to_json(orient="records")),
}
os.makedirs("outputs_dense", exist_ok=True)
with open("outputs_dense/underserved_stats.json", "w") as fh:
    json.dump(out, fh, indent=2)
print("saved outputs_dense/underserved_stats.json")
```
