## Supplemental code for "Scoring respondents on a semantic frame of reference identifies a group that is burdened across many questionnaires and reaches care less easily": pool_card_labels.py

```
"""Shared card labelling for the pool component grids.

Both the main-text card panel (fig1_pool.py) and the supplement's reproduction grid (fig_xenc.py)
draw the same 55 components, so they abbreviate construct names the same way. Abbreviations are a
presentation choice only. The underlying label is always the documented construct, never a grouping.
"""

ABBR = {"Neighborhood disorder": "Nbhd. disorder", "Neighborhood cohesion": "Nbhd. cohesion",
        "Interpersonal & sexual violence": "Sex/IP viol.", "Interpersonal violence": "IP violence",
        "Body-focused repetitive behavior": "Body-focused", "Adverse childhood experiences": "Childhood adv.",
        "Sleep-related impairment": "Sleep-rel. imp.", "Sleep disturbance": "Sleep distur.",
        "Quality of life: physical": "QoL: physical", "Quality of life: psychological": "QoL: psychol.",
        "Quality of life: environment": "QoL: environ.", "Quality of life: social": "QoL: social",
        "Quality of life: overall": "QoL: overall", "Post-traumatic stress": "PTSD",
        "Life-threatening events": "Life-threat.", "Everyday discrimination": "Everyday disc.",
        "Healthcare discrimination": "Healthc. disc.", "Major discrimination": "Major disc.",
        "Heightened vigilance": "Vigilance", "Subjective wellbeing": "Subj. wellb.",
        "Trauma exposure": "Trauma exp.", "Food insecurity": "Food insec.",
        "Somatic symptoms": "Somatic sympt.", "Emotion regulation": "Emotion reg.",
        "Social support": "Social supp.", "Emotional support": "Emot. supp.",
        "Social isolation": "Social isol.", "Perceived stress": "Perceived str.",
        "Life satisfaction": "Life satisf.", "Conscientiousness": "Conscient.",
        "Meaning in life": "Life meaning"}


def short(c, width=14):
    s = ABBR.get(c, c)
    return s if len(s) <= width else s[:width - 1] + "…"
```
