## Supplemental code for "Scoring respondents on a semantic frame of reference identifies a group that is burdened across many questionnaires and reaches care less easily": pool_colocation.py

```
#!/usr/bin/env python
"""Cross-instrument CO-LOCATION, the core-to-core read (PORTABILITY headline, NHB).

Framing (user, 2026-08-07): items are organized by SEMANTIC CONTENT, not by instrument. An
instrument's items are SUPPOSED to scatter across components when their content differs (PHQ-9 ->
depression / somatic / sleep is correct, not a failure). So we do NOT tally "% of an instrument on
one axis". The unit is the component's CORE.

Method. Fixed reference = pool W (577 external items, no AoU, stem-stripped).
  - pool's OWN core of component c  = fixed-point core of pool Z[:,c]  (outputs_pool/fixedpoint_cores)
      -> which POOL instruments DEFINE the axis (independent = one AoU never administered).
  - AoU's core of component c        = fixed-point core of P*[:,c], P* = (X_aou - mu_pool) @ Wᵀ
      (outputs_pool/aou_on_pool) -> which AoU items LAND on that same axis.
Both cores index the SAME pool component c. Cross-instrument co-location = AoU items land in the
core of a component whose pool core is DEFINED BY A DIFFERENT (independent) instrument of the SAME
content. That is meaning transfer across instruments, at the item level, with no instrument-cohesion
assumption.

Reads outputs_pool/{fixedpoint_cores,aou_on_pool}/<enc>_cores.csv (already computed).
Writes outputs_pool/colocation/{<enc>_colocation.csv, summary.csv}.
Run:  /data_gpu5/ROSS_Proj/env/bin/python pool_colocation.py   (pure CSV join, no embeddings)
"""
import csv, glob, os, re
from collections import Counter

HERE = os.path.dirname(os.path.abspath(__file__))
OUT = os.environ.get("ROSS_OUT_DIR", os.path.join(HERE, "outputs_pool"))
CMAP = os.path.join(HERE, "construct_map_151.csv")
POOL = os.path.join(HERE, "..", "global_pool", "pool_processed.csv")

_ALIASES = [
    {"anxiety", "generalized anxiety", "panic & phobia", "panic", "phobia"},
    {"depression", "depressive symptoms", "depressed mood"},
    {"loneliness", "social isolation"},
    {"social support", "emotional support", "perceived social support", "perceived emotional support"},
    {"food insecurity", "food insufficiency"},
    {"ptsd", "post-traumatic stress", "posttraumatic stress", "trauma exposure",
     "life-threatening events", "interpersonal & sexual violence", "interpersonal violence"},
    {"adverse childhood experiences", "childhood adversity", "childhood trauma"},
    {"perceived stress", "stress"},
    {"everyday discrimination", "discrimination", "major experiences of discrimination",
     "healthcare discrimination", "discrimination in medical settings", "heightened vigilance"},
    {"well-being", "subjective wellbeing", "well being", "subjective well-being",
     "quality of life", "quality of life: physical", "quality of life: psychological"},
    {"neighborhood social cohesion", "neighborhood cohesion"},
    {"neighborhood disorder & safety", "neighborhood disorder", "perceived neighborhood disorder"},
    {"openness", "open-mindedness"}, {"neuroticism", "negative emotionality"},
    {"psychosis", "prodromal symptoms", "psychotic experiences"},
    {"mania", "hypomania", "manic symptoms"},
    {"adult adhd", "adhd", "attention deficit"},
    {"daily spiritual experience", "spirituality", "daily spiritual experiences", "religious participation"},
    {"conscientiousness"}, {"extraversion"}, {"agreeableness"},   # Big-Five facets match exactly
    {"self-esteem"}, {"suicidality & self-harm", "suicidality"},
    {"somatic symptoms", "somatic"}, {"sleep disturbance", "insomnia", "sleep-related impairment"},
    {"anger"}, {"drug use", "substance use", "alcohol use"}, {"self-rated health"},
    {"emotion regulation"}, {"meaning in life", "meaning"}, {"resilience"}, {"optimism"},
]


def _n(s):
    return (s or "").strip().lower()


def fam(a, b):
    a, b = _n(a), _n(b)
    if not a or not b or "?" in (a, b):
        return False
    return a == b or any(a in g and b in g for g in _ALIASES)


def short(scale):
    m = re.findall(r"\(([^)]+)\)", scale)
    if m:
        return m[-1]
    s = scale.replace("PROMIS Item Bank v1.0 - Emotional Distress - ", "PROMIS-") \
             .replace("PROMIS Item Bank v2.0 - ", "PROMIS-").replace("PROMIS Item Bank v1.1 - ", "PROMIS-") \
             .replace("PROMIS Item Bank v1.0 - ", "PROMIS-")
    return s[:22]


def load_cores(path):
    out = {}
    for r in csv.DictReader(open(path)):
        out[int(r["component"])] = {"items": r["items"].split(),
                                    "modal": r["modal_construct"], "size": int(r["size"])}
    return out


def main():
    aou_instr = {r["item"]: r["instrument"] for r in csv.DictReader(open(CMAP))}
    aou_con = {r["item"]: r["construct"] for r in csv.DictReader(open(CMAP))}
    pool = list(csv.DictReader(open(POOL)))
    pscale = {r["item_id"]: r["scale"] for r in pool}
    prole = {r["item_id"]: r["role"] for r in pool}

    outdir = os.path.join(OUT, "colocation"); os.makedirs(outdir, exist_ok=True)
    summ = []
    for pcpath in sorted(glob.glob(os.path.join(OUT, "fixedpoint_cores", "*_cores.csv"))):
        m = os.path.basename(pcpath).replace("_cores.csv", "")
        poolc = load_cores(pcpath)
        aouc = load_cores(os.path.join(OUT, "aou_on_pool", f"{m}_cores.csv"))

        rows = []
        for c in sorted(set(poolc) | set(aouc)):
            pc, ac = poolc.get(c), aouc.get(c)
            pool_scales = [short(pscale.get(i, "?")) for i in pc["items"]] if pc else []
            pool_roles = [prole.get(i, "?") for i in pc["items"]] if pc else []
            pool_indep = pc and Counter(pool_roles).most_common(1)[0][0] == "independent"
            aou_items = ac["items"] if ac else []
            aou_ins = sorted(set(aou_instr.get(i, "?") for i in aou_items))
            content = bool(pc and ac and fam(pc["modal"], ac["modal"]))
            # cross-instrument: AoU content lands on an axis DEFINED by an independent pool instrument
            cross = bool(content and pool_indep)
            rows.append({
                "component": c,
                "pool_axis": pc["modal"] if pc else "", "pool_core_n": pc["size"] if pc else 0,
                "pool_core_instruments": " ".join(f"{s}[{r[0]}]" for s, r in zip(pool_scales, pool_roles)),
                "aou_axis": ac["modal"] if ac else "", "aou_core_n": ac["size"] if ac else 0,
                "aou_core_instruments": " ".join(aou_ins),
                "aou_core_items": " ".join(aou_items),
                "content_match": int(content), "cross_instrument": int(cross),
            })
        with open(os.path.join(outdir, f"{m}_colocation.csv"), "w", newline="") as fh:
            w = csv.DictWriter(fh, fieldnames=list(rows[0].keys())); w.writeheader(); w.writerows(rows)

        matched = [r for r in rows if r["content_match"]]
        cross = [r for r in rows if r["cross_instrument"]]
        aou_covered = set(i for r in matched for i in r["aou_core_items"].split())
        summ.append({"encoder": m, "k": len(poolc),
                     "content_matched_axes": len(matched), "cross_instrument_axes": len(cross),
                     "aou_items_on_matched_axes": len(aou_covered)})
        print(f"\n### {m}: {len(cross)} cross-instrument co-location axes "
              f"({len(matched)} content-matched, {len(aou_covered)} AoU items) ###")
        for r in sorted(cross, key=lambda x: -x["aou_core_n"])[:12]:
            print(f"  c{r['component']:<3d} {r['pool_axis'][:26]:26s} "
                  f"pool<-[{r['pool_core_instruments'][:52]}]  AoU<-[{r['aou_core_instruments']}]")

    with open(os.path.join(outdir, "summary.csv"), "w", newline="") as fh:
        w = csv.DictWriter(fh, fieldnames=list(summ[0].keys())); w.writeheader(); w.writerows(summ)
    print("\n->", os.path.join(outdir, "summary.csv"))


if __name__ == "__main__":
    main()
```
