## Supplemental code for "Scoring respondents on a semantic frame of reference identifies a group that is burdened across many questionnaires and reaches care less easily": pool_core_columns.py

```
#!/usr/bin/env python
"""Core COMPONENTS (columns) per item -- the dual of core ITEMS per column.

Motivation (NHB P1, generalizing use of the pool basis): to attribute an ARBITRARY
projected item to the pool's components, we need, for each item i, the small set of
components that DEFINE it -- the converse of the fixed-point core-items that define a
component.

Definition.
  Z = pool scores (items x components) from the operating projection P (NOT the
  variance-ordered basis W). P's columns carry heterogeneous scale (gte: std spread
  ~3.5x), so raw rows are not comparable across columns. Scores are already
  column-mean-zero (projection of mean-centered embeddings), so we STANDARDIZE each
  column by its magnitude (std): Ztil[:,c] = Z[:,c] / std(Z[:,c]).
    - core_items(c)  = fixed_point_core(Z[:,c])  == fixed_point_core(Ztil[:,c])
      (PR and within-column |.| ranking are scale-invariant -> UNCHANGED vs published).
    - core_cols(i)   = fixed_point_core(Ztil[i,:])  (the components on which item i is
      most distinctively extreme, in each column's own SD units -- the proper dual).

Reciprocity question: if c is a core column of i, is i a core item of c?
  E_cols  = {(i,c): c in core_cols(i)}      E_items = {(i,c): i in core_items(c)}
  forward (col->item) = |E_cols & E_items| / |E_cols|
  backward(item->col) = |E_cols & E_items| / |E_items|
  jaccard             = |E_cols & E_items| / |E_cols | E_items|

Writes outputs_pool/core_columns/<enc>_core_columns.csv (per item) + reciprocity.csv.
Run:  CUDA_VISIBLE_DEVICES="" /data_gpu5/ROSS_Proj/env/bin/python pool_core_columns.py
"""
import csv, os, sys
import numpy as np
from collections import Counter

HERE = os.path.dirname(os.path.abspath(__file__))
OUT = os.environ.get("ROSS_OUT_DIR", os.path.join(HERE, "outputs_pool"))
POOL = os.path.join(HERE, "..", "global_pool", "pool_processed.csv")
ENCODERS = sys.argv[1:] or ["gte-large-en-v1.5"]


def pr(v):
    s2 = float((v ** 2).sum()); s4 = float((v ** 4).sum())
    return s2 * s2 / s4 if s4 > 0 else 0.0


def fixed_point_core(v):
    idx = np.arange(len(v))
    while True:
        n = max(1, min(int(round(pr(v[idx]))), len(idx)))
        top = np.sort(idx[np.argsort(-np.abs(v[idx]))[:n]])
        if len(top) == len(idx) and (top == idx).all():
            return top
        idx = top


def fit_dir(enc):
    import glob
    g = glob.glob(os.path.join(OUT, "fits", f"{enc}__k*_ls1.0"))
    return g[0]


def main():
    con = {r["item_id"]: r["construct"] for r in csv.DictReader(open(POOL))}
    recdir = os.path.join(OUT, "core_columns"); os.makedirs(recdir, exist_ok=True)
    summary = []
    for enc in ENCODERS:
        fd = fit_dir(enc)
        Z = np.load(os.path.join(fd, "scores.npy"))                       # items x comps
        items = [str(x) for x in np.load(os.path.join(HERE, "data", f"pool_items_{enc}.npz"),
                                         allow_pickle=True)["items"]]
        n_item, k = Z.shape
        mu = Z.mean(axis=0); sd = Z.std(axis=0)
        print(f"\n### {enc}: {n_item} items x {k} comps | col-mean |max|={np.abs(mu).max():.2e} "
              f"(should be ~0) | std {sd.min():.3f}-{sd.max():.3f} ###")
        Ztil = (Z - mu) / sd                                              # standardized

        core_items = {c: set(fixed_point_core(Z[:, c]).tolist()) for c in range(k)}   # == published
        # modal construct of each component (from its core items), for labeling
        comp_modal = {}
        for c in range(k):
            cc = Counter(con.get(items[i], "?") for i in core_items[c])
            comp_modal[c] = cc.most_common(1)[0][0] if cc else "?"

        core_cols = {i: fixed_point_core(Ztil[i, :]) for i in range(n_item)}          # NEW dual

        # per-item CSV
        rows = []
        for i in range(n_item):
            cols = core_cols[i]
            cols = cols[np.argsort(-np.abs(Ztil[i, cols]))]                            # order by |z|
            recip = [int(i in core_items[int(c)]) for c in cols]
            rows.append({
                "item": items[i], "construct": con.get(items[i], "?"),
                "n_core_cols": len(cols),
                "core_cols": " ".join(f"C{c}" for c in cols),
                "core_col_constructs": " | ".join(comp_modal[int(c)] for c in cols),
                "core_col_absz": " ".join(f"{abs(Ztil[i, c]):.2f}" for c in cols),
                "reciprocal": " ".join(map(str, recip)),
                "n_reciprocal": int(sum(recip)),
            })
        with open(os.path.join(recdir, f"{enc}_core_columns.csv"), "w", newline="") as fh:
            w = csv.DictWriter(fh, fieldnames=list(rows[0].keys())); w.writeheader(); w.writerows(rows)

        # reciprocity
        E_cols = set((i, int(c)) for i in range(n_item) for c in core_cols[i])
        E_items = set((i, c) for c in range(k) for i in core_items[c])
        inter = E_cols & E_items
        fwd = len(inter) / len(E_cols)
        bwd = len(inter) / len(E_items)
        jac = len(inter) / len(E_cols | E_items)
        sizes = sorted(len(core_cols[i]) for i in range(n_item))
        med_sz = sizes[len(sizes) // 2]
        items_any = sum(1 for i in range(n_item) if any(i in core_items[int(c)] for c in core_cols[i]))
        top_recip = sum(1 for i in range(n_item)
                        if int(core_cols[i][np.argmax(np.abs(Ztil[i, core_cols[i]]))]) in
                        [c for c in range(k) if i in core_items[c]])
        print(f"  core_cols/item: median {med_sz}, range {sizes[0]}-{sizes[-1]}")
        print(f"  |E_cols|={len(E_cols)}  |E_items|={len(E_items)}  inter={len(inter)}")
        print(f"  forward  (of item's core cols, share where item is a core item): {fwd:.3f}")
        print(f"  backward (of col's core items, share where col is a core col)  : {bwd:.3f}")
        print(f"  jaccard(E_cols, E_items) = {jac:.3f}")
        print(f"  items with >=1 reciprocal core col: {items_any}/{n_item} ({items_any/n_item:.2f})")
        print(f"  items whose TOP core col is reciprocal: {top_recip}/{n_item} ({top_recip/n_item:.2f})")
        summary.append({"encoder": enc, "n_item": n_item, "k": k,
                        "median_core_cols": med_sz, "min_core_cols": sizes[0], "max_core_cols": sizes[-1],
                        "E_cols": len(E_cols), "E_items": len(E_items), "intersection": len(inter),
                        "forward_recip": round(fwd, 3), "backward_recip": round(bwd, 3),
                        "jaccard": round(jac, 3),
                        "items_ge1_recip": items_any, "items_top_recip": top_recip})
    with open(os.path.join(recdir, "reciprocity.csv"), "w", newline="") as fh:
        w = csv.DictWriter(fh, fieldnames=list(summary[0].keys())); w.writeheader(); w.writerows(summary)
    print("\n->", os.path.join(recdir, "reciprocity.csv"))


if __name__ == "__main__":
    main()
```
