## Supplemental code for "Scoring respondents on a semantic frame of reference identifies a group that is burdened across many questionnaires and reaches care less easily": pool_fixedpoint_cores.py

```
#!/usr/bin/env python
"""Fixed-point PR cores for the GLOBAL POOL fits (stage-3, the CORRECT core rule).

For each encoder's fit (scores Z, items x components), take the participation-ratio
**fixed-point** core of every component column: iterate keep-top-round(PR) until the
set stops changing (PR(S) <= |S| makes it non-increasing -> a small self-consistent
core). This is the SAME rule the manuscript used (make_core_items.fixed_point_core /
crossencoder_cores.fpk) -- NOT the single-pass round(PR) in prune_scores_pr.py, which
leaves ~50-item cores because PR of the spread-out dense column is large.

Writes outputs_pool/fixedpoint_cores/<encoder>_cores.csv (component, size, modal
construct, purity, items, constructs) + summary.csv.

Run:
    ROSS_NPZ_PREFIX=pool_items ROSS_OUT_DIR=.../outputs_pool \
      CUDA_VISIBLE_DEVICES="" /data_gpu5/ROSS_Proj/env/bin/python pool_fixedpoint_cores.py
"""
import csv, glob, os
import numpy as np
from collections import Counter

HERE = os.path.dirname(os.path.abspath(__file__))
OUT = os.environ.get("ROSS_OUT_DIR", os.path.join(HERE, "outputs_pool"))
POOL = os.path.join(HERE, "..", "global_pool", "pool_processed.csv")


def pr(v):
    s2 = float((v ** 2).sum()); s4 = float((v ** 4).sum())
    return s2 * s2 / s4 if s4 > 0 else 0.0


def fixed_point_core(v):
    """Iterate keep-top-round(PR) to convergence -> the fixed-point core indices."""
    idx = np.arange(len(v))
    while True:
        n = max(1, min(int(round(pr(v[idx]))), len(idx)))
        top = np.sort(idx[np.argsort(-np.abs(v[idx]))[:n]])
        if len(top) == len(idx) and (top == idx).all():
            return top
        idx = top


def main():
    meta = {r["item_id"]: r["construct"] for r in csv.DictReader(open(POOL))}
    outdir = os.path.join(OUT, "fixedpoint_cores"); os.makedirs(outdir, exist_ok=True)
    summ = []
    for fit in sorted(glob.glob(os.path.join(OUT, "fits", "*_ls1.0"))):
        m = os.path.basename(fit).split("__")[0]
        Z = np.load(os.path.join(fit, "scores.npy"))
        items = [str(x) for x in np.load(os.path.join(HERE, "data", f"pool_items_{m}.npz"),
                                         allow_pickle=True)["items"]]
        rows = []
        for c in range(Z.shape[1]):
            core = fixed_point_core(Z[:, c])
            core = core[np.argsort(-np.abs(Z[core, c]))]          # order by |score|
            cons = [meta.get(items[i], "?") for i in core]
            cc = Counter(cons); modal, mc = cc.most_common(1)[0]
            rows.append({"component": c, "size": len(core),
                         "modal_construct": modal, "purity": round(mc / len(core), 3),
                         "items": " ".join(items[i] for i in core),
                         "constructs": " | ".join(cons)})
        with open(os.path.join(outdir, f"{m}_cores.csv"), "w", newline="") as fh:
            w = csv.DictWriter(fh, fieldnames=list(rows[0].keys())); w.writeheader(); w.writerows(rows)
        sizes = sorted(r["size"] for r in rows)
        cov = len(set(w for r in rows for w in r["items"].split())) / len(items)
        purs = sorted(r["purity"] for r in rows)
        summ.append({"encoder": m, "k": len(rows), "median_size": sizes[len(sizes) // 2],
                     "min_size": sizes[0], "max_size": sizes[-1],
                     "coverage": round(cov, 3), "median_purity": purs[len(purs) // 2]})
        print(f"{m:24s} k={len(rows)} med_size={sizes[len(sizes)//2]} cov={cov:.2f} "
              f"med_purity={purs[len(purs)//2]:.2f}")
    with open(os.path.join(outdir, "summary.csv"), "w", newline="") as fh:
        w = csv.DictWriter(fh, fieldnames=list(summ[0].keys())); w.writeheader(); w.writerows(summ)
    print("->", os.path.join(outdir, "summary.csv"))


if __name__ == "__main__":
    main()
```
