## Supplemental code for "Scoring respondents on a semantic frame of reference identifies a group that is burdened across many questionnaires and reaches care less easily": pool_project_aou.py

```
#!/usr/bin/env python
"""Project the AoU-151 item embeddings onto the POOL's W (per encoder) and take the
fixed-point PR core of each resulting component column.

Tests whether the pool-derived meaning basis (learned with NO AoU involvement)
organizes AoU's own 151 items into construct-coherent cores. This is the REVERSE
of the manuscript framing (there AoU is the fixed reference; here the pool is).

Projection uses the pool's mean (the frame W was learned in):  P = (X_aou - mu_pool) @ W_poolᵀ.
Cores = iterated fixed_point_core on each column of P (same rule as pool_fixedpoint_cores.py).

CAVEATS recorded in output: (a) some AoU items ARE pool anchor instruments (their landing is a
sanity check, flagged via `aou_in_pool` by source-instrument overlap is NOT auto-computed here);
(b) AoU-151 embeddings carry item stems while pool W was learned on stem-stripped text.

Writes outputs_pool/aou_on_pool/<encoder>_cores.csv + summary.csv.
Run:  CUDA_VISIBLE_DEVICES="" /data_gpu5/ROSS_Proj/env/bin/python pool_project_aou.py
"""
import csv, glob, os
import numpy as np
from collections import Counter

HERE = os.path.dirname(os.path.abspath(__file__))
OUT = os.environ.get("ROSS_OUT_DIR", os.path.join(HERE, "outputs_pool"))
CMAP = os.path.join(HERE, "construct_map_151.csv")


def pr(v):
    s2 = float((v ** 2).sum()); s4 = float((v ** 4).sum())
    return s2 * s2 / s4 if s4 > 0 else 0.0


def fixed_point_core(v):
    idx = np.arange(len(v))
    while True:
        n = max(1, min(int(round(pr(v[idx]))), len(idx)))
        top = np.sort(idx[np.argsort(-np.abs(v[idx]))[:n]])
        if len(top) == len(idx) and (top == idx).all():
            return top
        idx = top


def main():
    cmap = {r["item"]: r["construct"] for r in csv.DictReader(open(CMAP))}
    outdir = os.path.join(OUT, "aou_on_pool"); os.makedirs(outdir, exist_ok=True)
    summ = []
    for fit in sorted(glob.glob(os.path.join(OUT, "fits", "*_ls1.0"))):
        m = os.path.basename(fit).split("__")[0]
        W = np.load(os.path.join(fit, "loadings.npy")); mu = np.load(os.path.join(fit, "mean.npy"))
        d = np.load(os.path.join(HERE, "data", f"core151_items_{m}.npz"), allow_pickle=True)
        items = [str(x) for x in d["items"]]; X = np.asarray(d["vectors"], float)
        P = (X - mu) @ W.T                                   # 151 x k_pool
        rows = []
        for c in range(P.shape[1]):
            core = fixed_point_core(P[:, c]); core = core[np.argsort(-np.abs(P[core, c]))]
            cons = [cmap.get(items[i], "?") for i in core]
            cc = Counter(cons); modal, mc = cc.most_common(1)[0]
            rows.append({"component": c, "size": len(core),
                         "modal_construct": modal, "purity": round(mc / len(core), 3),
                         "pure_axis": int(len(core) >= 2 and mc == len(core)),
                         "items": " ".join(items[i] for i in core),
                         "constructs": " | ".join(cons)})
        with open(os.path.join(outdir, f"{m}_cores.csv"), "w", newline="") as fh:
            w = csv.DictWriter(fh, fieldnames=list(rows[0].keys())); w.writeheader(); w.writerows(rows)
        sizes = sorted(r["size"] for r in rows)
        cov = len(set(w for r in rows for w in r["items"].split())) / len(items)
        purs = sorted(r["purity"] for r in rows)
        summ.append({"encoder": m, "k": len(rows), "median_size": sizes[len(sizes) // 2],
                     "coverage": round(cov, 3), "median_purity": purs[len(purs) // 2],
                     "pure_axes": sum(r["pure_axis"] for r in rows)})
        print(f"{m:24s} k={len(rows)} med_size={sizes[len(sizes)//2]} cov={cov:.2f} "
              f"med_purity={purs[len(purs)//2]:.2f} pure_axes={sum(r['pure_axis'] for r in rows)}")
    with open(os.path.join(outdir, "summary.csv"), "w", newline="") as fh:
        w = csv.DictWriter(fh, fieldnames=list(summ[0].keys())); w.writeheader(); w.writerows(summ)
    print("->", os.path.join(outdir, "summary.csv"))


if __name__ == "__main__":
    main()
```
