## Supplemental code for "Scoring respondents on a semantic frame of reference identifies a group that is burdened across many questionnaires and reaches care less easily": pool_xenc_hungarian.py

```
#!/usr/bin/env python
"""Cross-encoder reproducibility of pool components via Hungarian matching of cores (label-free).

For each encoder pair: Jaccard similarity between component cores (item sets) -> rectangular
linear_sum_assignment -> matched-pair Jaccards. Null = permute item identities in B, rematch.
tau = 95th pct of null matched Jaccard. Then anchor on gte: a component "reproduces in E" if its
matched Jaccard in E >= tau; "reproduces across all six" if it does so in all 5 other encoders.
Break down reproducibility by core mixedness (# distinct constructs), current 54-construct labels.
"""
import csv, glob, os
import numpy as np
from collections import Counter
from scipy.optimize import linear_sum_assignment

GP = "examples/global_pool"; OUTP = "examples/allofus/outputs_pool/fixedpoint_cores"
doc = {r['item_id']: r['documented_label'] for r in csv.DictReader(open(f"{GP}/item_construct_documented.csv"))}

enc_cores = {}
for f in sorted(glob.glob(f"{OUTP}/*_cores.csv")):
    enc = os.path.basename(f).replace("_cores.csv", "")
    comps = []
    for r in csv.DictReader(open(f)):
        items = set(str(r['items']).split())
        comps.append((items, len(set(doc.get(i, '?') for i in items))))
    enc_cores[enc] = comps
ENCS = list(enc_cores)
universe = sorted(set().union(*[it for e in ENCS for it, _ in enc_cores[e]]))
print("k per encoder:", {e: len(enc_cores[e]) for e in ENCS})

def jac(a, b): return len(a & b) / len(a | b) if (a | b) else 0.0
def match(A, B):
    J = np.array([[jac(a[0], b[0]) for b in B] for a in A])
    ri, ci = linear_sum_assignment(1.0 - J)
    return {int(i): float(J[i, j]) for i, j in zip(ri, ci)}

pairs = [(A, B) for i, A in enumerate(ENCS) for B in ENCS[i + 1:]]
real = np.array([v for A, B in pairs for v in match(enc_cores[A], enc_cores[B]).values()])

rng = np.random.RandomState(0)
null = []
for A, B in pairs:
    for _ in range(5):
        perm = dict(zip(universe, rng.permutation(universe)))
        Bp = [(set(perm.get(x, x) for x in it), nc) for it, nc in enc_cores[B]]
        null += list(match(enc_cores[A], Bp).values())
null = np.array(null); tau = np.percentile(null, 95)
print(f"\nmatched Jaccard  |  real median {np.median(real):.3f} mean {real.mean():.3f}"
      f"  |  null median {np.median(null):.3f}  tau(null p95) {tau:.3f}")
print(f"share of matched pairs with Jaccard >= tau: {(real >= tau).mean()*100:.0f}%")

# ---- encoder-agreement matrix (spot an outlier encoder) ----
Mpair = {}
for A, B in pairs:
    med = float(np.median(list(match(enc_cores[A], enc_cores[B]).values())))
    Mpair[(A, B)] = med; Mpair[(B, A)] = med
sh = {e: e.replace('-embedding', '').split('-')[0][:9] for e in ENCS}
print("\npairwise median matched Jaccard (encoder x encoder):")
print(" " * 11 + " ".join(f"{sh[e]:>9s}" for e in ENCS))
for A in ENCS:
    cells = " ".join((f"{Mpair[(A, B)]:9.2f}" if A != B else f"{'--':>9s}") for B in ENCS)
    mo = np.mean([Mpair[(A, B)] for B in ENCS if B != A])
    print(f"{sh[A]:>10s} {cells}   mean {mo:.3f}")

# ---- anchor gte: per-other match counts + reproducibility at several thresholds ----
ANCHOR = "gte-large-en-v1.5"; others = [e for e in ENCS if e != ANCHOR]
ga = enc_cores[ANCHOR]; ncv = np.array([nc for _, nc in ga])
Jby = {E: match(ga, enc_cores[E]) for E in others}
print("\nanchor gte: # of 55 components matching >= tau in each other encoder:")
for E in others:
    print(f"  {E:24s} {sum(1 for i in range(len(ga)) if Jby[E].get(i, 0.0) >= tau):2d}/55")
rep = np.array([sum(1 for E in others if Jby[E].get(i, 0.0) >= tau) for i in range(len(ga))])
print("\nreproducibility at several thresholds (of 6 encoders), with mixedness breakdown:")
for thr, name in [(5, 'all 6'), (4, '>=5/6'), (3, '>=4/6')]:
    p = rep >= thr
    bym = "  ".join(f"{b}c {int(p[(ncv>=5) if b=='5+' else (ncv==b)].sum())}/{int(((ncv>=5) if b=='5+' else (ncv==b)).sum())}"
                    for b in [1, 2, 3, 4, '5+'])
    print(f"  {name}: {p.sum():2d}/55   |  {bym}")
```
