## Supplemental code for "Scoring respondents on a semantic frame of reference identifies a group that is burdened across many questionnaires and reaches care less easily": pool_xenc_itemside_profiles.py

```
#!/usr/bin/env python
"""Item-side cross-encoder agreement, recomputed on construct PROFILES (D6-A).

Modal labelling is retired: a component is read by the SET of constructs of its core items, not by
the most common one. So an item's construct set under encoder e is

    S_g^e  =  union over i in F_g^e  of  { C_h : h in E_i^e }

    F_g^e  core components of item g   (fixed-point PR down its row of Z_G^e)
    E_i^e  core items of component i   (fixed-point PR down column i of Z_G^e)
    C_h    documented construct of item h (pool_processed.csv)

Agreement = mean pairwise Jaccard of S_g over the 15 encoder pairs, against a null that permutes
item identities (the same null as the modal version in sec:methods_xenc). The modal numbers are
recomputed alongside so the change is attributable to the labelling and nothing else.

Reports: overall, at the coarser domain level, split by whether an item anchors any core, and
stratified by the item's number of core components (the Supplementary Table S16 rows).

Run: CUDA_VISIBLE_DEVICES="" /data_gpu5/ROSS_Proj/env/bin/python pool_xenc_itemside_profiles.py
"""
import os, glob, csv
from itertools import combinations
import numpy as np
import pandas as pd

HERE = os.path.dirname(os.path.abspath(__file__))
GP = os.path.join(HERE, "..", "global_pool")
ENCS = ["gte-large-en-v1.5", "bge-m3", "bge-large-en-v1.5", "all-mpnet-base-v2",
        "e5-large-v2", "qwen3-embedding-0.6b"]
RNG = np.random.RandomState(0)
NPERM = 200


def pr(v):
    s2 = float((v**2).sum()); s4 = float((v**4).sum())
    return s2 * s2 / s4 if s4 > 0 else 0.0


def fpc(v):
    idx = np.arange(len(v))
    while True:
        n = max(1, min(int(round(pr(v[idx]))), len(idx)))
        top = np.sort(idx[np.argsort(-np.abs(v[idx]))[:n]])
        if len(top) == len(idx) and (top == idx).all():
            return top
        idx = top


def jac(a, b):
    if isinstance(a, dict):                              # weighted (Ruzicka) form
        keys = set(a) | set(b)
        num = sum(min(a.get(k, 0.0), b.get(k, 0.0)) for k in keys)
        den = sum(max(a.get(k, 0.0), b.get(k, 0.0)) for k in keys)
        return num / den if den else 1.0
    return len(a & b) / len(a | b) if (a | b) else 1.0


def mean_pairwise(sets_by_enc, perm=False):
    n = len(sets_by_enc[0]); vals = []
    for A, B in combinations(range(len(sets_by_enc)), 2):
        pb = RNG.permutation(n) if perm else np.arange(n)
        vals.append(np.mean([jac(sets_by_enc[A][i], sets_by_enc[B][pb[i]]) for i in range(n)]))
    return float(np.mean(vals))


def per_item(sets_by_enc):
    n = len(sets_by_enc[0])
    return np.array([np.mean([jac(sets_by_enc[A][i], sets_by_enc[B][i])
                              for A, B in combinations(range(len(sets_by_enc)), 2)]) for i in range(n)])


pp = pd.read_csv(os.path.join(GP, "pool_processed.csv"))
dom = dict(pd.read_csv(os.path.join(GP, "pool_construct_domain.csv")).values)
con = dict(zip(pp.item_id, pp.construct))
ids = list(pp.item_id)
pos = {g: j for j, g in enumerate(ids)}

prof_sets, prof_dom_sets, modal_sets, modal_dom_sets, wgt_sets, sizes, anchors = [], [], [], [], [], [], []
for enc in ENCS:
    fit = glob.glob(os.path.join(HERE, "outputs_pool", "fits", f"{enc}__k*_ls1.0"))[0]
    Z = np.load(os.path.join(fit, "scores.npy"))
    Zt = Z / Z.std(0)                                   # D2: pool sigma, columns already zero-mean
    rows = list(csv.DictReader(open(os.path.join(HERE, "outputs_pool", "fixedpoint_cores", f"{enc}_cores.csv"))))
    core_items = {int(r["component"]): str(r["items"]).split() for r in rows}
    modal = {int(r["component"]): r["modal_construct"] for r in rows}
    profile = {c: frozenset(con[i] for i in v) for c, v in core_items.items()}
    anchored = {i for v in core_items.values() for i in v}

    ps, pds, ms, md, ws, sz = [], [], [], [], [], []
    for g in ids:
        F = [int(c) for c in fpc(Zt[pos[g]])]
        u = frozenset().union(*[profile[c] for c in F])
        ps.append(u)
        pds.append(frozenset(dom[c] for c in u))
        ms.append(frozenset(modal[c] for c in F))
        md.append(frozenset(dom[modal[c]] for c in F))
        w = {}                                          # construct share, averaged over core components
        for c in F:
            items_c = core_items[c]
            for h in items_c:
                w[con[h]] = w.get(con[h], 0.0) + 1.0 / (len(items_c) * len(F))
        ws.append(w)
        sz.append(len(F))
    prof_sets.append(ps); prof_dom_sets.append(pds); modal_sets.append(ms)
    modal_dom_sets.append(md); wgt_sets.append(ws)
    sizes.append(sz); anchors.append(np.array([g in anchored for g in ids]))

sizes = np.array(sizes)
mean_size = sizes.mean(0)                              # as in Supplementary Table S16
anchor_any = np.array(anchors).mean(0) >= 0.5          # anchors a core under most encoders

print(f"pool {len(ids)} items, {len(ENCS)} encoders, {NPERM} permutations\n")
print(f"{'set':34s} {'mean |set|':>10s} {'real':>7s} {'null':>7s} {'chance-corrected':>17s}")
for name, sets in [("modal construct (retired)", modal_sets),
                   ("construct profile", prof_sets),
                   ("construct profile, weighted", wgt_sets),
                   ("modal construct, domain level", modal_dom_sets),
                   ("construct profile, domain level", prof_dom_sets)]:
    real = mean_pairwise(sets)
    null = np.mean([mean_pairwise(sets, perm=True) for _ in range(NPERM)])
    msz = np.mean([len(s) for e in sets for s in e])
    print(f"{name:34s} {msz:10.2f} {real:7.3f} {null:7.3f} {(real-null)/(1-null):17.3f}")

pi_prof, pi_modal, pi_wgt = per_item(prof_sets), per_item(modal_sets), per_item(wgt_sets)
print(f"\nanchors a core   n={int(anchor_any.sum()):3d}   profile {pi_prof[anchor_any].mean():.3f}"
      f"   (modal {pi_modal[anchor_any].mean():.3f})")
print(f"anchors none     n={int((~anchor_any).sum()):3d}   profile {pi_prof[~anchor_any].mean():.3f}"
      f"   (modal {pi_modal[~anchor_any].mean():.3f})")

print("\nstratified by the item's average number of core components (Supplementary Table S16 bins):")
print(f"{'core components':16s} {'items':>6s} {'profile':>9s} {'weighted':>10s} {'modal':>8s}")
for lo, hi, tag in [(0, 1.5, "1"), (1.5, 2.5, "2"), (2.5, 4.5, "3 to 4"), (4.5, 99, "5 or more")]:
    m = (mean_size >= lo) & (mean_size < hi)
    if m.sum():
        print(f"{tag:16s} {int(m.sum()):6d} {pi_prof[m].mean():9.3f} {pi_wgt[m].mean():10.3f} "
              f"{pi_modal[m].mean():8.3f}")
```
