## Supplemental code for "Scoring respondents on a semantic frame of reference identifies a group that is burdened across many questionnaires and reaches care less easily": porting_quality.py

```
#!/usr/bin/env python
"""How well each item of a ported battery ports, on three separable quantities.

Placing a battery on the fixed basis is not all-or-nothing, and "it ported" hides three different
things that can each fail on their own. For every item of every ported battery, under each of the six
encoders, this measures:

  COVERAGE   ||V^T (x_g - mu_G)||^2 / ||x_g - mu_G||^2, with V an orthonormal basis of the row space
             of the KEPT rows of W_G. How much of the item the basis holds at all. Same quantity as
             kept_variance.py, per item rather than per battery, at the pool's origin (D7).

  LANDING    where the retained part points: the constructs the POOL documents for the core items of
             the item's core components (fixed-point participation ratio down its own standardized
             row), reported as a description rather than scored. Nothing here compares a battery
             label with a pool label -- the two documentations name the same content differently, and
             forcing a match would need a pairing table this work avoids.

  STABILITY  mean pairwise overlap, over the 15 encoder pairs, of that landing description. Component
             indices are not comparable across encoders, so the constructs are the comparable object;
             this is the Figure 2b quantity.

  SPREAD     how many core components the item takes, and its largest |z|. A well-held item sits on
             one or two components at high |z|, a poorly held one spreads thinly over many.

The tobacco singles and the alcohol single from the same All of Us survey are printed as a worked
contrast: the pool fields alcohol through AUDIT and drugs through DAST-10 and holds no tobacco item
at all.

Reads saved artefacts only, recomputes no fit:
    outputs_pool/fits/<enc>__k*_ls1.0/{loadings,mean,scores}.npy
    outputs_pool/fixedpoint_cores/<enc>_cores.csv
    data/core151_items_<enc>.npz, core_items_151.csv, encoding_map_all151.csv
    ../randhrsdata/acidd_input/hrs_lb2022_<enc>.npz, hrs_lb2022_items.csv
    ../global_pool/pool_processed.csv

Run: CUDA_VISIBLE_DEVICES="" /data_gpu5/ROSS_Proj/env/bin/python porting_quality.py
"""
import csv
import glob
import itertools
import os

import numpy as np
import pandas as pd
from collections import Counter

from _common import keep_columns, pool_bands
from hrs_kept_components import fpc, ENCS, HERE, GP, HRSD

TOBACCO = ["electronicsmoking_electricsmokeparticipant", "smokelesstobacco_smokelesstobaccoparticipant",
           "smoking_100cigslifetime", "cigarsmoking_cigarsmokeparticipant",
           "hookahsmoking_hookahsmokeparticipant"]
ALCOHOL = "alcohol_alcoholparticipant"


def jac(a, b):
    return len(a & b) / len(a | b) if (a | b) else 0.0


def load_pool():
    pp = pd.read_csv(os.path.join(GP, "pool_processed.csv"))
    return list(pp.item_id), dict(zip(pp.item_id, pp.construct))


def battery(name):
    """item ids, embedding path template, and each item's documented construct."""
    if name == "All of Us":
        ai = pd.read_csv(os.path.join(HERE, "core_items_151.csv"))
        con = dict(zip(ai.item, ai.construct)) if "construct" in ai.columns else {}
        if not con:
            cm = pd.read_csv(os.path.join(HERE, "construct_map_151.csv"))
            con = dict(zip(cm.item, cm.construct))
        return os.path.join(HERE, "data", "core151_items_{enc}.npz"), con
    h = pd.read_csv(os.path.join(HRSD, "hrs_lb2022_items.csv"))
    return os.path.join(HRSD, "hrs_lb2022_{enc}.npz"), dict(zip(h.item, h.construct))


def per_encoder(enc, path_tmpl, bcon, pool_ids, pcon):
    fit = glob.glob(os.path.join(HERE, "outputs_pool", "fits", f"{enc}__k*_ls1.0"))[0]
    W = np.load(os.path.join(fit, "loadings.npy"))
    mu = np.load(os.path.join(fit, "mean.npy"))
    P = np.load(os.path.join(fit, "scores.npy"))
    sigma = P.std(0)
    Zpool = (P - P.mean(0)) / sigma
    cores = {int(r["component"]): str(r["items"]).split() for r in csv.DictReader(
        open(os.path.join(HERE, "outputs_pool", "fixedpoint_cores", f"{enc}_cores.csv")))}
    profile = {c: {pcon[h] for h in v} for c, v in cores.items()}

    d = np.load(path_tmpl.format(enc=enc), allow_pickle=True)
    ids = [str(x) for x in d["items"]]
    X = np.asarray(d["vectors"], float) - mu
    Z = (X @ W.T) / sigma

    kept = set(keep_columns(Z, pool_bands(enc, Zpool, pool_ids)))
    V = np.linalg.svd(W[sorted(kept)], full_matrices=False)[2]
    cov = ((X @ V.T) ** 2).sum(1) / (X ** 2).sum(1)

    rows = {}
    for i, g in enumerate(ids):
        c_mod = int(np.argmax(np.abs(Z[i])))
        F = [int(c) for c in fpc(Z[i])]
        prof = set().union(*[profile.get(c, set()) for c in F]) if F else set()
        lead = profile.get(c_mod, set())
        rows[g] = dict(coverage=float(cov[i]), profile=prof, modal=lead,
                       n_core=len(F), zmax=float(np.abs(Z[i]).max()))
    return rows


def kept_columns_table(name, enc="gte-large-en-v1.5"):
    """Every kept component, described in both vocabularies, with the energy it carries.

    Two descriptions of the same column, neither scored against the other: the constructs the POOL
    documents for the items that core it in the pool, and the constructs the BATTERY documents for
    its own items that core it there. Energy is the share of the battery's ported energy the column
    carries, sum_g z_gc^2 over the kept columns.
    """
    pool_ids, pcon = load_pool()
    path_tmpl, bcon = battery(name)
    fit = glob.glob(os.path.join(HERE, "outputs_pool", "fits", f"{enc}__k*_ls1.0"))[0]
    W = np.load(os.path.join(fit, "loadings.npy"))
    mu = np.load(os.path.join(fit, "mean.npy"))
    P = np.load(os.path.join(fit, "scores.npy"))
    sigma = P.std(0)
    Zpool = (P - P.mean(0)) / sigma
    cores = {int(r["component"]): str(r["items"]).split() for r in csv.DictReader(
        open(os.path.join(HERE, "outputs_pool", "fixedpoint_cores", f"{enc}_cores.csv")))}

    d = np.load(path_tmpl.format(enc=enc), allow_pickle=True)
    ids = [str(x) for x in d["items"]]
    X = np.asarray(d["vectors"], float) - mu
    Z = (X @ W.T) / sigma
    kept = sorted(set(keep_columns(Z, pool_bands(enc, Zpool, pool_ids))))

    # battery-side core items of a column: the fixed-point PR core down that column, within S
    bside = {c: [ids[j] for j in fpc(Z[:, c])] for c in kept}
    energy = (Z[:, kept] ** 2).sum(0)
    share = 100 * energy / energy.sum()

    def names(counter, n=3):
        return ", ".join(f"{k}" for k, _ in counter.most_common(n))

    print(f"\n{'='*112}\n{name}: the {len(kept)} kept components, both vocabularies ({enc})\n")
    print(f"  {'col':>4s} {'energy':>7s} {'n':>3s}  {'pool documents it as':44s}  battery documents it as")
    order = np.argsort(-share)
    for i in order:
        c = kept[i]
        pl = Counter(pcon[h] for h in cores.get(c, []))
        bl = Counter(bcon[g] for g in bside[c] if g in bcon)
        print(f"  C{c:<3d} {share[i]:6.1f}% {len(bside[c]):3d}  {names(pl)[:44]:44s}  {names(bl)[:44]}")
    print(f"\n  energy is the share of this battery's ported energy the column carries, and n the "
          f"number of battery items that core it")
    return [(kept[i], share[i], len(bside[kept[i]]),
             names(Counter(pcon[h] for h in cores.get(kept[i], []))),
             names(Counter(bcon[g] for g in bside[kept[i]] if g in bcon)))
            for i in order]


def summarize(name):
    pool_ids, pcon = load_pool()
    path_tmpl, bcon = battery(name)
    per = {e: per_encoder(e, path_tmpl, bcon, pool_ids, pcon) for e in ENCS}
    ids = list(per[ENCS[0]])

    T = pd.DataFrame({
        "item": ids,
        "coverage": [np.mean([per[e][g]["coverage"] for e in ENCS]) for g in ids],
        "n_core": [np.mean([per[e][g]["n_core"] for e in ENCS]) for g in ids],
        "zmax": [np.mean([per[e][g]["zmax"] for e in ENCS]) for g in ids],
        "stability": [np.mean([jac(per[a][g]["profile"], per[b][g]["profile"])
                               for a, b in itertools.combinations(ENCS, 2)]) for g in ids],
    }).set_index("item")

    print(f"\n{'='*92}\n{name}: {len(T)} items, six encoders\n")
    print(f"  coverage   median {100*T.coverage.median():.1f}%  "
          f"range {100*T.coverage.min():.1f}-{100*T.coverage.max():.1f}%")
    print(f"  stability  mean {T.stability.mean():.2f} (landing overlap across the 15 encoder pairs)")
    print(f"  spread     {T.n_core.mean():.1f} core components per item, largest |z| {T.zmax.mean():.1f} on average")

    T["tertile"] = pd.qcut(T.coverage, 3, labels=["low", "mid", "high"])
    print(f"\n  by coverage tertile\n  {'tertile':8s} {'n':>3s} {'coverage':>9s} {'stability':>10s} "
          f"{'core comps':>11s} {'|z|max':>8s}")
    for t, g in T.groupby("tertile", observed=True):
        print(f"  {str(t):8s} {len(g):3d} {100*g.coverage.mean():8.1f}% {g.stability.mean():10.2f} "
              f"{g.n_core.mean():11.1f} {g.zmax.mean():8.1f}")

    land = {g: ", ".join(sorted(per[ENCS[0]][g]["profile"])[:3]) for g in ids}
    print(f"\n  ten least-covered items, and where they land (gte-large)")
    for g, r in T.sort_values("coverage").head(10).iterrows():
        print(f"    {str(g)[:32]:32s} {100*r.coverage:5.1f}%  stab {r.stability:.2f}  "
              f"{r.n_core:3.1f} comps  |z|max {r.zmax:.1f}  -> {land[g][:52]}")
    print(f"\n  ten best-covered items, and where they land (gte-large)")
    for g, r in T.sort_values("coverage").tail(10).iterrows():
        print(f"    {str(g)[:32]:32s} {100*r.coverage:5.1f}%  stab {r.stability:.2f}  "
              f"{r.n_core:3.1f} comps  |z|max {r.zmax:.1f}  -> {land[g][:52]}")
    return T, land


def tobacco_contrast(T, land):
    print(f"\n  the five tobacco singles against the alcohol single from the same survey")
    for g in TOBACCO + [ALCOHOL]:
        if g not in T.index:
            print(f"    {g:32s} not in the battery")
            continue
        r = T.loc[g]
        print(f"    {g[:32]:32s} {100*r.coverage:5.1f}%  stab {r.stability:.2f}  "
              f"{r.n_core:3.1f} comps  |z|max {r.zmax:.1f}  -> {land[g][:52]}")


TEX = os.path.join(HERE, "..", "..", "manuscript_nhb", "porting_tables.tex")


def esc(s):
    for a, b in [("&", r"\&"), ("%", r"\%"), ("_", r"\_"), ("#", r"\#")]:
        s = str(s).replace(a, b)
    return s


def write_tex(TA, TH, colsA, colsH, land):
    def summary_rows(T, label):
        out = [f"{label} & all & {len(T)} & {100*T.coverage.median():.1f}\\% & "
               f"{100*T.coverage.min():.1f}--{100*T.coverage.max():.1f}\\% & {T.stability.mean():.2f} & "
               f"{T.n_core.mean():.1f} & {T.zmax.mean():.1f} \\\\"]
        for t, g in T.groupby("tertile", observed=True):
            out.append(f" & {t} & {len(g)} & {100*g.coverage.mean():.1f}\\% & --- & "
                       f"{g.stability.mean():.2f} & {g.n_core.mean():.1f} & {g.zmax.mean():.1f} \\\\")
        return out

    with open(TEX, "w") as f:
        f.write("% AUTO-GENERATED by examples/allofus/porting_quality.py -- do not edit\n")
        # ---- summary + tertiles
        f.write(r"\begin{table}[H]" "\n" r"\centering" "\n")
        f.write(r"\caption{\textbf{How much of each battery the basis holds, and how sharply it places "
                r"it.} \emph{Coverage} is the share of an item's energy about the pool's centre that "
                r"lies in the span of the kept components, averaged over the six encoders. "
                r"\emph{Stability} is the mean pairwise overlap, over the 15 encoder pairs, of the pool "
                r"constructs the item's core components carry. \emph{Core components} and $|z|_{\max}$ "
                r"describe how concentrated the placement is. Items are split into tertiles by "
                r"coverage.}\label{tab:porting_summary}" "\n")
        f.write(r"\small" "\n" r"\begin{tabular}{@{}l l r r c c c c@{}}" "\n" r"\toprule" "\n")
        f.write("Battery & items & $n$ & coverage & range & stability & core comps & $|z|_{\\max}$ \\\\\n")
        f.write(r"\midrule" "\n")
        for r in summary_rows(TA, "All of Us"):
            f.write(r + "\n")
        f.write(r"\midrule" "\n")
        for r in summary_rows(TH, "HRS"):
            f.write(r + "\n")
        f.write(r"\bottomrule" "\n" r"\end{tabular}" "\n" r"\end{table}" "\n\n")

        # ---- kept columns, both vocabularies
        for label, cols, lab in (("All of Us", colsA, "aou"), ("HRS", cols_h_lab := colsH, "hrs")):
            f.write(r"\begin{longtable}{@{}r r r >{\raggedright\arraybackslash}p{5.0cm} "
                    r">{\raggedright\arraybackslash}p{5.0cm}@{}}" "\n")
            f.write(r"\caption{\textbf{The components " + label + r" keeps, described in both "
                    r"documentations.} \emph{Energy} is the share of the battery's ported energy the "
                    r"component carries and $n$ the number of battery items that core it there. The "
                    r"last two columns are the leading constructs of the component's core items in the "
                    r"pool and in the battery, each taken from its own documentation and neither "
                    r"scored against the other (gte-large exemplar).}"
                    r"\label{tab:porting_cols_" + lab + r"}\\" "\n")
            f.write(r"\toprule" "\n"
                    "Component & energy & $n$ & the pool documents it as & " + label +
                    " documents it as \\\\\n" r"\midrule\endfirsthead" "\n")
            f.write(r"\toprule" "\n"
                    "Component & energy & $n$ & the pool documents it as & " + label +
                    " documents it as \\\\\n" r"\midrule\endhead" "\n"
                    r"\bottomrule\endfoot" "\n")
            for c, sh, n, pl, bl in cols:
                f.write(f"C{c} & {sh:.1f}\\% & {n} & {esc(pl)} & {esc(bl)} \\\\\n")
            f.write(r"\end{longtable}" "\n\n")

        # ---- tobacco contrast
        f.write(r"\begin{table}[H]" "\n" r"\centering" "\n")
        f.write(r"\caption{\textbf{Content the pool does not hold, against content it does.} The five "
                r"All of Us tobacco singles and the alcohol single administered in the same survey. "
                r"The pool fields alcohol through the AUDIT and drug use through the DAST-10 and "
                r"contains no item mentioning smoking, tobacco, cigarettes, vaping or nicotine. "
                r"Columns as in Table~\ref{tab:porting_summary}, with the landing taken under the "
                r"gte-large exemplar.}\label{tab:porting_tobacco}" "\n")
        # The All of Us variable names are long enough to run past the margin in an "l" column,
        # so the item column wraps and the name is allowed to break after each underscore.
        f.write(r"\footnotesize" "\n" r"\begin{tabular}{@{}>{\raggedright\arraybackslash}p{4.4cm} r c c c >{\raggedright\arraybackslash}p{4.0cm}@{}}"
                "\n" r"\toprule" "\n")
        f.write("Item & coverage & stability & core comps & $|z|_{\\max}$ & lands on \\\\\n")
        f.write(r"\midrule" "\n")
        for g in TOBACCO + [ALCOHOL]:
            if g not in TA.index:
                continue
            r = TA.loc[g]
            name = esc(g).replace(r"\_", r"\_\allowbreak ")
            f.write(f"\\texttt{{{name}}} & {100*r.coverage:.1f}\\% & {r.stability:.2f} & "
                    f"{r.n_core:.1f} & {r.zmax:.1f} & {esc(land[g])} \\\\\n")
        f.write(r"\bottomrule" "\n" r"\end{tabular}" "\n" r"\end{table}" "\n")
    print(f"\nwrote {os.path.normpath(TEX)}")


def main():
    A, land = summarize("All of Us")
    tobacco_contrast(A, land)
    colsA = kept_columns_table("All of Us")
    H, _ = summarize("HRS")
    colsH = kept_columns_table("HRS")
    write_tex(A, H, colsA, colsH, land)


if __name__ == "__main__":
    main()
```
