## Supplemental code for "Scoring respondents on a semantic frame of reference identifies a group that is burdened across many questionnaires and reaches care less easily": random_starts.py

```
#!/usr/bin/env python
"""Random-restart sweep for one encoder and one lambda_score.

100 random orthonormal initializations (seeds) of ROSS-Proj at a fixed
lambda_score, to probe whether the (non-convex) projection-sparsifying fit has a
single basin or many as a function of lambda_score. For each seed we save the
loadings W and the projection P (= FitResult.Z, the 151 x k item projection) so
VE and PR (one-time defining-set + iterated fixed-point) can be computed here and
re-derived downstream.

Only the initialization varies across seeds: with batch_size = n (full batch) and
subsample_fraction = 1.0 the robust scale and the single minibatch are
deterministic, so random_state feeds *only* _init_random_normalized.

Layout:
    <out>/<encoder>__k{k}_ls{ls}/seed{seed:03d}/W.npy      (k x D loadings)
    <out>/<encoder>__k{k}_ls{ls}/seed{seed:03d}/P.npy      (n x k projection)
    <out>/<encoder>__k{k}_ls{ls}/metrics.csv               (one row per seed)

Run (one GPU, one lambda):
    CUDA_VISIBLE_DEVICES=0 python random_starts.py --lambda-score 0.01 --seeds 0-99
See run_random_starts.sh for the 5-GPU fan-out.
"""
import argparse
import csv
import json
import os
import time

import numpy as np

from ross_proj import fit_projection
from ross_proj.defining_sets import participation_ratio  # (Sz^2)^2 / Sz^4 per component

from _common import EMBEDDERS, OUT_DIR, load_embedding


def _prk(sq, k):
    """PR over the top-k already-sorted (descending) squared loadings."""
    a = sq[:max(k, 1)]
    return a.sum() ** 2 / ((a ** 2).sum() + 1e-300)


def fixed_point_core_size(col):
    """PR fixed-point core size of one component column (== result2_figures.py).

    Start at round(PR over all items), iterate k <- round(PR of top-k by loading^2)
    to convergence. Non-increasing (PR(top-k) <= k), so it converges.
    """
    sq = np.sort(np.asarray(col, float) ** 2)[::-1]
    k = int(round(_prk(sq, len(sq))))
    for _ in range(200):
        kn = int(round(_prk(sq, k)))
        if kn == k:
            break
        k = kn
    return max(k, 1)


def variance_explained(X, mean, W, Z):
    """Joint VE at theta=0: 1 - ||Xc - Z W||^2 / ||Xc||^2  (W is k x D, Z is n x k)."""
    Xc = X - mean
    recon = Z @ W
    return 1.0 - np.sum((Xc - recon) ** 2) / np.sum(Xc ** 2)


def parse_seeds(spec):
    """'0-99' -> range(0,100); '0,5,7' -> [0,5,7]; '10' -> [10]."""
    if "-" in spec:
        a, b = spec.split("-")
        return list(range(int(a), int(b) + 1))
    return [int(s) for s in spec.split(",")]


def resolve_k(name, k_arg):
    if k_arg != "auto":
        return int(k_arg)
    path = os.path.join(OUT_DIR, "k_selection", name, "parallel_analysis.json")
    if not os.path.exists(path):
        raise SystemExit(f"missing {path} — run 1_select_k.py first, or pass --k <int>")
    return int(json.load(open(path))["k"])


def main():
    ap = argparse.ArgumentParser()
    ap.add_argument("--encoder", default="gte-large-en-v1.5", choices=list(EMBEDDERS))
    ap.add_argument("--lambda-score", type=float, required=True)
    ap.add_argument("--lambda-ortho", type=float, default=0.1)
    ap.add_argument("--k", default="auto",
                    help="'auto' (read stage-1 k) or an integer")
    ap.add_argument("--seeds", default="0-99", help="e.g. '0-99' or '0,5,7'")
    ap.add_argument("--total-steps", type=int, default=5000)
    ap.add_argument("--out-dir", default=os.path.join(OUT_DIR, "random_starts"))
    ap.add_argument("--overwrite", action="store_true",
                    help="refit seeds even if W.npy/P.npy already exist")
    args = ap.parse_args()

    name, ls = args.encoder, args.lambda_score
    items, X = load_embedding(name)
    n = X.shape[0]
    k = resolve_k(name, args.k)
    seeds = parse_seeds(args.seeds)

    tag = f"{name}__k{k}_ls{ls}"
    root = os.path.join(args.out_dir, tag)
    os.makedirs(root, exist_ok=True)
    metrics_path = os.path.join(root, "metrics.csv")
    new_file = not os.path.exists(metrics_path)
    mf = open(metrics_path, "a", newline="")
    writer = csv.writer(mf)
    if new_file:
        writer.writerow(["encoder", "k", "lambda_score", "seed", "ve",
                         "pr_onetime_median", "pr_onetime_mean",
                         "pr_fixed_median", "pr_fixed_mean", "n_steps", "secs"])
        mf.flush()

    print(f"[{tag}] {len(seeds)} seeds on {n}x{X.shape[1]}, k={k}, "
          f"lambda_score={ls}", flush=True)

    for seed in seeds:
        sd = os.path.join(root, f"seed{seed:03d}")
        wnpy, pnpy = os.path.join(sd, "W.npy"), os.path.join(sd, "P.npy")
        if not args.overwrite and os.path.exists(wnpy) and os.path.exists(pnpy):
            print(f"  seed {seed:3d}  skip (exists)", flush=True)
            continue
        os.makedirs(sd, exist_ok=True)

        t0 = time.time()
        res = fit_projection(
            X, n_components=k, lambda_score=ls, lambda_ortho=args.lambda_ortho,
            init="random_normalized", random_state=seed,
            batch_size=n, subsample_fraction=1.0, total_steps=args.total_steps,
            verbose=False, log_interval=0)
        secs = time.time() - t0

        W, P = np.asarray(res.W), np.asarray(res.Z)   # (k,D) loadings ; (n,k) projection
        np.save(wnpy, W)
        np.save(pnpy, P)

        ve = variance_explained(X, res.mean, W, P)
        pr1 = participation_ratio(P)                              # one-time PR per component
        prf = np.array([fixed_point_core_size(P[:, c]) for c in range(k)])
        n_steps = int(getattr(res.model, "n_steps_", -1))
        writer.writerow([name, k, ls, seed, f"{ve:.6f}",
                         f"{np.median(pr1):.4f}", f"{np.mean(pr1):.4f}",
                         f"{np.median(prf):.4f}", f"{np.mean(prf):.4f}",
                         n_steps, f"{secs:.1f}"])
        mf.flush()
        print(f"  seed {seed:3d}  VE={ve:.4f}  PR1_med={np.median(pr1):5.1f}  "
              f"PRfix_med={np.median(prf):4.1f}  [{secs:.1f}s]", flush=True)

    mf.close()
    print(f"[{tag}] done -> {root}", flush=True)


if __name__ == "__main__":
    main()
```
