## Supplemental code for "Scoring respondents on a semantic frame of reference identifies a group that is burdened across many questionnaires and reaches care less easily": README.md

# examples/allofus — the pipeline and the local generators

**The step-by-step is [`../../README.md`](../../README.md)**: the environment, the four stages in
order, and which of them need cohort access. This file only says what lives in this directory.

Everything here runs on published question text. Nothing here reads a response, so all of it is
reproducible without All of Us access. The scripts that need responses are in `../../all_of_us_codes/`
and run inside the Researcher Workbench.

## Two bases, and why the scripts look generic

The stage scripts (`0_embed.py`, `1_select_k.py`, `2_fit.py`) are basis-agnostic. They read
`data/<prefix>_<encoder>.npz` and write under `ROSS_OUT_DIR`, both switchable:

```bash
export ROSS_NPZ_PREFIX=pool_items
export ROSS_OUT_DIR=/data_gpu5/ROSS_Proj/examples/allofus/outputs_pool
```

**Set both.** Their defaults (`core151_items`, `outputs_core151`) are the **retired** 151-item own
basis, with 23 to 26 components — an object the manuscript no longer contains. The live object is the
577-item reference pool with 49 to 55 components, and every result in the paper is computed on it.

## What is here

| group | scripts |
|---|---|
| the basis | `0_embed.py`, `1_select_k.py`, `2_fit.py`, `random_starts.py`, `select_projection.py`, `pool_fixedpoint_cores.py`, `pool_core_columns.py`, `build_reference_basis_package.py` |
| the penalty weight | `lambda_sweep_pool.py`, `table_lambda_sweep.py` |
| placing a battery | `build_aou_poolw_projection.py`, `pool_project_aou.py`, `make_poolw_basis.py`, `make_core_items_pool.py`, `hrs_kept_components.py` |
| shared definitions | `_common.py` — `pool_bands` and `keep_columns` are the keep rule of record, and no script may carry its own copy |
| figures | `fig1_pool.py`, `fig1_panels.py`, `fig_pool_ribbon.py`, `fig_xenc.py`, `fig_aou_twoside_cores.py`, `fig_hrs_twoside_cores.py`, `fig_hrs_loadings.py`, `fig_corecomp.py`, `fig_coverage.py`, `fig_portability.py`, `fig_contentgrammar_3panel.py` |
| tables and data | `supp_pool_composition.py`, `table_dense_control.py`, `table_phrasing_xenc.py`, `table_faithfulness_disagreements.py`, `porting_quality.py` |
| analyses named in Methods | `construct_splitting.py`, `core_separation.py`, `dense_control.py`, `pool_xenc_itemside_profiles.py`, `pool_xenc_hungarian.py`, `hrs_xenc_itemside_profiles.py`, `pool_colocation.py` |
| content versus syntax | `content_grammar_manifest.py`, `generate_variants.py`, `content_grammar_poolbasis.py` |

`../../manuscript_nhb/ANALYSIS_SCRIPTS.md` maps each result in the paper to its script, and
`FIGURES.md` beside it maps each figure and table to its generator.

## Two traps

**`fig_aou_twoside_cores.py` and `fig_hrs_twoside_cores.py` write into
`../global_pool/figures/`**, not into the manuscript. Their output must be copied to
`../../manuscript_nhb/figures/` by hand after every run, or the manuscript silently keeps the old
image.

**Fits are device-dependent.** A GPU fit and a CPU fit of the same data give different bases. The
published basis is the CPU fit, so reproduce on CPU.

> This directory held 147 scripts until 2026-08-31. The 101 that no longer serve the manuscript —
> most of them from the retired own basis — are in `../../archive/superseded_20260831/allofus/`,
> with an inventory in its `MOVED.txt`. Nothing here imports them.
