## Supplemental code for "Scoring respondents on a semantic frame of reference identifies a group that is burdened across many questionnaires and reaches care less easily": select_projection.py

```
#!/usr/bin/env python
"""Select the operating projection P from the random-restart sweep (writes selection_best.json).

For each encoder we fit the concentrated ROSS-PCA projection from 100 random orthonormal
initializations at each penalty weight lambda_score in {0.01,0.05,0.1,0.5,1} (random_starts.py).
This script applies the manuscript's selection rule:

  among all fits that keep W orthonormal (max off-diagonal |cos|, rounded to 3 decimals, <= ORTHO_MAX)
  and preserve
  explained variance (VE >= VE_FRAC * VE_PCA, i.e. within 1% of the PCA maximum), retain the
  fit that MAXIMIZES the median core energy (fraction of a component's squared weight on its
  participation-ratio fixed-point core).

It writes selection_best.json with, per encoder: k, selected lambda_score (ls), seed, path, the
selected fit's (mean_core, coreE median, ve, ortho), the PCA-initialized lambda=0.01 baseline
(op_*), and the dense-PCA lambda=0 baseline (dense_*).

Run from examples/allofus/ :
  ROSS_SEL_OUT=/path/to/selection_best.json ../../env/bin/python select_projection.py
Default writes outputs_core151/random_starts/selection_best.json. Set ROSS_SEL_OUT to compare
without overwriting. Prints a comparison against any existing selection_best.json.
"""
import csv, glob, json, os, re
import numpy as np

HERE = os.path.dirname(os.path.abspath(__file__))
OUT_DIR = os.path.join(HERE, "outputs_core151")
RS_DIR = os.path.join(OUT_DIR, "random_starts")
FITS = os.path.join(OUT_DIR, "fits")
PCA_CSV = os.path.join(OUT_DIR, "pca_baseline.csv")
CANON = os.path.join(RS_DIR, "selection_best.json")
OUT = os.environ.get("ROSS_SEL_OUT", CANON)

ORTHO_MAX = 0.01      # W orthonormal: largest off-diagonal |cos|, rounded to 3 decimals, <= 0.01
VE_FRAC = 0.99        # explained variance within 1% of the PCA maximum
LAMBDAS = [0.01, 0.05, 0.1, 0.5, 1.0]
ENCODERS = ["bge-m3", "bge-large-en-v1.5", "all-mpnet-base-v2",
            "e5-large-v2", "gte-large-en-v1.5", "qwen3-embedding-0.6b"]


def participation_ratio(col):
    sq = col.astype(float) ** 2
    s1 = sq.sum()
    return 0.0 if s1 <= 0 else float(s1 * s1 / (sq * sq).sum())


def fixed_point_core_size(col):
    sq = np.sort(col.astype(float) ** 2)[::-1]
    k = max(1, min(int(round(participation_ratio(col))), len(sq)))
    for _ in range(1000):
        top = sq[:k]; s1 = top.sum()
        prk = 1.0 if s1 <= 0 else float(s1 * s1 / (top * top).sum())
        kn = max(1, min(int(round(prk)), len(sq)))
        if kn == k:
            break
        k = kn
    return k


def core_metrics(P):
    """(median core energy, mean core size) over the columns of P."""
    ce, cs = [], []
    for c in range(P.shape[1]):
        col = P[:, c]; sq = col.astype(float) ** 2; tot = sq.sum()
        s = fixed_point_core_size(col)
        cs.append(s)
        ce.append(0.0 if tot <= 0 else float(np.sort(sq)[::-1][:s].sum() / tot))
    return float(np.median(ce)), float(np.mean(cs))


def ortho_maxcos(W):
    k = W.shape[0]
    Wn = W / np.linalg.norm(W, axis=1, keepdims=True)
    C = Wn @ Wn.T
    return float(np.abs(C[~np.eye(k, dtype=bool)]).max())


def load_pca():
    ve_pca, k_of = {}, {}
    with open(PCA_CSV) as f:
        for r in csv.DictReader(f):
            ve_pca[r["embedder"]] = float(r["ve_PCA"]); k_of[r["embedder"]] = int(r["k"])
    return ve_pca, k_of


def baseline_metrics(enc, k):
    """(coreE_median, mean_core, ve, ortho) for a fits/ projection, or None if absent."""
    def one(tag, ve_key=None):
        d = os.path.join(FITS, f"{enc}__k{k}_ls{tag}")
        P = np.load(os.path.join(d, "scores.npy"))
        W = np.load(os.path.join(d, "loadings.npy"))
        cfg = json.load(open(os.path.join(d, "config.json")))
        ce, mc = core_metrics(P)
        return dict(coreE=ce, mean_core=mc, ve=float(cfg.get("ve_subspace", np.nan)), ortho=ortho_maxcos(W))
    return one("0.0"), one("0.01")


def main():
    ve_pca, k_of = load_pca()
    existing = json.load(open(CANON)) if os.path.exists(CANON) else {}
    out = {}
    print(f"{'encoder':22s} {'sel ls':>6s} {'seed':>4s} {'coreE':>6s} {'core':>5s} {'ve':>6s} {'ortho':>7s}"
          f"  | {'passers':>7s}  vs current")
    for enc in ENCODERS:
        k = k_of[enc]
        dense, op = baseline_metrics(enc, k)
        best, n_pass, n_tot = None, 0, 0
        for ls in LAMBDAS:
            lsdir = os.path.join(RS_DIR, f"{enc}__k{k}_ls{ls}")   # dir uses str(float): 1.0 -> "1.0"
            mfile = os.path.join(lsdir, "metrics.csv")
            ve_by_seed = {}
            if os.path.exists(mfile):
                for r in csv.DictReader(open(mfile)):
                    ve_by_seed[int(r["seed"])] = float(r["ve"])
            for sd in sorted(glob.glob(os.path.join(lsdir, "seed*"))):
                seed = int(re.search(r"seed(\d+)", sd).group(1))
                P = np.load(os.path.join(sd, "P.npy"))
                W = np.load(os.path.join(sd, "W.npy"))
                n_tot += 1
                ve = ve_by_seed.get(seed, np.nan)
                orth = ortho_maxcos(W)
                if not (round(orth, 3) <= ORTHO_MAX and ve >= VE_FRAC * ve_pca[enc]):
                    continue
                n_pass += 1
                ce, mc = core_metrics(P)
                cand = dict(k=k, ls=ls, seed=seed, mean_core=mc, coreE=ce, ve=ve, ortho=orth,
                            path=os.path.relpath(sd, os.path.join(HERE, "..", "..")))
                if best is None or ce > best["coreE"]:
                    best = cand
        best.update(op_mean_core=op["mean_core"], op_coreE=op["coreE"], op_ve=op["ve"], op_ortho=op["ortho"],
                    dense_coreE=round(dense["coreE"], 4), dense_mean_core=round(dense["mean_core"], 4),
                    dense_ve=round(ve_pca[enc], 4))
        out[enc] = best
        cur = existing.get(enc, {})
        same = cur.get("ls") == best["ls"] and cur.get("seed") == best["seed"]
        flag = "SAME" if same else f"CHANGED (was ls{cur.get('ls')} s{cur.get('seed')})"
        print(f"{enc:22s} {best['ls']:>6g} {best['seed']:>4d} {best['coreE']:>6.3f} {best['mean_core']:>5.2f} "
              f"{best['ve']:>6.3f} {best['ortho']:>7.4f}  | {n_pass:>3d}/{n_tot:<3d}  {flag}")

    json.dump(out, open(OUT, "w"), indent=2)
    print(f"\nwrote {OUT}")


if __name__ == "__main__":
    main()
```
