## Supplemental code for "Scoring respondents on a semantic frame of reference identifies a group that is burdened across many questionnaires and reaches care less easily": supp_pool_composition.py

```
#!/usr/bin/env python
"""Supplementary tables documenting the external reference pool (Supplementary Data 4).

Reads the shipped supplementary data only -- no fits, no hardcoded counts -- and writes
the LaTeX table that supplement.tex \\input's:

  pool_core_summary_table.tex  the per-encoder component summary

The per-instrument table this also used to write was retired: it restated
supplementary_data/global_pool/pool_instruments.csv, which ships as Supplementary Data 4, so
Supplementary Section S18 now points at that file instead.

Run:
    cd examples/allofus
    CUDA_VISIBLE_DEVICES="" /data_gpu5/ROSS_Proj/env/bin/python supp_pool_composition.py
"""
import csv
import os

HERE = os.path.dirname(os.path.abspath(__file__))
DATA = os.path.join(HERE, "..", "..", "manuscript_nhb", "supplementary_data", "global_pool")
OUT = os.path.join(HERE, "..", "..", "manuscript_nhb")

ENCODER_ORDER = [
    "gte-large-en-v1.5",
    "bge-large-en-v1.5",
    "bge-m3",
    "e5-large-v2",
    "all-mpnet-base-v2",
    "qwen3-embedding-0.6b",
]


def tex(s):
    """Escape the LaTeX specials that occur in instrument names and construct labels."""
    for a, b in [("\\", r"\textbackslash{}"), ("&", r"\&"), ("%", r"\%"), ("$", r"\$"),
                 ("#", r"\#"), ("_", r"\_"), ("{", r"\{"), ("}", r"\}"), ("~", r"\textasciitilde{}"),
                 ("^", r"\textasciicircum{}")]:
        s = s.replace(a, b)
    # House rule: no semicolons anywhere in the manuscript. The construct profile separator
    # becomes a comma, which stays unambiguous because every construct carries its own count.
    return s.replace(";", ",")


def read(name):
    with open(os.path.join(DATA, name)) as fh:
        return list(csv.DictReader(fh))


def core_summary_table(rows):
    by_enc = {r["encoder"]: r for r in rows}
    order = [e for e in ENCODER_ORDER if e in by_enc] + [
        e for e in sorted(by_enc) if e not in ENCODER_ORDER
    ]
    lines = [
        r"\begin{table}[H]",
        r"\centering",
        r"\caption{\textbf{The pool's components under each encoder.} The number of components, the"
        r" size of their cores, the fraction of the 577 pool items that are a core item of at least"
        r" one component, and how many distinct documented constructs a core spans. The per-component"
        r" composition, with the full text of every core item, is Supplementary Data~4"
        r" (\texttt{cores/}).}\label{tab:pool_core_summary}",
        r"\small",
        r"\begin{tabular}{@{}l r c r r c@{}}",
        r"\toprule",
        r"Encoder & Components & Core size & Coverage & Constructs & Single-construct \\",
        r"& & median [range] & & per core & cores \\",
        r"\midrule",
    ]
    for e in order:
        r = by_enc[e]
        lines.append(
            f"\\texttt{{\\small {tex(e)}}} & {r['n_components']} & "
            f"{r['median_core']} [{r['min_core']}--{r['max_core']}] & "
            f"{float(r['coverage']):.2f} & {r['median_constructs']} & {r['n_single_construct']} \\\\"
        )
    lines += [r"\bottomrule", r"\end{tabular}", r"\end{table}"]
    return "\n".join(lines)


def main():
    items = read("pool_items.csv")
    instruments = read("pool_instruments.csv")
    summary = read("pool_core_summary.csv")

    for name, body in [
        ("pool_core_summary_table.tex", core_summary_table(summary)),
    ]:
        path = os.path.join(OUT, name)
        with open(path, "w") as fh:
            fh.write("% generated by examples/allofus/supp_pool_composition.py -- do not edit\n")
            fh.write(body + "\n")
        print("wrote", os.path.relpath(path, os.path.join(HERE, "..", "..")))

    # The counts quoted in the prose of the section, printed so they can be checked against it.
    def tally(rows, key):
        out = {}
        for r in rows:
            out[r[key]] = out.get(r[key], 0) + 1
        return dict(sorted(out.items(), key=lambda kv: -kv[1]))

    print("\nprose check")
    print("  items                ", len(items))
    print("  instruments          ", len(instruments))
    print("  documented constructs", len({r["documented_construct"] for r in items}))
    print("  item role            ", tally(items, "role"))
    print("  instrument role      ", tally(instruments, "role"))
    print("  label level          ", tally(items, "label_level"))
    print("  provenance           ", tally(items, "provenance"))
    print("  verification         ", tally(instruments, "verification"))
    print("  stems removed        ", sum(1 for r in items if r["stem_removed"].strip()))


if __name__ == "__main__":
    main()
```
