## Supplemental code for "Scoring respondents on a semantic frame of reference identifies a group that is burdened across many questionnaires and reaches care less easily": table_dense_control.py

```
#!/usr/bin/env python
"""Supplementary table: the dense-basis control, under all six encoders.

dense_control.py fits, for one encoder, a dense basis of the same size on the same pool items and
scores both bases for faithfulness against their own nulls (Methods "Dense-basis control"). It writes
one JSON per encoder. This script only collects those six files into the supplementary table, so it
refits nothing and recomputes nothing.

Reads:  outputs_pool/dense_control_<enc>.json   (written by dense_control.py, one run per encoder)
Writes: manuscript_nhb/dense_control_table.tex, and prints the ranges Supplementary Section S20 quotes.

Run: /data_gpu5/ROSS_Proj/env/bin/python table_dense_control.py
"""
import os, json
import numpy as np

from hrs_kept_components import ENCS, HERE

OUT = os.path.join(HERE, "..", "..", "manuscript_nhb", "dense_control_table.tex")
ENC_LABEL = {"gte-large-en-v1.5": "gte-large", "bge-m3": "bge-m3",
             "bge-large-en-v1.5": "bge-large", "all-mpnet-base-v2": "all-mpnet",
             "e5-large-v2": "e5-large", "qwen3-embedding-0.6b": "qwen3"}


def main():
    rows = []
    for enc in ENCS:
        path = os.path.join(HERE, "outputs_pool", f"dense_control_{enc}.json")
        d = json.load(open(path))
        s, dn = d["sparse"], d["dense"]
        rows.append(dict(
            enc=enc, k=d["k"],
            s_margin=100 * s["all"]["margin"], d_margin=100 * dn["all"]["margin"],
            s_single=s["constructs_per_component"]["single_construct"],
            d_single=dn["constructs_per_component"]["single_construct"],
            s_cpc=s["constructs_per_component"]["mean"],
            d_cpc=dn["constructs_per_component"]["mean"]))

    print(f"{'encoder':12s} {'k':>3s} {'sparse':>7s} {'dense':>7s}   single-construct   constructs/component")
    for r in rows:
        print(f"{ENC_LABEL[r['enc']]:12s} {r['k']:3d} {r['s_margin']:6.1f}% {r['d_margin']:6.1f}%   "
              f"{r['s_single']:2d} vs {r['d_single']:2d} of {r['k']:2d}    "
              f"{r['s_cpc']:.2f} vs {r['d_cpc']:.2f}")
    print(f"\nsparse margin {min(r['s_margin'] for r in rows):.1f} to {max(r['s_margin'] for r in rows):.1f}, "
          f"dense {min(r['d_margin'] for r in rows):.1f} to {max(r['d_margin'] for r in rows):.1f}")
    print(f"single-construct components: sparse {min(r['s_single'] for r in rows)} to "
          f"{max(r['s_single'] for r in rows)}, dense {min(r['d_single'] for r in rows)} to "
          f"{max(r['d_single'] for r in rows)}")

    with open(OUT, "w") as f:
        f.write("% AUTO-GENERATED by examples/allofus/table_dense_control.py -- do not edit\n")
        f.write(r"\begin{table}[H]" "\n" r"\centering" "\n")
        f.write(r"\caption{\textbf{The dense-basis control, under each encoder.} For each encoder, a "
                r"dense basis of the same size is fit on the same pool items and given the same item "
                r"budget, and both bases are scored for faithfulness by the same rule "
                r"(Methods~\S\ref{sec:methods_dense}). \emph{Margin} is the faithfulness rate above "
                r"that basis's own label-shuffle chance rate, which is the comparable quantity because "
                r"a basis whose components carry more constructs attains a higher raw rate by "
                r"construction. \emph{Single-construct} counts the components whose naming items all "
                r"carry one documented construct, and \emph{constructs per component} averages how many "
                r"each names.}\label{tab:dense_control}" "\n")
        f.write(r"\small" "\n" r"\begin{tabular}{@{}l c c c c c c@{}}" "\n" r"\toprule" "\n")
        f.write(r"& & \multicolumn{2}{c}{Margin} & \multicolumn{2}{c}{Single-construct} & "
                r"\multicolumn{1}{c}{Constructs/component} \\" "\n")
        f.write(r"\cmidrule(lr){3-4}\cmidrule(lr){5-6}\cmidrule(l){7-7}" "\n")
        f.write("Encoder & $k$ & sparse & dense & sparse & dense & sparse vs dense \\\\\n")
        f.write(r"\midrule" "\n")
        for r in rows:
            f.write(f"{ENC_LABEL[r['enc']]} & {r['k']} & {r['s_margin']:.1f}\\% & {r['d_margin']:.1f}\\% & "
                    f"{r['s_single']} & {r['d_single']} & {r['s_cpc']:.2f} vs {r['d_cpc']:.2f} \\\\\n")
        f.write(r"\bottomrule" "\n" r"\end{tabular}" "\n" r"\end{table}" "\n")
    print("\nwrote", os.path.normpath(OUT))


if __name__ == "__main__":
    main()
```
