## Supplemental code for "Scoring respondents on a semantic frame of reference identifies a group that is burdened across many questionnaires and reaches care less easily": table_faithfulness_disagreements.py

```
#!/usr/bin/env python
"""Supplementary table: items whose core components do not name their own documented construct.

Faithfulness, at the construct level only (no broader grouping): an item is faithful when its
documented construct appears among the constructs of the core items of its core components. The
item's own membership of a core is excluded, so an item that anchors a core is not counted faithful
merely for being there.

    F_g     core components of item g        fixed-point PR down its row of Z_G
    E_i     core items of component i        fixed-point PR down column i of Z_G
    faithful(g)  <=>  C_g in { C_h : i in F_g, h in E_i, h != g }

Reads only saved artefacts - no item ids, constructs or counts are hardcoded:
    outputs_pool/fits/<enc>__k*_ls1.0/scores.npy
    outputs_pool/fixedpoint_cores/<enc>_cores.csv
    ../global_pool/pool_processed.csv

Writes manuscript_nhb/supplementary_data/faithfulness_disagreements.csv (Supplementary Data 6: every
item whose documented construct is absent from the constructs of its core components, flagged by
whether it anchors a core) and prints every summary number Supplementary Section S12 quotes. This was
Supplementary Table S6, which listed only the never-core subset.

Run: CUDA_VISIBLE_DEVICES="" /data_gpu5/ROSS_Proj/env/bin/python table_faithfulness_disagreements.py
"""
import os, glob, csv
import numpy as np
import pandas as pd

HERE = os.path.dirname(os.path.abspath(__file__))
REPO = os.path.abspath(os.path.join(HERE, "..", ".."))
GP = os.path.join(HERE, "..", "global_pool")
OUT = os.path.join(REPO, "manuscript_nhb", "supplementary_data", "faithfulness_disagreements.csv")
ENC = "gte-large-en-v1.5"
NPERM = 200


def pr(v):
    s2 = float((v ** 2).sum()); s4 = float((v ** 4).sum())
    return s2 * s2 / s4 if s4 > 0 else 0.0


def fpc(v):
    idx = np.arange(len(v))
    while True:
        n = max(1, min(int(round(pr(v[idx]))), len(idx)))
        top = np.sort(idx[np.argsort(-np.abs(v[idx]))[:n]])
        if len(top) == len(idx) and (top == idx).all():
            return top
        idx = top


def main():
    pp = pd.read_csv(os.path.join(GP, "pool_processed.csv"))
    con = dict(zip(pp.item_id, pp.construct))
    txt = dict(zip(pp.item_id, pp.prompt_raw))
    ids = list(pp.item_id)
    fit = glob.glob(os.path.join(HERE, "outputs_pool", "fits", f"{ENC}__k*_ls1.0"))[0]
    Z = np.load(os.path.join(fit, "scores.npy"))
    Zt = Z / Z.std(0)
    core = {int(r["component"]): str(r["items"]).split() for r in
            csv.DictReader(open(os.path.join(HERE, "outputs_pool", "fixedpoint_cores", f"{ENC}_cores.csv")))}
    anchored = {h for v in core.values() for h in v}
    F = {g: [int(c) for c in fpc(Zt[j])] for j, g in enumerate(ids)}

    def constructs_of(g, lab):
        return {lab[h] for c in F[g] for h in core[c] if h != g}

    def rate(sub, lab):
        return float(np.mean([lab[g] in constructs_of(g, lab) for g in sub]))

    groups = [("all items", ids), ("anchors a core", [g for g in ids if g in anchored]),
              ("anchors none", [g for g in ids if g not in anchored])]
    rng = np.random.RandomState(0)
    print(f"{ENC}\n{'group':16s} {'n':>4s} {'faithful':>9s} {'chance':>8s}")
    for name, sub in groups:
        null = np.mean([rate(sub, dict(zip(ids, rng.permutation(list(con.values())))))
                        for _ in range(NPERM)])
        print(f"{name:16s} {len(sub):4d} {100*rate(sub, con):8.1f}% {100*null:7.1f}%")

    disagree = [g for g in ids if con[g] not in constructs_of(g, con)]
    never = [g for g in disagree if g not in anchored]
    print(f"\ndisagreements {len(disagree)} of {len(ids)}   anchoring {len(disagree)-len(never)}   never-core {len(never)}")
    print("\nnever-core disagreements by instrument:")
    print(pd.Series([pp.set_index('item_id').scale[g] for g in never]).value_counts().to_string())

    rows = []
    for g in sorted(disagree, key=lambda g: (con[g], g)):
        seen, names = set(), []
        for c in sorted(F[g], key=lambda c: -abs(Zt[ids.index(g), c])):
            counts = pd.Series([con[h] for h in core[c]]).value_counts()
            lead = "/".join(list(counts.index[:2]))
            if lead not in seen:
                seen.add(lead); names.append(lead)
        rows.append({
            "item_id": g,
            "item": str(txt[g]).strip(),
            "instrument": pp.set_index("item_id").scale[g],
            "documented_construct": con[g],
            "anchors_a_core": int(g in anchored),
            "constructs_of_core_components": ", ".join(names),
        })
    df = pd.DataFrame(rows)
    df.to_csv(OUT, index=False)
    print("\nwrote", OUT, f"| {len(df)} disagreements, "
          f"{int((df.anchors_a_core == 0).sum())} of them never-core")


if __name__ == "__main__":
    main()
```
