## Supplemental code for "Scoring respondents on a semantic frame of reference identifies a group that is burdened across many questionnaires and reaches care less easily": table_lambda_sweep.py

```
#!/usr/bin/env python
"""Supplementary table: the sparsity-weight sweep behind lambda_score = 1.

Reads outputs_pool/lambda_sweep.csv (written by lambda_sweep_pool.py) and emits a LaTeX longtable.
Nothing is hardcoded: every number, the encoder list and the lambda grid come from the CSV.

The faithfulness columns are reported, never used to choose the weight -- see lambda_sweep_pool.py.

Out: manuscript_nhb/lambda_sweep_table.tex
Run: CUDA_VISIBLE_DEVICES="" /data_gpu5/ROSS_Proj/env/bin/python table_lambda_sweep.py
"""
import os
import pandas as pd

HERE = os.path.dirname(os.path.abspath(__file__))
REPO = os.path.abspath(os.path.join(HERE, "..", ".."))
SRC = os.path.join(HERE, "outputs_pool", "lambda_sweep.csv")
OUT = os.path.join(REPO, "manuscript_nhb", "lambda_sweep_table.tex")

SHORT = {"all-mpnet-base-v2": "mpnet", "bge-large-en-v1.5": "bge-large", "bge-m3": "bge-m3",
         "e5-large-v2": "e5-large", "gte-large-en-v1.5": "gte-large",
         "qwen3-embedding-0.6b": "qwen3"}


def main():
    df = pd.read_csv(SRC).sort_values(["encoder", "lambda_score"])
    lams = sorted(df.lambda_score.unique())
    encs = sorted(df.encoder.unique())

    with open(OUT, "w") as f:
        f.write("% AUTO-GENERATED by examples/allofus/table_lambda_sweep.py -- do not edit\n")
        f.write("{\\small\n\\setlength{\\LTcapwidth}{\\textwidth}\n")
        f.write("\\begin{longtable}{@{}l r r r r r r r@{}}\n")
        f.write("\\caption{\\textbf{The sparsity-weight sweep.} For each encoder the basis is refit "
                "independently from the principal components at each $\\lambda_{\\mathrm{score}}$, with "
                "$\\lambda_{\\mathrm{ortho}}=0.1$ and $k$ from parallel analysis. \\emph{ortho} is the "
                "largest off-diagonal cosine between the columns of $W$, \\emph{core} the median core "
                "size in items, \\emph{energy} the median core energy, and \\emph{VE/PCA} the variance "
                "the $k$ components retain relative to principal component analysis at the same $k$. "
                "These four use the embeddings alone and are what the weight was chosen on. The "
                "faithfulness columns, the rate at which an item's documented construct appears among "
                "the constructs of its core components and the chance rate from a label shuffle, are "
                "reported as a consequence of the weight and took no part in choosing it.}\n")
        f.write("\\label{tab:lambda_sweep}\\\\\n\\toprule\n")
        head = ("Encoder & $\\lambda_{\\mathrm{score}}$ & ortho & core & energy & VE/PCA & "
                "faithful & chance \\\\\n")
        f.write(head + "\\midrule\\endfirsthead\n")
        f.write("\\multicolumn{8}{@{}l}{\\textit{Table \\thetable\\ (continued)}}\\\\\n\\toprule\n")
        f.write(head + "\\midrule\\endhead\n\\bottomrule\\endfoot\n")
        for enc in encs:
            g = df[df.encoder == enc]
            for i, (_, r) in enumerate(g.iterrows()):
                name = SHORT.get(enc, enc) + f" ($k={int(r.k)}$)" if i == 0 else ""
                bold = r.lambda_score == 1.0
                fmt = (lambda s: "\\textbf{" + s + "}") if bold else (lambda s: s)
                f.write(f"{name} & {fmt(f'{r.lambda_score:g}')} & {fmt(f'{r.ortho_max_offdiag:.4f}')} & "
                        f"{fmt(f'{r.core_size_median:.1f}')} & {fmt(f'{r.core_energy_median:.3f}')} & "
                        f"{fmt(f'{r.explained_var_ratio_to_pca:.4f}')} & "
                        f"{fmt(f'{r.faith_rate:.3f}')} & {fmt(f'{r.faith_chance:.3f}')} \\\\\n")
            f.write("\\midrule\n" if enc != encs[-1] else "")
        f.write("\\end{longtable}}\n")
    print(f"wrote {OUT} | {len(encs)} encoders x {len(lams)} weights")


if __name__ == "__main__":
    main()
```
