## Supplemental code for "Scoring respondents on a semantic frame of reference identifies a group that is burdened across many questionnaires and reaches care less easily": table_phrasing_xenc.py

```
#!/usr/bin/env python
"""Supplementary table: placement under a difference in syntax, under all six encoders.

Both batteries ask questions the pool also asks. Matching an item of a battery to its pool version by
text alone splits the matched pairs into two groups, the ones the two sources word identically and the
ones whose syntax differs (a stem the pool strips, punctuation, or a bare fragment against a full
interrogative). For each pair we compare the set of core components the two versions reach on the same
fixed basis by the Jaccard index, against a baseline that pairs each battery item with a random pool
item. This is the procedure of Methods "Robustness to syntax", run once per encoder rather than on the
exemplar alone.

The gte-large column reproduces the numbers quoted in the main text, and the matching rules are the
ones in hrs_kept_components.py: All of Us prompts shorter than 20 normalized characters are skipped,
because a short tail matches too many pool items, and HRS text is compared after case and punctuation
are removed, because HRS administers bare fragments.

Reads only saved artefacts - no numbers or item ids are hardcoded:
    outputs_pool/fits/<enc>__k*_ls1.0/{loadings,mean,scores}.npy   the pool basis and projection
    data/core151_items_<enc>.npz                                   the All of Us battery embeddings
    ../randhrsdata/acidd_input/hrs_lb2022_<enc>.npz                the HRS battery embeddings
    ../global_pool/pool_processed.csv, core_items_151.csv,
    ../randhrsdata/acidd_input/hrs_lb2022_items.csv                the published question text

Writes manuscript_nhb/phrasing_xenc_table.tex and prints every number Supplementary Section S19 quotes.

Run: CUDA_VISIBLE_DEVICES="" /data_gpu5/ROSS_Proj/env/bin/python table_phrasing_xenc.py
"""
import os, glob, re
import numpy as np
import pandas as pd

from hrs_kept_components import fpc, ENCS, HERE, GP, HRSD

OUT = os.path.join(HERE, "..", "..", "manuscript_nhb", "phrasing_xenc_table.tex")
ENC_LABEL = {"gte-large-en-v1.5": "gte-large", "bge-m3": "bge-m3",
             "bge-large-en-v1.5": "bge-large", "all-mpnet-base-v2": "all-mpnet",
             "e5-large-v2": "e5-large", "qwen3-embedding-0.6b": "qwen3"}
NPERM = {"aou": 100, "hrs": 200}          # as in hrs_kept_components.phrasing_aou / phrasing


def tail(t):
    return re.sub(r"[^a-z]", "", str(t).lower())[-45:]


def norm(t):
    return re.sub(r"[^a-z ]", "", str(t).lower()).strip()


def jac(a, b):
    return len(a & b) / len(a | b) if (a | b) else 0.0


def battery_text(battery):
    if battery == "aou":
        ai = pd.read_csv(os.path.join(HERE, "core_items_151.csv"))
        return dict(zip(ai.item, ai.prompt))
    hrs = pd.read_csv(os.path.join(HRSD, "hrs_lb2022_items.csv"))
    return dict(zip(hrs.item, hrs.prompt))


def battery_vectors(battery, enc):
    if battery == "aou":
        d = np.load(os.path.join(HERE, "data", f"core151_items_{enc}.npz"), allow_pickle=True)
    else:
        d = np.load(os.path.join(HRSD, f"hrs_lb2022_{enc}.npz"), allow_pickle=True)
    return [str(x) for x in d["items"]], np.asarray(d["vectors"], float)


def run(battery, enc, pp, ids, btxt):
    fit = glob.glob(os.path.join(HERE, "outputs_pool", "fits", f"{enc}__k*_ls1.0"))[0]
    W = np.load(os.path.join(fit, "loadings.npy"))
    mu = np.load(os.path.join(fit, "mean.npy"))
    P = np.load(os.path.join(fit, "scores.npy"))
    sigma = P.std(0)                                  # the pool's column scale, D2
    Zp = P / sigma
    bid, V = battery_vectors(battery, enc)
    Zb = ((V - mu) @ W.T) / sigma
    pool_by_tail = {tail(t): (i, t) for i, t in zip(ids, pp.prompt_clean)}
    same, diff = [], []
    for i, it in enumerate(bid):
        k = tail(btxt.get(it, ""))
        if battery == "aou" and len(k) < 20:
            continue
        m = next((pool_by_tail[q] for q in pool_by_tail if q.endswith(k) or k.endswith(q)), None)
        if m is None:
            continue
        A = {int(c) for c in fpc(Zb[i])}
        B = {int(c) for c in fpc(Zp[ids.index(m[0])])}
        identical = (str(btxt[it]).strip() == str(m[1]).strip() if battery == "aou"
                     else norm(btxt[it]) == norm(m[1]))
        (same if identical else diff).append((jac(A, B), len(A & B) > 0, A == B))
    rng = np.random.RandomState(0)
    null = float(np.mean([jac({int(c) for c in fpc(Zb[i])},
                              {int(c) for c in fpc(Zp[rng.randint(len(ids))])})
                          for i in range(len(bid)) for _ in range(NPERM[battery])]))
    return dict(n_same=len(same), same=float(np.mean([x[0] for x in same])),
                n_diff=len(diff), diff=float(np.mean([x[0] for x in diff])),
                share=sum(x[1] for x in diff), exact=sum(x[2] for x in diff), null=null)


def main():
    pp = pd.read_csv(os.path.join(GP, "pool_processed.csv"))
    ids = list(pp.item_id)
    rows = {}
    for battery, name in (("aou", "All of Us"), ("hrs", "HRS")):
        btxt = battery_text(battery)
        rows[battery] = [(enc, run(battery, enc, pp, ids, btxt)) for enc in ENCS]
        r0 = rows[battery][0][1]
        print(f"\n{name}: {r0['n_same']} identical, {r0['n_diff']} differing in syntax")
        print(f"{'encoder':12s} {'identical':>9s} {'differing':>9s} {'share>=1':>9s} {'same set':>9s} {'random':>7s}")
        for enc, r in rows[battery]:
            print(f"{ENC_LABEL[enc]:12s} {r['same']:9.2f} {r['diff']:9.2f} "
                  f"{r['share']:5d}/{r['n_diff']:<3d} {r['exact']:5d}/{r['n_diff']:<3d} {r['null']:7.3f}")
        d = [r['diff'] for _, r in rows[battery]]
        print(f"  differing-syntax overlap runs {min(d):.2f} to {max(d):.2f} across the six")

    with open(OUT, "w") as f:
        f.write("% AUTO-GENERATED by examples/allofus/table_phrasing_xenc.py -- do not edit\n")
        f.write(r"\begin{table}[H]" "\n" r"\centering" "\n")
        f.write(r"\caption{\textbf{Placement under a difference in syntax, under each encoder.} "
                r"For each battery, the questions the reference pool also asks are split by whether "
                r"the two sources word them identically or differ in syntax, and each pair is scored "
                r"by the overlap of the core components the two versions reach on the same fixed basis "
                r"(1 is full overlap and 0 is none). \emph{shares one} counts the differing pairs "
                r"reaching at least one core component in common and \emph{same set} those reaching "
                r"exactly the same components. \emph{random} pairs each battery item with a random "
                r"pool item. Identical text must place identically, so that column is a check on the "
                r"procedure rather than a result "
                r"(Methods~\S\ref{sec:methods_variants}).}\label{tab:phrasing_xenc}" "\n")
        f.write(r"\small" "\n" r"\begin{tabular}{@{}l c c c c c@{}}" "\n" r"\toprule" "\n")
        f.write("Encoder & identical & differing & shares one & same set & random \\\\\n")
        for battery, name in (("aou", "All of Us"), ("hrs", "HRS")):
            n_same = rows[battery][0][1]["n_same"]
            n_diff = rows[battery][0][1]["n_diff"]
            f.write(r"\midrule" "\n")
            f.write(f"\\multicolumn{{6}}{{@{{}}l}}{{\\emph{{{name}}}, {n_same} identical and "
                    f"{n_diff} differing in syntax}} \\\\[1pt]\n")
            for enc, r in rows[battery]:
                f.write(f"{ENC_LABEL[enc]} & {r['same']:.2f} & {r['diff']:.2f} & "
                        f"{r['share']}/{r['n_diff']} & {r['exact']}/{r['n_diff']} & {r['null']:.3f} \\\\\n")
        f.write(r"\bottomrule" "\n" r"\end{tabular}" "\n" r"\end{table}" "\n")
    print("\nwrote", os.path.normpath(OUT))


if __name__ == "__main__":
    main()
```
