## Supplemental code for "Scoring respondents on a semantic frame of reference identifies a group that is burdened across many questionnaires and reaches care less easily": build_deck.py

```
#!/usr/bin/env python
import sys
sys.path.insert(0, "/data_gpu5/ROSS_Proj/.pylibs")
from pptx import Presentation
from pptx.util import Inches, Pt, Emu
from pptx.dml.color import RGBColor
from pptx.enum.text import PP_ALIGN, MSO_ANCHOR
from pptx.enum.shapes import MSO_SHAPE
from pptx.chart.data import CategoryChartData
from pptx.enum.chart import XL_CHART_TYPE, XL_LEGEND_POSITION, XL_LABEL_POSITION

# palette
INK="16233B"; INK2="21395E"; TEAL="1C7293"; SEA="2F9C8B"; AMBER="E7A33E"
CORAL="D9695E"; CLOUD="F3F6FA"; WHITE="FFFFFF"; TINT="E7EFF5"; SLATE="56657E"
BODY="1E2C46"; LINE="D3DEE8"; MUTE1="C9D6E6"; MUTE2="9FB2C7"
HF="Cambria"; BF="Calibri"

from PIL import Image
def C(h): return RGBColor.from_string(h)

prs = Presentation()
prs.slide_width = Inches(13.33); prs.slide_height = Inches(7.5)
BLANK = prs.slide_layouts[6]
W=13.33; MX=0.6
_PAGE=[0]
FIGDIR="/data_gpu5/ROSS_Proj/examples/global_pool/figures"

def slide(bg):
    _PAGE[0]+=1
    s = prs.slides.add_slide(BLANK)
    r = s.shapes.add_shape(MSO_SHAPE.RECTANGLE, Inches(0),Inches(0),Inches(W),Inches(7.5))
    r.fill.solid(); r.fill.fore_color.rgb=C(bg); r.line.fill.background(); r.shadow.inherit=False
    r.name="bg"
    return s

def rrect(s,x,y,w,h,fill,line=None,lw=1.0,rad=0.09):
    sh=s.shapes.add_shape(MSO_SHAPE.ROUNDED_RECTANGLE,Inches(x),Inches(y),Inches(w),Inches(h))
    try: sh.adjustments[0]=rad
    except Exception: pass
    sh.fill.solid(); sh.fill.fore_color.rgb=C(fill)
    if line: sh.line.color.rgb=C(line); sh.line.width=Pt(lw)
    else: sh.line.fill.background()
    sh.shadow.inherit=False
    return sh

def rect(s,x,y,w,h,fill):
    sh=s.shapes.add_shape(MSO_SHAPE.RECTANGLE,Inches(x),Inches(y),Inches(w),Inches(h))
    sh.fill.solid(); sh.fill.fore_color.rgb=C(fill); sh.line.fill.background(); sh.shadow.inherit=False
    return sh

def oval(s,x,y,w,h,fill,line=None,lw=1.0):
    sh=s.shapes.add_shape(MSO_SHAPE.OVAL,Inches(x),Inches(y),Inches(w),Inches(h))
    sh.fill.solid(); sh.fill.fore_color.rgb=C(fill)
    if line: sh.line.color.rgb=C(line); sh.line.width=Pt(lw)
    else: sh.line.fill.background()
    sh.shadow.inherit=False
    return sh

def hline(s,x,y,w,color,wt=1.5):
    ln=s.shapes.add_connector(2,Inches(x),Inches(y),Inches(x+w),Inches(y))
    ln.line.color.rgb=C(color); ln.line.width=Pt(wt); ln.shadow.inherit=False
    return ln

def txt(s,x,y,w,h,runs,align=PP_ALIGN.LEFT,anchor=MSO_ANCHOR.TOP,wrap=True,
        space=1.0,m0=True):
    tb=s.shapes.add_textbox(Inches(x),Inches(y),Inches(w),Inches(h)); tf=tb.text_frame
    tf.word_wrap=wrap; tf.vertical_anchor=anchor
    if m0:
        tf.margin_left=0; tf.margin_right=0; tf.margin_top=0; tf.margin_bottom=0
    if isinstance(runs,str): runs=[(runs,{})]
    para=tf.paragraphs[0]; para.alignment=align
    try: para.line_spacing=space
    except Exception: pass
    for t,o in runs:
        r=para.add_run(); r.text=t; f=r.font
        f.name=o.get("font",BF); f.size=Pt(o.get("size",14))
        f.bold=o.get("bold",False); f.italic=o.get("italic",False)
        f.color.rgb=C(o.get("color",BODY))
    return tb

def notes(s,t): s.notes_slide.notes_text_frame.text=t

def head(s,kick,title,tcolor=INK):
    txt(s,MX,0.5,12,0.32,[(kick.upper(),{"color":TEAL,"font":BF,"size":12,"bold":True})])
    txt(s,MX,0.82,12.1,0.9,[(title,{"color":tcolor,"font":HF,"size":34,"bold":True})],anchor=MSO_ANCHOR.TOP)

def foot(s,n,tag):
    txt(s,MX,7.06,9,0.3,[("Global item pool   ·   "+tag,{"color":SLATE,"font":BF,"size":9})],anchor=MSO_ANCHOR.MIDDLE)
    txt(s,12.4,7.06,0.45,0.3,[(str(_PAGE[0]),{"color":SLATE,"font":BF,"size":9})],align=PP_ALIGN.RIGHT,anchor=MSO_ANCHOR.MIDDLE)

def image_slide(kick,title,img,tag,caption=None):
    s=slide(CLOUD); head(s,kick,title)
    wpx,hpx=Image.open(img).size; aspect=wpx/hpx
    x0,top,boxw,boxh=0.5,1.74,12.33,4.95
    w=boxw; h=w/aspect
    if h>boxh: h=boxh; w=h*aspect
    x=x0+(boxw-w)/2; y=top+(boxh-h)/2
    s.shapes.add_picture(img,Inches(x),Inches(y),width=Inches(w))
    if caption:
        txt(s,MX,6.5,12.1,0.4,[(caption,{"color":SLATE,"font":BF,"size":11,"italic":True})],align=PP_ALIGN.CENTER)
    foot(s,0,tag); return s

def style_chart(chart,colors,pct=True,dsize=10,legend=False):
    chart.has_title=False
    if legend:
        chart.has_legend=True; chart.legend.position=XL_LEGEND_POSITION.TOP
        chart.legend.include_in_layout=False; chart.legend.font.size=Pt(11)
        chart.legend.font.name=BF; chart.legend.font.color.rgb=C(SLATE)
    else:
        chart.has_legend=False
    plot=chart.plots[0]; plot.gap_width=60
    for i,ser in enumerate(plot.series):
        ser.format.fill.solid(); ser.format.fill.fore_color.rgb=C(colors[i%len(colors)])
        ser.format.line.fill.background()
    plot.has_data_labels=True
    dl=plot.data_labels
    dl.font.size=Pt(dsize); dl.font.name=BF; dl.font.color.rgb=C(BODY)
    dl.position=XL_LABEL_POSITION.OUTSIDE_END
    if pct: dl.number_format='0"%"'; dl.number_format_is_linked=False
    va=chart.value_axis; va.visible=False; va.has_major_gridlines=False
    va.minimum_scale=0; va.maximum_scale=100
    ca=chart.category_axis; ca.has_major_gridlines=False
    ca.tick_labels.font.size=Pt(11); ca.tick_labels.font.name=BF
    ca.tick_labels.font.color.rgb=C(BODY)
    try: ca.format.line.color.rgb=C(LINE)
    except Exception: pass

# ============================ 1 TITLE
s=slide(INK)
for (x,y,c) in [(10.4,1.0,SEA),(11.0,0.7,TEAL),(11.6,1.15,AMBER),(10.9,1.55,TEAL),
                (11.7,1.75,SEA),(12.3,1.25,CORAL),(12.1,0.65,TEAL),(10.5,1.9,AMBER)]:
    oval(s,x,y,0.34,0.34,c,line=INK,lw=1)
txt(s,MX,2.35,11,0.4,[("MEASUREMENT PORTABILITY",{"color":AMBER,"font":BF,"size":15,"bold":True})])
txt(s,MX,2.8,11.4,1.9,[("A global item pool for text-derived\npsychosocial measurement",
    {"color":WHITE,"font":HF,"size":42,"bold":True})],space=1.02)
txt(s,MX,4.8,10.6,0.9,[("Do meaning-based components learned from an external item bank recover the structure of All of Us — and transfer across instruments?",
    {"color":MUTE1,"font":BF,"size":16})],space=1.05)
meta=[("577","pool items"),("50","instruments"),("53","constructs"),("6","text encoders")]
for i,(n,l) in enumerate(meta):
    x=MX+i*2.5
    txt(s,x,5.9,1.1,0.55,[(n,{"color":SEA,"font":HF,"size":30,"bold":True})])
    txt(s,x+1.15,6.02,1.3,0.5,[(l,{"color":MUTE2,"font":BF,"size":11})],anchor=MSO_ANCHOR.MIDDLE)
notes(s,"A cross-instrument pool of 577 validated psychosocial items assembled from published item text alone. Core question: does a meaning basis learned from this external bank recover All of Us structure and transfer across instruments.")

# ============================ 2 BUILDING THE POOL
s=slide(CLOUD)
head(s,"Step 1 · Assembly","Building a global item pool")
txt(s,MX,1.62,11.8,0.5,[("Validated psychosocial questionnaire items, assembled from their published item text alone — no respondents required.",
    {"color":SLATE,"font":BF,"size":14})])
stats=[("577","items across the pool"),("50","validated instruments"),("53","target constructs"),("6","embedding encoders")]
sw=2.75; gap=(12.13-4*sw)/3
for i,(n,l) in enumerate(stats):
    x=MX+i*(sw+gap); rrect(s,x,2.25,sw,1.25,WHITE,line=LINE)
    txt(s,x,2.4,sw,0.7,[(n,{"color":TEAL,"font":HF,"size":38,"bold":True})],align=PP_ALIGN.CENTER)
    txt(s,x+0.15,3.06,sw-0.3,0.4,[(l,{"color":SLATE,"font":BF,"size":11.5})],align=PP_ALIGN.CENTER)
cy=3.8; ch=2.15
rrect(s,MX,cy,5.86,ch,WHITE,line=LINE)
txt(s,MX+0.3,cy+0.25,5.2,0.4,[("36 independent instruments",{"bold":True,"color":INK,"size":17,"font":HF}),
    ("   476 items",{"color":SEA,"size":13,"font":BF,"bold":True})])
txt(s,MX+0.3,cy+0.75,5.3,1.2,[("A different instrument for the construct than All of Us used — the real transfer test (CES-D, PROMIS, IPIP-50, De Jong Gierveld, USDA).",
    {"color":BODY,"font":BF,"size":13})],space=1.05)
x2=MX+6.27
rrect(s,x2,cy,5.86,ch,WHITE,line=LINE)
txt(s,x2+0.3,cy+0.25,5.2,0.4,[("14 shared instruments",{"bold":True,"color":INK,"size":17,"font":HF}),
    ("   101 items",{"color":AMBER,"size":13,"font":BF,"bold":True})])
txt(s,x2+0.3,cy+0.75,5.3,1.2,[("The same instrument All of Us administered — a sanity check. All 17 AoU validated instruments are represented in the pool.",
    {"color":BODY,"font":BF,"size":13})],space=1.05)
txt(s,MX,6.28,12,0.4,[("Time-frame stems stripped before embedding — 162 of 577 items — so items cluster by content, not recall window.",
    {"color":SLATE,"font":BF,"size":12,"italic":True})])
foot(s,2,"Assembly")
notes(s,"577 items, 50 instruments, 53 constructs, 6 encoders. 36 independent instruments (476 items) are the real transfer test; 14 shared instruments (101 items) reuse AoU's own instruments. Stems stripped so clustering is by content.")

# ============================ 3 METHOD
s=slide(CLOUD)
head(s,"Step 2 · Method","From item text to a shared meaning basis")
txt(s,MX,1.62,11.9,0.5,[("A ROSS-Proj sparse-component basis is learned from the pool embeddings — then All of Us items are projected onto it. All CPU.",
    {"color":SLATE,"font":BF,"size":14})])
steps=[("1","Embed","6 text encoders on stem-stripped item text"),
       ("2","Select k","Horn's parallel analysis — k = 49–55"),
       ("3","Fit basis","ROSS-Proj components W (λ = 1.0)"),
       ("4","Cores","Iterated fixed-point PR core per component"),
       ("5","Project AoU","Place the 151 AoU items onto pool W")]
cw=2.14; cg=(12.13-5*cw)/4; cyy=2.7; chh=2.4
for i,(n,t,d) in enumerate(steps):
    x=MX+i*(cw+cg); dark=(i==4)
    rrect(s,x,cyy,cw,chh,INK if dark else WHITE,line=None if dark else LINE)
    oval(s,x+cw/2-0.32,cyy+0.28,0.64,0.64,AMBER if dark else TEAL)
    txt(s,x+cw/2-0.32,cyy+0.28,0.64,0.64,[(n,{"color":INK if dark else WHITE,"font":HF,"size":24,"bold":True})],align=PP_ALIGN.CENTER,anchor=MSO_ANCHOR.MIDDLE)
    txt(s,x+0.1,cyy+1.05,cw-0.2,0.4,[(t,{"color":WHITE if dark else INK,"font":HF,"size":16,"bold":True})],align=PP_ALIGN.CENTER)
    txt(s,x+0.14,cyy+1.48,cw-0.28,0.85,[(d,{"color":MUTE1 if dark else SLATE,"font":BF,"size":11})],align=PP_ALIGN.CENTER,space=1.02)
    if i<4:
        txt(s,x+cw+cg/2-0.2,cyy+0.55,0.4,0.9,[("›",{"color":TEAL,"font":BF,"size":30,"bold":True})],align=PP_ALIGN.CENTER,anchor=MSO_ANCHOR.MIDDLE)
txt(s,MX,5.75,12,0.6,[("Method note.  ",{"bold":True,"color":INK,"font":BF,"size":12}),
    ("Cores use the iterated fixed-point participation ratio (not single-pass defining sets). W is never pruned — projection is on the dense basis.",{"color":SLATE,"font":BF,"size":12})])
foot(s,3,"Method")
notes(s,"Pipeline: embed with 6 encoders, select k via Horn's parallel analysis (49-55), fit ROSS-Proj basis at lambda=1.0, take iterated fixed-point PR cores, then project AoU's 151 items onto the pool basis. All CPU.")

# ============================ 4 POOL CORES
s=slide(CLOUD)
head(s,"Result · Pool cores","The pool organizes itself by construct")
txt(s,MX,1.62,11.9,0.5,[("Fixed-point cores are construct-coherent and stable across encoders — and independent instruments of the same construct co-locate.",
    {"color":SLATE,"font":BF,"size":14})])
labels=["MPNet","BGE-lg","BGE-m3","E5","GTE","Qwen3"]
cd=CategoryChartData(); cd.categories=labels
cd.add_series("Coverage",(62,70,67,69,71,64))
cd.add_series("Median purity",(67,68,64,56,67,67))
gf=s.shapes.add_chart(XL_CHART_TYPE.COLUMN_CLUSTERED,Inches(MX),Inches(2.35),Inches(6.5),Inches(3.9),cd)
style_chart(gf.chart,[TEAL,AMBER],pct=True,dsize=9,legend=True)
txt(s,MX,6.18,6.5,0.3,[("% by encoder — cores vs the 53 fine-grained constructs",{"color":SLATE,"font":BF,"size":10,"italic":True})],align=PP_ALIGN.CENTER)
rx=7.5
rrect(s,rx,2.35,5.23,1.05,INK)
txt(s,rx+0.3,2.35,4.7,1.05,[("k = 49–55",{"color":WHITE,"font":HF,"size":20,"bold":True}),
    ("  components   ·   ",{"color":MUTE2,"font":BF,"size":12}),
    ("median core ≈ 7",{"color":AMBER,"font":HF,"size":20,"bold":True}),
    ("  items",{"color":MUTE2,"font":BF,"size":12})],anchor=MSO_ANCHOR.MIDDLE)
txt(s,rx,3.28,5.2,0.3,[("Independent instruments that share a core:",{"color":INK,"font":BF,"size":12,"bold":True})])
ex=[("Depression","CES-D + PROMIS-Depression"),
    ("Anxiety","PROMIS-Anxiety + OASIS"),
    ("Sleep","PROMIS-Sleep + Insomnia Severity Index"),
    ("Discrimination","Everyday + Medical-Settings instruments")]
ey=3.6; eh=0.62; eg=0.06
for i,(c,d) in enumerate(ex):
    y=ey+i*(eh+eg); rrect(s,rx,y,5.23,eh,WHITE,line=LINE)
    oval(s,rx+0.18,y+eh/2-0.075,0.15,0.15,SEA)
    txt(s,rx+0.45,y,1.75,eh,[(c,{"color":INK,"font":HF,"size":13,"bold":True})],anchor=MSO_ANCHOR.MIDDLE)
    txt(s,rx+2.2,y,2.95,eh,[(d,{"color":BODY,"font":BF,"size":11.5})],anchor=MSO_ANCHOR.MIDDLE)
foot(s,4,"Pool cores")
notes(s,"Standalone pool cores: k 49-55, median core ~7 items, coverage 0.62-0.71, purity 0.56-0.68 vs 53 fine constructs. Independent instruments of the same construct co-locate on one core.")

image_slide("Result · Pool cores","Different instruments merge into shared components",
            FIGDIR+"/fig_pool_ribbon.png","Pool merge",
            "Instrument → construct → pool component by fixed-point core membership (gte-large). Blue = cross-instrument merge onto a shared component.")
image_slide("Result · Pool cores","What each pool component is",
            FIGDIR+"/fig_pool_cards_main.png","Pool components",
            "One card per component: its core's constructs and items-in-core / items-in-pool. The 26 clearest of 55 (full set in the appendix).")

# ============================ 5 AoU TRANSFER
s=slide(CLOUD)
head(s,"Result · Transferability","All of Us items are recovered by the external basis")
txt(s,MX,1.62,12,0.5,[("Project the 151 AoU items onto the pool basis, then core each component. AoU items fall into construct-coherent cores.",
    {"color":SLATE,"font":BF,"size":14})])
rrect(s,MX,2.4,4.5,3.85,INK)
txt(s,MX+0.1,2.72,4.3,1.1,[("88–94%",{"color":AMBER,"font":HF,"size":60,"bold":True})],align=PP_ALIGN.CENTER)
txt(s,MX+0.2,3.85,4.1,0.4,[("of AoU's 151 items covered",{"color":WHITE,"font":BF,"size":15,"bold":True})],align=PP_ALIGN.CENTER)
hline(s,MX+0.7,4.55,3.1,INK2,1.5)
txt(s,MX+0.2,4.72,4.1,0.7,[("5–9",{"color":SEA,"font":HF,"size":34,"bold":True}),
    ("  pure axes / encoder",{"color":MUTE1,"font":BF,"size":14})],align=PP_ALIGN.CENTER,anchor=MSO_ANCHOR.MIDDLE)
txt(s,MX+0.2,5.62,4.1,0.4,[("Consistent across all six encoders",{"color":MUTE2,"font":BF,"size":12,"italic":True})],align=PP_ALIGN.CENTER)
txt(s,5.55,2.4,7.2,0.35,[("AoU coverage by encoder",{"color":INK,"font":BF,"size":13,"bold":True})])
cd=CategoryChartData(); cd.categories=labels; cd.add_series("Coverage",(90,92,88,89,92,94))
gf=s.shapes.add_chart(XL_CHART_TYPE.COLUMN_CLUSTERED,Inches(5.5),Inches(2.8),Inches(7.25),Inches(3.45),cd)
style_chart(gf.chart,[TEAL],pct=True,dsize=12,legend=False)
foot(s,5,"Transferability")
notes(s,"Projecting AoU onto pool W and coring each component: 88-94% of AoU's 151 items covered, 5-9 pure axes per encoder, consistent across all six encoders.")

image_slide("Result · Transferability","AoU items transfer onto the external pool basis",
            FIGDIR+"/fig_aou_on_pool_ribbon.png","AoU transfer",
            "Red = AoU instrument NOT in the pool (genuine external transfer: ACE-BRFSS, MOS-SS, BFI-2-XS). Grey = instrument shared with the pool (sanity check). Right label = the pool instrument defining each component.")
image_slide("Result · Transferability","What AoU lands on each pool component",
            FIGDIR+"/fig_aou_on_pool_cards_main.png","AoU components",
            "Same pool component IDs as the pool cards, now showing which AoU constructs landed there.")
image_slide("Result · Other cohorts","HRS items form the cores of the pool components they cover",
            FIGDIR+"/fig_hrs_on_pool_ribbon.png","HRS transfer",
            "HRS 2022 Leave-Behind social battery (35 items) projected onto the pool. Discrimination, loneliness and neighborhood transfer cleanly; PSS is diffuse and scatters. Red = instrument not in the pool. Components HRS doesn't measure stay empty (expected coverage gap).")
image_slide("Result · Other cohorts","A clinical cohort (Xinxiang) on the pool",
            FIGDIR+"/fig_xinxiang_on_pool_ribbon.png","Xinxiang transfer",
            "Xinxiang (PHQ-9, GAD-7, PSS-14, ISI; 37 items embedded independently) projected onto the pool. ISI→sleep, GAD→anxiety, PHQ→depression/somatic land cleanly; PSS is diffuse. All four are pool instruments — a coverage / wording-robustness check.")

# ============================ 6 STEM ROBUSTNESS
s=slide(CLOUD)
head(s,"Robustness · Item stems","Landing survives a stem mismatch")
txt(s,MX,1.62,12.1,0.7,[("AoU items carry their time-frame stems. The pool basis was learned on stem-stripped text. They still land on the right component — the asymmetry is the robustness result.",
    {"color":SLATE,"font":BF,"size":14})],space=1.03)
by=2.9
rrect(s,MX,by,3.75,2.5,WHITE,line=LINE)
txt(s,MX+0.25,by+0.22,3.3,0.35,[("AoU item — with stem",{"color":AMBER,"font":BF,"size":12,"bold":True})])
txt(s,MX+0.25,by+0.72,3.3,1.1,[("“In the past 7 days, ",{"color":CORAL,"italic":True,"font":BF,"size":14,"bold":True}),
    ("I felt worthless.”",{"color":BODY,"italic":True,"font":BF,"size":14})],space=1.05)
txt(s,MX+0.25,by+1.98,3.3,0.3,[("time-frame stem in red",{"color":SLATE,"font":BF,"size":10,"italic":True})])
txt(s,MX+3.85,by+0.8,0.5,0.9,[("›",{"color":TEAL,"font":BF,"size":34,"bold":True})],align=PP_ALIGN.CENTER,anchor=MSO_ANCHOR.MIDDLE)
x2=MX+4.45
rrect(s,x2,by,3.75,2.5,INK)
txt(s,x2+0.25,by+0.22,3.3,0.35,[("Pool basis — stem-stripped",{"color":SEA,"font":BF,"size":12,"bold":True})])
txt(s,x2+0.25,by+0.72,3.3,1.4,[("Learned on content only. 162 of 577 pool items had a leading time phrase removed.",
    {"color":MUTE1,"font":BF,"size":13.5})],space=1.12)
txt(s,x2+3.85,by+0.8,0.5,0.9,[("›",{"color":TEAL,"font":BF,"size":34,"bold":True})],align=PP_ALIGN.CENTER,anchor=MSO_ANCHOR.MIDDLE)
x3=x2+4.45
rrect(s,x3,by,3.53,2.5,WHITE,line=LINE)
txt(s,x3+0.25,by+0.22,3.1,0.35,[("Lands correctly",{"color":TEAL,"font":BF,"size":12,"bold":True})])
oval(s,x3+1.42,by+0.72,0.7,0.7,SEA)
txt(s,x3+1.42,by+0.72,0.7,0.7,[("✓",{"color":WHITE,"font":BF,"size":26,"bold":True})],align=PP_ALIGN.CENTER,anchor=MSO_ANCHOR.MIDDLE)
txt(s,x3+0.2,by+1.58,3.1,0.9,[("on the depression core — no stem-stripped re-embed of AoU required.",
    {"color":BODY,"font":BF,"size":12.5})],align=PP_ALIGN.CENTER,space=1.08)
txt(s,MX,5.75,12.1,0.7,[("Why it matters.  ",{"bold":True,"color":INK,"font":BF,"size":13}),
    ("Re-embedding AoU stem-stripped would erase the very contrast that demonstrates content-based transfer. The with-stem → stripped-basis result already carries it.",
    {"color":SLATE,"font":BF,"size":13})])
foot(s,6,"Stem robustness")
notes(s,"AoU items keep their stems; the pool basis is stem-stripped; they still land on the correct core. That asymmetry is itself the cross-stem robustness result, so no stem-stripped AoU re-embed is needed.")

# ============================ 7 CROSS-INSTRUMENT
s=slide(CLOUD)
head(s,"Key result","Cross-instrument co-location")
txt(s,MX,1.6,9.0,0.7,[("AoU items land in cores defined by a different, independent instrument of the same content — replicating in every encoder.",
    {"color":SLATE,"font":BF,"size":14})],space=1.03)
rrect(s,9.75,1.55,2.98,1.5,INK)
txt(s,9.95,1.68,2.65,0.55,[("13–19",{"color":AMBER,"font":HF,"size":24,"bold":True}),
    ("  cross-instrument axes",{"color":MUTE1,"font":BF,"size":11})],anchor=MSO_ANCHOR.MIDDLE)
txt(s,9.95,2.35,2.65,0.55,[("76–101",{"color":SEA,"font":HF,"size":24,"bold":True}),
    ("  of 151 AoU items",{"color":MUTE1,"font":BF,"size":11})],anchor=MSO_ANCHOR.MIDDLE)
rows=[("Childhood adversity","ACE-BRFSS","Felitti ACE"),
      ("Psychosis","CIDI-5 / MHQ","PQ-B"),
      ("Loneliness / isolation","UCLA-V3","PROMIS Social Isolation"),
      ("Social support","MOS-SS","ISEL-12 / MSPSS"),
      ("Food insecurity","Hunger Vital Sign","USDA HFSSM"),
      ("Trauma / IPV","ACE / MHQ","LEC-5 / E-HITS"),
      ("Suicidality","PHQ-9 (item 9)","SBQ-R")]
ty=3.25; rh=0.44; rg=0.055
txt(s,MX+0.15,ty-0.32,2.6,0.3,[("CONSTRUCT",{"color":SLATE,"font":BF,"size":10,"bold":True})])
txt(s,MX+3.05,ty-0.32,2.9,0.3,[("AoU INSTRUMENT",{"color":SLATE,"font":BF,"size":10,"bold":True})])
txt(s,MX+6.6,ty-0.32,4.0,0.3,[("POOL INSTRUMENT (INDEPENDENT)",{"color":SLATE,"font":BF,"size":10,"bold":True})])
txt(s,MX+10.75,ty-0.32,1.3,0.3,[("ENCODERS",{"color":SLATE,"font":BF,"size":10,"bold":True})],align=PP_ALIGN.CENTER)
for i,(c,a,po) in enumerate(rows):
    y=ty+i*(rh+rg)
    rrect(s,MX,y,12.13,rh,WHITE if i%2 else TINT,line=LINE,lw=0.75,rad=0.06)
    txt(s,MX+0.2,y,2.75,rh,[(c,{"color":INK,"font":HF,"size":13,"bold":True})],anchor=MSO_ANCHOR.MIDDLE)
    txt(s,MX+3.05,y,2.9,rh,[(a,{"color":BODY,"font":BF,"size":12.5})],anchor=MSO_ANCHOR.MIDDLE)
    txt(s,MX+6.05,y,0.4,rh,[("›",{"color":TEAL,"font":BF,"size":18,"bold":True})],align=PP_ALIGN.CENTER,anchor=MSO_ANCHOR.MIDDLE)
    txt(s,MX+6.6,y,4.0,rh,[(po,{"color":TEAL,"font":BF,"size":12.5,"bold":True})],anchor=MSO_ANCHOR.MIDDLE)
    rrect(s,MX+10.9,y+0.07,0.95,rh-0.14,SEA,rad=0.14)
    txt(s,MX+10.9,y+0.07,0.95,rh-0.14,[("6 / 6",{"color":WHITE,"font":BF,"size":11,"bold":True})],align=PP_ALIGN.CENTER,anchor=MSO_ANCHOR.MIDDLE)
txt(s,MX,6.78,12.13,0.35,[("Also in 4–5 / 6 encoders:  ",{"bold":True,"color":INK,"font":BF,"size":11.5}),
    ("anxiety (GAD-7 → PROMIS-Anxiety) · personality (BFI-2-XS → IPIP-50) · depression (PHQ-9 → PROMIS-Depression) · quality of life (→ WHOQOL-BREF) · mania (→ MDQ)",
    {"color":SLATE,"font":BF,"size":11.5})],anchor=MSO_ANCHOR.MIDDLE)
foot(s,7,"Cross-instrument")
notes(s,"Core-to-core join: AoU items land in cores defined by a different independent instrument of the same content. Seven construct families replicate in all six encoders; five more in 4-5/6. Per encoder 13-19 cross-instrument axes, 76-101 of 151 AoU items.")

# ============================ 8 TAKEAWAYS
s=slide(INK)
txt(s,MX,0.65,11,0.4,[("WHAT THIS SHOWS",{"color":AMBER,"font":BF,"size":14,"bold":True})])
txt(s,MX,1.05,11.8,1.0,[("Measurement meaning transfers across instruments",{"color":WHITE,"font":HF,"size":33,"bold":True})])
items=[("Portability","A basis learned from 577 external items — with no All of Us input — recovers 88–94% of AoU's 151 items into construct-coherent cores."),
       ("Cross-instrument transfer","AoU items co-locate with different-instrument pool cores of the same content. Seven construct families replicate in all six encoders."),
       ("Robust by design","Landing survives the stem mismatch and holds across six encoders. Multi-content instruments correctly scatter by content, not by label."),
       ("Honest texture","Cores are ~0.6 pure — the seven all-encoder families are the clean, reportable set, stated alongside the mixed axes rather than hidden.")]
y0=2.5; rh=1.02
for i,(t,d) in enumerate(items):
    y=y0+i*rh
    oval(s,MX,y,0.6,0.6,SEA if i<3 else AMBER)
    txt(s,MX,y,0.6,0.6,[(str(i+1),{"color":INK,"font":HF,"size":22,"bold":True})],align=PP_ALIGN.CENTER,anchor=MSO_ANCHOR.MIDDLE)
    txt(s,MX+0.9,y-0.05,11.2,0.95,[(t+"   ",{"bold":True,"color":WHITE,"font":HF,"size":16}),
        (d,{"color":MUTE1,"font":BF,"size":13.5})],anchor=MSO_ANCHOR.MIDDLE,space=1.05)
txt(s,MX,6.78,12,0.4,[("Next: restructure the manuscript with portability as the lead result.",
    {"color":MUTE2,"font":BF,"size":12,"italic":True})])
notes(s,"Takeaways: portability, cross-instrument transfer replicating across six encoders, robustness by design, and honest reporting of core purity. Next step: manuscript restructure with portability as the lead.")

out="/data_gpu5/ROSS_Proj/examples/global_pool/global_pool_results.pptx"
prs.save(out)
print("saved", out, "slides:", len(prs.slides._sldIdLst))
```
