## Supplemental code for "Scoring respondents on a semantic frame of reference identifies a group that is burdened across many questionnaires and reaches care less easily": build_processed_pool.py

```
import csv, glob, re
BASE='/data_gpu5/ROSS_Proj/examples/global_pool/'

# (regex anchored at start, replacement, stem_removed label).
# Strip the leading TIME phrase only; KEEP the "how often have you / have you / have you ever"
# response wrappers so cleaned items stay whole sentences (no fragments).
RULES = [
    (r'^In the last month,\s*', '', 'In the last month,'),                 # PSS-10  (keeps "how often have you...")
    (r'^In the past month,\s*', '', 'In the past month,'),                 # PC-PTSD-5 (keeps "have you...")
    (r'^In your life,\s*', '', 'In your life,'),                           # MHQ / CIDI (keeps "have you ever...")
    (r'^How often during the last year have you\s+', 'How often have you ', 'during the last year'),  # AUDIT 4-8
    (r'^Has there been a time when you were not your usual self and\s*', '', 'Has there been a time when you were not your usual self and'),  # MDQ (result is a whole clause)
    (r'^In the past 7 days,?\s*', '', 'In the past 7 days'),               # PROMIS banks / Global Health
    (r'^Over the last two weeks,?\s*', '', 'Over the last two weeks,'),    # WHO-5
    (r'^Within the past 12 months,?\s*', '', 'Within the past 12 months,'),# HVS
    (r'^In the last 12 months,?\s*', '', 'In the last 12 months,'),        # USDA
    (r'^In the past week,?\s*', '', 'In the past week,'),                  # OASIS (keeps "how often have you...")
]
RULES = [(re.compile(p, re.IGNORECASE), rep, lab) for p, rep, lab in RULES]

def recap(s):
    return re.sub(r'^([^A-Za-z]*)([a-z])', lambda m: m.group(1)+m.group(2).upper(), s, count=1)

def clean(text):
    for pat, rep, lab in RULES:
        if pat.match(text):
            return recap(pat.sub(rep, text, count=1)), lab
    return text, ''

roles={r['scale']:r['role'] for r in csv.DictReader(open(BASE+'instrument_roles.csv'))}

rows=[]
for f in sorted(glob.glob(BASE+'*.csv')):
    rdr=csv.DictReader(open(f))
    if not {'item_id','prompt','source','scale','construct'} <= set(rdr.fieldnames or []): continue
    for r in rdr:
        cl,rm=clean(r['prompt'])
        rows.append(dict(item_id=r['item_id'], source=r['source'], scale=r['scale'],
                         construct=r['construct'], role=roles.get(r['scale'],'?'),
                         stripped=('yes' if rm else 'no'),
                         prompt_raw=r['prompt'], prompt_clean=cl, stem_removed=rm))

with open(BASE+'pool_processed.csv','w',newline='') as fh:
    w=csv.DictWriter(fh, fieldnames=['item_id','source','scale','construct','role','stripped',
                                     'prompt_raw','prompt_clean','stem_removed'])
    w.writeheader(); w.writerows(rows)

changed=[r for r in rows if r['stem_removed']]
print('TOTAL items:', len(rows), '| stem stripped:', len(changed), '| unchanged:', len(rows)-len(changed))
from collections import Counter
print('stripped by role:', dict(Counter(r['role'] for r in changed)))
print('\n=== stems removed ===')
for k,v in Counter(r['stem_removed'] for r in changed).most_common(): print('  %3d  %r' % (v,k))
print('\n=== wrapper-kept examples (was fragment before) ===')
for src in ['Cohen-PSS10','PC-PTSD-5','AUDIT','MHQ','WMH-CIDI','OASIS']:
    ex=[r for r in changed if r['source']==src]
    if ex:
        print('  [%s] %r' % (src, ex[0]['prompt_raw']))
        print('        -> %r' % (ex[0]['prompt_clean'],))
```
