## Supplemental code for "Scoring respondents on a semantic frame of reference identifies a group that is burdened across many questionnaires and reaches care less easily": build_sources.py

```
import csv, glob
from collections import OrderedDict

BASE='/data_gpu5/ROSS_Proj/examples/global_pool/'

# verification + reference by source tag
V = {
 'PROMIS-CDE': ('pre-existing curation','curated PROMIS item banks (pre-existing)'),
 'PROMIS':     ('pre-existing curation','curated PROMIS item bank (pre-existing)'),
 'PROMIS-GH':  ('AoU items_full.csv','manuscript/supplementary_data/items_full.csv'),
 'IPIP-50':    ('web-verified','https://ipip.ori.org/New_IPIP-50-item-scale.htm'),
 'Williams-EDS':('pre-existing curation','Williams EDS curation (pre-existing)'),
 'Williams-MED':('pre-existing curation','Williams MED curation (pre-existing)'),
 'Williams-HV': ('pre-existing curation','Williams HV curation (pre-existing)'),
 'PQ-B':       ('web-verified','https://psytests.org/diag/pqben-bl.html'),
 'CES-D':      ('standard public-domain','Radloff 1977 standard wording (2 items confirmed via web search)'),
 'PHQ-9':      ('standard public-domain','PHQ-9 standard public-domain (web search)'),
 'GAD-7':      ('standard public-domain','GAD-7 standard public-domain (web search)'),
 'AUDIT':      ('web-verified','https://nida.nih.gov/sites/default/files/files/AUDIT.pdf'),
 'DAST-10':    ('web-verified','https://oasas.ny.gov (DAST-10, web search)'),
 'ISEL-12':    ('web-verified','https://db.arabpsychology.com/scales/interpersonal-support-evaluation-list-12/'),
 'ACE':        ('web-verified','Felitti ACE standard wording (web search)'),
 'Cohen-PSS10':('standard public-domain','Cohen PSS-10 standard public-domain (web search)'),
 'RSES':       ('web-verified','https://www.apa.org/obesity-guideline/rosenberg-self-esteem.pdf'),
 'Hager-HVS':  ('standard public-domain','Hager 2010 Hunger Vital Sign (web search)'),
 'USDA-HFSSM': ('standard public-domain','USDA HFSSM 6-item Short Form (web search)'),
 'Sampson-1997':('web-verified','https://sparqtools.org/mobility-measure/collective-efficacy-scale/'),
 'E-HITS':     ('web-verified','https://portal.ct.gov/-/media/DPH/Maternal-Mortality/E-HITS-screening.pdf'),
 'PC-PTSD-5':  ('web-verified','https://www.ptsd.va.gov/professional/assessment/documents/pc-ptsd5-screen.pdf'),
 'WHO-5':      ('web-verified','https://cdn.who.int/media/docs/default-source/mental-health/who-5_english-original.pdf'),
 'Hughes-UCLA3':('standard public-domain','Hughes 2004 UCLA 3-item (web search)'),
 'MDQ':        ('web-verified','https://scales.arabpsychology.com/s/mood-disorder-questionnaire-mdq/'),
 'ASRS':       ('web-verified','https://add.org/wp-content/uploads/2015/03/adhd-questionnaire-ASRS111.pdf'),
 'NDS':        ('AoU items_full.csv','manuscript/supplementary_data/items_full.csv'),
 'DMS':        ('AoU items_full.csv','manuscript/supplementary_data/items_full.csv'),
 'DSES':       ('AoU items_full.csv','manuscript/supplementary_data/items_full.csv'),
 'MHQ':        ('AoU items_full.csv','manuscript/supplementary_data/items_full.csv'),
 'WMH-CIDI':   ('AoU items_full.csv','manuscript/supplementary_data/items_full.csv'),
 'OASIS':      ('web-verified','https://depts.washington.edu/uwhatc (OASIS)'),
 'ISI':        ('web-verified','https://www.med.upenn.edu/snaplab/assets/user-content/documents/insomnia-severity-index.pdf'),
 'MSPSS':      ('web-verified','https://emerge.ucsd.edu/r_2awuzr8vphwvy2o/'),
 'DJGLS-6':    ('web-verified','https://research.vu.nl (de Jong Gierveld 2006 6-item)'),
 'SWLS':       ('web-verified','http://www.recoveryanswers.org/assets/satisfaction_with_life_scale.pdf'),
 'WHOQOL-BREF':('web-verified','http://www.recoveryanswers.org/assets/whoqol-bref.pdf'),
 'PCL-5':      ('web-verified','https://www.ptsd.va.gov/professional/assessment/documents/PCL5_Standard_form.PDF'),
 'LEC-5':      ('web-verified','https://www.ptsd.va.gov/professional/assessment/documents/LEC5_Standard_Self-report.pdf'),
 'SBQ-R':      ('web-verified','https://youthsuicideprevention.nebraska.edu/.../SBQ-R.pdf'),
 'PHQ-15':     ('web-verified','https://providers.bcidaho.com (PHQ-15)'),
 'BRS':        ('web-verified','https://ogg.osu.edu (Brief Resilience Scale)'),
 'LOT-R':      ('web-verified','https://www.psy.miami.edu/faculty/ccarver/lot-r.html'),
 'MLQ':        ('web-verified','https://cic.edu (Meaning in Life Questionnaire)'),
 'ERQ':        ('web-verified','https://db.arabpsychology.com/scales/emotion-regulation-questionnaire-erq/'),
}

# item counts per (source, scale) and constructs
counts=OrderedDict()
for f in sorted(glob.glob(BASE+'*.csv')):
    if f.endswith('instrument_roles.csv') or f.endswith('sources.csv'): continue
    for r in csv.DictReader(open(f)):
        k=(r['source'], r['scale'])
        counts.setdefault(k, {'n':0,'cons':set(),'file':f.split('/')[-1]})
        counts[k]['n']+=1; counts[k]['cons'].add(r['construct'])

roles={(r['source'],r['scale']):r['role'] for r in csv.DictReader(open(BASE+'instrument_roles.csv'))}

with open(BASE+'sources.csv','w',newline='') as fh:
    w=csv.writer(fh)
    w.writerow(['source','scale','role','n_items','constructs','verification','source_ref','file'])
    miss_v=[]; miss_r=[]
    for (src,scale),d in counts.items():
        role=roles.get((src,scale),'??')
        if role=='??': miss_r.append((src,scale))
        ver,ref=V.get(src,('??','??'))
        if ver=='??': miss_v.append(src)
        w.writerow([src,scale,role,d['n'],'; '.join(sorted(d['cons'])),ver,ref,d['file']])
print('sources.csv rows:', len(counts))
print('missing role:', miss_r or 'none')
print('missing verification:', sorted(set(miss_v)) or 'none')
# role/verif rollup
from collections import Counter
vc=Counter(V.get(src,('??',''))[0] for (src,scale) in counts)
print('verification rollup (by scale):', dict(vc))
```
