## Supplemental code for "Scoring respondents on a semantic frame of reference identifies a group that is burdened across many questionnaires and reaches care less easily": README.md

# Global item pool

A cross-instrument pool of validated psychosocial questionnaire items, assembled from their
published item text alone. **Purpose:** a portability / meaning-transfer demonstration for the
NHB manuscript — hold the All of Us (AoU) text-derived components fixed, place these
external-instrument items on them from wording alone, and show that items land on the component
their construct defines (and, where independent instruments measure the same construct, that
they co-locate). Pool-level analogue of the HRS transfer (`../randhrsdata/`), without needing
respondents.

**Current size: 577 items · 50 scales · 53 constructs · 39 item files** (14 anchor scales /
36 independent). All `item_id`s unique, all files parse, every scale covered by the role map.

## Files in this folder
- `*_items.csv` (39 files) — the item pool, one instrument (or instrument family) per file.
- **`instrument_roles.csv`** — authoritative anchor/independent map, keyed by scale
  (`source, scale, construct, role, aou_match, note`). Roles split *within* the discrimination
  and food-insecurity files, so this is keyed at scale level, not file level.
- **`sources.csv`** — provenance/verification per scale
  (`source, scale, role, n_items, constructs, verification, source_ref, file`).
- **`pool_processed.csv`** — combined 577-item pool with time-frame boilerplate stems stripped
  (`item_id, source, scale, construct, role, stripped, prompt_raw, prompt_clean, stem_removed`).
  See "Stem-stripping" below. Regenerate with `python3 build_processed_pool.py`.
- **`build_processed_pool.py`** / **`build_sources.py`** — regenerate `pool_processed.csv` /
  `sources.csv` from the item files.
- `README.md` — this file.

## Schema (item files)
One row per item. Columns (5, `QUOTE_MINIMAL`): `item_id` (unique across the pool),
`prompt` (canonical item text, self-contained — the only field that gets embedded),
`source` (short provenance tag), `scale` (full instrument name),
`construct` (grouping / ground-truth label — **not embedded**, used only as the analysis-time
grouping key). `promis_items.csv` additionally carries an empty `stem` column (harmless legacy).

## Design: strong (independent) vs anchor (AoU-overlapping)
The strong transfer test uses instruments **AoU never administered** — landing those correctly
proves the components generalize beyond the fitting data. AoU-matched instruments are kept as
**anchors** (sanity check), prunable for a strictly independent pool.

**Roles are keyed at the INSTRUMENT level against AoU Table 2** (the 17 validated scales in the
151 AoU items) — not the construct level. A construct can be in AoU via one instrument while the
pool uses a *different* instrument for it (e.g. CES-D vs AoU's PHQ-9 for depression): that is
still an independent/strong test. An item is an *anchor* only when AoU embedded that same item
text (or a literal subset, like UCLA-3 ⊂ ULS-8).

- **Anchor scales (14):** Williams EDS, PHQ-9, GAD-7, PSS-10, Hunger Vital Sign, Neighborhood
  Social Cohesion (=AoU SCNS), UCLA-3 loneliness (⊂ ULS-8), ASRS (ADHD), NDS (neighborhood
  disorder), DMS (medical-setting discrimination), DSES (spirituality), PROMIS Global Health,
  MHQ, WMH-CIDI. The AoU-sourced anchors use AoU's own text from
  `../../manuscript/supplementary_data/items_full.csv`. MHQ and WMH-CIDI are multi-construct and
  carry AoU's own IPV / psychosis / mania items — the direct anchors for E-HITS / PQ-B / MDQ.
  **All 17 AoU Table-2 scales are represented.**
- **Independent scales (36):** everything else — see `instrument_roles.csv`.

Multi-instrument constructs (strongest test — independent + AoU anchor should co-locate):
Depression = CES-D / PROMIS-Dep (indep) + PHQ-9 (anchor); Anxiety = PROMIS-Anx / OASIS (indep) +
GAD-7 (anchor); Big Five = IPIP-50 (indep) vs AoU BFI-2-XS; Loneliness = PROMIS Social Isolation /
DJGLS-6 (indep) + UCLA-3 (anchor); Social support = ISEL-12 / MSPSS (indep) vs AoU mMOS-SS;
Food insecurity = USDA (indep) + HVS (anchor); PTSD = PC-PTSD-5 / PCL-5 (both indep).

## Inventory (all 39 files)

| file | items | role | scale(s) (n) |
|------|------:|------|--------------|
| `promis_items.csv` | 136 | indep | 7 PROMIS banks: Depression (28), Anxiety (29), Anger (22), Sleep Disturbance (27), Sleep-Related Impairment (16), Social Isolation (8), Emotional Support (6) |
| `ipip50_items.csv` | 50 | indep | IPIP Big-Five Factor Markers (50) |
| `quality_of_life_whoqol_items.csv` | 26 | indep | WHOQOL-BREF (26) |
| `discrimination_items.csv` | 23 | mixed | Everyday Discrimination Scale (9, anchor); Major Experiences of Discrimination (9, indep); Heightened Vigilance (5, indep) |
| `psychosis_items.csv` | 21 | indep | Prodromal Questionnaire-Brief (PQ-B) (21) |
| `depression_cesd_items.csv` | 20 | indep | CES-D (20) |
| `ptsd_pcl5_items.csv` | 20 | indep | PTSD Checklist for DSM-5 (PCL-5) (20) |
| `substance_use_items.csv` | 20 | indep | AUDIT (10); DAST-10 (10) |
| `trauma_exposure_lec_items.csv` | 17 | indep | Life Events Checklist for DSM-5 (LEC-5) (17) |
| `phq_gad_items.csv` | 16 | anchor | PHQ-9 (9); GAD-7 (7) |
| `somatic_phq15_items.csv` | 15 | indep | PHQ-15 (15) |
| `mania_items.csv` | 13 | indep | Mood Disorder Questionnaire (MDQ) (13) |
| `neighborhood_disorder_items.csv` | 13 | anchor | Perceived Neighborhood Disorder (AoU NDS) (13) |
| `mhq_items.csv` | 12 | anchor | UK Biobank Mental Health Questionnaire (12, multi-construct) |
| `social_support_items.csv` | 12 | indep | ISEL-12 (12) |
| `social_support_mspss_items.csv` | 12 | indep | MSPSS (12) |
| `ace_items.csv` | 10 | indep | ACE Questionnaire (Felitti) (10) |
| `emotion_regulation_erq_items.csv` | 10 | indep | Emotion Regulation Questionnaire (ERQ) (10) |
| `meaning_mlq_items.csv` | 10 | indep | Meaning in Life Questionnaire (MLQ) (10) |
| `perceived_stress_items.csv` | 10 | anchor | PSS-10 (10) |
| `self_esteem_items.csv` | 10 | indep | Rosenberg Self-Esteem Scale (10) |
| `cidi_items.csv` | 9 | anchor | WHO World Mental Health CIDI (9, multi-construct) |
| `food_insecurity_items.csv` | 8 | mixed | Hunger Vital Sign (2, anchor); USDA HFSSM 6-item (6, indep) |
| `insomnia_isi_items.csv` | 7 | indep | Insomnia Severity Index (ISI) (7) |
| `medical_discrimination_items.csv` | 7 | anchor | Discrimination in Medical Settings (AoU DMS) (7) |
| `adhd_items.csv` | 6 | anchor | ASRS v1.1 6-item screener (6) |
| `loneliness_djg_items.csv` | 6 | indep | De Jong Gierveld Loneliness Scale (6-item) (6) |
| `optimism_lotr_items.csv` | 6 | indep | LOT-R (6 scored) |
| `resilience_brs_items.csv` | 6 | indep | Brief Resilience Scale (BRS) (6) |
| `spirituality_items.csv` | 6 | anchor | Daily Spiritual Experiences (AoU DSES) (6) |
| `anxiety_oasis_items.csv` | 5 | indep | OASIS (5) |
| `ipv_items.csv` | 5 | indep | Extended HITS (E-HITS) (5) |
| `life_satisfaction_swls_items.csv` | 5 | indep | Satisfaction with Life Scale (SWLS) (5) |
| `neighborhood_items.csv` | 5 | anchor | Neighborhood Social Cohesion (AoU SCNS) (5) |
| `ptsd_items.csv` | 5 | indep | PC-PTSD-5 (5) |
| `wellbeing_items.csv` | 5 | indep | WHO-5 Well-Being Index (5) |
| `suicidality_sbqr_items.csv` | 4 | indep | Suicidal Behaviors Questionnaire-Revised (SBQ-R) (4) |
| `loneliness_items.csv` | 3 | anchor | UCLA-3 Loneliness (⊂ AoU ULS-8) (3) |
| `promis_globalhealth_items.csv` | 3 | anchor | PROMIS Global Health (AoU) (3) |

## Provenance / verification
Full per-scale provenance is in `sources.csv`. Summary of how wording was obtained:
**web-verified against a cited public source — 27 scales** (IPIP-50, PQ-B, ISEL-12, Neighborhood
Cohesion, E-HITS, AUDIT, DAST-10, PC-PTSD-5, WHO-5, Rosenberg, MDQ, ASRS, OASIS, ISI, MSPSS,
DJGLS-6, SWLS, WHOQOL-BREF, PCL-5, LEC-5, SBQ-R, PHQ-15, BRS, LOT-R, MLQ, ERQ, and one more).
**AoU's own item text from `items_full.csv` — 6 scales** (NDS, DMS, DSES, PROMIS Global Health,
MHQ, WMH-CIDI). **standard public-domain wording (web-searched, not full-PDF-verified) — 7 scales**
(CES-D, PHQ-9, GAD-7, PSS-10, UCLA-3, HVS, USDA-6). **pre-existing curation — 10 scales**
(the 7 PROMIS banks + Williams EDS/MED/HV). A final spot-check against source PDFs is still
advisable before freezing the pool for the paper.

## Decisions on record
- **Excluded on review** (unsuitable for text embedding): **QIDS-SR-16** (items are bare topic
  headings), **ASRM** (verbatim is the score-0 "no symptom" pole), **PANAS** (single mood
  adjectives embed ambiguously). **LOT-R** fillers (items 2, 5, 6, 8) dropped.
- **AUDIT-10 replaced AUDIT-C** in `substance_use_items.csv`.
- **ACE is independent** — AoU used the BRFSS ACE, the pool uses Felitti wording (different item
  text). Kept Felitti rather than swapping to BRFSS.
- **EDS_1/2 ≡ DMS_1/2** are textually identical across the Everyday Discrimination Scale (anchor)
  and Discrimination in Medical Settings (anchor). Kept intentionally — identical text should land
  in the same place. More broadly, EDS and DMS share several near-identical stems, so
  everyday-vs-healthcare discrimination is barely separable by wording (by design).
- **IPIP:** `../IPIP.txt` = the entire IPIP master bank (~3,320 items, unlabeled). The pool uses
  the labeled **IPIP-50 Big-Five Factor Markers** over 20-item Mini-IPIP. Optional future work:
  project all ~3,320 IPIP items onto the fixed AoU components as an unlabeled large-scale
  organization test (needs IPIP scale keys to label; not built).

## AoU coverage — complete
All 17 AoU Table-2 validated scales are represented (as anchors with AoU's own text and/or paired
with an independent counterpart). The 13 AoU standalone single items are not individually mirrored
(they are not validated multi-item scales). AoU instruments carried as broad multi-construct
anchors: MHQ, WMH-CIDI.

## Construct-label harmonization TODO (small, analysis-time)
Construct labels are metadata (never embedded), so these do not affect landing — they only matter
as the grouping key when scoring co-location. A few near-duplicate labels to merge then:
`Interpersonal & sexual violence` (MHQ) ≈ `Interpersonal violence` (E-HITS);
`Subjective wellbeing` (MHQ) ≈ `Well-being` (WHO-5); optionally
`Life-threatening events` (MHQ/CIDI) ⊂ `Trauma exposure` (LEC-5), and
`Insomnia` (ISI) vs the PROMIS sleep banks. The manuscript construct map uses "Interpersonal &
sexual violence".

## Stem-stripping (processed pool)
`pool_processed.csv` removes repeated **time-frame / recall-window** boilerplate that would
otherwise make items cluster by frame instead of content. **162 of 577 items** had a leading
time phrase removed. Both `prompt_raw` and `prompt_clean` are kept (reversible, auditable via
`stem_removed`; `stripped` = yes/no). **Only the leading time phrase is removed — the
`how often have you` / `have you` / `have you ever` response wrappers are kept, so every cleaned
item stays a whole sentence (no fragments).** Verified: 0 fragments; every cleaned prompt begins
with a subject/aux/wh-word.
- **Removed** (leading time phrase only): `In the past 7 days` (PROMIS/PROMIS-GH, 107),
  `Has there been a time when you were not your usual self and` (MDQ, 13 — result is still a whole
  clause, "You felt so good or hyper that…"), `In the last month,` (PSS-10), `In your life,`
  (MHQ/CIDI), `In the past week,` (OASIS), `In the past month,` (PC-PTSD-5), `during the last year`
  (AUDIT 4-8, internal), `Over the last two weeks,` (WHO-5), `Within the past 12 months,` (HVS),
  `In the last 12 months,` (USDA).
- **Kept deliberately** (load-bearing): `While you were growing up,` (ACE — childhood is the
  construct), `How often does your partner` (E-HITS — partner violence), and the WHOQOL/UCLA
  response frames.
- **Anchors are embedded stripped too** (per decision): 19 of the stripped items are AoU anchors.
  Embed `prompt_clean` for all items (anchors included); the `role=anchor` and `stem_removed`
  columns record which anchors were stripped and what came off. `prompt_raw` is kept for audit.
- **Not applied (optional level 2):** stripping response frames from WHOQOL/UCLA to expose bare
  content — more aggressive, left off by default.

## Analysis — see `RESULTS.md` for the organized results
Full results (numbers, per-encoder tables, output map, caveats) are in **`RESULTS.md`**. In brief,
the `../allofus` pipeline repointed at the pool (`ROSS_NPZ_PREFIX=pool_items`,
`ROSS_OUT_DIR=examples/allofus/outputs_pool`, all CPU, embed `prompt_clean`):
`0_embed_pool.sh → 1_select_k.py → 2_fit.py --lambda-score 1.0 → pool_fixedpoint_cores.py →
pool_project_aou.py → pool_colocation.py`. Outputs under `../allofus/outputs_pool/`.

- **k (Horn's PA)** 49–55 (median 52). **Fit** at `lambda_score=1.0` (dense Z, expected).
- **Cores = iterated fixed-point PR** everywhere (never `defining_sets`, never CPP, W un-pruned).
- **Pool self-organizes** by construct (median core 7, purity ~0.6–0.68).
- **AoU→pool:** 88–94% of AoU items covered, 5–9 pure axes/encoder, robust to AoU's stems.
- **★ Cross-instrument co-location (core-to-core, `pool_colocation.py`):** AoU items land in cores
  *defined by different independent instruments* of the same content — **7 families in all 6 encoders**
  (childhood adversity, psychosis, loneliness, social support, food insecurity, trauma/IPV,
  suicidality) plus 5 more in 4–5/6. Multi-content instruments correctly scatter by content.
  **Supersedes the old argmax "90/87/78/77"** (wrong lens — tallied per instrument).

## Figures, cohorts, deck — DONE (see `RESULTS.md` §7–8)
Core-based figures (gte exemplar, no domain colour): pool ribbon + cards, AoU→pool ribbon (red = the 3
external instruments ACE-BRFSS/MOS-SS/BFI-2-XS, grey = anchor) + cards, MHQ split, and **cohort transfer
ribbons for HRS and Xinxiang**. Generators `fig_*.py` in `../allofus/`, PNGs in `figures/`. Xinxiang got
its own item-text file (`../Xinxiang_data/xinxiang_items.csv`, embedded fresh). Slide deck
`global_pool_results.pptx` (14 slides), generator `build_deck.py` here.

## Not yet done
Restructure `manuscript_nhb/` with **portability as the lead** (see `RESULTS.md` §9). Decided against:
re-embedding AoU stem-stripped (the with-stem → stripped-pool asymmetry already *is* the robustness
result), and the reverse direction (pool onto fixed AoU components) — portability, AoU-onto-pool, is
the lead instead.
