## Supplemental code for "Scoring respondents on a semantic frame of reference identifies a group that is burdened across many questionnaires and reaches care less easily": pyproject.toml

```
[build-system]
requires = ["setuptools>=64"]
build-backend = "setuptools.build_meta"

[project]
name = "ross-proj"
version = "0.1.0"
description = "Sparse-projection PCA: a smooth penalty that sparsifies the component scores Z = X_c·W, for interpretability."
readme = "README.md"
requires-python = ">=3.10"
license = { text = "UNLICENSED" }
authors = [{ name = "ROSS-Proj authors" }]
keywords = ["sparse-pca", "interpretability", "jax", "single-cell"]
classifiers = [
    "Programming Language :: Python :: 3",
    "Intended Audience :: Science/Research",
    "Topic :: Scientific/Engineering",
]

# Core runtime dependencies (CPU). For GPU, install the CUDA build of JAX,
# e.g. `pip install "jax[cuda12]"`.
dependencies = [
    "numpy",
    "scipy",
    "scikit-learn",
    "jax",
    "optax",
    "tqdm",
]

[project.optional-dependencies]
# Single-cell data loading (.h5ad) for cpp_z on real data.
data = ["scanpy", "anndata"]
dev = ["pytest"]

[project.scripts]
ross-proj-select-k = "ross_proj.parallel_analysis:main"
ross-proj-fit = "ross_proj.fit:main"
ross-proj-cpp-z = "ross_proj.cpp_z:main"

[tool.setuptools]
packages = ["ross_proj"]
```
