## Supplemental code for "Scoring respondents on a semantic frame of reference identifies a group that is burdened across many questionnaires and reaches care less easily": README.md

# Scoring respondents on a semantic frame of reference

Code for the manuscript *"Scoring respondents on a semantic frame of reference identifies a group
burdened across many instruments and reaching care less easily"* (`manuscript_nhb/`).

The method builds one **reference basis** from the published text of 577 questions drawn from 50
instruments, holds it fixed, places any further questionnaire battery on it from that battery's item
text alone, and scores the people who answered — giving each respondent one position, one distance,
and an attribution of that distance to named components.

No responses enter the basis. Everything in stages A to C below runs on published question text and
can be reproduced without access to any cohort. Stage D needs individual-level All of Us responses,
which cannot leave the Researcher Workbench.

---

## Layout

| path | what it is |
|---|---|
| `ross_proj/` | the fitting library: sparse-projection PCA, parallel analysis, core sets (method write-up: [`docs/METHOD.md`](docs/METHOD.md)) |
| `examples/global_pool/` | assembling the 577-item reference pool from published instruments |
| `examples/allofus/` | the pipeline, and every figure and table generator that runs locally |
| `all_of_us_codes/` | the screen, scoring and attribution — these run **inside the Workbench** |
| `manuscript_nhb/` | the manuscript, supplement, cover letter, and the figure map |
| `docs/` | background: the method write-up, the instrument citation log, the prior-work digest |
| `archive/` | superseded work, kept for provenance and referenced by nothing |

## Environment

```bash
CUDA_VISIBLE_DEVICES="" /data_gpu5/ROSS_Proj/env/bin/python <script>.py
```

The environment is `env/` (see `environment.yml`). Everything below is CPU-only and reproducible on
a laptop except stage A2, the embedding, which is faster with a GPU. **Fits are device-dependent:**
a GPU fit and a CPU fit of the same data give different bases, and the published basis is the CPU
fit — so reproduce on CPU if you want the published numbers.

Stages A to C are run from `examples/allofus/` unless stated. Two environment variables switch the
pipeline from its default onto the pool:

```bash
export ROSS_NPZ_PREFIX=pool_items
export ROSS_OUT_DIR=/data_gpu5/ROSS_Proj/examples/allofus/outputs_pool
```

---

## Stage A — build the reference basis

Run once. The output is the portable object every later stage reads.

| # | command | produces |
|---|---|---|
| A1 | `python examples/global_pool/build_processed_pool.py` | `pool_processed.csv` — the 577 pool items, with the leading recall-window phrase stripped (162 of them) |
| A2 | `python 0_embed.py` | `data/pool_items_<encoder>.npz`, one per encoder, items as samples |
| A3 | `python 1_select_k.py` | `k` per encoder by Horn's parallel analysis on column-permuted embeddings (49 to 55) |
| A4 | `python 2_fit.py --lambda-score 1.0` | the sparse projection `P` per encoder |
| A5 | `python random_starts.py` then `python select_projection.py` | the operating projection, chosen by fixed-point core energy over random restarts |
| A6 | `python pool_fixedpoint_cores.py` | `outputs_pool/fixedpoint_cores/<encoder>_cores.csv` — each component's core, the fixed point of the participation-ratio pruning |
| A7 | `python pool_core_columns.py` | the dual object: each item's core components |
| A8 | `python build_reference_basis_package.py` | `reference_basis/<encoder>.npz` — `W`, `mu`, `sigma`, `mu_scores`, `band`. This folder is self-contained and is what the Workbench needs |

The penalty weight is set once by `lambda_sweep_pool.py` (Supplementary Note S15); the published fits
use λ_score = 1.0 with λ_ortho = 0.1.

## Stage B — place a battery on the fixed basis

Nothing is refitted here. A battery is embedded with the same encoder, projected onto the stored
components, and divided by the stored per-column standard deviations.

| command | produces |
|---|---|
| `python build_aou_poolw_projection.py` | the All of Us battery placed on the basis, and the kept components |
| `python make_core_items_pool.py` | Supplementary Data 3: the battery items forming each kept component, with instrument, construct and question text |
| `python hrs_kept_components.py` | the same for the HRS battery, and the keep decisions across encoders |

The keep rule lives in `_common.py` (`pool_bands`, `keep_columns`) and is the definition of record —
downstream scripts must not carry their own copy.

## Stage C — figures and tables that need no responses

Each generator reads saved artefacts and hardcodes no result. `manuscript_nhb/FIGURES.md` maps every
figure and table to its generator; the main ones:

```bash
python fig1_pool.py && python fig1_panels.py   # Figure 1 panels, then sh manuscript_nhb/build_panels.sh
python fig_pool_ribbon.py                      # Figure 2b
python fig_xenc.py                             # Figure 3, Figs S5 and S6, Supplementary Data 5
python fig_aou_twoside_cores.py                # Figure 4
python fig_hrs_twoside_cores.py --all          # Figs S9 to S14
python supp_pool_composition.py                # Table S12
```

Two generators write into `examples/global_pool/figures/`, so their output must be copied into
`manuscript_nhb/figures/` by hand: `fig_aou_twoside_cores.py` and `fig_hrs_twoside_cores.py`.
`fig_xenc.py` writes straight into the manuscript's figure directory.

## Stage D — the screen, inside the Researcher Workbench

Individual-level responses cannot leave All of Us, so these run there. Copy `all_of_us_codes/` and
the `reference_basis/` folder from A8 into the Workbench and run in order:

| # | command | produces |
|---|---|---|
| D1 | `python screen_battery.py` | per-encoder distances and `consensus_outliers.csv` — the 510 flagged under all six encoders |
| D2 | `python cutoffs.py` | `cutoff_flags.csv` — each participant against the eight validated criteria |
| D3 | `python attribution.py` and `attribution_components.py` | the distance partitioned over components, with the joint/marginal split and integration |
| D4 | `python joint_only.py` | the neutralization test behind Supplementary Note S9 |
| D5 | `python five_items_to_drop.py` | the five clinically invisible outliers, and how many items unflag them |
| D6 | `python response_style.py` | content extremity and endpoint use, the response-style controls |
| D7 | `python underserved_setup.py` then `underserved.py` | the health-record and access-survey linkage |
| D8 | `python fig_screen.py`, `fig_component_ranks.py`, `fig_jaccard_threshold.py` | Figures 5, 6 and S2 |

Only aggregate outputs leave the Workbench. `table_five.py`, `five_xenc.py`, `severity_baseline.py`
and `zero_breadth.py` produce the remaining tables and reported numbers from what D1 to D7 wrote.

---

## Reproducing a number from the manuscript

Every reported number has a generator, and no generator hardcodes a result. To check one, find it in
`manuscript_nhb/FIGURES.md` (figures and tables) or in the Methods, which names the generator for
each analysis — for example `construct_splitting.py`, `core_separation.py`, `dense_control.py`,
`lambda_sweep_pool.py`, and `all_of_us_codes/five_items_to_drop.py`.

## Data availability

The 577 pool items and the 151 All of Us items are published question text and ship with the
manuscript as Supplementary Data 1 and 4, so stages A to C are fully reproducible. Individual-level
All of Us responses are available only to authorized investigators through the controlled-tier
Researcher Workbench, so stage D can be re-run only there.
