## Supplemental code for "Scoring respondents on a semantic frame of reference identifies a group that is burdened across many questionnaires and reaches care less easily": __init__.py

```
"""ROSS-Proj — sparse-projection PCA.

A sibling of ROSS-PCA that adds a smooth penalty on the *projections*
Z = X_c·W, so each component's scores are sparse (each factor active for only
a few samples). Aim: interpretability — component j ↔ a small set of
high-scoring samples. Exact sparsity comes from a pruning step on Z.

Modular pipeline (each stage returns a typed result; see :mod:`ross_proj.results`)

1. Choose k — :func:`ross_proj.select_k`  (permutation Parallel Analysis)
       CLI: ``python -m ross_proj.parallel_analysis``
2. Fit     — :func:`ross_proj.fit_projection`  (X, k, λ -> mean, W, Z)
       CLI: ``python -m ross_proj.fit``
3. Prune   — :func:`ross_proj.prune_and_report`  (cosine-preserving + participation)
       CLI: ``python -m ross_proj.cpp_z``

``select_k`` + ``fit_projection`` are also bundled as
:func:`ross_proj.parallel_analysis.fit_with_selected_k`.

Estimator: :class:`SparseProjectionPCA` (scikit-learn style). For a full worked
example see ``examples/allofus/``.

Example
-------
>>> import numpy as np
>>> from ross_proj import select_k, fit_projection, prune_and_report
>>> X = np.random.randn(2000, 50)              # UNCENTERED; fit centers internally
>>> k = select_k(X, n_perm=200).k              # objective model order
>>> fit = fit_projection(X, n_components=k, lambda_score=1.0)
>>> rep = prune_and_report(X, fit.W, fit.mean) # rep.Z_pruned, rep.participation_ratio
"""

from .sparse_projection import SparseProjectionPCA
from .fit import fit_projection
from .parallel_analysis import select_k, parallel_analysis, fit_with_selected_k
from .cpp_z import (
    run_sweep,
    cpp_precompute,
    cpp_prune_fast,
    select_elbow,
    select_dtheta_ds,
    participation_ratio,
    prune_and_report,
)
from .defining_sets import defining_sets, overlap_stats, DefiningSets
from .io import load_data, load_model
from .results import FitResult, KResult, PruneResult

__version__ = "0.1.0"

__all__ = [
    # estimator
    "SparseProjectionPCA",
    # stage 1: choose k
    "select_k",
    "parallel_analysis",
    "fit_with_selected_k",
    # stage 2: fit
    "fit_projection",
    # stage 3: prune + report
    "run_sweep",
    "prune_and_report",
    "participation_ratio",
    "cpp_precompute",
    "cpp_prune_fast",
    "select_elbow",
    "select_dtheta_ds",
    # stage 3 (interpretation): defining sets via participation ratio
    "defining_sets",
    "overlap_stats",
    "DefiningSets",
    # io + result types
    "load_data",
    "load_model",
    "FitResult",
    "KResult",
    "PruneResult",
    "__version__",
]
```
