## Supplemental code for "Scoring respondents on a semantic frame of reference identifies a group that is burdened across many questionnaires and reaches care less easily": cpp_z.py

```
#!/usr/bin/env python
"""cpp_z.py — Cosine-Preserving Pruning of the *projections* Z (scores).

Sibling of ROSS-PCA's CPP, but the pruned object is the score matrix
Z = X_c·W rather than the loadings W. For interpretability we want each
component's scores concentrated on a few samples, so we prune the columns of
Z (equivalently, the rows of Zᵀ) cosine-preservingly — dropping the smallest
scores while keeping each component's sample-activation direction within angle
θ — then reconstruct with the trained orthonormal W as decoder:

    Z   = X_c W                       (X_c = X − mean)
    Z_p = cosine_prune(Zᵀ, θ).T       (per-column / per-component)
    X̂_c = Z_p Wᵀ                      (decode pruned scores; W stays fixed)
    VE(θ) = 1 − ‖X_c − X̂_c‖² / ‖X_c‖²

At θ=0 nothing is pruned, so the curve starts at the k-subspace VE and decays
as scores are zeroed. The operating point is chosen by the same dθ/ds
curvature elbow used by ROSS-PCA.

Output: JSON with the (Z-sparsity, reconstruction, VE) Pareto frontier.

Usage:
    python -m ross_proj.cpp_z \
        --model out/model.pkl --data X.npz --output-dir out/cpp_z/
"""

import argparse
import json
import os
import pickle
import time

import numpy as np

import jax
jax.config.update("jax_enable_x64", True)
import jax.numpy as jnp
from scipy import sparse


# ──────────────────────────────────────────────────────────────────────
# Cosine-preserving pruning (generic; operates on rows of a matrix)
# ──────────────────────────────────────────────────────────────────────

def cpp_precompute(M):
    """Precompute sort order and cumulative energy for each row of M.

    M is (n_rows, n_cols). For CPP-on-Z we pass Zᵀ, so each row is one
    component's scores across all samples.
    """
    n_rows, n_cols = M.shape
    sort_orders = np.zeros((n_rows, n_cols), dtype=np.intp)
    cum_removed = np.zeros((n_rows, n_cols), dtype=np.float64)
    energy_totals = np.zeros(n_rows, dtype=np.float64)

    for j in range(n_rows):
        energy_totals[j] = np.sum(M[j, :] ** 2)
        sort_orders[j, :] = np.argsort(np.abs(M[j, :]))  # smallest first
        sq_sorted = M[j, sort_orders[j, :]] ** 2
        cum_removed[j, :] = np.cumsum(sq_sorted)

    return sort_orders, cum_removed, energy_totals


def cpp_prune_fast(M, cos_t, sort_orders, cum_removed, energy_totals):
    """Prune each row of M to the smallest support within angle θ (cos_t)."""
    n_rows, n_cols = M.shape
    M_pruned = M.copy()
    n_kept_per_row = np.zeros(n_rows, dtype=int)

    energy_thresholds = cos_t ** 2 * energy_totals

    for j in range(n_rows):
        if energy_totals[j] == 0:
            M_pruned[j, :] = 0.0
            n_kept_per_row[j] = 0
            continue
        energy_remaining = energy_totals[j] - cum_removed[j, :]
        can_zero = np.searchsorted(-energy_remaining, -energy_thresholds[j])
        M_pruned[j, sort_orders[j, :can_zero]] = 0.0
        n_kept_per_row[j] = n_cols - can_zero

    norms_orig = np.sqrt(energy_totals)
    norms_pruned = np.sqrt(np.sum(M_pruned ** 2, axis=1))
    cosines = np.where(norms_orig > 0, norms_pruned / norms_orig, 1.0)
    sparsity_per_row = 1.0 - n_kept_per_row / n_cols

    stats = {
        "cos_t": float(cos_t),
        "sparsity_mean": float(sparsity_per_row.mean()),
        "sparsity_std": float(sparsity_per_row.std()),
        "n_kept_mean": float(n_kept_per_row.mean()),
        "n_kept_std": float(n_kept_per_row.std()),
        "n_kept_min": int(n_kept_per_row.min()),
        "n_kept_max": int(n_kept_per_row.max()),
        "cosine_min": float(cosines.min()),
        "cosine_mean": float(cosines.mean()),
        "overall_sparsity": float((M_pruned == 0).sum() / M_pruned.size),
    }
    return M_pruned, stats


# ──────────────────────────────────────────────────────────────────────
# Operating-point selection (max |dθ/ds| curvature elbow on the VE curve)
# ──────────────────────────────────────────────────────────────────────

def select_elbow(pareto):
    """Max distance-to-chord on the normalized (sparsity, VE) frontier."""
    if len(pareto) <= 2:
        return 0
    sparsity = np.array([p["sparsity"] for p in pareto])
    ve = np.array([p["var_explained"] for p in pareto])
    s_min, s_max = sparsity.min(), sparsity.max()
    v_min, v_max = ve.min(), ve.max()
    if s_max - s_min < 1e-12 or v_max - v_min < 1e-12:
        return 0
    s_norm = (sparsity - s_min) / (s_max - s_min)
    v_norm = (ve - v_min) / (v_max - v_min)
    x0, y0 = s_norm[0], v_norm[0]
    x1, y1 = s_norm[-1], v_norm[-1]
    dx, dy = x1 - x0, y1 - y0
    line_len = np.sqrt(dx**2 + dy**2)
    if line_len < 1e-12:
        return 0
    dist = np.abs(dy * s_norm - dx * v_norm + x1 * y0 - y1 * x0) / line_len
    return int(np.argmax(dist))


def select_dtheta_ds(pareto):
    """Select operating point via max |dθ/ds| on the normalized VE curve."""
    from scipy.signal import savgol_filter

    if len(pareto) < 7:
        return select_elbow(pareto)

    sparsity = np.array([p["sparsity"] for p in pareto])
    ve = np.array([p["var_explained"] for p in pareto])

    mask = np.ones(len(sparsity), dtype=bool)
    for i in range(1, len(sparsity)):
        if abs(sparsity[i] - sparsity[i-1]) < 1e-15 and abs(ve[i] - ve[i-1]) < 1e-15:
            mask[i] = False
    sp_c, ve_c = sparsity[mask], ve[mask]
    idx_map = np.where(mask)[0]
    if len(sp_c) < 7:
        return select_elbow(pareto)

    sp_min, sp_max = sp_c.min(), sp_c.max()
    ve_min, ve_max = ve_c.min(), ve_c.max()
    if sp_max - sp_min < 1e-15 or ve_max - ve_min < 1e-15:
        return select_elbow(pareto)
    sn = (sp_c - sp_min) / (sp_max - sp_min)
    vn = (ve_c - ve_min) / (ve_max - ve_min)

    sp_mask = np.ones(len(sn), dtype=bool)
    for i in range(1, len(sn)):
        if abs(sn[i] - sn[i-1]) < 1e-15:
            sp_mask[i] = False
    sn2, vn2 = sn[sp_mask], vn[sp_mask]
    sp_idx_map = np.arange(len(sn))[sp_mask]
    if len(sn2) < 7:
        return select_elbow(pareto)

    win = min(len(sn2) // 3, 21)
    if win % 2 == 0:
        win -= 1
    win = max(win, 5)
    if win > len(sn2):
        win = len(sn2) if len(sn2) % 2 == 1 else len(sn2) - 1

    vn_smooth = savgol_filter(vn2, window_length=win, polyorder=3)
    d1 = np.gradient(vn_smooth, sn2)
    d2 = np.gradient(d1, sn2)
    dtheta_ds = d2 / (1.0 + d1**2)
    margin = max(2, len(dtheta_ds) // 20)
    dtheta_ds[:margin] = 0
    dtheta_ds[-margin:] = 0
    best_sp_idx = int(np.argmin(dtheta_ds))
    return int(idx_map[sp_idx_map[best_sp_idx]])


# ──────────────────────────────────────────────────────────────────────
# Model / data loading — moved to ross_proj.io; re-exported here for the CLI
# and back-compat (e.g. `from ross_proj.cpp_z import load_model`).
# ──────────────────────────────────────────────────────────────────────

from .io import load_data, load_model  # noqa: F401,E402


def _get_batch(X, start, end):
    """Return rows [start:end) of X as a dense float array."""
    batch = X[start:end]
    if sparse.issparse(batch):
        batch = batch.toarray()
    return np.asarray(batch)


# ──────────────────────────────────────────────────────────────────────
# Scores + batched reconstruction (JAX)
# ──────────────────────────────────────────────────────────────────────

@jax.jit
def _batch_scores(X_batch, mean, W):
    return jnp.dot(X_batch - mean, W)                 # (b, k)


@jax.jit
def _batch_ss_data(X_batch, mean):
    Xc = X_batch - mean
    return jnp.sum(Xc * Xc)


@jax.jit
def _batch_ss_resid(X_batch, mean, Z_p_batch, components):
    Xc = X_batch - mean
    recon = jnp.dot(Z_p_batch, components)            # (b, k)·(k, p) = (b, p)
    diff = Xc - recon
    return jnp.sum(diff * diff)


def compute_scores(X, mean_j, W_j, n, batch_size, dtype):
    """Z = (X − mean) @ W, computed in row batches. Returns (n, k) host array."""
    k = W_j.shape[1]
    Z = np.zeros((n, k), dtype=np.float64)
    for s in range(0, n, batch_size):
        e = min(s + batch_size, n)
        Xb = jax.device_put(jnp.asarray(_get_batch(X, s, e), dtype=dtype))
        Z[s:e] = np.asarray(_batch_scores(Xb, mean_j, W_j), dtype=np.float64)
    return Z


# ──────────────────────────────────────────────────────────────────────
# Sweep
# ──────────────────────────────────────────────────────────────────────

def run_sweep(X, components, mean, thetas_deg, batch_size=20000, precision=32,
              verbose=True):
    """Cosine-prune Z over the θ sweep; return Pareto list + chosen index.

    components: (k, p) = Wᵀ (sklearn convention). mean: (p,).
    """
    dtype = jnp.float32 if precision == 32 else jnp.float64
    n = X.shape[0]
    components = np.asarray(components, dtype=np.float64)
    W = components.T                                   # (p, k)
    k, p = components.shape

    mean_j = jax.device_put(jnp.asarray(mean, dtype=dtype))
    W_j = jax.device_put(jnp.asarray(W, dtype=dtype))
    comp_j = jax.device_put(jnp.asarray(components, dtype=dtype))

    # Scores and ‖X_c‖² (one pass each).
    Z = compute_scores(X, mean_j, W_j, n, batch_size, dtype)   # (n, k)
    ss_data = 0.0
    for s in range(0, n, batch_size):
        e = min(s + batch_size, n)
        Xb = jax.device_put(jnp.asarray(_get_batch(X, s, e), dtype=dtype))
        ss_data += float(_batch_ss_data(Xb, mean_j))
    if ss_data <= 0:
        raise ValueError("‖X_c‖² is zero — is the data already centered to 0?")

    # Precompute cosine pruning on Zᵀ (each row = one component's scores).
    ZT = np.ascontiguousarray(Z.T)                     # (k, n)
    sort_orders, cum_removed, energy_totals = cpp_precompute(ZT)

    pareto = []
    for theta in thetas_deg:
        cos_t = float(np.cos(np.deg2rad(theta)))
        ZT_p, stats = cpp_prune_fast(ZT, cos_t, sort_orders, cum_removed,
                                     energy_totals)
        Z_p = np.ascontiguousarray(ZT_p.T)             # (n, k)

        ss_resid = 0.0
        for s in range(0, n, batch_size):
            e = min(s + batch_size, n)
            Xb = jax.device_put(jnp.asarray(_get_batch(X, s, e), dtype=dtype))
            Zb = jax.device_put(jnp.asarray(Z_p[s:e], dtype=dtype))
            ss_resid += float(_batch_ss_resid(Xb, mean_j, Zb, comp_j))

        rel_recon = float(np.sqrt(ss_resid / ss_data))
        ve = float(1.0 - ss_resid / ss_data)
        entry = {
            "theta_deg": float(theta),
            "sparsity": stats["overall_sparsity"],
            "relative_recon_loss": rel_recon,
            "var_explained": ve,
            "n_kept_mean": stats["n_kept_mean"],
            "n_kept_std": stats["n_kept_std"],
            "cosine_min": stats["cosine_min"],
        }
        pareto.append(entry)
        if verbose:
            print(f"  θ={theta:6.2f}° | Z-sparsity {entry['sparsity']:7.1%} | "
                  f"rel_recon {rel_recon:.6f} | VE {ve:7.4%} | "
                  f"kept/comp {stats['n_kept_mean']:.0f}", flush=True)

    best_idx = select_dtheta_ds(pareto) if pareto else -1
    return pareto, best_idx


# ──────────────────────────────────────────────────────────────────────
# Reporting: participation ratio + prune-at-operating-point
# ──────────────────────────────────────────────────────────────────────

def participation_ratio(Z):
    """Per-component participation ratio (Σz²)²/Σz⁴ over the columns of Z.

    The "effective number of items" carrying each component: L equal-magnitude
    entries → L; one dominant entry → 1; all equal → n. Computed on the dense
    scores, so it measures the concentration the fit produced.
    """
    Z = np.asarray(Z, dtype=np.float64)
    z2 = (Z ** 2).sum(0)
    z4 = (Z ** 4).sum(0)
    return z2 ** 2 / (z4 + 1e-300)


def prune_and_report(X, components, mean, theta="auto", thetas_deg=None,
                     labels=None, top_n=12, batch_size=20000, precision=32):
    """Cosine-prune the scores at an operating point and report (PruneResult).

    Sweeps θ (for the operating-point selector and the Pareto curve), prunes the
    scores Z at the chosen θ ('auto' = the dθ/ds point, or a float in degrees),
    and returns the joint VE there, per-component participation ratios on the
    dense scores, and — if `labels` is given — the top items per component
    (signed cosine weights = score / original column norm).
    """
    from .results import PruneResult
    components = np.asarray(components, dtype=np.float64)        # (k, p)
    W = components.T                                            # (p, k)
    k, p = components.shape
    n = X.shape[0]
    if thetas_deg is None:
        thetas_deg = np.linspace(0.0, 20.0, 81)

    pareto, best_idx = run_sweep(X, components, mean, thetas_deg,
                                 batch_size=batch_size, precision=precision,
                                 verbose=False)
    elbow_idx = select_elbow(pareto)
    op = pareto[best_idx] if best_idx >= 0 else pareto[0]
    elbow = pareto[elbow_idx]
    ve0 = pareto[0]["var_explained"]
    theta_star = float(op["theta_deg"]) if theta == "auto" else float(theta)

    # Dense scores Z and ‖X_c‖² (same JAX helpers as run_sweep).
    dtype = jnp.float32 if precision == 32 else jnp.float64
    mean_j = jax.device_put(jnp.asarray(mean, dtype=dtype))
    W_j = jax.device_put(jnp.asarray(W, dtype=dtype))
    comp_j = jax.device_put(jnp.asarray(components, dtype=dtype))
    Z = compute_scores(X, mean_j, W_j, n, batch_size, dtype)    # (n, k)
    ss_data = 0.0
    for s in range(0, n, batch_size):
        e = min(s + batch_size, n)
        Xb = jax.device_put(jnp.asarray(_get_batch(X, s, e), dtype=dtype))
        ss_data += float(_batch_ss_data(Xb, mean_j))

    # Prune Z at theta_star (cosine-preserving on Zᵀ).
    ZT = np.ascontiguousarray(Z.T)
    so, cr, et = cpp_precompute(ZT)
    ZT_p, stats = cpp_prune_fast(ZT, float(np.cos(np.deg2rad(theta_star))),
                                 so, cr, et)
    Z_p = np.ascontiguousarray(ZT_p.T)                          # (n, k)

    # Joint VE at theta_star (reconstruct Z_p Wᵀ), consistent with run_sweep.
    ss_resid = 0.0
    for s in range(0, n, batch_size):
        e = min(s + batch_size, n)
        Xb = jax.device_put(jnp.asarray(_get_batch(X, s, e), dtype=dtype))
        Zb = jax.device_put(jnp.asarray(Z_p[s:e], dtype=dtype))
        ss_resid += float(_batch_ss_resid(Xb, mean_j, Zb, comp_j))
    ve = float(1.0 - ss_resid / ss_data)

    pr = participation_ratio(Z)

    top_items = None
    if labels is not None:
        labels = np.asarray(labels).astype(str)
        z_norms = np.sqrt(np.maximum(et, 1e-300))               # original column norms
        top_items = []
        for j in range(k):
            col = Z_p[:, j]
            nz = np.flatnonzero(col)
            order = nz[np.argsort(-np.abs(col[nz]))][:top_n]
            top_items.append([(labels[i], float(col[i] / z_norms[j])) for i in order])

    return PruneResult(
        theta_deg=theta_star, sparsity=float(stats["overall_sparsity"]),
        var_explained=ve, ve_subspace=float(ve0), Z_pruned=Z_p,
        participation_ratio=pr, pareto=pareto, operating_point=op, elbow=elbow,
        top_items=top_items)


# ──────────────────────────────────────────────────────────────────────
# CLI
# ──────────────────────────────────────────────────────────────────────

def parse_args():
    ap = argparse.ArgumentParser(
        description="Cosine-Preserving Pruning of projections Z (scores).")
    ap.add_argument("--model", required=True, help="Fitted model .pkl")
    ap.add_argument("--data", required=True, help=".npz (key 'X') or .h5ad")
    ap.add_argument("--output-dir", required=True)
    ap.add_argument("--theta-max", type=float, default=10.0,
                    help="Max pruning angle in degrees")
    ap.add_argument("--theta-steps", type=int, default=100)
    ap.add_argument("--batch-size", type=int, default=20000)
    ap.add_argument("--precision", type=int, default=32, choices=[32, 64])
    return ap.parse_args()


def main():
    args = parse_args()
    os.makedirs(args.output_dir, exist_ok=True)

    print(f"Loading model: {args.model}")
    model = load_model(args.model)
    components = np.asarray(model.components_)         # (k, p)
    mean = np.asarray(model.mean_)                     # (p,)
    k, p = components.shape
    print(f"  components {components.shape}, mean {mean.shape}")

    print(f"Loading data: {args.data}")
    t0 = time.time()
    X = load_data(args.data)
    print(f"  {X.shape[0]:,} × {X.shape[1]:,} [{time.time()-t0:.1f}s]")
    assert X.shape[1] == p, f"data has {X.shape[1]} features, model has {p}"

    thetas = np.linspace(0.0, args.theta_max, args.theta_steps)
    print(f"Sweeping {len(thetas)} angles in [0, {args.theta_max}]°:")
    pareto, best_idx = run_sweep(
        X, components, mean, thetas,
        batch_size=args.batch_size, precision=args.precision)

    results = {
        "n_samples": int(X.shape[0]),
        "n_features": int(p),
        "n_components": int(k),
        "theta_max": float(args.theta_max),
        "theta_steps": int(args.theta_steps),
        "pareto": pareto,
        "best_operating_point": (
            {"index": int(best_idx), **pareto[best_idx]} if best_idx >= 0 else None),
    }
    out_json = os.path.join(args.output_dir, "cpp_z.json")
    with open(out_json, "w") as f:
        json.dump(results, f, indent=2, default=str)
    print(f"\nResults -> {out_json}")
    if best_idx >= 0:
        bp = pareto[best_idx]
        print(f"Selected: θ={bp['theta_deg']:.2f}° | "
              f"Z-sparsity {bp['sparsity']:.1%} | VE {bp['var_explained']:.4%}")


if __name__ == "__main__":
    main()
```
