## Supplemental code for "Scoring respondents on a semantic frame of reference identifies a group that is burdened across many questionnaires and reaches care less easily": defining_sets.py

```
"""Defining sets: which items each component (column of Z) focuses on.

.. warning::

   **This is the ONE-SHOT selection, and it is NOT the object the manuscript reports.**
   The published cores are the *participation-ratio fixed point*: apply the selection
   below, recompute PR on the survivors, and re-select, iterating to convergence. The
   set stops shrinking because PR(S) <= |S| for any set S. One pass and the fixed point
   differ whenever the first selection changes the column's PR, which is the usual case.

   The reference implementation is ``fixed_point_core`` in
   ``examples/allofus/pool_fixedpoint_cores.py``, which writes the shipped
   ``outputs_pool/fixedpoint_cores/<encoder>_cores.csv`` that every downstream analysis
   reads. Reproducing the paper from this module's one-shot output will give cores of a
   different size. This function remains as the library's one-pass primitive, and for
   ``participation_ratio``, which the fixed-point iteration itself calls. Its former callers
   were archived in the 2026-08-31 cleanup, so nothing in the live tree depends on it.

Pure **interpretation** — no reconstruction, no pruning sweep. For each component
`j` (a column of the scores `Z`), the participation ratio

    PR_j = (Σ_i Z_ij²)² / Σ_i Z_ij⁴

is its *effective number of items*. The **defining set** of component `j` is the
`round(PR_j)` items with the largest `|score|`, and we report the fraction of the
column's score energy those items carry — the justification for the selection:

    energy_retained_j = Σ_{i ∈ set} Z_ij²  /  Σ_i Z_ij² .

This is the same "drop the smallest |score| first" ordering as Cosine-Preserving
Pruning (`ross_proj.cpp_z`), but with a **parameter-free, per-component stopping
count** (`round(PR_j)`) instead of a globally swept angle θ. Equivalently, the
set retains `energy_retained` of the column, i.e. it matches CPP at the implied
angle `θ = arccos(√energy_retained)`. Self-contained (no JAX).
"""
from dataclasses import dataclass
from typing import List, Optional

import numpy as np


def participation_ratio(Z):
    """Per-component participation ratio (Σz²)²/Σz⁴ over the columns of `Z`."""
    Z = np.asarray(Z, dtype=np.float64)
    z2 = (Z ** 2).sum(0)
    z4 = (Z ** 4).sum(0)
    return z2 ** 2 / (z4 + 1e-300)


@dataclass
class DefiningSets:
    pr: np.ndarray                 # (k,)   participation ratio per component
    counts: np.ndarray            # (k,)   round(PR_j), clipped to [1, n]
    membership: np.ndarray        # (n, k) bool: item i is in component j's set
    energy_retained: np.ndarray   # (k,)   Σ_set z² / Σ z²  (the justification)
    components: List[dict]        # per component: pr, count, energy_retained, items


def defining_sets(Z, labels=None):
    """Per-component defining set from the dense scores `Z` (n × k).

    ONE PASS, so this is NOT the manuscript's core. The published object iterates
    {select, recompute PR} to convergence -- see this module's docstring and
    ``fixed_point_core`` in ``examples/allofus/pool_fixedpoint_cores.py``.

    Returns a `DefiningSets`. Each `components[j]` is a dict with `component`,
    `pr`, `count`, `energy_retained`, and `items` — the selected items ordered by
    descending `|score|`, each `{index, label, score}` (signed weight).
    """
    Z = np.asarray(Z, dtype=np.float64)
    n, k = Z.shape
    z2 = Z ** 2
    col_energy = z2.sum(0)
    pr = col_energy ** 2 / ((z2 ** 2).sum(0) + 1e-300)
    counts = np.clip(np.rint(pr).astype(int), 1, n)
    if labels is not None:
        labels = np.asarray(labels).astype(str)

    membership = np.zeros((n, k), dtype=bool)
    energy = np.zeros(k, dtype=np.float64)
    comps = []
    for j in range(k):
        order = np.argsort(-np.abs(Z[:, j]))          # largest |score| first
        sel = order[:counts[j]]
        membership[sel, j] = True
        energy[j] = z2[sel, j].sum() / (col_energy[j] + 1e-300)
        items = [{"index": int(i),
                  "label": (labels[i] if labels is not None else str(int(i))),
                  "score": float(Z[i, j])} for i in sel]
        comps.append({"component": int(j), "pr": float(pr[j]),
                      "count": int(counts[j]), "energy_retained": float(energy[j]),
                      "items": items})
    return DefiningSets(pr=pr, counts=counts, membership=membership,
                        energy_retained=energy, components=comps)


def overlap_stats(membership):
    """How much the defining sets overlap across components (partition check).

    Defining sets are *not* a clean item→component partition: an item can define
    several components. Returns counts/fractions characterizing that overlap.
    """
    membership = np.asarray(membership, dtype=bool)
    n, k = membership.shape
    comps_per_item = membership.sum(1)                 # (n,)
    used = comps_per_item > 0
    nu = int(used.sum())
    return {
        "n_items": int(n), "k": int(k), "items_used": nu,
        "mean_set_size": float(membership.sum(0).mean()),
        "frac_items_multicomponent": float((comps_per_item > 1).sum() / max(nu, 1)),
        "mean_components_per_item": float(comps_per_item[used].mean()) if nu else 0.0,
        "max_components_per_item": int(comps_per_item.max()) if n else 0,
    }
```
