## Supplemental code for "Scoring respondents on a semantic frame of reference identifies a group that is burdened across many questionnaires and reaches care less easily": fit.py

```
#!/usr/bin/env python
"""fit.py — core ROSS-Proj fit.

The first pipeline stage: given data `X` and the sparsity weights, fit the
estimator and return everything downstream stages need (`mean`, loadings `W`,
scores `Z`). A thin functional wrapper around :class:`SparseProjectionPCA` that
returns a :class:`FitResult`.

Usage (library):
    from ross_proj import fit_projection
    res = fit_projection(X, n_components=k, lambda_score=1.0, lambda_ortho=0.1)
    res.mean, res.W, res.Z      # X_mean, loadings (k×p), scores (n×k)

Usage (CLI):
    python -m ross_proj.fit --data X.npz --n-components 39 \
        --lambda-score 1.0 --lambda-ortho 0.1 --output-dir out/
"""
import argparse
import time

import numpy as np

from .sparse_projection import SparseProjectionPCA
from .results import FitResult
from .io import load_data, save_fit


def fit_projection(X, n_components, lambda_score=1.0, lambda_ortho=0.1, **kwargs):
    """Fit ROSS-Proj on `X` (n_samples × n_features) and return a FitResult.

    Pass UNCENTERED `X`; the estimator centers internally. Extra keyword args are
    forwarded to :class:`SparseProjectionPCA` (e.g. ``batch_size``,
    ``total_steps``, ``subsample_fraction``, ``random_state``, ``verbose``).
    """
    model = SparseProjectionPCA(
        n_components=n_components, lambda_score=lambda_score,
        lambda_ortho=lambda_ortho, **kwargs).fit(X)
    Z = model.transform(X)
    return FitResult(
        mean=np.asarray(model.mean_), W=np.asarray(model.components_), Z=Z,
        n_components=int(n_components), lambda_score=float(lambda_score),
        lambda_ortho=float(lambda_ortho),
        pca_recon_loss=float(model.pca_recon_loss_), model=model)


def parse_args():
    ap = argparse.ArgumentParser(
        description="Core ROSS-Proj fit (X -> mean, loadings W, scores Z).")
    ap.add_argument("--data", required=True, help=".npz (key 'X') or .h5ad")
    ap.add_argument("--output-dir", required=True)
    ap.add_argument("--n-components", type=int, required=True)
    ap.add_argument("--lambda-score", type=float, default=1.0)
    ap.add_argument("--lambda-ortho", type=float, default=0.1)
    ap.add_argument("--batch-size", type=int, default=None,
                    help="minibatch size (default: all n samples — full batch)")
    ap.add_argument("--total-steps", type=int, default=5000)
    ap.add_argument("--subsample-fraction", type=float, default=None,
                    help="fraction of rows for the robust scale (default: 1.0 if "
                         "n<10000 else estimator default)")
    ap.add_argument("--random-state", type=int, default=42)
    return ap.parse_args()


def main():
    args = parse_args()
    X = load_data(args.data)
    X = X.toarray() if hasattr(X, "toarray") else np.asarray(X, dtype=np.float64)
    n, p = X.shape
    kw = dict(total_steps=args.total_steps, random_state=args.random_state,
              verbose=False, log_interval=0)
    kw["batch_size"] = args.batch_size or n
    kw["subsample_fraction"] = (args.subsample_fraction
                                if args.subsample_fraction is not None
                                else (1.0 if n < 10000 else 0.05))
    t0 = time.time()
    res = fit_projection(X, args.n_components, args.lambda_score,
                         args.lambda_ortho, **kw)
    save_fit(res, args.output_dir)
    print(f"fit {n}×{p} k={args.n_components} λs={args.lambda_score} "
          f"λo={args.lambda_ortho} [{time.time()-t0:.1f}s] -> {args.output_dir}")


if __name__ == "__main__":
    main()
```
