## Supplemental code for "Scoring respondents on a semantic frame of reference identifies a group that is burdened across many questionnaires and reaches care less easily": io.py

```
"""I/O helpers: data loading and (de)serialization of pipeline results.

Kept separate so the compute modules stay pure. Data loading is shared by every
stage's CLI; the save/load helpers let the per-stage example scripts hand
results to one another (fit -> prune).
"""
import json
import os
import pickle

import numpy as np
from scipy import sparse


# ──────────────────────────────────────────────────────────────────────
# Data loading
# ──────────────────────────────────────────────────────────────────────

def load_data(path):
    """Load `.npz` (scipy.sparse, or dense under key 'X' / first key) or `.h5ad`.

    Returns a dense array or a scipy.sparse matrix (n_samples × n_features).
    """
    ext = os.path.splitext(path)[1].lower()
    if ext == ".npz":
        try:
            return sparse.load_npz(path)
        except Exception:
            d = np.load(path, allow_pickle=True)
            key = "X" if "X" in d.files else d.files[0]
            return np.asarray(d[key], dtype=np.float64)
    elif ext == ".h5ad":
        import scanpy as sc
        return sc.read_h5ad(path).X
    raise ValueError(f"Unsupported data format: {ext}")


# ──────────────────────────────────────────────────────────────────────
# Lenient model loading (tolerant of an un-importable model class)
# ──────────────────────────────────────────────────────────────────────

class _AttrBag:
    """Stand-in for un-importable model classes; only needs attributes."""
    def __setstate__(self, state):
        self.__dict__.update(state)


class _LenientUnpickler(pickle.Unpickler):
    def find_class(self, module, name):
        try:
            return super().find_class(module, name)
        except (ImportError, AttributeError):
            return _AttrBag


def load_model(model_path):
    """Load a fitted model; needs only `.components_` (k, p) and `.mean_` (p,)."""
    with open(model_path, "rb") as f:
        return _LenientUnpickler(f).load()


# ──────────────────────────────────────────────────────────────────────
# Result (de)serialization
# ──────────────────────────────────────────────────────────────────────

def save_fit(result, output_dir):
    """Persist a FitResult: loadings/scores/mean as .npy, meta .json, model .pkl."""
    os.makedirs(output_dir, exist_ok=True)
    np.save(os.path.join(output_dir, "loadings.npy"), result.W)
    np.save(os.path.join(output_dir, "scores.npy"), result.Z)
    np.save(os.path.join(output_dir, "mean.npy"), result.mean)
    meta = {
        "n_components": int(result.n_components),
        "lambda_score": float(result.lambda_score),
        "lambda_ortho": float(result.lambda_ortho),
        "pca_recon_loss": float(result.pca_recon_loss),
    }
    with open(os.path.join(output_dir, "fit.json"), "w") as f:
        json.dump(meta, f, indent=2)
    if result.model is not None:
        with open(os.path.join(output_dir, "model.pkl"), "wb") as f:
            pickle.dump(result.model, f)


def load_fit(output_dir):
    """Reload a saved fit into a FitResult (model via lenient unpickler if present)."""
    from .results import FitResult
    meta = json.load(open(os.path.join(output_dir, "fit.json")))
    model_path = os.path.join(output_dir, "model.pkl")
    model = load_model(model_path) if os.path.exists(model_path) else None
    return FitResult(
        mean=np.load(os.path.join(output_dir, "mean.npy")),
        W=np.load(os.path.join(output_dir, "loadings.npy")),
        Z=np.load(os.path.join(output_dir, "scores.npy")),
        n_components=meta["n_components"], lambda_score=meta["lambda_score"],
        lambda_ortho=meta["lambda_ortho"], pca_recon_loss=meta["pca_recon_loss"],
        model=model,
    )


def save_kresult(result, output_dir):
    """Persist a KResult as parallel_analysis.json."""
    os.makedirs(output_dir, exist_ok=True)
    out = {
        "k": int(result.k), "k_cross": int(result.k_cross),
        "k_count": int(result.k_count), "k_mean": int(result.k_mean),
        "d80": int(result.d80), "d90": int(result.d90),
        "n_perm": int(result.n_perm), "percentile": float(result.percentile),
        "real_eigs": np.asarray(result.real_eigs).tolist(),
        "null_percentile": np.asarray(result.null_percentile).tolist(),
        "null_mean": np.asarray(result.null_mean).tolist(),
    }
    with open(os.path.join(output_dir, "parallel_analysis.json"), "w") as f:
        json.dump(out, f, indent=2)


def save_prune(result, output_dir):
    """Persist a PruneResult: pruned scores .npy + sweep/operating-point .json."""
    os.makedirs(output_dir, exist_ok=True)
    np.save(os.path.join(output_dir, "scores_pruned.npy"), result.Z_pruned)
    out = {
        "theta_deg": float(result.theta_deg), "sparsity": float(result.sparsity),
        "var_explained": float(result.var_explained),
        "ve_subspace": float(result.ve_subspace),
        "participation_ratio": np.asarray(result.participation_ratio).tolist(),
        "operating_point": result.operating_point, "elbow": result.elbow,
        "pareto": result.pareto,
    }
    with open(os.path.join(output_dir, "cpp_report.json"), "w") as f:
        json.dump(out, f, indent=2, default=str)
```
