## Supplemental code for "Scoring respondents on a semantic frame of reference identifies a group that is burdened across many questionnaires and reaches care less easily": parallel_analysis.py

```
#!/usr/bin/env python
"""parallel_analysis.py — choose the number of components k (Parallel Analysis).

Stage that fixes the model order. Horn's Parallel Analysis (1965), permutation
variant (Buja & Eyuboglu 1992): compare the data's covariance eigenvalues to a
null built by shuffling each feature independently across samples (same
per-feature marginals, no cross-feature structure), and keep only the components
whose eigenvalue beats the null.

This is **upstream of and independent of** ``lambda_score``: it fixes how many
principal directions are real, and ROSS-Proj then finds the sparse rotation of
that k-dimensional subspace. The null needs only the **PCA eigenvalue spectrum**
of each permutation — not a full ROSS-Proj fit (the subspace dimension is a
covariance property the sparse fit only rotates within), so the permutations are
cheap SVDs and the core fit runs once, at the selected k.

What the eigenvalues are. With X_c = Z Wᵀ, each component's eigenvalue is its
variance ‖Z_j‖² (loadings W_j are unit-norm directions). So PA thresholds the
score variances; k is the count of (W_j, Z_j) components above the noise floor.

Usage (library):
    from ross_proj import select_k
    kres = select_k(X, n_perm=200)          # KResult; kres.k is the recommended k
    # or select-then-fit in one call:
    from ross_proj.parallel_analysis import fit_with_selected_k
    kres, fit = fit_with_selected_k(X, lambda_score=1.0, lambda_ortho=0.1)

Usage (CLI):
    python -m ross_proj.parallel_analysis --data X.npz --output-dir out/ \
        --n-perm 200 --percentile 95
"""
import argparse
import os
import time

import numpy as np

from .io import load_data, save_kresult
from .results import KResult


# ──────────────────────────────────────────────────────────────────────
# Core
# ──────────────────────────────────────────────────────────────────────

def _eigenvalues(Xc):
    """Descending covariance eigenvalues of centered Xc (∝ squared singulars)."""
    s = np.linalg.svd(np.asarray(Xc, dtype=np.float64), compute_uv=False)
    return s ** 2


def _permute_columns(X, rng):
    """Independently shuffle each feature (column) across samples (rows)."""
    idx = np.argsort(rng.random(X.shape), axis=0)
    return np.take_along_axis(X, idx, axis=0)


def select_k(X, n_perm=200, percentile=95.0, seed=0, center=True):
    """Permutation parallel analysis -> :class:`KResult`.

    `X` is (n_samples, n_features), in the orientation you fit (for ROSS-Proj on
    item embeddings: items as samples, embedding dims as features). Returns the
    real/null spectra and the recommended k (first-crossing) plus alternatives.
    """
    X = np.asarray(X.toarray() if hasattr(X, "toarray") else X, dtype=np.float64)
    rng = np.random.default_rng(seed)
    Xc = X - X.mean(0) if center else X
    real = _eigenvalues(Xc)
    r = real.size

    null = np.empty((n_perm, r), dtype=np.float64)
    for b in range(n_perm):
        Xp = _permute_columns(X, rng)
        null[b] = _eigenvalues(Xp - Xp.mean(0) if center else Xp)

    null_pct = np.percentile(null, percentile, axis=0)
    null_mean = null.mean(0)

    below = np.where(real <= null_pct)[0]
    k_cross = int(below[0]) if below.size else r
    k_count = int(np.sum(real > null_pct))
    below_mean = np.where(real <= null_mean)[0]
    k_mean = int(below_mean[0]) if below_mean.size else r

    cum = np.cumsum(real) / real.sum()
    d80 = int(np.searchsorted(cum, 0.80) + 1)
    d90 = int(np.searchsorted(cum, 0.90) + 1)

    return KResult(
        k=k_cross, k_cross=k_cross, k_count=k_count, k_mean=k_mean,
        real_eigs=real, null_percentile=null_pct, null_mean=null_mean,
        d80=d80, d90=d90, n_perm=int(n_perm), percentile=float(percentile))


# Back-compat alias (dict-free): same as select_k.
parallel_analysis = select_k


def fit_with_selected_k(X, lambda_score=1.0, lambda_ortho=0.1, n_perm=200,
                        percentile=95.0, seed=0, **fit_kwargs):
    """Select k by parallel analysis, then run the core fit once at that k.

    Returns ``(KResult, FitResult)`` — the "PA returns mean/Z/W/null/selected-k"
    entry point. ``fit_kwargs`` are forwarded to :func:`ross_proj.fit.fit_projection`.
    """
    from .fit import fit_projection
    kres = select_k(X, n_perm=n_perm, percentile=percentile, seed=seed)
    fit = fit_projection(X, n_components=kres.k, lambda_score=lambda_score,
                         lambda_ortho=lambda_ortho, **fit_kwargs)
    return kres, fit


# ──────────────────────────────────────────────────────────────────────
# Optional scree plot (lazy matplotlib import; skipped if unavailable)
# ──────────────────────────────────────────────────────────────────────

def save_scree(kres, path, title=None, max_components=90):
    """Write a real-vs-null scree plot; no-op (returns False) if no matplotlib."""
    try:
        import matplotlib
        matplotlib.use("Agg")
        import matplotlib.pyplot as plt
    except Exception:
        return False
    real, null_p = kres.real_eigs, kres.null_percentile
    m = int(min(max_components, real.size))
    x = np.arange(1, m + 1)
    fig, ax = plt.subplots(figsize=(6.5, 4))
    ax.plot(x, real[:m], "-", color="#cc3333", lw=1.8, label="real")
    ax.plot(x, null_p[:m], "--", color="#3366cc", lw=1.3,
            label="null p%g" % kres.percentile)
    ax.axvline(kres.k, color="#222", ls=":", lw=1.2, label="k=%d" % kres.k)
    ax.set_yscale("log")
    ax.set_xlabel("component"); ax.set_ylabel("eigenvalue")
    ax.set_title(title or "Permutation parallel analysis  (k=%d)" % kres.k)
    ax.legend(fontsize=8); ax.grid(alpha=0.3, which="both")
    fig.tight_layout(); fig.savefig(path, dpi=130); plt.close(fig)
    return True


# ──────────────────────────────────────────────────────────────────────
# CLI
# ──────────────────────────────────────────────────────────────────────

def parse_args():
    ap = argparse.ArgumentParser(
        description="Parallel Analysis (permutation) for choosing k.")
    ap.add_argument("--data", required=True, help=".npz (key 'X') or .h5ad")
    ap.add_argument("--output-dir", required=True)
    ap.add_argument("--n-perm", type=int, default=200)
    ap.add_argument("--percentile", type=float, default=95.0)
    ap.add_argument("--seed", type=int, default=0)
    ap.add_argument("--no-center", action="store_true",
                    help="do not mean-center features (default: center)")
    ap.add_argument("--no-plot", action="store_true")
    return ap.parse_args()


def main():
    args = parse_args()
    os.makedirs(args.output_dir, exist_ok=True)

    print(f"Loading data: {args.data}")
    t0 = time.time()
    X = load_data(args.data)
    print(f"  {X.shape[0]:,} samples × {X.shape[1]:,} features [{time.time()-t0:.1f}s]")

    print(f"Parallel analysis: {args.n_perm} permutations, p{args.percentile:g} null...")
    kres = select_k(X, n_perm=args.n_perm, percentile=args.percentile,
                    seed=args.seed, center=not args.no_center)
    print(f"  k (first-crossing) = {kres.k} | k_count = {kres.k_count} | "
          f"k_mean = {kres.k_mean} | d80 = {kres.d80} | d90 = {kres.d90}")

    save_kresult(kres, args.output_dir)
    print(f"Results -> {os.path.join(args.output_dir, 'parallel_analysis.json')}")
    if not args.no_plot:
        png = os.path.join(args.output_dir, "parallel_analysis.png")
        if save_scree(kres, png):
            print(f"Scree    -> {png}")


if __name__ == "__main__":
    main()
```
