## Supplemental code for "Scoring respondents on a semantic frame of reference identifies a group that is burdened across many questionnaires and reaches care less easily": results.py

```
"""Typed result containers passed between the ROSS-Proj pipeline stages.

The three modules each return one of these, so stages compose cleanly:

    select_k        (parallel_analysis) -> KResult
    fit_projection  (sparse_projection) -> FitResult
    prune_and_report(cpp_z)             -> PruneResult
"""
from dataclasses import dataclass
from typing import List, Optional

import numpy as np


@dataclass
class KResult:
    """Output of permutation parallel analysis (k-selection)."""
    k: int                            # recommended k (= k_cross)
    k_cross: int                      # first-crossing count (conservative)
    k_count: int                      # total ranks above the null percentile
    k_mean: int                       # ranks above the null mean (Horn's liberal criterion)
    real_eigs: np.ndarray             # (r,) real covariance eigenvalues, descending
    null_percentile: np.ndarray       # (r,) null eigenvalue at `percentile`, per rank
    null_mean: np.ndarray             # (r,) null eigenvalue mean, per rank
    d80: int                          # components for 80% cumulative variance (heuristic)
    d90: int                          # components for 90% cumulative variance (heuristic)
    n_perm: int
    percentile: float


@dataclass
class FitResult:
    """Output of the core ROSS-Proj fit."""
    mean: np.ndarray                  # (p,)   X_mean (per-feature)
    W: np.ndarray                     # (k, p) loadings (= components_)
    Z: np.ndarray                     # (n, k) scores  (= (X - mean) @ Wᵀ)
    n_components: int
    lambda_score: float
    lambda_ortho: float
    pca_recon_loss: float             # calibration constant (penalty scale)
    model: object = None              # fitted SparseProjectionPCA (for transform / pickle)


@dataclass
class PruneResult:
    """Output of cosine-preserving pruning of the scores + reporting."""
    theta_deg: float                  # operating-point angle used for the pruned scores
    sparsity: float                   # overall fraction of zeroed scores at theta
    var_explained: float              # joint VE at theta (1 - ‖X_c - Z_p Wᵀ‖²/‖X_c‖²)
    ve_subspace: float                # VE at θ=0 (the k-subspace ceiling)
    Z_pruned: np.ndarray              # (n, k) scores pruned at theta
    participation_ratio: np.ndarray   # (k,) per-component PR on the DENSE scores
    pareto: List[dict]                # full (sparsity, VE) sweep
    operating_point: dict             # dθ/ds-selected point
    elbow: dict                       # distance-to-chord elbow point
    top_items: Optional[list] = None  # per-component [(label, signed_weight), ...] if labels given
```
