## Supplemental code for "Scoring respondents on a semantic frame of reference identifies a group that is burdened across many questionnaires and reaches care less easily": sparse_projection.py

```
# ross_pca/sparse_pca.py
import jax
import jax.numpy as jnp
import optax
import numpy as np
from scipy import sparse
from sklearn.base import BaseEstimator, TransformerMixin
from sklearn.utils.extmath import randomized_svd
from tqdm import tqdm as _tqdm
from functools import partial


class _NoOpPbar:
    """Silent no-op replacement for tqdm progress bar."""
    def __init__(self, *a, **kw): pass
    def update(self, *a, **kw): pass
    def set_postfix(self, *a, **kw): pass
    def clear(self, *a, **kw): pass
    def close(self, *a, **kw): pass
    def __enter__(self): return self
    def __exit__(self, *a): pass
    def __iter__(self): return self
    def __next__(self): raise StopIteration


def _noop_tqdm(iterable=None, *a, **kw):
    """Silent replacement for tqdm that just passes through the iterable."""
    if iterable is not None:
        return iterable
    return _NoOpPbar()

# 1. ENABLE FLOAT64
jax.config.update("jax_enable_x64", True)

# Consistency constant: MAD * 1.4826 ≈ std for Gaussian data
_MAD_TO_STD = 1.4826

# -----------------------------------------------------------------------------
# 2. Math Helpers
# -----------------------------------------------------------------------------

def _safe_to_dense(batch):
    """Convert scipy.sparse to dense; pass numpy arrays through unchanged."""
    if sparse.issparse(batch):
        return batch.toarray()
    return batch


def log_sparsity_penalty(x, epsilon_sq):
    """Smooth log penalty for sparsity: log(1 + x²/ε²).
    
    A fully smooth, doubly-differentiable approximation to the classical
    log(|x| + ε) penalty (Candès, Wakin, Boyd 2008), adapted for
    gradient-based optimization.
    
    Behavior (controlled by ratio |x|/ε):
        |x| << ε: penalty ≈ x²/ε² (quadratic, strong push to zero)
        |x| >> ε: penalty ≈ 2·log(|x|/ε) (log, gentle on large entries)
        Transition around |x| ≈ ε.
    
    Gradient: 2x / (ε² + x²)
        |x| << ε: gradient ≈ 2x/ε² (linear, strong)
        |x| = ε:  gradient = 1/ε (peak pressure)
        |x| >> ε: gradient ≈ 2/x (decays, leaves large entries alone)
    
    The concavity naturally provides adaptive lasso behavior without
    explicit reweighting: small entries receive strong pressure while
    large entries are left alone.
    
    Args:
        x: Input array (entries of column-normalized direction matrix).
        epsilon_sq: ε² (squared threshold). Precomputed to avoid repeated
            squaring inside JIT-compiled functions.
    
    Returns:
        Element-wise log(1 + x²/ε²).
    """
    return jnp.log1p(x**2 / epsilon_sq)


def otsu_threshold(values):
    """Otsu's method: find threshold that maximizes between-class variance.
    
    Operates on a 1D array of non-negative values (e.g., |W̃| entries).
    Finds the optimal split point between two populations (near-zero
    entries to kill vs. signal entries to keep).
    
    For a bimodal distribution, this finds the valley between the modes.
    For a uniform distribution, this returns approximately the mean.
    
    Uses histogram-based approach with 256 bins for efficiency.
    
    Args:
        values: 1D numpy array of non-negative values.
    
    Returns:
        Optimal threshold (float64).
    """
    values = np.asarray(values, dtype=np.float64).ravel()
    
    n_bins = 256
    v_min, v_max = values.min(), values.max()
    if v_max - v_min < 1e-15:
        return float(v_max)
    
    counts, bin_edges = np.histogram(values, bins=n_bins)
    bin_centers = 0.5 * (bin_edges[:-1] + bin_edges[1:])
    
    total = counts.sum()
    probs = counts.astype(np.float64) / total
    
    omega = np.cumsum(probs)
    mu = np.cumsum(probs * bin_centers)
    mu_total = mu[-1]
    
    valid = (omega > 1e-10) & (omega < 1 - 1e-10)
    sigma_b_sq = np.zeros(n_bins, dtype=np.float64)
    sigma_b_sq[valid] = (mu_total * omega[valid] - mu[valid])**2 / \
                         (omega[valid] * (1.0 - omega[valid]))
    
    best_idx = np.argmax(sigma_b_sq)
    threshold = float(bin_centers[best_idx])
    
    return threshold


def cosine_annealing(current_step, total_steps):
    """Cosine annealing schedule: 0.0 → 1.0 over total_steps."""
    if current_step >= total_steps:
        return 1.0
    progress = current_step / total_steps
    return 0.5 * (1.0 - jnp.cos(jnp.pi * progress))


# -----------------------------------------------------------------------------
# 3. Data Preprocessing: Global Mean + Robust Scale (Single Pass)
# -----------------------------------------------------------------------------

def compute_global_mean_and_scale(input_matrix, chunk_size=10000,
                                  subsample_fraction=0.1, seed=None,
                                  verbose=True):
    """Compute global mean (full data) and robust variance (random subsample).
    
    Two-pass approach:
      Pass 1: Stream full data to accumulate global mean.
      Pass 2: Load a single random subsample (pre-selected indices) to compute
              per-feature MAD centered around the global mean.
    
    The subsample is loaded as one contiguous block (sorted indices for cache
    efficiency), avoiding the peak RAM spike from concatenating per-chunk lists.
    Maximum subsample RAM = n_sub × n_features × 8 bytes (float64).
    
    Returns:
        global_mean: (n_features,) array, float64
        sigma_sq_robust: scalar, (median nonzero MAD × 1.4826)²
        mad_per_feature: (n_features,) array, per-feature MAD values
    """
    _print = print if verbose else (lambda *a, **kw: None)
    _tbar = _tqdm if verbose else _noop_tqdm
    
    n_samples, n_features = input_matrix.shape
    rng = np.random.default_rng(seed)
    
    n_sub = max(2, int(n_samples * subsample_fraction))
    sub_idx = np.sort(rng.choice(n_samples, size=n_sub, replace=False))
    sub_size_gb = n_sub * n_features * 8 / 1e9
    
    _print(f"Computing global mean and robust scale ({n_samples:,} samples, "
           f"{n_features:,} features)...")
    _print(f"  Subsample: {n_sub:,} rows ({100*n_sub/n_samples:.1f}%), "
           f"~{sub_size_gb:.2f} GB float64", flush=True)
    
    # --- Pass 1: Global mean ---
    mean_acc = np.zeros(n_features, dtype=np.float64)
    for i in _tbar(range(0, n_samples, chunk_size), desc="Pass 1 (mean)"):
        end = min(i + chunk_size, n_samples)
        chunk = _safe_to_dense(input_matrix[i:end]).astype(np.float64)
        mean_acc += chunk.sum(axis=0)
    global_mean = mean_acc / n_samples
    del mean_acc
    
    # --- Pass 2: MAD from subsample, centered around global mean ---
    _print(f"  Pass 2: loading subsample for MAD...", flush=True)
    sub_chunks = []
    for i in _tbar(range(0, n_sub, chunk_size), desc="Pass 2 (MAD subsample)"):
        idx_chunk = sub_idx[i:i + chunk_size]
        sub_chunks.append(_safe_to_dense(input_matrix[idx_chunk]).astype(np.float64))
    subsample = np.concatenate(sub_chunks, axis=0)
    del sub_chunks
    
    n_subsample = subsample.shape[0]
    subsample -= global_mean
    np.abs(subsample, out=subsample)
    mad_per_feature = np.median(subsample, axis=0)
    del subsample
    
    nonzero_mads = mad_per_feature[mad_per_feature > 0]
    n_nonzero_mads = len(nonzero_mads)
    if n_nonzero_mads > 0:
        median_mad = float(np.median(nonzero_mads))
    else:
        median_mad = 1.0
        print("  WARNING: all per-feature MADs are zero, falling back to sigma_sq_robust=1.0")
    sigma_sq_robust = (median_mad * _MAD_TO_STD) ** 2
    
    _print(f"  Mean computed on {n_samples:,} samples.")
    _print(f"  MAD computed on {n_subsample:,} subsampled rows "
           f"({100 * n_subsample / n_samples:.1f}%).")
    _print(f"  Nonzero MAD features: {n_nonzero_mads}/{len(mad_per_feature)} "
           f"({100 * n_nonzero_mads / len(mad_per_feature):.1f}%)")
    _print(f"  σ²_robust = {sigma_sq_robust:.6f} "
           f"(median nonzero MAD = {median_mad:.6f})")
    
    return global_mean, sigma_sq_robust, mad_per_feature


# -----------------------------------------------------------------------------
# 4. Initialization
# -----------------------------------------------------------------------------

def _init_pca(input_matrix, global_mean, n_components, random_state,
              chunk_size=10000, verbose=True):
    """PCA initialization via randomized SVD (Halko et al. 2009).
    
    Operates on the full centered data. randomized_svd handles large matrices
    via matrix-vector products (O(n × p × k) cost), works with sparse input.
    Returns right singular vectors as (n_features, n_components).
    """
    _print = print if verbose else (lambda *a, **kw: None)
    _print(f"PCA initialization: randomized_svd for {n_components} components...")
    
    n_samples, n_features = input_matrix.shape
    
    if sparse.issparse(input_matrix):
        from scipy.sparse.linalg import LinearOperator
        
        mean_vec = global_mean.astype(np.float64)
        
        def centered_matvec(v):
            """Compute (X - 1μᵀ) @ v without materializing centered matrix."""
            result = np.zeros(n_samples, dtype=np.float64)
            for start in range(0, n_samples, chunk_size):
                end_idx = min(start + chunk_size, n_samples)
                batch = _safe_to_dense(input_matrix[start:end_idx]).astype(np.float64)
                result[start:end_idx] = batch @ v
            result -= mean_vec @ v
            return result
        
        def centered_rmatvec(u):
            """Compute (X - 1μᵀ)ᵀ @ u without materializing centered matrix."""
            result = np.zeros(n_features, dtype=np.float64)
            for start in range(0, n_samples, chunk_size):
                end_idx = min(start + chunk_size, n_samples)
                batch = _safe_to_dense(input_matrix[start:end_idx]).astype(np.float64)
                result += batch.T @ u[start:end_idx]
            result -= mean_vec * u.sum()
            return result
        
        X_centered_op = LinearOperator(
            shape=(n_samples, n_features),
            matvec=centered_matvec,
            rmatvec=centered_rmatvec,
            dtype=np.float64
        )
        
        U, S, Vt = randomized_svd(X_centered_op, n_components=n_components,
                                   random_state=random_state, n_iter=5)
    else:
        X_centered = input_matrix.astype(np.float64) - global_mean
        U, S, Vt = randomized_svd(X_centered, n_components=n_components,
                                   random_state=random_state, n_iter=5)
    
    W_init = Vt.T.astype(np.float64)
    
    if n_components == 1:
        _print(f"  Top singular value: [{S[0]:.2f}]")
    else:
        _print(f"  Top {n_components} singular values: "
               f"[{S[0]:.2f}, {S[1]:.2f}, ..., {S[-1]:.2f}]")
    
    return W_init


def _init_random_normalized(n_features, n_components, random_state,
                            verbose=True):
    """Random orthonormal initialization (unit-norm columns via QR).

    Columns are orthonormal, matching the scale of the PCA init
    (``_init_pca`` returns orthonormal ``Vt.T``), so PCA and random restarts
    start on the same footing. The tied-weight reconstruction ``X_c W Wᵀ`` is
    a true projection only when ``W`` is orthonormal, so no data-scale
    (MAD) rescaling is applied.
    """
    _print = print if verbose else (lambda *a, **kw: None)
    _print("Random orthonormal initialization: QR (unit columns)...")

    rng = np.random.default_rng(random_state)
    R = rng.standard_normal(size=(n_features, n_components))
    Q, _ = np.linalg.qr(R)
    W_init = Q.astype(np.float64)

    _print(f"  Orthonormal columns: {n_components} of dim {n_features}")

    return W_init


def _validate_custom_init(W_init, n_features, n_components):
    """Validate user-provided initialization array."""
    W_init = np.asarray(W_init, dtype=np.float64)
    if W_init.shape != (n_features, n_components):
        raise ValueError(
            f"Custom init shape {W_init.shape} does not match expected "
            f"({n_features}, {n_components}). Ensure init array has shape "
            f"(n_features, n_components)."
        )
    if not np.all(np.isfinite(W_init)):
        raise ValueError("Custom init contains non-finite values (NaN or Inf).")
    return W_init


# -----------------------------------------------------------------------------
# 5. Minibatch Generator
# -----------------------------------------------------------------------------

def minibatch_generator(input_matrix, batch_size, shuffle=True, seed=None):
    """Yield dense float64 minibatches from a (possibly sparse) matrix."""
    n_samples = input_matrix.shape[0]
    indices = np.arange(n_samples)
    if shuffle:
        rng = np.random.default_rng(seed)
        rng.shuffle(indices)
    for start_idx in range(0, n_samples, batch_size):
        end_idx = min(start_idx + batch_size, n_samples)
        batch_indices = indices[start_idx:end_idx]
        X_batch = _safe_to_dense(input_matrix[batch_indices]).astype(np.float64)
        yield X_batch


# -----------------------------------------------------------------------------
# 6. JAX Training Step
# -----------------------------------------------------------------------------

@partial(jax.jit, static_argnames=['optimizer', 'is_feature_mode',
                                   'use_robust_weighting', 'use_ema_scale'])
def train_step(params, opt_state, X_batch, global_mean,
               lambda_sparsity, lambda_ortho, lambda_score,
               sigma_sq_robust, epsilon_sq, epsilon_z_sq,
               annealing_factor, stability_eps,
               pca_recon_loss, score_scale_ema,
               optimizer, is_feature_mode, use_robust_weighting,
               use_ema_scale):
    """Single training step: reconstruction + W sparsity + ortho + Z sparsity.

    Loss = recon / σ²_robust
         + λ_sparsity × L_pca × mean(log(1 + W̃²/ε_W²)) × annealing
         + λ_ortho    × L_pca × mean((W_norm.T @ W_norm - I)²)
         + λ_score    × L_pca × mean(log(1 + Z̃²/ε_z²)) × annealing      ← NEW

    where Z = X_c·W are the projections (scores) and Z̃ is column-normalized
    (unit-L2 columns over the batch) so the penalty is scale-free and
    size-independent, mirroring the W penalty. ε_z is Otsu-adaptive, floored
    at 1/√b. The smooth log penalty gives intrinsic adaptive-lasso behavior.

    The score normalizer is either the per-batch column norm (default) or,
    when use_ema_scale is True, sqrt(b) × score_scale_ema (a running estimate
    of each component's per-entry score scale — lower variance than per-batch).

    pca_recon_loss (L_pca) scales every penalty to the recon magnitude.

    Aux returns the per-batch RMS score scale (z_rms) so the caller can update
    the EMA.
    """

    def loss_fn(p):
        n_features, n_components = p.shape
        X_centered = X_batch - global_mean
        scores = jnp.dot(X_centered, p)
        residuals = X_centered - jnp.dot(scores, p.T)

        # --- RECONSTRUCTION (normalized by robust variance) ---
        if use_robust_weighting:
            sample_errors = jnp.sqrt(jnp.mean(residuals**2, axis=1) + stability_eps)
            # standardize the per-sample residual by the robust scale so the
            # tanh (log-cosh) influence knee sits at ~1 robust sigma, not at a
            # data-scale-dependent absolute residual of ~1
            u = sample_errors / jnp.sqrt(sigma_sq_robust)
            raw_weights = jnp.tanh(u) / u
            batch_weights = jax.lax.stop_gradient(raw_weights / jnp.mean(raw_weights))
            mse_loss = jnp.mean((residuals ** 2) * batch_weights[:, None])
        else:
            mse_loss = jnp.mean(residuals ** 2)
        recon_loss = mse_loss / sigma_sq_robust

        # --- COLUMN NORMALIZATION ---
        col_norms = jnp.sqrt(jnp.sum(p**2, axis=0, keepdims=True) + stability_eps)
        p_normalized = p / col_norms

        col_norms_sg = jax.lax.stop_gradient(col_norms)
        p_direction = p / col_norms_sg

        # --- W SPARSITY (log penalty with adaptive ε via Otsu) ---
        if is_feature_mode:
            row_norms = jnp.sqrt(jnp.sum(p_direction**2, axis=1) + stability_eps)
            sparsity_loss = jnp.mean(
                log_sparsity_penalty(row_norms, epsilon_sq)
            ) * annealing_factor
        else:
            sparsity_loss = jnp.mean(
                log_sparsity_penalty(p_direction, epsilon_sq)
            ) * annealing_factor

        # --- SCORE SPARSITY (log penalty on column-normalized projections) ---
        b = scores.shape[0]
        z_col_norms = jnp.sqrt(jnp.sum(scores**2, axis=0, keepdims=True)
                               + stability_eps)            # (1, k), unit-L2 scale
        z_rms = z_col_norms[0] / jnp.sqrt(b)               # (k,), per-entry scale
        if use_ema_scale:
            z_scale = jax.lax.stop_gradient(
                score_scale_ema[None, :] * jnp.sqrt(b) + stability_eps)
        else:
            z_scale = jax.lax.stop_gradient(z_col_norms)
        Z_tilde = scores / z_scale
        score_sparsity_loss = jnp.mean(
            log_sparsity_penalty(Z_tilde, epsilon_z_sq)
        ) * annealing_factor

        # --- ORTHOGONALITY (on normalized columns) ---
        gram_diff = jnp.dot(p_normalized.T, p_normalized) - jnp.eye(n_components, dtype=p.dtype)
        ortho_loss = jnp.mean(gram_diff**2)

        total_loss = (recon_loss
                      + lambda_sparsity * pca_recon_loss * sparsity_loss
                      + lambda_score    * pca_recon_loss * score_sparsity_loss
                      + lambda_ortho    * pca_recon_loss * ortho_loss)
        return total_loss, (recon_loss, sparsity_loss, ortho_loss,
                            score_sparsity_loss, z_rms)

    (loss, metrics), grads = jax.value_and_grad(loss_fn, has_aux=True)(params)
    updates, new_opt_state = optimizer.update(grads, opt_state, params)
    new_params = optax.apply_updates(params, updates)

    return new_params, new_opt_state, loss, metrics


# Memoized compiled scan runners. The optax optimizer is a fresh (stateless)
# object each fit, so passing it as a jit static arg would miss the cache and
# recompile the (expensive) scan every fit. We instead build one jitted runner
# per training CONFIG, closing over the optimizer, and reuse it across all fits
# with that config — opt_state (the only per-fit state) is passed as an arg.
_SCAN_FN_CACHE = {}


def get_scan_fn(optimizer, is_feature_mode, robust_weighting, use_ema_scale,
                sparsity_warmup_steps, cache_key):
    """Return a cached jitted runner that advances n_steps via lax.scan.

    Fast path for the full-batch, no-patience, no-history case: one device
    dispatch runs n_steps optimizer steps instead of one Python->device round
    trip per step (the dispatch overhead that dominates tiny fits). epsilon_sq /
    epsilon_z_sq are constant within a chunk (Otsu updates run on the host
    between chunks), the per-step annealing is recomputed from the absolute step
    index to match cosine_annealing(step, sparsity_warmup_steps) bit-for-bit
    (divide first, then multiply by pi), and the optax schedule advances through
    opt_state so the LR schedule is preserved.
    """
    cached = _SCAN_FN_CACHE.get(cache_key)
    if cached is not None:
        return cached

    warm = float(sparsity_warmup_steps)

    @partial(jax.jit, static_argnames=('n_steps',))
    def run(params, opt_state, ema_vec, X_batch, global_mean, start_step,
            n_steps, lambda_sparsity, lambda_ortho, lambda_score,
            sigma_sq_robust, epsilon_sq, epsilon_z_sq, stability_eps,
            pca_recon_loss, score_beta):

        def body(carry, i):
            p, ostate, ema = carry
            step = start_step + i
            progress = step.astype(jnp.float64) / warm
            anneal = jnp.where(
                step >= sparsity_warmup_steps, jnp.float64(1.0),
                0.5 * (1.0 - jnp.cos(jnp.pi * progress)))
            new_p, new_ostate, loss, (r, s, o, sc, zrms) = train_step(
                p, ostate, X_batch, global_mean,
                lambda_sparsity, lambda_ortho, lambda_score,
                sigma_sq_robust, epsilon_sq, epsilon_z_sq,
                anneal, stability_eps, pca_recon_loss, ema,
                optimizer, is_feature_mode, robust_weighting, use_ema_scale)
            if use_ema_scale:
                ema = score_beta * ema + (1.0 - score_beta) * zrms
            return (new_p, new_ostate, ema), loss

        (params, opt_state, ema_vec), losses = jax.lax.scan(
            body, (params, opt_state, ema_vec), jnp.arange(n_steps))
        return params, opt_state, ema_vec, losses

    _SCAN_FN_CACHE[cache_key] = run
    return run


# -----------------------------------------------------------------------------
# 6b. PCA Baseline Reconstruction Loss
# -----------------------------------------------------------------------------

def compute_pca_recon_loss(pca_components, input_matrix, global_mean,
                           sigma_sq_robust, epsilon=1e-12,
                           batch_size=5000, n_batches=5, seed=42,
                           robust_weighting=True):
    """Compute reconstruction loss of PCA components under the training loss.
    
    Evaluates the exact same loss_fn used in train_step (same robust sample
    weighting, same sigma_sq_robust normalization) on PCA components before
    any training. Used to set the scale for sparsity and ortho penalties.
    """
    n_samples = input_matrix.shape[0]
    rng = np.random.default_rng(seed)
    
    p = jax.device_put(jnp.array(pca_components, dtype=jnp.float64))
    mean_jax = jax.device_put(global_mean)
    sigma_jax = jnp.float64(sigma_sq_robust)
    
    @jax.jit
    def recon_loss_robust(p, X_batch_jax):
        X_centered = X_batch_jax - mean_jax
        scores = jnp.dot(X_centered, p)
        residuals = X_centered - jnp.dot(scores, p.T)
        sample_errors = jnp.sqrt(jnp.mean(residuals**2, axis=1) + epsilon)
        # standardize by the robust scale so the tanh knee matches train_step
        u = sample_errors / jnp.sqrt(sigma_jax)
        raw_weights = jnp.tanh(u) / u
        batch_weights = jax.lax.stop_gradient(raw_weights / jnp.mean(raw_weights))
        mse_loss = jnp.mean((residuals**2) * batch_weights[:, None])
        return mse_loss / sigma_jax
    
    @jax.jit
    def recon_loss_plain(p, X_batch_jax):
        X_centered = X_batch_jax - mean_jax
        scores = jnp.dot(X_centered, p)
        residuals = X_centered - jnp.dot(scores, p.T)
        mse_loss = jnp.mean(residuals**2)
        return mse_loss / sigma_jax
    
    loss_fn = recon_loss_robust if robust_weighting else recon_loss_plain
    
    losses = []
    for _ in range(n_batches):
        idx = rng.choice(n_samples, size=min(batch_size, n_samples), replace=False)
        X_batch = _safe_to_dense(input_matrix[idx]).astype(np.float64)
        X_jax = jax.device_put(X_batch)
        losses.append(float(loss_fn(p, X_jax)))
    
    return float(np.mean(losses))


# -----------------------------------------------------------------------------
# 7. Estimator Class
# -----------------------------------------------------------------------------

class SparseProjectionPCA(BaseEstimator, TransformerMixin):
    """Sparse-projection PCA: a smooth penalty that sparsifies the *scores*.

    A sibling of ROSS-PCA (SmoothSparsePCA). The reconstruction stays centered
    PCA — minimizing ‖X_c − X_c W Wᵀ‖² with orthonormal W maximizes captured
    variance — but a smooth log penalty is added on the projections
    Z = X_c·W so that each component's scores are sparse (each factor active
    for only a few samples). The aim is interpretability: component j ↔ a small
    set of high-scoring samples.

    The score penalty mirrors the W penalty's dimensionless construction:
    log(1 + Z̃²/ε_z²) on column-normalized Z̃ (unit-L2 columns over the batch,
    so scale-free), averaged over entries (size-free), with ε_z Otsu-adaptive
    floored at 1/√b, and ×pca_recon_loss for commensurability.

    Orthogonality on W is retained as the identifiability anchor: with
    orthonormal W, reconstruction depends only on the subspace W Wᵀ, so
    sparsifying Z is a rotation of the basis within the principal subspace —
    nearly free in reconstruction. W-sparsity (lambda_sparsity) defaults to 0;
    lambda_score is the primary knob.

    Exact sparsity (hard zeros / per-component active sets) comes from a
    pruning step on Z afterward — see ross_proj.cpp_z (Cosine-Preserving
    Pruning of Zᵀ, reconstructed as Z_p Wᵀ). Training shrinks; pruning zeros.

    Parameters
    ----------
    n_components : int, default=100
        Number of components to extract.
    
    init : str or ndarray, default='pca'
        Initialization strategy:
        - 'pca': Truncated SVD via randomized_svd (Halko et al. 2009).
        - 'random_normalized': QR-orthonormalized random matrix scaled by
          median(MAD) × 1.4826.
        - ndarray of shape (n_features, n_components): Custom initialization.
    
    sparsity_method : str, default='element'
        - 'element': Penalize each loading entry independently via
          log(1 + x²/ε²) on column-normalized entries.
        - 'feature': Group lasso on row norms of column-normalized W,
          encouraging entire features to drop out.
    
    lambda_sparsity : float, default=0.0
        Weight of the sparsity penalty on the loadings W. Off by default in
        this variant (sparsity is targeted on the scores, not W); raise it for
        a doubly-sparse model.

    lambda_score : float, default=1.0
        Weight of the sparsity penalty on the projections Z = X_c·W. The
        primary knob of this method. Dimension-independent, scaled by
        pca_recon_loss like the other terms.

    score_scale_ema : float or None, default=None
        If None, Z is column-normalized per batch. If a decay in [0,1) (e.g.
        0.99), each component's score scale is tracked by an exponential moving
        average across batches (lower variance than the per-batch estimate),
        used as the normalizer. Set this if the per-batch normalization makes
        training jittery.

    lambda_ortho : float, default=0.1
        Orthogonality penalty weight (the identifiability anchor). Computed on
        column-normalized W.

    batch_size : int, default=10000
        Number of samples per gradient step.
    
    total_steps : int, default=5000
        Total number of gradient steps.
    
    sparsity_warmup_steps : int, default=2500
        Steps over which sparsity penalty is cosine-annealed from 0 to full.
        During warmup, ε is fixed at 1/√p. After warmup, ε is updated via
        Otsu at each epsilon_update_steps checkpoint.
    
    epsilon_update_steps : int, default=500
        Update ε via Otsu every this many steps. Replaces reweight_steps.
        During warmup (before sparsity_warmup_steps), ε stays fixed at 1/√p.
    
    patience : int, default=10
        After the scheduled total_steps, continue training at lr_max/10
        until neither Gini nor reconstruction improves for this many
        consecutive checkpoints (each checkpoint = epsilon_update_steps).
        Set to 0 to disable continuation (stop at total_steps).
    
    min_delta_gini_rel : float, default=0.005
        Minimum relative improvement in Gini to count as progress.
        Improvement = (gini_new - gini_best) / gini_best.
    
    min_delta_recon_rel : float, default=0.005
        Minimum relative improvement in reconstruction loss to count as
        progress. Improvement = (recon_best - recon_new) / recon_best.
    
    lr_max : float, default=0.005
        Peak learning rate. LR warmup starts at lr_max/100, cosine decay
        ends at lr_max/10. This single parameter controls the entire LR
        schedule.
    
    lr_warmup_steps : int or None, default=None
        Steps for LR linear warmup from lr_max/100 to lr_max. If None,
        defaults to sparsity_warmup_steps // 2.
    
    weight_decay : float, default=1e-4
        AdamW weight decay.
    
    grad_clip_norm : float, default=1.0
        Global gradient norm clipping.
    
    log_interval : int, default=100
        Print lightweight loss summary every this many steps. 0 to disable.
    
    pca_components : ndarray of shape (n_features, n_components) or None
        Reference PCA loading matrix (columns are components) used only to
        calibrate the penalty scale: lambda_sparsity / lambda_ortho are scaled
        by this baseline's reconstruction loss (pca_recon_loss_) so they carry
        consistent meaning across datasets. This is separate from `init`.
        If None, fit() computes a PCA internally for calibration
        ("self-calibration"); provide it to reuse a PCA you already have (e.g.
        a fixed reference, or to skip recomputation on large data). When
        init='pca' and pca_components is given, the same matrix is reused as
        the initialization.
    
    stability_eps : float, default=1e-12
        Small constant for numerical stability.
    
    subsample_fraction : float, default=0.05
        Fraction of data subsampled for MAD computation.
    
    random_state : int, default=42
        Random seed for reproducibility.
    
    Attributes
    ----------
    components_ : ndarray of shape (n_components, n_features)
        Learned sparse loading matrix. sklearn convention.
    
    mean_ : ndarray of shape (n_features,)
        Per-feature mean of training data.
    
    sigma_sq_robust_ : float
        Robust variance estimate (median nonzero MAD × 1.4826)².
    
    mad_per_feature_ : ndarray of shape (n_features,)
        Per-feature MAD values from training data.
    
    pca_recon_loss_ : float
        Reconstruction loss of PCA components under training loss.
    
    epsilon_history_ : list of (int, float)
        (step, ε) pairs at each Otsu update for diagnostics.
    """
    
    def __init__(self, n_components=100, init='pca', sparsity_method='element',
                 lambda_sparsity=0.0, lambda_ortho=0.1,
                 lambda_score=1.0, score_scale_ema=None,
                 batch_size=10000, total_steps=5000,
                 sparsity_warmup_steps=2500, epsilon_update_steps=500,
                 patience=5, patience_check_steps=100, max_continuation_steps=5000,
                 min_delta_gini_rel=0.005, min_delta_recon_rel=0.005,
                 lr_max=0.005, lr_warmup_steps=None,
                 weight_decay=1e-4, grad_clip_norm=1.0,
                 stability_eps=1e-12, subsample_fraction=0.05, random_state=42,
                 log_interval=100, pca_components=None, verbose=True,
                 record_history=False, robust_weighting=True, precision=64,
                 scan=True):

        self.n_components = n_components
        self.init = init
        self.sparsity_method = sparsity_method
        self.lambda_sparsity = lambda_sparsity
        self.lambda_ortho = lambda_ortho
        self.lambda_score = lambda_score
        self.score_scale_ema = score_scale_ema
        self.batch_size = batch_size
        self.total_steps = total_steps
        self.sparsity_warmup_steps = sparsity_warmup_steps
        self.epsilon_update_steps = epsilon_update_steps
        self.patience = patience
        self.patience_check_steps = patience_check_steps
        self.max_continuation_steps = max_continuation_steps
        self.min_delta_gini_rel = min_delta_gini_rel
        self.min_delta_recon_rel = min_delta_recon_rel
        self.lr_max = lr_max
        self.lr_warmup_steps = lr_warmup_steps
        self.weight_decay = weight_decay
        self.grad_clip_norm = grad_clip_norm
        self.stability_eps = stability_eps
        self.subsample_fraction = subsample_fraction
        self.random_state = random_state
        self.log_interval = log_interval
        self.pca_components = pca_components
        self.verbose = verbose
        self.record_history = record_history
        self.robust_weighting = robust_weighting
        if precision not in (32, 64):
            raise ValueError(f"precision must be 32 or 64, got {precision}")
        self.precision = precision
        self._np_dtype = np.float32 if precision == 32 else np.float64
        self._jnp_dtype = jnp.float32 if precision == 32 else jnp.float64
        self.scan = scan
        self.components_ = None
        self.mean_ = None
        self.sigma_sq_robust_ = None
        self.mad_per_feature_ = None
        self.pca_recon_loss_ = None
        self.epsilon_history_ = []
        self.epsilon_z_history_ = []
        self.history_ = None

    def _compute_epsilon(self, params, n_features, is_feature):
        """Compute ε via Otsu on |W̃| entries (or row norms for feature mode).
        
        Floored at 1/√p (the initial value) to prevent ε from collapsing
        below the natural scale of direction matrix entries. If Otsu returns
        a smaller threshold, the distribution hasn't separated and the
        initial scale is still the best estimate.
        
        Returns:
            epsilon: max(Otsu threshold, 1/√p).
        """
        col_norms = jnp.sqrt(
            jnp.sum(params**2, axis=0, keepdims=True) + self.stability_eps)
        p_direction = params / col_norms
        
        if is_feature:
            row_norms = jnp.sqrt(
                jnp.sum(p_direction**2, axis=1) + self.stability_eps)
            values = np.array(row_norms)
        else:
            values = np.abs(np.array(p_direction))
        
        eps_otsu = otsu_threshold(values)
        eps_floor = 1.0 / np.sqrt(n_features)
        eps = max(eps_otsu, eps_floor)

        return eps

    def _compute_epsilon_z(self, params, X_batch, global_mean, score_scale=None):
        """Compute ε_z via Otsu on |Z̃| (column-normalized projections).

        Z = (X_batch - mean) @ params; columns are normalized by their L2 norm
        over the batch (or by sqrt(b)·score_scale if an EMA scale is given), so
        |Z̃| entries are scale-free. Floored at 1/√b, mirroring _compute_epsilon.

        Returns:
            epsilon_z: max(Otsu threshold on |Z̃|, 1/√b).
        """
        X_c = np.asarray(X_batch, dtype=np.float64) - np.asarray(global_mean)
        scores = X_c @ np.asarray(params, dtype=np.float64)
        b = scores.shape[0]
        if score_scale is not None:
            z_scale = np.asarray(score_scale) * np.sqrt(b) + self.stability_eps
        else:
            z_scale = np.sqrt(np.sum(scores**2, axis=0) + self.stability_eps)
        z_tilde = scores / z_scale[None, :]
        eps_otsu = otsu_threshold(np.abs(z_tilde))
        eps_floor = 1.0 / np.sqrt(b)
        return max(eps_otsu, eps_floor)

    def _log(self, msg, **kwargs):
        """Print only if verbose is True."""
        if self.verbose:
            print(msg, **kwargs)

    def fit(self, input_matrix):
        import resource
        
        def _rss_gb():
            try:
                return resource.getrusage(resource.RUSAGE_SELF).ru_maxrss / 1e6
            except Exception:
                return float('nan')
        
        def _mem(label):
            if self.verbose:
                print(f"  [MEM] {label}: RSS={_rss_gb():.2f} GB", flush=True)
        
        _log = self._log
        tqdm = _tqdm if self.verbose else _noop_tqdm
        n_samples, n_features = input_matrix.shape
        k = self.n_components
        
        _log(f"\n=== SparseProjectionPCA fit: {n_samples:,} × {n_features:,}, "
             f"k={k}, λ_score={self.lambda_score} ===", flush=True)
        
        params_gb = n_features * k * 8 / 1e9
        adam_gb = params_gb * 2
        _log(f"  Predicted allocations: params={params_gb:.2f} GB, "
             f"Adam moments={adam_gb:.2f} GB "
             f"(total ~{params_gb + adam_gb:.2f} GB)", flush=True)
        _mem("start of fit")
        
        # --- Step 1: Global mean + robust scale ---
        if self.mean_ is None or self.sigma_sq_robust_ is None:
            _log("\n[Step 1] Computing global mean + robust scale...", flush=True)
            self.mean_, self.sigma_sq_robust_, self.mad_per_feature_ = \
                compute_global_mean_and_scale(
                    input_matrix, subsample_fraction=self.subsample_fraction,
                    seed=self.random_state, verbose=self.verbose
                )
        else:
            _log("\n[Step 1] Using cached mean + robust scale.", flush=True)
        _mem("after mean+MAD")
        
        global_mean_jax = jax.device_put(self.mean_.astype(self._np_dtype))
        sigma_sq_jax = self._jnp_dtype(self.sigma_sq_robust_)
        _mem("after jax.device_put(mean)")
        
        # --- Step 1b: PCA components for calibration ---
        # Always compute PCA for the calibration constant pca_recon_loss.
        # This is a property of the data, not the initialization strategy.
        # If pca_components was provided externally, use those instead.
        if self.pca_components is not None:
            pca_W = np.asarray(self.pca_components, dtype=np.float64)
            if pca_W.shape != (n_features, self.n_components):
                raise ValueError(
                    f"pca_components shape {pca_W.shape} != "
                    f"({n_features}, {self.n_components})"
                )
            _log("[Step 1b] Using provided pca_components for calibration.",
                  flush=True)
        else:
            _log("[Step 1b] Computing PCA for calibration...", flush=True)
            pca_W = _init_pca(input_matrix, self.mean_,
                              self.n_components, self.random_state,
                              verbose=self.verbose)
        
        self.pca_recon_loss_ = compute_pca_recon_loss(
            pca_W, input_matrix, self.mean_, self.sigma_sq_robust_,
            epsilon=self.stability_eps, batch_size=self.batch_size,
            seed=self.random_state,
            robust_weighting=self.robust_weighting
        )
        _log(
            f"  PCA baseline recon loss: {self.pca_recon_loss_:.4f} "
            f"(sparsity and ortho scaled by this value)",
            flush=True
        )
        pca_recon_loss_jax = self._jnp_dtype(self.pca_recon_loss_)
        _mem("after PCA calibration")
        
        # --- Step 2: Initialization ---
        _log(f"\n[Step 2] Initialization: "
              f"{self.init if isinstance(self.init, str) else 'custom array'}...",
              flush=True)
        if isinstance(self.init, str):
            if self.init == 'pca':
                W_init = pca_W  # reuse PCA components already computed
                _log("  Reusing PCA components from calibration step.",
                      flush=True)
            elif self.init == 'random_normalized':
                W_init = _init_random_normalized(
                    n_features, self.n_components,
                    self.random_state, verbose=self.verbose
                )
            else:
                raise ValueError(
                    f"Unknown init '{self.init}'. "
                    f"Use 'pca', 'random_normalized', or a custom array."
                )
        else:
            W_init = _validate_custom_init(self.init, n_features,
                                           self.n_components)
        _mem("after W_init computed")
        
        _log(f"  Sending params to JAX device ({params_gb:.2f} GB)...",
              flush=True)
        params = jax.device_put(jnp.array(W_init, dtype=self._jnp_dtype))
        del W_init
        _mem("after params on device")
        
        # --- Step 3: Initial ε = 1/√p ---
        epsilon = 1.0 / np.sqrt(n_features)
        epsilon_sq = self._jnp_dtype(epsilon ** 2)
        self.epsilon_history_ = [(0, epsilon)]
        _log(f"\n[Step 3] Initial ε = 1/√p = {epsilon:.6f}", flush=True)

        # --- Step 3b: Score-penalty ε_z and (optional) EMA score scale ---
        epsilon_z = 1.0 / np.sqrt(self.batch_size)
        epsilon_z_sq = self._jnp_dtype(epsilon_z ** 2)
        self.epsilon_z_history_ = [(0, epsilon_z)]
        use_ema_scale = self.score_scale_ema is not None
        if use_ema_scale:
            # Seed the EMA from the initial projection on a sample batch.
            seed_idx = np.random.default_rng(self.random_state).choice(
                n_samples, size=min(self.batch_size, n_samples), replace=False)
            X_seed = _safe_to_dense(input_matrix[seed_idx]).astype(np.float64)
            Z_seed = (X_seed - self.mean_) @ np.asarray(params)
            score_scale_ema_vec = jnp.asarray(
                np.sqrt(np.mean(Z_seed**2, axis=0) + self.stability_eps),
                dtype=self._jnp_dtype)
            _log(f"[Step 3b] Initial ε_z = 1/√b = {epsilon_z:.6f} | "
                 f"EMA score scale on (β={self.score_scale_ema})", flush=True)
        else:
            score_scale_ema_vec = jnp.zeros(self.n_components, dtype=self._jnp_dtype)
            _log(f"[Step 3b] Initial ε_z = 1/√b = {epsilon_z:.6f} | "
                 f"per-batch score scale", flush=True)

        # --- Step 4: LR schedule + Optimizer ---
        _log(f"\n[Step 4] Building optimizer + Adam state (~{adam_gb:.2f} GB)...",
              flush=True)
        lr_warmup_steps = (self.lr_warmup_steps
                           if self.lr_warmup_steps is not None
                           else self.sparsity_warmup_steps // 2)
        
        # Two-phase LR schedule derived from lr_max alone:
        #   Phase 1 [0, lr_warmup_steps]:        linear warmup lr_max/100 → lr_max
        #   Phase 2 [lr_warmup_steps, total]:     cosine decay lr_max → lr_max/10
        lr_warmup_start = self.lr_max / 100
        lr_decay_floor = self.lr_max / 10
        decay_steps = max(1, self.total_steps - lr_warmup_steps)
        
        phase1 = optax.linear_schedule(
            init_value=lr_warmup_start,
            end_value=self.lr_max,
            transition_steps=lr_warmup_steps,
        )
        phase2 = optax.cosine_decay_schedule(
            init_value=self.lr_max,
            decay_steps=decay_steps,
            alpha=lr_decay_floor / self.lr_max,  # = 0.1
        )
        lr_schedule = optax.join_schedules(
            schedules=[phase1, phase2],
            boundaries=[lr_warmup_steps],
        )
        optimizer = optax.chain(
            optax.clip_by_global_norm(self.grad_clip_norm),
            optax.adamw(learning_rate=lr_schedule,
                        weight_decay=self.weight_decay),
        )
        opt_state = optimizer.init(params)
        _mem("after optimizer.init")
        
        _log(f"\n[Step 5] Training configuration:", flush=True)
        _log(f"  LR schedule: warmup {lr_warmup_steps} steps "
              f"({lr_warmup_start:.2e} → {self.lr_max:.2e}), "
              f"cosine decay over {decay_steps} steps "
              f"→ {lr_decay_floor:.2e}")
        _log(f"  Sparsity warmup: {self.sparsity_warmup_steps} steps")
        _log(f"  ε update every: {self.epsilon_update_steps} steps "
              f"(Otsu, after warmup)")
        _log(f"  Grad clip norm: {self.grad_clip_norm}")
        _log(f"  Log interval:   {self.log_interval} steps | "
              f"Batch size: {self.batch_size}")
        if self.patience > 0:
            _log(f"  Patience: {self.patience} checkpoints after schedule "
                  f"(δ_gini={self.min_delta_gini_rel:.3f}, "
                  f"δ_recon={self.min_delta_recon_rel:.3f})")
        else:
            _log(f"  Patience: disabled (stop at total_steps)")
        _log(f"  PCA recon loss: {self.pca_recon_loss_:.4f} "
              f"(sparsity/ortho scale factor)")
        
        # Gini at initialization
        abs_p0 = jnp.abs(params)
        abs_sorted0 = jnp.sort(abs_p0, axis=0)
        n0 = n_features
        w0 = jnp.arange(1, n0 + 1, dtype=params.dtype)
        col_sums0 = jnp.sum(abs_sorted0, axis=0) + self.stability_eps
        gini0 = (2.0 * jnp.sum(w0[:, None] * abs_sorted0, axis=0)
                 / (n0 * col_sums0) - (n0 + 1.0) / n0)
        _log(f"  Gini at init: mean={float(jnp.mean(gini0)):.4f} "
              f"± {float(jnp.std(gini0)):.4f} "
              f"(target: increasing over training)", flush=True)
        
        # --- Step 5: Steps-based training loop ---
        is_feature = (self.sparsity_method == 'feature')
        
        if self.sparsity_method not in ('element', 'feature'):
            raise ValueError(
                f"Unknown sparsity_method '{self.sparsity_method}'. "
                f"Use 'element' or 'feature'."
            )

        # --- Step 5a: fast full-batch path (one dispatch per chunk) ---
        # When the whole dataset is a single batch and per-step history is off,
        # run the SCHEDULED phase with lax.scan (chunks between Otsu ε-updates)
        # instead of a Python loop — this removes the per-step Python->device
        # dispatch that dominates tiny fits. The patience CONTINUATION (a small
        # tail) then runs as a plain Python per-step loop, identical to the loop
        # below. The full per-step loop is kept unchanged for minibatch / history.
        use_scan = (self.scan and self.batch_size >= n_samples
                    and not self.record_history)
        if use_scan:
            X_full = jax.device_put(
                _safe_to_dense(input_matrix[0:n_samples]).astype(self._np_dtype))
            beta = (self._jnp_dtype(self.score_scale_ema)
                    if use_ema_scale else self._jnp_dtype(0.0))
            step0 = 0
            chunk = self.epsilon_update_steps
            # One compiled scan runner per training config, reused across every
            # fit sharing that config (so the scan is compiled once, not per fit).
            cache_key = (round(float(self.lr_max), 12), self.total_steps,
                         lr_warmup_steps, round(float(self.weight_decay), 12),
                         round(float(self.grad_clip_norm), 12), is_feature,
                         self.robust_weighting, use_ema_scale, self.precision,
                         self.sparsity_warmup_steps, n_features, self.n_components)
            scan_fn = get_scan_fn(optimizer, is_feature, self.robust_weighting,
                                  use_ema_scale, self.sparsity_warmup_steps,
                                  cache_key)
            _log(f"\n[Step 5] Fast full-batch scan: {self.total_steps} steps "
                 f"in chunks of {chunk}", flush=True)
            chunk_losses = None
            while step0 < self.total_steps:
                nstep = int(min(chunk, self.total_steps - step0))
                params, opt_state, score_scale_ema_vec, chunk_losses = scan_fn(
                    params, opt_state, score_scale_ema_vec, X_full,
                    global_mean_jax, step0, nstep,
                    self.lambda_sparsity, self.lambda_ortho, self.lambda_score,
                    sigma_sq_jax, epsilon_sq, epsilon_z_sq, self.stability_eps,
                    pca_recon_loss_jax, beta)
                step0 += nstep
                # Otsu ε / ε_z update at the ε-update boundaries (multiples of
                # epsilon_update_steps), after warmup — matching the loop path.
                if (step0 >= self.sparsity_warmup_steps
                        and step0 % self.epsilon_update_steps == 0):
                    epsilon = self._compute_epsilon(params, n_features, is_feature)
                    epsilon_sq = self._jnp_dtype(epsilon ** 2)
                    self.epsilon_history_.append((step0, epsilon))
                    ema_arg = (np.asarray(score_scale_ema_vec)
                               if use_ema_scale else None)
                    epsilon_z = self._compute_epsilon_z(
                        params, X_full, global_mean_jax, ema_arg)
                    epsilon_z_sq = self._jnp_dtype(epsilon_z ** 2)
                    self.epsilon_z_history_.append((step0, epsilon_z))
            global_step = step0

            # --- Step 5b: patience continuation, also via scan ---
            # Run patience_check_steps per scan dispatch, then decide patience on
            # the host from the chunk's per-step losses (mean over the window ==
            # the loop's mean over patience_loss_acc). Same logic as the loop's
            # continuation, but ~one dispatch per check instead of per step, so
            # patience adds almost nothing. ε-updates land exactly on their
            # boundaries when patience_check_steps divides epsilon_update_steps.
            if self.patience > 0:
                cont_opt = optax.chain(
                    optax.clip_by_global_norm(self.grad_clip_norm),
                    optax.adamw(learning_rate=lr_decay_floor,
                                weight_decay=self.weight_decay))
                opt_state = cont_opt.init(params)
                cont_scan = get_scan_fn(
                    cont_opt, is_feature, self.robust_weighting, use_ema_scale,
                    self.sparsity_warmup_steps, cache_key + ("cont",))
                best_loss = float(chunk_losses[-1])
                patience_counter = 0
                pcs = self.patience_check_steps
                next_epsilon_update = ((self.total_steps // self.epsilon_update_steps
                                        + 1) * self.epsilon_update_steps)
                _log(f"\n  === Continuation (scan chunks of {pcs}): "
                     f"patience={self.patience} (best {best_loss:.5f}) ===",
                     flush=True)
                while True:
                    params, opt_state, score_scale_ema_vec, closs = cont_scan(
                        params, opt_state, score_scale_ema_vec, X_full,
                        global_mean_jax, global_step, pcs,
                        self.lambda_sparsity, self.lambda_ortho, self.lambda_score,
                        sigma_sq_jax, epsilon_sq, epsilon_z_sq, self.stability_eps,
                        pca_recon_loss_jax, beta)
                    global_step += pcs
                    if (global_step >= next_epsilon_update
                            and global_step >= self.sparsity_warmup_steps):
                        epsilon = self._compute_epsilon(params, n_features, is_feature)
                        epsilon_sq = self._jnp_dtype(epsilon ** 2)
                        self.epsilon_history_.append((global_step, epsilon))
                        ema_arg = (np.asarray(score_scale_ema_vec)
                                   if use_ema_scale else None)
                        epsilon_z = self._compute_epsilon_z(
                            params, X_full, global_mean_jax, ema_arg)
                        epsilon_z_sq = self._jnp_dtype(epsilon_z ** 2)
                        self.epsilon_z_history_.append((global_step, epsilon_z))
                        next_epsilon_update += self.epsilon_update_steps
                    mean_loss = float(jnp.mean(closs))
                    improved = ((best_loss - mean_loss) / max(abs(best_loss), 1e-10)
                                > self.min_delta_recon_rel)
                    if improved:
                        patience_counter = 0
                        best_loss = mean_loss
                    else:
                        patience_counter += 1
                    if patience_counter >= self.patience:
                        _log(f"  [scan] early stop at {global_step} "
                             f"(best {best_loss:.5f})", flush=True)
                        break
                    if global_step - self.total_steps >= self.max_continuation_steps:
                        _log(f"  [scan] max continuation at {global_step}",
                             flush=True)
                        break

            self.n_steps_ = global_step
            self.components_ = np.array(params).T
            self.score_scale_ = (np.asarray(score_scale_ema_vec)
                                 if use_ema_scale else None)
            _log(f"  [scan] done: {global_step} steps, "
                 f"{len(self.epsilon_history_) - 1} ε-updates", flush=True)
            return self

        global_step = 0
        next_epsilon_update = self.epsilon_update_steps
        
        if self.record_history:
            self.history_ = []
        
        # Infinite minibatch stream
        def batch_stream():
            epoch = 0
            while True:
                gen = minibatch_generator(
                    input_matrix, self.batch_size, shuffle=True,
                    seed=self.random_state + epoch
                )
                for batch in gen:
                    yield batch
                epoch += 1
        
        stream = batch_stream()
        
        # Logging accumulators
        interval_losses   = []
        interval_recon    = []
        interval_sparsity = []
        interval_ortho    = []
        interval_score    = []

        log_losses   = []
        log_recon    = []
        log_sparsity = []
        log_ortho    = []
        log_score    = []

        # Patience tracking (used after scheduled phase)
        patience_counter = 0
        in_continuation = False
        next_patience_check = self.total_steps + self.patience_check_steps
        patience_loss_acc = []
        
        # Upper bound for progress bar (will extend if continuation kicks in)
        pbar = tqdm(total=self.total_steps, desc="Training (scheduled)")
        
        while True:
            X_batch = next(stream)
            X_batch_jax = jax.device_put(X_batch.astype(self._np_dtype))
            
            annealing_factor = cosine_annealing(global_step,
                                                self.sparsity_warmup_steps)
            
            params, opt_state, loss_val, (r_val, s_val, o_val, sc_val, zrms_val) = \
                train_step(
                    params, opt_state, X_batch_jax, global_mean_jax,
                    self.lambda_sparsity, self.lambda_ortho, self.lambda_score,
                    sigma_sq_jax, epsilon_sq, epsilon_z_sq,
                    annealing_factor, self.stability_eps,
                    pca_recon_loss_jax, score_scale_ema_vec,
                    optimizer, is_feature, self.robust_weighting,
                    use_ema_scale
                )

            # EMA update of the per-component score scale (if enabled)
            if use_ema_scale:
                beta = self.score_scale_ema
                score_scale_ema_vec = (beta * score_scale_ema_vec
                                       + (1.0 - beta) * zrms_val)

            global_step += 1
            # Append the raw device arrays (no float()) so JAX can dispatch
            # steps asynchronously instead of blocking on a host sync every
            # step. These accumulators are reduced (np.mean) only at the log /
            # ε-update checkpoints, which already sync. This is logging-only and
            # does not affect the fit.
            interval_losses.append(loss_val)
            interval_recon.append(r_val)
            interval_sparsity.append(s_val)
            interval_ortho.append(o_val)
            interval_score.append(sc_val)
            log_losses.append(loss_val)
            log_recon.append(r_val)
            log_sparsity.append(s_val)
            log_ortho.append(o_val)
            log_score.append(sc_val)
            
            # Per-step history (only if enabled)
            if self.record_history:
                current_lr_hist = float(lr_schedule(global_step)) if not in_continuation else lr_decay_floor
                self.history_.append({
                    "step": global_step,
                    "lr": current_lr_hist,
                    "annealing": float(annealing_factor),
                    "total_loss": float(loss_val),
                    "recon_loss": float(r_val),
                    "sparsity_loss_raw": float(s_val),
                    "score_loss_raw": float(sc_val),
                    "ortho_loss_raw": float(o_val),
                    "epsilon": epsilon,
                    "epsilon_z": epsilon_z,
                })
            
            if self.verbose:
                pbar.update(1)
                pbar.set_postfix({'Loss': f"{float(loss_val):.4f}"})
            
            # Lightweight log
            if self.log_interval > 0 and global_step % self.log_interval == 0:
                current_lr = float(lr_schedule(global_step)) if not in_continuation else lr_decay_floor
                pbar.clear()
                phase_tag = "cont" if in_continuation else "sched"
                _log(f"  Step {global_step} [{phase_tag}] | "
                      f"LR: {current_lr:.2e} | "
                      f"Anneal: {float(annealing_factor):.2f} | "
                      f"Total: {np.mean(log_losses):.5f} | "
                      f"Recon: {np.mean(log_recon):.5f} | "
                      f"Sparse_W: {np.mean(log_sparsity):.5f} | "
                      f"Sparse_Z: {np.mean(log_score):.5f} | "
                      f"Ortho: {np.mean(log_ortho):.5f} | "
                      f"ε: {epsilon:.6f} | ε_z: {epsilon_z:.6f} | "
                      f"BS: {self.batch_size}", flush=True)
                log_losses.clear()
                log_recon.clear()
                log_sparsity.clear()
                log_ortho.clear()
                log_score.clear()
            
            # ε update checkpoint
            if global_step == next_epsilon_update:
                pbar.clear()
                
                # Sparsity tracking
                col_norms_track = jnp.sqrt(
                    jnp.sum(params**2, axis=0, keepdims=True) + self.stability_eps)
                p_direction_track = params / col_norms_track
                if is_feature:
                    row_norms_dir = jnp.sqrt(
                        jnp.sum(p_direction_track**2, axis=1) + self.stability_eps)
                    sp_below_eps = float(jnp.mean(row_norms_dir < epsilon) * 100)
                    sp_below_half = float(jnp.mean(row_norms_dir < epsilon * 0.5) * 100)
                    sp_label = "rows"
                else:
                    abs_entries = jnp.abs(p_direction_track)
                    sp_below_eps = float(jnp.mean(abs_entries < epsilon) * 100)
                    sp_below_half = float(jnp.mean(abs_entries < epsilon * 0.5) * 100)
                    sp_label = "entries"
                
                # Gini coefficient
                abs_params = jnp.abs(params)
                abs_sorted = jnp.sort(abs_params, axis=0)
                n = n_features
                weights = jnp.arange(1, n + 1, dtype=params.dtype)
                col_sums = jnp.sum(abs_sorted, axis=0) + self.stability_eps
                gini_per_col = (
                    2.0 * jnp.sum(weights[:, None] * abs_sorted, axis=0)
                    / (n * col_sums)
                    - (n + 1.0) / n
                )
                gini_mean = float(jnp.mean(gini_per_col))
                gini_std  = float(jnp.std(gini_per_col))
                
                # Update ε via Otsu (only after sparsity warmup)
                if global_step >= self.sparsity_warmup_steps:
                    epsilon_new = self._compute_epsilon(
                        params, n_features, is_feature)
                    epsilon_old = epsilon
                    epsilon = epsilon_new
                    epsilon_sq = self._jnp_dtype(epsilon ** 2)
                    self.epsilon_history_.append((global_step, epsilon))
                    # Update ε_z via Otsu on |Z̃| using the current batch.
                    ema_arg = (np.asarray(score_scale_ema_vec)
                               if use_ema_scale else None)
                    epsilon_z_old = epsilon_z
                    epsilon_z = self._compute_epsilon_z(
                        params, X_batch_jax, global_mean_jax, ema_arg)
                    epsilon_z_sq = self._jnp_dtype(epsilon_z ** 2)
                    self.epsilon_z_history_.append((global_step, epsilon_z))
                    eps_msg = (f"ε: {epsilon_old:.6f} → {epsilon:.6f} | "
                               f"ε_z: {epsilon_z_old:.6f} → {epsilon_z:.6f} (Otsu)")
                else:
                    eps_msg = f"ε: {epsilon:.6f} | ε_z: {epsilon_z:.6f} (fixed, warmup)"

                current_lr = float(lr_schedule(global_step)) if not in_continuation else lr_decay_floor
                mean_recon = float(np.mean(interval_recon)) if interval_recon else float('nan')
                
                phase_tag = "cont" if in_continuation else "sched"
                patience_msg = ""
                if in_continuation:
                    patience_msg = f" | patience: {patience_counter}/{self.patience}"
                
                _log(f"  Step {global_step} [{phase_tag}] | "
                      f"Anneal: {float(annealing_factor):.2f} | "
                      f"LR: {current_lr:.2e} | "
                      f"Total: {np.mean(interval_losses):.5f} | "
                      f"Recon: {mean_recon:.5f} | "
                      f"Sparse_W: {np.mean(interval_sparsity):.5f} | "
                      f"Sparse_Z: {np.mean(interval_score):.5f} | "
                      f"Ortho: {np.mean(interval_ortho):.5f} | "
                      f"{sp_label} <ε: {sp_below_eps:.1f}% <ε/2: {sp_below_half:.1f}% | "
                      f"{eps_msg} | "
                      f"Gini: {gini_mean:.4f} ± {gini_std:.4f}"
                      f"{patience_msg}")

                interval_losses.clear()
                interval_recon.clear()
                interval_sparsity.clear()
                interval_ortho.clear()
                interval_score.clear()
                log_losses.clear()
                log_recon.clear()
                log_sparsity.clear()
                log_ortho.clear()
                log_score.clear()
                
                next_epsilon_update += self.epsilon_update_steps
            
            # --- Transition to continuation phase ---
            if global_step == self.total_steps and not in_continuation:
                if self.patience > 0:
                    in_continuation = True
                    # Switch to constant LR optimizer at decay floor
                    optimizer = optax.chain(
                        optax.clip_by_global_norm(self.grad_clip_norm),
                        optax.adamw(learning_rate=lr_decay_floor,
                                    weight_decay=self.weight_decay),
                    )
                    opt_state = optimizer.init(params)
                    
                    # Initialize best from recent loss
                    best_loss = float(np.mean(patience_loss_acc)) if patience_loss_acc else float(loss_val)
                    patience_counter = 0
                    patience_loss_acc = []
                    next_patience_check = global_step + self.patience_check_steps
                    
                    pbar.close()
                    pbar = tqdm(desc="Training (continuation)", unit="step")
                    _log(f"\n  === Continuation phase: LR={lr_decay_floor:.2e}, "
                          f"patience={self.patience} × {self.patience_check_steps} steps "
                          f"(best loss={best_loss:.5f}) ===", flush=True)
                else:
                    break
            
            # --- Patience check (separate from epsilon updates) ---
            if in_continuation:
                patience_loss_acc.append(loss_val)  # raw; reduced at the check
                if global_step >= next_patience_check:
                    mean_loss = float(np.mean(patience_loss_acc))
                    loss_improved = (best_loss - mean_loss) / max(abs(best_loss), 1e-10) > self.min_delta_recon_rel
                    
                    if loss_improved:
                        patience_counter = 0
                        best_loss = mean_loss
                    else:
                        patience_counter += 1
                    
                    _log(f"  Step {global_step} [cont] | "
                          f"mean_loss={mean_loss:.5f} "
                          f"(best={best_loss:.5f}) | "
                          f"patience: {patience_counter}/{self.patience}")
                    
                    patience_loss_acc = []
                    next_patience_check += self.patience_check_steps
                    
                    if patience_counter >= self.patience:
                        _log(f"\n  Early stopping: no improvement for "
                              f"{self.patience} × {self.patience_check_steps} steps "
                              f"(best loss={best_loss:.5f})")
                        break
                    
                    if global_step - self.total_steps >= self.max_continuation_steps:
                        _log(f"\n  Max continuation reached: "
                              f"{self.max_continuation_steps} steps "
                              f"(best loss={best_loss:.5f})")
                        break
        
        pbar.close()
        self.n_steps_ = global_step
        self.components_ = np.array(params).T  # (n_components, n_features) — sklearn convention
        self.score_scale_ = (np.asarray(score_scale_ema_vec)
                             if use_ema_scale else None)
        return self

    def transform(self, input_matrix):
        """Project data onto sparse components: (X - μ) @ Wᵀ. sklearn convention."""
        if self.components_ is None:
            raise RuntimeError("Model not fitted. Call fit() first.")
        n_samples = input_matrix.shape[0]
        projections = np.zeros((n_samples, self.n_components), dtype=np.float64)
        components_jax = jax.device_put(self.components_)
        mean_jax = jax.device_put(self.mean_)
        
        tqdm = _tqdm if self.verbose else _noop_tqdm
        for start in tqdm(range(0, n_samples, self.batch_size),
                         desc="Transform"):
            end = min(start + self.batch_size, n_samples)
            X_batch = _safe_to_dense(input_matrix[start:end]).astype(np.float64)
            X_centered = jax.device_put(X_batch) - mean_jax
            projections[start:end] = np.array(jnp.dot(X_centered, components_jax.T))
        return projections

    def inverse_transform(self, X_transformed, batch_size=None):
        """Reconstruct data from scores: Z @ W + μ. sklearn convention."""
        if self.components_ is None:
            raise RuntimeError("Model not fitted. Call fit() first.")
        n_samples = X_transformed.shape[0]
        n_features = self.components_.shape[1]
        batch_size = batch_size or self.batch_size
        
        try:
            X_recon = np.zeros((n_samples, n_features), dtype=np.float64)
        except MemoryError:
            raise MemoryError(
                f"Cannot allocate ({n_samples}, {n_features}) float64 array "
                f"for reconstruction. Use smaller batch_size or subsample."
            )
        
        components_jax = jax.device_put(self.components_)
        mean_jax = jax.device_put(self.mean_)
        
        tqdm = _tqdm if self.verbose else _noop_tqdm
        for start in tqdm(range(0, n_samples, batch_size), desc="Inverse"):
            end = min(start + batch_size, n_samples)
            Z_batch = X_transformed[start:end]
            Z_batch_jax = jax.device_put(Z_batch)
            recon_batch = jnp.dot(Z_batch_jax, components_jax) + mean_jax
            X_recon[start:end] = np.array(recon_batch)
        return X_recon
```
